# Inductive reasoning with large language models: a simulated randomized controlled trial for epilepsy

**DOI:** 10.1101/2024.03.18.24304493

**Authors:** Daniel M. Goldenholz, Shira R. Goldenholz, Sara Habib, M. Brandon Westover

**Affiliations:** Department of Neurology, Harvard Medical School, Boston USA; Department of Neurology, Beth Israel Deaconess Medical Center, Boston USA

**Keywords:** artificial intelligence, large language models, epilepsy, randomized clinical trials

## Abstract

**Importance:** The analysis of electronic medical records at scale to learn from clinical experience is currently very challenging. The integration of artificial intelligence (AI), specifically foundational large language models (LLMs), into an analysis pipeline may overcome some of the current limitations of modest input sizes, inaccuracies, biases, and incomplete knowledge bases.

**Objective:** To explore the effectiveness of using an LLM for generating realistic clinical data and other LLMs for summarizing and synthesizing information in a model system, simulating a randomized clinical trial (RCT) in epilepsy to demonstrate the potential of inductive reasoning via medical chart review.

**Design:** An LLM-generated simulated RCT based on a RCT for treatment with an anti-seizure medication, cenobamate, including a placebo arm and a full-strength drug arm, evaluated by an LLM-based pipeline versus a human reader.

**Setting:** Simulation based on realistic seizure diaries, treatment effects, reported symptoms and clinical notes generated by LLMs with multiple different neurologist writing styles.

**Participants:** Simulated cohort of 240 patients, divided 1:1 into placebo and drug arms.

**Intervention:** Utilization of LLMs for the generation of clinical notes and for the synthesis of data from these notes, aiming to evaluate the efficacy and safety of cenobamate in seizure control either with a human evaluator or AI-pipeline.

**Measures:** The AI and human analysis focused on identifying the number of seizures, symptom reports, and treatment efficacy, with statistical analysis comparing the 50%-responder rate and median percentage change between the placebo and drug arms, as well as side effect rates in each arm.

**Results:** AI closely mirrored human analysis, demonstrating the drug’s efficacy with marginal differences (<3%) in identifying both drug efficacy and reported symptoms.

**Conclusions and Relevance:** This study showcases the potential of LLMs accurately simulate and analyze clinical trials. Significantly, it highlights the ability of LLMs to reconstruct essential trial elements, identify treatment effects, and recognize reported symptoms, within a realistic clinical framework. The findings underscore the relevance of LLMs in future clinical research, offering a scalable, efficient alternative to traditional data mining methods without the need for specialized medical language training.

**Key Points:** *Question:* Can large language models (LLMs) effectively simulate and analyze a randomized clinical trial, accurately summarizing and synthesizing clinical data to evaluate drug efficacy and identify relevant reported symptoms?

*Findings:* In a simulated study using LLMs to generate and analyze clinical notes for a trial comparing a drug to a placebo in epilepsy treatment, AI-driven analyses were found to closely match human expert evaluations. The process demonstrated the ability of LLMs to accurately capture treatment effects and identify reported symptoms, with minimal differences in outcomes between the human and LLM analyses.

*Meaning:* The use of LLMs in simulating and analyzing clinical trials offers a promising approach to developing inductive reasoning systems based on electronic medical records. This could revolutionize the way clinical trials are conducted and analyzed, enabling rapid, accurate assessments of therapeutic efficacy and safety without the need for specialized medical language training.

## Introduction

It is very challenging to extract knowledge from the electronic medical system^1^. Various approaches, including the use of structured data^2^, natural language processing toolboxes^3–5^, and others have been shown to hold some promise. Nevertheless, the dream of an AI ingesting hundreds of millions of patient charts to develop “clinical judgement” is currently still not practical. With the advent of highly capable foundational large language models (LLMs)^6–8^, this dream may be closer to reality than ever before. Current generation systems are plagued with a variety of constraints, including very modest input size limits confabulations (a.k.a. “hallucinations”), inaccuracies, biases, and incomplete knowledge bases^6^. Despite these limitations, modern LLMs have made important strides both in the realm of generative AI for producing artificial documents, as well as in information extraction and summarization.

In this study, we set out to explore a model system of using an LLM (Figure 1, LLM A) to generate clinical data and other LLMs (Figure 2, LLMs B and C) to summarize and synthesize information. The hypothesis was that a simulated randomized clinical trial could be generated, summarized, and accurately evaluated with the help of LLMs. *Inductive reasoning*, defined here as generalizing knowledge based on a set of observations, is submitted as one of the ways clinicians learn. The purpose of this task was to demonstrate the power of AI-enhanced inductive reasoning applied to medical chart review.

**Figure 1:**
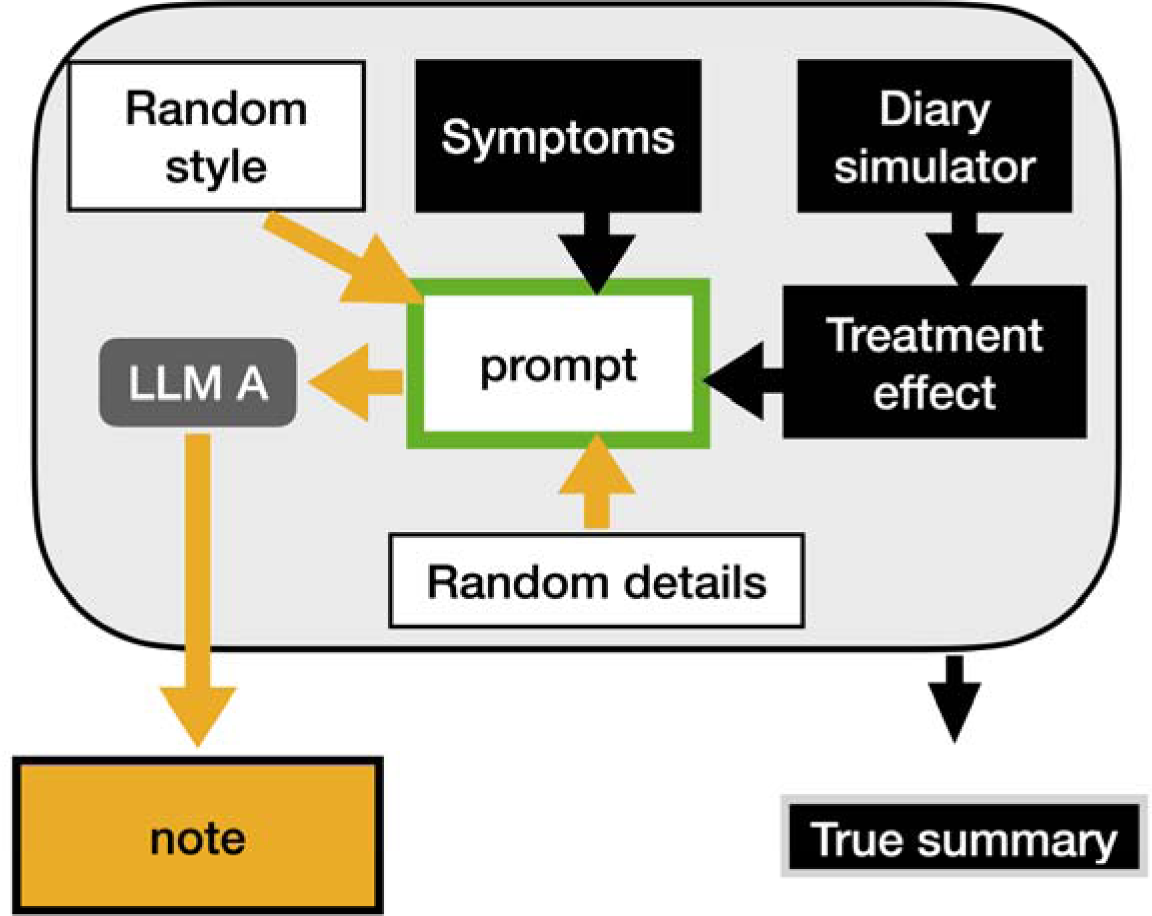
Generation of clinical notes. The diary simulator (CHOCOLATES) was used to produce a realistic seizure diary. This was modulated by the treatment effect (0% in placebo arm, and 39% in drug arm) during the experimental maintenance stage. One of four writing styles were chosen, and a random set of reported symptoms were selected (based on previously reported incidence of symptoms for that arm). These items were used to generate the prompt submitted to LLM A (Llama 2:7b). The LLM generated the clinical note. The true summary was generated based on the original elements used to produce the prompt.

**Figure 2:**
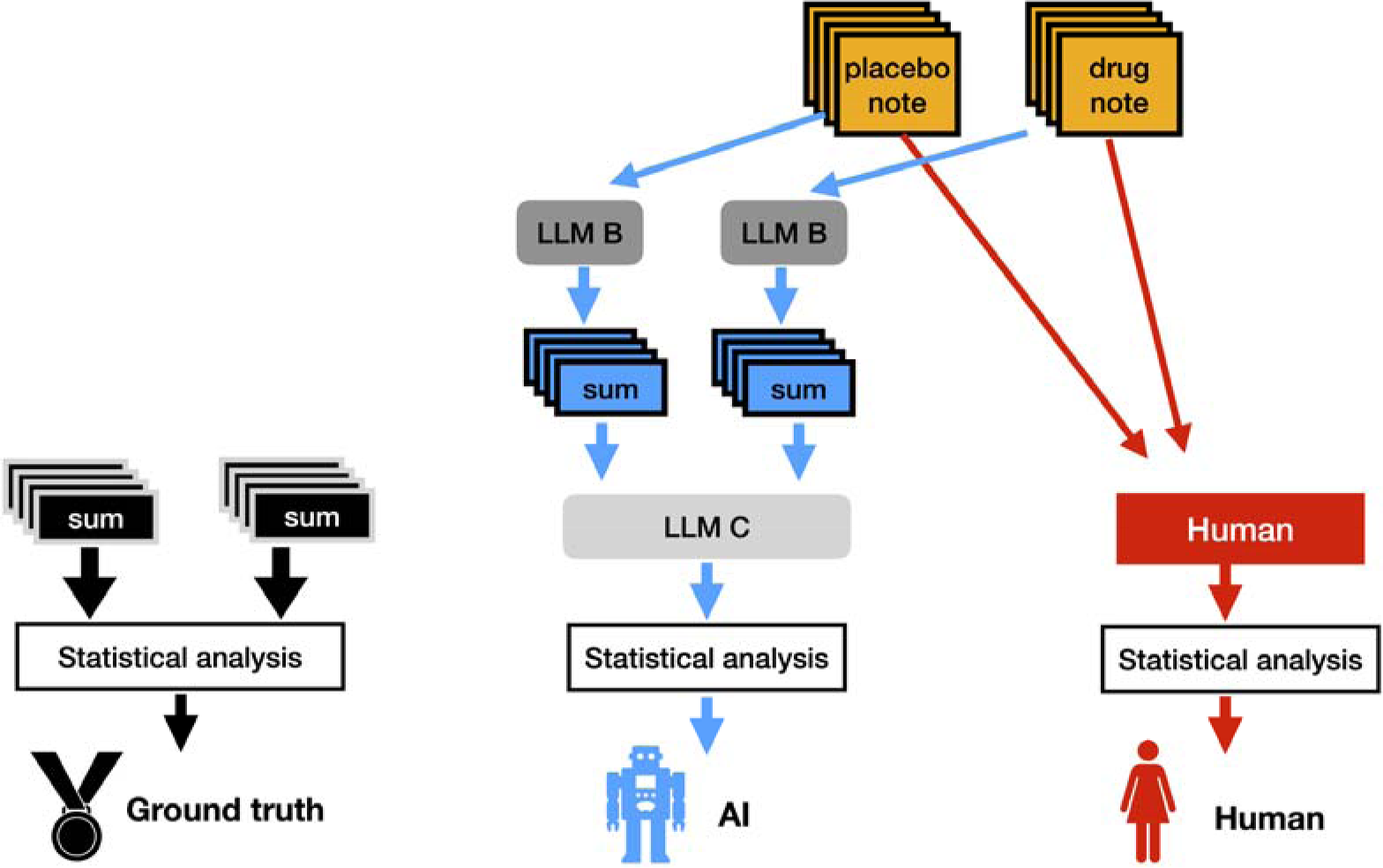
Analyzing the trial. The Ground truth summaries (Figure 1) were used directly as a data table. The AI pathway took the clinical notes (Figure 1), and then LLM B (Mistral) produced a summary that indicated the number of seizures and symptoms reported. LLM C (Claude 2) was used to further summarize and synthesize the brief summaries from LLM B into a complete data table. The clinical notes (Figure 1) were manually assessed by the Human to build a data table. The data tables from the Ground truth, the AI and the Human were analyzed in the standard statistical fashion.

## Methods

### Building the dataset

A simulated randomized clinical trial was constructed (Figure 1) based on an actual clinical trial in epilepsy for cenobamate^9^. In that trial, there were 2 months of baseline, and 3 months of maintenance at steady state for the drug. In our simulation, there was a placebo arm and a full-strength drug arm (corresponding to 400mg/day cenobamate). Similar to the cenobamate trial, we included 120 patients per arm. To generate a realistic cohort of simulated patients, a previously validated simulator (CHOCOLATES) was used^10^. CHOCOLATES was designed to account for heterogeneity in seizure frequencies across patients^11^, the “L-relationship” power law within dairies^12^, seizure clustering^13,14^, seizure susceptibility cycles^15,16^, and maximum allowable seizure rates^17^. Based on multiple lines of evidence^10,18–24^, we assumed that placebo did not have any intrinsic effect and any measured effect would be due to natural variability and regression to the mean^25^. Similar to the RCT^9^, simulated patients needed to have an average rate of 4 seizures per month to be included in the simulated study. Like cenobamate, the simulated drug was 39% more effective than placebo^9^. The precise symptom reporting rates in the placebo and drug arms of the cenobamate trial were simulated as well^9^. A well-characterized, open-source LLM^26^ called Llama2:7b was used to generate clinical notes with the temperature parameter set to 1.0 (values >0 increase creativity). The creativity, as well as LLM hallucination, were intentionally part of this study to properly simulate “noise” caused by inaccurate patient reporting and inaccurate documentation. One of four neurologist writing styles was randomly selected at the time of each clinical note generation: 1) a terse minimalist style using bullet points, 2) a complete but brief style, 3) a narrative style in 2-4 paragraphs, and 4) an erudite academic professor with many extraneous details. Each simulated patient had two notes generated, one after the baseline period and one after the blinded maintenance period (480 notes total). Additional random details about the patients’ past medical history were added randomly but kept consistent within each patient. In addition to a complete note, each encounter also generated a “ground truth” entry in a data table that indicates what information was used in the prompt to the LLM to generate the clinical note.

### AI analysis of the notes

An AI pipeline for analysis of the RCT was constructed as follows (Figure 2). Each note was fed individually (due to input size constraints) to a second open-source LLM^27^ (Mistral 7B v0.1) set to a temperature of 0.0 to increase precision and decrease extraneous detail. This LLM was selected because it was produced independently of Llama2, and thus would not have the luxury of expecting certain styles or methods of writing. The LLM was asked to summarize the note, specifically indicating the number of seizures during the observation period and what symptoms were reported by the patient. Due to inaccuracies and incomplete responses from typical open-source LLMs, it was not feasible for the LLM to build the final data table required for statistical analysis. Thus, a set of somewhat poorly formatted but mostly complete summaries was obtained from the second LLM.

A third LLM (Claude 2), was also used. This LLM has an extended data input limit and is able to ingest large numbers of summaries at once, resulting in the ability to produce a well formatted data table, and correctly make synthesis inferences correctly. Claude 2 is freely available via web interface, but the application programming interface (API) requires a paid account. In addition to improving the formatting, the third LLM was asked to indicate, on each row, the number of seizures during each period of the study; it was also asked to indicate if there were symptoms reported in the second encounter that differed from the first encounter (representing new symptoms that started along with the experimental treatment).

### Human analysis of the notes

The set of 480 generated clinical notes were assessed by one of the authors (SH), a trained neurologist. The relevant features, namely, the number of seizures during the observation period and any symptoms reported, were manually extracted and organized into a data table.

### Statistical analysis of data tables

Three data tables (the ground truth, the AI, and the human) were analyzed in the same way. The percentage change between average monthly seizure rate during baseline to average monthly seizure rate during the maintenance period was computed for each patient^28^. These percentage change values were used to tally the fraction of 50%-responders in each arm, and then to compute a Fisher Exact test to compare arms (RR50). The same percentage change values were also used to compute a median percentage change (MPC) and the Mann-Whitney U test was used to non-parametrically compare the two arms. Uniquely reported symptoms were tallied up in each arm, and these were summarized.

The TRIPOD reporting checklist^29^ is provided (Appendix). Code was prepared in python using langchain and ollama. Open-source code is available at https://github.com/GoldenholzLab/LLM-rct.git.

## Results

Computational time for generation and summarization of notes combined took roughly 20 hours on a single computer; this time would of course be reduced with increased computational resources. The complete set of notes are available for review (Appendix). The human review of the 480 notes required roughly 5 hours. In the placebo group, 9 patients were identified as not having any value reported for seizures in either the baseline or maintenance periods. In the drug group there were an additional 8 such patients. These failures can be attributed to the generative LLM A (Figure 1) that produced the notes. These were not corrected, as these represented examples of undesirable “noise” that prevented perfect reconstruction of the ground truth. When computing the statistics for efficacy, patients with missing numbers were excluded. All patients were included when computing symptom report summaries.

The treatment effect sizes reported for the 50%-responder rate (RR50) and median percentage change (MPC) are shown in Figure 3. The marginal efficacy between drug and placebo are shown in Table 1. All comparisons were statistically significant. The AI and human marginal efficacies differed by 1% in both RR50 and MPC.

**Figure 3:**
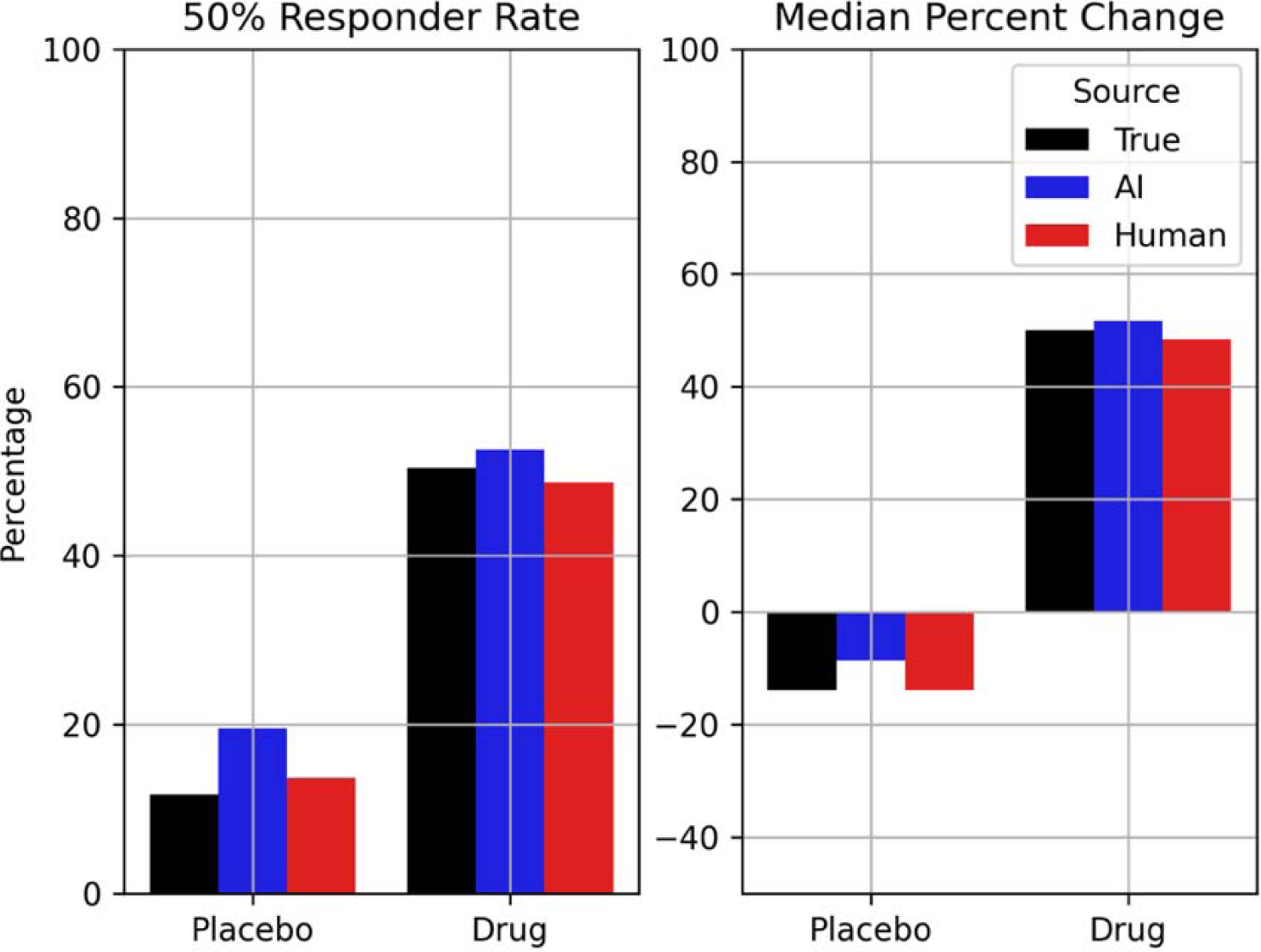
Treatment effect. Shown here are the 50% responder rate (RR50) and the median percentage change (MPC) from the placebo and drug arms of the simulated study. Three colors are shown: ground truth (black), AI estimated (blue), and human reviewed (red). All three were similar though not identical. Nevertheless, both the AI and the human would conclude that the drug is dramatically better than placebo.

**Table 1:**
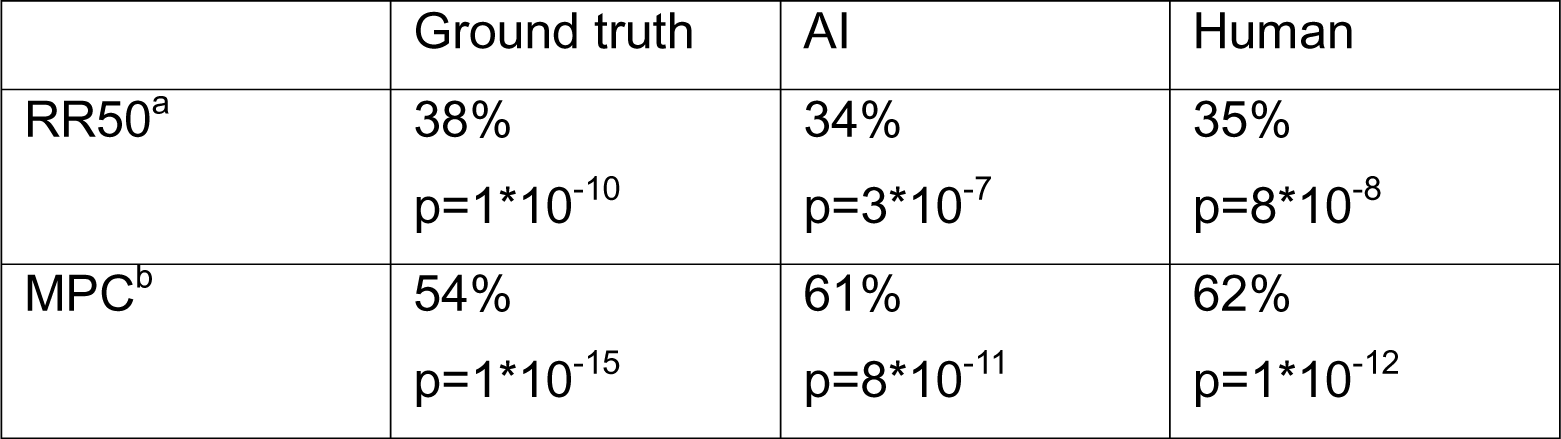
The marginal difference between placebo and drug efficacy using the 50%-responder method (RR50) or the median percentage change (MPC) methods. ^a^ RR50p values are computed using Fisher Exact Test. ^b^ MPC p values were computed using Mann-Whitney U test.

The reported symptoms identified from each of the data tables are shown in Figure 4. The maximum differences in symptom rates between tables were: AI vs. truth – 2%, human vs truth – 2%, AI vs. human – 3%.

**Figure 4:**
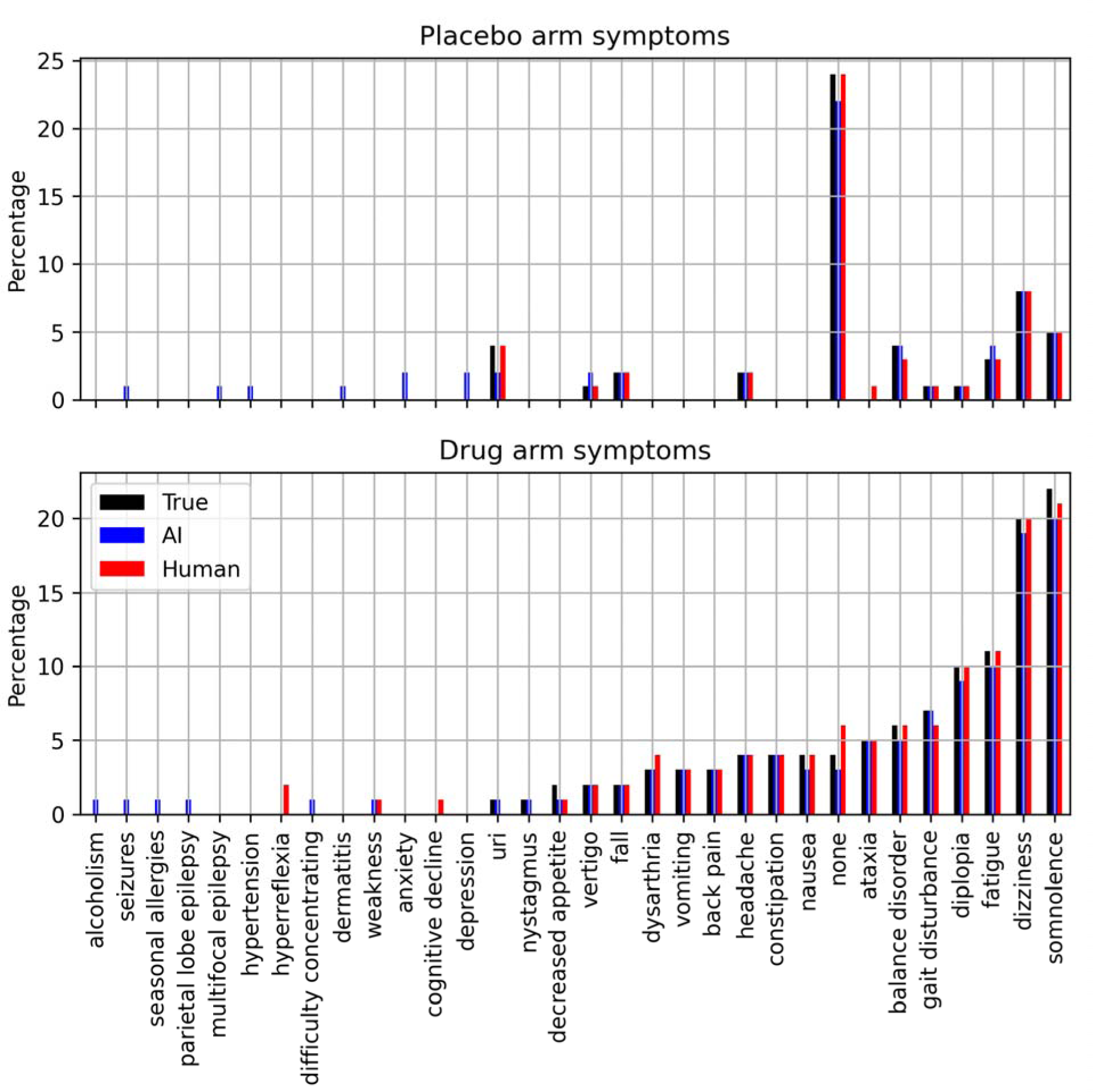
Symptom list. Shown here are the symptoms found in either drug or placebo groups. The ground truth (black), AI derived (blue), and human reviewed (red) bars indicate the fraction of each group that reported the specific symptom. Not all bars match, however the general trend is that they are within 3% of each other.

## Discussion

This study simulated a realistic trial modelled after a recently published randomized drug trial^9^, and using AI, was able to reconstruct the important elements that were reported quantitatively and qualitatively in the clinical notes. The AI pipeline was able to correctly show the marginal drug efficacy (drug vs. placebo) differing from human review by no more than 1%. Similarly, the pipeline was able to identify the relevant symptoms reported in drug and placebo arms, differing with the human by no more than 3%. The use of generative AI allowed us to intentionally inject “noise” (distracting and/or incorrect elements) into our experiment. This deliberate addition was made to help determine if we could teach AI system to learn medical information by induction in the presence of noise. In typical clinical situations, there is virtually always some “noise” generated, whether due to inaccurate reporting by patients or caregivers, or inaccurate recording by clinicians. Our system was able to correctly show a strong effect of the simulated drug and found the appropriate common side effects without being taught to look for something specific. These achievements are all the more remarkable when considering an important point: this entire project did not make use of *any* LLMs specially trained in medical language^30^. Moreover, advanced APIs, necessitating expensive and computationally prohibitive setups, were not required.

Future versions of the present pipeline might employ only a single LLM if it was computationally efficient, reliable, inexpensive and had a very large token size. The advantage of the current approach is that it is not necessary to wait for such advances to be made available.

Like any simulation, this study is only as good as the assumptions made. We assumed we have an adequate model for seizure diaries and trial simulation based on prior work^10,21,24,31,32^. We also assumed that generative LLM clinical notes can represent a first approximation for true clinical notes, and that the conditions presented here are relevant to other inductive learning tasks of interest in clinical settings. Another limitation of this study was a linguistic one: our study was conducted entirely in English. Multilingual open-source models^33^ are available to extend the present work to many other languages.

It must also be noted that extremely rare side effects in a randomized controlled trial might be missed by the type of system developed here – for example, if an investigational drug causes a systemic inflammatory reaction in only 1 patient for the whole study, this fact must not be missed by trialists. Whereas the system proposed here may miss such rare side effects, our goal is to look for larger trends and not “outlier” rare results. Indeed, if such rare reactions were only noted in postmarketing studies, it could take a long time for regulators (and therefore clinicians) to become aware of them, yet an inductively learning AI system could flag situations like this if they happen in low fractions of patients beyond expected levels.

The longer-term purpose of building clinical inductive learning tools is to develop real-time systems that can learn from very large populations and apply this knowledge to uncertain situations. For instance, if a new drug is approved, physicians develop a certain personal clinical “experience” with that drug, and after this they base their prescribing habits on that experience. That personal experience sometimes matches the clinical trials, while sometimes there is a mismatch. This “clinical experience” is one of the ingredients that makes seasoned clinicians more effective at choosing from an uncertain set of choices. If an AI-enhanced system can develop such clinical experience across populations, it will be able to rapidly assist countless clinicians with the most updated experience base possible – vastly larger than any one clinician can accrue in their personal practice.

In conclusion, we demonstrated that known (but hidden) knowledge could be learned by induction with a moderate sample of patient charts. Further studies are needed to expand this capability to broader medical knowledge acquisition and applications.

## Data Sharing Statement

Open-source code and data is available on: https://github.com/GoldenholzLab/LLM-rct.git.

DMG – project design and oversight, data interpretation, manuscript writing and editing

SRG – manuscript editing

SH – data analysis, manuscript editing

MBW – project oversight, manuscript editing

## Data Availability

All data produced in the present work are contained in the manuscript

https://github.com/GoldenholzLab/LLM-rct.git

## Acknowledgements

Funding for this work came in part from NIH K23NS124656. The authors wish to thank the open-source community for sharing and distributing large language models such as llama2 and mistral, as these tools can help advance biomedical science.

