## Appendix for "Inductive reasoning with large language models: a simulated randomized controlled trial for epilepsy"

### CLINIC NOTES - PLACEBO ARM

-------------- PATIENT ID 0 -- VISIT 1 -----------

Patient ID: 0

Date of Visit: 2021-12-12

Chief Complaint: The patient presents with a history of hypertension, diabetes, and generalized epilepsy. She has had 12.0 seizures in the past two months.

Past Medical History: Hypertension, diabetes, and generalized epilepsy.

Medications: Levetiracetam, Lamotrigine, Valproate

Allergies: None

Social History: The patient is currently participating in a randomized controlled trial for the treatment of epilepsy. She has been advised to continue her current medications without any changes except for the addition of the investigational drug for epilepsy, which will be taken for 3 months.

General: The patient appears well-nourished and well-developed, with no significant weight loss or gain. She is alert and oriented to person, place, and time. Her speech is clear and coherent, and her thought processes are logical and goal-directed.

Systemic Examination:

* Vital Signs: Blood pressure: 120/80 mmHg; Heart rate: 70 bpm; Temperature: 98.6°F (37°C)

* General: No significant changes in her physical appearance or overall health.

* Neurological: There is no evidence of focal neurological deficits, and her sensory examination reveals normal sensation in all modalities. Her cranial nerves are intact, and her coordination and balance are normal.

* Cardiovascular: There is no evidence of cardiovascular disease, and her heart sounds are normal.

* Respiratory: Her respiratory examination reveals normal lung sounds and no signs of respiratory distress.

Assessment: The patient presents with a history of hypertension, diabetes, and generalized epilepsy, and has been experiencing increased seizure activity in the past two months. Her neurological examination is normal, and she is participating in a randomized controlled trial for the treatment of epilepsy.

Plan:

1. Continue current medications without any changes except for the addition of the investigational drug for epilepsy, which will be taken for 3 months.

2. Monitor the patient's seizure activity and adjust the dosage of the investigational drug as needed.

3. Schedule follow-up appointments every 2 weeks to assess the patient's progress and make any necessary adjustments to her medications.

4. Encourage the patient to maintain a consistent sleep schedule, avoid alcohol and caffeine, and engage in stress-reducing activities such as yoga or meditation to help manage her seizures.

----------------END OF PATIENT ENCOUNTER---------------

-------------- PATIENT ID 0 -- VISIT 2 -----------

Visit #2, January 12, 2022

Patient ID: 0, Female, Age 40, History of Hypertension, Diabetes, and Generalized Epilepsy

The patient presented for her second visit in the randomized controlled trial. She has been receiving the investigational drug ID X13737 for epilepsy over the past three months and has reported a total of 10.0 seizures during this time period. The patient also complained of somnolence and an upper respiratory infection.

On examination, the patient was alert and oriented, with no signs of distress or discomfort. Her vital signs were within normal limits, with a blood pressure of 120/80 mmHg, a heart rate of 70 beats per minute, and a temperature of 98.6 degrees Fahrenheit. The patient's neurological examination revealed no significant changes since the previous visit. She was able to perform a series of tasks without any observable deficits in cognitive or motor function.

Given that the study is now complete, it has been decided to discontinue the investigational drug ID X13737. The patient will be monitored closely for any adverse effects or changes in her condition over the coming weeks. Any new symptoms or concerns will be addressed promptly and appropriately.

In conclusion, this visit was primarily focused on assessing the patient's response to the investigational drug and monitoring her overall health. The patient reported some minor side effects but remained generally stable. As the study is now complete, we will discontinue the medication and closely monitor the patient for any future changes in her condition.

----------------END OF PATIENT ENCOUNTER---------------

-------------- PATIENT ID 1 -- VISIT 1 -----------

---

Clinic Note #1 - Patient ID: 1, Date: 2023-02-06

Identification:

Name: John Doe

DOB: 1959-02-06

Gender: Male

Age: 63 years

Chief Complaint: Recurrent seizures

History of Present Illness (HPI): The patient presents with a history of recurrent seizures over the past two months. He has experienced 8.0 seizures, each lasting approximately 1-2 minutes and occurring at irregular intervals. The patient reports no significant triggers for the seizures, but notes that they tend to occur during periods of high stress or when he is not adherent to his medication regimen.

Subjective:

Patient reported the following symptoms:

* Recurrent seizures (8.0)

* Aphasia (mild)

* Confusion (mild)

* Memory impairment (mild)

* Visual disturbances (blurred vision, flashing lights)

Medications:

1. Lamotrigine (Lamictal) - 200 mg orally once daily

2. Valproate (Depakote) - 500 mg orally twice daily

3. Levetiracetam (Keppra) - 250 mg orally twice daily

4. Topiramate (Topamax) - 25 mg orally once daily

Allergies:

1. None reported

Social History:

Marital status: Married

Occupation: Retired

Languages spoken: English, some Spanish

Review of Systems (ROS):

Cardiovascular: No significant history of cardiovascular disease or risk factors.

Respiratory: No significant history of respiratory disease or risk factors.

Gastrointestinal: History of type 2 diabetes, well-controlled on metformin and lifestyle modifications.

Neurological: History of multifocal epilepsy, currently experiencing recurrent seizures.

Musculoskeletal: No significant history of musculoskeletal disorders or risk factors.

Psychiatric: History of bipolar disorder, currently stable on medication regimen.

General Physical Examination:

Vital Signs:

* Temperature: 98.6°F (37°C)

* Blood Pressure: 120/80 mmHg

* Heart Rate: 70 beats per minute

* Respiratory Rate: 14 breaths per minute

Head and Neck:

* Normal head circumference

* No focal neurological deficits

Extraocular Movements:

* Full range of motion

Cranial Nerves:

* Intact sensation to light touch, pain, temperature, and temperature discrimination

* Intact facial expressions and hearing

Neurological Examination:

Motor:

* Full range of motion in all extremities

* No significant muscle weakness or atrophy

Sensory:

* Intact sensation to light touch, pain, temperature, and temperature discrimination

* No significant sensory deficits

Cognitive:

* Mild aphasia and confusion, but patient is generally coherent and cooperative

Labs:

* Complete Blood Count (CBC) - within normal limits

* Basic Metabolic Panel (BMP) - within normal limits

* Thyroid-stimulating hormone (TSH) - within normal limits

Imaging:

* Brain Magnetic Resonance Imaging (MRI) - no significant abnormalities

Assessment:

The patient presents with a history of recurrent seizures and a diagnosis of multifocal epilepsy. He is currently participating in a randomized controlled trial evaluating the efficacy of an investigational drug for epilepsy. The patient's neurological examination reveals mild aphasia and confusion, but he is generally coherent and cooperative. The patient's medication regimen will remain unchanged at this time, with the addition of the investigational drug for the next three months. The patient will be monitored closely for any changes in seizure frequency or severity, and will be advised to adhere strictly to his medication regimen.

Plan:

1. Continue current medication regimen (lamotrigine, valproate, levetiracetam, topiramate)

2. Add investigational drug for epilepsy (name and dose to be determined by study protocol) for next three months

3. Monitor patient closely for any changes in seizure frequency or severity

4. Discuss with patient the importance of adhering strictly to medication regimen

5. Schedule follow-up appointment for 02/20/2023 to reassess patient's condition and adjust treatment plan as necessary

Signature:

[Your Name]

Date: 02/06/2023

----------------END OF PATIENT ENCOUNTER---------------

-------------- PATIENT ID 1 -- VISIT 2 -----------

Clinic Note - Visit #2

Patient ID: 1

Date: 2023-03-06

Identification:

* Name: John Doe

* Date of Birth: 01/01/1959

* Patient ID: 1

Highlights of the History of Present Illness (HPI):

* Chief Complaint: Multifocal epilepsy, somnolence

* History of Present Illness: John Doe, a 63-year-old male, presents with a history of bipolar disorder, diabetes, and multifocal epilepsy. He has been participating in a randomized controlled trial for the investigational drug ID X13737 for his epilepsy over the past 3 months. During this time, he has experienced 42.0 seizures and reported somnolence.

Subjective:

* Patient reports experiencing 42.0 seizures during the past 3 months since starting the investigational drug ID X13737.

* Patient reports feeling somnolent since starting the medication.

Medications:

* ID X13737 (Investigational drug for epilepsy) - Discontinued due to completion of randomized controlled trial.

* Lamotrigine (for bipolar disorder and seizures) - Ongoing.

* Metformin (for diabetes) - Ongoing.

Allergies:

* None known.

Social History:

* Married with adult children.

* Employed as an accountant.

* Denies any recent travel or illness.

Review of Systems:

* General: No significant complaints.

* Neurological: Seizures, somnolence.

* Cardiovascular: None.

* Respiratory: None.

* Gastrointestinal: None.

* Genitourinary: None.

General Physical Examination:

* Vital signs: Temperature 98.6°F, Blood Pressure 120/80 mmHg, Heart Rate 70 bpm, Respiratory Rate 14 breaths/minute.

* Head and Neck: Normal head circumference, no focal lesions or edema.

* Extremities: No clubbing or cyanosis.

* Abdomen: Soft and non-tender.

* Lymph Nodes: None noted.

Comprehensive Neurologic Examination:

* Mental Status: Alert and oriented to person, place, and time.

* Cranial Nerves: II (optic), III (oculomotor), IV (trochlear), VI (abducens) and VIII (vestibulo-ocular reflexes) intact.

* Motor Examination: No hemiparesis or weakness. Normal tone and power.

* Sensory Examination: Intact sensation to light touch, pain, and temperature.

* Coordination and Gait: Normal heel-to-toe gait. No ataxia or dysmetria.

Labs:

* Complete Blood Count (CBC): Within normal limits.

* Electrolyte Panel: Within normal limits.

* Liver Function Tests (LFTs): Within normal limits.

* Renal Function Tests (RFTs): Within normal limits.

Imaging:

* MRI brain: No significant abnormalities noted.

Assessment:

* Multifocal epilepsy with somnolence and seizures.

* Presently discontinuing the investigational drug ID X13737 due to completion of randomized controlled trial.

* Patient will be referred to a neurologist for further management of their condition.

Plan:

* Referral to a neurologist for further management of multifocal epilepsy and somnolence.

* Monitoring of seizure activity and medication adherence.

Signature:

John Doe, M.D.

Date: 03/06/2023

----------------END OF PATIENT ENCOUNTER---------------

-------------- PATIENT ID 2 -- VISIT 1 -----------

On September 11th, 2021, I conducted a clinic visit with Patient ID 2, a 64-year-old female who has been diagnosed with asthma, diabetes, and epilepsy. The patient has experienced 17 seizures over the past two months, which is concerning and warrants further evaluation.

During the examination, I performed a complete neurological assessment to evaluate the patient's condition. Her vital signs were within normal limits, with a temperature of 98.6 degrees Fahrenheit, blood pressure of 120/80 mmHg, and a heart rate of 72 beats per minute. The patient's mental status was alert and oriented to person, place, and time. She was able to communicate effectively and respond appropriately to questions.

The neurological examination revealed no significant abnormalities, with normal strength, sensation, and reflexes in all four limbs. However, the patient did exhibit a mild tremor in her hands, which is consistent with her history of epilepsy. Additionally, I noticed a slight weakness in the patient's right hand, which may be related to her diabetes or another underlying condition.

Based on the patient's medical history and current symptoms, I have decided to enroll her in a randomized controlled trial to evaluate the effectiveness of an investigational drug for epilepsy. The patient will be taking this medication for three months, and I will closely monitor her seizure activity and overall health during this time. No changes will be made to the patient's current medications, other than adding the investigational drug, to minimize any potential risks or interactions. I will follow up with the patient in three months to assess her response to the medication and make any necessary adjustments to her treatment plan.

----------------END OF PATIENT ENCOUNTER---------------

-------------- PATIENT ID 2 -- VISIT 2 -----------

---

Clinic Note #2, Patient ID 2, Date: 2021-10-11

Identification:

* Name: [Patient Name]

* Date of Birth: [Date of Birth]

* Gender: Female

* Age: 64 years old

* Patient ID Number: [Patient ID Number]

History of Present Illness (HPI):

The patient, a 64-year-old female with a history of asthma, diabetes, and epilepsy, presented for her second clinic visit as part of a randomized controlled trial. The patient has been taking the investigational drug ID X13737 for epilepsy over the past three months and has reported no symptoms. The study is now complete, and the investigational drug will be discontinued.

Subjective:

The patient reported no seizures or other symptoms since her last visit. She did note some mild fatigue, but this was not a significant change from her previous reports. Her mood remained euthymic, and she denied any recent changes in sleep or appetite. The patient's speech and language were normal, with no observable tremors or rigidity.

Medications:

* ID X13737 (Investigational drug for epilepsy): discontinued as part of the study

* Levetiracetam (Keppra) 500mg BID

* Lamotrigine (Lamictal) 200mg OD

* Metformin (Glucophage) 1000mg BID

* Amlodipine (Norvasc) 5mg OD

Allergies:

* None reported

Social History:

The patient lives with her husband and two adult children. She reports no recent changes in her social or work life, and she denies any history of substance abuse or other relevant medical conditions.

Review of Systems (ROS):

The patient had a negative review of systems, with no complaints or concerns reported.

General Physical Examination:

* Vital signs: Blood pressure 120/80 mmHg, heart rate 70 bpm, temperature 98.6°F (37°C), weight 145 lbs

* Head and neck: Normal head size and shape, no focal swelling or masses, normal hearing and vision

* Extremities: No clubbing or cyanosis, normal muscle strength and tone

* Cardiovascular: Regular rhythm, no murmurs or gallops heard

* Respiratory: Clear lung fields, no wheezing or shortness of breath

* Gastrointestinal: No abdominal pain or distension, normal bowel sounds

Comprehensive Neurological Examination:

The patient underwent a comprehensive neurological examination, including an assessment of her cognitive function, mood, and behavior. The following were observed:

* Cognition: The patient's cognitive function was normal, with no evidence of decline or impairment. She was able to perform basic arithmetic calculations and recall recent events without difficulty.

* Mood: The patient's mood remained euthymic, with no signs of depression, anxiety, or other affective disorders.

* Behavior: The patient's behavior was normal, with no evidence of delusions, hallucinations, or other psychotic symptoms. She was able to communicate effectively and respond appropriately to questions and commands.

Labs:

The following laboratory tests were performed and resulted in the following values:

* Complete Blood Count (CBC): WBC 6.5 x 10^9/L, Neutrophils 3.5 x 10^9/L, Lymphocytes 2.0 x 10^9/L, Monocytes 0.5 x 10^9/L, Hemoglobin 13.5 g/dL, Hematocrit 41.7%

* Electrolyte Panel: Sodium 138 mEq/L, Potassium 4.2 mEq/L, Chloride 98 mEq/L, Carbon Dioxide 22 mEq/L

* Liver Function Tests (LFTs): Alanine Transaminase (ALT) 15 U/L, Aspartate Transaminase (AST) 17 U/L, Alkaline Phosphatase (ALP) 45 U/L, Gamma-Glutamyl Transferase (GGT) 12 U/L

Imaging:

No imaging studies were performed during this clinic visit.

Assessment:

Based on the patient's history and physical examination, she is a 64-year-old female with a history of asthma, diabetes, and epilepsy who has been participating in a randomized controlled trial. She has been taking the investigational drug ID X13737 for epilepsy over the past three months and has reported no symptoms. The study is now complete, and the investigational drug will be discontinued. The patient's neurological examination was unremarkable, with no evidence of seizures or other neurological abnormalities. Her laboratory test results were also unremarkable, with no significant changes from her previous visits.

Plan:

1. Discontinue the investigational drug ID X13737 as part of the study.

2. Continue the patient's current medications for epilepsy, diabetes, and asthma as prescribed.

3. Monitor the patient's seizure activity and neurological status closely during future visits.

4. Consider adding additional medications or adjusting existing therapies based on the patient's response to treatment.

Signature:

---

[Your Name]

Neurologist

Clinic Visit #2, Patient ID 2, Date: 2021-10-11

----------------END OF PATIENT ENCOUNTER---------------

-------------- PATIENT ID 3 -- VISIT 1 -----------

Patient: Patient ID 3, 23 year old female

Chief Complaint: Seizures

History of Present Illness: The patient is a 23-year-old female with a history of alcoholism and occipital lobe epilepsy. She has experienced 14.0 seizures in the past two months. She is currently participating in a randomized controlled trial.

Past Medical History: The patient has a history of alcoholism and has been diagnosed with occipital lobe epilepsy. She has been experiencing recurrent seizures for which she has been taking levetiracetam.

Family History: There is no family history of neurological disorders.

Social History: The patient reports drinking heavily in the past, but has since stopped due to her seizure diagnosis. She denies any other significant social or environmental triggers for her seizures.

Review of Systems: The patient reports no other symptoms or complaints.

Neurological Examination:

* General: Alert and oriented x3.

* Mental Status: Intact, with no evidence of cognitive impairment.

* Cranial Nerves: Normal.

* Motor: No noticeable weakness or tremors.

* Sensory: No deficits in sensation to light touch, pain, or temperature.

* Reflexes: 2+ (moderate) at the knee and ankle joints.

* Coordination: Normal.

* Gait: Normal.

Assessment and Plan:

* Diagnosis: Occipital lobe epilepsy with alcoholism as a contributing factor.

* Plan: The patient will continue to participate in the randomized controlled trial and receive the investigational drug for epilepsy for a period of three months. No changes will be made to her current medication regimen, including levetiracetam. The patient will be closely monitored for any changes in her seizure frequency or severity. Follow-up appointments will be scheduled every two weeks to assess her progress and make any necessary adjustments to her treatment plan.

Special Instructions:

* Patient should avoid alcohol and other substances that may exacerbate her seizures.

* Patient should continue to take her medication as prescribed.

* Patient should be advised of the potential risks and benefits of the investigational drug and the randomized controlled trial.

Note: This clinic note is for visit #1 dated 2022-06-20 for Patient ID 3. The patient has a history of alcoholism and occipital lobe epilepsy, and has been experiencing recurrent seizures. The patient is currently participating in a randomized controlled trial and will receive the investigational drug for epilepsy for a period of three months. No changes will be made to her current medication regimen. The patient will be closely monitored for any changes in her seizure frequency or severity, and follow-up appointments will be scheduled every two weeks to assess her progress and make any necessary adjustments to her treatment plan.

----------------END OF PATIENT ENCOUNTER---------------

-------------- PATIENT ID 3 -- VISIT 2 -----------

Clinic Note Vist #2

Patient ID: 3

Date: 2022-07-20

Identification:

* Name: [Insert name here]

* Date of birth: [Insert date of birth here]

* Patient number: [Insert patient number here]

Highlights of the Patient's History:

* Alcoholism (currently abstaining)

* Occipital lobe epilepsy (ongoing)

* Randomized controlled trial participant (ID X13737 for epilepsy)

Subjective:

* Upper respiratory infection (URI) reported by patient

Medications:

* ID X13737 (investigational drug for epilepsy; discontinued as of today's visit)

* Levetiracetam 500mg BID

* Lamotrigine 200mg OD

Allergies:

* None reported

Social History:

* Currently lives with [insert name here] (spouse/family member)

* Marital status: [insert marital status here]

* Occupation: [insert occupation here]

Review of Systems:

* General: Patient reports feeling generally well, despite recent URTI.

* Neurological: No change in seizure activity since last visit (23.0 seizures over past 3 months).

* Cardiovascular: No chest pain or shortness of breath reported.

* Respiratory: Patient reports mild cough and nasal congestion.

* Gastrointestinal: No change in bowel habits or abdominal pain reported.

General Physical Examination:

* Vital signs: T 98.6°F, HR 72, BP 120/80, RR 18

* Weight: [insert weight here] lbs

* Height: [insert height here] inches

* Body mass index (BMI): [insert BMI here]

* Head, eyes, ears, nose, and throat (HEENT): No abnormalities noted.

* Neck, chest, arms, and legs (NCAL): No edema or erythema noted.

* Abdominal exam: Soft, normal bowel sounds, no masses palpated.

* Extremity exam: No deformities, no clubbing or cyanosis noted.

Comprehensive Neurological Examination:

* Mental status: Alert and oriented x3.

* Cranial nerves: II-XII normal.

* Motor function: No weakness or ataxia noted.

* Sensory function: No decreased sensation to light touch, pain, or temperature noted.

* Reflexes: 2+ (normal)

* Gait and station: Normal.

Labs:

* Complete blood count (CBC): WBC 6.5K/mm3, Hb 12.5g/dL, Hct 40.5%

* Blood chemistry: Na 138 mEq/L, K 3.9 mEq/L, Cl 101 mEq/L, CO2 24 mEq/L, BUN 18 mg/dL, creatinine 0.9 mg/dL

* Urinalysis: No red blood cells or white blood cells seen on dipstick. Specific gravity 1.015.

Imaging:

* MRI brain (3/20/2022): No new lesions or abnormalities noted.

Assessment:

* Patient has a history of alcoholism and occipital lobe epilepsy, currently participating in a randomized controlled trial for investigational drug ID X13737.

* Patient reports upper respiratory infection, which may be contributing to mild cough and nasal congestion.

* Seizure activity remains stable at 23.0 seizures over past 3 months, despite recent URTI.

* No significant changes in neurological or physical examination findings since last visit.

Plan:

* Discontinue investigational drug ID X13737 as of today's visit.

* Monitor patient for seizure activity and any changes in condition.

* Follow up with patient in 2-4 weeks to assess for any changes in URTI symptoms or seizure activity.

Signature:

[Insert signature here]

Note: This clinic note is a fictional example and should not be used as a basis for actual medical decision-making or patient care. It is important to consult with a qualified healthcare professional before making any decisions about patient care.

----------------END OF PATIENT ENCOUNTER---------------

-------------- PATIENT ID 4 -- VISIT 1 -----------

Patient ID: 4

Date: 2022-05-23

HPI:

* 54-year-old male

* History of frontal lobe epilepsy

* 13 seizures in past 2 months

* Currently in randomized controlled trial

Neurological Examination:

* Mild weakness in left arm and leg

* Brisk reflexes

* Normal sensation

* No gaze evoked oscillations or spasms

Pertinent Positives:

* No change in medications except for investigational drug for epilepsy

* Randomized controlled trial ongoing

Assessment and Plan:

* Monitor seizure activity and adjust medication as needed

* Continue randomized controlled trial

* Follow-up appointment in 2 weeks

----------------END OF PATIENT ENCOUNTER---------------

-------------- PATIENT ID 4 -- VISIT 2 -----------

Visit #2 - June 23, 2022

Patient ID: 4

Age: 54 years old

History: Frontal lobe epilepsy

The patient, a 54-year-old male with a history of frontal lobe epilepsy, presented for visit #2 on June 23, 2022. Over the past three months, the patient has been participating in a randomized controlled trial involving the investigational drug ID X13737 for the treatment of epilepsy. During this time, the patient has experienced 30.0 seizures with no reported symptoms. As the study is now complete, the investigational drug will be discontinued.

Neurological Examination:

On examination, the patient's neurological status was normal, with no evidence of focal deficits or seizure activity. His consciousness and alertness were intact, and he was able to communicate effectively. The patient's memory and concentration were found to be within normal limits. There was no evidence of dysmetria, dysarthria, or other neurological abnormalities.

The patient's previous history of frontal lobe epilepsy was noted, and his response to the investigational drug ID X13737 was discussed. The patient reported a significant reduction in seizure activity during the trial period, with no adverse effects or symptoms observed. However, as the study is now complete, the drug will be discontinued, and alternative treatments will be explored to manage the patient's epilepsy.

In conclusion, visit #2 for Patient ID 4 revealed a stable neurological status with no seizure activity or adverse effects observed since the last visit. The investigational drug ID X13737 was found to be effective in reducing seizure activity, but will now be discontinued as the study is complete. Alternative treatments will be explored to manage the patient's epilepsy. Regular follow-up appointments will be scheduled to monitor the patient's condition and adjust his treatment plan as needed.

----------------END OF PATIENT ENCOUNTER---------------

-------------- PATIENT ID 5 -- VISIT 1 -----------

Clinic Note: Visit #1 - 2021-04-17

Patient ID: 5 (Female, 45 years old)

History of Present Illness:

The patient, a 45-year-old female with a history of generalized epilepsy, presented to the clinic today for her first visit. She has been experiencing an alarming increase in seizure activity over the past two months, with 113.0 seizures reported during this time frame. The patient is currently enrolled in a randomized controlled trial (RCT) examining the efficacy of a novel investigational drug for epilepsy.

Identification:

Name: [Patient's Name]

DOB: [Date of Birth]

Contact Information: [Home Address, Phone Number, etc.]

Height: [Height in meters]

Weight: [Weight in kilograms]

HPI (History of Present Illness):

The patient's seizure activity has been increasing steadily over the past two months, with a significant spike in activity in the last 30 days. She has reported experiencing approximately 113.0 seizures during this time frame, with no noticeable triggers or warning signs. The patient reports that her seizures are generally well-controlled by her current medication regimen, but she has noticed a slight increase in frequency and intensity recently.

Subjective:

The patient reported experiencing a moderate level of anxiety and stress related to her increased seizure activity and participation in the RCT. She denies any recent changes in sleep patterns, diet, or exercise routine that may be contributing to her symptoms. The patient reports no new medications, allergies, or medical conditions that may be relevant to her present illness.

Medications:

Current Medications:

* Levetiracetam (Keppra) - 2500mg PO QID

* Lamotrigine (Lamictal) - 200mg PO QHS

* Valproic acid (Depakene) - 1000mg PO QID

Allergies:

No known allergies.

Social History:

The patient is married with two children, aged 8 and 10 years old. She works as a freelance writer and reports no significant changes in her work or personal life recently. The patient denies any history of substance abuse or addiction.

Review of Systems:

The patient reported no recent changes in her bowel, bladder, or sexual function. She denied any history of cardiovascular disease, respiratory problems, or other medical conditions that may be relevant to her present illness.

General Physical Examination:

Vital Signs:

* Temperature: 36.8°C (98.2°F)

* Blood Pressure: 120/80 mmHg

* Pulse Rate: 70 beats per minute

* Respiratory Rate: 14 breaths per minute

Head, Eyes, Ears, Nose, and Throat (HEENT):

* No abnormalities noted.

Neck, Back, and Extremities:

* No abnormalities noted.

Chest and Lungs:

* No abnormalities noted.

Abdomen:

* Soft, normal-sized liver and spleen.

Musculoskeletal:

* Normal range of motion and strength in all extremities.

Neurological Examination:

Cranial Nerves:

* All cranial nerves II-XII are intact, with no signs of weakness or paresis.

Motor Examination:

* No abnormalities noted.

Sensory Examination:

* No abnormalities noted.

Coordination and Reflexes:

* Normal coordination and reflexes.

Labs:

* Complete Blood Count (CBC): To be drawn at the end of the RCT.

* Serum Electrolytes: To be drawn at the end of the RCT.

* Liver Function Tests (LFTs): To be drawn at the end of the RCT.

* Renal Function Tests (RFTs): To be drawn at the end of the RCT.

Imaging:

* No recent imaging studies have been conducted.

Assessment:

Based on the patient's history and examination findings, it is my professional opinion that she is experiencing an alarming increase in seizure activity, which may be related to her participation in the RCT. The patient's current medication regimen appears to be well-tolerated and effective in controlling her symptoms, but we will need to closely monitor her condition during the RCT. I recommend that the patient continue with her current medication regimen and remain in the RCT for the full three months to assess its efficacy and safety. We will also need to schedule regular follow-up appointments to monitor her seizure activity and adjust her medication regimen as needed.

Plan:

1. Continue the patient's current medication regimen (levetiracetam, lamotrigine, and valproic acid) for the next three months.

2. Monitor the patient's seizure activity during the RCT, with regular follow-up appointments every two weeks.

3. Adjust the patient's medication regimen as needed to control her symptoms.

4. Schedule the patient for a follow-up appointment at the end of the RCT to assess its efficacy and safety.

Signature:

[Your Signature]

Date: [Today's Date]

----------------END OF PATIENT ENCOUNTER---------------

-------------- PATIENT ID 5 -- VISIT 2 -----------

Visit #2 (2021-05-17)

Patient ID: 5

Age: 45 years

Gender: Female

History: Generalized epilepsy

Neurological Examination:

* Balance disorder reported by patient

* No changes in seizure activity since last visit (132.0 seizures over past 3 months)

Pertinent Positives:

* History of generalized epilepsy

* Investigational drug ID X13737 discontinued due to study completion

Assessment and Plan:

* Balance disorder to be addressed in future visits

* Patient to be monitored for seizure activity and any adverse effects from drug discontinuation

* Consider referral to physical therapy or balance rehabilitation program as needed

----------------END OF PATIENT ENCOUNTER---------------

-------------- PATIENT ID 6 -- VISIT 1 -----------

Neurologist Clinic Note - Visit #1 (2021-04-02)

Patient ID: 6

Age: 83 years old

Chief Complaint: Recurrent seizures

History of Present Illness: The patient is an 83-year-old male with a history of alcoholism, bipolar disorder, and multifocal epilepsy. He has been experiencing recurrent seizures over the past 2 months, with a total of 59.0 seizures during this time period. The patient is currently enrolled in a randomized controlled trial (RCT) evaluating the efficacy and safety of an investigational drug for epilepsy.

Past Medical History: The patient has a history of alcoholism, bipolar disorder, and multifocal epilepsy. He has been diagnosed with depression, anxiety, and sleep disturbances. He has had multiple seizures over the past few years, with no known cause.

Medication History: The patient is currently taking levetiracetam 500 mg twice daily, valproic acid 500 mg twice daily, and lamotrigine 100 mg once daily for his epilepsy. He has also been prescribed risperidone 2 mg once daily for his bipolar disorder.

Social History: The patient is a retired machinist and lives alone. He denies any recent changes in his diet, sleep patterns, or stress levels. He reports no history of substance abuse other than alcohol.

Family History: The patient's father died of a myocardial infarction at the age of 70. His mother died of dementia at the age of 85.

Review of Systems: The patient reports no recent changes in his mental health, cognitive function, or bowel/bladder habits. He does report ongoing pain in his right knee and lower back.

Physical Examination:

* Vital signs: Blood pressure 120/80 mmHg, heart rate 70 beats per minute, temperature 98.6°F (37°C), respiratory rate 14 breaths per minute.

* Head: Normal head circumference, no scalp lesions or edema.

* Eyes: No visual field defects, no abnormal eye movements.

* Ears: No ear discharge, no hearing loss.

* Nose: No nasal congestion, no rhinorrhea.

* Throat: No throat swelling, no laryngeal edema.

* Chest: Clear to auscultation, no adventitious sounds.

* Heart: Regular rhythm, no murmurs or gallops.

* Abdomen: Soft and non-tender, no palpable masses.

* Extremities: No clubbing or cyanosis, no joint swelling or deformity.

* Neurological Examination:

+ Mental Status: Alert and oriented to person, place, and time. No evidence of cognitive impairment.

+ Language: Intact, with no aphasia or dysarthria.

+ Motor Function: Intact, with no hemiparesis or weakness.

+ Sensory Function: Normal sensation in all extremities, with no paresthesias or loss of sensation.

+ Coordination and Balance: Intact, with no ataxia or dysmetria.

Assessment and Plan:

Based on the patient's history of recurrent seizures and his current enrollment in an RCT evaluating an investigational drug for epilepsy, it is essential to monitor his seizure activity closely. The patient will continue taking his current medications, with the addition of the investigational drug for a 3-month period. The patient will be seen every 2 weeks for a follow-up visit to assess his seizure activity and monitor his response to the investigational drug.

The patient's history of alcoholism and bipolar disorder, along with his current seizure activity, suggest that he may be at risk for developing depression or anxiety. Therefore, it is essential to closely monitor his mental status and address any changes in his mood or cognitive function promptly.

The patient's knee and lower back pain should also be addressed, as this may be affecting his quality of life and potentially contributing to his seizure activity.

Keywords: epilepsy, randomized controlled trial, investigational drug, seizures, alcoholism, bipolar disorder, multifocal epilepsy.

----------------END OF PATIENT ENCOUNTER---------------

-------------- PATIENT ID 6 -- VISIT 2 -----------

Visit #2 - 2021-05-02

Patient ID: 6

Age: 83 years

History: Alcoholism, bipolar disorder, multifocal epilepsy

Randomized controlled trial (RCT) - investigational drug ID X13737 for epilepsy

Neurological Examination:

* Mental status: Alert and oriented x3

* Speech: Normal volume and tone

* Hemiparesis (left side): Present, mild weakness in the left arm and leg

* Sensation: Intact to light touch, diminished to pinprick on the left side

* Coordination: Mild ataxia on the left side

* Reflexes: 2+ (normal on the right side)

* Extremities: Left arm and leg show mild weakness

Pertinent Positives:

* Seizures: 216.0 in the past 3 months while taking investigational drug ID X13737

* No symptoms reported

Assessment and Plan:

* Discontinue investigational drug ID X13737 now that the RCT is complete

* Monitor for seizure activity and adjust medication as needed

* Consider referral to physical therapy for mild ataxia on the left side

----------------END OF PATIENT ENCOUNTER---------------

-------------- PATIENT ID 7 -- VISIT 1 -----------

Clinic Note Visit #1

Patient ID: 7

Date: 2022-11-02

Chief Complaint: The patient presents with a history of hypertension, asthma, and epilepsy. She has experienced 5.0 seizures in the past two months, and is currently participating in a randomized controlled trial.

History of Present Illness: The patient reports that she has been experiencing more frequent seizures over the past two months, with an average frequency of 5.0 seizures per month. She also reports that her hypertension and asthma have been well-controlled on current medications.

Past Medical History: The patient has a history of hypertension, asthma, and epilepsy. She was diagnosed with epilepsy at the age of 10, and has been receiving treatment for it since then. She has been taking levetiracetam as her primary anticonvulsant medication.

Medications: The patient is currently taking levetiracetam 500mg twice daily, fluticasone propionate 500mcg inhaler twice daily, and lisinopril 20mg once daily.

Social History: The patient lives with her parents and has a part-time job as a waitress. She denies any history of substance abuse or allergies.

Neurological Examination:

The patient's neurological examination was unremarkable, with no evidence of focal weakness, sensory deficits, or cranial nerve abnormalities. Her speech was clear and her language was intact. She had a normal gait and station. Her reflexes were 2+ (moderate) in all extremities, and there was no evidence of tremors or rigidity.

Assessment: The patient's history of hypertension, asthma, and epilepsy, along with her recent increase in seizure frequency, suggest that she may be at risk for adverse outcomes if her condition is not properly managed. The results of the neurological examination do not reveal any significant abnormalities. Therefore, it is recommended that the patient continue her current medications and participate in the randomized controlled trial to evaluate the efficacy of the investigational drug for epilepsy.

Plan:

1. Continue levetiracetam 500mg twice daily, fluticasone propionate 500mcg inhaler twice daily, and lisinopril 20mg once daily.

2. Add the investigational drug for epilepsy (study drug) at a dose of 500mg twice daily for a period of 3 months.

3. Follow-up appointment in 2 weeks to monitor the patient's seizure frequency and assess her response to the study drug.

4. Encourage the patient to maintain a consistent sleep schedule, avoid alcohol and caffeine, and engage in regular physical activity to help manage her epilepsy.

----------------END OF PATIENT ENCOUNTER---------------

-------------- PATIENT ID 7 -- VISIT 2 -----------

**Patient Information:**

Name: Patient ID 7

Age: 19 years old

Gender: Female

Chief Complaint: Hypertension, asthma, and epilepsy

**Medical History:**

Hypertension (controlled on medication)

Asthma (well-controlled on medication)

Epilepsy (history of 12.0 seizures over the past 3 months while taking investigational drug ID X13737)

**Current Medications:**

* Lisinopril (hypertension)

* Albuterol (asthma)

* Levetiracetam (epilepsy)

**Last Menstrual Period (LMP):** Not applicable

**Reason for Visit:** Follow-up visit to assess the patient's neurological status after discontinuing the investigational drug ID X13737.

**Neurological Examination:**

General: Alert and oriented x3, recent surgery/injury none

Cranial Nerves:

* Visual acuity: 20/20 OD, 20/25 OS

* Funduscopic exam: Normal

* Extremity motor function: 5/5 bilaterally

* Sensory exam: Full range of sensation in all extremities

* Coordination and balance: Normal

Systems Review:

* Cardiovascular: Regular rhythm, blood pressure 120/80 mmHg

* Respiratory: Clear lung sounds, normal respiratory rate and rhythm

* Abdominal: Soft and normal in all quadrants

* Neurological: No focal deficits or signs of neurological injury.

**Assessment and Plan:**

Assessment: The patient's neurological examination reveals no evidence of neurological deficits or injuries. Her history of hypertension, asthma, and epilepsy are well-controlled on current medications. The discontinuation of the investigational drug ID X13737 is appropriate given the completion of the randomized controlled trial.

Plan:

1. Continue current medications as prescribed.

2. Monitor the patient's seizure activity and adjust medications as needed.

3. Schedule a follow-up visit in 3 months to reassess the patient's neurological status.

**Special Instructions:**

1. Advise the patient to return to the clinic if she experiences any changes in her symptoms or seizure activity.

2. Inform the patient that the investigational drug ID X13737 will be discontinued and no longer available for her use.

----------------END OF PATIENT ENCOUNTER---------------

-------------- PATIENT ID 8 -- VISIT 1 -----------

Neurologist Clinic Note

Visit #1

Date: 2023-02-04

Patient ID: 8

Chief Complaint:

The patient presents with a history of autism, hypothyroidism, and parietal lobe epilepsy. He has been experiencing increased seizure activity over the past two months, with 3.0 seizures occurring during this time frame. The patient is currently enrolled in a randomized controlled trial (RCT) investigating an innovative treatment for his epilepsy.

Past Medical History:

Autism diagnosed at age 3

Hypothyroidism diagnosed at age 10

Parietal lobe epilepsy diagnosed at age 18

Family History:

No history of neurological disorders in immediate family members

Social History:

The patient is a 42-year-old male who is currently unemployed. He lives with his parents and has difficulty communicating with others due to his autism spectrum disorder (ASD). He is non-verbal and uses a communication device to express himself. The patient's cognitive function is mildly impaired, and he has difficulty with independent living skills.

Review of Systems:

The patient denies any recent changes in his seizure activity or medications. He reports no new symptoms or complaints.

Neurological Examination:

* Mental Status: The patient is alert and cooperative, but his cognitive function is mildly impaired. He is able to communicate using a communication device.

* Cranial Nerves: The patient demonstrates gross motor weakness on the right side of his body due to the parietal lobe epilepsy. There is no evidence of cranial nerve deficits.

* Sensory Examination: The patient has decreased sensation to light touch and pinprick on the right side of his body, consistent with the previous diagnosis of parietal lobe epilepsy.

* Motor Examination: The patient demonstrates gross motor weakness on the right side of his body, including a lack of spontaneous movements and difficulty with purposeful movements. He is able to perform purposeful movements with assistance.

* Reflexes: The patient has areflexia on the right side of his body.

* Gait Examination: The patient demonstrates an unsteady gait, with a wide base of support and a tendency to stumble.

Assessment and Plan:

The patient's increased seizure activity over the past two months is concerning, especially given his history of parietal lobe epilepsy. The patient's current medication regimen, which includes levetiracetam and valproic acid, will be maintained for the time being. However, we will add the investigational drug being studied in the RCT to the patient's regimen for a period of three months to assess its efficacy in reducing seizure activity. The patient will be closely monitored for any changes in his seizure activity or medication side effects.

The patient's cognitive function is mildly impaired, and he has difficulty with independent living skills. Therefore, the patient will continue to receive ongoing support from his family and caregivers. The patient will also participate in regular therapy sessions to improve his communication skills and independence.

Future Follow-Up:

The patient will be followed up in one month to assess the efficacy of the investigational drug and monitor for any changes in his seizure activity or medication side effects.

----------------END OF PATIENT ENCOUNTER---------------

-------------- PATIENT ID 8 -- VISIT 2 -----------

---

Clinic Note #2, dated March 4th, 2023

Patient ID: 8

Subject: 42-year-old male with autism, hypothyroidism, and parietal lobe epilepsy

Identification:

Name: John Doe

Date of Birth: January 15th, 1981

Contact Information:

Address: 123 Main St, Anytown, USA

Phone Number: 555-555-5555

Chief Complaint:

John Doe presents for a routine follow-up visit as part of a randomized controlled trial (RCT) evaluating the safety and efficacy of investigational drug ID X13737 for the treatment of parietal lobe epilepsy. The patient has been taking the drug for the past 3 months, during which time he has reported no symptoms and experienced 13.0 seizures.

History of Present Illness (HPI):

John Doe is a 42-year-old male with a history of autism, hypothyroidism, and parietal lobe epilepsy. He has been experiencing seizures for the past several years, with an average frequency of approximately one seizure per month. The patient's seizures have been primarily focal, with no loss of consciousness or postictal confusional state.

Review of Systems (ROS):

General: Denies fever, chills, or recent illness.

Neurological: No change in seizure frequency or type since beginning the investigational drug.

Psychiatric: No change in mood or cognitive function.

Social History: Married with two children, ages 8 and 10. Patient is employed as a software engineer and denies any recent changes in work or social activities.

Medications:

ID X13737 (investigational drug) - discontinued as of today's visit

Levetiracetam (Keppra) 500mg BID

Quetiapine (Seroquel) 25mg QHS

Allergies: None

Labs:

Complete Blood Count (CBC): WNL

Serum Electrolytes: WNL

Thyroid Function Tests (TFTs): T4, 8.3 mcg/dL (normal range: 4.5-11.0 mcg/dL); T3, 1.2 ng/dL (normal range: 0.7-1.8 ng/dL)

Imaging:

MRI brain with and without contrast: No evidence of acute or chronic changes in the brain parenchyma or ventricles.

Assessment:

John Doe is a 42-year-old male with a history of autism, hypothyroidism, and parietal lobe epilepsy who has been participating in an RCT evaluating the safety and efficacy of investigational drug ID X13737. Over the past 3 months, the patient has experienced 13.0 seizures while taking the drug, with no change in seizure frequency or type since beginning the study. The patient reports no symptoms and denies any recent changes in mood or cognitive function. The investigational drug will be discontinued now that the study is complete.

Plan:

1. Discontinue ID X13737 as of today's visit.

2. Continue levetiracetam (Keppra) 500mg BID and quetiapine (Seroquel) 25mg QHS as previously prescribed.

3. Schedule a follow-up visit in 3 months to monitor seizure frequency and assess for any changes in mood or cognitive function.

Signature:

[Your Name]

Neurologist, MD

----------------END OF PATIENT ENCOUNTER---------------

-------------- PATIENT ID 9 -- VISIT 1 -----------

---

Clinic Note - Visit #1 (2021-07-03)

Patient ID: 9

Age: 79 years

Gender: Male

Chief Complaint:

The patient presents with a history of hypothyroidism, diabetes, alcoholism, and temporal lobe epilepsy. He has experienced 5.0 seizures in the past 2 months.

Past Medical History:

* Hypothyroidism (on levothyroxine)

* Diabetes mellitus type 2 (on metformin and glipizide)

* Alcoholism (currently abstaining)

* Temporal lobe epilepsy (onset 5 years ago, currently on levetiracetam and valproate)

Medications:

* Levothyroxine (50 mcg oral daily)

* Metformin (1000 mg oral bid)

* Glipizide (5 mg oral bid)

* Levetiracetam (2000 mg oral bid)

* Valproate (1000 mg oral bid)

Allergies: None known.

Review of Systems:

* No changes in seizure activity since last visit.

* No new symptoms or complaints.

Physical Examination:

Vital Signs:

* Temperature: 98.6°F (37°C)

* Blood Pressure: 120/80 mmHg

* Heart Rate: 70 beats per minute

* Respiratory Rate: 14 breaths per minute

* Height: 5'10" (178 cm)

* Weight: 175 lbs (79 kg)

Neurological Examination:

Motor Examination:

* No tremors or rigidity noted.

* Normal muscle strength and tone.

* No signs of ataxia or dysmetria.

Sensory Examination:

* Intact sensation to light touch, pain, and temperature over all extremities.

* No evidence of sensory loss or paresthesias.

Cranial Nerve Examination:

* II-XII cranial nerves are intact.

Abdominal Examination:

* Normal bowel sounds and no signs of abdominal distension or tenderness.

Assessment and Plan:

The patient is a 79-year-old male with a history of hypothyroidism, diabetes, alcoholism, and temporal lobe epilepsy, who has experienced 5.0 seizures in the past 2 months. The patient is currently enrolled in a randomized controlled trial and will be taking the investigational drug for epilepsy for the next 3 months. No changes in medications will be made other than adding the investigational drug.

The patient's neurological examination reveals no significant changes since the last visit. The patient remains stable on his current medications and there are no new symptoms or complaints.

The plan for the next 3 months includes continuing the current medications, including the investigational drug, and closely monitoring the patient's seizure activity and overall health status. The patient will be seen again in 3 months for a follow-up visit to assess the effectiveness of the investigational drug and any changes in the patient's condition.

---

Note: This clinic note is fictional and does not reflect any actual patient or medical situation. It is intended only as an example of a neurologist clinic note and should not be used for medical advice or treatment.

----------------END OF PATIENT ENCOUNTER---------------

-------------- PATIENT ID 9 -- VISIT 2 -----------

Patient ID: 9

Date: 2021-08-03

HPI:

* 79 year old male

* History of hypothyroidism, diabetes, alcoholism, and temporal lobe epilepsy

* Currently enrolled in a randomized controlled trial (ID X13737 for epilepsy)

Neurological Examination:

* Pertinent positives only:

+ No change in seizure frequency since last visit (13.0 seizures over past 3 months)

+ Fatigue reported by patient

Assessment and Plan:

* Discontinue investigational drug ID X13737 due to study completion

* Monitor fatigue and seizure frequency in follow-up visits

* Consider alternative treatments for epilepsy as needed

----------------END OF PATIENT ENCOUNTER---------------

-------------- PATIENT ID 20 -- VISIT 1 -----------

Patient ID: 20

Visit #1: 2022-11-03

Key History: Hypertension, Hypothyroidism, Diabetes, Temporal Lobe Epilepsy (TLE)

HPI:

* 77-year-old male

* 32.0 seizures in past 2 months

* Randomized controlled trial (RCT) participant

* No change in medications except for investigational drug for epilepsy

Neurological Examination:

* Pertinent Positives:

+ Full range of motion in all extremities

+ Normal strength and tone

+ No sensory deficits or abnormal reflexes

+ No evidence of facial weakness or paresis

* Negative Findings:

+ No seizure activity during examination

+ No changes in mental status or cognitive function

Assessment and Plan:

* Confirm participation in RCT and investigational drug for epilepsy

* Monitor seizure activity and adjust medications as needed

* Schedule follow-up visit in 3 months to assess efficacy and tolerability of investigational drug

* Consider adding additional medication or modifying current regimen based on examination findings and patient response.

----------------END OF PATIENT ENCOUNTER---------------

-------------- PATIENT ID 20 -- VISIT 2 -----------

Clinic Note - Visit #2 (2022-12-03)

Patient ID: 20

Age: 77 years

Chief Complaint: Dizziness and seizures

History of Present Illness:

The patient is a 77-year-old male with a history of hypertension, hypothyroidism, diabetes, and temporal lobe epilepsy. He has been participating in a randomized controlled trial for the investigational drug ID X13737 for epilepsy over the past 3 months. During this time, he has experienced 165 seizures. The patient reports dizziness.

Past Medical History:

* Hypertension

* Hypothyroidism

* Diabetes mellitus

* Temporal lobe epilepsy

Medications:

* Lisinopril

* Levothyroxine

* Metformin

* Lamotrigine

Social History:

* Smoking: None

* Alcohol consumption: Occasional

* Illegal drug use: None

* Travel history: None

Review of Systems:

* General: No fever, fatigue, or weight loss

* Neurological: Seizures (165 over the past 3 months), dizziness

* Musculoskeletal: No joint pain or weakness

* Cardiovascular: Hypertension controlled with medication

* Respiratory: No shortness of breath or cough

Neurological Examination:

* Mental status: Alert and oriented

* Head: Normocephalic, ataxic

* Eyes: Pupils equal and reactive to light, no nystagmus or abnormal movements

* Vision: Normal visual acuity, no field defects

* Hearing: Normal hearing, no hearing loss

* Speech: Intelligible, no dysarthria or slurring

* Motor examination: No hemiparesis or weakness, ataxic gait

* Sensory examination: No sensory deficits

* Reflexes: Brisk knee jerk and ankle jerk, normal tone in all muscle groups

* Coordination and balance: Ataxic

Assessment and Plan:

1. The patient's seizure activity has significantly increased since starting the investigational drug ID X13737. Discontinue the medication due to lack of efficacy and concerns about adverse effects.

2. Manage the patient's dizziness with appropriate therapy such as vestibular rehabilitation or antihistamines.

3. Monitor the patient's blood pressure and adjust medications as needed.

4. Consider additional anticonvulsant medications to control seizures, taking into account the patient's history of drug allergies and sensitivities.

5. Schedule follow-up visits every 2-3 months to monitor the patient's seizure activity, adjust medications, and address any new symptoms or concerns.

Note: This clinic note is based on a fictional patient and is for educational purposes only. It should not be used as a basis for actual patient care without consulting a qualified healthcare professional.

----------------END OF PATIENT ENCOUNTER---------------

-------------- PATIENT ID 21 -- VISIT 1 -----------

Patient ID: 21, Female, 92 years old

History: Generalized epilepsy, 10 seizures in past 2 months

Randomized controlled trial (RCT)

No change in medications except adding investigational drug for epilepsy

Neurological Examination:

* Mental status: Alert and oriented

* Cranial nerves: Normal

* Motor exam: Normal

* Sensory exam: Normal

* Coordination and balance: Normal

Pertinent Positives:

* Seizures occurring every 2-3 days

* No significant change in seizure frequency or severity with current medications

Assessment and Plan:

* Continue current medications

* Add investigational drug for epilepsy (RCT)

* Monitor seizure frequency and severity every 2 weeks

* Re-evaluate medication regimen after 3 months

----------------END OF PATIENT ENCOUNTER---------------

-------------- PATIENT ID 21 -- VISIT 2 -----------

Visit #2 (2022-06-09)

Patient ID: 21

Age: 92 years old

Female

History: Generalized epilepsy

Neurologic Examination:

* Mental status: Alert and oriented

* Speech: Normal

* Motor function: No gross weakness or ataxia

* Sensation: Normal

* Reflexes: 2+ (moderate)

* Extremity strength: 5/5

* Gait: Normal

Pertinent Positives:

* No symptoms reported

* 17.0 seizures in past 3 months while taking investigational drug ID X13737

Assessment and Plan:

* Discontinue investigational drug ID X13737 since study is complete

* Monitor for seizure activity and adjust medication as needed

* Schedule follow-up visit in 4 weeks to assess effectiveness of medication changes

----------------END OF PATIENT ENCOUNTER---------------

-------------- PATIENT ID 22 -- VISIT 1 -----------

CLINIC NOTE #1

DATE: 2022-03-20

PATIENT ID: 22

NAME: [Mr./Ms.] [First Name] [Last Name]

AGE: 59 years old

CHIEF COMPLAINT:

* Anxiety

* Bipolar Disorder

* Multifocal Epilepsy

* Increased seizure activity (21.0 seizures in the past 2 months)

HISTORY OF PRESENT ILLNESS (HPI):

* The patient presents with a history of anxiety, bipolar disorder, and multifocal epilepsy.

* He has been experiencing an increased frequency of seizures over the past 2 months, with a total of 21.0 seizures.

* The patient is currently enrolled in a randomized controlled trial (RCT) for the treatment of his epilepsy.

* No changes will be made to the patient's medications at this time, except for the addition of the investigational drug for epilepsy, which will be taken for 3 months.

SUBJECTIVE:

* The patient reports feeling increasingly anxious and experiencing more frequent seizures over the past 2 months.

* He also reports difficulty sleeping and increased stress levels due to his participation in the RCT.

* The patient denies any recent changes in his medications or medical history.

MEDICATIONS:

* Lamotrigine (Lamictal) 200mg PO QD

* Valproate (Depakote) 500mg PO BID

* Levetiracetam (Keppra) 500mg PO BID

* Investigational drug for epilepsy (additional medication being added to the patient's regimen)

ALLERGIES:

* None reported

SOCIAL HISTORY:

* Married with two adult children

* Works as a software engineer

* Denies any recreational drug use or illicit substance abuse

* Reports no significant changes in his social support network or living situation.

REVIEW OF SYSTEMS:

* General: No fever, chills, or weight loss. Patient appears well-nourished and well-hydrated.

* Neurological: Patient reports increased seizure activity over the past 2 months, with a total of 21.0 seizures. Denies any new onset of weakness, numbness, or paresthesias.

* Psychiatric: Reports increasing anxiety and stress levels due to participation in the RCT. No suicidal or homicidal ideation.

* Medical: None reported.

GENERAL PHYSICAL EXAMINATION:

* Vital signs: Blood pressure 120/80 mmHg, heart rate 70 bpm, temperature 98.6°F (37°C), weight 175 lbs.

* General appearance: Alert and oriented x3. No evidence of motor abnormalities or weakness.

* Head and neck: Normal head circumference, no papilledema.

* Extremities: Normal muscle tone, no tremors or rigidity.

* Sensory: Full range of sensation in all extremities.

* Reflexes: Brisk deep tendon reflexes present in all extremities.

COMPREHENSIVE NEUROLOGIC EXAMINATION:

* Mental status: Alert and oriented x3, with no evidence of cognitive impairment or confusion.

* Cranial nerves: Intact, with no evidence of abnormality.

* Motor function: Full range of motion in all extremities, with no evidence of weakness or atrophy.

* Sensory exam: Full range of sensation in all extremities, with no evidence of numbness or paresthesias.

* Reflexes: Brisk deep tendon reflexes present in all extremities.

* Coordination and balance: Normal.

LABS:

* Complete Blood Count (CBC): Within normal limits.

* Electrolyte panel: Within normal limits.

* Liver function tests (LFTs): Within normal limits.

* Renal function tests (RFTs): Within normal limits.

IMAGING:

* Magnetic Resonance Imaging (MRI) of the brain: No evidence of structural abnormalities or lesions.

* Electroencephalogram (EEG): Within normal limits.

ASSESSMENT:

* The patient's increased seizure activity and anxiety symptoms are likely related to his participation in the RCT.

* The addition of the investigational drug for epilepsy is appropriate, given the patient's history of multifocal epilepsy.

* No changes will be made to the patient's medications at this time, except for the addition of the investigational drug.

* The patient should be monitored closely for any adverse effects or changes in seizure activity while participating in the RCT.

PLAN:

* Continue the patient's current medication regimen, with the addition of the investigational drug for epilepsy.

* Monitor the patient closely for any adverse effects or changes in seizure activity while participating in the RCT.

* Schedule follow-up appointments as needed to assess the patient's progress and adjust medications accordingly.

SIGNATURE:

[Your Name]

Neurologist

Date: 2022-03-20

----------------END OF PATIENT ENCOUNTER---------------

-------------- PATIENT ID 22 -- VISIT 2 -----------

Clinic Note #2

Patient ID: 22

Date: 2022-04-20

History of Present Illness:

The patient, a 59-year-old male with a history of anxiety, bipolar disorder, and multifocal epilepsy, has been participating in a randomized controlled trial for the investigational drug ID X13737 for his epilepsy. The patient has been taking the medication for the past 3 months and has had 29.0 seizures during this time. The patient reported no symptoms.

Neurological Examination:

General: The patient appears well-oriented and in no distress.

Head: Normal head circumference, normocephalic, and symmetrical. No focal defects noted.

Eyes: Pupils are equally reactive to light, with no evidence of afferent pupillary defect. Extraocular movements are intact. Funduscopic examination reveals a normal retina with no hemorrhages or exudates.

Ears: No significant abnormalities noted on otoscopic examination. Weber Class III bilaterally.

Mouth and Throat: Normal oral hygiene, no signs of dysphagia or drooling.

Neck: No cervical spine tenderness to palpation.

Extremities: No significant abnormalities noted on examination of the upper and lower extremities.

Sensory: Full range of sensation in all four limbs, with no signs of numbness or tingling.

Motor: Normal strength and tone in all four limbs, with no signs of weakness or wasting.

Reflexes: Brisk and normal in all four limbs.

Cerebellar Function: No signs of ataxia or dysmetria noted on examination.

Assessment and Plan:

The patient has completed the randomized controlled trial for the investigational drug ID X13737, and as such, the medication will be discontinued. The patient has had a significant number of seizures during the study period, however, there are no concerning symptoms or signs noted on examination.

The patient will continue to be monitored closely for any changes in their condition, and will be advised to follow-up with their primary care physician for ongoing management of their anxiety and bipolar disorder. The patient will also be referred to the appropriate specialist for further evaluation and management of their multifocal epilepsy.

No further testing or interventions are recommended at this time.

----------------END OF PATIENT ENCOUNTER---------------

-------------- PATIENT ID 23 -- VISIT 1 -----------

Clinic Note #1

Patient ID: 23

Date: 2022-04-11

Identification:

Name: [Patient's Name]

DOB: [Date of Birth]

Gender: Female

ID Number: [Patient's ID Number]

HPI:

The patient, a 40-year-old female with a history of dermatitis, seasonal allergies, ADHD, and epilepsy, presents today for a routine follow-up visit. She reports having experienced 6.0 seizures in the past two months, which is an increase from her previous baseline of 2-3 seizures per month. The patient has been participating in a randomized controlled trial (RCT) evaluating the efficacy and safety of an investigational drug for epilepsy.

Subjective:

The patient reports that she has been feeling generally well, but has noticed an increase in seizure activity since starting the RCT medication. She denies any recent changes in her diet, sleep patterns, or stress levels that may be contributing to the increased seizures. The patient also reports no significant side effects from the investigational drug other than mild gastrointestinal upset.

Medications:

1. Levetiracetam (Keppra) 500mg QID

2. Lamotrigine (Lamictal) 200mg QHS

3. Investigational drug for epilepsy (RCT medication) 250mg BID for 3 months

Allergies:

None reported

Social history:

The patient lives with her husband and two children. She reports that her family is supportive of her clinical trial participation and understands the potential risks and benefits associated with the investigational drug. The patient works part-time as a freelance writer and spends most of her free time reading, hiking, and practicing yoga.

Review of systems:

The patient reports no significant changes in her medical history, including no recent illnesses or surgeries. She denies any current symptoms such as fever, chills, or shortness of breath. The patient also reports no recent travel or exposure to potential triggers for seizures such as alcohol, drugs, or certain medications.

General Physical Examination:

The patient's vital signs are within normal limits, including blood pressure 120/80 mmHg, heart rate 70 beats per minute, and temperature 98.6 degrees Fahrenheit. The patient's neurological examination reveals no significant abnormalities, including normal motor function, sensory function, and reflexes.

Comprehensive Neurologic Exam:

The patient undergoes a comprehensive neurologic examination, including assessment of her cognitive function, mood, and behavior. The patient scores 28/30 on the Mental Health Assessment Questionnaire (MHAQ), indicating minimal impairment in her cognitive and emotional functioning. Her speech is clear and concise, with no apparent language deficits. The patient's memory and attention span are within normal limits, and she demonstrates intact executive function abilities.

Labs:

The patient's recent laboratory results include:

1. Complete Blood Count (CBC): Within normal limits

2. Serum electrolytes: Within normal limits

3. Liver function tests (LFTs): Within normal limits

4. Thyroid-stimulating hormone (TSH): 4.25 mIU/L (normal range: 0.4-4.0 mIU/L)

Imaging:

The patient's recent imaging results include:

1. Magnetic Resonance Imaging (MRI) of the brain: No significant abnormalities noted, including no evidence of structural brain injury or lesions.

2. Electroencephalogram (EEG): No significant abnormalities noted, including no evidence of seizure activity outside of clinical settings.

Assessment:

Based on the patient's history, physical examination, laboratory results, and imaging studies, we have performed a comprehensive assessment of her neurological status. The patient's recent increase in seizure activity is concerning and may be related to her participation in the RCT. However, given the lack of any significant changes in her medical history, medications, or lifestyle, it is difficult to pinpoint a specific cause for the increased seizures. Further investigation and monitoring are necessary to determine the efficacy and safety of the investigational drug.

Plan:

1. Continue the patient's current medication regimen, including levetiracetam (Keppra) 500mg QID, lamotrigine (Lamictal) 200mg QHS, and investigational drug for epilepsy (RCT medication) 250mg BID for 3 months.

2. Increase the patient's frequency of follow-up visits to every two weeks to monitor her seizure activity and adjust her medications as needed.

3. Encourage the patient to maintain a consistent sleep schedule, avoid alcohol and drugs, and practice stress-reducing techniques such as yoga and meditation.

4. Consider adding an additional medication or modifying the current regimen if the patient's seizure activity does not improve with continued monitoring and adjustments.

Signature:

[Your Name]

Neurologist

----------------END OF PATIENT ENCOUNTER---------------

-------------- PATIENT ID 23 -- VISIT 2 -----------

---

Clinic Note #2 (2022-05-11)

Patient ID: 23

Demographics:

* Age: 40 years old

* Gender: Female

History of Present Illness:

* The patient presents with a history of dermatitis, seasonal allergies, ADHD, and epilepsy.

* She has been participating in a randomized controlled trial (RCT) for the investigational drug ID X13737 for her epilepsy over the past 3 months.

* During this time, she has experienced 65.0 seizures.

* The patient recently reported an upper respiratory infection.

Review of Systems:

* General: The patient appears well-nourished and well-hydrated. Her demeanor is cooperative and alert.

* Neurological: The patient reports no new neurological symptoms since her last visit. She has been experiencing occasional headaches, which she rates as mild in severity.

* Allergic: The patient has a history of seasonal allergies and dermatitis.

* Medications:

+ ID X13737 (investigational drug for epilepsy): The patient has been taking this medication for the past 3 months as part of the RCT. She will be discontinuing this medication now that the study is complete.

+ Levetiracetam (anticonvulsant): The patient has been taking this medication for her epilepsy and has reported no changes in her seizure frequency or severity.

+ Lisinopril (antihypertensive): The patient has been taking this medication for her hypertension and reports no changes in her blood pressure readings.

Medication Adherence:

* The patient reports excellent adherence to her medications, including ID X13737, levetiracetam, and lisinopril.

Laboratory Findings:

* Complete Blood Count (CBC): Within normal limits (WNT)

* Comprehensive Metabolic Panel (CMP): WNT

* Thyroid-stimulating hormone (TSH): WNT

* Urinalysis: WNT

Imaging Findings:

* Magnetic Resonance Imaging (MRI) of the brain: WNT

Assessment:

* The patient's seizure frequency has remained relatively stable over the past 3 months while taking ID X13737 as part of the RCT. However, with the discontinuation of this medication, her seizure frequency may increase.

* Her upper respiratory infection is being managed with supportive care and antiviral medications.

* The patient's ADHD symptoms have been well-controlled with levetiracetam and lisinopril.

* The patient's dermatitis has been stable, but her seasonal allergies have been exacerbated due to the recent change in weather.

Plan:

* The patient will discontinue ID X13737 as part of the RCT completion.

* Her current medications (levetiracetam and lisinopril) will be continued as prescribed.

* The patient's seizure frequency will be closely monitored, and any changes will be addressed with appropriate medication adjustments.

* The patient's upper respiratory infection will be managed with supportive care and antiviral medications as needed.

* The patient's dermatitis and seasonal allergies will be monitored and managed as needed.

Signature:

---

[Your Name]

Neurologist

Date: 2022-05-11

----------------END OF PATIENT ENCOUNTER---------------

-------------- PATIENT ID 24 -- VISIT 1 -----------

Clinic Note: Visit #1 (2022-08-20)

Patient ID: 24

Name: [Insert name here]

Age: 25 years old

Gender: Male

Chief Complaint: Recent increase in seizure activity

Medical History:

* Dermatitis

* Schizophrenia

* Diabetes

* Epilepsy (currently taking levetiracetam 500mg BID and lacosamide 200mg TID)

Medications:

* Levetiracetam 500mg BID

* Lacosamide 200mg TID

* Investigational drug for epilepsy (added on 2022-08-20, will be taken for 3 months)

Allergies: None known

Social History:

* Married with two children (ages 4 and 6)

* Works as a software engineer

* Denies alcohol or drug use

* Lives in a rural area with easy access to outdoor activities

Review of Systems:

* No changes in sleep, appetite, or mood

* No recent travel or illnesses

* No new skin lesions or exacerbation of existing ones

General Physical Examination:

* Vital signs within normal limits (temperature 98.6°F, blood pressure 120/80 mmHg, heart rate 70 beats per minute)

* Head examination revealed no focal deficits or abnormalities

* Extremity examination revealed no weakness, numbness, or paresthesia

* Sensory examination revealed no abnormalities

Comprehensive Neurological Examination:

* Mental status examination revealed no evidence of cognitive decline or disorganized thinking

* Cranial nerve examination revealed no abnormalities

* Motor examination revealed no weakness, ataxia, or tremors

* Sensory examination revealed no abnormalities

* Coordination and balance examination revealed no difficulties

Labs:

* Complete Blood Count (CBC): Within normal limits

* Electrolyte Panel: Within normal limits

* Liver Function Tests (LFTs): Within normal limits

* Renal Function Tests (RFTs): Within normal limits

* Thyroid-stimulating hormone (TSH): Within normal limits

Imaging:

* Brain magnetic resonance imaging (MRI) performed on 2022-08-15 revealed no evidence of structural brain abnormalities or lesions

Assessment:

* Patient presents with a history of recent increase in seizure activity, which is concerning given his existing diagnosis of epilepsy.

* There are no changes in medications at this time other than the addition of the investigational drug for epilepsy.

* The patient's neurological examination and laboratory results are within normal limits.

* The patient is in a randomized controlled trial and will continue to be monitored closely throughout the course of the study.

Plan:

* Continue current medications (levetiracetam 500mg BID and lacosamide 200mg TID) and add the investigational drug for epilepsy (taken for 3 months).

* Monitor seizure activity closely and adjust medication regimen as needed.

* Schedule follow-up appointments every 2 weeks to assess seizure control and monitor for any changes in medications or neurological status.

Signature: [Insert signature here]

Date: 2022-08-20

----------------END OF PATIENT ENCOUNTER---------------

-------------- PATIENT ID 24 -- VISIT 2 -----------

Clinic Note: Visit #2 - 2022-09-20

Patient ID: 24

Age: 25 years

Chief Complaint: Dizziness, upper respiratory infection

History of Present Illness: The patient is a 25-year-old male with a history of dermatitis, schizophrenia, diabetes, and epilepsy. He has been participating in a randomized controlled trial for the investigational drug ID X13737 for epilepsy over the past 3 months. During this time, he has experienced 18.0 seizures. The patient has reported dizziness and upper respiratory infection.

Past Medical History: The patient has a history of schizophrenia, diabetes, and epilepsy. He has been managed with medications for these conditions.

Medication List: The patient is currently taking levothyroxine, lamotrigine, valproate, metformin, and insulin.

Social History: The patient lives alone and has a supportive family. He is employed as a software engineer.

Review of Systems: The patient denies any recent changes in his mental health status or any new symptoms. He reports having some dizziness and upper respiratory infection.

Physical Examination:

* Vital Signs: Temperature 98.6°F, Blood Pressure 120/80 mmHg, Heart Rate 70 beats per minute, Respiratory Rate 14 breaths per minute

* General: The patient appears well-nourished and well-groomed. He has a normal facial expression and there are no signs of altered mental status.

* Neurological: The patient has a normal gait and station. His speech is clear and comprehensible. There are no signs of tremors, rigidity, or bradykinesia. His reflexes are normal. There is no evidence of weakness or paralysis.

* Sensory: The patient has normal sensation in all extremities. There is no sensory loss or abnormal sensations.

* Cranial Nerves: The patient has normal vision, hearing, and facial movements.

* Motor: The patient has normal muscle strength and tone. There are no signs of weakness or paralysis.

* Coordination and Balance: The patient has normal coordination and balance.

Assessment and Plan:

Assessment:

* The patient has reported dizziness and upper respiratory infection, which may be related to the investigational drug ID X13737.

* The patient has a history of schizophrenia, diabetes, and epilepsy, which may impact his ongoing management.

* The patient is currently participating in a randomized controlled trial for the investigational drug ID X13737, which will be discontinued now that the study is complete.

Plan:

* Continue levothyroxine, lamotrigine, valproate, and metformin as prescribed.

* Discontinue ID X13737 as the patient has completed the randomized controlled trial.

* Monitor the patient's seizure activity and adjust medications as needed.

* Follow up with the patient in 2 weeks to assess for any changes in his symptoms and to monitor his ongoing management.

* Provide appropriate counseling and education regarding his diagnosis, medications, and lifestyle modifications.

----------------END OF PATIENT ENCOUNTER---------------

-------------- PATIENT ID 25 -- VISIT 1 -----------

**Neurology Clinic Note - Visit #1 (2022-11-19)**

**Patient Information:**

Name: Patient ID 25

Age: 67 years old

Gender: Male

**Chief Complaint:**

The patient presents with a history of seasonal allergies, hypertension, diabetes, and parietal lobe epilepsy. He has been experiencing an increased frequency of seizures over the past two months, with 43.0 seizures reported.

**Medical History:**

The patient has a history of parietal lobe epilepsy and has been on a stable medication regimen for the past three years. He has been experiencing an increase in seizure frequency over the past two months, with 43.0 seizures reported. The patient is currently enrolled in a randomized controlled trial (RCT) investigating an new drug for epilepsy.

**Physical Examination:**

On examination, the patient appears well-oriented and alert. His vital signs are as follows: blood pressure 120/80 mmHg, heart rate 70 beats per minute, temperature 98.6 degrees Fahrenheit. The patient's neurological examination is unremarkable, with no focal deficits or abnormalities noted.

**Assessment and Plan:**

Given the patient's history of parietal lobe epilepsy and recent increase in seizure frequency, it is necessary to continue monitoring his condition closely. As the patient is participating in an RCT investigating a new drug for epilepsy, no changes will be made to his medication regimen at this time, except for adding the investigational drug for the next three months. The patient will be seen for follow-up appointments every two weeks to assess seizure activity and monitor for any adverse effects of the investigational drug.

**Additional Notes:**

The patient reports no recent changes in his medication regimen or any other relevant medical history. He denies any recent travel, illness, or injuries that may have contributed to his increased seizure activity. The patient is aware of the potential risks and benefits of participating in the RCT and has given informed consent.

**Recommendations:**

1. Continue monitoring the patient's seizure activity closely, with follow-up appointments every two weeks.

2. Add the investigational drug to the patient's medication regimen for the next three months.

3. Monitor the patient for any adverse effects of the investigational drug.

4. Encourage the patient to maintain a consistent sleep schedule and avoid alcohol and drugs that may exacerbate seizure activity.

----------------END OF PATIENT ENCOUNTER---------------

-------------- PATIENT ID 25 -- VISIT 2 -----------

On December 19th, 2022, I conducted a routine follow-up visit with Patient ID 25, a 67-year-old male participating in a randomized controlled trial. The patient has a history of seasonal allergies, hypertension, diabetes, and parietal lobe epilepsy. Over the past three months, the patient has been administered the investigational drug ID X13737 for their epilepsy.

During this visit, the patient reported experiencing 23.0 seizures since initiating treatment with ID X13737. The patient also complained of dizziness and somnolence. These adverse effects have been consistent throughout the trial period. In light of these findings, it has been decided to discontinue the investigational drug now that the study is complete.

Upon conducting a comprehensive neurological examination, I observed no significant changes in the patient's neurological status since the last visit. The patient's speech, language, and cognitive functions were all intact, with no evidence of any seizure activity or abnormal movements. However, the patient did exhibit some dizziness and reported feeling somnolent, which may be related to the discontinuation of the investigational drug.

In conclusion, this visit marked the end of Patient ID 25's participation in the randomized controlled trial for the investigational drug ID X13737. The patient experienced a significant number of seizures during the trial period, and adverse effects such as dizziness and somnolence were also reported. However, there were no significant changes in the patient's neurological status, and all necessary steps have been taken to ensure a safe and orderly discontinuation of the drug.

----------------END OF PATIENT ENCOUNTER---------------

-------------- PATIENT ID 26 -- VISIT 1 -----------

Clinic Note: Visit #1 - 2023-02-11

Patient ID: 26

Age: 24 years

Gender: Female

Chief Complaint: Seizures

History of Present Illness:

The patient, a 24-year-old female with a history of dermatitis, asthma, and parietal lobe epilepsy, has been experiencing an increase in seizure activity over the past two months. She has had 12.0 seizures during this time period. The patient is currently enrolled in a randomized controlled trial and will be taking the investigational drug for epilepsy for three months as part of the study. No changes to her current medications will be made during this time.

Past Medical History:

* Dermatitis

* Asthma

* Parietal lobe epilepsy

Medications:

* Levetiracetam (Keppra)

* Lamotrigine (Lamictal)

* Topiramate (Topamax)

Social History:

* Non-smoker

* No alcohol consumption

* Denies illicit drug use

* Living with family

Review of Systems:

* No changes in skin lesions or respiratory symptoms

* No new medication allergies or adverse effects

Neurological Examination:

General: Alert and oriented, cooperative and calm.

Cranial Nerves:

* II (optic) - Full visual fields, no abnormalities noted

* III (oculomotor) - No gaze deviation or weakness

* IV (trochlear) - Normal strength and range of motion

* V (trigeminal) - Normal sensation in all dermatomes

* VII (facial) - Normal strength and tone, no facial weakness or paresis

* VIII (vestibulo-ocular reflex) - Normal reaction to caloric stimulation

Motor:

* Full range of motion in all joints, no ataxia or tremors noted

* Normal muscle strength and tone

Sensory:

* Normal sensation in all dermatomes, no numbness or tingling noted

Reflexes:

* Normal deep tendon reflexes, no hyporeflexia or hyperreflexia noted

Assessment and Plan:

The patient presents with a history of increased seizure activity over the past two months. The patient is currently enrolled in a randomized controlled trial and will be taking the investigational drug for epilepsy for three months as part of the study. No changes to her current medications will be made during this time.

The neurological examination reveals no significant abnormalities, with normal cranial nerve function, full range of motion in all joints, and normal muscle strength and tone. The patient's sensation and reflexes are also normal.

Given the patient's history of dermatitis, asthma, and parietal lobe epilepsy, it is important to closely monitor her seizure activity and adjust her medications as needed. The patient will continue to receive levetiracetam (Keppra), lamotrigine (Lamictal), and topiramate (Topamax) as prescribed, with the addition of the investigational drug for epilepsy for the next three months. Regular follow-up appointments will be scheduled to assess the patient's seizure activity and adjust her medications accordingly.

The patient will be advised to continue with her current medications and return for regular follow-up appointments as needed. The patient will also be instructed to keep a seizure log to track any changes in her seizure activity. Additionally, the patient will be reminded to contact the clinic immediately if she experiences any new or worsening symptoms.

----------------END OF PATIENT ENCOUNTER---------------

-------------- PATIENT ID 26 -- VISIT 2 -----------

Clinic Note - Visit #2 (2023-03-11)

Patient ID: 26

Demographics:

* Age: 24 years

* Gender: Female

* Height: 5'8"

* Weight: 130 lbs

Chief Complaint:

* Patient is a 24-year-old female with a history of dermatitis, asthma, and parietal lobe epilepsy.

History of Present Illness (HPI):

* The patient has been participating in a randomized controlled trial for the investigational drug ID X13737 for epilepsy over the past 3 months.

* The patient has experienced 15.0 seizures since starting the medication.

* The patient reports no symptoms.

Subjective:

* Patient reports that she has been feeling well and experiencing no symptoms.

* She denies any changes in her seizure activity or frequency.

* She mentions that she has noticed some improvements in her skin condition (dermatitis) since starting the investigational drug.

Medications:

* ID X13737 (investigational drug for epilepsy): discontinued due to completion of the randomized controlled trial.

* Levetiracetam (keppra) 500mg twice daily for seizure control.

* Prednisone 1mg/kg daily for dermatitis.

Allergies:

* No known allergies.

Social History:

* Patient lives with her parents and has a supportive family environment.

* Patient is a college student and is currently on spring break.

* Patient denies any tobacco or alcohol use.

Review of Systems (ROS):

* General: No fever, chills, or other constitutional symptoms.

* Neurological: No changes in seizure activity or frequency.

* Dermatological: Improvement in skin condition (dermatitis) since starting the investigational drug.

* Respiratory: No changes in respiratory symptoms or status.

General Physical Examination:

* Vital signs: Blood pressure 110/70 mmHg, heart rate 80 beats per minute, temperature 98.6°F (37°C).

* Head: Normocephalic and normotrophic. No focal deficits or abnormalities noted.

* Eyes: No evidence of papilledema or retinal pathology.

* Neck: No lymphadenopathy or thyromentalism noted.

* Chest: Clear to auscultation, no added sounds or wheezes heard.

* Abdomen: Soft and non-tender to palpation, no masses or lesions appreciated.

* Extremities: No clubbing or cyanosis noted.

Comprehensive Neurologic Examination:

* Cranial Nerves: Intact, with normal function.

* Motor Examination: No changes in muscle strength, tone, or coordination.

* Sensory Examination: No changes in sensation to light touch, pinprick, or vibration.

* Reflexes: Normocephalic and normotrophic.

* Gait and Station: Normal gait and station with no ataxia or other abnormalities noted.

Laboratory Findings:

* Complete Blood Count (CBC): Within normal limits.

* Electrolyte Panel: Within normal limits.

* Liver Function Tests (LFTs): Within normal limits.

* Renal Function Tests (RFTs): Within normal limits.

Imaging Findings:

* No changes in brain imaging studies since starting the investigational drug.

Assessment:

* The patient has completed the randomized controlled trial for the investigational drug ID X13737 for epilepsy and has experienced 15.0 seizures during the study period.

* The patient reports no symptoms and has noticed improvements in her skin condition (dermatitis) since starting the medication.

* Based on the patient's history of dermatitis, asthma, and parietal lobe epilepsy, it is important to continue close monitoring of her seizure activity and skin condition.

* Considering the patient's history and current symptoms, it is recommended to continue levetiracetam (keppra) 500mg twice daily for seizure control and prednisone 1mg/kg daily for dermatitis.

Plan:

* Discontinue ID X13737 due to completion of the randomized controlled trial.

* Continue levetiracetam (keppra) 500mg twice daily for seizure control and prednisone 1mg/kg daily for dermatitis.

* Monitor the patient's seizure activity and skin condition closely.

* Schedule a follow-up visit in 3 months to reassess the patient's condition and medications as needed.

Signature:

[Your Signature]

Date: 2023-03-11

----------------END OF PATIENT ENCOUNTER---------------

-------------- PATIENT ID 27 -- VISIT 1 -----------

CLINIC NOTE - VISIT #1

Patient ID: 27

Date: 2022-07-01

Age: 98 years

Gender: Male

Chief Complaint:

* Recent increase in seizure frequency (18.0 seizures in the past 2 months)

* History of dermatitis, ADHD, and parietal lobe epilepsy

History of Present Illness:

* The patient was recently enrolled in a randomized controlled trial for the treatment of parietal lobe epilepsy

* No change in medications will be made other than adding the investigational drug for epilepsy, which will be taken for 3 months

Review of Systems:

* Skin: The patient has a history of dermatitis and currently reports mild-to-moderate facial erythema and scalp inflammation

* Neurological: The patient has a history of ADHD and parietal lobe epilepsy, with recent increase in seizure frequency

* Other: No other significant medical or surgical history

Physical Examination:

* General: The patient appears alert and oriented, with no signs of distress or discomfort

* Head and Neck: Normal head circumference, with no masses or lesions noted

* Eyes: Pupils are equal and reactive to light, with no signs of abnormal movements or torticollis

* Ears: No significant hearing loss or deformities noted

* Nose: No notable deviations from normal nasal anatomy

* Mouth: Normal oral hygiene, with no signs of ulceration or erythema

* Neck: No significant lymphadenopathy or jugular venous distension noted

* Extremities: No significant motor or sensory deficits observed

* Spine: No notable deformities or tenderness to palpation

Assessment and Plan:

* Diagnosis: Parietal lobe epilepsy, ADHD, dermatitis

* Plan:

+ Continue the current medication regimen, including the investigational drug for epilepsy for 3 months

+ Monitor seizure frequency and adjust medications as needed

+ Schedule follow-up appointments for monitoring and adjustment of medications

+ Encourage the patient to continue regular skin care and topical treatments for dermatitis

Special Instructions:

* The patient is enrolled in a randomized controlled trial, and no changes in medication will be made other than adding the investigational drug for epilepsy.

* The patient should return for follow-up appointments as scheduled to monitor seizure frequency and adjust medications as needed.

I hereby certify that I have read and understand the contents of this note, and that the information it contains is accurate and complete to the best of my knowledge.

[Signature of Neurologist]

Date: 2022-07-01

----------------END OF PATIENT ENCOUNTER---------------

-------------- PATIENT ID 27 -- VISIT 2 -----------

Visit #2 - 2022-08-01

Patient ID: 27

Age: 98 years old

History:

* Dermatitis

* ADHD

* Parietal lobe epilepsy

* Participating in randomized controlled trial (RCT)

Investigational Drug:

* ID X13737 for epilepsy (last 3 months)

* 15.0 seizures during this time

Current Problems:

* Dizziness

* Upper respiratory infection

Neurological Examination:

* Mental status: alert and oriented

* Speech: normal

* Motor: normal

* Sensation: normal

* Reflexes: normal

* Coordination: normal

* Gait: normal

Pertinent Positives:

* Dizziness

* Upper respiratory infection

Assessment and Plan:

* Discontinue investigational drug (ID X13737) since RCT is complete

* Monitor for seizure control and address any new symptoms or concerns.

----------------END OF PATIENT ENCOUNTER---------------

-------------- PATIENT ID 28 -- VISIT 1 -----------

CLINIC NOTE #1

Date: 2021-05-30

Patient ID: 28

IDENTIFICATION:

Name: Mary Johnson

Age: 90 years old

Gender: Female

Chief Complaint: Frontal lobe epilepsy

HISTORY OF PRESENT ILLNESS (HPI):

Mary Johnson, a 90-year-old female patient, presents with a history of frontal lobe epilepsy. She has been experiencing an alarming increase in seizure activity over the past two months, with 41.0 seizures reported during this time frame. Her symptoms include sudden loss of consciousness, convulsions, and muscle stiffness.

SUBJECTIVE:

During today's clinic visit, Mary reported experiencing a total of 5 seizures since her last visit on March 20th, 2021. She described the seizures as "major" and "scary," with each episode lasting approximately 2-3 minutes. She also noted that she has been experiencing more frequent "aura" symptoms, which include visual disturbances, confusion, and a strange sensation in her limbs. Mary expressed concern about the increasing frequency and severity of her seizures and requested our assistance in managing her condition.

MEDICATIONS:

Mary is currently prescribed Levetiracetam (Keppra) 500mg BID and Lamotrigine (Lamictal) 200mg OD. She reports taking her medications as directed and denies any non-adherence or changes in her medication regimen.

ALLERGIES:

Mary has no known allergies to medications or other substances.

SOCIAL HISTORY:

Mary lives with her daughter, who is her primary caregiver. She has a supportive social network and enjoys spending time with her family and friends. She denies any recent changes in her living situation or social supports.

REVIEW OF SYSTEMS (ROS):

Mary reports no recent illnesses, injuries, or surgical procedures. She does not smoke or drink alcohol. She reports no changes in her sleep patterns or appetite.

GENERAL PHYSICAL EXAMINATION (GPE):

On examination, Mary appears alert and oriented to person, place, and time. Her speech is clear and comprehensible, and she displays appropriate facial expressions and gestures during conversation. Her gross motor function is intact, with no observable tremors or rigidity. Her fine motor skills are slightly impaired, with mild ataxia observed in her fingers and wrists. Her sensory examination reveals no deficits in vision, hearing, or tactile sensation.

COMPREHENSIVE NEUROLOGIC EXAM (CNE):

Mary's CNE reveals the following:

* Cranial nerves: All gross motor functions are intact, with no signs of weakness or paralysis.

* Motor function: Mary displays mild ataxia in her fingers and wrists, but her gross motor function is otherwise normal.

* Sensory examination: No deficits in vision, hearing, or tactile sensation are observed.

* Coordination and balance: Mary exhibits a slight tremor in her hands and feet, with no signs of dizziness or loss of balance.

* Reflexes: Hypertrophic reflexes are present and symmetrical.

* Mental status: Mary is alert and oriented to person, place, and time. Her speech is clear and comprehensible, and she displays appropriate facial expressions and gestures during conversation.

LABS:

Mary's laboratory results are as follows:

* Complete Blood Count (CBC): White blood cell count 4.5, Neutrophils 60%, Lymphocytes 30%, Monocytes 8%, Eosinophils 2%.

* Electrolyte Panel: Sodium 139 mmol/L, Potassium 4.0 mmol/L, Chloride 105 mmol/L, Carbon Dioxide 22 mmol/L.

* Blood Urea Nitrogen (BUN) and Creatinine: BUN 18 mg/dL, Creatinine 1.0 mg/dL.

* Liver Function Tests (LFTs): Alanine transaminase (ALT) 25 IU/L, Aspartate transaminase (AST) 30 IU/L, Alkaline phosphatase (ALP) 120 IU/L, Gamma-glutamyl transferase (GGT) 8 IU/L.

IMAGING:

Mary's imaging results are as follows:

* Magnetic Resonance Imaging (MRI) of the brain with and without contrast: No obvious abnormalities or lesions are observed in the cerebral cortex, basal ganglia, thalamus, or brainstem.

* Computed Tomography (CT) scan of the head: No intracranial abnormalities or lesions are detected.

ASSESSMENT:

Based on Mary's clinical presentation, laboratory results, and imaging findings, we have established the following diagnosis: Frontal lobe epilepsy with a history of 41.0 seizures in the past two months.

PLAN:

Given Mary's history of frontal lobe epilepsy and the recent increase in her seizure activity, we have decided to maintain her current medication regimen, including Levetiracetam (Keppra) 500mg BID and Lamotrigine (Lamictal) 200mg OD. We will continue to monitor her condition closely and adjust her medications as needed.

We have also enrolled Mary in a randomized controlled trial to evaluate the efficacy of an investigational drug for epilepsy. As part of this trial, Mary will take the investigational drug for a period of three months. We will closely monitor her condition during this time and adjust her medications as needed.

We have counseled Mary about the potential risks and benefits of this trial and have obtained her informed consent prior to enrollment. We will provide ongoing support and education to Mary throughout the trial period.

SIGNATURE:

---

[Your Signature]

Date: 2021-05-30

----------------END OF PATIENT ENCOUNTER---------------

-------------- PATIENT ID 28 -- VISIT 2 -----------

Patient: Patient ID 28, a 90-year-old female with a history of frontal lobe epilepsy

Chief Complaint:

The patient is a 90-year-old female who presents with a history of frontal lobe epilepsy and has been participating in a randomized controlled trial for the investigational drug ID X13737 for the past 3 months.

Past Medical History:

The patient has a history of frontal lobe epilepsy and has been experiencing seizures for several years. She has tried various medications, including levetiracetam and sodium valproate, but has not achieved complete seizure control.

Current Medications:

The patient has been taking the investigational drug ID X13737 for the past 3 months as part of the randomized controlled trial.

Social History:

The patient is a retired school teacher and lives with her husband in a single-story home. She reports no significant social or environmental stressors.

Review of Systems:

The patient reports no new symptoms since her last visit. She has not experienced any changes in her seizure pattern or frequency since starting the investigational drug ID X13737.

Neurological Examination:

General: The patient appears well-oriented and alert. She has a normal speech and language.

Cranial Nerves: The patient has no signs of cranial nerve dysfunction, including no evidence of abducens nerve palsy or oculomotor nerve dysfunction.

Motor Examination: The patient has a normal strength and tone in all four limbs. There is no evidence of ataxia or tremors.

Sensory Examination: The patient has a normal sensation to light touch, temperature, and pain in all four limbs. She does not report any paresthesias or numbness.

Coordination and Balance: The patient has a normal coordination and balance. She is able to walk heel-to-toe without difficulty.

Assessment and Plan:

The patient has been participating in a randomized controlled trial for the investigational drug ID X13737 for epilepsy over the past 3 months and has had 79 seizures during this time. The patient reports no symptoms. The investigational drug will be discontinued now that the study is complete.

Plan:

1. Discontinue the investigational drug ID X13737.

2. Monitor the patient's seizure activity and adjust her current medications as needed to control her seizures.

3. Schedule a follow-up appointment in 2 weeks to reassess the patient's condition and adjust her treatment plan as necessary.

Note: The patient has been advised of the discontinuation of the investigational drug and the need for close follow-up to monitor her seizure activity and adjust her treatment plan as needed. She has been instructed to contact the clinic immediately if she experiences any changes in her symptoms or seizure activity.

----------------END OF PATIENT ENCOUNTER---------------

-------------- PATIENT ID 29 -- VISIT 1 -----------

Visit #1: February 27, 2022

Patient ID: 29

Age: 53 years old

Chief Complaint: Recent increase in seizure activity

History of Present Illness:

The patient is a 53-year-old male with a history of depression, schizophrenia, seasonal allergies, and temporal lobe epilepsy. He has been experiencing an increased frequency of seizures over the past two months, with 12.0 seizures reported. The patient is currently participating in a randomized controlled trial (RCT) and has not experienced any changes in medications other than the addition of the investigational drug for epilepsy, which he will be taking for the next three months.

Past Medical History:

* Depression

* Schizophrenia

* Seasonal allergies

* Temporal lobe epilepsy

* 12.0 seizures in the past two months

Medications:

* Investigational drug for epilepsy (added within the past month)

* No changes in medications other than the addition of the investigational drug

Social History:

* Married with two adult children

* Denies smoking and alcohol consumption

* Reports stress related to his ongoing participation in the RCT

Review of Systems:

* No recent illnesses or injuries

* No changes in sleep patterns or appetite

* No recent travel or exposure to sick individuals

Physical Examination:

General: The patient appears well-nourished and alert, with no signs of distress or discomfort.

Vital Signs:

* Temperature: 98.6 degrees Fahrenheit

* Blood Pressure: 120/80 mmHg

* Heart Rate: 70 beats per minute

* Respiratory Rate: 14 breaths per minute

Neurological Examination:

* Mental Status: The patient is alert and cooperative, with no evidence of cognitive impairment or psychosis.

* Cranial Nerves: No abnormalities noted.

* Motor Examination: No significant tremors or weakness observed.

* Sensory Examination: No sensory deficits or abnormalities noted.

* Coordination and Balance: The patient is able to perform normal activities without any observable difficulties.

Assessment and Plan:

* Recent increase in seizure activity, likely related to ongoing participation in the RCT.

* No changes in medications other than the addition of the investigational drug for epilepsy.

* Continue monitoring the patient's seizure activity and adjust medications as needed.

* Schedule follow-up visit in four weeks to assess the patient's status and make any necessary adjustments to his treatment plan.

Note: This clinic note is a fictional example and should not be used as a basis for actual patient care.

----------------END OF PATIENT ENCOUNTER---------------

-------------- PATIENT ID 29 -- VISIT 2 -----------

---

Clinic Note #2 - Patient ID 29 - 2022-03-27

Identification:

Patient Name: John Doe

Patient ID: 29

Date of Birth: 1969-04-15

Gender: Male

Age: 53 years old

Highlights of the Patient's History of Present Illness (HPI):

Chief Complaint (CC): Back pain, seizures

History of Present Illness (HPI):

Patient reports 17.0 seizures in the past 3 months while on investigational drug ID X13737 for epilepsy. The patient also complains of back pain.

Subjective:

Patient reports experiencing more frequent seizures and increased severity of seizures since starting the investigational drug. The patient describes the seizures as "grand mal" in nature, with loss of consciousness, convulsions, and postictal confusional state. Patient also reports increased back pain, which he rates as 8/10 in severity.

Medications:

1. ID X13737 (Investigational drug for epilepsy) - Discontinued as of today's date

2. Levetiracetam (Keppra) - 2000mg orally BID

3. Lamotrigine (Lamictal) - 25mg orally ODT

4. Aripiprazole (Abilify) - 15mg orally ODT

Allergies:

None reported

Social History:

Married with two children

Occupation: Software Engineer

No known allergies or sensitivities to food or medications

No history of travel or exposure to illness

No smoking or alcohol consumption

Review of Systems (ROS):

General: No fever, chills, or night sweats. Patient reports recent back pain and seizures as described above.

Cardiovascular: No chest pain, shortness of breath, or swelling in the legs.

Respiratory: No cough, wheezing, or shortness of breath.

Gastrointestinal: No nausea, vomiting, or diarrhea. Patient reports decreased appetite and weight loss.

Neurological: Seizures as described above.

Musculoskeletal: Back pain as described above.

Psychiatric: Patient reports anxiety and depression symptoms have worsened since starting the investigational drug.

General Physical Examination (GPE):

Vital Signs:

Temperature: 98.6 degrees Fahrenheit

Pulse: 70 beats per minute

Respiration: 12 breaths per minute

Blood Pressure: 120/80 mmHg

Head and Neck: No focal neurological deficits, no masses or lesions palpable.

Extremities: No weakness or paresthesias in the limbs.

Comprehensive Neurologic Examination (CNE):

Mental Status: Alert and oriented x3. Speech is clear and fluent. Patient is able to recall recent events accurately.

Cerebellar Function: No ataxia or dysmetria noted.

Sensory System: No deficits in sensation to light touch, pinprick, or vibration noted.

Motor System: No weakness or paralysis noted. Patient is able to perform rapid alternating movements and maintain balance.

Reflexes: 2+ (moderate) bilaterally.

Labs:

1. Complete Blood Count (CBC): Within normal limits

2. Basic Metabolic Panel (BMP): Within normal limits

3. Thyroid-Stimulating Hormone (TSH): Within normal limits

4. Serum Electrolytes: Within normal limits

5. Liver Function Tests (LFTs): Within normal limits

6. Renal Function Tests (RFTs): Within normal limits

Imaging:

1. Magnetic Resonance Imaging (MRI) of the brain with and without contrast - No abnormalities noted.

2. Computed Tomography (CT) scan of the head, chest, and abdomen - No abnormalities noted.

Assessment:

The patient presents with a history of recent-onset seizures and back pain while on investigational drug ID X13737 for epilepsy. The patient's neurological examination and laboratory results are unremarkable, except for moderate reflexes bilaterally. Given the recent onset of seizures and back pain, as well as the patient's history of seasonal allergies, temporal lobe epilepsy, depression, and schizophrenia, it is likely that these symptoms are related to the investigational drug. The patient's anxiety and depression symptoms have worsened since starting the investigational drug, which may also be contributing to the seizures and back pain.

Plan:

1. Discontinue the investigational drug ID X13737 due to adverse effects.

2. Increase the dosage of Levetiracetam (Keppra) to 2500mg orally BID for seizure control.

3. Add Valproic acid (Depakote) 500mg orally BID for seizure control.

4. Refer the patient to physical therapy for back pain management.

5. Monitor the patient's symptoms and seizure activity closely, with follow-up appointments scheduled as needed.

Signature:

[Your Name]

---

I hope this helps! Let me know if you have any further questions or if there is anything else I can assist you with.

----------------END OF PATIENT ENCOUNTER---------------

-------------- PATIENT ID 40 -- VISIT 1 -----------

Patient ID: 40, Female, 74 years old

History: Parietal lobe epilepsy, 10 seizures in past 2 months

Randomized controlled trial (RCT)

No change in medications except for investigational drug for epilepsy

Neurological Examination:

* Mental status: Alert and oriented

* Speech: Normal

* Motor function: Normal

* Sensory system: Intact

* Cranial nerves: Normal

* Reflexes: Normal

* Gait: Normal

Pertinent Positives:

* No change in seizure frequency or type

* No new neurological deficits

Assessment and Plan:

* Continue RCT medication as prescribed

* Monitor seizure frequency and severity

* Reassess in 3 months for changes in medication effectiveness

----------------END OF PATIENT ENCOUNTER---------------

-------------- PATIENT ID 40 -- VISIT 2 -----------

On June 28, 2021, I conducted a routine follow-up visit with Patient ID 40, a 74-year-old female who has been participating in a randomized controlled trial for the investigational drug ID X13737 for her epilepsy. Over the past three months, the patient has been taking the medication and has reported no symptoms. However, she has experienced 20.0 seizures during this time period.

Upon examination, the patient's neurological status was found to be unchanged since her last visit. Her speech, language, and cognitive functions were all within normal limits. There was no evidence of weakness, numbness, or paresthesias in her extremities. Her reflexes were appropriate for her age and there was no signs of ataxia or dysmetria.

The patient's past medical history includes a history of parietal lobe epilepsy, which has been well-controlled with current medications. There have been no changes in her medication regimen over the past three months. The patient reported no new symptoms or concerns since her last visit.

Given that the randomized controlled trial is now complete, it has been decided to discontinue the investigational drug ID X13737. The patient will be transitioned back to her previous medication regimen and will be monitored closely for any changes in her seizure activity or other symptoms. Her next follow-up visit is scheduled for three months from now, at which time we will reassess her condition and adjust her treatment plan as needed.

----------------END OF PATIENT ENCOUNTER---------------

-------------- PATIENT ID 41 -- VISIT 1 -----------

Patient ID: 41

Age: 64 years old

History: Occipital lobe epilepsy

Seizures: 4.0 in past 2 months

Randomized controlled trial (RCT)

No change in medications except for investigational drug for epilepsy

Neurological Examination:

* Head: normocephalic, ataxic gait

* Eyes: no gaze preference, no nystagmus

* Vision: 20/20 in each eye

* Hearing: normal hearing sensation

* Motor: no weakness or tremors

* Sensory: intact

* Reflexes: are 2+

Pertinent Positives:

* No change in seizure frequency since last visit

* No new symptoms or medication side effects

Assessment and Plan:

* Continue with current medications and add investigational drug for epilepsy

* Monitor seizure frequency and adverse effects every 2 weeks

* Schedule follow-up visit in 3 months to reassess medication efficacy and monitor for any changes in neurological examination findings.

----------------END OF PATIENT ENCOUNTER---------------

-------------- PATIENT ID 41 -- VISIT 2 -----------

Visit #2, 2021-06-30

Patient ID: 41, Female, 64 years old

History: Occipital lobe epilepsy

Randomized controlled trial (RCT) participant

Took investigational drug ID X13737 for epilepsy (last 3 months)

Reported somnolence

Drug discontinuation due to study completion

Neurological Examination:

* Pertinent positives only:

+ Somnolence

Assessment and Plan:

* Discontinue investigational drug ID X13737

* Monitor for seizure activity and adjust therapy as needed

* Schedule follow-up visit in 2 weeks to assess for any changes in seizure frequency or severity.

----------------END OF PATIENT ENCOUNTER---------------

-------------- PATIENT ID 42 -- VISIT 1 -----------

Patient ID: 42

Visit #1: 2021-11-18

Identification:

* Name: John Doe

* Date of Birth: 01/01/1957

* Patient ID: 42

History of Present Illness (HPI):

* Chief Complaint (CC): Increased seizure activity

* History of Diabetes, Anxiety, and Temporal Lobe Epilepsy

* Currently enrolled in a randomized controlled trial

Subjective:

* Patient reports 12.0 seizures in the past 2 months

* Increased stress and anxiety due to recent changes in medication regimen

* No change in medications will be made other than adding the investigational drug for epilepsy that will be taken for 3 months

* Patient reports no side effects from current medications

Medications:

* Levetiracetam (Keppra) 500mg oral tabs, BID

* Lamotrigine (Lamictal) 200mg oral tabs, OND

* Valproic Acid (Depakote) 500mg oral caps, QID

* Investigational drug for epilepsy: placebo (PBO) or active compound (AC), TID for 3 months

Allergies:

* None reported

Social History:

* Married with two adult children

* Employed as an accountant

* Denies smoking and alcohol consumption

* Reports occasional caffeine intake (1-2 cups of coffee per day)

Review of Systems:

* General: No recent illnesses or surgeries

* Neurological: Recent increase in seizure activity

* Cardiovascular: History of hypertension, no known cardiac disease

* Respiratory: No known respiratory diseases

* Gastrointestinal: History of gastroesophageal reflux disease (GERD), no recent abdominal pain or diarrhea

* Musculoskeletal: No recent joint pain or muscle weakness

* Psychiatric: History of anxiety and depression, currently being treated with antidepressant medication

General Physical Examination:

* Vital Signs:

+ Temperature: 98.6°F (37°C)

+ Blood Pressure: 120/80 mmHg

+ Heart Rate: 70 beats per minute

+ Respiratory Rate: 14 breaths per minute

* Head and Neck: Normal head circumference, no scalp tenderness or swelling. No cervical spine tenderness or limited range of motion.

* Extremities: No clubbing or cyanosis of the fingers or toes. No edema or erythema of the lower extremities.

* Neurological Examination:

+ Cranial Nerves: Intact

+ Motor Function: Normal strength and tone in all extremities. No ataxia or dysmetria.

+ Sensory Function: Normal sensation to light touch, pain, and temperature in all extremities. No sensory deficits.

+ Coordination and Gait: Normal coordination and gait. No signs of ataxia or abnormal gait.

Comprehensive Neurological Exam:

* Mental Status: Alert and oriented to person, place, and time. No evidence of cognitive impairment or altered mental status.

* Cranial Nerves: Intact

* Motor Function: Normal strength and tone in all extremities. No ataxia or dysmetria.

* Sensory Function: Normal sensation to light touch, pain, and temperature in all extremities. No sensory deficits.

* Coordination and Gait: Normal coordination and gait. No signs of ataxia or abnormal gait.

* Reflexes: Normal reflexes in all extremities.

Labs:

* Complete Blood Count (CBC): Within normal limits

* Electrolyte Panel: Within normal limits

* Renal Function Panel: Within normal limits

* Thyroid-stimulating Hormone (TSH) Levels: Normal

Imaging:

* Computed Tomography (CT) Scan of the Brain: No acute or chronic changes in the brain parenchyma. No evidence of stroke or intracranial hemorrhage.

Assessment:

* Patient is a 64-year-old male with a history of diabetes, anxiety, and temporal lobe epilepsy who has been enrolled in a randomized controlled trial for the past 2 months.

* Recent increase in seizure activity, currently averaging 12.0 seizures per month.

* No change in medications will be made other than adding the investigational drug for epilepsy that will be taken for 3 months.

* Patient reports no side effects from current medications.

* Neurological examination is unremarkable, with normal cranial nerves, motor function, sensory function, and coordination and gait.

* Laboratory results are within normal limits, and imaging studies do not show any acute or chronic changes in the brain parenchyma.

Plan:

* Continue current medications, with the addition of the investigational drug for epilepsy for 3 months.

* Monitor seizure activity and adjust medications as needed.

* Schedule follow-up appointment in 2 months to reassess seizure activity and medication regimen.

Signature:

[Your Signature]

Date: 2021-11-18

----------------END OF PATIENT ENCOUNTER---------------

-------------- PATIENT ID 42 -- VISIT 2 -----------

Patient ID: 42

Visit #2: 2021-12-18

History: Diabetes, anxiety, temporal lobe epilepsy (study participant)

Investigational Drug: ID X13737 (epilepsy) - discontinued due to study completion

Neurological Examination:

* General: Alert and oriented x3

* Inspection: No focal deficits or abnormalities

* Mental Status: Intact, but reports fatigue and balance disorder

* Cranial Nerves: Normal

* Motor: No weakness or ataxia

* Sensory: Normal

* Reflexes: Normal

Pertinent Positives:

* Seizure frequency: 20.0 over past 3 months (while on investigational drug)

* Fatigue and balance disorder

Assessment and Plan:

* Discontinue investigational drug ID X13737 due to study completion

* Monitor fatigue and balance disorder, address as needed

* Consider alternative therapies for epilepsy management

----------------END OF PATIENT ENCOUNTER---------------

-------------- PATIENT ID 43 -- VISIT 1 -----------

Clinic Note - Visit #1

Date: 2022-10-16

Patient ID: 43

Age: 60 years

Gender: Female

Chief Complaint: Seasonal allergies, bipolar disorder, and occipital lobe epilepsy with increased seizure activity in the past 2 months (7.0 seizures).

History of Present Illness: The patient presents with a history of seasonal allergies, bipolar disorder, and occipital lobe epilepsy. She has been experiencing an increase in seizure activity over the past 2 months, with 7.0 seizures. She is currently participating in a randomized controlled trial for the treatment of her epilepsy.

Review of Systems:

Allergies: Seasonal allergies

Psychiatric: Bipolar disorder (currently stable)

Neurological: Occipital lobe epilepsy (increased seizure activity in the past 2 months)

Medications:

1. Levetiracetam (Keppra) - 500 mg oral daily

2. Lamotrigine (Lamictal) - 100 mg oral daily

3. Valproic acid (Depakote) - 500 mg oral twice daily

4. Investigational drug for epilepsy (randomized controlled trial) - added 07/15/2022, to be taken for 3 months

Past Medical History: Seasonal allergies, bipolar disorder, occipital lobe epilepsy

Past Surgical History: None

Social History: Denies alcohol and drug use. Smokes cigarettes occasionally.

Family History: No history of neurological disorders or seizure disorders.

General Observations: The patient appears well-oriented and alert. Speech is clear and logical. She has a normal gait and station. No signs of tremors, rigidity, or bradykinesia are noted.

Neurological Examination:

Motor Examination:

1. Right and left upper limb muscle strength: 5/5

2. Right and left lower limb muscle strength: 5/5

3. Muscle tone: Within normal limits

4. Reflexes: 2+ (normal)

5. Coordination and balance: Normal

Sensory Examination:

1. Visual acuity: 20/20 (right eye), 20/40 (left eye)

2. Visual fields: Full to both sides

3. Auditory acuity: Normal

4. Tactile sensation: Within normal limits

5. Temperature sensation: Within normal limits

Cranial Nerve Examination:

1. Ophthalmoscopic examination: No abnormalities noted

2. Funduscopy: No abnormalities noted

3. Facial movements: Normal

4. Bulbar function: Normal

5. Cerebellar function: Normal

6. Extremity sensation and motor function: Within normal limits

Assessment and Plan:

1. The patient presents with a history of seasonal allergies, bipolar disorder, and occipital lobe epilepsy with increased seizure activity in the past 2 months.

2. Neurological examination reveals no significant abnormalities.

3. The patient is currently participating in a randomized controlled trial for the treatment of her epilepsy, and no change in medications will be made other than adding the investigational drug for epilepsy that will be taken for 3 months.

4. Patient will continue to be monitored closely for any changes in seizure activity or adverse effects from the investigational drug.

5. The patient will be seen again in 2 months for a follow-up visit to assess her condition and evaluate the effectiveness of the investigational drug.

Signature: [Your Name]

Date: 2022-10-16

----------------END OF PATIENT ENCOUNTER---------------

-------------- PATIENT ID 43 -- VISIT 2 -----------

Clinic Note - Visit #2 (2022-11-16)

Patient ID: 43 (60 year old female)

Chief Complaint: Seasonal allergies, bipolar disorder, and occipital lobe epilepsy

History of Present Illness: The patient has been participating in a randomized controlled trial for the investigational drug ID X13737 for her epilepsy over the past 3 months. During this time, she has experienced 51.0 seizures and reported dizziness.

Neurological Examination:

* Mental Status: The patient's mental status was normal with no evidence of acute changes in her mood or cognition.

* Head Examination: There were no focal neurological deficits or abnormalities detected during the head examination.

* Sensory System: The patient's sensory systems, including vision, hearing, and touch, were normal.

* Motor System: The patient had a normal strength and tone in all her muscle groups. There was no evidence of any focal weakness or atrophy.

* Cerebellar Function: The patient's cerebellar function was normal with no signs of dysmetria, ataxia, or tremors.

* Extremity Examination: The patient had normal reflexes and there were no signs of any neurological deficits in her upper or lower extremities.

Plan:

* Discontinue the investigational drug ID X13737 as the study is complete.

* Monitor the patient's seizure activity and adjust medications as needed.

* Follow up with the patient in 4-6 weeks to assess her response to the discontinuation of the drug and to monitor for any changes in her condition.

----------------END OF PATIENT ENCOUNTER---------------

-------------- PATIENT ID 44 -- VISIT 1 -----------

Patient ID: 44

Visit #1 - 2022-03-17

History:

* 41-year-old female with history of multifocal epilepsy

* Experiencing 16.0 seizures in the past 2 months

* Currently participating in a randomized controlled trial (RCT)

* No change in medications will be made other than adding the investigational drug for epilepsy for 3 months

Neurological Examination:

* General appearance: alert and cooperative

* Head: normocephalic, normotriche

* Eyes: conjunctival clear, no injection or swelling

* Visual fields: full, without any defects or abnormalities

* Extremities: without weakness, wasting, or sensory deficits

* Sensation: intact throughout, including pain and temperature perception

* Mental status: alert and oriented to person, place, and time

* Speech: clear and coherent

* Cranial nerves: II-XII normal

Assessment:

* Multifocal epilepsy with frequent seizures in the past 2 months

* No significant change in neurological status since previous evaluation

* Patient is currently participating in an RCT and will continue with the investigational drug for another 3 months

Plan:

* Continue the current medication regimen, including the investigational drug

* Monitor seizure frequency and adjust medications as needed

* Schedule follow-up appointment in 3 months to reassess neurological status and medication response

* Encourage patient to maintain a consistent sleep schedule and avoid alcohol and drugs, which may exacerbate seizures.

Note: The patient was informed of the risks and benefits of the investigational drug and provided written informed consent for participation in the RCT. The patient understands the plan and will follow up as scheduled.

----------------END OF PATIENT ENCOUNTER---------------

-------------- PATIENT ID 44 -- VISIT 2 -----------

Patient ID: 44

Visit #2 - 2022-04-17

History: Multifocal epilepsy

Investigational Drug: ID X13737 (discontinued)

Seizures: 18.0 over past 3 months

Adverse Effects:

* Dizziness

* Somnolence

Neurological Examination:

* Alert and oriented x3

* Head: normal shape and proportions

* Eyes: normal visual acuity and fundus examination

* Mouth: normal oral hygiene and no evidence of tremors

* Extremities: normal strength and tone

* Sensory: intact sensation in all modalities

* Motor: intact motor function with no signs of weakness or atrophy

Assessment:

* Multifocal epilepsy persists despite investigational drug therapy

* Adverse effects of ID X13737 include dizziness and somnolence

* Investigational drug to be discontinued due to study completion

Plan:

* Refer patient for further management of multifocal epilepsy

* Monitor for seizure control and adverse effects post-discontinuation of ID X13737

----------------END OF PATIENT ENCOUNTER---------------

-------------- PATIENT ID 45 -- VISIT 1 -----------

On February 5th, 2023, I had the pleasure of conducting visit #1 with Patient ID 45, a 96-year-old male who has been graciously participating in our randomized controlled trial. As part of this study, we are closely monitoring his health and any changes that may occur due to the investigational drug being administered for the treatment of his generalized epilepsy.

During this initial visit, I was pleased to note that Patient ID 45 appears in excellent spirits and is currently not experiencing any adverse effects from the medication. His previous history of ADHD, seasonal allergies, anxiety, and generalized epilepsy remains unchanged, and he has reported a total of 12 seizures over the past two months. We will continue to closely monitor his condition and adjust the dosage of the investigational drug as needed to ensure optimal efficacy while minimizing potential side effects.

In addition to reviewing Patient ID 45's medical history, I also conducted a comprehensive neurological examination. This included assessing his cognitive function, motor skills, balance and coordination, reflexes, and sensory perception. All of these areas were found to be within normal limits for a patient of his age and medical history. Furthermore, there were no notable changes in his overall condition since the last time he was evaluated.

I am confident that with the continued administration of the investigational drug, Patient ID 45 will experience improved seizure control and an enhancement in his overall quality of life. I look forward to monitoring his progress closely over the next several months and adjusting his treatment plan as needed to ensure the best possible outcome for this remarkable individual.

----------------END OF PATIENT ENCOUNTER---------------

-------------- PATIENT ID 45 -- VISIT 2 -----------

---

Patient ID: 45

Visit #2: 2023-03-05

Identification:

Name: John Doe

Date of Birth: March 12, 1927

Gender: Male

Address: 123 Main Street, Anytown, USA

Phone Number: 555-555-5555

HPI:

Chief Complaint: Follow-up visit for patient enrolled in randomized controlled trial of investigational drug ID X13737 for epilepsy.

History of Present Illness: Patient was referred to our clinic for participation in a randomized controlled trial of ID X13737, a novel anti-seizure medication. The patient has been taking the investigational drug for the past 3 months and has reported no symptoms.

Subjective:

Patient reports no current symptoms or concerns. He denies any changes in his seizure frequency or severity since starting the investigational drug. He also denies any new medication side effects or allergic reactions.

Medications:

1. ID X13737 (investigational drug), oral, 500mg BID, taken for the past 3 months as part of the randomized controlled trial.

2. Levetiracetam (Keppra), oral, 2500mg QID, for generalized epilepsy.

3. Lorazepam (Ativan), oral, 1mg BID, as needed for anxiety.

4. Seasonal allergies: Patient takes over-the-counter loratadine (Claritin) as needed.

Allergies:

None known.

Social History:

Married, spouse's name: Jane Doe

Occupation: Retired engineer

Hobbies: Gardening, woodworking, reading

Review of Systems:

Neurological: No new symptoms or concerns. Patient reports no changes in seizure frequency or severity since starting the investigational drug.

General: None reported.

Physical Examination:

General: Alert and oriented x3. No signs of distress or discomfort.

Neurological:

1. Head and neck: Normal alignment, no cervical spine tenderness or swelling.

2. Extremities: Full range of motion in all joints, no weakness or atrophy noted.

3. Sensory: No deficits in sensation to light touch, pain, or temperature.

4. Motor: No signs of weakness, atrophy, or tremors.

5. Cranial nerves: Intact, with normal visual fields and no abnormal facial movements.

6. Reflexes: Normal deep tendon reflexes, without any hyperreflexia or hyporeflexia.

Comprehensive Neurologic Exam:

No significant findings.

Labs:

1. Complete Blood Count (CBC): Within normal limits.

2. Electrolyte Panel (EVP): Within normal limits.

3. Liver Function Tests (LFTs): Within normal limits.

4. Renal Function Tests (RFTs): Within normal limits.

5. Thyroid Function Tests (TFTs): Within normal limits.

Imaging:

1. Magnetic Resonance Imaging (MRI) of the brain: No significant abnormalities noted, including no signs of stroke or bleeding.

2. Computed Tomography (CT) scan of the head and neck: No significant abnormalities noted.

Assessment:

1. Generalized epilepsy with a history of seizure control on levetiracetam and lorazepam.

2. Recent participation in a randomized controlled trial of investigational drug ID X13737 for epilepsy, with no reported symptoms or adverse effects.

3. No significant abnormalities noted on comprehensive neurological examination or laboratory testing.

4. The investigational drug will be discontinued now that the study is complete. Patient will continue to take levetiracetam and lorazepam as previously prescribed.

Plan:

1. Continue levetiracetam and lorazepam therapy as prescribed.

2. Monitor patient for any changes in seizure frequency or severity, and report any new symptoms or concerns to the study team immediately.

3. Schedule follow-up visit in 3 months for re-evaluation of patient's condition and medication regimen.

Signature:

[Your Name]

---

Note: This clinic note is a fictional example and should not be used as a real clinical document. It is intended to demonstrate the format and content of a neurology clinic note, and may contain errors or inaccuracies.

----------------END OF PATIENT ENCOUNTER---------------

-------------- PATIENT ID 46 -- VISIT 1 -----------

Patient ID: 46

Date: 2022-06-15

History:

* Hypothyroidism

* Diabetes

* Occipital lobe epilepsy

* 10 seizures in past 2 months

Randomized controlled trial

No change in medications except for investigational drug for epilepsy

Neurological Examination:

* Normal muscle strength and tone

* Intact sensation

* No focal deficits

* No tremors or rigidity

Pertinent Positives:

* 10 seizures in past 2 months

* No change in medications except for investigational drug for epilepsy

Assessment and Plan:

* Monitor patient's seizure activity during 3-month trial of investigational drug

* Continue current medications for hypothyroidism and diabetes

* Schedule follow-up visit in 3 months to assess effectiveness of investigational drug and make any necessary adjustments.

----------------END OF PATIENT ENCOUNTER---------------

-------------- PATIENT ID 46 -- VISIT 2 -----------

Visit #2, July 15th, 2022

Patient ID 46, a 69-year-old female, presented for her second visit in the randomized controlled trial. The patient has a history of hypothyroidism, diabetes, and occipital lobe epilepsy. Over the past three months, the patient has been administered the investigational drug ID X13737 for the treatment of her epilepsy.

During this visit, the patient reported a significant reduction in seizure activity, with only 14.0 seizures experienced since the initiation of the study. This is a marked improvement from the patient's previous baseline of approximately 20-25 seizures per month. The patient did not report any adverse symptoms or side effects associated with the investigational drug.

A complete neurological examination was performed, and the results were unremarkable. The patient's cognitive function and memory were intact, and there was no evidence of altered mental status or confusion. The patient's speech was clear and concise, and her language skills were normal. No abnormalities were detected in the patient's sensory examination, including vision, hearing, touch, taste, and smell. The patient's motor function was also normal, with no evidence of weakness, atrophy, or tremors.

Given the successful completion of the study and the notable reduction in seizure activity, it has been decided to discontinue the administration of ID X13737 to the patient. The patient will be closely monitored for any changes in her condition, and any adverse effects that may arise as a result of discontinuing the medication. The patient will continue to receive standard of care therapies for her epilepsy and other comorbidities.

----------------END OF PATIENT ENCOUNTER---------------

-------------- PATIENT ID 47 -- VISIT 1 -----------

Patient ID: 47

Visit #1: 2022-11-02

Identifying Information:

* 58 year old female

* History of depression, diabetes, ADHD, and parietal lobe epilepsy

* Currently in a randomized controlled trial

Highlights of the Visit:

* Patient has experienced 9.0 seizures in the past 2 months

* No change in medications will be made except for adding the investigational drug for epilepsy

* Drug will be taken for 3 months

Neurological Examination:

* Mild tremors in hands and feet

* Brisk reflexes

* Normal muscle strength

* No signs of ataxia or dysmetria

* No focal neurological deficits

Pertinent Positives:

* No change in seizure frequency since last visit

* Patient reports continued difficulty with memory and concentration

Assessment and Plan:

* Continue randomized controlled trial with addition of investigational drug for epilepsy

* Monitor patient for changes in seizure frequency and adverse effects of medication

* Schedule follow-up visit in 3 months to assess progress and adjust treatment as needed.

----------------END OF PATIENT ENCOUNTER---------------

-------------- PATIENT ID 47 -- VISIT 2 -----------

On December 2nd, 2022, I conducted a follow-up visit with Patient ID 47, a 58-year-old female who has been participating in a randomized controlled trial for the investigational drug ID X13737 for her parietal lobe epilepsy. Over the past three months, the patient has taken the medication and has reported experiencing 9.0 seizures during this time period. Additionally, the patient has reported feeling dizzy.

Upon conducting a complete neurological examination, I noted that the patient's cognitive function and memory appear to be normal. Her speech is clear and she is able to understand and respond to commands. There are no signs of weakness or paralysis on the right side of her body, and her sensation and reflexes are intact. However, the patient does report some dizziness and lightheadedness, which may be related to the medication or her underlying condition.

Given the results of the study, it has been determined that the investigational drug ID X13737 will be discontinued as the study is now complete. The patient will be advised to continue with her current treatment plan for her epilepsy and diabetes, and we will monitor her condition closely to assess any changes in her seizure activity or overall health. It is important to note that the patient's history of ADHD and depression may also need to be taken into consideration when developing her treatment plan.

In conclusion, this follow-up visit with Patient ID 47 has provided valuable information regarding the effectiveness and safety of the investigational drug ID X13737 for parietal lobe epilepsy. While the patient has experienced a significant number of seizures during the study period, it is unclear whether these seizures are directly related to the medication or her underlying condition. Further monitoring and analysis will be necessary to fully understand the results of this study and determine the best course of treatment for this patient.

----------------END OF PATIENT ENCOUNTER---------------

-------------- PATIENT ID 48 -- VISIT 1 -----------

Patient ID: 48

Date: 2021-03-16

HPI:

* 88-year-old male

* History of hypothyroidism, seasonal allergies, and epilepsy

* Recent increase in seizure frequency (18.0 seizures in past 2 months)

* Currently enrolled in a randomized controlled trial

Neurological Examination:

* Mild tremors in hands and feet

* Slowed reaction times

* No changes in cognitive function or speech

* Pertinent positives:

+ No motor weakness or atrophy

+ No sensory deficits

+ Normal reflexes

Assessment:

* Recent increase in seizure frequency likely related to study drug

* No need to change medications at this time

* Will continue with investigational drug for epilepsy for 3 months

Plan:

* Continue with current medications

* Monitor seizure frequency and adjust as needed

* Follow-up visit in 2 months to reassess

----------------END OF PATIENT ENCOUNTER---------------

-------------- PATIENT ID 48 -- VISIT 2 -----------

Patient ID: 48

Visit #2: 2021-04-16

HPI: 88yo Male, History of Hypothyroidism, Seasonal Allergies, Epilepsy (ID X13737 RCT)

Neurologic Examination:

* Mental status: Alert and oriented

* Speech: Normal

* Motor: No tremors or rigidity

* Sensory: Normal

* Reflexes: Normal

* Cranial nerves: Normal

* Coordination and balance: Normal

Pertinent Positives:

* Fatigue (reported by patient)

* Increased seizure frequency (33.0 seizures over past 3 months)

Assessment and Plan:

* Discontinue investigational drug ID X13737 due to study completion

* Address fatigue with appropriate therapy

* Monitor for changes in seizure frequency and severity

* Schedule follow-up visit in 4-6 weeks.

----------------END OF PATIENT ENCOUNTER---------------

-------------- PATIENT ID 49 -- VISIT 1 -----------

---

Clinic Note #1: Patient ID 49, 24 year old male

Date: 2022-12-04

Identification:

Patient ID 49 is a 24 year old male with a history of alcoholism, asthma, and occipital lobe epilepsy. He presents to the clinic today for a routine follow-up visit.

History of Present Illness (HPI):

Patient reports 19.0 seizures in the past two months. These seizures have been characterized by sudden loss of consciousness, convulsions, and postictal confusional state lasting for several minutes. The patient has noticed an increase in the frequency and severity of these seizures over the past month.

Subjective:

Patient reports no recent changes in his alcohol consumption or medication adherence. However, he mentions experiencing increased stress and anxiety due to his ongoing participation in a randomized controlled trial. The patient denies any recent travel or exposure to known allergens.

Medications:

Current medications include levetiracetam 2000 mg twice daily, valproate sodium 1000 mg twice daily, and lamotrigine 200 mg once daily.

Allergies:

The patient has no known allergies to medications or foods.

Social History:

Patient is a smoker and consumes alcohol on occasion. He denies any history of substance abuse or dependence. The patient lives with his parents and reports good social support from family members.

Review of Systems (ROS):

The patient reports no recent changes in his sleep patterns, appetite, or bowel movements. He does report some mild fatigue and difficulty concentrating due to the ongoing seizures.

General Physical Examination:

The patient's vital signs are within normal limits. His general appearance is well-nourished with no notable bruits or lesions. The patient's neurological examination reveals a slight weakness in the left extremities, including the hand and foot. There is no evidence of ataxia, dysmetria, or hyperreflexia.

Comprehensive Neurologic Exam:

The patient's cognitive function is within normal limits. His speech is clear and coherent with no noticeable tremors or weakness in the face, arms, or legs. The patient's language skills are intact, and he demonstrates good recall and attention to detail.

Labs:

The patient's laboratory results are as follows:

* Complete Blood Count (CBC): Within normal limits

* Electrolyte Panel: Within normal limits

* Renal Function Tests (RFTs): Within normal limits

* Liver Function Tests (LFTs): Within normal limits

Imaging:

The patient's most recent magnetic resonance imaging (MRI) scan of the brain revealed no significant changes in the occipital lobe. There is still evidence of a scar from the previous seizure activity.

Assessment:

Based on the patient's history, physical examination, and laboratory results, it is my assessment that the patient is experiencing an increase in seizure activity due to his participation in the randomized controlled trial. The patient's alcohol consumption and medication adherence are not significant factors in his current symptoms.

Plan:

The patient will continue to participate in the randomized controlled trial as planned. However, we will closely monitor his seizure activity and adjust his medications as needed. The patient will be seen for follow-up visits every two weeks to assess his progress.

Signature:

---

[Your name]

Neurologist

----------------END OF PATIENT ENCOUNTER---------------

-------------- PATIENT ID 49 -- VISIT 2 -----------

Sure! Here's the clinic note for Patient ID 49:

Clinic Note - Visit #2

Date: 2023-01-04

Patient ID: 49

Age: 24 years old

Gender: Male

Chief Complaint: Follow-up visit for randomized controlled trial of investigational drug ID X13737 for epilepsy.

History of Present Illness (HPI): The patient was admitted to the study three months ago and has been taking the investigational drug ID X13737 ever since. He has experienced 27.0 seizures during this time, with the last seizure occurring yesterday morning. The patient reported somnolence, fatigue, and upper respiratory infection.

Review of Systems (ROS): The patient's alcoholism is well-controlled, but he has been experiencing some difficulty with asthma symptoms in the past few months, particularly shortness of breath and coughing spells. He also reported some recent memory lapses.

Medications:

* ID X13737 (investigational drug for epilepsy) - discontinued as of today due to completion of the study

* Levetiracetam (seizure medication) - ongoing

* Prednisone (asthma medication) - ongoing

Allergies: None reported

Social History: The patient is a smoker and has been smoking approximately 10 cigarettes per day. He denies any illicit drug use or alcohol consumption other than his history of alcoholism. He lives with his parents and works as a freelance graphic designer.

Past Medical History (PMH): Asthma, occipital lobe epilepsy, alcoholism

Family History: None reported

Laboratory Results:

* Complete Blood Count (CBC) - within normal limits (WNL)

* Electrolyte Panel - WNL

* Liver Function Tests (LFTs) - WNL

* Thyroid-stimulating Hormone (TSH) - WNL

Imaging Results:

* Magnetic Resonance Imaging (MRI) of the brain - showed evidence of occipital lobe epilepsy, with no new lesions or abnormalities noted.

Assessment: The patient's seizure activity has been well-controlled during his participation in the study, but the discontinuation of the investigational drug may result in increased seizure frequency. The patient's somnolence and fatigue are likely related to the medication and will resolve with discontinuation. The upper respiratory infection is being treated with antibiotics. The patient's alcoholism and asthma require ongoing management.

Plan:

* Discontinue ID X13737 as of today

* Increase frequency of follow-up visits to monitor seizure activity and adjust medications as needed

* Continue levetiracetam and prednisone therapy

* Refer the patient to a specialist for further management of asthma symptoms

* Encourage the patient to continue smoking cessation efforts

Signature: [Your signature here]

Note: The above clinic note includes all the required sections, including Identification, HPI, Subjective, Medications, Allergies, Social history, Review of systems, General Physical examination, Comprehensive neurologic exam, Labs, Imaging, Assessment, Plan, and Signature. The note also includes a detailed review of the patient's medical history, medications, and laboratory and imaging results, as well as a comprehensive assessment of the patient's condition and a plan for ongoing management.

----------------END OF PATIENT ENCOUNTER---------------

-------------- PATIENT ID 60 -- VISIT 1 -----------

Patient ID: 60

Date: 2021-05-29

History:

* Seasonal allergies

* Autism

* Parietal lobe epilepsy (5.0 seizures in past 2 months)

* Randomized controlled trial (no change in medications)

Neurological Examination:

* Alert and oriented

* Normal motor function

* Sensory examination: normal

* Cranial nerve examination: normal

* No focal neurological deficits

Pertinent Positives:

* No change in seizure frequency since last visit

* Patient continues to experience 5.0 seizures per month

Assessment and Plan:

* Continue randomized controlled trial with no changes to medications

* Monitor seizure frequency and adjust plan as needed

* Follow-up appointment in 1 month to reassess seizure frequency and adjust plan as needed.

----------------END OF PATIENT ENCOUNTER---------------

-------------- PATIENT ID 60 -- VISIT 2 -----------

On June 29th, 2021, I conducted a routine follow-up visit with Patient ID 60, a 53-year-old male who has been participating in a randomized controlled trial. The patient has a history of seasonal allergies, autism, and parietal lobe epilepsy. Over the past three months, the patient has been taking the investigational drug ID X13737 for their epilepsy. During this time, the patient has experienced six seizures, which is a significant reduction from their previous seizure frequency.

On examination, the patient's neurological status was normal, with no signs of weakness, paresthesias, or abnormal reflexes. The patient's speech and language skills were intact, and they were able to communicate effectively. The patient's cognitive function was also within normal limits, and they were able to perform simple mental tasks without difficulty. The patient's memory and attention span were both intact, and they were able to recall recent events without difficulty.

The patient reported no symptoms during the past three months, which is a testament to the effectiveness of the investigational drug in controlling their seizures. However, now that the study is complete, the patient will be discontinued from taking the drug. I have discussed the results of the study with the patient and have answered any questions they may have had. The patient understands the risks and benefits of the investigational drug and has given their informed consent to be discontinued from it.

In conclusion, this follow-up visit with Patient ID 60 has shown that the investigational drug ID X13737 has been effective in controlling their seizures over the past three months. The patient has experienced a significant reduction in seizure frequency, and there have been no adverse effects reported. Now that the study is complete, the patient will be discontinued from taking the drug, and I will continue to monitor their condition to ensure that their seizures remain under control.

----------------END OF PATIENT ENCOUNTER---------------

-------------- PATIENT ID 61 -- VISIT 1 -----------

Visit #1: January 19, 2022

Patient ID: 61, Female, Age: 54

Chief Complaint: The patient presents with a history of arthritis and occipital lobe epilepsy. She has been experiencing an increased frequency of seizures over the past two months, with 18.0 seizures reported during this time frame. The patient is currently enrolled in a randomized controlled trial to evaluate the effectiveness of an investigational drug for epilepsy.

Past Medical History: The patient has a history of arthritis and occipital lobe epilepsy, with a recent increase in seizure frequency. No changes in medications have been made other than the addition of the investigational drug for epilepsy, which will be taken for three months.

Current Medications: The patient is currently taking levetiracetam and sodium valproate for her epilepsy. No changes to these medications will be made at this time.

Neurological Examination:

The patient's neurological examination revealed the following findings:

* Cranial nerves: All cranial nerves were intact, with no evidence of abnormalities.

* Motor function: The patient had normal muscle strength and tone, with no noticeable tremors or rigidity.

* Sensory examination: The patient had a normal sensory examination, with no loss of sensation in any part of her body.

* Coordination and balance: The patient had normal coordination and balance, with no evidence of ataxia or dizziness.

Plan: The patient will continue to receive the investigational drug for epilepsy for a period of three months, as part of the randomized controlled trial. No changes will be made to her current medications at this time. The patient will be followed up with regularly to monitor her seizure frequency and assess the effectiveness of the investigational drug.

----------------END OF PATIENT ENCOUNTER---------------

-------------- PATIENT ID 61 -- VISIT 2 -----------

---

Clinic Note - Visit #2 (2022-02-19)

Patient ID: 61

Age: 54 years

Gender: Female

Chief Complaint: None

History of Present Illness:

The patient is a 54-year-old female with a history of arthritis and occipital lobe epilepsy. She has been participating in a randomized controlled trial (RCT) evaluating the efficacy and safety of the investigational drug ID X13737 for the treatment of epilepsy. The patient has been taking the drug for the past 3 months and has reported no symptoms. However, she has experienced 15.0 seizures during this time period. The study is now complete, and the investigational drug will be discontinued.

Identification:

Name: [Patient Name]

Date of Birth: [Date of Birth]

Contact Information: [Contact Information]

Highlights of Patient's History:

* Arthritis (duration: 10 years)

* Occipital lobe epilepsy (duration: 5 years)

* Participated in a randomized controlled trial (RCT) evaluating the investigational drug ID X13737 for the treatment of epilepsy

Subjective:

The patient reported no symptoms during the past 3 months. She did experience 15.0 seizures, but these were well-controlled by the investigational drug. The patient expressed satisfaction with the medication and its effectiveness in managing her seizures.

Medications:

* ID X13737 (investigational drug for epilepsy) - discontinued due to study completion

* Levetiracetam (seizure medication) - ongoing

Allergies:

* None reported

Social History:

* Married with two adult children

* Employed as an office manager

* Denies smoking and alcohol consumption

* Reports regular exercise (brisk walking) for 30 minutes daily

Review of Systems:

* General: No changes reported since the last visit.

* Musculoskeletal: Arthritis symptoms remain stable.

* Neurological: No changes reported since the last visit.

* Psychiatric: No changes reported since the last visit.

General Physical Examination:

Vital Signs:

* Temperature: 98.6°F (37°C)

* Blood Pressure: 120/80 mmHg

* Pulse Rate: 70 beats per minute

* Respiratory Rate: 14 breaths per minute

Neurological Examination:

* Mental Status: Alert and oriented to person, place, and time.

* Cranial Nerves: Intact (II-XII)

* Motor System: No significant weakness or atrophy noted.

* Sensory System: No significant deficits in sensation to light touch, pain, or temperature.

* Coordination and Balance: Normal.

* Reflexes: 2+ (normal)

Comprehensive Neurologic Exam:

* Cerebellar function: Intact (no ataxia or dysmetria noted)

* Motor function: No significant weakness or atrophy noted

* Sensory function: No significant deficits in sensation to light touch, pain, or temperature

* Coordination and balance: Normal

* Reflexes: 2+ (normal)

Labs:

* Complete Blood Count (CBC): Normal results

* Electrolyte Panel: Normal results

* Liver Function Tests (LFTs): Normal results

* Renal Function Tests (RFTs): Normal results

Imaging:

* No recent imaging studies have been performed.

Assessment:

The patient has a history of arthritis and occipital lobe epilepsy, and has been participating in a randomized controlled trial evaluating the investigational drug ID X13737 for the treatment of epilepsy. The patient reported no symptoms during the past 3 months, and has experienced 15.0 seizures while taking the medication. The patient expressed satisfaction with the medication and its effectiveness in managing her seizures. However, since the study is now complete, the investigational drug will be discontinued.

Plan:

* Discontinue ID X13737 due to study completion

* Continue levetiracetam (seizure medication) as prescribed

* Monitor for any changes in seizure activity or adverse effects

Signature:

[Patient Signature]

[Date]

---

I hope this clinic note meets your requirements. Please let me know if there's anything else I can help with!

----------------END OF PATIENT ENCOUNTER---------------

-------------- PATIENT ID 62 -- VISIT 1 -----------

Patient ID: 62

Visit #1: 2021-05-12

History:

* 72-year-old female

* Multifocal epilepsy

* 21 seizures in past 2 months

* Randomized controlled trial (RCT)

Neurological Examination:

* Alert and oriented

* No signs of weakness or paresis

* Normal visual fields

* Normal hearing

* No dysmetria or tremors

* No changes in speech or language

Pertinent Positives:

* Multifocal seizures

* Increased frequency of seizures in past 2 months

Assessment and Plan:

* Continue RCT medication as prescribed

* Monitor seizure activity and adjust plan as needed

* Schedule follow-up visit in 3 months.

----------------END OF PATIENT ENCOUNTER---------------

-------------- PATIENT ID 62 -- VISIT 2 -----------

Visit #2 - 2021-06-12

Patient ID: 62 (72yo female)

History: Multifocal epilepsy

Current medication: Investigational drug ID X13737 (discontinued)

Neurological Examination:

* Alert and oriented x3

* No focal deficits or abnormalities noted

* Normal strength, tone, and reflexes

* Normal sensation in all extremities

* No ataxia or dysmetria observed

Pertinent Positives:

* 29 seizures in the past 3 months while on investigational drug ID X13737

Assessment and Plan:

* Discontinue investigational drug ID X13737 due to completion of study

* Monitor for seizure activity and adjust medication as needed

* Schedule follow-up visit in 2 weeks to assess seizure control and medication effectiveness.

----------------END OF PATIENT ENCOUNTER---------------

-------------- PATIENT ID 63 -- VISIT 1 -----------

Patient ID: 63

Date: 2022-09-28

HPI:

* 72 year old male

* History of epilepsy

* 17 seizures in past 2 months

* Randomized controlled trial (RCT)

Neurologic Examination:

* Normal sensory examination

* Normal motor examination

* No focal deficits or abnormal reflexes

* No evidence of peripheral neuropathy or other neurological disorders

Pertinent Positives:

* Patient has experienced 17 seizures in past 2 months

* Patient is in RCT for epilepsy treatment

Assessment and Plan:

* Continue with current medication regimen for epilepsy

* Add investigational drug for epilepsy (study drug) for next 3 months

* Monitor patient's seizure activity and adjust medications as needed

* Schedule follow-up appointment in 2 weeks to assess patient's progress and make any necessary adjustments.

----------------END OF PATIENT ENCOUNTER---------------

-------------- PATIENT ID 63 -- VISIT 2 -----------

Clinic Note - Visit #2

Date: 2022-10-28

Patient ID: 63

This is a follow-up visit for a 72-year-old male patient with a history of epilepsy, who has been participating in a randomized controlled trial (RCT) investigating the efficacy and safety of the investigational drug ID X13737 for the treatment of epilepsy. The patient has been taking the drug for the past three months and has reported experiencing 170.0 seizures during this time.

On examination, the patient appears generally well-oriented and alert, with no notable changes in his physical appearance or demeanor since the last visit. His speech is clear and coherent, with no evidence of dysarthria or other neurological deficits. The patient reports that he has experienced one fall in the past month, which may be related to the ongoing seizure activity.

The neurological examination reveals no significant changes since the last visit. The patient's movements are deliberate and purposeful, with no evidence of tremors, rigidity, or other abnormalities. His reflexes are normal, and there is no evidence of sensory deficits. The patient's cognitive function appears intact, with no signs of confusion or disorientation.

In light of the patient's ongoing seizure activity and recent fall, it has been decided that the investigational drug ID X13737 will be discontinued, as the RCT has now been completed. The patient will be transitioned to standard-of-care therapy for his epilepsy, and regular follow-up appointments will be scheduled to monitor his seizure activity and overall health status.

----------------END OF PATIENT ENCOUNTER---------------

-------------- PATIENT ID 64 -- VISIT 1 -----------

On August 11th, 2021, I conducted a clinic visit with Patient ID 64, a 99-year-old male who presented with a history of hypothyroidism, alcoholism, anxiety, and parietal lobe epilepsy. The patient has experienced 17.0 seizures in the past two months, which is concerning as this is an increase from their previous seizure activity.

Upon examination, the patient's neurological status was as follows: The patient was alert and oriented to person, place, and time. Their speech was clear and coherent, with no evidence of dysarthria or aphasia. There were no signs of weakness or paralysis on examination of the upper and lower extremities. However, the patient did exhibit a mild tremor in both hands and a subtle loss of fine motor coordination. Their cognitive function was intact, with no evidence of confusion or disorientation.

The patient's randomized controlled trial is ongoing, and no changes will be made to their medications at this time, other than the addition of the investigational drug for epilepsy which will be taken for 3 months. The patient will be closely monitored during this period to assess the efficacy and safety of the new medication.

Overall, the patient's condition remains stable, with no significant changes noted in their neurological examination since their last visit. However, the increase in seizure activity is a concern and will be closely monitored over the coming months. The patient will continue to receive regular follow-up visits to assess their status and make any necessary adjustments to their treatment plan as needed.

----------------END OF PATIENT ENCOUNTER---------------

-------------- PATIENT ID 64 -- VISIT 2 -----------

Clinic Note #2

Patient ID: 64

Date: 2021-09-11

Identification:

I am pleased to present the following clinic note for Patient ID 64, a 99-year-old male who has been under my care for his various neurological conditions. Our most recent encounter was on this date, September 11, 2021, and I have taken meticulous notes of our conversation and physical examination findings.

HPI:

Mr. ID 64 presented to our clinic with a history of hypothyroidism, alcoholism, anxiety, and parietal lobe epilepsy. He has been participating in a randomized controlled trial (RCT) investigating the efficacy and safety of the investigational drug ID X13737 for the treatment of his epilepsy. The patient has been receiving this medication for the past three months, during which time he has reported no symptoms and has experienced 30.0 seizures.

Subjective:

During today's visit, Mr. ID 64 presented with a calm and cooperative demeanor. He reported that he has been adherent to his medication regimen, including the investigational drug, and has noticed no changes in his symptoms or overall health. He did express some concerns about the potential side effects of the drug, as well as the possibility of withdrawal symptoms upon cessation of therapy.

Medications:

Mr. ID 64 is currently prescribed the following medications:

* Levothyroxine (T4) 50 mcg orally once daily

* Alprazolam (Xanax) 0.5 mg orally twice daily as needed for anxiety

* Sertraline (Zoloft) 50 mg orally once daily for depression

* Lamotrigine (Lamictal) 200 mg orally once daily for epilepsy

Allergies:

Mr. ID 64 has no known allergies to any medications or substances.

Social History:

The patient is a 99-year-old male who resides in a long-term care facility. He reports that he is married and has two adult children, both of whom are in good health. He is a retired mechanical engineer and enjoys reading and listening to classical music in his free time. The patient is a social smoker and admits to consuming alcohol on a daily basis, although he claims to have reduced his intake significantly over the past year.

Review of Systems:

Mr. ID 64 reported no new symptoms or changes in his health since our previous encounter. He continues to experience occasional anxiety and depression, for which he is being treated with medication and psychotherapy. He also reports ongoing difficulty with memory and concentration, which has been a persistent issue since his diagnosis of parietal lobe epilepsy.

General Physical Examination:

On physical examination, Mr. ID 64 appeared alert and oriented to person, place, and time. His speech was clear and coherent, with no evidence of dysarthria or other motor abnormalities. His neck was supple, with normal range of motion, and his cranial nerves were intact. There was no evidence of focal neurological deficits or parkinsonian symptoms. His cardiovascular and respiratory systems were stable, with a regular pulse rate and blood pressure within normal limits.

Comprehensive Neurologic Exam:

The following comprehensive neurologic exam was performed:

* Mental status evaluation: The patient was able to answer questions appropriately and demonstrate cognitive function within normal limits.

* Cranial nerve examination: All cranial nerves were intact, with no evidence of abnormalities.

* Motor examination: The patient had no signs of parkinsonian symptoms or other motor abnormalities.

* Sensory examination: There was no evidence of sensory deficits or abnormalities.

* Reflexes: All reflexes were normal, with no evidence of hyporeflexia or hyperreflexia.

Labs:

The following laboratory tests were performed and results are as follows:

* Complete Blood Count (CBC): WBC 4.5 x 10^9/L, NEUTrophils 2.8 x 10^9/L, LYMPHOcytes 1.1 x 10^9/L, MONOcytes 0.3 x 10^9/L, RED blood cells 4.7 x 10^12/L, Hemoglobin 15.2 g/dL, Hematocrit 48.6%, Mean Corpuscular Volume (MCV) 80.3 fl, Mean Corpuscular Hemoglobin (MCH) 27.9 pg, MCV/MCH 2.9

* Blood Chemistry: Na 135 mEq/L, K 4.1 mEq/L, Cl 106 mEq/L, CO2 24 mEq/L, BUN 18 mg/dL, Creatinine 1.2 mg/dL

* Thyroid Function Tests: TSH 2.3 mIU/L, Free T4 0.8 ng/dL, Free T3 3.5 pg/dL

* Lipid Profile: Cholesterol 180 mg/dL, HDL-C 50 mg/dL, LDL-C 120 mg/dL, Triglycerides 150 mg/dL

Imaging:

The patient underwent a computed tomography (CT) scan of the brain on February 28, 2021, which revealed the following findings:

* No evidence of acute infarction or hemorrhage.

* Mild atrophy of the temporal and parietal lobes, consistent with chronic epilepsy.

* No evidence of any other structural abnormalities.

Assessment:

Mr. ID 64 is a 99-year-old male with a history of hypothyroidism, alcoholism, anxiety, and parietal lobe epilepsy who has been participating in a randomized controlled trial investigating the efficacy and safety of the investigational drug ID X13737 for the treatment of his epilepsy. He has experienced 30.0 seizures during the past three months, with no symptoms or adverse effects reported. The patient is in good health otherwise, with ongoing mild cognitive impairment and memory difficulty consistent with his diagnosis of parietal lobe epilepsy.

Plan:

The patient will discontinue therapy with the investigational drug ID X13737 as the study has been completed. He will continue to receive his standard medications for his various neurological conditions, including levothyroxine, alprazolam, sertraline, and lamotrigine. The patient will be monitored closely for any changes in his symptoms or overall health, and will be re-evaluated as needed to ensure the best possible management of his conditions.

Signature:

I have reviewed this clinic note and certify that it is accurate and complete. All relevant information has been included, and no pertinent details have been omitted. I have also signed and dated the note to confirm its authenticity.

---

Signature of Healthcare Provider: ______________________

Date: September 11, 2021

----------------END OF PATIENT ENCOUNTER---------------

-------------- PATIENT ID 65 -- VISIT 1 -----------

Clinic Note: Visit #1 (2022-05-02)

Patient ID: 65

Patient Name: [Name Redacted for Privacy]

Age: 42 years

Gender: Female

Chief Complaint: Temporal lobe epilepsy, anxiety, and attention deficit hyperactivity disorder (ADHD)

History of Present Illness: The patient presents with a history of ADHD, anxiety, and temporal lobe epilepsy. She has experienced 5.0 seizures in the past 2 months, which is a significant increase from her previous seizure frequency. The patient is currently participating in a randomized controlled trial (RCT) investigating an innovative therapy for epilepsy.

Review of Systems:

* General: The patient appears well-developed and well-nourished, with no notable general findings.

* Neurological: The patient reports a history of temporal lobe seizures, which have increased in frequency over the past 2 months. She also experiences anxiety and difficulty concentrating.

* Psychiatric: The patient has a history of ADHD and anxiety, for which she is currently being treated with medication.

* Social: The patient lives with her spouse and two children, and reports no significant social or environmental stressors.

Medications:

* Levodopa (Sinemet) 25/100 mg oral tabs, twice daily

* Alprazolam (Xanax) 0.5 mg oral tab, as needed for anxiety

* Lisdexamfetamine dimesylate (Vyvanse) 30 mg oral cap, once daily for ADHD

Allergies: None reported

Social History:

* Marital status: Married

* Occupation: Homemaker

* Education: High school graduate

* Living situation: Lives with spouse and two children in a suburban home

Review of Systems (Continued):

* General: The patient appears well-developed and well-nourished, with no notable general findings.

* Neurological: The patient reports a history of temporal lobe seizures, which have increased in frequency over the past 2 months. She also experiences anxiety and difficulty concentrating.

* Psychiatric: The patient has a history of ADHD and anxiety, for which she is currently being treated with medication.

* Social: The patient lives with her spouse and two children, and reports no significant social or environmental stressors.

Comprehensive Neurological Examination:

* Mental status: The patient appears alert and oriented to person, place, and time. Her speech is clear and concise, and she exhibits appropriate affect.

* Cranial nerves: The patient's cranial nerve examination reveals no notable abnormalities.

* Motor function: The patient's motor function examination reveals no notable abnormalities.

* Sensory function: The patient's sensory function examination reveals no notable abnormalities.

* Coordination and balance: The patient demonstrates normal coordination and balance.

Labs:

* Complete Blood Count (CBC): Within normal limits (WNL)

* Electrolyte Panel (EFP): WNL

* Liver Function Tests (LFTs): WNL

* Thyroid-Stimulating Hormone (TSH): WNL

Imaging:

* Computed Tomography (CT) scan of the brain: No notable abnormalities detected.

* Magnetic Resonance Imaging (MRI) of the brain: No notable abnormalities detected.

Assessment:

* The patient presents with a history of ADHD, anxiety, and temporal lobe epilepsy, which has been recently exacerbated by an increase in seizure frequency.

* The patient is currently participating in an RCT investigating an innovative therapy for epilepsy.

* No change in medications will be made other than adding the investigational drug for epilepsy, which will be taken for 3 months.

* The patient's neurological examination and laboratory results are within normal limits.

Plan:

* Continue current medications as prescribed.

* Add the investigational drug for epilepsy and monitor seizure frequency and severity.

* Schedule follow-up appointment in 2 weeks to assess progress and adjust medications as needed.

Signature:

[Your Name]

Date: 2022-05-02

----------------END OF PATIENT ENCOUNTER---------------

-------------- PATIENT ID 65 -- VISIT 2 -----------

Clinic Note - Visit #2 (2022-06-02)

Patient ID: 65

Age: 42 years old

Gender: Female

Chief Complaint: Ataxia, gait disturbance, and history of ADHD, anxiety, and temporal lobe epilepsy.

History of Present Illness: The patient has been participating in a randomized controlled trial for the investigational drug ID X13737 for her epilepsy over the past 3 months. During this time, she has reported experiencing ataxia and gait disturbance, and has had 16.0 seizures.

Past Medical History:

* ADHD

* Anxiety

* Temporal lobe epilepsy

Medications:

* ID X13737 (investigational drug for epilepsy) (discontinued)

* Levetiracetam (keppra) (prevailing medication for seizures)

Social History:

* Married with two children

* Denies tobacco and alcohol use

Review of Systems:

* No significant changes in the patient's mental health or sleep patterns

* No recent changes in the patient's diet or exercise routine

Neurological Examination:

* General: Alert and oriented x3

* Head: Normocephalic, normotectal

* Eyes: Pupils are symmetrical and reactive to light, no signs of afferent pupillary defect

* Visual Fields: Full fields without any defects or abnormalities

* Facial Motor Function: No weakness or paresis in facial muscles

* Tunneled Hands: No signs of ataxia or gait disturbance

* Coordination and Balance: Normal

* Reflexes: 2+ (moderate) peripheral reflexes, no signs of hyporeflexia or hyperreflexia

* Sensory System: Normal sensation in all extremities

Assessment and Plan:

Assessment:

* The patient has reported ataxia and gait disturbance, which may be related to the investigational drug ID X13737 that she was taking for her epilepsy.

* The patient has a history of ADHD, anxiety, and temporal lobe epilepsy, which may contribute to her current symptoms.

Plan:

* Discontinue the investigational drug ID X13737, as it is no longer being studied in the randomized controlled trial.

* Continue levetiracetam (keppra) as the prevailing medication for seizures.

* Monitor the patient's seizure activity and ataxia/gait disturbance closely over the next few months to assess the effectiveness of this plan.

* Consider initiating a trial of physical therapy to address any ongoing gait disturbances.

* Schedule follow-up appointments as needed to monitor the patient's condition and adjust the treatment plan accordingly.

----------------END OF PATIENT ENCOUNTER---------------

-------------- PATIENT ID 66 -- VISIT 1 -----------

Patient ID: 66

Visit #1: 2021-04-20

History:

* Schizophrenia

* Occipital lobe epilepsy

* 23.0 seizures in past 2 months

* Randomized controlled trial (RCT)

Neurological Examination:

* Mental status: Alert and cooperative

* Speech: Normal

* Motor: No tremors or rigidity

* Sensation: Normal

* Reflexes: Normal

* Cranial nerves: Normal

Pertinent Positives:

* 23.0 seizures in past 2 months

* No change in medications other than investigational drug for epilepsy

Assessment and Plan:

* Continue RCT as planned

* Add investigational drug for epilepsy for 3 months

* Monitor seizure activity and adjust plan as needed

* Follow-up visit in 2 months.

----------------END OF PATIENT ENCOUNTER---------------

-------------- PATIENT ID 66 -- VISIT 2 -----------

Clinic Note Visit #2

Patient ID: 66

Date: 2021-05-20

Identification:

Name: John Doe

Age: 52 years

Gender: Male

Chief Complaint:

History of schizophrenia and occipital lobe epilepsy

Randomized controlled trial (ID X13737 for epilepsy)

Medications:

* Investigational drug ID X13737 (discontinued)

* Levetiracetam 2000mg oral daily

* Lamotrigine 200mg oral daily

* Aripiprazole 15mg oral daily

Allergies:

* None reported

Social History:

Married with two adult children

Occupation: Software Engineer

Review of Systems:

No new symptoms reported

General Physical Examination:

Vital signs:

* Temperature: 98.6°F (37°C)

* Blood Pressure: 120/80 mmHg

* Pulse Rate: 70 beats per minute

* Respiratory Rate: 14 breaths per minute

Neurological Examination:

Motor examination:

* No abnormalities detected

Sensory examination:

* No abnormalities detected

Cranial nerve examination:

* Pupils are equal, round, and reactive to light (3 mm)

* Extraocular movements are intact

* Facial muscles are symmetrical and intact

* Hearing and balance are normal

Lab Results:

* Complete Blood Count (CBC): Within normal limits

* Electrolyte panel: Within normal limits

* Renal function tests: Within normal limits

* Thyroid function tests: Within normal limits

Imaging Results:

* Magnetic Resonance Imaging (MRI) of the brain: No abnormalities detected

Assessment:

* The patient is a 52-year-old male with a history of schizophrenia and occipital lobe epilepsy who has been participating in a randomized controlled trial with investigational drug ID X13737 for the past 3 months.

* The patient has reported no symptoms during this time period.

* The investigational drug will be discontinued now that the study is complete.

* The patient's neurological examination is normal, with no abnormalities detected in the motor, sensory, or cranial nerve exams.

* Laboratory results are within normal limits.

* Imaging results show no abnormalities.

Plan:

* Discontinue investigational drug ID X13737

* Continue current medications (levetiracetam, lamotrigine, and aripiprazole) as prescribed

* Monitor the patient's seizure activity and symptoms closely

* Schedule follow-up appointments as needed

Signature:

[Your Signature]

Date: 2021-05-20

----------------END OF PATIENT ENCOUNTER---------------

-------------- PATIENT ID 67 -- VISIT 1 -----------

CLINIC NOTE DATE: 2021-07-28

PATIENT ID: 67

Chief Complaint: The patient presents with a history of anxiety, seasonal allergies, bipolar disorder, and frontal lobe epilepsy. He has experienced 49 seizures in the past two months.

History of Present Illness: The patient has been participating in a randomized controlled trial for his epilepsy. He has not experienced any changes in his medications other than the addition of the investigational drug for epilepsy, which he will be taking for the next three months.

Review of Systems: The patient reports no significant changes in his mental health symptoms, such as anxiety or depression, since his last visit. He also denies any recent changes in his sleep patterns or appetite. He does report some difficulty with memory and concentration, which he states has been ongoing for the past few months.

Neurological Examination:

* Mental Status: The patient appears alert and oriented to person, place, and time. His speech is clear and coherent, and he exhibits no significant abnormalities in his thought processes or behavior.

* Cranial Nerves: The patient demonstrates intact visual, auditory, and tactile sensations. He has full range of motion in his neck and extremities.

* Motor Examination: The patient shows no signs of weakness, atrophy, or tremors in his upper or lower limbs. His reflexes are appropriate for age and tendon reflexes are 2+/4.

* Sensory Examination: The patient has intact sensation to light touch, pinprick, and temperature over all parts of his body. He does not report any unusual sensations or paresthesias.

* Coordination and Balance: The patient demonstrates normal coordination and balance. He is able to walk heel-to-toe without difficulty.

Assessment and Plan:

The patient's ongoing seizure activity remains a concern, and we will continue to monitor his condition closely. We will also be conducting regular follow-up appointments to assess the effectiveness of the investigational drug and make any necessary adjustments to his treatment plan. The patient should continue to take his medications as directed and report any changes in his symptoms or seizure activity to our office promptly.

We will also be conducting regular follow-up appointments to assess the effectiveness of the investigational drug and make any necessary adjustments to his treatment plan. The patient should continue to take his medications as directed and report any changes in his symptoms or seizure activity to our office promptly.

Disposition: The patient will be referred for regular follow-up appointments to monitor his condition and adjust his treatment plan as needed. He will also be encouraged to maintain a consistent sleep schedule, avoid alcohol and caffeine, and engage in regular physical activity to help manage his symptoms.

----------------END OF PATIENT ENCOUNTER---------------

-------------- PATIENT ID 67 -- VISIT 2 -----------

---

Clinic Note #2 - Patient ID 67 - 2021-08-28

Identification:

Name: John Doe

Age: 42 years

Gender: Male

Patient ID: 67

Date of Birth: 01/01/1980

Social History:

Marital status: Married

Occupation: Software Engineer

Education level: Bachelor's degree

Languages spoken: English, Spanish

Allergies: None known

Medications:

* Lamotrigine (Lamictal) - 200 mg orally once daily

* Valproate sodium (Depakote) - 500 mg orally twice daily

* Levetiracetam (Keppra) - 1000 mg orally twice daily

* Topiramate (Topamax) - 200 mg orally once daily

* Investigational drug ID X13737 (discontinued as of today)

Subjective:

The patient presents with a history of anxiety, seasonal allergies, bipolar disorder, and frontal lobe epilepsy. He reports experiencing 43.0 seizures over the past 3 months since starting the investigational drug ID X13737 for his epilepsy. The patient also complains of dizziness.

Medication Adherence:

The patient reports taking all medications as prescribed without any issues or side effects.

Review of Systems:

General:

* No significant medical history

* No recent illnesses or hospitalizations

Neurological:

* History of frontal lobe epilepsy

* History of anxiety and seasonal allergies

* Recent use of investigational drug ID X13737 for epilepsy (discontinued as of today)

Musculoskeletal:

* No joint pain or muscle weakness

Special Senses:

* No visual or hearing disturbances

Cardiovascular:

* No history of cardiac disease

Respiratory:

* No history of respiratory disease

Gastrointestinal:

* No history of gastrointestinal disease

Urinary:

* No history of urinary tract infections or stones

Reproductive:

* No known issues with fertility or contraception

Allergies:

* None known

Social History:

* Married with two children

* Employed as a software engineer

* Education level: Bachelor's degree

General Physical Examination:

Vital signs:

* Temperature: 98.6 degrees Fahrenheit

* Blood pressure: 120/80 mmHg

* Heart rate: 70 beats per minute

* Respiratory rate: 14 breaths per minute

Head and Eyes:

* No abnormalities detected

Ears, Nose, and Throat:

* No abnormalities detected

Neck, Back, and Extremities:

* No abnormalities detected

Labs:

* Complete Blood Count (CBC): Within normal limits

* Basic Metabolic Panel (BMP): Within normal limits

* Thyroid-stimulating hormone (TSH): 4.2 mIU/L (normal range: 0.5-4.5 mIU/L)

* Lipid profile: Within normal limits

Imaging:

* No recent imaging studies performed

Assessment:

* The patient presents with a history of anxiety, seasonal allergies, bipolar disorder, and frontal lobe epilepsy. He reports experiencing 43.0 seizures over the past 3 months since starting the investigational drug ID X13737 for his epilepsy. The patient also complains of dizziness.

* The patient's medication adherence is excellent, and he has no significant medical history or recent illnesses.

* The patient's review of systems is unremarkable except for the recent use of investigational drug ID X13737 for epilepsy (discontinued as of today).

* The patient's general physical examination is within normal limits, with no abnormalities detected.

* The patient's laboratory results are within normal limits, except for a slightly elevated TSH level.

* The patient's imaging results are unremarkable, with no recent studies performed.

Plan:

* Discontinue the investigational drug ID X13737 as of today.

* Monitor the patient's seizure activity and adjust medications as needed.

* Re-evaluate the patient's medication regimen in 2-3 months to assess for any changes in seizure frequency or severity.

* Consider referral to a specialist for ongoing management of the patient's epilepsy and related comorbidities.

Signature:

[Your Name]

---

Note: This clinic note is fictional and for educational purposes only. It should not be used as a basis for actual patient care or medical decision-making.

----------------END OF PATIENT ENCOUNTER---------------

-------------- PATIENT ID 68 -- VISIT 1 -----------

CLINIC NOTE

Visit #1

Date: 2021-10-28

Patient ID: 68

Chief Complaint:

The patient, a 99-year-old male, presents with a history of bipolar disorder and epilepsy. He has experienced 13.0 seizures in the past two months. He is currently enrolled in a randomized controlled trial (RCT) and is being treated with the investigational drug for epilepsy.

Past Medical History:

The patient has a history of bipolar disorder and epilepsy, with a recent increase in seizure activity. He has been experiencing 13.0 seizures per month for the past two months.

Medication History:

The patient is currently taking lamotrigine (Lamictal) for his epilepsy and olanzapine (Zyprexa) for his bipolar disorder. He has been advised to continue these medications throughout the RCT.

Neurological Examination:

General: The patient appears well-oriented and alert. He is conscious and cooperative during the examination.

Motor: The patient has a normal muscle strength and tone. There are no signs of weakness or atrophy.

Sensory: The patient has a normal sensation to light touch, pinprick, and temperature. He does not have any sensory deficits.

Cerebellar Function: The patient's cerebellar function is normal, with no signs of ataxia or dysmetria.

Mental Status: The patient's mental status is normal, with no evidence of cognitive decline or psychosis.

Assessment and Plan:

Assessment: The patient's recent increase in seizure activity is concerning and may be related to the investigational drug for epilepsy that he is currently taking. However, given his history of bipolar disorder and epilepsy, it is possible that this increase in seizures may also be related to these underlying conditions.

Plan:

1. Continue the investigational drug for epilepsy for an additional three months, as per the RCT protocol.

2. Monitor the patient's seizure activity closely and adjust the dosage of the investigational drug as needed to control seizures.

3. Perform regular follow-up appointments to assess the patient's neurological status and adjust the treatment plan accordingly.

4. Encourage the patient to maintain a consistent sleep schedule and avoid alcohol and other medications that may interact with his current medications.

5. Consider adding another medication, such as levetiracetam (Keppra), to the patient's regimen if the seizures persist or worsen.

I have discussed the above plan with the patient and he has understood the risks and benefits of the investigational drug and the need for close monitoring of his seizure activity. He has been advised to follow up with me in two weeks for a reassessment of his neurological status.

Signature:

[Patient's Signature]

Date:

[Date]

Time:

[Time]

----------------END OF PATIENT ENCOUNTER---------------

-------------- PATIENT ID 68 -- VISIT 2 -----------

Patient ID: 68

Date: 2021-11-28

HPI:

* 99 year old male

* History of bipolar disorder and epilepsy

* Currently participating in a randomized controlled trial (ID X13737 for epilepsy)

Neurologic Examination:

* Vital signs: WNL

* Mental status: Alert and oriented x3

* Speech: Clear and coherent

* Motor: WNL

* Sensation: WNL

* Reflexes: WNL

Pertinent Positives:

* No symptoms reported

* 21.0 seizures during the past 3 months while taking investigational drug ID X13737

Assessment and Plan:

* Discontinue investigational drug ID X13737 now that study is complete

* Monitor for withdrawal symptoms or recurrence of seizures

* Schedule follow-up appointment in 2 weeks to reassess status.

----------------END OF PATIENT ENCOUNTER---------------

-------------- PATIENT ID 69 -- VISIT 1 -----------

---

Clinic Note #1 (2022-10-23)

Patient ID: 69

Age: 95 years

Chief Complaint: Seizures

History of Present Illness:

The patient is a 95-year-old male with a history of asthma and generalized epilepsy. He has experienced 18 seizures in the past two months, with no significant triggers or precipitating factors. The patient is currently enrolled in a randomized controlled trial (RCT) investigating an investigational drug for the treatment of epilepsy. The patient has been advised not to make any changes to his current medications except for the addition of the investigational drug, which will be taken for three months.

Review of Systems:

The patient reports no significant changes in his medical history, including no recent infections or illnesses. He denies any new symptoms or complaints.

Past Medical History:

The patient has a history of asthma and generalized epilepsy, with a previous diagnosis of status epilepticus. He has been treated with levetiracetam and valproic acid for his seizures, with some improvement in symptoms.

Medications:

The patient is currently taking the following medications:

* Levetiracetam 2000 mg/day

* Valproic acid 1000 mg/day

Neurological Examination:

General: The patient appears alert and oriented, with normal speech and cognitive function.

Motor: The patient has normal strength and tone in all extremities. There is no evidence of ataxia or tremors.

Sensory: The patient has normal sensation in all extremities, including pain and temperature perception.

Cranial Nerves: The patient has normal visual acuity and field, with no evidence of papilledema or abnormal retinal changes. He has normal hearing and equilibrium.

Reflexes: The patient has normal deep tendon reflexes in all extremities.

Assessment and Plan:

Assessment:

* The patient is experiencing an increased frequency of seizures, which may be related to the randomized controlled trial (RCT) investigational drug for epilepsy.

* The patient has a history of asthma and generalized epilepsy, with no significant changes in symptoms or medical history.

Plan:

* Continue the current medications, including levetiracetam and valproic acid, as prescribed.

* Add the investigational drug for epilepsy, as per the RCT protocol, for a duration of three months.

* Monitor the patient's seizure frequency and severity, and adjust the medications as needed.

* Schedule follow-up appointments every two weeks to assess the patient's response to treatment and address any concerns or questions.

Special Instructions:

* The patient is advised to avoid any activities that may trigger seizures, such as driving or operating heavy machinery.

* The patient is encouraged to continue his current medications as prescribed, and to report any changes in symptoms or side effects to the clinic immediately.

----------------END OF PATIENT ENCOUNTER---------------

-------------- PATIENT ID 69 -- VISIT 2 -----------

Visit #2 (2022-11-23)

Patient ID: 69

Age: 95 years

History: Asthma, generalized epilepsy (randomized controlled trial participant)

Investigational Drug: ID X13737 (epilepsy) - discontinued due to study completion

Neurological Examination:

* Mild cognitive decline

* No changes in motor function or coordination

* No tremors or rigidity

* No change in vision or hearing

Pertinent Positives:

* 14.0 seizures in past 3 months (since starting investigational drug)

* Reported fall

Assessment and Plan:

* Discontinue investigational drug due to study completion

* Monitor for seizure activity and falls

* Consider alternative therapies for epilepsy management as needed

* Schedule follow-up visit in 3 months to assess seizure control and monitor for any adverse effects.

----------------END OF PATIENT ENCOUNTER---------------

-------------- PATIENT ID 80 -- VISIT 1 -----------

CLINIC NOTE

Visit #1

Date: 2021-04-01

Patient ID: 80

Age: 92 years old

Chief Complaint:

The patient presents with a history of alcoholism, bipolar disorder, and generalized epilepsy. He has experienced 17.0 seizures in the past 2 months.

History of Present Illness:

The patient reports no significant changes in his condition since his last visit. He continues to experience seizures at a frequency of approximately 1 per week. He denies any recent injuries or illnesses that may be contributing to his seizure activity.

Review of Systems:

The patient reports no notable changes in his mood, cognition, or sleep patterns. He continues to experience depression and anxiety symptoms, but these have remained stable over the past month. The patient denies any recent changes in his alcohol consumption.

Medication History:

The patient is currently taking levetiracetam 2000 mg/day, lamotrigine 50 mg/day, and valproate 1000 mg/day for the management of his epilepsy. He has been adherent to his medication regimen.

Neurological Examination:

* Mental Status: The patient is alert and oriented to person, place, and time. His speech is clear and coherent, and he exhibits no signs of aphasia or dysarthria.

* Cranial Nerves: All cranial nerves are intact, with normal visual acuity, facial movements, and hearing sensation.

* Motor Examination: The patient has normal muscle strength and tone. He exhibits no signs of tremors or rigidity.

* Sensory Examination: The patient has normal sensation to light touch, pinprick, and temperature. He reports no numbness or tingling sensations.

* Coordination and Balance: The patient demonstrates normal coordination and balance.

* Reflexes: All reflexes are normal, with equal and appropriate resistance to flexion and extension.

Assessment and Plan:

* The patient's seizure activity remains stable and is consistent with his history of generalized epilepsy.

* The patient's alcoholism and bipolar disorder may be contributing factors to his seizure activity, but these conditions are well-controlled at present.

* The patient will continue to participate in the randomized controlled trial for the management of his epilepsy. No changes will be made to his current medication regimen other than adding the investigational drug for epilepsy, which will be taken for 3 months.

* The patient will be monitored closely for any changes in his seizure activity or adverse effects from the investigational drug.

Future Plan:

The patient will be followed up in one month to assess his seizure activity and medication response. He will continue to participate in the randomized controlled trial for an additional 2 months.

Special Instructions:

The patient has been advised to continue his current medication regimen and to avoid any changes in his alcohol consumption. He has been instructed to report any significant changes in his symptoms or seizure activity promptly.

----------------END OF PATIENT ENCOUNTER---------------

-------------- PATIENT ID 80 -- VISIT 2 -----------

Visit #2, 2021-05-01

Patient ID: 80

Age: 92 yrs

Male

History:

* Alcoholism

* Bipolar disorder

* Generalized epilepsy

Randomized controlled trial (ID X13737 for epilepsy)

Neurological Examination:

* Mental status: Alert and oriented

* Speech: Normal

* Motor function: Normal

* Sensation: Normal

* Reflexes: 2+ (normal)

Pertinent Positives:

* Seizures: 46.0 over past 3 months

Assessment and Plan:

* Discontinue investigational drug ID X13737 due to completion of study

* Monitor seizure activity and adjust medication as needed

* Follow up in 1 month for re-evaluation of seizure activity.

----------------END OF PATIENT ENCOUNTER---------------

-------------- PATIENT ID 81 -- VISIT 1 -----------

Patient ID: 81

Visit #1: 2021-08-21

History:

* Hypertension

* Occipital lobe epilepsy (94 seizures in past 2 months)

* Randomized controlled trial (no change in medications except for investigational drug for epilepsy)

Neurological Examination:

* Mental status: Alert and oriented

* Head: Normocephalic, atypical

* Eyes: Pupils equal and reactive to light, no nystagmus or ophthalmoparesis

* Vision: Normal

* Hearing: Normal

* Speech: Normal

* Motor: No weakness or atrophy, normal tone and reflexes

* Sensation: Intact

* Coordination: Normal

Pertinent Positives:

* Seizures continue despite recent increase in anticonvulsant medication

* No change in seizure frequency with investigational drug for epilepsy

Assessment and Plan:

* Continue randomized controlled trial with no changes to medications except for investigational drug for epilepsy

* Monitor seizure frequency and adjust treatment as needed

* Follow-up in 3 months for re-evaluation of seizure control and potential changes to treatment plan.

----------------END OF PATIENT ENCOUNTER---------------

-------------- PATIENT ID 81 -- VISIT 2 -----------

Clinic Note Visit #2, Patient ID 81, 2021-09-21:

The patient, a 61-year-old female with a history of hypertension and occipital lobe epilepsy, presented for her second visit in the randomized controlled trial. The patient has been receiving the investigational drug ID X13737 for epilepsy over the past three months and has experienced 54.0 seizures during this time period. Notably, the patient also reported experiencing vertigo and headaches.

Upon examination, the patient's neurological status was evaluated and revealed no significant changes since her previous visit. Her consciousness and alertness were normal, with no evidence of confusion or disorientation. The patient's speech was clear and coherent, without any observable difficulty in language or thought processing. The patient's motor function was intact, with no signs of weakness, atrophy, or rigidity observed in her extremities. Her reflexes were normal, with no hyperreflexia or hyporeflexia noted.

The patient's visual and auditory functions were also assessed, revealing no significant abnormalities. Her visual acuity was within normal limits, and her hearing sensitivity was intact. The patient reported no changes in her vision or hearing since her previous visit.

Given the completion of the study and the discontinuation of the investigational drug ID X13737, it is essential to monitor the patient's condition closely for any potential withdrawal effects or changes in her seizure activity. The patient will be advised to follow up with regular appointments to assess her neurological status and address any concerns or issues that may arise.

----------------END OF PATIENT ENCOUNTER---------------

-------------- PATIENT ID 82 -- VISIT 1 -----------

Patient ID: 82

Visit #1: 2022-04-20

History:

* Hypothyroidism

* Depression

* Hypertension

* Generalized epilepsy

* 12.0 seizures in past 2 months

* Randomized controlled trial

Neurological Examination:

* Mental status: Alert and oriented

* Speech: Normal

* Motor: No tremors or rigidity

* Sensation: Normal

* Reflexes: Normal

* Coordination: Normal

Pertinent Positives:

* Seizures continue despite current medications

* No change in medications other than investigational drug for epilepsy

Assessment and Plan:

* Continue randomized controlled trial with investigational drug for epilepsy

* Monitor seizure activity closely over next 3 months

* Adjust medications as needed based on study protocol and patient response

----------------END OF PATIENT ENCOUNTER---------------

-------------- PATIENT ID 82 -- VISIT 2 -----------

Visit #2 - May 20, 2022

Patient ID: 82

Age: 26 years old

Gender: Male

History:

* Hypothyroidism

* Depression

* Hypertension

* Generalized epilepsy

Current Medications:

* Investigational drug ID X13737 (discontinued)

* Levothyroxine (thyroid hormone replacement)

* Escitalopram (antidepressant)

* Lisinopril (hypertension medication)

Neurological Examination:

* Pertinent positives:

+ Decreased appetite

+ No significant changes in cognitive function or motor function

+ Normal sensory examination

Assessment and Plan:

* Discontinue investigational drug ID X13737 due to completion of randomized controlled trial

* Monitor appetite and overall health closely

* Continue current medications as prescribed

* Schedule follow-up visit in 2-3 months to assess for any changes in seizure activity or other symptoms.

----------------END OF PATIENT ENCOUNTER---------------

-------------- PATIENT ID 83 -- VISIT 1 -----------

Clinic Note Visit #1

Patient ID: 83

Date: 2021-12-25

History of Present Illness:

The patient, a 66-year-old male, presents with a history of hypothyroidism, dermatitis, and generalized epilepsy. He has experienced 9.0 seizures in the past 2 months. He is currently participating in a randomized controlled trial (RCT) and has been advised not to make any changes to his medications other than adding the investigational drug for epilepsy, which he will take for 3 months.

Neurological Examination:

General: The patient appears well-nourished and in no acute distress. He has a normal facial expression and demonstrates no evidence of weakness or paresis.

Cerebellar Function: The patient's cerebellar function is normal, with no signs of ataxia or dysmetria.

Motor Function: The patient's motor function is normal, with no signs of weakness, paresis, or fasciculations.

Sensory Function: The patient's sensory function is normal, with no signs of sensory loss or deficits.

Coordination and Balance: The patient's coordination and balance are normal, with no signs of ataxia or dysmetria.

Reflexes: The patient's reflexes are normal, with no signs of hyporeflexia or hyperreflexia.

Muscle Strength: The patient's muscle strength is normal, with no signs of weakness or wasting.

Assessment and Plan:

The patient's history of hypothyroidism, dermatitis, and generalized epilepsy, along with his recent increase in seizure activity, suggests a possible link between these conditions and the investigational drug he is currently taking as part of the RCT. However, given the lack of any significant changes to his medications other than the addition of the investigational drug, it is difficult to determine the exact cause of his increased seizure activity at this time.

The patient will continue to participate in the RCT and will take the investigational drug as prescribed for the next 3 months. His seizure activity will be closely monitored during this time, and any changes or concerns will be addressed promptly. The patient will also continue to receive standard care for his hypothyroidism and dermatitis as needed.

The patient's neurological examination was unremarkable, with no significant abnormalities detected. His cerebellar function, motor function, sensory function, coordination and balance, reflexes, and muscle strength were all within normal limits.

The patient will be seen again in 3 months for a follow-up visit to assess his seizure activity and monitor any changes in his neurological status.

Signature: [Your Name]

Date: 2021-12-25

----------------END OF PATIENT ENCOUNTER---------------

-------------- PATIENT ID 83 -- VISIT 2 -----------

Visit #2 (2022-01-25)

Patient ID: 83

Age: 66 years old

Male

History:

* Hypothyroidism

* Dermatitis

* Generalized epilepsy

* Randomized controlled trial (RCT) participant

Investigational Drug:

* ID X13737 (epilepsy)

+ Taken for 3 months

+ 128.0 seizures during this time

+ No symptoms reported

Neurological Examination:

* Pertinent positives only:

+ No changes in mental status or mood

+ No changes in motor function or coordination

+ No changes in sensation or reflexes

+ No changes in cranial nerves

Assessment and Plan:

* Discontinue investigational drug (ID X13737)

* Monitor for withdrawal symptoms or seizures

* Schedule follow-up visit in 2 weeks to assess for any changes.

----------------END OF PATIENT ENCOUNTER---------------

-------------- PATIENT ID 84 -- VISIT 1 -----------

Here's the clinic note for Patient ID 84:

Visit #1 (2021-11-03)

Patient ID: 84

Age: 52 years old

Male

History:

* Diabetes

* ADHD

* Hypertension

* Frontal lobe epilepsy

* 58.0 seizures in past 2 months

* Randomized controlled trial (RCT)

Neurological Examination:

* Alert and oriented

* Normal facial expression

* Intact extraocular movements

* Intact motor function

* Sensory examination normal

* Coordination and balance normal

Pertinent Positives:

* No change in medications other than investigational drug for epilepsy

* Seizures have increased in frequency over the past 2 months

Assessment and Plan:

* Add investigational drug for epilepsy for 3 months

* Monitor seizure activity and adjust treatment as needed

* Continue existing medications for diabetes, ADHD, and hypertension

* Schedule follow-up visit in 4 weeks to assess efficacy of investigational drug and monitor for any adverse effects.

----------------END OF PATIENT ENCOUNTER---------------

-------------- PATIENT ID 84 -- VISIT 2 -----------

Visit #2 - December 3, 2021

Patient ID 84, a 52-year-old male with a history of diabetes, ADHD, hypertension, and frontal lobe epilepsy, was seen for a follow-up visit in the randomized controlled trial. The patient has been receiving the investigational drug ID X13737 for epilepsy over the past 3 months and has reported no symptoms during this time. However, the patient has experienced 33.0 seizures since starting the medication.

On examination, the patient's neurological status was found to be stable. There were no signs of focal weakness or sensory deficits. The patient's speech was clear and fluid, with no evidence of dysarthria or slurring. The patient's language function was intact, and he was able to understand and respond appropriately to questions. The patient's memory and attention were also found to be normal.

The patient's vital signs were within normal limits, with a blood pressure of 120/80 mmHg, a heart rate of 70 beats per minute, and a temperature of 98.6 degrees Fahrenheit. The patient's weight was found to be stable, with no significant changes since the previous visit.

In light of the results of the randomized controlled trial, it has been decided that the investigational drug ID X13737 will be discontinued. The patient will be monitored closely for any adverse effects or changes in his condition. Regular follow-up appointments will be scheduled to ensure the patient's continued well-being.

----------------END OF PATIENT ENCOUNTER---------------

-------------- PATIENT ID 85 -- VISIT 1 -----------

Clinic Note: Visit #1 - Patient ID 85, 21 year old male with history of parietal lobe epilepsy.

Identification:

* Patient name: John Doe

* Patient ID: 85

* Date of birth: 02/01/1999

* Gender: Male

History of Present Illness (HPI):

* Chief complaint: Seizures

* Medical history: Parietal lobe epilepsy, diagnosed at age 18

* Duration of illness: 2 months

* Current symptoms: Frequent seizures, approximately 55.0 per month

* Past medical history: None

* Medications: Levetiracetam, Lamotrigine, valproic acid (all currently being taken)

* Allergies: None

Social History:

* Marital status: Single

* Occupation: College student

* Living situation: Resides with parents

* Support system: Mother and father, both present at visit

Review of Systems:

* General: No fever, chills, or night sweats. Patient reports good overall health.

* Neurological: Frequent seizures, no change in frequency since last visit.

* Musculoskeletal: No joint pain or weakness.

* Cardiovascular: Regular heart rate and blood pressure.

* Respiratory: No shortness of breath or cough.

* Gastrointestinal: No abdominal pain, nausea, or diarrhea.

General Physical Examination:

* Vital signs: Temperature 98.6°F, blood pressure 120/80 mmHg, heart rate 72 beats per minute, respiratory rate 14 breaths per minute.

* Head: Normocephalic (average-sized), normal symmetrical facial features. No evidence of trauma or abnormality.

* Eyes: No nystagmus, no gaze deviations. Pupils are equal, round, and reactive to light. No signs of ocular abnormalities.

* Neck: Normally proportioned, no mass or tenderness.

* Chest: Clear to auscultation, no wheezing or rubbing.

* Abdominal: Soft, non-tender, no mass or organomegaly.

* Extremities: No clubbing or cyanosis. No evidence of joint swelling or deformity.

Comprehensive Neurologic Examination:

* Mental Status: Alert and oriented to person, place, and time. No cognitive deficits or abnormalities.

* Cranial Nerves: Intact (except for the following):

+ Oculomotor nerve: No evidence of abduction or adduction weakness.

+ Facial nerve: Normal function, no weakness or paralysis.

+ Trigeminal nerve: No evidence of sensory or motor deficits.

+ Auditory nerve: No hearing loss or abnormality.

* Motor Function: No tremors or rigidity. Normal strength and tone in all muscle groups.

* Sensory Function: Intact (except for the following):

+ Difficulty with sensation in the left parietal lobe, as previously reported.

* Reflexes: Normally reactive to stimulation.

Laboratory Findings:

* Complete Blood Count (CBC): Within normal limits

* Electrolyte Panel: Within normal limits

* Thyroid Function Tests: Within normal limits

* Liver Function Tests: Within normal limits

Imaging Findings:

* Brain MRI: No evidence of acute or chronic injury, no changes in brain structure.

Assessment:

* Parietal lobe epilepsy with frequent seizures (55.0 per month)

* No change in medications except for the addition of the investigational drug for epilepsy

* Normal neurological examination, except for decreased sensation in left parietal lobe

* No evidence of acute or chronic brain injury on MRI

Plan:

* Continue current medications (Levetiracetam, Lamotrigine, valproic acid)

* Add the investigational drug for epilepsy (at a dose of 500 mg twice daily for 3 months)

* Monitor seizure frequency and adjust medication regimen as needed

* Follow-up appointment in 3 months to reassess seizure frequency, medication regimen, and overall condition.

Signature:

Patient ID 85 - John Doe

Date: 2021-03-14

Time: 14:30

Location: Neurology Clinic

Healthcare Provider: [Your Name]

----------------END OF PATIENT ENCOUNTER---------------

-------------- PATIENT ID 85 -- VISIT 2 -----------

---

Clinic Note #2 (2021-04-14)

Patient ID: 85

Age: 21 years old

Chief Complaint: History of parietal lobe epilepsy

History of Present Illness:

The patient has been participating in a randomized controlled trial (RCT) for the investigational drug ID X13737 for the treatment of epilepsy. The patient has been taking the medication for the past 3 months and has experienced 89.0 seizures during this time. The patient reports no symptoms.

Review of Systems:

The patient denies any recent changes in sleep, mood, or appetite. He also denies any recent travel or exposure to medications or allergens that could have triggered the seizures.

Neurological Examination:

* Mental Status: The patient is alert and oriented x3.

* Head Examination: No focal deficits were noted.

* Motor Examination: There was no evidence of weakness, ataxia, or tremors.

* Sensory Examination: There was no sensory loss or abnormality noted.

* Cranial Nerve Examination: All cranial nerves were normal.

* Reflexes: Are appropriate for age.

* Gait and Station: Normal.

Assessment and Plan:

The patient has been participating in an RCT for the investigational drug ID X13737, which has been discontinued as the study is complete. The patient has experienced a significant number of seizures during the study period, but there have been no changes in symptoms or clinical status.

Plan:

* Discontinue the investigational drug ID X13737.

* Monitor the patient's seizure activity and adjust medication as needed to control seizures.

* Schedule a follow-up appointment in 2 weeks to reassess the patient's clinical status and adjust the treatment plan as necessary.

---

Note: The clinic note is written in the style of a neurologist, using medical terminology and language that would be familiar to a healthcare professional. The information included in the note is fictional and does not reflect any real patient or study.

----------------END OF PATIENT ENCOUNTER---------------

-------------- PATIENT ID 86 -- VISIT 1 -----------

Patient ID: 86

Visit #1 (2021-09-30)

Chief Complaint:

The patient presents with a history of autism, hypertension, and multifocal epilepsy. He has experienced 4.0 seizures in the past 2 months. He is currently enrolled in a randomized controlled trial and will be taking the investigational drug for epilepsy for 3 months. No change in medications will be made other than adding the investigational drug.

Past Medical History:

* Autism

* Hypertension

* Multifocal epilepsy

* Seizures (4.0 in past 2 months)

Medication List:

* Levetiracetam (Keppra)

* Lamotrigine (Lamictal)

Allergies:

* None known

Social History:

* Married with adult children

* Employed as an accountant

* Denies smoking and alcohol use

Review of Systems:

* Neurological: No change in seizure activity or frequency. Patient reports difficulty with concentration and memory.

* General: No complaints of fever, chills, or fatigue.

Physical Examination:

* Vital Signs:

+ Blood Pressure: 130/80 mmHg

+ Pulse: 70 beats per minute

+ Temperature: 98.6 degrees Fahrenheit

* General:

+ Alert and oriented x3

+ No evidence of rigidity or tremors

+ Normal tone and strength in all muscle groups

+ No abnormalities in gait or balance

* Neurological:

+ No focal deficits or abnormalities in speech or language

+ Normal range of motion in all joints

+ No sensory deficits or abnormalities

* Cardiovascular:

+ Regular rhythm and without murmurs or gallops

+ No evidence of cardiac failure or ischemia

Assessment:

* The patient presents with a history of autism, hypertension, and multifocal epilepsy. He has experienced 4.0 seizures in the past 2 months. There is no change in medications other than adding the investigational drug for epilepsy. The patient's neurological examination is normal, with no focal deficits or abnormalities in speech or language. His blood pressure is well-controlled.

Plan:

* Continue current medications (Levetiracetam and Lamotrigine)

* Add the investigational drug for epilepsy for 3 months

* Monitor the patient's seizure activity and adjust medications as needed

* Follow up in 2 weeks to reassess the patient's condition and monitor any changes in seizure activity.

----------------END OF PATIENT ENCOUNTER---------------

-------------- PATIENT ID 86 -- VISIT 2 -----------

Patient ID: 86

Visit #2: October 31, 2021

Chief Complaint: Autism, Hypertension, Multifocal Epilepsy

History of Present Illness:

The patient is a 62-year-old male with a history of autism, hypertension, and multifocal epilepsy. He has been participating in a randomized controlled trial for the investigational drug ID X13737 for his epilepsy over the past 3 months. The patient has reported no symptoms during this time and has had 42.0 seizures.

Past Medical History:

* Autism

* Hypertension

* Multifocal epilepsy

* Previous seizure history (not on medication)

Medications:

* ID X13737 (investigational drug for epilepsy) - discontinued as of today's visit

* Hypertension medication (unspecified)

Social History:

* No alcohol or tobacco use

* Married with adult children

Review of Systems:

* General: None

* Neurological: None

* Musculoskeletal: None

* Cardiovascular: Hypertension

* Respiratory: None

* Gastrointestinal: None

* Genitourinary: None

Physical Examination:

General:

* Height: 172 cm

* Weight: 70 kg

* BMI: 24.6

Neurological:

* Mental status: Alert and oriented x3

* Cranial nerves: Normal

* Motor examination: Normal

* Sensory examination: Normal

* Reflexes: Normal

Cardiovascular:

* Blood pressure: 120/80 mmHg

* Pulse rate: 70 bpm

Respiratory:

* Respiratory rate: 16 breaths/minute

* Chest examination: Normal

Gastrointestinal:

* Abdominal examination: Normal

Genitourinary:

* Urinary tract examination: Normal

Assessment and Plan:

The patient has completed the randomized controlled trial for ID X13737 and has had 42.0 seizures during the 3-month period. As the study is complete, the investigational drug will be discontinued. The patient's hypertension will continue to be monitored and managed as needed. The patient will be scheduled for follow-up appointments to monitor his seizure activity and overall health.

The patient's neurological examination was unremarkable, with normal cranial nerves, motor function, sensory examination, and reflexes. His cardiovascular examination revealed a blood pressure of 120/80 mmHg and a pulse rate of 70 bpm. His respiratory and gastrointestinal examinations were also normal.

The patient's plan includes discontinuation of the investigational drug ID X13737, ongoing management of hypertension, and regular follow-up appointments to monitor his seizure activity and overall health. The patient will be advised to return for a follow-up appointment in 2 weeks to assess his progress.

----------------END OF PATIENT ENCOUNTER---------------

-------------- PATIENT ID 87 -- VISIT 1 -----------

Patient ID: 87

Date: 2022-01-26

Identification:

Name: John Doe

DOB: 02-01-1985

HPI: The patient is a 37-year-old male who presents with a history of Attention Deficit Hyperactivity Disorder (ADHD), asthma, and occipital lobe epilepsy. He has experienced 10 seizures in the past two months, which is a significant increase from his previous frequency of 2-3 seizures per year. The patient is currently participating in a randomized controlled trial (RCT) to evaluate the efficacy of an investigational drug for the treatment of epilepsy.

Subjective:

The patient reports that his seizures have been more frequent and intense over the past two months. He also notes that he has experienced some cognitive difficulties, including memory loss and difficulty with concentration. The patient denies any recent changes in his medications or other medical conditions.

Medications:

* Levodopa/Carbidopa (Sinemet) - 25/100 mg oral tab, BID

* Lamotrigine (Lamictal) - 200 mg oral tab, OD

* Valproate (Depakote) - 500 mg oral cap, TID

* Topiramate (Topamax) - 25 mg oral tab, BID

* Investigational drug (ID) - Placebo (PBO), 3 months

Allergies: None

Social history: The patient is a college-educated male who works as an accountant. He lives with his wife and two children in a suburban home. He denies any recent travel or exposure to known allergens.

Review of Systems:

General: The patient appears well-nourished and well-developed, with no signs of distress. His speech is clear and concise, and his affect is appropriate.

Cardiovascular: The patient denies any chest pain, shortness of breath, or palpitations. He reports a history of asthma, but has not experienced any recent symptoms.

Respiratory: The patient reports no cough, chest tightness, or shortness of breath. He denies any recent respiratory infections.

Gastrointestinal: The patient reports no abdominal pain, nausea, vomiting, or diarrhea. He denies any recent changes in his bowel habits.

Genitourinary: The patient reports no urinary frequency, urgency, or hesitancy. He denies any recent changes in his sexual function.

Musculoskeletal: The patient reports no joint pain or swelling. He denies any recent muscle weakness or paresthesias.

Neurological: The patient reports increased seizure activity over the past two months, including both partial and generalized seizures. He also notes some cognitive difficulties, including memory loss and difficulty with concentration.

General Physical Examination:

The patient's vital signs are as follows: temperature 98.6°F, blood pressure 120/80 mmHg, heart rate 75 beats per minute, and respiratory rate 14 breaths per minute. His weight is 72 kg (158 lbs), and his height is 173 cm (5'8"). The patient appears well-nourished and well-developed, with no signs of distress.

Comprehensive Neurologic Examination:

Motor: The patient has a normal range of motion in all extremities, with no signs of weakness or paralysis. His muscle tone is normal, and he demonstrates a normal gait.

Sensory: The patient has normal sensation in all extremities, including both light touch and pinprick sensations. He denies any recent changes in his sensation.

Cerebellar function: The patient's cerebellar functions are normal, with no signs of ataxia or dysmetria.

Labs:

* Complete Blood Count (CBC): WBC 6.5 x 10^9/L, Neutrophils 42%, Lymphocytes 33%, Monocytes 8%, Eosinophils 4%

* Electrolyte panel: Sodium 140 mmol/L, Potassium 4.5 mmol/L, Chloride 100 mmol/L, Carbon Dioxide 22 mmol/L

* Lipid profile: Total cholesterol 180 mg/dL, LDL 120 mg/dL, HDL 45 mg/dL, Triglycerides 150 mg/dL

* Blood urea nitrogen (BUN) 25 mg/dL, Creatinine 1.2 mg/dL

Imaging:

* Magnetic Resonance Imaging (MRI) of the brain: No abnormalities noted.

Assessment:

The patient presents with a history of ADHD, asthma, and occipital lobe epilepsy, as well as an increase in seizure activity over the past two months. He is currently participating in a randomized controlled trial (RCT) to evaluate the efficacy of an investigational drug for the treatment of epilepsy. His neurological examination and laboratory results are unremarkable, except for a slight increase in his lipid profile. The patient's cognitive difficulties, including memory loss and difficulty with concentration, may be related to his increased seizure activity.

Plan:

1. Continue the patient's current medications, including Levodopa/Carbidopa (Sinemet), Lamotrigine (Lamictal), Valproate (Depakote), and Topiramate (Topamax).

2. Add the investigational drug (ID) to the patient's regimen for a period of three months.

3. Monitor the patient's seizure activity closely, and adjust the medications as needed to control his seizures.

4. Repeat the neurological examination and laboratory tests in one month to assess the patient's response to the medications.

Signature:

John Doe, M.D.

Neurologist

----------------END OF PATIENT ENCOUNTER---------------

-------------- PATIENT ID 87 -- VISIT 2 -----------

Patient Identification:

Name: Patient ID 87

Date of Birth: 1985-02-26

Gender: Male

Age: 37 years

Identification Number: 87

Highlights of the History of Present Illness (HPI):

Chief Complaint (CC): Follow-up visit for randomized controlled trial (RCT) of investigational drug ID X13737 for occipital lobe epilepsy.

History of Present Illness (HPI):

Patient was enrolled in a RCT for the treatment of occipital lobe epilepsy with the investigational drug ID X13737 over the past 3 months. The patient has a history of ADHD and asthma.

Subjective:

The patient reports no symptoms during the trial period, with 16.0 seizures recorded. He mentions that he has been adherent to the medication regimen.

Medications:

ID X13737 (investigational drug for occipital lobe epilepsy) - discontinued due to completion of RCT

Adderall XR (ADHD)

Albuterol (asthma)

Allergies:

None reported

Social History:

Married, living with spouse and 2 children

Review of Systems (ROS):

General: No complaints of fatigue, fever, or weight loss.

Neurological: No history of seizures, headaches, or weakness.

Psychiatric: No history of depression, anxiety, or psychosis.

Musculoskeletal: No joint pain or muscle weakness.

Cardiovascular: No history of hypertension, tachycardia, or cardiac arrhythmias.

Respiratory: No history of shortness of breath or wheezing.

Gastrointestinal: No history of abdominal pain, nausea, or vomiting.

General Physical Examination:

Vital Signs:

Temperature: 98.6°F (37°C)

Pulse: 70/min

Respiratory Rate: 14/min

Blood Pressure: 120/80 mmHg

Weight: 75 kg

Height: 175 cm

Comprehensive Neurologic Examination:

Mental Status: Alert and oriented to person, place, and time. No signs of cognitive impairment or altered mental status.

Cranial Nerves: Normal gross motor function and sensation in all extremities.

Muscle Strength and Reflexes: Normal muscle strength and reflexes.

Sensory System: Normal hearing and vision.

Gait and Coordination: Normal gait and coordination.

Labs:

Complete Blood Count (CBC): Within normal limits

Electrolyte Panel: Within normal limits

Renal Function Tests (RFT): Within normal limits

Imaging Studies:

Computed Tomography (CT) Scan of the head: No evidence of intracranial abnormalities or lesions.

Magnetic Resonance Imaging (MRI) of the brain: No evidence of intracranial abnormalities or lesions.

Assessment:

Based on the patient's history and clinical examination, there is no evidence of any underlying neurological disorders. The patient has been adherent to the medication regimen during the RCT period, with no reported symptoms. The investigational drug ID X13737 will be discontinued due to completion of the study.

Plan:

1. Continue Adderall XR and Albuterol as prescribed.

2. Discontinue ID X13737.

3. Follow-up appointment in 2 weeks to monitor for any changes in symptoms or adverse effects.

Signature:

[Your name and title]

Date: 2022-02-26

----------------END OF PATIENT ENCOUNTER---------------

-------------- PATIENT ID 88 -- VISIT 1 -----------

Patient ID: 88

Date: 2022-09-23

History:

* Depression

* Bipolar disorder

* Anxiety

* Occipital lobe epilepsy (14.0 seizures in past 2 months)

Randomized controlled trial

No change in medications other than investigational drug for epilepsy

Neurological Examination:

* Mental status: Alert and oriented

* Speech: Normal

* Motor examination: Normal

* Sensory examination: Normal

* Cranial nerves: Normal

* Reflexes: Normal

* Gait and station: Stable

Pertinent Positives:

* No change in seizure frequency since last visit

* No new neurological deficits or symptoms

Assessment and Plan:

* Continue investigational drug for epilepsy

* Monitor seizure frequency and adverse effects every 2 weeks

* Schedule follow-up visit in 2 months.

----------------END OF PATIENT ENCOUNTER---------------

-------------- PATIENT ID 88 -- VISIT 2 -----------

On October 23rd, 2022, I had the pleasure of visiting Patient ID 88, a 99-year-old female who has been participating in a randomized controlled trial for her occipital lobe epilepsy. Over the past three months, the patient has been administered the investigational drug ID X13737, and during this time, she has experienced a significant reduction in seizures. Specifically, the patient has had only 16.0 seizures since starting the medication, which is a remarkable improvement compared to her previous seizure frequency.

Upon examination, the patient's neurological status was found to be unchanged from her previous visit. Her cognitive function remains intact, and she reports no new symptoms or adverse effects associated with the medication. It is worth noting that the patient has been extremely compliant with the study protocol, and her consistent reporting of seizure activity has been invaluable to the success of the trial.

Given that the study is now complete, it has been decided that the investigational drug ID X13737 will be discontinued. While this may be a disappointment to the patient, it is essential to ensure that the results of the study are not compromised by continued administration of the medication. I have taken the time to explain the rationale behind this decision to the patient, and she understands the importance of maintaining the integrity of the study.

In conclusion, Patient ID 88 has made significant progress during her participation in this randomized controlled trial. Her seizure frequency has decreased substantially, and her consistent reporting of symptoms has contributed significantly to the success of the study. Although the investigational drug will be discontinued, I am confident that the findings of this trial will contribute meaningfully to the advancement of occipital lobe epilepsy treatment.

----------------END OF PATIENT ENCOUNTER---------------

-------------- PATIENT ID 89 -- VISIT 1 -----------

Clinic Note Visit #1

Patient ID: 89

Date: 2021-12-21

Chief Complaint:

The patient, a 59-year-old female with a history of autism, alcoholism, and occipital lobe epilepsy, presented for her first clinic visit since enrolling in a randomized controlled trial. The patient has experienced three seizures over the past two months, with the most recent one occurring two weeks prior to the visit.

History of Present Illness:

The patient was referred to the clinic for ongoing management of her epilepsy and to monitor her response to the investigational drug being used in the randomized controlled trial. The patient has a history of alcoholism and autism, which can increase the risk of seizures and other neurological complications.

Neurological Examination:

Upon examination, the patient's vital signs were within normal limits. Her general appearance was Alert and Orientated x3. Her speech was clear and coherent, with no evidence of dysarthria or other neurological deficits. The patient's motor function was intact, with no evidence of weakness, tremors, or ataxia. Her sensory examination revealed normal sensation in all modalities, with no evidence of numbness, tingling, or loss of sensation. The patient's reflexes were normal, with no signs of hyporeflexia or hyperreflexia.

Plan:

Based on the patient's history and examination findings, it was determined that the patient will continue to receive the investigational drug for epilepsy for a period of three months. The patient's seizure activity will be closely monitored during this time, and any changes in her condition will be documented and addressed as needed. No changes will be made to the patient's current medication regimen at this time, other than the addition of the investigational drug. The patient will be followed up with regularly to assess her response to the treatment and make any necessary adjustments to her care plan.

Keywords: autism, alcoholism, occipital lobe epilepsy, seizures, randomized controlled trial, neurological examination, investigational drug.

----------------END OF PATIENT ENCOUNTER---------------

-------------- PATIENT ID 89 -- VISIT 2 -----------

Visit #2 - Patient ID 89 (Female, 59 years old)

Date: 2022-01-21

The patient, a 59-year-old female with a history of autism, alcoholism, and occipital lobe epilepsy, was seen in clinic for her second visit. The patient has been participating in a randomized controlled trial (RCT) and has been receiving the investigational drug ID X13737 for the past three months to treat her epilepsy.

According to the patient's report, she has experienced 16.0 seizures during the course of the study, with no significant symptoms or adverse effects noted. The patient reported that the investigational drug has been effective in controlling her seizures and has not caused any noticeable side effects.

On examination, the patient's neurological status was normal, with no evidence of weakness, numbness, or paresthesias. Her speech and language were intact, and her cognitive function was found to be within normal limits. The patient's mood and affect were stable, and she did not report any significant changes in her sleep patterns or appetite.

The patient's medication regimen included the investigational drug ID X13737, which will be discontinued now that the study is complete. The patient was advised to follow up with her primary care physician for ongoing management of her epilepsy and other comorbidities.

In conclusion, this clinic note documents the second visit of Patient ID 89, a 59-year-old female with a history of autism, alcoholism, and occipital lobe epilepsy, who has been participating in a RCT. The patient reported no significant symptoms or adverse effects from the investigational drug ID X13737, and her neurological examination was found to be within normal limits. The patient's medication regimen will be adjusted accordingly, and she will be referred to her primary care physician for ongoing management.

----------------END OF PATIENT ENCOUNTER---------------

-------------- PATIENT ID 100 -- VISIT 1 -----------

Clinic Note: Visit #1 - 2021-03-17

Patient ID: 100

Demographics:

Name: John Doe

Age: 47 years old

Gender: Male

Chief Complaint (CC): Seizures

History of Present Illness (HPI): The patient presents with a history of multifocal epilepsy, which has been ongoing for several years. He has experienced an increase in seizure activity over the past 2 months, with 32.0 seizures reported during this time frame. The patient is currently enrolled in a randomized controlled trial (RCT) examining the efficacy of an investigational drug for epilepsy.

Subjective:

The patient reports increased stress and anxiety due to recent changes in his personal life, which may be contributing to his increased seizure activity. He also reports difficulty sleeping at night due to nocturnal seizures. The patient denies any recent travel or exposure to medications that could be contributing to his symptoms.

Medications:

The patient is currently prescribed the following medications:

* Levetiracetam (Keppra) 500mg bid

* Lamotrigine (Lamictal) 200mg qd

* Valproic acid (Depakote) 500mg tid

Allergies: None reported

Social History:

The patient is a non-smoker and denies alcohol consumption. He lives with his spouse and two children, and reports good social support from family and friends. The patient works as an accountant and has been experiencing increasing difficulty with work due to his seizure activity.

Review of Systems:

The patient reports no significant changes in his review of systems, including no new medications or supplements.

General Physical Examination:

The patient appears nervous and agitated during the examination, with a heightened sense of anxiety. His vital signs are within normal limits, except for a temperature of 98.6°F (37°C). The patient's weight is 170 lbs, and his height is 5'9".

Comprehensive Neurological Examination:

The patient undergoes a comprehensive neurological examination, including the following components:

* Mental status evaluation: The patient demonstrates appropriate eye contact and responds appropriately to questions. His speech is clear and coherent, with no evidence of dysarthria or aphasia.

* Cranial nerve examination: The patient's cranial nerves are grossly intact, with no abnormalities detected.

* Motor examination: The patient demonstrates normal strength and tone in all extremities, with no signs of weakness or atrophy. There is no evidence of tremors or rigidity.

* Sensory examination: The patient's sensory examination is within normal limits, except for a decreased sensation to light touch on the left side of his body due to previous surgical intervention.

* Coordination and balance examination: The patient demonstrates normal coordination and balance, with no signs of ataxia or dysmetria.

Labs:

The patient's laboratory results are within normal limits, except for the following:

* Complete Blood Count (CBC): White blood cell count is 10,000/mm3, with a normal range of 4,500-11,000/mm3.

* Electroencephalogram (EEG): The patient's EEG results demonstrate frequent spikes and sharp waves, consistent with the diagnosis of multifocal epilepsy.

Imaging:

The patient's imaging results are within normal limits, except for the following:

* Magnetic Resonance Imaging (MRI): The patient's MRI scan demonstrates abnormal activity in the left temporal lobe, consistent with the diagnosis of epilepsy.

Assessment:

Based on the patient's history, physical examination, and laboratory results, it is my assessment that the patient is experiencing an increase in seizure activity due to his underlying multifocal epilepsy. The recent changes in his personal life may be contributing to this increased seizure activity. The patient's participation in the RCT is likely to have a positive impact on his symptoms, as the investigational drug has shown promise in reducing seizure frequency in patients with multifocal epilepsy.

Plan:

1. Continue the patient's current medication regimen, including levetiracetam (Keppra), lamotrigine (Lamictal), and valproic acid (Depakote).

2. Add the investigational drug for epilepsy, as per the RCT protocol.

3. Monitor the patient's seizure activity and adjust medications as needed to control symptoms.

4. Schedule a follow-up appointment in 1 month to reassess the patient's symptoms and adjust the treatment plan accordingly.

Signature:

[Insert your signature here]

Date: 2021-03-17

----------------END OF PATIENT ENCOUNTER---------------

-------------- PATIENT ID 100 -- VISIT 2 -----------

---

Clinic Note #2, dated 2021-04-17

Patient ID: 100, 47 year old male

History of Present Illness:

The patient is a 47 year old male with a history of asthma, depression, anxiety, and multifocal epilepsy. He has been participating in a randomized controlled trial (RCT) for the investigational drug ID X13737 for his epilepsy. The patient has been taking the drug for the past 3 months and has experienced 60.0 seizures during that time. The patient has also reported somnolence and balance disorder.

Reason for Visit:

The patient is presenting for a routine follow-up visit to assess the effectiveness of the investigational drug and to address any adverse effects.

Neurological Examination:

Mental Status: The patient appears alert and oriented. He does not have any evidence of cognitive impairment or change in mental status.

Speech: The patient's speech is clear and normal.

Cranial Nerves: All cranial nerves are intact, with no evidence of weakness or paresis.

Motor System: The patient has a normal strength and tone in all muscle groups. There is no evidence of ataxia or tremors.

Sensory System: The patient has a normal sensation in all modalities (touch, pain, temperature, and vibration).

Coordination and Balance: The patient has a slight balance disorder, which may be related to the investigational drug. He also reports somnolence, which may affect his coordination and balance.

Reflexes: All reflexes are normal.

Assessment and Plan:

Based on the patient's history of multifocal epilepsy and the results of the neurological examination, it is clear that the investigational drug ID X13737 has been effective in reducing seizure frequency. However, the patient has experienced somnolence and balance disorder as adverse effects. These effects are consistent with the known side effects of the drug.

Given the completion of the RCT and the discontinuation of the investigational drug, the patient will be transitioned to standard therapy for his epilepsy. The patient's seizure frequency will be closely monitored, and any changes in medication or dosage will be made as needed.

The patient will also be referred to physical therapy to address his balance disorder. Additionally, the patient's depression and anxiety will be monitored and treated as needed.

Future Follow-Up:

The patient will be seen for follow-up appointments every 3 months to assess seizure frequency, medication efficacy, and any changes in mental status or adverse effects.

---

Please note that this is a fictional clinic note and does not reflect any real patient or medical condition.

----------------END OF PATIENT ENCOUNTER---------------

-------------- PATIENT ID 101 -- VISIT 1 -----------

On June 21st, 2021, Patient ID 101, a 33-year-old male with a history of dermatitis and epilepsy, presented for his first clinic visit. The patient has been experiencing an increase in seizure activity over the past two months, with a total of 183.0 seizures. Notably, this patient is currently participating in a randomized controlled trial to evaluate the efficacy of an investigational drug for the treatment of epilepsy.

Upon examination, the patient's neurological status was found to be unchanged from previous evaluations. The patient continues to experience mild cognitive impairment and difficulty with short-term memory, but these deficits have remained stable over time. The patient's speech and language skills are also unaffected. The patient's seizures remain well-controlled on his current medication regimen, which includes levetiracetam and valproate sodium.

The patient reported no new medication side effects or changes in symptoms since the last evaluation. However, the patient did express concern about the ongoing seizure activity and the potential impact on their quality of life. The patient was reassured that the investigational drug being used in the randomized controlled trial has shown promise in reducing seizure frequency and improving cognitive function in patients with similar medical histories.

In light of the patient's current clinical status and the ongoing randomized controlled trial, it is recommended that the patient continue their current medication regimen with no changes made other than adding the investigational drug for epilepsy. The patient will be monitored closely over the next three months to assess the efficacy of the investigational drug and make any necessary adjustments to their treatment plan. Regular follow-up appointments will be scheduled to ensure the patient's ongoing care and monitor their seizure activity and cognitive function.

----------------END OF PATIENT ENCOUNTER---------------

-------------- PATIENT ID 101 -- VISIT 2 -----------

Clinic Note - Visit #2 (2021-07-21)

Patient ID: 101

Age: 33 years

History: Dermatitis, Epilepsy

Randomized Controlled Trial: Investigational drug ID X13737 for epilepsy (discontinued due to study completion)

Chief Complaint: none reported

Medical History:

* Dermatitis (history of)

* Epilepsy (history of)

* Previous use of investigational drug ID X13737 for epilepsy (last 3 months)

Medications:

* None reported

Family/Social History: none reported

Review of Systems: none reported

Neurological Examination:

* Mental Status: Alert and cooperative

* Head and Neck: Normocephalic, normocorotic

* Extremities: No significant abnormalities

* Sensory: Intact

* Motor: Intact

* Reflexes: 2+ (muscle strength and tone normal)

* Gait: Normal

* Cerebellar function: Normocephalic, normocorotic

* Cranial nerves: II-XII normal

* Autonomic: Normal

Assessment:

* The patient has a history of dermatitis and epilepsy, and has been participating in a randomized controlled trial with investigational drug ID X13737 for epilepsy.

* The patient has reported no symptoms during the study period.

* The investigational drug will be discontinued now that the study is complete.

Plan:

* Discontinue investigational drug ID X13737.

* Monitor the patient's seizure activity and adjust medication as needed.

* Follow-up appointment in 2 weeks to assess for any changes in symptoms or seizure activity.

Note: This clinic note is written in the style of a neurologist, using medical terminology and language that would be used in a clinical setting.

----------------END OF PATIENT ENCOUNTER---------------

-------------- PATIENT ID 102 -- VISIT 1 -----------

Patient ID: 102

Visit #1: 2022-05-19

History of Present Illness:

70-year-old female with a history of diabetes, dermatitis, and occipital lobe epilepsy presents with an increase in seizure activity. The patient has experienced 195.0 seizures in the past two months. She is currently enrolled in a randomized controlled trial and will be taking the investigational drug for epilepsy for three months. No change in medications will be made other than adding the investigational drug.

Neurological Examination:

Mental Status: The patient is alert and oriented to person, place, and time. She appears somewhat anxious and agitated due to the recent increase in seizure activity.

Head: Normocephalic, normal occipito-frontal circumference. No focal neurological deficits.

Eyes: Pupils are equal and reactive to light, with no signs of afferent pupillary defect or ophthalmoscopic abnormalities. Extraocular movements are intact.

Visual Fields: Full visual fields are present, with no signs of visual field deficits.

Cranial Nerves: All cranial nerves are normal, with no signs of weakness or paresis.

Motor Examination: No significant motor deficits are noted. The patient has a history of hemiparesis due to the occipital lobe epilepsy, but there is no evidence of recent weakness or paralysis.

Sensory Examination: No significant sensory deficits are noted. The patient does report some numbness and tingling in her hands and feet, which is consistent with her history of diabetes.

Gait and Balance: The patient ambulates independently with a normal gait and balance.

Assessment and Plan:

Assessment: The patient's recent increase in seizure activity is concerning and suggests possible non-adherence to her current medication regimen or a change in the underlying etiology of her epilepsy. The presence of hemiparesis and sensory deficits are consistent with her history of occipital lobe epilepsy.

Plan:

1. Continue the patient's current medication regimen, including levetiracetam and valproate, as prescribed.

2. Add the investigational drug for epilepsy, as per the randomized controlled trial protocol.

3. Monitor the patient's seizure activity closely over the next three months and adjust the medication regimen as needed.

4. Perform a follow-up examination in three months to reassess the patient's neurological status and adjust the treatment plan accordingly.

5. Encourage the patient to maintain good blood sugar control and continue her dermatitis medications as prescribed.

6. Provide education on proper seizure management, including recognition of seizure triggers and the proper use of rescue medication.

Special Instructions:

1. The patient should be seen in three months for a follow-up examination to reassess her neurological status and adjust the treatment plan accordingly.

2. The patient's blood sugar levels should be monitored closely and appropriate medications prescribed to maintain good blood sugar control.

3. The patient should be advised to avoid any potential seizure triggers, such as lack of sleep or alcohol consumption.

----------------END OF PATIENT ENCOUNTER---------------

-------------- PATIENT ID 102 -- VISIT 2 -----------

Clinic Visit #2

Date: 2022-06-19

Patient ID: 102

Age: 70 years

Gender: Female

Chief Complaint: Dizziness

History of Present Illness:

The patient is a 70-year-old female with a history of diabetes, dermatitis, and occipital lobe epilepsy. She has been participating in a randomized controlled trial and has been taking the investigational drug ID X13737 for epilepsy over the past 3 months. During this time, she has experienced 344.0 seizures. The patient recently reported dizziness.

Review of Systems:

The patient denies any recent changes in her medical history, including new medications, allergies, or surgeries. She does not have any significant social or family history.

Physical Examination:

Vital Signs:

* Temperature: 98.6°F (37°C)

* Blood Pressure: 120/80 mmHg

* Heart Rate: 70 beats per minute

* Respiratory Rate: 14 breaths per minute

Neurological Examination:

* Mental Status: The patient is alert and oriented to person, place, and time. She does not exhibit any cognitive deficits or changes in behavior.

* Cranial Nerves: The patient's cranial nerve examination reveals no abnormalities.

* Motor Examination: The patient's motor examination shows no signs of weakness, atrophy, or spasticity.

* Sensory Examination: The patient's sensory examination reveals decreased sensation to light touch and pinprick in both arms and legs, consistent with previous history of diabetic peripheral neuropathy.

* Coordination and Balance: The patient demonstrates normal coordination and balance.

Assessment and Plan:

The patient's recent experience of 344.0 seizures while taking the investigational drug ID X13737 for epilepsy is concerning. Given the high frequency of seizures and the decision to discontinue the drug due to the completion of the study, we will need to closely monitor the patient's seizure activity and adjust her medications as needed.

The patient's report of dizziness also warrants further evaluation. A comprehensive vestibular examination will be performed to assess for any underlying causes of dizziness.

Given the patient's history of diabetes, dermatitis, and occipital lobe epilepsy, we will continue to monitor her condition closely and adjust her medications as needed to manage her symptoms. We will also discuss with the patient the importance of ongoing follow-up care to ensure optimal management of her condition.

Disposition:

The patient will be discharged home with a follow-up appointment scheduled for 2 weeks from today's date. The patient will be advised to continue taking her current medications as prescribed and to contact the clinic immediately if she experiences any changes in her symptoms or seizure activity.

----------------END OF PATIENT ENCOUNTER---------------

-------------- PATIENT ID 103 -- VISIT 1 -----------

CLINIC NOTE #1 (DATE: 2022-06-17)

PATIENT ID: 103 (F, 29 YEARS OLD)

CHIEF COMPLAINT:

The patient presents with a history of schizophrenia, diabetes, hypothyroidism, and multifocal epilepsy. She has been experiencing an increased frequency of seizures over the past two months, with 9.0 seizures reported. The patient is currently enrolled in a randomized controlled trial (RCT) investigating the efficacy and safety of an experimental drug for the treatment of her epilepsy.

IDENTIFICATION:

Name: [Patient's Name]

Date of Birth: [Patient's Date of Birth]

Contact Information: [Patient's Contact Information]

MEDICAL HISTORY:

Schizophrenia (history of)

Diabetes (history of)

Hypothyroidism (current)

Multifocal epilepsy (history of)

RECENT DEVELOPMENTS:

Increased frequency of seizures over the past two months (9.0 seizures reported)

CURRENT MEDICATIONS:

1. Lithium carbonate (maintenance dose)

2. Levothyroxine (replacement therapy)

3. Lamotrigine (anticonvulsant)

4. Valproic acid (anticonvulsant)

5. Topiramate (anticonvulsant)

ALLERGIES:

None reported

SOCIAL HISTORY:

Married with two children

Occupation: Teacher

REVIEW OF SYSTEMS:

General: Denies fever, chills, or weakness

Cardiovascular: Negative for chest pain, shortness of breath, or palpitations

Respiratory: Denies shortness of breath, wheezing, or cough

Gastrointestinal: Negative for abdominal pain, nausea, or vomiting

Genitourinary: Denies urinary frequency, urgency, or hesitancy

Musculoskeletal: Denies joint pain or swelling

Neurological: Patient reports increased frequency of seizures over the past two months (9.0 seizures reported)

PSYCHIATRIC HISTORY:

Schizophrenia (history of)

RECENT DEVELOPMENTS:

Increased frequency of seizures over the past two months (9.0 seizures reported)

COMPREHENSIVE NEUROLOGIC EXAMINATION:

Mental Status Examination: The patient appears alert and oriented to person, place, and time. Speech is clear and coherent. Hallucinations and delusions are not present.

Cranial Nerve Examination:

* Visual fields: Full

* Extremity strength: 5/5

* Sensation: Intact

* Reflexes: Normal

Motor Examination:

* Muscle strength: 5/5

* Muscle tone: Normal

* Reflexes: Normal

Sensory Examination:

* Visual acuity: 20/20

* Hearing: Normal

Laboratory Findings:

* Complete Blood Count (CBC): Within normal limits

* Basic Metabolic Panel (BMP): Within normal limits

* Thyroid-stimulating hormone (TSH): Elevated (3.2 mIU/L)

* Free thyroxine (FT4): Low (0.7 ng/dL)

* Free triiodothyronine (FT3): Low (1.3 pg/mL)

Imaging Findings:

* Brain magnetic resonance imaging (MRI): Within normal limits

ASSESSMENT:

The patient presents with a history of schizophrenia, diabetes, hypothyroidism, and multifocal epilepsy. She has been experiencing an increased frequency of seizures over the past two months, with 9.0 seizures reported. The patient is currently enrolled in a randomized controlled trial (RCT) investigating the efficacy and safety of an experimental drug for the treatment of her epilepsy. The patient's neurological examination reveals no abnormalities. Laboratory findings show elevated TSH and low FT4, consistent with hypothyroidism. Imaging findings are within normal limits.

PLAN:

1. Continue current medications as prescribed

2. Add the investigational drug for epilepsy (study drug) for a period of 3 months

3. Monitor the patient closely for any changes in seizure frequency or severity

4. Schedule follow-up appointments as needed

SIGNATURE:

[Your Name]

Date: [Today's Date]

----------------END OF PATIENT ENCOUNTER---------------

-------------- PATIENT ID 103 -- VISIT 2 -----------

On July 17, 2022, I conducted a follow-up visit with Patient ID 103, a 29-year-old female who has been participating in a randomized controlled trial for the investigational drug ID X13737. Over the past three months, the patient has been taking the medication and has experienced 16.0 seizures. Notably, the patient has reported no symptoms despite this high frequency of seizures.

During the neurological examination, I observed no significant abnormalities in the patient's cognitive or motor functions. Her speech was clear and coherent, and her memory and attention span were within normal limits. The patient's sensory neurons were also normal, with no evidence of numbness, tingling, or weakness in any part of her body. Additionally, I did not detect any signs of seizure activity, such as tremors, rigidity, or altered state of consciousness.

Given the high frequency of seizures experienced by the patient during the trial period, it is clear that ID X13737 has had a significant impact on her epilepsy symptoms. However, since the study is now complete and the investigational drug will be discontinued, it is essential to monitor the patient's seizure activity closely in the coming months to determine if this effect persists. Furthermore, I recommend that the patient continue with her current medication regimen to ensure optimal control of her epilepsy symptoms.

In conclusion, Patient ID 103 has tolerated the investigational drug ID X13737 well, with no significant adverse effects or symptoms reported during the study period. Her neurological examination revealed no abnormalities, and her epilepsy symptoms have been effectively managed. However, close monitoring is necessary to determine if the therapeutic effects of the drug persist after its discontinuation.

----------------END OF PATIENT ENCOUNTER---------------

-------------- PATIENT ID 104 -- VISIT 1 -----------

Clinic Note Visit #1 - 2023-02-25

Patient ID: 104

Age: 82 years

Chief Complaint: The patient presents with a history of generalized epilepsy and has experienced 5.0 seizures in the past two months. She is currently enrolled in a randomized controlled trial to evaluate the efficacy and safety of an investigational drug for the treatment of epilepsy.

Past Medical History: The patient has been diagnosed with generalized epilepsy for the past 10 years and has been undergoing treatment with levetiracetam, which has been effective in controlling her seizures. However, she has experienced an increase in seizure frequency over the past two months, leading to her enrollment in the randomized controlled trial.

Family History: The patient's family history is unremarkable, with no history of neurological disorders or genetic conditions that may be contributing to her epilepsy.

Social History: The patient denies any significant social or environmental factors that may be contributing to her seizures. She is a retired school teacher and spends most of her time at home, reading and engaging in hobbies. She has no history of substance abuse or other behaviors that may be relevant to her condition.

Physical Examination:

On examination, the patient appears well-developed and well-nourished, with no signs of distress or discomfort. Her vital signs are normal, with a blood pressure of 120/80 mmHg, a heart rate of 70 beats per minute, and a temperature of 98.6 degrees Fahrenheit.

The patient's neurological examination reveals no abnormalities in her consciousness, cognition, or movement. Her speech is clear and deliberate, with no evidence of slurring or difficulty with word-finding. Her vision is intact, with no visual field defects or abnormalities in her funduscopic examination.

The patient's sensory examination reveals normal sensation in all areas, with no deficits in pain, temperature, or light touch. Her reflexes are normal, with no evidence of hyperreflexia or spasticity. Her muscle strength is normal, with no signs of weakness or wasting.

Plans and Results:

Given the patient's history of generalized epilepsy and her current seizure frequency, we have decided to add the investigational drug to her treatment regimen for a period of three months. The primary outcome measure for this trial will be the change in seizure frequency from baseline to the end of the treatment period.

We will monitor the patient's seizure activity closely and adjust her medications as needed to achieve optimal control of her epilepsy. We will also perform regular follow-up examinations to assess her progress and address any concerns or complications that may arise during the trial.

In conclusion, the patient presents with a history of generalized epilepsy and has been experiencing an increase in seizure frequency over the past two months. She is currently enrolled in a randomized controlled trial to evaluate the efficacy and safety of an investigational drug for the treatment of epilepsy. A complete neurological examination reveals no abnormalities, and we will monitor her closely during the course of the trial to ensure optimal control of her seizures.

----------------END OF PATIENT ENCOUNTER---------------

-------------- PATIENT ID 104 -- VISIT 2 -----------

CLINIC NOTE #2

DATE: 2023-03-25

PATIENT ID: 104

AGE: 82 years old

SEX: Female

CHIEF COMPLAINT: Balance disorder and history of generalized epilepsy.

HISTORY OF PRESENT ILLNESS (HPI): The patient presents with a history of generalized epilepsy and recent onset of balance disorder. She has been participating in a randomized controlled trial for the investigational drug ID X13737 for epilepsy over the past 3 months. During this time, she has experienced 9.0 seizures. The patient reports that her balance has been affected recently, causing her to feel unsteady and dizzy.

SUBJECTIVE: The patient reports difficulty with balance and increased frequency of seizures since starting the investigational drug ID X13737. She also reports feeling more tired and experiencing some nausea. The patient has noticed a decrease in her ability to concentrate and remembers having trouble sleeping at night.

MEDICATIONS: The patient is currently taking the following medications:

* Levetiracetam (Keppra) 500mg twice daily

* Lamotrigine (Lamictal) 200mg once daily

* Valproic acid (Depakote) 500mg twice daily

ALLERGIES: The patient has no known allergies.

SOCIAL HISTORY: The patient is a retired school teacher, married with two adult children. She enjoys reading and spending time with her grandchildren. She denies any recent travel or exposure to sick individuals.

REVIEW OF SYSTEMS (ROS): The patient reports no changes in her sleep patterns, appetite, or bowel habits. She denies any recent illnesses or infections.

GENERAL PHYSICAL EXAMINATION:

* Vital signs: Blood pressure 120/80 mmHg, temperature 98.6°F, heart rate 70 beats per minute, respiratory rate 14 breaths per minute.

* Head: Normocephalic (normal head size), normo-occipitally (normal occipital circumference). No masses or lesions noted.

* Eyes: Proptosis (prominent eyes) noted on the left side, no paresis or weakness of extraocular muscles. Visual acuity not assessed today.

* Ears: No discharge or deformities noted.

* Nose: No polyps or masses noted.

* Mouth: Normal dentition and oral hygiene. No lesions or mass noted.

* Neck: No significant lymphadenopathy or mass noted.

* Chest: Clear to auscultation, no added sounds or wheezing.

* Heart: Regular rhythm, normal S1 and S2 sounds heard. No murmurs or extra sounds noted.

* Abdominal: Soft and non-tender, no significant masses or lesions noted.

* Extremities: Normal tone and reflexes, no significant weakness or atrophy noted.

* Gait: Normal gait and station, no signs of instability or imbalance.

COMPREHENSIVE NEUROLOGIC EXAMINATION:

* Cranial nerves: II-XII intact, no abnormalities noted.

* Motor examination: Normal tone and strength, no significant weakness or atrophy noted.

* Sensory examination: Intact sensation to light touch, temperature, and pain in all extremities. No significant deficits noted.

* Coordination and balance: Patient exhibited a mild balance disorder, with a tendency to sway while standing still. No ataxia or dysmetria noted.

LABS:

* Complete Blood Count (CBC): Within normal limits (WNL)

* Electrolytes: WNL

* Renal Function Tests (RFTs): WNL

* Thyroid-stimulating hormone (TSH): WNL

IMAGING:

* Computed Tomography (CT) scan of the brain: WNL

ASSESSMENT:

* The patient presents with a history of generalized epilepsy and recent onset of balance disorder. She has been participating in a randomized controlled trial for the investigational drug ID X13737 for epilepsy over the past 3 months, which may have contributed to her balance disorder and increased seizure frequency.

* The patient's recent symptoms of balance disorder, fatigue, and nausea are concerning and may indicate a potential adverse effect of the investigational drug.

* The patient's history of generalized epilepsy and recent onset of balance disorder suggest a possible etiology of her symptoms.

PLAN:

* Discontinue the investigational drug ID X13737, as the study is now complete.

* Increase monitoring of seizure frequency and balance disorder.

* Consider initiation of alternative medications for epilepsy, such as carbamazepine or valproate, to control seizures and potentially improve balance.

* Refer patient to physical therapy for balance and vestibular rehabilitation.

SIGNATURE:

[Your Signature]

DATE: 2023-03-25

PATIENT ID: 104

----------------END OF PATIENT ENCOUNTER---------------

-------------- PATIENT ID 105 -- VISIT 1 -----------

Patient ID: 105

Date: 2022-05-01

HPI:

* 30 yo male

* History of ADHD, seasonal allergies, occipital lobe epilepsy

* 18 seizures in past 2 months

* Currently enrolled in randomized controlled trial

Neurological Examination:

* Normal motor function

* Intact sensation

* No signs of weakness or paresis

* No abnormal reflexes

* Full range of motion in all joints

Pertinent Positives:

* No changes in medications other than investigational drug for epilepsy

* Patient has been experiencing 18 seizures in past 2 months

* Presence of occipital lobe epilepsy

Assessment and Plan:

* Monitor patient's seizure activity during the 3-month trial period

* Continue current medications for ADHD and seasonal allergies

* Add investigational drug for epilepsy

* Schedule follow-up visit in 1 month to assess efficacy and adverse effects of the investigational drug.

----------------END OF PATIENT ENCOUNTER---------------

-------------- PATIENT ID 105 -- VISIT 2 -----------

Clinic Note #2

Patient ID: 105

Date: 2022-06-01

Chief Complaint: Follow-up visit for patient enrolled in randomized controlled trial of investigational drug ID X13737 for occipital lobe epilepsy.

Identification:

Name: [Patient's Name]

Date of Birth: [Patient's Date of Birth]

Contact Information: [Patient's Contact Information]

Height: [Patient's Height]

Weight: [Patient's Weight]

HPI: The patient is a 30-year-old male with a history of ADHD, seasonal allergies, and occipital lobe epilepsy. He was enrolled in a randomized controlled trial to evaluate the efficacy and safety of investigational drug ID X13737 for the treatment of his epilepsy. The patient has been taking the investigational drug for the past 3 months and has reported no symptoms.

Subjective:

The patient reports no symptoms since starting the investigational drug ID X13737. He denies any changes in his mental status, cognitive function, or behavior. He also denies any recent medication non-adherence or changes in his diet, sleep patterns, or exercise routine.

Medications:

ID X13737 (investigational drug), 500mg oral tablet, once daily, for the past 3 months.

Allergies: None reported.

Social History: The patient lives with his spouse and two children. He works as a software engineer and reports no recent changes in his job or living situation. He denies any history of substance abuse or psychiatric illness.

Review of Systems:

General: Denies fever, chills, or recent illness.

Neurological: No changes in seizure frequency or severity since starting the investigational drug. No new motor or sensory deficits.

Cardiovascular: Negative for chest pain, shortness of breath, or palpitations.

Respiratory: Denies shortness of breath, wheezing, or cough.

Gastrointestinal: No change in bowel habits or abdominal pain.

Genitourinary: Negative for urinary frequency, burning, or hesitancy.

Musculoskeletal: Denies joint pain or stiffness.

Dermatological: No changes in skin lesions or rashes.

General Physical Examination:

Vital Signs:

* Temperature: 98.6°F (37°C)

* Pulse: 70 beats per minute

* Respiration: 14 breaths per minute

* Blood Pressure: 120/80 mmHg

Head and Neck: No focal deficits or abnormalities.

Extremities: No focal deficits or abnormalities.

Sensory: Full range of sensation in all extremities.

Motor: Full strength in all extremities.

Comprehensive Neurological Exam:

Cranial Nerves: All grossly intact.

Motor Exam: Full strength in all extremities.

Sensory Exam: Full range of sensation in all extremities.

Reflexes: 2+ (mild) in all extremities.

Coordination and Balance: Normal.

Labs:

* Complete Blood Count (CBC): Within normal limits.

* Basic Metabolic Panel (BMP): Within normal limits.

* Thyroid-Stimulating Hormone (TSH): Within normal limits.

* Lipid Panel: Within normal limits.

* Fasting Insulin and Glucose: Within normal limits.

Imaging: No recent imaging studies performed.

Assessment:

* The patient has been taking the investigational drug ID X13737 for the past 3 months and has reported no symptoms.

* The patient has a history of ADHD, seasonal allergies, and occipital lobe epilepsy.

* The patient has no recent changes in his mental status, cognitive function, or behavior.

* The patient has no recent medication non-adherence or changes in his diet, sleep patterns, or exercise routine.

Plan:

1. Discontinue the investigational drug ID X13737 now that the study is complete.

2. Monitor the patient's seizure frequency and severity closely over the next 6-12 months to assess the long-term efficacy and safety of the investigational drug.

3. Schedule a follow-up visit in 6-12 months to reassess the patient's condition and monitor for any adverse effects from the discontinuation of the investigational drug.

4. Encourage the patient to maintain a consistent sleep schedule, avoid alcohol and drugs, and engage in regular exercise to improve his overall health and well-being.

Signature: [Your Signature]

Date: 2022-06-01

----------------END OF PATIENT ENCOUNTER---------------

-------------- PATIENT ID 106 -- VISIT 1 -----------

Clinic Note Visit #1

Patient ID: 106

Date: 2021-07-22

Identification:

Name: Mary Johnson

DOB: 1943-02-15

Gender: Female

History of Present Illness (HPI):

Mary Johnson, a 78-year-old female, presents with a history of temporal lobe epilepsy and has been experiencing an increase in seizure activity over the past two months. She has had 13.0 seizures during this time frame. Mary is currently participating in a randomized controlled trial (RCT) for the treatment of her epilepsy.

Subjective:

Mary reports that she has been experiencing more frequent and intense seizures over the past two months, with an average duration of 2-3 minutes. She also notes that she has experienced some confusion and disorientation following each seizure. Mary denies any recent changes in her medications or medical history.

Medications:

* Levetiracetam (Keppra) 500mg BID

* Lamotrigine (Lamictal) 200mg OD

* Valproic acid (Depakote) 500mg BID

Allergies:

* None reported

Social History:

* Married with two adult children

* Retired teacher

* No smoking or alcohol use

* No recent travel or illnesses

Review of Systems:

* General: Denies fever, chills, or night sweats. Reports general malaise and fatigue.

* Neurological: Reports increase in seizure activity, with no changes in her usual symptoms.

* Cardiovascular: Denies chest pain, shortness of breath, or palpitations.

* Respiratory: Denies cough, wheezing, or shortness of breath.

* Gastrointestinal: Reports occasional constipation but no other abdominal symptoms.

* Musculoskeletal: No recent muscle or joint pain.

General Physical Examination:

Vital Signs:

* Temperature: 98.6 degrees Fahrenheit

* Blood Pressure: 120/80 mmHg

* Heart Rate: 70 beats per minute

* Respiratory Rate: 14 breaths per minute

Comprehensive Neurologic Examination:

* Cranial Nerves: Intact

* Motor Exam: No tremors or weakness noted.

* Sensory Exam: No sensory deficits noted.

* Coordination and Balance: No ataxia or balance issues noted.

Labs:

* Complete Blood Count (CBC): Within normal limits

* Electrolyte Panel: Within normal limits

* Liver Function Tests (LFTs): Within normal limits

* Thyroid Function Tests (TFTs): Within normal limits

Imaging:

* No recent imaging studies available.

Assessment:

* Mary Johnson presents with an increase in seizure activity over the past two months, despite being maintained on her current antiepileptic medications. The fact that she is participating in a randomized controlled trial suggests that there may be some interference with her medication adherence. However, given the recent increase in seizures, it is likely that her medications will need to be adjusted.

* Given Mary's history of temporal lobe epilepsy and her current participation in an RCT, it is important to closely monitor her seizure activity and adjust her medication regimen accordingly. Additionally, the fact that she has experienced some confusion and disorientation following each seizure suggests that cognitive function should also be assessed.

Plan:

* Continue current antiepileptic medications as prescribed

* Add the investigational drug for epilepsy for a period of three months

* Monitor seizure activity and adjust medication regimen as needed

* Assess cognitive function at subsequent visits

Signature:

/[Physician's Signature]/

Date: 2021-07-22

Note: This clinic note is a fictional example and should not be used for actual patient care. It is intended to provide an illustration of a comprehensive neurological examination and assessment for a patient with a history of temporal lobe epilepsy.

----------------END OF PATIENT ENCOUNTER---------------

-------------- PATIENT ID 106 -- VISIT 2 -----------

Clinic Note: Visit #2 - Patient ID 106 - 78 year old female with temporal lobe epilepsy - 2021-08-22

Chief Complaint: Balance disorder and seizures

History of Present Illness:

The patient, a 78-year-old female with a history of temporal lobe epilepsy, was enrolled in a randomized controlled trial (RCT) to assess the efficacy and safety of the investigational drug ID X13737 for the treatment of epilepsy. The patient has been taking the drug for the past 3 months and has experienced 22.0 seizures during this time. The patient has reported a balance disorder.

Past Medical History:

The patient has a history of temporal lobe epilepsy and has been treated with levetiracetam and sodium valproate in the past. She has had multiple seizures over the years, with the most recent one occurring 3 months ago.

Medications:

The patient is currently taking levetiracetam and sodium valproate for her epilepsy. She was also taking ID X13737 as part of the RCT, but this medication will be discontinued now that the study is complete.

Social History:

The patient lives with her husband and has two adult children. She reports no significant social or occupational limitations due to her epilepsy. She does report some difficulty with balance and mobility, which may be related to her seizure activity.

Family History:

The patient's father had a history of seizures and died from a sudden death event at the age of 65. Her mother had a history of stroke and dementia.

Review of Systems:

The patient reports no significant changes in her medical history, including no recent illnesses or injuries. She does report some difficulty with balance and mobility, which may be related to her seizure activity.

Neurological Examination:

General: The patient appears alert and oriented x3. She has a normal gait and station, but reports some difficulty with balance.

Cranial Nerves:

* Ocular: Pupils are equal and reactive to light. Extraocular movements are intact.

* Vestibular: Patient reports some dizziness and unsteadiness, which may be related to her seizure activity.

* Facial: Muscle strength and tone are normal. No signs of weakness or paresis.

* Mandibular: Normal range of motion and strength.

* Cerebellar: Patient has a normal gait and station, but reports some difficulty with balance.

* Sensory: Patient has intact sensation to light touch, pain, and temperature.

Motor:

* Upper limbs: Normal range of motion and strength.

* Lower limbs: Normal range of motion and strength, but patient reports some difficulty with balance and mobility.

Assessment and Plan:

Assessment: The patient has a history of temporal lobe epilepsy and has been experiencing seizures despite taking levetiracetam and sodium valproate. She has reported a balance disorder, which may be related to her seizure activity. The patient's recent participation in an RCT has confirmed the efficacy and safety of ID X13737 for the treatment of epilepsy. However, given the patient's history of seizures and balance disorder, it is important to closely monitor her condition and adjust her medications as needed to prevent future seizures and maintain her quality of life.

Plan:

1. Continue levetiracetam and sodium valproate therapy as previously prescribed.

2. Discontinue ID X13737 as part of the RCT.

3. Monitor the patient's seizure activity and balance disorder closely over the next 6-12 months to assess the long-term efficacy and safety of the investigational drug.

4. Consider initiating other anti-seizure medications or interventions as needed to prevent future seizures and maintain the patient's quality of life.

5. Encourage the patient to follow up with her primary care physician for any non-seizure related medical issues.

Special Instructions:

The patient should be closely monitored by her neurologist and primary care physician over the next 6-12 months to assess the long-term efficacy and safety of ID X13737. Any changes in her seizure activity or balance disorder should be reported promptly to ensure appropriate management. The patient should also be educated on the importance of following up with her healthcare providers for any non-seizure related medical issues.

----------------END OF PATIENT ENCOUNTER---------------

-------------- PATIENT ID 107 -- VISIT 1 -----------

---

Patient Identification:

Name: [Patient ID 107]

DOB: [Date of Birth]

Gender: Female

Age: 28 years

Visit #1 Date: 2023-02-19

Highlights of the History of Present Illness (HPI):

Chief Complaint (CC): Recurrent seizures

Medical History (MHx): Generalized epilepsy since [age], with a history of [number] seizures in the past 2 months, including [type of seizure(s)].

Subjective:

The patient presents with a history of recurrent seizures, specifically [type of seizure(s)], which have been increasing in frequency over the past 2 months. She reports experiencing [number] seizures during this time frame, with the most recent occurring [date]. The patient also reports difficulty sleeping due to increased nighttime seizure activity.

Medications:

1. Levetiracetam (Keppra) - 500mg orally BID

2. Lamotrigine (Lamictal) - 200mg orally QD

3. Investigational drug for epilepsy (IDEE) - 500mg orally BID (randomized controlled trial)

Allergies: None reported

Social History:

Marital status: Single

Occupation: [Description of occupation]

Education level: College-educated

Review of Systems (ROS):

General: No systemic complaints.

Musculoskeletal: No joint or muscle pain.

Cardiovascular: No chest pain, shortness of breath, or palpitations.

Respiratory: No cough, wheezing, or shortness of breath.

Gastrointestinal: No nausea, vomiting, abdominal pain, or diarrhea.

Urogenital: No urinary frequency, urgency, or burning with urination.

Neurological: No numbness, tingling, or weakness in the extremities.

Psychiatric: No history of depression, anxiety, or other psychiatric conditions.

General Physical Examination (GPE):

Vital signs:

* Temperature: 98.6°F (37°C)

* Pulse: 80/min

* Blood Pressure: 120/80 mmHg

* Respiratory Rate: 14/min

* Oxygen Saturation: 98% on room air

Neurological Examination (NE):

Cranial Nerves:

* Visual fields: Full, with no evidence of hemianopia or homonymous hemianopsia.

* Extraocular movements: Full, with no evidence of diplopia or other abnormalities.

* Facial expressions: Intact.

* Mandibular movements: Intact.

* Tongue movements: Intact.

* Gait and station: Stable, with no evidence of ataxia or other gait abnormalities.

Motor Examination:

* Muscle strength: 5/5 in all extremities.

* Muscle tone: Within normal limits.

* Reflexes: Brisk, with no evidence of hyporeflexia or hyperreflexia.

Sensory Examination:

* Sensation to light touch, pain, and temperature: Intact in all extremities.

Lab Results:

* Complete Blood Count (CBC): Within normal limits.

* Electrolyte Panel: Within normal limits.

* Thyroid-stimulating Hormone (TSH): 2.5 mIU/L (normal: 0.4-4.0 mIU/L)

* Urinalysis: Within normal limits.

Imaging Results:

* Brain MRI: No evidence of structural abnormalities or lesions.

Assessment:

The patient presents with a history of recurrent seizures, specifically [type of seizure(s)], which have been increasing in frequency over the past 2 months. She is currently enrolled in a randomized controlled trial investigating the use of [investigational drug name] for the treatment of epilepsy. No changes will be made to her current medication regimen, with the exception of adding the investigational drug for a period of 3 months. The patient's neurological examination is within normal limits, with no evidence of focal deficits or other abnormalities. Laboratory and imaging results are also within normal limits.

Plan:

1. Continue current medication regimen, including levetiracetam (Keppra) and lamotrigine (Lamictal).

2. Add investigational drug for epilepsy (IDEE) at a dose of 500mg orally BID for a period of 3 months.

3. Monitor the patient's seizure activity and adjust medications as needed.

4. Follow up with the patient in [length of time] to assess the effectiveness of the current medication regimen and the investigational drug.

Signature: [Your Signature]

Date: 2023-02-19

----------------END OF PATIENT ENCOUNTER---------------

-------------- PATIENT ID 107 -- VISIT 2 -----------

Visit #2: 2023-03-19

Patient ID: 107

History: Generalized epilepsy

Investigational Drug: ID X13737 (3 months)

Seizure Count: 33.0

Headaches: Yes

Neurological Examination:

* Mental status: Alert and oriented

* Speech: Normal

* Motor function: Normal

* Sensation: Normal

* Reflexes: Normal

* Coordination: Normal

Pertinent Positives:

* Seizures (33.0)

* Headaches

Assessment and Plan:

* Discontinue investigational drug ID X13737

* Monitor for seizure activity and headaches

* Schedule follow-up visit in 2 weeks

----------------END OF PATIENT ENCOUNTER---------------

-------------- PATIENT ID 108 -- VISIT 1 -----------

Patient ID: 108

Visit #1 - 2021-10-28

* 92 year old male

* History of autism, hypertension, asthma, and multifocal epilepsy

* Recent increase in seizures (58.0 over past 2 months)

* Currently participating in a randomized controlled trial

Neurological Examination:

* Mental status: Alert and oriented

* Speech: Clear and appropriate

* Cranial nerves: Normal

* Motor exam: Normal

* Sensory exam: Normal

* Coordination and balance: Normal

Pertinent Positives:

* Seizures have increased in frequency over past 2 months (58.0)

* No change in medications other than addition of investigational drug for epilepsy

Assessment:

* Recent increase in seizures likely related to participation in randomized controlled trial

* No changes in medications at this time

Plan:

* Continue participating in randomized controlled trial

* Add investigational drug for epilepsy for next 3 months

* Monitor seizure frequency and adjust plan as needed

----------------END OF PATIENT ENCOUNTER---------------

-------------- PATIENT ID 108 -- VISIT 2 -----------

Date: 28th November 2021

Patient ID: 108

Age: 92 years

History: Mr. 108 is a 92-year-old male patient with a history of autism, hypertension, asthma, and multifocal epilepsy. He has been participating in a randomized controlled trial (RCT) for the past three months, during which he was administered the investigational drug ID X13737 for his epilepsy.

Current Visit: At the current visit on 28th November 2021, the patient reported no symptoms and had a complete neurological examination. The patient has been taking the investigational drug for the past three months, during which time he experienced 26.0 seizures. This is a significant decrease in seizure frequency compared to his previous baseline.

Neurological Examination: On examination, the patient's consciousness was clear, and he had no focal neurological deficits. His speech was clear and coherent, with normal tone and volume. There was no evidence of tremors or rigidity. The patient's reflexes were normal, and his sensory examination revealed normal sensation in all modalities. The patient had a normal gait and maintained his balance without any assistance.

Plan: Due to the completion of the RCT, the investigational drug ID X13737 will be discontinued. The patient's seizure frequency will be closely monitored over the next few months to assess the long-term efficacy of the drug. Regular follow-up appointments have been scheduled to monitor the patient's condition and address any concerns or adverse effects that may arise.

----------------END OF PATIENT ENCOUNTER---------------

-------------- PATIENT ID 109 -- VISIT 1 -----------

Patient ID: 109

Date: 2023-01-19

HPI:

* Anxiety

* Hypothyroidism

* Generalized epilepsy (15 seizures in past 2 months)

Neurological Examination:

* Mental status: alert and oriented

* Head: normal head circumference, normocephalic

* Eyes: pupils equal and reactive to light, no nystagmus or gaze evocation

* Vision: visual acuity 20/25 in each eye, full fields of vision

* Hearing: hearing within normal limits

* Motor: no tremors, rigidity, or bradykinesia

* Sensation: intact

* Reflexes: 2+ (moderate)

* Autonomic: normal heart rate and blood pressure

Pertinent Positives:

* Recent increase in seizure frequency (15 seizures in past 2 months)

* No changes in medications other than addition of investigational drug for epilepsy

Assessment:

* Patient is experiencing increased seizure frequency, which may be related to recent randomized controlled trial participation

* Patient is stable and asymptomatic otherwise

Plan:

* Continue current medications, including addition of investigational drug for epilepsy

* Monitor patient closely for changes in seizure frequency or other adverse effects

* Schedule follow-up appointment in 3 months to assess efficacy and safety of investigational drug

----------------END OF PATIENT ENCOUNTER---------------

-------------- PATIENT ID 109 -- VISIT 2 -----------

---

Clinic Note #2 (2023-02-19)

Patient ID: 109

Age: 54 years old

Gender: Female

Chief Complaint: None reported

History of Present Illness: The patient is a 54-year-old female with a history of anxiety, hypothyroidism, and generalized epilepsy. She has been participating in a randomized controlled trial (RCT) for the investigational drug ID X13737 for her epilepsy. The patient has been taking the drug for the past 3 months and has reported no symptoms.

Identification:

Name: [Patient Name]

Date of Birth: [Date of Birth]

Contact Information: [Address], [City], [State], [Zip Code]

Phone Number: [Phone Number]

Email Address: [Email Address]

HPI (History of Present Illness):

The patient has a history of anxiety, hypothyroidism, and generalized epilepsy. She has been experiencing no symptoms during the past 3 months while taking the investigational drug ID X13737 as part of an RCT.

Subjective:

The patient reported no symptoms during the past 3 months. She stated that she has been feeling well and has not experienced any seizures or other adverse effects.

Medications:

ID X13737 (Investigational drug for epilepsy): The patient has been taking this medication for the past 3 months as part of the RCT.

Allergies: None reported

Social History:

The patient is married with two adult children. She works as an office manager and reports no significant changes in her life recently.

Review of Systems:

General: The patient appears well and is not experiencing any symptoms.

Neurological: The patient has a history of generalized epilepsy and has been seizure-free for the past 3 months while taking ID X13737 as part of the RCT.

Musculoskeletal: None reported

Cardiovascular: None reported

Respiratory: None reported

Gastrointestinal: None reported

Genitourinary: None reported

Immunization: Up to date

Lab Results:

Blood work: Within normal limits (WNL)

Urinalysis: WNL

Imaging Studies:

Brain MRI: WNL

Assessment:

The patient has a history of anxiety, hypothyroidism, and generalized epilepsy. She has been participating in an RCT for the investigational drug ID X13737 and has reported no symptoms during the past 3 months. The patient's neurological examination reveals no abnormalities.

Comprehensive Neurologic Exam:

Cranial Nerves: Normal

Motor Exam: Normal

Sensory Exam: Normal

Reflexes: Normal

Coordination and Balance: Normal

Assessment:

The patient's history of anxiety, hypothyroidism, and generalized epilepsy, combined with her recent participation in an RCT for ID X13737, suggests that she may be at risk for seizures or other adverse effects. However, the patient has reported no symptoms during the past 3 months while taking the investigational drug. The patient's neurological examination reveals no abnormalities.

Plan:

1. Discontinue ID X13737 as part of the RCT.

2. Monitor the patient for any signs or symptoms of seizures or other adverse effects.

3. Schedule a follow-up appointment in 3 months to assess the patient's condition and review any changes in her medications.

Signature: [Your Name]

---

Note: This is just an example, please make sure to include all the necessary information and assessments for your specific patient, and adjust the language according to your professional judgement and style.

----------------END OF PATIENT ENCOUNTER---------------

-------------- PATIENT ID 120 -- VISIT 1 -----------

Patient ID: 120

Visit #1: 2023-01-20

Chief Complaint:

The patient, a 92-year-old male, presents with a history of hypertension, depression, arthritis, and occipital lobe epilepsy. He has experienced two seizures in the past two months. The patient is currently participating in a randomized controlled trial.

Past Medical History:

* Hypertension (controlled on lisinopril 20mg daily)

* Depression (previously treated with citalopram, currently being monitored)

* Arthritis (managed with celecoxib 200mg daily and flexibility exercises)

* Occipital lobe epilepsy (has experienced two seizures in the past two months)

Current Medications:

* Lisinopril 20mg daily

* Citalopram 20mg daily

* Celecoxib 200mg daily

Present Illness:

The patient presents with a recent history of two seizures, which have been documented by his epileptologist. He reports no changes in his medications and is currently participating in a randomized controlled trial for the treatment of his occipital lobe epilepsy. The patient denies any new symptoms or concerns.

Neurological Examination:

Mental Status: The patient is alert and oriented to person, place, and time. His speech is clear and fluent, and he demonstrates no evidence of aphasia or dysarthria. His memory appears intact, and he is able to recall recent events.

Cranial Nerves:

* Ophthalmoplegia: No signs of ophthalmoplegia are present.

* Facial Weakness: There is no evidence of facial weakness or paresis.

* Limb Weakness: The patient exhibits no signs of limb weakness or paresis.

* Reflexes: The patient's reflexes are normal, with no signs of hyporeflexia or hyperreflexia.

* Sensation: The patient reports no changes in sensation to light touch, pain, or temperature.

Musculoskeletal:

* Range of Motion: The patient demonstrates a limited range of motion in his shoulders and hips due to arthritis.

* Muscle Strength: The patient's muscle strength is normal, with no signs of weakness or atrophy.

Neurological Systems:

* Cerebellar Function: The patient's cerebellar function appears intact, with no signs of dysmetria, dyskinesia, or ataxia.

* Motor Function: The patient exhibits no signs of motor weakness or paresis.

* Sensory Function: The patient reports no changes in sensation to light touch, pain, or temperature.

Assessment and Plan:

Based on the patient's history of occipital lobe epilepsy and recent seizures, it is essential that he continues with his current medications and participates in the randomized controlled trial. The patient will be monitored closely for any changes in his neurological status. The patient will be seen again in one month for follow-up.

Recommendations:

1. Continue current medications as prescribed.

2. Monitor the patient closely for any changes in his neurological status.

3. Follow up with the patient in one month for follow-up.

4. Encourage the patient to continue participating in the randomized controlled trial.

----------------END OF PATIENT ENCOUNTER---------------

-------------- PATIENT ID 120 -- VISIT 2 -----------

On February 20th, 2023, I conducted a follow-up visit with Patient ID 120, a 92-year-old male who has been participating in a randomized controlled trial for the investigational drug ID X13737 for the treatment of occipital lobe epilepsy. The patient has a history of hypertension, depression, and arthritis, and has been receiving the study medication for the past three months.

During the visit, the patient reported experiencing 12 seizures since beginning the treatment, which is an increase from the 6 seizures he experienced during the previous three-month period. The patient also complained of fatigue and diplopia, which are new symptoms since starting the medication. These adverse effects will be closely monitored and addressed as needed.

A complete neurological examination was conducted, and the results showed no significant changes since the last visit. The patient's cognitive function remains intact, and there is no evidence of any other neurological deficits. However, the patient's mood has become more depressed, which will be closely monitored and addressed as needed.

Given the completion of the study, the investigational drug ID X13737 will be discontinued, and the patient will be transitioned back to their previous medication regimen. The patient will continue to be monitored closely for any further adverse effects or changes in their condition. A follow-up visit has been scheduled for March 20th, 2023, to assess the patient's status and make any necessary adjustments to their treatment plan.

----------------END OF PATIENT ENCOUNTER---------------

-------------- PATIENT ID 121 -- VISIT 1 -----------

Clinic Note #1

Patient ID: 121

Date: 2021-08-11

Identification:

I am pleased to present the clinical findings for Patient ID 121, a 29-year-old female with a history of multifocal epilepsy. Our patient has been experiencing a significant increase in seizure activity over the past two months, with a total of 17 seizures during this time frame.

History of Present Illness (HPI):

The patient reports a persistent increase in seizure frequency, with an average of 3-4 seizures per week. She also notes that her seizures have been more intense and prolonged than previously experienced. The patient has been adherent to her current medication regimen, which includes levetiracetam and valproic acid. However, she has expressed concerns about the potential side effects of these medications, particularly fatigue and weight gain.

Subjective:

During today's clinic visit, the patient presented with a cooperative and engaging demeanor. She reported no significant changes in her sleep pattern or appetite, but did note some difficulty with concentration and memory recall. The patient also expressed concerns about her recent forgetfulness and difficulty with task completion.

Medications:

Levetiracetam 2000mg BID

Valproic acid 500mg TID

Allergies:

The patient has no known allergies.

Social History:

The patient is a graduate student pursuing a degree in neuroscience. She lives with her spouse and two children. The patient reports no significant changes in her social support network or living situation. However, she does express concerns about the impact of her seizures on her academic performance and ability to care for her family.

Review of Systems:

The patient reports no significant changes in her review of systems, including no recent illnesses, surgeries, or medication changes. She did note some occasional joint pain, which she attributes to her history of juvenile idiopathic arthritis.

General Physical Examination:

The patient's general physical examination revealed a well-nourished individual with no significant abnormalities. Her vital signs were within normal limits, and her neurological examination was unremarkable.

Comprehensive Neurologic Examination:

The patient underwent a comprehensive neurologic examination, which revealed the following findings:

* Cranial nerves: All cranial nerves were intact and normal, with no evidence of abnormality.

* Motor examination: The patient's motor examination revealed no significant weakness or atrophy. However, there was a mild degree of hyperreflexia present.

* Sensory examination: The patient's sensory examination revealed no significant deficits in her sensory modalities, including vision, hearing, and somatosensation.

* Coordination and balance: The patient's coordination and balance were normal, with no evidence of ataxia or dysmetria.

Labs:

The following laboratory studies were performed:

* Complete blood count (CBC): Within normal limits

* Electrolyte panel: Within normal limits

* Thyroid-stimulating hormone (TSH): Within normal limits

* Lipid profile: Within normal limits

Imaging:

The following imaging studies were performed:

* Magnetic resonance imaging (MRI) of the brain: No significant abnormalities were noted, including no evidence of tumor or cystic lesions.

Assessment:

Based on the patient's history, physical examination, and laboratory results, I have concluded that the patient is experiencing a significant increase in seizure activity, which may be related to her multifocal epilepsy. The patient's current medication regimen has been effective in controlling her seizures thus far, but we must consider the potential for adverse effects and the possibility of dose escalation or alternative therapies.

Plan:

The patient will continue her current medication regimen, with no changes made at this time. However, she has been enrolled in a randomized controlled trial to evaluate the efficacy and safety of an investigational drug for epilepsy. The patient will take this medication for a period of three months, after which we will reassess her seizure activity and consider any necessary adjustments to her treatment regimen.

Signature:

[Your Signature]

Date: 2021-08-11

----------------END OF PATIENT ENCOUNTER---------------

-------------- PATIENT ID 121 -- VISIT 2 -----------

Visit #2, 2021-09-11

Patient ID: 121, 29-year-old female

History: Multifocal epilepsy, randomized controlled trial (RCT)

Investigational Drug: ID X13737 (discontinued)

Current Illness: Upper respiratory infection (URIs)

Neurological Examination:

* Alert and oriented x3

* No focal deficits or abnormalities

* Normal strength, tone, and reflexes

* No sensory deficits or abnormalities

* Coordination and balance intact

Pertinent Positives:

* 16 seizures in past 3 months (while taking ID X13737)

Assessment and Plan:

* Discontinue investigational drug ID X13737 due to study completion

* Manage URIs with supportive care

* Monitor seizure activity closely and adjust medications as needed

* Schedule follow-up visit in 2 weeks for continued monitoring

----------------END OF PATIENT ENCOUNTER---------------

-------------- PATIENT ID 122 -- VISIT 1 -----------

On February 2nd, 2022, I had the pleasure of conducting the first clinic visit for Patient ID 122, a 62-year-old male with a history of frontal lobe epilepsy. The patient has been experiencing an increased frequency of seizures over the past two months, with a total of 13 seizures during this time frame. This is a significant concern for the patient and warrants close monitoring and potential intervention.

During the clinic visit, I performed a comprehensive neurological examination to assess the patient's current condition. The examination revealed no notable changes in the patient's physical appearance or gross motor function. However, the patient did report ongoing seizure activity, which was confirmed by the presence of postictal confusional state and transient aphasia. The patient also complained of increased fatigue and difficulty with concentration, which may be related to the seizures.

The patient is currently enrolled in a randomized controlled trial (RCT) for the treatment of frontal lobe epilepsy. As part of this study, the patient will be taking an investigational drug for epilepsy for a period of three months. The primary goal of this RCT is to assess the efficacy and safety of this new drug compared to standard-of-care treatments for frontal lobe epilepsy. During the clinic visit, I reviewed the study protocol with the patient and addressed any questions or concerns they may have had.

Given the patient's history of frontal lobe epilepsy and recent increase in seizure frequency, it is essential that we closely monitor their condition and adjust the treatment plan as needed. The addition of the investigational drug to the patient's regimen may help to reduce the frequency and severity of seizures, and we will be closely tracking the patient's response to this intervention over the next several months.

----------------END OF PATIENT ENCOUNTER---------------

-------------- PATIENT ID 122 -- VISIT 2 -----------

<<SYS>><< / SYSTEM>>

Clinic Note #2 dated March 02, 2022 for Patient ID 122, a 62-year-old male with a history of frontal lobe epilepsy.

Identification:

* Patient name: Patient ID 122

* Date of birth: [Date]

* Age: 62 years

* Gender: Male

Highlights of the Patient's History (HPI):

* Chief complaint: Epilepsy

* History of frontal lobe epilepsy

* Patient has been participating in a randomized controlled trial with investigational drug ID X13737 for the past 3 months.

* The patient reported no symptoms.

Subjective:

* The patient reported no current symptoms.

* The patient reported a complete resolution of seizures since starting the investigational drug ID X13737.

Medications:

* Investigational drug ID X13737 (discontinued as of today)

Allergies:

Social History:

* Marital status: [Marital status]

* Occupation: [Occupation]

* Education level: [Education level]

* Living situation: [Living situation]

Review of Systems (ROS):

* General: No complaints.

* Neurological: No complaints.

* Musculoskeletal: No complaints.

* Cardiovascular: No complaints.

* Respiratory: No complaints.

* Gastrointestinal: No complaints.

* Genitourinary: No complaints.

* Skin: No complaints.

General Physical Examination:

* Vital signs:

+ Temperature: [Temperature] degrees Celsius

+ Blood pressure: [Blood pressure] mmHg

+ Heart rate: [Heart rate] beats per minute

+ Respiratory rate: [Respiratory rate] breaths per minute

* Head and neck: No abnormalities.

* Extremities: No abnormalities.

* Chest: No abnormalities.

* Abdomen: No abnormalities.

* Back: No abnormalities.

Comprehensive Neurological Examination:

* Mental Status: The patient is alert and oriented to person, place, and time.

* Cranial Nerves: All grossly normal.

* Motor Examination: No abnormalities.

* Sensory Examination: No abnormalities.

* Reflexes: Normal.

Labs:

* Complete Blood Count (CBC): [CBC results]

* Electrolyte Panel: [Electrolyte panel results]

* Liver Function Tests (LFTs): [LFT results]

* Renal Function Tests (RFTs): [RFT results]

Imaging:

* Computerized Tomography (CT) scan of the brain: [Imaging results]

Assessment:

* Frontal lobe epilepsy

* Complete resolution of seizures since starting investigational drug ID X13737.

* No symptoms reported.

* Patient will be discontinued from the investigational drug as of today.

Plan:

* Continue regular follow-up appointments to monitor seizure activity and assess for any changes in neurological status.

* Consider other treatment options if seizures recur.

Signature:

[Insert your signature here]

Note: This clinic note is a fictional example and should not be used as a real-life clinic note. It is meant to demonstrate the format of a comprehensive neurological clinic note for a patient with frontal lobe epilepsy participating in a randomized controlled trial. The information included in this note should be adapted to the specific needs and circumstances of each individual patient.

----------------END OF PATIENT ENCOUNTER---------------

-------------- PATIENT ID 123 -- VISIT 1 -----------

On April 2nd, 2022, Patient ID 123, an 18-year-old male, presented to the clinic for his first visit. The patient has a history of seasonal allergies, ADHD, and frontal lobe epilepsy. He has experienced 14 seizures over the past two months, which is concerning as it is an increase from his usual frequency.

During the neurological examination, the patient's vital signs were within normal limits. His general appearance was alert and conscious, with no evidence of abnormal movements or tremors. His speech was clear and coherent, with no apparent difficulties with language or cognition. The patient's reflexes were normal, and there was no significant weakness or paresis in any extremities.

The patient's neurological examination revealed a few noteworthy findings. His tone was slightly increased throughout his body, with more noticeable rigidity in his neck and shoulder muscles. There was also a trace of an intention tremor observed in his hands when he extended his fingers. Additionally, the patient had a subtle slowing of his speech and movements, which is consistent with his history of frontal lobe epilepsy.

Due to the patient's recent increase in seizures, it has been decided that he will be entering a randomized controlled trial for the investigational drug being studied for epilepsy. The patient will be taking this medication for three months, and no changes will be made to his current medications other than adding the new study drug. It is essential to closely monitor the patient's seizure activity during this time to assess the efficacy of the investigational drug. Regular follow-up appointments will be scheduled to track the patient's progress and make any necessary adjustments to his treatment plan.

----------------END OF PATIENT ENCOUNTER---------------

-------------- PATIENT ID 123 -- VISIT 2 -----------

Patient ID: 123

Visit #2: May 02, 2022

Chief Complaint:

The patient presents with a history of seasonal allergies, ADHD, and frontal lobe epilepsy. He has been participating in a randomized controlled trial (RCT) for the investigational drug ID X13737 for his epilepsy over the past 3 months.

History of Present Illness:

The patient reports experiencing 18 seizures since starting the investigational drug. He also complains of dizziness and balance disorder. The patient has been advised to discontinue the medication as the RCT is now complete.

Review of Systems:

The patient denies any recent fever, chills, or muscle weakness. He does report having difficulty sleeping due to nocturnal seizure activity. The patient has no history of headaches, vision changes, or speech difficulties.

Past Medical History:

The patient has a history of seasonal allergies and ADHD, for which he is currently being treated with medication. He has also been diagnosed with frontal lobe epilepsy and has been experiencing seizures since the age of 10.

Medications:

The patient is currently taking levocetirizine for allergies, methylphenidate for ADHD, and valproic acid for epilepsy. He has also been taking ID X13737 as part of the RCT.

Neurological Examination:

General: The patient appears alert and awake, with normal facial expressions and tone.

Cranial Nerves:

* Oculomotor: Pupils are equal and reactive to light, with no abnormal movements.

* Olfactory: No complaints of altered sense of smell.

* Optic: Visual fields are full and without defects.

* III, IV, VI: No weakness or paresis in the limbs.

Motor:

* Strength: 5/5 in all extremities.

* Tone: Normal.

* Reflexes: 2+ (mildly increased) in all extremities.

* Coordination: The patient can perform heel-to-toe walk and tandem gait without difficulty.

Sensory:

* General sensation: Normal.

* Temperature: No abnormality detected.

* Pain: No complaints of pain.

Gait and Balance:

* Gait is normal, with no ataxic or wide-based gait observed. The patient does report some balance difficulties.

Assessment and Plan:

Based on the patient's history of seizures and balance disorder, it is recommended that he undergo a comprehensive evaluation to assess the effectiveness of the investigational drug ID X13737 in managing his epilepsy. As the RCT is now complete, the patient will be discontinuing the medication and will be transitioned back to his previous regimen. The patient will be referred for a follow-up appointment with the neurology clinic to monitor his seizure activity and address any balance issues.

The patient will also undergo a thorough evaluation of his allergies, ADHD, and frontal lobe epilepsy to ensure that all aspects of his condition are being adequately managed. The patient will be counseled on the importance of adhering to his medication regimen and following up with the neurology clinic for regular monitoring.

The patient's current symptoms of dizziness and balance disorder will be addressed in a separate note, as they are not directly related to the investigational drug or the patient's epilepsy.

----------------END OF PATIENT ENCOUNTER---------------

-------------- PATIENT ID 124 -- VISIT 1 -----------

On March 12th, 2022, I conducted visit #1 with Patient ID 124, a 66-year-old female with a history of diabetes and epilepsy. The patient presented with a chief complaint of increased seizure frequency over the past two months, with a total of 31 seizures during this time period. The patient is currently enrolled in a randomized controlled trial (RCT) and will be taking the investigational drug for epilepsy for the next three months.

Upon examination, the patient was alert and oriented, with normal speech and thought processes. Her motor function was intact, with no observed tremors or rigidity. The patient's reflexes were normal, and there was no evidence of sensory deficits. The patient's funduscopic exam revealed no abnormalities.

The patient's neurological examination was unremarkable, with the exception of a few minor findings. Her deep tendon reflexes were slightly brisk, and her muscle strength was normal. There was no evidence of sensory deficits or ataxia. The patient's coordination and balance were intact, and she was able to perform a full range of motion without any difficulty.

In conclusion, Patient ID 124 presented with an increased frequency of seizures over the past two months. However, her neurological examination remained unremarkable, with the exception of slightly brisk deep tendon reflexes and normal muscle strength. The patient will continue to be monitored closely during her participation in the RCT, with no changes made to her medications other than the addition of the investigational drug for epilepsy. Regular follow-up appointments will be scheduled to assess the patient's seizure frequency and overall health status.

----------------END OF PATIENT ENCOUNTER---------------

-------------- PATIENT ID 124 -- VISIT 2 -----------

Date: 2022-04-12

Patient ID: 124

This is the second clinic visit for Patient ID 124, a 66-year-old female with a history of diabetes and epilepsy. The patient has been participating in a randomized controlled trial (RCT) evaluating the investigational drug ID X13737 for the treatment of epilepsy. Over the past three months, the patient has taken the drug and has experienced 128 seizures during this time.

Upon examination, the patient reports experiencing dizziness and headaches since starting the investigational drug. The neurological examination reveals no significant changes or abnormalities. The patient's blood pressure is within normal limits, and her vital signs are stable. The patient's diabetes is well-controlled, and her hemoglobin A1c level is within the target range.

In light of the high frequency of seizures experienced by the patient during the RCT, it has been decided to discontinue the investigational drug. The study has now been completed, and the patient will be transitioned back to standard care for her epilepsy. The patient will be closely monitored for any changes in her condition, and any necessary adjustments will be made to her treatment plan as needed.

In conclusion, this clinic visit reveals that Patient ID 124 has experienced a high frequency of seizures during the RCT evaluating ID X13737 for epilepsy. Although the patient reported dizziness and headaches, there were no significant changes or abnormalities noted on neurological examination. The investigational drug will be discontinued, and the patient will be transitioned back to standard care for her epilepsy. Close monitoring and adjustments as needed will ensure the best possible outcome for this patient.

----------------END OF PATIENT ENCOUNTER---------------

-------------- PATIENT ID 125 -- VISIT 1 -----------

Clinic Note Visit #1

Patient ID: 125

Date: 2022-09-14

Identification:

I am pleased to present the following clinic note for Patient ID 125, a 72-year-old male with an illustrious history of bipolar disorder, ADHD, and generalized epilepsy. Our esteemed patient has graced us with his presence on this auspicious day, September 14th, 2022.

Historical Perspective (HP):

Our distinguished patient has been grappling with the affliction of bipolar disorder for the past three decades, and has exhibited remarkable resilience in the face of this formidable challenge. In addition, he has been diagnosed with ADHD and generalized epilepsy, which have further compounded his burden.

Subjective:

Upon inquiring about the patient's subjective experience, he disclosed that he has been enduring an unprecedented level of seizure activity over the past two months, with a total of four seizures. These seizures have been characterized by sudden losses of consciousness and convulsions, causing significant distress and impairment in his daily life.

Medications:

Our patient has been prescribed an impressive array of medications to manage his conditions, including lithium carbonate, methylphenidate, and valproic acid. He has been adhering to these regimens with commendable dedication, as evident from the meticulous records kept by his caregivers.

Allergies:

Upon thorough probing, our patient has revealed no allergies or sensitivities to any medications or substances.

Social History:

Our esteemed patient has been blessed with a loving spouse and two grown children, who have been providing invaluable support throughout his journey. He has been working part-time as an artist, which he finds incredibly fulfilling. However, his condition has forced him to reduce his workload considerably.

Review of Systems (ROS):

During the review of systems, our patient reported experiencing no significant complaints other than those related to seizure activity. He did, however, express some mild cognitive difficulties, which he attributes to the burden of his multiple conditions.

General Physical Examination (GPE):

Upon performing a thorough general physical examination, I observed no remarkable findings other than a subtle tremor in the patient's hands. His vital signs were within normal limits, and his overall physical condition was found to be stable.

Comprehensive Neurologic Exam (CNE):

The comprehensive neurologic exam revealed no obvious abnormalities in the patient's cranial nerves, motor function, or sensory perception. However, he did exhibit a subtle degree of rigidity and bradykinesia, which are common manifestations of Parkinson's disease. His speech was found to be clear and coherent, with no evidence of dysarthria or other communication disorders.

Labs:

Recent laboratory tests have confirmed the patient's diagnosis of generalized epilepsy, with elevated levels of sodium and potassium in his serum. His renal function is within normal limits, and his liver enzymes are slightly elevated, consistent with prior episodes of seizure activity.

Imaging:

A thorough review of the patient's imaging studies has revealed no notable abnormalities, including no evidence of structural brain lesions or other anatomical alterations. His MRI scan did reveal some subtle atrophy in the temporal lobe, which is consistent with his long-standing history of seizure activity.

Assessment:

Based on the patient's comprehensive neurological examination and laboratory findings, I have reached the following assessments:

1. Bipolar disorder: The patient has been grappling with this condition for several decades, and his current symptoms are consistent with a manic episode.

2. ADHD: The patient's attention deficit hyperactivity disorder has been well-controlled by his current medication regimen.

3. Generalized epilepsy: The patient's recent seizure activity is a manifestation of this condition, and the results of his laboratory tests and imaging studies are consistent with this diagnosis.

4. Parkinson's disease: The patient's subtle tremor and rigidity are indicative of this condition, which is not uncommon in patients with long-standing epilepsy.

Plan:

In light of the above assessments, I have devised a comprehensive plan for our esteemed patient, as follows:

1. Continue the patient's existing medication regimen, including lithium carbonate, methylphenidate, and valproic acid, with careful monitoring of his serum levels and any adjustments as needed.

2. Add an investigational drug for epilepsy, which will be taken for a period of three months to assess its efficacy in reducing seizure activity.

3. Schedule regular follow-up appointments to monitor the patient's condition and adjust his medication regimen accordingly.

Signature:

I hereby certify that I have reviewed and agree with the content of this clinic note, and that all information contained herein is accurate and complete to the best of my knowledge and belief.

Date: 2022-09-14

Signature: _______________________________

Patient ID: 125

----------------END OF PATIENT ENCOUNTER---------------

-------------- PATIENT ID 125 -- VISIT 2 -----------

Visit #2, October 14th, 2022

Patient ID: 125, 72-year-old male

History: Bipolar disorder, ADHD, generalized epilepsy

Randomized controlled trial (ID X13737 for epilepsy)

Neurological Examination:

* Mild tremors in the hands and feet

* Slowed reaction time

* Slurred speech

* Difficulty with concentration and memory

Pertinent Positives:

* 11 seizures in the past 3 months while on ID X13737

* Reported fall

Assessment and Plan:

* Discontinue investigational drug ID X13737 due to study completion

* Monitor for seizure activity and adjust medication as needed

* Schedule follow-up visit in 4 weeks to assess seizure control and medication efficacy.

----------------END OF PATIENT ENCOUNTER---------------

-------------- PATIENT ID 126 -- VISIT 1 -----------

Clinic Note - Visit #1

Patient ID: 126

Date: 2022-12-31

Chief Complaint: The patient presents with a history of alcoholism and epilepsy, and has experienced 46 seizures in the past two months. He is currently enrolled in a randomized controlled trial (RCT) for the treatment of his epilepsy.

History of Present Illness: The patient reports that he has been experiencing an increase in seizure frequency over the past two months, with an average of 2-3 seizures per week. He denies any recent changes in his alcohol consumption or medication regimen. The patient's last seizure was on December 28th, and he reports that it was a complex partial seizure that lasted for approximately 15 minutes.

Past Medical History: The patient has a history of alcoholism and epilepsy, and has been receiving treatment for both conditions for several years. He has tried various medications for his epilepsy, including levetiracetam, valproate, and lamotrigine, but has not experienced significant improvement in seizure control. The patient has also reported experiencing withdrawal symptoms during periods of alcohol abstinence.

Family History: The patient's father and grandfather both had a history of seizures, and the patient's mother has a history of depression.

Social History: The patient reports that he has been drinking heavily for the past several years, with an average of 1-2 liters of alcohol per day. He denies any recent changes in his social or occupational status.

Review of Systems: The patient reports no other symptoms or complaints.

Neurological Examination:

* General: The patient appears alert and oriented, with a normal mood and affect.

* Cranial Nerves: There is no evidence of abnormality in the cranial nerves.

* Motor: The patient has a normal strength and tone, with no evidence of ataxia or tremors.

* Sensory: There is no evidence of sensory loss or abnormality.

* Coordination: The patient has a normal coordination and balance.

* Reflexes: The patient has a normal reflex response.

Assessment:

* The patient's history of alcoholism and epilepsy, combined with his recent increase in seizure frequency, suggests that he may be experiencing withdrawal symptoms or a worsening of his epilepsy.

* The patient's participation in an RCT for the treatment of his epilepsy adds a layer of complexity to his care, and it is important to ensure that any changes to his medication regimen are carefully monitored and documented.

Plan:

* Continue the patient's current medication regimen, including levetiracetam, valproate, and lamotrigine, for the treatment of his epilepsy.

* Add the investigational drug to the patient's medication regimen for a period of 3 months, as part of the RCT.

* Monitor the patient's seizure frequency and any changes in his neurological status closely during this time, and make any necessary adjustments to his medication regimen based on his response.

* Encourage the patient to continue abstaining from alcohol, and provide him with resources for alcohol support groups and counseling as needed.

* Schedule a follow-up appointment for February 28th to assess the patient's progress and make any necessary adjustments to his medication regimen.

----------------END OF PATIENT ENCOUNTER---------------

-------------- PATIENT ID 126 -- VISIT 2 -----------

Visit #2: Patient ID 126, 37 year old male

* History: Alcoholism, epilepsy (randomized controlled trial)

* Investigational drug: ID X13737 (epilepsy) - discontinued due to study completion

* Seizure frequency: 47 seizures over past 3 months

* Patient reported no symptoms

Neurological Examination:

* Mental status: Alert and oriented

* Speech: Normal

* Motor function: Normal

* Sensory system: Normal

* Reflexes: Normal

* Cranial nerves: Normal

Pertinent Positives:

* No new symptoms reported

* No changes in seizure frequency or type

Assessment and Plan:

* Discontinue investigational drug ID X13737

* Monitor for any changes in seizure frequency or type

* Schedule follow-up visit in 2 weeks to reassess patient's condition.

----------------END OF PATIENT ENCOUNTER---------------

-------------- PATIENT ID 127 -- VISIT 1 -----------

Clinic Note Visit #1

Patient ID: 127

Date: 2021-04-08

---

Identification:

Name: Sarah Johnson

Age: 64 years

Gender: Female

Contact Information:

Address: 123 Main St, Anytown, USA

---

HPI (History of Present Illness):

Sarah Johnson, a 64-year-old female, presents with a history of anxiety, depression, and occipital lobe epilepsy. She has experienced four seizures in the past two months, with the most recent one occurring three days ago. The patient reports no significant changes in her medication regimen, except for the addition of an investigational drug for epilepsy as part of a randomized controlled trial.

Subjective:

Patient reports feeling anxious and depressed, with a moderate level of stress due to her ongoing health issues. She denies any recent changes in her mood or mental status. Patient reports that she has been experiencing more frequent seizures over the past two months, which has caused significant distress and impacted her daily activities.

Medications:

1. Lamotrigine (Lamictal) - 200mg PO QD

2. Levetiracetam (Keppra) - 500mg PO BID

3. Investigational drug for epilepsy (Experimental Medication XYZ) - 500mg PO QD (added three days ago)

Allergies:

1. None reported

Social History:

Patient is a retired nurse and lives with her husband in a single-story home. She has two adult children who live nearby. Patient denies any history of substance abuse or alcohol use. She reports exercising regularly, including walking and yoga, and maintains a balanced diet.

Review of Systems:

1. General: No significant changes in overall health.

2. Neurological: Recent seizures (4 in past 2 months) with no aura or warning signs.

3. Psychiatric: Anxiety and depression, moderate level of stress.

4. Social: Married with two adult children.

5. Family: No significant medical history in first-degree relatives.

6. Allergies: None reported.

General Physical Examination:

Vital signs:

* Temperature: 98.6°F (37°C)

* Blood pressure: 120/80 mmHg

* Heart rate: 70 beats per minute

* Respiratory rate: 14 breaths per minute

Head-to-Toe Assessment:

1. Head: Normocephalic, normal occipito-frontal circumference. No focal neurological deficits or abnormalities.

2. Eyes: Pupils are symmetrical and reactive to light, with no evidence of afferent pupillary defect. Visual acuity is 20/25 in each eye.

3. Ears: No significant abnormalities detected.

4. Nose: No significant abnormalities detected.

5. Throat: No significant abnormalities detected.

6. Chest: Normal respiratory sounds, with no wheezing or added sounds.

7. Heart: Regular rhythm, normal S1 and S2 heart sounds, with no murmurs or extra sounds.

8. Abdomen: Soft and non-tender, with no significant abnormalities detected.

9. Extremities: Normal muscle strength and tone, with no significant abnormalities detected.

Comprehensive Neurologic Examination:

1. Mental Status: Alert and oriented to person, place, and time. No significant changes in her mental status.

2. Cranial Nerves: Intact, with no focal deficits or abnormalities.

3. Motor Function: Normal strength and tone, with no significant deficits or abnormalities.

4. Sensory Function: Normocephalic, with no significant deficits or abnormalities.

5. Reflexes: Normal reflexes, with no hyporeflexia or hyperreflexia.

6. Gait and Coordination: Normal gait and coordination, with no significant deficits or abnormalities.

Labs:

1. Complete Blood Count (CBC) - WNL (within normal limits)

2. Basic Metabolic Panel (BMP) - WNL

3. Thyroid-stimulating hormone (TSH) - 2.50 mIU/L (normal range: 0.40-4.00 mIU/L)

4. Lipid profile - WNL

5. Renal function tests - WNL

Imaging:

1. Computed Tomography (CT) scan of the head - WNL

2. Magnetic Resonance Imaging (MRI) of the brain - WNL

Assessment:

1. Anxiety and depression with a moderate level of stress due to ongoing health issues.

2. Recent seizures (4 in past 2 months) with no significant changes in her neurological examination.

3. No evidence of any focal neurological deficits or abnormalities.

4. Normal results from recent laboratory and imaging studies.

Plan:

1. Continue current medication regimen, including Lamotrigine (Lamictal), Levetiracetam (Keppra), and Investigational drug for epilepsy (Experimental Medication XYZ).

2. Monitor seizure activity and adjust medications as needed.

3. Reassess mental status, mood, and level of stress at subsequent visits.

4. Consider adding additional medications or therapies as needed to manage anxiety and depression.

5. Follow-up visit in two weeks to reassess patient's condition and adjust treatment plan as necessary.

Signature:

---

[Your Name]

Neurologist

----------------END OF PATIENT ENCOUNTER---------------

-------------- PATIENT ID 127 -- VISIT 2 -----------

May 8th, 2021

Patient ID: 127

Age: 64 years old

Gender: Female

Chief Complaint: Anxiety, Depression, and Occipital Lobe Epilepsy

History of Present Illness:

The patient, a 64-year-old female, has been participating in a randomized controlled trial for the investigational drug ID X13737 for her epilepsy. Over the past three months, she has taken the medication and has experienced 10 seizures. However, she has reported no symptoms. The study is now complete, and the investigational drug will be discontinued.

Neurological Examination:

On examination, the patient's vital signs are within normal limits. Her mental status is clear, and she is able to concentrate and follow instructions. Her speech is normal, and her language is intact. She has no evidence of aphasia or dysarthria. Her extraocular movements are full, and her funduscopic examination reveals no abnormalities. Her cranial nerve examination is normal, with no signs of weakness or paresis. Her motor exams show no signs of weakness, atrophy, or fasciculations. Her sensory exams are intact, with no signs of deficits in sensation to light touch, pinprick, or temperature. Her reflexes are normal, with no signs of spasticity or hyporeflexia.

Plan:

The patient will be discontinued from the investigational drug ID X13737 now that the study is complete. She will be referred to her primary care physician for ongoing management of her anxiety and depression. Her occipital lobe epilepsy will be monitored with regular follow-up appointments to assess for any changes in seizure activity.

----------------END OF PATIENT ENCOUNTER---------------

-------------- PATIENT ID 128 -- VISIT 1 -----------

Patient ID: 128

Visit #1: 2022-01-28

History: Seasonal allergies, parietal lobe epilepsy (13 seizures in past 2 months)

Randomized controlled trial

No change in medications except adding investigational drug for epilepsy for 3 months

Neurological Examination:

* Head: Normocephalic, normotectal

* Eyes: No ocular abnormalities

* Visual Fields: Full fields with no defects

* Extremities: Normal tone and strength

* Sensation: Intact

* Mental Status: Alert and oriented

Pertinent Positives:

* Seizures continue despite no changes in medications

* Enrolled in randomized controlled trial for investigational drug for epilepsy

Assessment and Plan:

* Monitor seizure activity closely during 3-month trial period

* Evaluate efficacy and safety of investigational drug

* Maintain current medication regimen

* Follow-up visit in 3 months.

----------------END OF PATIENT ENCOUNTER---------------

-------------- PATIENT ID 128 -- VISIT 2 -----------

Clinic Note - Visit #2

Date: 2022-02-28

Patient ID: 128

History of Present Illness:

The patient is a 42-year-old male with a history of seasonal allergies and parietal lobe epilepsy. He has been participating in a randomized controlled trial and has been receiving the investigational drug ID X13737 for the past three months. During this time, he has had 18 seizures with no reported symptoms. The study is now complete, and the investigational drug will be discontinued.

Neurological Examination:

General:

* Alert and oriented x3

* No signs of distress or discomfort

Head and Neck:

* Normal facial expressions and tone

* No hemiparesis or weakness noted

* No abnormalities in the oropharynx or neck

Extraocular Movements:

* Full range of motion in all axes

* No nystagmus or oscillations noted

Cranial Nerves:

* Intact sensation to light touch and pinprick over the face and scalp

* Pupils are equal, round, and reactive to light

* No abnormalities in the funduscopic examination

Motor System:

* Full range of motion in all limbs

* No hemiparesis or weakness noted

* No ataxia or tremors observed

Sensory System:

* Normal sensation to light touch and pinprick over the extremities

* No evidence of any sensory deficits or abnormalities

Assessment and Plan:

* The patient has a history of parietal lobe epilepsy and has been participating in a randomized controlled trial with the investigational drug ID X13737. During the study, he has had 18 seizures without any reported symptoms.

* The study is now complete, and the investigational drug will be discontinued.

* The patient's neurological examination reveals no significant abnormalities.

* We will monitor the patient closely for any changes in his condition and adjust the treatment plan as needed.

* The patient will continue to receive standard antiepileptic medications as prescribed by his primary care physician.

* Follow-up appointment is scheduled for [insert date].

Note: This clinic note is a fictional example and should not be used in actual patient care. It is intended only as an educational tool to demonstrate the format and content of a neurology clinic note.

----------------END OF PATIENT ENCOUNTER---------------

-------------- PATIENT ID 129 -- VISIT 1 -----------

---

Clinic Note #1 (2022-07-06)

Patient ID: 129

Age: 27 years

Gender: Female

Chief Complaint: Multifocal epilepsy with increased seizure frequency in the past 2 months.

History of Present Illness (HPI): The patient has a history of multifocal epilepsy, and has been experiencing an increase in seizure frequency over the past 2 months. She has had 24 seizures during this time period. The patient is currently participating in a randomized controlled trial for the treatment of her epilepsy.

Review of Systems (ROS):

* General: No changes in overall health or constitutional symptoms.

* Neurological: The patient reports no new neurological symptoms, but does note an increase in seizure frequency as mentioned above.

* Psychiatric: No recent changes in mood or cognitive function.

* Social: The patient denies any recent changes in her social support system or living situation.

Medications:

* Levetiracetam (Keppra) 500mg QID (previously prescribed)

* Lamotrigine (Lamictal) 200mg QHS (previously prescribed)

Allergies:

* None reported.

Social History:

* Marital status: Single.

* Occupation: Student.

* Living situation: Resides with parents.

* Recreational activities: Enjoys hiking and reading.

Review of Laboratory Findings:

* Complete Blood Count (CBC): Within normal limits.

* Basic Metabolic Panel (BMP): Within normal limits.

* Thyroid-stimulating hormone (TSH): Within normal limits.

* Vitamin D level: Low (20 ng/mL).

Review of Imaging Findings:

* Magnetic Resonance Imaging (MRI) of the brain with and without contrast: No significant abnormalities seen.

Assessment:

* The patient's increased seizure frequency is concerning, and may be related to the ongoing randomized controlled trial.

* The patient's current medications are appropriate for her condition, but the addition of an investigational drug for epilepsy may be beneficial.

* The patient's overall health and cognitive function appear stable, but further monitoring is needed to assess the efficacy and safety of the investigational drug.

Plan:

* Continue current medications (Levetiracetam and Lamotrigine) as prescribed.

* Add an investigational drug for epilepsy (E2022-13) at a dose of 500mg QID for a period of 3 months.

* Monitor the patient's seizure frequency and overall health closely during the course of the randomized controlled trial.

* Schedule follow-up appointments as needed to assess the patient's condition and adjust the treatment plan accordingly.

Signature:

---

[Your name and title]

Neurologist

----------------END OF PATIENT ENCOUNTER---------------

-------------- PATIENT ID 129 -- VISIT 2 -----------

CLINIC NOTE #2

Date: August 6, 2022

Patient ID: 129

Age: 27 years old

Gender: Female

Chief Complaint: Vertigo and multifocal epilepsy

History of Present Illness:

The patient is a 27-year-old female with a history of multifocal epilepsy. Over the past 3 months, she has been participating in a randomized controlled trial (RCT) and has been receiving the investigational drug ID X13737 for her epilepsy. During this time, she has experienced 42 seizures. The patient has also reported vertigo.

Past Medical History:

The patient has a history of multifocal epilepsy and has been experiencing seizures since the age of 10. She has tried various anticonvulsant medications, including levetiracetam, valproic acid, and lamotrigine, but has not achieved complete seizure control.

Current Medications:

The patient is currently taking levetiracetam 500 mg twice daily, valproic acid 2000 mg twice daily, and lamotrigine 200 mg once daily. She has also been receiving the investigational drug ID X13737 500 mg twice daily as part of the RCT.

Social History:

The patient is a non-smoker and does not drink alcohol. She lives with her husband and two children. She reports no significant stressors or changes in her life recently.

Review of Systems:

General: The patient denies fever, chills, or fatigue.

Neurological: The patient reports vertigo and has had 42 seizures over the past 3 months. She also experiences frequent headaches and difficulty with memory and concentration.

Psychiatric: The patient denies any psychiatric symptoms.

Musculoskeletal: The patient denies any musculoskeletal symptoms.

Other: The patient reports no other symptoms or concerns.

Physical Examination:

General: The patient appears well-nourished and well-oriented. Her speech is clear and her affect is appropriate.

Neurological: The patient has a normal gross motor examination. She has a slight limb weakness on the right side of her body, which is consistent with her history of multifocal epilepsy. Her sensory examination reveals decreased sensation to light touch and pinprick on the left side of her body, consistent with her reported vertigo. Her reflexes are normal.

Cranial Nerve Examination:

The patient has normal visual acuity and fields. Her extraocular movements are full and normal. Her oculomotor and trochlear muscles are intact. She has a slight weakness of her abducens nerve, consistent with her history of multifocal epilepsy.

Mental Status Examination:

The patient's mental status is normal. Her memory and concentration appear to be intact. She denies any hallucinations or delusions.

Assessment and Plan:

Based on the patient's history of multifocal epilepsy and her recent experience with 42 seizures over the past 3 months, it is clear that she continues to experience significant seizure activity despite being on multiple anticonvulsant medications. The investigational drug ID X13737 has not been effective in controlling her seizures, and its discontinuation is appropriate now that the RCT is complete.

The patient's vertigo and slight limb weakness on the right side of her body are consistent with her history of multifocal epilepsy and suggest ongoing seizure activity. Her decreased sensation to light touch and pinprick on the left side of her body may be related to her reported vertigo.

The patient's mental status is normal, and she denies any hallucinations or delusions. However, her history of multifocal epilepsy and recent experience with seizures suggest that she may be at risk for cognitive impairment or psychiatric symptoms.

Plan:

1. Discontinue the investigational drug ID X13737.

2. Increase the dose of levetiracetam to 1000 mg twice daily and valproic acid to 2500 mg twice daily to further optimize seizure control.

3. Add lamotrigine 500 mg once daily to the patient's regimen to provide additional seizure control.

4. Monitor the patient closely for any changes in her seizure activity or cognitive function over the next 6-12 months.

5. Consider referring the patient to a neuropsychologist or cognitive behavioral therapist to address any potential cognitive impairment or psychiatric symptoms.

----------------END OF PATIENT ENCOUNTER---------------

-------------- PATIENT ID 140 -- VISIT 1 -----------

Patient ID: 140

Visit #1: 2022-08-23

Age: 75 yrs

History: Depression, anxiety, hypertension, generalized epilepsy

Seizures: 14 in past 2 months

Randomized controlled trial (RCT)

Investigational drug for epilepsy added for 3 months

Neurological Examination:

* Alert and oriented x3

* Normal muscle strength and tone

* No sensory deficits

* Coordination and balance intact

* Reflexes are normal

Pertinent Positives:

* No changes in medications other than investigational drug for epilepsy

* Seizure activity persists (14 seizures in past 2 months)

Assessment:

* Persistent seizure activity despite current medication regimen

* Randomized controlled trial (RCT) underway to evaluate efficacy of investigational drug for epilepsy

Plan:

* Continue existing medication regimen

* Add investigational drug for epilepsy for 3 months

* Monitor seizure activity and adjust medications as needed

* Schedule follow-up visit in 3 months to reassess efficacy of treatment and make any necessary changes.

----------------END OF PATIENT ENCOUNTER---------------

-------------- PATIENT ID 140 -- VISIT 2 -----------

Clinic Note - Visit #2

Patient ID: 140

Date: 2022-09-23

Chief Complaint:

The patient is a 75-year-old male with a history of depression, anxiety, hypertension, and generalized epilepsy. He is currently participating in a randomized controlled trial for the investigational drug ID X13737 for his epilepsy.

History of Present Illness:

The patient has been taking the investigational drug ID X13737 for the past 3 months and has experienced 7 seizures during this time. He reports no other symptoms. The study is now complete and the investigational drug will be discontinued.

Review of Systems:

The patient denies any new symptoms or changes in his condition since his last visit. He continues to experience anxiety and depression, but these symptoms have not worsened.

Neurological Examination:

Mental Status: The patient is alert and oriented to person, place, and time. His speech is clear and concise. He exhibits no signs of disorganized thinking or psychosis.

Cranial Nerves:

* Ocular: Pupils are equal and reactive to light. There is no evidence of afferent pupillary defect.

* Vestibulo-ocular Reflex: Present and intact.

* Facial: There is no evidence of facial weakness or paresis.

* Mandibular: No difficulty with mandibular movement or tremor.

* Tongue: Normal tongue mobility and strength.

Motor Examination:

* Upper Limb: The patient has normal range of motion and strength in his upper limbs. There is no evidence of weakness, paresis, or atrophy.

* Lower Limb: The patient has normal range of motion and strength in his lower limbs. There is no evidence of weakness, paresis, or atrophy.

* Muscle Strength: The patient has normal muscle strength in all extremities.

Sensory Examination:

* Somatosensory: The patient has normal sensation to light touch and pinprick over all extremities. There is no evidence of hypoesthesia or hypersensitivity.

* Dysesthesias: No unusual sensations are reported.

Reflexes:

* Deep Tendon Reflexes: Present and normal.

* Superficial Tendon Reflexes: Present and normal.

Assessment and Plan:

The patient has completed the randomized controlled trial for the investigational drug ID X13737. He has experienced 7 seizures during this time, but reports no other symptoms. The study is now complete and the investigational drug will be discontinued. The patient's neurological examination is normal, with no evidence of new symptoms or changes in his condition since his last visit. His anxiety and depression continue to be managed with ongoing psychotherapy and medication.

Plan:

1. Discontinue the investigational drug ID X13737.

2. Continue current medications for anxiety and depression as prescribed.

3. Monitor the patient's seizure activity and adjust medications as needed.

4. Schedule follow-up visits as necessary to monitor the patient's condition and adjust treatment as needed.

----------------END OF PATIENT ENCOUNTER---------------

-------------- PATIENT ID 141 -- VISIT 1 -----------

On August 11, 2021, I attended a clinic visit with Patient ID 141, an 18-year-old female who has been experiencing increased seizure activity over the past two months. The patient has a history of hypothyroidism, asthma, and frontal lobe epilepsy, which we have been closely monitoring and managing with medication.

During this visit, I conducted a comprehensive neurological examination to assess the patient's current condition. Her vital signs were within normal limits, and she appeared alert and focused. On inspection, there were no abnormalities noted in her head, face, or neck. Her speech was clear and coherent, and her language skills were intact. The patient's cognitive function was also within normal limits, and she was able to perform simple calculations and recall recent events without any difficulties.

The patient reported that she had experienced eight seizures in the past two months, which is a significant increase from her previous baseline. She also mentioned that she has been experiencing some mild dizziness and blurred vision, but these symptoms are not severe enough to interfere with her daily activities. I reviewed the patient's medication regimen and confirmed that no changes will be made at this time, other than adding the investigational drug for epilepsy that will be taken for three months as part of a randomized controlled trial. The patient understands the purpose and potential risks and benefits of this trial and has provided her informed consent.

Overall, Patient ID 141 is in good health and her neurological examination revealed no significant abnormalities. However, her increased seizure activity remains a concern and we will continue to monitor her closely and adjust her medication regimen as needed to manage her condition effectively.

----------------END OF PATIENT ENCOUNTER---------------

-------------- PATIENT ID 141 -- VISIT 2 -----------

**Clinic Note, Visit #2: September 11, 2021**

Patient ID: 141

Age: 18 years

Gender: Female

Chief Complaint: Balance disorder, history of hypothyroidism, asthma, and frontal lobe epilepsy

History of Present Illness:

The patient has been participating in a randomized controlled trial for the investigational drug ID X13737 for the treatment of frontal lobe epilepsy. She has been taking the medication for the past 3 months and has experienced 29 seizures during this time. The patient has also reported a balance disorder.

Past Medical History:

* Hypothyroidism (diagnosed at age 15)

* Asthma (diagnosed at age 8)

* Frontal lobe epilepsy (diagnosed at age 12)

Medications:

* Levothyroxine (thyroid hormone replacement)

* Albuterol (asthma medication)

* Lamotrigine (anticonvulsant medication)

Social History:

* No known allergies

* Non-smoker

* Denies alcohol or illicit drug use

Review of Systems:

* Headache: None

* Dizziness/Vertigo: Yes, patient reports balance disorder

* Fatigue: None

* Muscle weakness: None

* Paresthesias: None

Physical Examination:

* Vital Signs:

+ Temperature: 98.6°F (37°C)

+ Blood Pressure: 120/80 mmHg

+ Pulse Rate: 70 beats per minute

+ Respiratory Rate: 14 breaths per minute

* General: Alert and oriented, cooperative and pleasant

* Neurological:

+ Cranial Nerves: All grossly normal

+ Motor Exam: All extremities are normocephalic and normochromic. No evidence of weakness or atrophy.

+ Sensory Exam: Patient reports decreased sensation in the right arm and leg, which is consistent with her history of frontal lobe epilepsy.

+ Coordination and Balance: Patient has difficulty maintaining balance and exhibits a wide-based gait.

* Special Senses: No abnormalities noted in vision or hearing.

Assessment and Plan:

The patient's history of frontal lobe epilepsy, hypothyroidism, and asthma, along with her recent experience of 29 seizures during the investigational drug trial, suggest that she is at high risk for seizure activity. The patient's current balance disorder is likely related to her seizure activity and may require further evaluation and management.

The patient will be advised to discontinue the investigational drug, ID X13737, as it is no longer necessary for the study. The patient's thyroid hormone replacement medication will be re-evaluated to ensure that her hypothyroidism is adequately controlled. Additionally, the patient's asthma medication will be reviewed to ensure that it is appropriate for her condition.

The patient will be scheduled for a follow-up appointment in 1 month to monitor her seizure activity and to reassess her balance disorder. In the meantime, the patient should avoid any activities that may exacerbate her balance problems and should report any changes in her symptoms or seizure activity to our office immediately.

Documentation:

* The patient has been advised of the risks and benefits of the investigational drug trial and has given informed consent for participation.

* The patient has been prescribed levothyroxine and albuterol for her pre-existing conditions.

* The patient has been advised to discontinue the investigational drug, ID X13737, as it is no longer necessary for the study.

----------------END OF PATIENT ENCOUNTER---------------

-------------- PATIENT ID 142 -- VISIT 1 -----------

Clinic Note: Visit #1 (2021-03-19)

Patient ID: 142

Age: 69 years

Chief Complaint: The patient presents with a history of bipolar disorder and epilepsy, with recent onset of increased seizure activity (9 seizures in the past 2 months).

History of Present Illness: The patient reports a worsening of seizure activity over the past 2 months, with an average of 1-2 seizures per week. The patient has been participating in a randomized controlled trial for epilepsy and has not experienced any changes in medications other than the addition of the investigational drug for the past 3 months.

Past Medical History: Bipolar disorder, epilepsy (controlled on levetiracetam), hypertension, hyperlipidemia, obesity

Medication List: Levetiracetam, lithium carbonate, atorvastatin, hydrochlorothiazide, olanzapine

Social History: The patient denies any recent travel or exposure to sick contacts. He reports a stable living situation with his spouse and adult children. The patient is a retired accountant and enjoys reading and gardening in his free time.

Review of Systems: The patient reports no significant changes in his mood, cognition, or sleep patterns. He denies any recent weight changes or musculoskeletal pain.

Physical Examination:

Vital Signs:

* Temperature: 98.6°F (37°C)

* Blood Pressure: 120/80 mmHg

* Pulse: 70 bpm

* Respiratory Rate: 14 breaths/min

Neurological Examination:

* Mental Status: The patient is alert and oriented, with no evidence of cognitive impairment.

* Speech and Language: The patient's speech is clear and normal, with no evidence of aphasia or dysarthria.

* Motor Examination: The patient demonstrates a normal range of motion in all extremities, with no evidence of weakness, ataxia, or tremors.

* Sensory Examination: The patient reports no significant changes in his sensation, including visual, auditory, or tactile sensations.

* Reflexes: The patient's reflexes are normal and symmetrical.

* Cranial Nerve Examination: The patient demonstrates a normal examination of the cranial nerves, with no evidence of abnormalities in visual, olfactory, or auditory functions.

Assessment and Plan:

* The patient's recent increase in seizure activity is concerning and may be related to the randomized controlled trial medication.

* The patient will continue to participate in the randomized controlled trial, with no changes in medications other than the addition of the investigational drug for epilepsy.

* The patient will be monitored closely for any changes in seizure activity or adverse effects from the medication.

* The patient will follow up in 2 weeks for a re-evaluation of his seizure activity and medication regimen.

Orders:

* Continue current medications (levetiracetam, lithium carbonate, atorvastatin, hydrochlorothiazide, olanzapine)

* Add investigational drug for epilepsy (as per randomized controlled trial protocol)

* Follow up in 2 weeks for re-evaluation of seizure activity and medication regimen.

----------------END OF PATIENT ENCOUNTER---------------

-------------- PATIENT ID 142 -- VISIT 2 -----------

Neurology Clinic Note - Visit #2 (2021-04-19)

Patient Information:

* Patient ID: 142

* Age: 69 years old

* Gender: Male

* History of: Bipolar disorder, epilepsy

Reason for Visit:

* The patient is in a randomized controlled trial and has been taking the investigational drug ID X13737 for epilepsy over the past 3 months.

Chief Complaint:

* Somnolence

Past Medical History:

* Bipolar disorder

* Epilepsy (history of 24 seizures during the past 3 months while taking ID X13737)

Medication List:

* ID X13737 (discontinued as of today's visit)

Neurological Examination:

* Mental Status: Alert and oriented x3

* Speech: Clear and coherent

* Motor Function: No tremors or rigidity observed. No evidence of parkinsonism or dystonia. Normal gait and station.

* Sensory Exam: Normal sensation in all extremities. No loss of pain or temperature sensation.

* Cranial Nerves: All cranial nerves are normal, with no signs of abnormality in the funduscopic exam.

* Reflexes: Are present and normal.

* Coordination and Balance: Normal.

Assessment:

* The patient's somnolence is likely related to the investigational drug ID X13737, as it has been discontinued.

* The patient has had 24 seizures over the past 3 months while taking ID X13737.

Plan:

* Discontinue ID X13737 as of today's visit.

* Monitor the patient for withdrawal symptoms from the investigational drug and provide appropriate management as needed.

* Increase the patient's current antiepileptic medication dose to control seizures.

* Schedule a follow-up appointment in 2 weeks to assess the patient's seizure control and monitor for any withdrawal symptoms.

----------------END OF PATIENT ENCOUNTER---------------

-------------- PATIENT ID 143 -- VISIT 1 -----------

---

Patient ID: 143 | Date: 2021-09-14

Identification:

* Name: Mary Johnson

* Age: 96 years

* Gender: Female

History of Present Illness (HPI):

Mary presents with a history of recurrent seizures over the past two months. She has experienced 15 seizures, each lasting approximately 2-3 minutes and occurring at irregular intervals. Mary's seizures are characterized by confusion, loss of consciousness, and postictal confusion. She reports a strong family history of epilepsy, with her mother and grandmother both diagnosed with the condition.

Subjective:

Mary reports experiencing auras before each seizure, which include a feeling of dread and a "funny" smell. She also notes that she has been experiencing more frequent episodes of forgetfulness and difficulty with word-finding. Mary's mood is generally depressed, but she denies any suicidal ideation or intent.

Medications:

* Levetiracetam (Keppra) 500mg BID

* Lamotrigine (Lamictal) 200mg OD

Allergies:

* None reported

Social History:

* Married with adult children

* Retired teacher

* Active in local community theater

Review of Systems:

* General: denies fever, chills, or weight loss

* Neurological: reports continued seizure activity despite current medications

* Musculoskeletal: denies joint pain or weakness

* Cardiovascular: denies chest pain or shortness of breath

General Physical Examination:

* Vital signs: temperature 98.6°F, blood pressure 120/80 mmHg, heart rate 70 beats per minute

* Head and neck: normal examination

* Extremities: normal examination

* Neurological: normal examination, with the exception of a mild left-sided hemiparesis

Comprehensive Neurologic Exam:

* Cranial nerves: intact

* Motor function: mild left-sided hemiparesis, 4/5 strength in the left upper and lower extremities

* Sensory function: intact

* Coordination and balance: normal

* Reflexes: normal

Labs:

* Complete Blood Count (CBC): WBC 6,000 (normal 4,500-11,000), HGB 12.5 g/dL (normal 11.5-15.5 g/dL), HCT 38% (normal 37%-52%)

* Electrolyte Panel: normal

* Thyroid Function Tests: normal

Imaging:

* CT scan of the brain with and without contrast: no acute abnormalities seen

Assessment:

* Mary Johnson is a 96-year-old female with a history of parietal lobe epilepsy and recurrent seizures over the past two months. She is currently enrolled in a randomized controlled trial investigating the use of an investigational drug for epilepsy. No change in medications will be made other than adding the investigational drug for the next three months. Mary's neurological examination reveals a mild left-sided hemiparesis, and her laboratory results are within normal limits.

Plan:

* Continue Levetiracetam (Keppra) 500mg BID and Lamotrigine (Lamictal) 200mg OD as prescribed

* Add investigational drug for epilepsy, per study protocol

* Monitor seizure activity and adjust medications as needed

* Follow up in one month to assess for any changes in medication or seizure activity

Signature:

---

[Your name and title]

Neurologist

----------------END OF PATIENT ENCOUNTER---------------

-------------- PATIENT ID 143 -- VISIT 2 -----------

Patient ID: 143

Visit #2: 2021-10-14

HPI:

* 96-year-old female

* History of parietal lobe epilepsy

* Currently enrolled in a randomized controlled trial (ID X13737) for epilepsy

* Took investigational drug for 3 months

* Experienced 22 seizures during that time

* Reported balance disorder

Neurologic Examination:

* Mental status: Alert and oriented

* Speech: Normal

* Motor: Normal

* Sensation: Normal

* Balance: Poor (reported by patient)

* Coordination: Normal

* Reflexes: Normal

Pertinent Positives:

* Seizures (22) during the past 3 months while taking investigational drug ID X13737

* Balance disorder reported by patient

Assessment and Plan:

* Discontinue investigational drug ID X13737 due to completed study

* Monitor seizure activity and balance issues closely

* Consider alternative treatments for epilepsy and balance disorders as needed.

----------------END OF PATIENT ENCOUNTER---------------

-------------- PATIENT ID 144 -- VISIT 1 -----------

Patient ID: 144

Date of Visit: 2022-02-22

Chief Complaint:

The patient, a 60-year-old female, presents with a history of hypothyroidism, autism, bipolar disorder, and occipital lobe epilepsy. She has experienced 17 seizures in the past two months, which is a significant increase from her usual frequency of 2-3 seizures per month. The patient is currently enrolled in a randomized controlled trial (RCT) investigating the efficacy and safety of an investigational drug for epilepsy.

Past Medical History:

The patient has a history of hypothyroidism, autism, bipolar disorder, and occipital lobe epilepsy. She has been managed with levothyroxine, lithium carbonate, and valproic acid. The patient has had a seizure-free period for the past 5 years, but has recently experienced an increase in seizure frequency.

Current Medications:

* Levothyroxine (T4) 25 mcg daily

* Lithium carbonate (Li) 900 mg daily

* Valproic acid (VPA) 1000 mg TID

* Investigational drug for epilepsy (ID-E) (randomized controlled trial)

Family History:

The patient's family history is positive for autism and bipolar disorder. Her mother and maternal grandmother both had a history of bipolar disorder, and her father had a history of alcoholism.

Social History:

The patient lives with her husband and adult daughter. She has a good support system and is able to manage her daily activities with some difficulty. The patient reports no significant changes in her social or occupational functioning since her last visit.

Neurological Examination:

General:

* Alert and oriented x3

* No focal neurological deficits

Cranial Nerves:

* Visual acuity: 20/20 OD, 20/40 OS (right eye)

* Fundoscopy: Normal

* Extremities: Normal motor tone and reflexes

Sensory:

* No sensory deficits

Motor:

* No significant motor weakness or atrophy

Reflexes:

* 2+ patellar reflexes

Autonomic:

* No significant autonomic dysfunction

Assessment and Plan:

Based on the patient's history of hypothyroidism, autism, bipolar disorder, and occipital lobe epilepsy, as well as her recent increase in seizure frequency, it is necessary to continue her current medications and add the investigational drug for epilepsy as part of the RCT. The patient will be monitored closely over the next 3 months to assess the efficacy and safety of the investigational drug.

During this time, the patient should continue to take her usual medications as prescribed, including levothyroxine, lithium carbonate, and valproic acid. She will also be administered the investigational drug for epilepsy twice daily. The patient will be seen for follow-up visits at 1 month, 2 months, and 3 months to assess her response to the medication and monitor for any adverse effects.

It is important for the patient to maintain a consistent sleep schedule and avoid alcohol and drugs, as these can trigger seizures. The patient should also be aware of her surroundings at all times and seek medical attention immediately if she experiences any new or worsening symptoms.

The patient will be contacted by the study team to discuss the results of the RCT and any necessary adjustments to her medication regimen.

Additionally, the patient should be aware of the potential risks and benefits associated with the investigational drug, as well as the possibility of withdrawal from the study if necessary. The patient will be monitored closely over the next 3 months to ensure her safety and efficacy of the investigational drug.

----------------END OF PATIENT ENCOUNTER---------------

-------------- PATIENT ID 144 -- VISIT 2 -----------

Visit #2 - 2022-03-22

Patient ID 144, a 60-year-old female with a history of hypothyroidism, autism, bipolar disorder, and occipital lobe epilepsy, was seen in the clinic for her second visit. The patient has been participating in a randomized controlled trial (RCT) for the investigational drug ID X13737, which she had been taking for the past three months. During this time, the patient reported experiencing 31 seizures. Despite this, the patient reported no symptoms.

On examination, the patient's neurological status was assessed, and there were no significant changes noted. The patient's cognitive function remained intact, and her mood was euthymic. There was no evidence of dyskinesias or other adverse effects. The patient's vital signs were within normal limits, including her temperature, blood pressure, and heart rate.

The investigational drug ID X13737 will be discontinued now that the study is complete. The patient will be closely monitored for any potential withdrawal symptoms or recurrence of seizures. The patient's medication regimen will be adjusted as needed to manage her occipital lobe epilepsy and other comorbid conditions.

In conclusion, Patient ID 144 has completed the RCT for ID X13737 and has experienced a significant number of seizures during the study period. However, the patient reported no symptoms and remained stable neurologically. The investigational drug will be discontinued, and the patient's medication regimen will be adjusted as needed to ensure her ongoing care and management.

----------------END OF PATIENT ENCOUNTER---------------

-------------- PATIENT ID 145 -- VISIT 1 -----------

Visit #1: 2021-03-11

Patient ID: 145

Age: 82 yrs

History: Multifocal epilepsy (12 seizures in past 2 months)

Randomized controlled trial

No change in medications except for investigational drug for epilepsy (3 months)

Neurological Examination:

* General: Alert and oriented

* Mental status: Normal

* Speech: Clear and coherent

* Motor: No weakness or tremors noted

* Sensation: Intact

* Reflexes: 2+ (normal)

* Coordination and balance: Normal

Pertinent Positives:

* Multifocal epilepsy

* Increased seizure frequency in past 2 months

Assessment and Plan:

* Continue randomized controlled trial with no changes to medications except for investigational drug for epilepsy (3 months)

* Monitor seizure frequency and severity during trial

* Follow-up visit in 2 weeks to assess tolerance and efficacy of investigational drug.

----------------END OF PATIENT ENCOUNTER---------------

-------------- PATIENT ID 145 -- VISIT 2 -----------

Clinic Note - Visit #2

Date: 2021-04-11

Patient ID: 145

Age: 82 years old

This is a follow-up visit for Patient ID 145, an 82-year-old male patient with a history of multifocal epilepsy. The patient has been participating in a randomized controlled trial (RCT) and has been taking the investigational drug ID X13737 for epilepsy over the past three months.

During the past three months, the patient has experienced 10 seizures. However, the patient reported no symptoms or adverse effects related to the investigational drug. The patient's neurological examination revealed no significant changes in their condition since the previous visit.

The patient was instructed to discontinue taking ID X13737 as the study is now complete. The patient will be followed up with regular appointments to monitor their condition and manage any ongoing seizures. The patient's response to the investigational drug and any long-term effects will be closely monitored and documented in future clinic notes.

Neurological Examination:

* Mental status: Alert and oriented x3

* Memory: Normal

* Speech: Clear and coherent

* Motor function: No notable abnormalities or weakness

* Sensation: Intact

* Reflexes: Normocal

* Coordination and balance: Normal

In conclusion, Patient ID 145 has completed the RCT and has been taking the investigational drug ID X13737 for epilepsy over the past three months. Although the patient experienced 10 seizures during this time, there were no significant symptoms or adverse effects related to the drug. The patient will be discontinued from taking ID X13737 and will be followed up with regular appointments to monitor their condition and manage any ongoing seizures.

----------------END OF PATIENT ENCOUNTER---------------

-------------- PATIENT ID 146 -- VISIT 1 -----------

**Patient Information:**

Name: Patient ID 146

Age: 61 years old

Gender: Female

Chief Complaint: Temporal lobe epilepsy with increased seizure frequency in the past 2 months.

**Past Medical History:**

* Depression

* Temporal lobe epilepsy (TLE)

* Previous history of stroke

* History of alcohol abuse

**Current Medications:**

* Levetiracetam 500mg twice daily

* Lamotrigine 200mg once daily

* Valproate sodium 500mg twice daily

* Investigational drug for epilepsy (addicted to the trial for 3 months)

**Present Illness:**

The patient presents with a history of increased seizure frequency in the past 2 months. She has experienced 27 seizures during this time, with no notable triggers or warning signs. The patient reports no changes in her medications since her last visit.

**Neurological Examination:**

* Mental Status: The patient is alert and oriented to person, place, and time. She displays no evidence of cognitive impairment or altered mental status.

* Cranial Nerve Examination: All cranial nerves are intact, with no signs of abnormality.

* Motor Examination: The patient has normal strength and tone in all muscle groups. There is no evidence of ataxia, tremors, or other motor abnormalities.

* Sensory Examination: The patient displays normal sensation to light touch, pain, and temperature on all areas examined. There is no evidence of sensory loss or altered sensation.

* Coordination and Balance Examination: The patient demonstrates normal coordination and balance. She is able to walk normally and does not display any signs of ataxia or unsteadiness.

**Assessment and Plan:**

Based on the patient's history of temporal lobe epilepsy and increased seizure frequency, I assess that the patient is appropriate for continuation in the randomized controlled trial. No changes will be made to her current medications at this time, with the exception of adding the investigational drug for epilepsy for a 3-month trial period.

The patient will be monitored closely throughout the trial period, with regular follow-up appointments scheduled every 2 weeks to assess seizure frequency and medication tolerability. Additionally, the patient will be advised to maintain a detailed seizure log to track any changes in seizure activity.

If the patient experiences any significant side effects or changes in seizure frequency during the trial period, appropriate adjustments to her medication regimen will be made as necessary. The patient will also be referred for regular EEG monitoring to assess for any changes in seizure activity.

I have discussed the risks and benefits of continuing in the randomized controlled trial with the patient, and she has been advised to contact me immediately if she experiences any significant side effects or changes in her condition.

**Planned Follow-up:**

The patient will be followed up every 2 weeks for a period of 3 months to assess seizure frequency, medication tolerability, and any other changes in her condition. Additionally, the patient will be referred for regular EEG monitoring throughout the trial period.

----------------END OF PATIENT ENCOUNTER---------------

-------------- PATIENT ID 146 -- VISIT 2 -----------

Visit #2 - 2022-06-25

Patient ID: 146

Age: 61 years

Gender: Female

History:

* Depression

* Temporal lobe epilepsy

* Randomized controlled trial (ID X13737 for epilepsy)

Current Illness:

* Upper respiratory infection

Pertinent Positives:

* 26 seizures in the past 3 months while taking investigational drug ID X13737

Neurological Examination:

* Alert and oriented x3

* No focal deficits or abnormalities detected

Assessment and Plan:

* Discontinue investigational drug ID X13737 due to completion of study

* Monitor for seizure activity and treat as needed

* Manage upper respiratory infection with antiviral medication as appropriate

* Follow up in 2-4 weeks for continued monitoring

----------------END OF PATIENT ENCOUNTER---------------

-------------- PATIENT ID 147 -- VISIT 1 -----------

Visit #1 - 2022-04-01

Patient ID: 147

Age: 84 years old

History:

* ADHD

* Anxiety

* Bipolar disorder

* Generalized epilepsy

* 9 seizures in past 2 months

* Randomized controlled trial (RCT)

Neurological Examination:

* Mental status: Alert and oriented

* Speech: Normal

* Motor: Normal

* Sensation: Normal

* Reflexes: Normal

* Gait: Normal

Pertinent Positives:

* No change in medications other than investigational drug for epilepsy

* Investigational drug to be taken for 3 months

Assessment and Plan:

* Monitor seizure activity during RCT

* Continue existing medications for ADHD, anxiety, and bipolar disorder

* Add investigational drug for epilepsy and monitor for efficacy and adverse effects

* Schedule follow-up visit in 3 months to reassess seizure activity and medication regimen.

----------------END OF PATIENT ENCOUNTER---------------

-------------- PATIENT ID 147 -- VISIT 2 -----------

Visit #2, May 1, 2022

Patient ID: 147

Age: 84 years old

Female

History: ADHD, anxiety, bipolar disorder, generalized epilepsy

Randomized controlled trial (ID X13737 for epilepsy)

Current Medications: None

Previous Medications: ID X13737 (discontinued)

Neurological Examination:

* Mental status: Alert and oriented

* Cranial nerves: Intact

* Motor exam: Normal tone, no ataxia or tremors

* Sensory exam: Intact

* Reflexes: 2+ (moderate)

* Gait: Normal

Pertinent Positives:

* No seizure activity reported since last visit

* 17 seizures experienced during the past 3 months while on ID X13737

Assessment and Plan:

* Discontinue ID X13737 per study protocol

* Monitor for seizure recurrence and adjust medication as needed

* Schedule follow-up visit in 2 weeks to assess for any changes in seizure activity.

----------------END OF PATIENT ENCOUNTER---------------

-------------- PATIENT ID 148 -- VISIT 1 -----------

Visit #1 - 2022-11-30

Patient ID: 148

Age: 24 years

History:

* Anxiety

* Depression

* Asthma

* Epilepsy (103 seizures in past 2 months)

* Randomized controlled trial (no change in medications except for investigational drug for epilepsy)

Neurological Examination:

* Head: normocephalic, normal facial expressions

* Eyes: PERRLA, no nystagmus or diplopia

* Visual Fields: full fields, no defects

* Extremities: no weakness or atrophy

* Sensation: intact

* Reflexes: 2+ (normal)

Pertinent Positives:

* Seizures: 103 in past 2 months

* No change in medications except for investigational drug for epilepsy

Assessment and Plan:

* Monitor seizure activity and adjust medications as needed

* Continue randomized controlled trial with no changes to medications except for investigational drug for epilepsy

* Follow up in 1 month for re-evaluation of seizure activity and medication adjustments as needed.

----------------END OF PATIENT ENCOUNTER---------------

-------------- PATIENT ID 148 -- VISIT 2 -----------

Patient ID: 148

Visit #2: 2022-12-30

History:

* Anxiety

* Depression

* Asthma

* Epilepsy (randomized controlled trial)

Investigational Drug:

* ID X13737 (epilepsy) - discontinued due to study completion

Neurological Examination:

* Mental status: alert and oriented

* Speech: clear and coherent

* Motor exam: normal strength and tone, no ataxia or tremors

* Sensory exam: intact

* Cranial nerve exam: normal

* Reflexes: normal

* Dizziness: reported

* Fall: reported

Pertinent Positives:

* Seizures (61) over past 3 months while taking investigational drug

* Dizziness and fall reported

Assessment:

* Discontinue investigational drug due to study completion

* Monitor for seizure activity and address any concerns with patient

Plan:

* Schedule follow-up appointment as needed

* Monitor patient's seizure activity and address any concerns

----------------END OF PATIENT ENCOUNTER---------------

-------------- PATIENT ID 149 -- VISIT 1 -----------

Clinic Note #1

Date: 2022-07-05

Patient ID: 149

Name: [Patient's Name]

Age: 42 years old

Gender: Female

Chief Complaint (CC): Multifocal epilepsy with increased seizure activity in the past two months.

History of Present Illness (HPI): The patient presents with a history of seasonal allergies and diabetes, which have been well-controlled for several years. However, she has experienced a significant increase in seizure activity over the past two months, with 86 seizures reported during this time frame. The patient is currently enrolled in a randomized controlled trial (RCT) investigating the efficacy of an investigational drug for the treatment of epilepsy. No changes will be made to her current medications other than the addition of the investigational drug, which she will take for three months.

Review of Systems (ROS): The patient reports no significant changes in her medical history, including no recent illnesses or injuries. She denies any recent travel or exposure to potential triggers for her seizures. She has been adherent to her medication regimen and has not missed any doses.

Medications:

* Levetiracetam (Keppra) 500mg PO BID

* Lamotrigine (Lamictal) 200mg PO QHS

* Investigational drug for epilepsy (RCT): Placebo (for 3 months)

Allergies:

* Seasonal allergies (pollen, dust mites)

Social History:

* Married with two children (ages 9 and 11)

* Employed as a teacher at a local elementary school

* Denies any history of substance abuse or alcohol use

Subjective: The patient reports an increase in seizure activity over the past two months, with an average of 4-5 seizures per week. She describes the seizures as being more frequent and longer in duration compared to previous years. She also reports increased fatigue and difficulty concentrating due to the seizures.

Objective:

* General Physical Examination (GPE): The patient appears well-nourished and well-developed, with no notable physical findings.

* Comprehensive Neurologic Exam (CNE): The patient exhibits a normal range of motion in all extremities, with no evidence of weakness or atrophy. Her sensory exam is essentially normal, with no deficits noted. She does not exhibit any abnormal movements or tremors.

* Laboratory Findings:

+ Complete Blood Count (CBC): Within normal limits (WNL)

+ Electrolyte Panel: WNL

+ Thyroid-stimulating Hormone (TSH): WNL

+ Lipid Profile: WNL

+ Urinalysis: WNL

* Imaging Findings:

+ Magnetic Resonance Imaging (MRI) of the brain with and without contrast: No evidence of structural abnormalities or lesions.

+ Computed Tomography (CT) scan of the head: No evidence of intracranial abnormalities.

Assessment:

* The patient's increased seizure activity over the past two months is concerning and may be related to the ongoing RCT investigational drug for epilepsy.

* The patient's history of seasonal allergies and diabetes may also be contributing factors to her seizure activity.

* The patient's current medication regimen, including levetiracetam and lamotrigine, appears to be effective in controlling her seizures, but the addition of the investigational drug for three months may help to further reduce seizure frequency and improve her quality of life.

* The patient's social history and employment as a teacher are positive factors that may contribute to her overall well-being and stress management.

Plan:

* Continue the patient's current medication regimen, including levetiracetam and lamotrigine.

* Add the investigational drug for epilepsy (placebo) for three months as part of the ongoing RCT.

* Monitor the patient's seizure activity closely during the three-month trial period.

* Schedule follow-up appointments every two weeks to assess the patient's response to the investigational drug and make any necessary adjustments to her medication regimen.

Signature:

[Your Signature]

Date: 2022-07-05

Time: [Time of Visit]

----------------END OF PATIENT ENCOUNTER---------------

-------------- PATIENT ID 149 -- VISIT 2 -----------

Sure, here's the clinic note for Patient ID 149, a 42-year-old female with a history of diabetes, seasonal allergies, and multifocal epilepsy:

Identification:

Patient ID: 149

Date of Birth: March 15, 1980

Gender: Female

HPI (History of Present Illness):

The patient presents with a history of diabetes, seasonal allergies, and multifocal epilepsy. She has been participating in a randomized controlled trial for the investigational drug ID X13737 for epilepsy over the past 3 months. During this time, she has experienced 108 seizures. The patient also reports balance disorder.

Subjective:

The patient reports feeling generally unwell and experiencing persistent seizures since starting the investigational drug. She also complains of balance difficulties, which have been ongoing for the past month. The patient describes her seizures as "like a thousand needles stabbing my brain at once." She has noticed that her seizures tend to occur more frequently during periods of high stress and when she is not adherent to her medication regimen.

Medications:

* ID X13737 (Investigational drug for epilepsy): Discontinued due to completion of the randomized controlled trial

* Levetiracetam (Keppra) 500mg BID

* Lamotrigine (Lamictal) 200mg OD

* Metformin (Glucophage) 1000mg BID

Allergies:

None reported

Social history:

The patient is a non-smoker and does not drink alcohol. She lives with her husband and two children. The patient reports that she has been experiencing increased stress due to the recent loss of her job. She also reports that she has been having difficulty sleeping at night due to her seizures.

Review of Systems:

Neurological: The patient reports persistent seizures, balance difficulties, and "brain fog"

Cardiovascular: The patient denies any cardiovascular symptoms

Respiratory: The patient reports mild shortness of breath

Gastrointestinal: The patient reports nausea and vomiting

General Physical Examination:

The patient appears thin and pale, with a height of 5'8" and a weight of 130 lbs. Her blood pressure is 120/80 mmHg, and her heart rate is 88 beats per minute. The patient's head is normocephalic and ataxic. Her speech is clear and coherent. Her extraocular movements are intact. Her hearing and vision are within normal limits. The patient's reflexes are decreased bilaterally. Her sensory examination reveals decreased sensation to light touch and pinprick over the left hemi-body.

Comprehensive Neurologic Exam:

The patient presents with a history of multifocal epilepsy, which is consistent with her neurological examination findings. She exhibits decreased sensation to light touch and pinprick over the left hemi-body, indicating a possible seizure focus in this area. Her speech is clear and coherent, but she reports difficulty with balance and "brain fog." The patient's reflexes are decreased bilaterally, which may indicate peripheral neuropathy or a seizure disorder.

Labs:

* Glucose (fasting): 120 mg/dL

* Hemoglobin A1c (HbA1c): 7.5%

* Thyroid-stimulating hormone (TSH): 4.2 μIU/mL

* Lipid profile: Within normal limits

* Complete blood count (CBC): Within normal limits

Imaging:

* MRI brain: No evidence of structural abnormalities or lesions

Assessment:

The patient's persistent seizures, balance difficulties, and "brain fog" are concerning. The recent loss of her job and difficulty sleeping may be contributing to her increased stress levels, which may be exacerbating her seizure disorder. The patient's decreased sensation to light touch and pinprick over the left hemi-body may indicate a possible seizure focus in this area. Her history of multifocal epilepsy and decreased reflexes suggest a possible peripheral neuropathy or seizure disorder.

Plan:

1. The patient will be referred to an epileptologist for further evaluation and management of her seizure disorder.

2. The patient's investigational drug, ID X13737, will be discontinued due to completion of the randomized controlled trial.

3. The patient will be prescribed a new medication, levetiracetam (Keppra), to be titrated up to 1000mg BID to help control her seizures.

4. The patient will follow up with neurology clinic in two weeks for reassessment of her seizure control and to discuss possible future medication options.

5. The patient will be referred to physical therapy to address her balance difficulties.

Signature:

[Your name and title]

Neurologist

Date: August 5, 2022

----------------END OF PATIENT ENCOUNTER---------------

-------------- PATIENT ID 160 -- VISIT 1 -----------

Patient ID: 160, Male, 37 years old

History: Arthritis, seasonal allergies, temporal lobe epilepsy (TLE)

Currently on: Levetiracetam 500mg twice daily, Lamotrigine 200mg once daily

Recent seizure history: 14 seizures in past 2 months

Randomized controlled trial (RCT): Investigational drug for epilepsy added for 3 months

Neurological examination:

* Mild limb rigidity

* Hyperreflexia

* Mild tremors

* No significant sensory or motor deficits

Pertinent positives:

* No changes in seizure frequency or type since last visit

* Patient reports no side effects from current medications

Assessment and plan:

* Continue Levetiracetam and Lamotrigine as prescribed

* Add investigational drug for epilepsy for 3 months

* Monitor seizure frequency and adjust medication regimen as needed

* Schedule follow-up visit in 4 weeks to assess efficacy and tolerability of new medication.

----------------END OF PATIENT ENCOUNTER---------------

-------------- PATIENT ID 160 -- VISIT 2 -----------

On March 24, 2023, I conducted a follow-up visit with Patient ID 160, a 37-year-old male who has been participating in a randomized controlled trial for the investigational drug ID X13737 for his temporal lobe epilepsy. Over the past three months, the patient has taken the medication and has experienced 14 seizures during this time period. Additionally, the patient reported experiencing somnolence while taking the drug.

Upon conducting a complete neurological examination, I noted that the patient's cognitive function and memory were intact, and there were no noticeable changes in his motor function or coordination. However, I did observe some mild tremors in the patient's hands and feet, which is consistent with the patient's history of arthritis. The patient's sensory functions, including vision and hearing, were also within normal limits.

Given the patient's experience of somnolence and the 14 seizures that occurred during the study period, it has been decided that the investigational drug ID X13737 will be discontinued. While the drug showed promise in controlling the patient's epilepsy, the risks associated with its use have outweighed the benefits. The patient will be closely monitored and managed with alternative medications to ensure his continued well-being.

In conclusion, this follow-up visit with Patient ID 160 has revealed some valuable insights into the effects of the investigational drug ID X13737 on his temporal lobe epilepsy. While the drug showed some promise, it is clear that its use is not without risk and alternative medications will be used to manage the patient's condition going forward.

----------------END OF PATIENT ENCOUNTER---------------

-------------- PATIENT ID 161 -- VISIT 1 -----------

**Patient Information:**

Name: Patient ID 161

Age: 45 years old

Gender: Male

**Chief Complaint:**

The patient presents with a history of diabetes, autism, and generalized epilepsy. He has experienced 42 seizures in the past 2 months.

**History of Present Illness:**

The patient has been participating in a randomized controlled trial for the treatment of his epilepsy. He has been taking the investigational drug for the past 3 months and has experienced an increase in seizure activity.

**Review of Systems:**

The patient denies any recent changes in his medications or substance use. He reports no recent travel or exposure to illness.

**Physical Examination:**

General: The patient appears well-nourished and well-developed, with no signs of distress or discomfort.

Head and Eyes: The patient's head is normocephalic and symmetrical. His eyes are appropriately spaced and aligned, with no evidence of strabismus or other abnormalities.

Ears, Nose, and Throat: The patient's ears, nose, and throat are normal in appearance and function.

Neck, Back, and Extremities: The patient's neck, back, and extremities are normal in alignment and movement, with no signs of weakness or instability.

Skin and Appendages: The patient's skin is smooth and unremarkable, with no rashes, lesions, or other abnormalities. His nails and hair are normal in appearance.

**Neurological Examination:**

Cranial Nerves: The patient's cranial nerves are intact, with no signs of dysfunction or weakness.

Motor Examination: The patient's motor function is normal, with no signs of weakness, atrophy, or other abnormalities.

Sensory Examination: The patient's sensory function is intact, with no signs of dysfunction or loss.

Coordination and Balance: The patient's coordination and balance are normal, with no signs of instability or difficulty with movement.

**Assessment and Plan:**

Based on the patient's history and physical examination, it is clear that he is experiencing an increase in seizure activity despite being in a randomized controlled trial for the treatment of his epilepsy. The patient will continue to participate in the trial, with the addition of the investigational drug for the next 3 months. No changes will be made to the patient's current medications.

The patient will be closely monitored during this time, with regular follow-up appointments scheduled to assess his seizure activity and response to treatment. Additionally, the patient will be educated on proper seizure management techniques and strategies to reduce the risk of injury or complications related to his seizures.

Date: 2022-05-21

Patient ID: 161

----------------END OF PATIENT ENCOUNTER---------------

-------------- PATIENT ID 161 -- VISIT 2 -----------

Clinic Note - Visit #2

Patient ID: 161

Date: 2022-06-21

Chief Complaint: Gait disturbance

History of Present Illness:

The patient, a 45-year-old male with a history of diabetes, autism, and generalized epilepsy, has been participating in a randomized controlled trial for the investigational drug ID X13737 for epilepsy. The patient has been taking the drug for the past three months and has experienced 76 seizures during this time. The patient has reported gait disturbance as a new symptom.

Past Medical History:

The patient has a history of diabetes, autism, and generalized epilepsy. He has been previously treated with levetiracetam and lamotrigine for his seizures. The patient has also experienced several episodes of status epilepticus.

Medications:

The patient is currently taking levetiracetam and lamotrigine for his seizures. He has been prescribed ID X13737 as part of the randomized controlled trial, which will be discontinued now that the study is complete.

Neurological Examination:

General: The patient appears alert and awake, but exhibits a slight tremor in his hands. He has a normal facial expression and does not demonstrate any abnormalities in his speech or language.

Cranial Nerves:

* Oculomotor: The patient's eyes are deviated slightly towards the left, with a mild restriction of adduction.

* Olfactory: The patient does not have any olfactory dysfunction.

* Optic: The patient's visual acuity is normal, but he does exhibit a slight afferent pupillary defect on the right.

* Vestibulo-ocular reflex: The patient's vestibulo-ocular reflex is intact.

Motor:

* Upper extremities: The patient has a mild tremor in his right hand, with a slightly weaker grip strength compared to the left side. There is no evidence of atrophy or weakness.

* Lower extremities: The patient exhibits a wide-based, unsteady gait with a slight limp. He has difficulty maintaining balance and requires assistance with walking.

Sensory:

* Somatosensory: The patient's sensation is intact, with no evidence of numbness or tingling.

* Dysesthesia: The patient does not have any dysesthesia or pain.

Reflexes:

* Extensor: The patient's reflexes are normal.

* Flexor: The patient's reflexes are slightly brisk on the right side.

Assessment and Plan:

Based on the patient's history of diabetes, autism, and generalized epilepsy, as well as his recent experience with ID X13737, it is likely that the patient's gait disturbance is related to his seizure activity. The patient's mild tremor and weaker grip strength on the right side may be indicative of a peripheral neuropathy or nerve damage.

The patient will be referred to physical therapy for further evaluation and treatment of his gait disturbance and balance issues. His medications, including levetiracetam and lamotrigine, will be reviewed and adjusted as necessary. The patient's seizure activity will be closely monitored, and alternative treatments will be considered if the patient's seizures persist or worsen.

The patient will be discontinued from the ID X13737 randomized controlled trial, as it is no longer necessary for his treatment. The patient will be provided with information regarding the results of the study and any further treatment options that may be available to him.

----------------END OF PATIENT ENCOUNTER---------------

-------------- PATIENT ID 162 -- VISIT 1 -----------

Clinic Note - Visit #1 (2022-02-11)

Patient ID: 162

Age: 33 years

Gender: Female

Chief Complaint: The patient presents with a history of depression, dermatitis, schizophrenia, and multifocal epilepsy. She has experienced 24 seizures in the past two months.

Past Medical History:

* Schizophrenia (diagnosed at age 25)

* Dermatitis (ongoing)

* Depression (ongoing)

* Multifocal epilepsy (diagnosed at age 20)

Medications:

* Risperidone (for schizophrenia)

* Lamotrigine (for epilepsy)

* Amitriptyline (for depression)

Social History:

* Married with two children (ages 8 and 10)

* Works as a teacher

Family History:

* No history of neurological disorders

Review of Systems:

* No recent changes in mood, cognition, or sleep

* No new skin lesions or exacerbation of existing ones

* No changes in appetite or weight

* No recent travel or illnesses

Neurologic Examination:

* General appearance: alert and cooperative

* Head: normocephalic, atrophic scar on the left parietal lobe

* Eyes: normal visual acuity, no evidence of gaze palsy or oculomotor abnormalities

* Cranial nerves: intact (I-XII)

* Motor examination: normal tone and strength, no ataxia or dyskinesias

* Sensory examination: decreased sensation to light touch and pinprick over the left hemiparesis, intact sensation on the right side

* Coordination and balance: normal

* Reflexes: 2+ (moderate) bilaterally

Assessment:

* The patient's history of schizophrenia, dermatitis, and multifocal epilepsy, along with her recent increase in seizure activity, suggests a possible exacerbation of her underlying neurological disorders.

* The presence of an atrophic scar on the left parietal lobe and decreased sensation on the left hemiparesis are consistent with previous history of seizures and stroke-like episodes.

* No significant changes in the patient's mood, cognition, or sleep were noted during this visit.

Plan:

1. Continue current medications (risperidone, lamotrigine, and amitriptyline) with no changes at this time.

2. Add the investigational drug for epilepsy (name to be provided by study staff) for a period of 3 months as part of the ongoing randomized controlled trial.

3. Monitor seizure activity and adjust medications as needed.

4. Schedule follow-up visit in 2 weeks to assess for any changes in symptoms or seizure activity.

5. Encourage patient to continue with her current treatment plan and to report any changes in her symptoms or seizure activity promptly.

----------------END OF PATIENT ENCOUNTER---------------

-------------- PATIENT ID 162 -- VISIT 2 -----------

Clinic Note - Visit #2 (2022-03-11)

Patient ID: 162, Female, 33 years old

History of Present Illness:

The patient is a 33-year-old female with a history of depression, dermatitis, schizophrenia, and multifocal epilepsy. She has been participating in a randomized controlled trial (RCT) for the investigational drug ID X13737, which she has been taking for the past three months. During this time, she has experienced 82 seizures and reported somnolence. The RCT is now complete, and the investigational drug will be discontinued.

Neurological Examination:

The patient's neurological examination revealed no significant changes since her last visit. She continues to experience multifocal seizures, with a average of 82 seizures per month. Her somnolence has improved slightly, but she still reports feeling drowsy and lethargic throughout the day. Her mood remains depressed, and she continues to experience dermatitis on her arms and legs.

Assessment and Plan:

Based on the patient's history of multifocal epilepsy and her recent experience with ID X13737, it is clear that she requires close monitoring and management of her seizures. The investigational drug will be discontinued now that the RCT is complete, and we will work towards finding an alternative treatment for her seizures. Her depression and dermatitis will also need to be closely managed. We will continue to monitor her condition closely and adjust her treatment plan as needed.

Recommendations:

1. Discontinue ID X13737 and transition to an alternative medication for seizure management.

2. Continue to monitor the patient's seizures and make any necessary adjustments to her treatment plan.

3. Address her ongoing depression and dermatitis with appropriate therapies and medications.

4. Schedule follow-up visits as needed to monitor the patient's condition and adjust her treatment plan accordingly.

----------------END OF PATIENT ENCOUNTER---------------

-------------- PATIENT ID 163 -- VISIT 1 -----------

Patient ID 163, a 33-year-old male with a history of autism, ADHD, and parietal lobe epilepsy, was seen in clinic on 2022-07-15 for a routine follow-up visit. The patient has had 32 seizures in the past two months, which is an increase from his previous seizure activity. The patient is currently participating in a randomized controlled trial (RCT) to assess the efficacy of an investigational drug for epilepsy.

On examination, the patient's neurological status was as follows: His height and weight were within normal limits. He had a normal head and neck examination, with no focal deficits or abnormalities noted. His cranial nerves were intact, and his motor function was normal, with no signs of weakness or paresis. The patient's sensory examination revealed no abnormalities in his sensation to light touch, pain, or temperature. His reflexes were also normal, with no hyperexplexia or hyperreflexia noted.

The patient's seizure activity has been persistent and frequent over the past two months, with an average of 2-3 seizures per week. The patient has experienced both partial onset seizures and secondary generalization seizures, with no significant change in the frequency or severity of these seizures since his last visit. The patient's mood and cognitive function remain stable, with no changes in his demeanor or behavior noted during the examination.

The patient will continue to participate in the RCT for the next three months, during which time he will receive the investigational drug as well as his standard anticonvulsant medications. No changes in his medication regimen will be made at this time, other than the addition of the investigational drug. The patient will be closely monitored for any changes in his seizure activity or neurological status, and will be seen again in clinic after three months to assess his response to the trial medication.

----------------END OF PATIENT ENCOUNTER---------------

-------------- PATIENT ID 163 -- VISIT 2 -----------

On August 15, 2022, I had the pleasure of conducting a follow-up visit with Patient ID 163, a 33-year-old male who has been participating in a randomized controlled trial for the investigational drug ID X13737 for epilepsy. Over the past three months, the patient has been taking this medication and has experienced 81 seizures during this time. Despite this, the patient reported no symptoms or adverse effects from the medication.

Upon conducting a complete neurological examination, I observed that the patient's cognitive function and speech were unremarkable. His motor functions were also intact, with no evidence of weakness or ataxia. However, I did notice some mild tremors in his hands, which could be related to the medication or his underlying condition. The patient's vision and hearing were also normal.

As the study is now complete, it has been decided that the investigational drug ID X13737 will be discontinued. I discussed this decision with the patient and he understood the reason for discontinuing the medication. I also provided him with information on other treatment options available for his condition and addressed any questions or concerns he may have had.

In conclusion, Patient ID 163 has tolerated the investigational drug ID X13737 well over the past three months, despite experiencing a high frequency of seizures during this time. His neurological examination revealed no significant adverse effects from the medication, and he reported no symptoms. As the study is now complete, it is important that we continue to monitor his condition closely and provide appropriate follow-up care to ensure his ongoing health and well-being.

----------------END OF PATIENT ENCOUNTER---------------

-------------- PATIENT ID 164 -- VISIT 1 -----------

Neurologist Clinic Note - Visit #1

Date: 2021-12-13

Patient ID: 164

Age: 83 years old

Gender: Female

History of Present Illness: The patient is an 83-year-old female with a history of parietal lobe epilepsy. She has experienced three seizures in the past two months, and is currently enrolled in a randomized controlled trial (RCT) for the treatment of her condition. No changes will be made to her current medications except for the addition of the investigational drug for epilepsy, which she will take for the next three months.

Past Medical History: The patient has a history of parietal lobe epilepsy and has been experiencing recurrent seizures over the past few years. She has tried several different medications and has undergone various diagnostic tests, including MRI and EEG studies, to determine the cause of her seizures.

Current Medications: The patient is currently taking levetiracetam 2500 mg/day, lamotrigine 200 mg/day, and valproate 1000 mg/day. She has also been prescribed the investigational drug for epilepsy, which she will take for the next three months as part of the RCT.

Social History: The patient lives with her husband and adult daughter. She enjoys spending time with her family and participating in community activities, such as gardening and volunteering at her local church. She denies any recent changes in her social support network or living situation that may have contributed to her seizures.

Review of Systems: The patient reports no other symptoms or concerns, such as fever, headache, or fatigue, that may be related to her seizures or medications. She denies any recent travel or exposure to potential triggers for her epilepsy.

Neurological Examination:

General: The patient appears alert and well-oriented. Her speech is clear and deliberate, and her affect is appropriate.

Cranial Nerves: All cranial nerves are intact, with no evidence of weakness or paresis.

Motor: The patient's motor function is normal, with no signs of weakness, atrophy, or tremors.

Sensory: Her sensation is intact, with no signs of numbness, tingling, or loss of sensation.

Coordination and Balance: The patient's coordination and balance are normal, with no signs of instability or dizziness.

Reflexes: Her reflexes are normal, with no signs of hyporeflexia or hyperreflexia.

Assessment and Plan:

Assessment: Based on the patient's history and examination findings, it is assessed that she is experiencing recurrent seizures related to her parietal lobe epilepsy. The recent seizures may be related to the ongoing RCT, but this cannot be definitively determined without further follow-up. The patient's current medications are effective in controlling her seizures, and no changes will be made at this time except for the addition of the investigational drug for epilepsy.

Plan: The patient will continue to take her current medications as prescribed, with the addition of the investigational drug for epilepsy for the next three months as part of the RCT. She will follow up with the neurologist in one month to assess her response to the new medication and to monitor any changes in her seizure activity. The patient will also be advised to return immediately if she experiences any new or worsening symptoms, such as increased seizure frequency or severity, or any other concerning signs or symptoms.

----------------END OF PATIENT ENCOUNTER---------------

-------------- PATIENT ID 164 -- VISIT 2 -----------

On January 13, 2022, I conducted a clinic visit with Patient ID 164, an 83-year-old female with a history of parietal lobe epilepsy. The patient has been participating in a randomized controlled trial (RCT) and has taken the investigational drug ID X13737 for epilepsy over the past three months. During this time, the patient has experienced 12 seizures, but has reported no other symptoms.

Upon examination, the patient's neurological status was stable and there were no changes in her condition since the last visit. The patient's cognitive function was intact and she was able to communicate effectively. Her motor function was normal, with no signs of weakness or paresis. Her sensory examination revealed no deficits in hearing, vision, or sensation.

The patient's randomized controlled trial is now complete, and the investigational drug ID X13737 will be discontinued. The patient will be monitored closely for any changes in her condition and will continue to receive standard care for her epilepsy.

In conclusion, Patient ID 164 has completed her participation in the RCT and has experienced 12 seizures during the three-month treatment period with ID X13737. The patient's neurological status is stable and there are no changes in her condition since the last visit. The investigational drug will be discontinued, and the patient will continue to receive standard care for her epilepsy.

----------------END OF PATIENT ENCOUNTER---------------

-------------- PATIENT ID 165 -- VISIT 1 -----------

Here's the clinic note for Patient ID 165, a 35-year-old female with a history of epilepsy:

Patient ID: 165

Date: 2021-10-20

Chief Complaint: Increased seizure activity

History:

* 35-year-old female

* History of epilepsy

* Previous AEDs: Levetiracetam, Lamotrigine

* Currently taking: Levetiracetam, Lamotrigine

* Recent seizures: 49 in past 2 months

* Randomized controlled trial (RCT) participant

Neurological Examination:

* General: Alert and oriented

* Mental Status: Normal

* Cranial Nerves: Normal

* Motor: No weakness or tremors

* Sensory: Normal

* Coordination and Balance: Normal

* Reflexes: Normal

Pertinent Positives:

* Increased seizure activity (49 seizures in past 2 months)

* Participating in RCT with investigational drug for epilepsy

Assessment and Plan:

* Continue Levetiracetam and Lamotrigine as prescribed

* Add investigational drug for epilepsy per RCT protocol

* Monitor seizure activity closely during 3-month trial period

* Follow up in 1 month to assess tolerability and efficacy of investigational drug.

----------------END OF PATIENT ENCOUNTER---------------

-------------- PATIENT ID 165 -- VISIT 2 -----------

Patient ID: 165

Date: 2021-11-20

HPI:

* Female, 35 years old

* History of epilepsy

* Enrolled in randomized controlled trial (RCT) for investigational drug ID X13737

* Took drug for 3 months

* Reported 38 seizures during that time

Neurologic Examination:

* Vitals normal

* Head: normocephalic, atraumatic

* Eyes: no abnormalities

* Ears: no abnormalities

* Mouth: normal

* Neck: normal

* Extremities: normal

* Sensory: normal

* Motor: normal

* Reflexes: normal

Pertinent Positives:

* No symptoms reported

* 38 seizures during 3-month period of taking investigational drug ID X13737

Assessment and Plan:

* Discontinue investigational drug ID X13737 due to completion of RCT

* Monitor for any changes in seizure frequency or severity

* Schedule follow-up visit in 2 weeks to reassess patient's condition

----------------END OF PATIENT ENCOUNTER---------------

-------------- PATIENT ID 166 -- VISIT 1 -----------

Patient ID: 166

Date of Visit: 2022-11-23

Identification:

Name: Mary Johnson

Age: 73 years

Gender: Female

HPI (History of Present Illness):

Chief Complaint: The patient presents with a history of increased seizure activity over the past two months, with 15 seizures occurring during this time. She reports no significant changes in her medical history, medications, or social history.

Subjective:

The patient reports feeling "extremely anxious" and experiencing "increased stress" due to the recent increase in seizure activity. She denies any visual hallucinations or changes in mood. The patient reports difficulty sleeping due to increased seizure activity at night.

Medications:

The patient is currently prescribed:

* Levetiracetam 500mg BID

* Lamotrigine 200mg OD

* Valproate 500mg TID

Allergies:

The patient has no known allergies.

Social History:

The patient is a retired school teacher and lives with her husband. She reports no significant changes in her social history or support system.

Review of Systems:

General: The patient appears well-nourished and well-developed, with no signs of distress.

Neurological: The patient reports no numbness, tingling, or weakness in her extremities. She denies any recent headaches or changes in vision.

Psychiatric: The patient reports no significant changes in her mood or thought patterns.

Family History: The patient's father had a history of seizures and died from a seizure-related cause at the age of 65.

Assessment:

* The patient's recent increase in seizure activity is concerning, particularly given her history of multifocal epilepsy.

* The patient's current medications appear to be effective in controlling her seizures, and no changes will be made at this time other than the addition of the investigational drug for epilepsy.

* The patient's anxiety and stress levels are elevated due to the recent increase in seizure activity.

* The patient's sleep disturbances may be related to her increased seizure activity.

Comprehensive Neurologic Exam:

Motor: The patient has normal strength and tone in all extremities, with no signs of weakness or paralysis.

Sensory: The patient has normal sensation in all extremities, with no signs of numbness or tingling.

Cerebellar Function: The patient has normal coordination and balance, with no signs of ataxia or dysmetria.

Memory and Cognition: The patient appears to have normal memory and cognitive function, with no signs of decline or confusion.

Labs:

* Complete Blood Count (CBC): Within normal limits

* Electrolyte Panel: Within normal limits

* Liver Function Tests (LFTs): Within normal limits

Imaging:

* Magnetic Resonance Imaging (MRI) of the brain with and without contrast: No abnormalities were noted.

Assessment:

* The patient's recent increase in seizure activity is concerning, particularly given her history of multifocal epilepsy.

* The patient's current medications appear to be effective in controlling her seizures, and no changes will be made at this time other than the addition of the investigational drug for epilepsy.

* The patient's anxiety and stress levels are elevated due to the recent increase in seizure activity.

* The patient's sleep disturbances may be related to her increased seizure activity.

Plan:

1. Continue current medications, including levetiracetam, lamotrigine, and valproate.

2. Add the investigational drug for epilepsy, with careful monitoring of seizure activity and medication side effects.

3. Increase patient education on proper medication adherence and seizure management techniques.

4. Consider referral to a mental health professional for management of anxiety and stress.

5. Encourage the patient to maintain a consistent sleep schedule and practice good sleep hygiene.

Signature:

[Your Name]

Neurologist

Date: 2022-11-23

----------------END OF PATIENT ENCOUNTER---------------

-------------- PATIENT ID 166 -- VISIT 2 -----------

Patient ID: 166

Visit Date: 2022-12-23

The patient, a 73-year-old female with a history of asthma, autism, depression, and multifocal epilepsy, presented for her second visit in the randomized controlled trial. Over the past three months, the patient has been administered the investigational drug ID X13737 for her epilepsy, and during this time, she has experienced 37 seizures. The patient also reported experiencing dizziness.

On examination, the patient's neurological status was as follows:

* Mental Status: The patient's cognitive function was normal, with no evidence of decline or impairment.

* Speech and Language: The patient's speech was clear and coherent, with no noticeable difficulties or abnormalities.

* Motor Function: The patient's motor function was generally intact, with no significant tremors, rigidity, or weakness observed. However, there was a mild degree of dizziness noted during the examination.

* Sensory Function: The patient's sensory function was largely normal, with no deficits in vision, hearing, or sensation noted.

* Reflexes: The patient's reflexes were generally brisk and appropriate to the stimuli applied.

Given the patient's history of multifocal epilepsy and the significant number of seizures experienced during the study period, it is apparent that the investigational drug ID X13737 has had a limited impact on the patient's condition. Additionally, the patient's report of dizziness suggests a potential adverse effect of the medication. In light of these findings and the completion of the study, it is recommended that the investigational drug be discontinued, and alternative therapies be considered to address the patient's ongoing seizure activity.

----------------END OF PATIENT ENCOUNTER---------------

-------------- PATIENT ID 167 -- VISIT 1 -----------

Clinic Note Visit #1

Patient ID: 167

Date: 2022-01-19

Identification:

* Name: Mary Johnson

* Date of Birth: March 15, 1958

* Contact Information: (home) 123 Main St, Anytown, USA; (cell) 555-1234; (work) 456-7890

* Marital Status: Married

* Occupation: Retired Teacher

History of Present Illness (HPI):

Mary Johnson, a 64-year-old female, presented to the clinic with a history of hypertension and frontal lobe epilepsy. She has experienced 13 seizures in the past two months, which have been well-documented by her family members and caregivers. The patient reports no significant changes in her medication regimen or lifestyle factors that may have contributed to the increase in seizure activity.

Subjective:

Mrs. Johnson reported a persistent feeling of anxiety and fear associated with the recent increase in seizures. She also mentioned experiencing more frequent memory lapses and difficulty concentrating, which have affected her daily activities and social interactions. The patient's mood was observed to be anxious and depressed, with tearfulness and irritability noted during the encounter.

Medications:

* Levothyroxine (T4) 50 mcg oral daily

* Lisinopril 20 mg oral daily

* Valproic acid 500 mg oral BID

* Lamotrigine 100 mg oral daily

Allergies:

* None reported

Social History:

* Married with two adult children and four grandchildren

* Retired teacher with no known allergies or sensitivities

* Denies smoking, alcohol, or illicit drug use

* No recent travel or exposure to potential seizure triggers

Review of Systems:

* General: No fever, chills, or weakness reported

* Neurological: Recent seizures with loss of consciousness and postictal confusion (13 episodes in past 2 months)

* Cardiovascular: Hypertension (blood pressure: 140/90 mmHg)

* Respiratory: No reported respiratory symptoms or abnormalities

* Gastrointestinal: No reported gastrointestinal symptoms or abnormalities

General Physical Examination:

* Vital Signs: Temperature 98.6°F, Blood Pressure 140/90 mmHg, Heart Rate 70 bpm, Respiratory Rate 12 cpm

* Head and Eyes: Pupils equal and reactive to light, no visual field defects or oculomotor abnormalities noted

* Neck, Back, and Extremities: No significant muscle wasting or weakness observed

* Sensory Examination: Intact sensation to light touch, pinprick, and temperature over all body regions

* Coordination and Balance: Normal gait and station with no obvious ataxia or dyskinesias noted

Comprehensive Neurological Examination:

* Cranial Nerves: II-XII grossly intact, no evidence of focal deficits or weakness

* Motor Examination: Normal tone and strength in all muscle groups, with no evidence of atrophy or weakness

* Sensory Examination: Intact sensation to light touch, pinprick, and temperature over all body regions

* Reflexes: 3+/5 (normal)

Labs:

* Complete Blood Count (CBC): Within normal limits

* Basic Metabolic Panel (BMP): Within normal limits

* Thyroid-Stimulating Hormone (TSH): 4.2 mIU/L (normal 0.5-4.5 mIU/L)

Imaging:

* Computed Tomography (CT) scan of the brain with and without contrast: No acute intracranial abnormalities or lesions noted

* Magnetic Resonance Imaging (MRI) of the brain with and without contrast: No acute intracranial abnormalities or lesions noted

Assessment:

* Mrs. Johnson presents with a history of hypertension and frontal lobe epilepsy, with recent increase in seizure activity.

* Neurological examination revealed no focal deficits or weakness, with intact sensation and reflexes.

* Laboratory results were within normal limits, except for slightly elevated TSH levels.

* Imaging studies did not reveal any acute intracranial abnormalities or lesions.

Plan:

1. Continue current medications (levothyroxine, lisinopril, valproic acid, and lamotrigine) with close monitoring of seizure activity and medication adherence.

2. Add the investigational drug for epilepsy (drug name: [insert drug name here], dosage: [insert dosage here], duration of treatment: [insert duration here]) to the patient's regimen for a period of 3 months, with careful assessment of seizure activity and medication adherence.

3. Schedule follow-up appointments every 2 weeks to monitor seizure activity, medication adherence, and potential side effects of the investigational drug.

4. Encourage patient and family members to maintain a detailed seizure log to track the frequency and severity of seizures during the trial period.

5. Counsel patient on the importance of adherence to medication regimen and the need for regular follow-up appointments to monitor seizure activity and medication side effects.

Signature:

[Insert your signature here]

Date: 2022-01-19

----------------END OF PATIENT ENCOUNTER---------------

-------------- PATIENT ID 167 -- VISIT 2 -----------

Clinic Note - Visit #2

Date: 2022-02-19

Patient ID: 167

Age: 64 years old

Gender: Female

Chief Complaint: Hypertension, frontal lobe epilepsy

History of Present Illness: The patient has been participating in a randomized controlled trial for the investigational drug ID X13737 for her epilepsy over the past 3 months. During this time, she has experienced 25 seizures and reported headaches. The study is now complete, and the investigational drug will be discontinued.

Neurological Examination:

The patient's neurological examination was within normal limits, with no evidence of focal deficits or abnormalities in sensation, strength, or reflexes. Her speech and language were intact, and she had no difficulty with tandem gait or maintaining her balance. The patient did report experiencing frequent headaches, which may be related to the investigational drug.

Assessment and Plan:

Based on the patient's history of hypertension and frontal lobe epilepsy, as well as her recent experience with the investigational drug ID X13737, we have concluded that the study has been successful in demonstrating the safety and efficacy of this new treatment option. However, due to the high frequency of seizures and headaches experienced by the patient during the study period, we have decided to discontinue the investigational drug at this time. The patient will be followed closely in the coming months to monitor for any changes in her condition and to ensure that her hypertension remains well-controlled. Regular follow-up appointments will be scheduled to assess her progress and make any necessary adjustments to her treatment plan.

----------------END OF PATIENT ENCOUNTER---------------

-------------- PATIENT ID 168 -- VISIT 1 -----------

On August 17, 2021, Patient ID 168, a 78-year-old female with a history of frontal lobe epilepsy, presented to our clinic for her first visit in the randomized controlled trial for the treatment of her condition. The patient has experienced 10 seizures over the past two months, and we have decided not to make any changes to her current medications other than adding the investigational drug for epilepsy that she will take for three months.

Upon examination, the patient was alert and oriented, with no evidence of focal neurological deficits or abnormalities in her speech, language, or motor functions. Her mood and affect were appropriate to her age and condition, and there were no signs of anxiety or depression. The results of her neurological examination were as follows:

* Mental status: Alert and oriented x3

* Memory: Intact

* Attention and concentration: Normal

* Language: Intact

* Motor function: Normal

* Sensation: Intact

* Reflexes: Normal

The patient's medical history, including her frontal lobe epilepsy and recent seizure activity, was reviewed in detail. She has been experiencing an average of two seizures per month over the past two months, which is consistent with her previous history. The patient reports no significant changes in her symptoms or quality of life since her last visit.

Based on the results of the neurological examination and the patient's medical history, we have decided to proceed with the randomized controlled trial for the treatment of her frontal lobe epilepsy. The patient will be administered the investigational drug for a period of three months, during which time we will closely monitor her seizure activity and adjust her medications as needed. We will also perform regular follow-up examinations to assess her progress and make any necessary adjustments to her treatment plan.

----------------END OF PATIENT ENCOUNTER---------------

-------------- PATIENT ID 168 -- VISIT 2 -----------

On September 17th, 2021, I conducted a routine clinic visit with Patient ID 168, a 78-year-old female who has been participating in a randomized controlled trial for the investigational drug ID X13737 for frontal lobe epilepsy. Over the past three months, the patient has taken the medication and has experienced eight seizures. However, the patient has reported no other symptoms during this time.

Upon conducting a complete neurological examination, I observed that the patient's cognitive function remains stable, with no significant changes in her memory, attention, or executive function. Her speech and language skills are also unchanged. Additionally, there is no evidence of any motor weakness or atrophy. The patient's vision and hearing are both normal, and she has not experienced any visual or auditory hallucinations.

Given the results of the study, the investigational drug ID X13737 will be discontinued, as it is no longer necessary for the patient's treatment. I will continue to monitor the patient's condition closely and adjust her medication regimen as needed to ensure optimal seizure control. It is essential that the patient continues to receive regular follow-up care to assess the efficacy of the discontinued medication and address any potential side effects or complications.

In conclusion, this clinic visit has provided valuable insights into the effectiveness and safety of the investigational drug ID X13737 for frontal lobe epilepsy. The patient's stable cognitive function and lack of significant symptoms indicate that the medication has been well-tolerated, and the study has successfully achieved its objectives. As we move forward, it is crucial that we continue to monitor the patient's condition closely to ensure the best possible outcome for her treatment.

----------------END OF PATIENT ENCOUNTER---------------

-------------- PATIENT ID 169 -- VISIT 1 -----------

On May 27th, 2022, I conducted a clinic visit with Patient ID 169, a 56-year-old female with a rich medical history of diabetes, depression, hypothyroidism, and generalized epilepsy. The patient has been experiencing increased seizure activity in the past two months, with six seizures reported during this timeframe. As part of a randomized controlled trial, the patient will continue her current medications without any changes, with the addition of an investigational drug specifically designed to treat her epilepsy for a period of three months.

During the neurological examination, the patient's vital signs were within normal limits. Her consciousness was clear, and her speech was normal. The patient's visual fields were full, and she had no signs of weakness or paresthesias in her limbs. Her sensory examination revealed a decrease in pain sensation to light touch and pinprick over the left hemiparesis area, consistent with her previous history of seizure activity. The patient's cranial nerves were intact, and her coordination and balance were normal.

The patient reported no changes in her seizure activity since our last visit, and her mood remained stable without any notable changes in her depression symptoms. Her hypothyroidism is currently well-controlled, and her diabetes management has been consistent with regular monitoring and appropriate medication adjustments. The patient was informed about the randomized controlled trial and the added investigational drug for epilepsy, and she expressed her willingness to participate in the study.

In conclusion, this clinic visit revealed no significant changes in Patient ID 169's medical condition, with her seizure activity remaining a concern. The addition of the investigational drug for epilepsy is expected to provide better control over her seizures, and we will closely monitor her progress throughout the trial period. Regular follow-up appointments have been scheduled to ensure proper care and adjust medications as needed.

----------------END OF PATIENT ENCOUNTER---------------

-------------- PATIENT ID 169 -- VISIT 2 -----------

Clinic Note Visit #2 dated 2022-06-27 for Patient ID 169:

Patient ID 169, a 56-year-old female, presented for her second visit in the randomized controlled trial. The patient has a history of diabetes, depression, hypothyroidism, and generalized epilepsy. Over the past three months, the patient has been taking the investigational drug ID X13737 for her epilepsy. During this time, the patient has experienced seven seizures. Additionally, the patient reported feeling fatigued.

On examination, the patient's vital signs were within normal limits. Her neurological examination revealed no significant abnormalities, with normal strength, sensation, and reflexes in all four limbs. However, the patient did report ongoing fatigue, which was noted in her medical history. The patient's speech was clear and coherent, and her mental status was intact.

The investigational drug ID X13737 will be discontinued now that the study is complete. The patient will be transitioned back to her usual medications for epilepsy management. The patient will be closely monitored for any changes in her seizure activity and overall health. Regular follow-up appointments will be scheduled to assess her condition and adjust her treatment plan as needed.

In conclusion, Patient ID 169 has completed the randomized controlled trial and has experienced seven seizures during the three-month period of taking the investigational drug ID X13737. The patient reported ongoing fatigue, which will be closely monitored and managed accordingly. The drug will be discontinued, and the patient will be transitioned back to her usual medications for epilepsy management. Regular follow-up appointments will be scheduled to assess her condition and adjust her treatment plan as needed.

----------------END OF PATIENT ENCOUNTER---------------

-------------- PATIENT ID 180 -- VISIT 1 -----------

Clinic Note #1

Patient ID: 180

Date: 2023-01-22

Chief Complaint: The patient presents with a history of schizophrenia and multifocal epilepsy. She has had 12 seizures in the past 2 months.

Past Medical History: Schizophrenia, multifocal epilepsy, hypertension, hyperlipidemia

Past Surgical History: None

Medications: Lithium (600 mg/day), Levetiracetam (2000 mg/day), Lamotrigine (200 mg/day), Valproic acid (1000 mg/day)

Family History: None

Social History: The patient is a 94-year-old female who lives independently and has a support system in place. She denies any alcohol or tobacco use.

Review of Systems: No significant changes in the patient's mental status, mood, or cognitive function. No changes in her seizure activity since her last visit.

Neurological Examination:

* Mental Status: The patient is alert and oriented to person, place, and time. Her speech is clear and coherent. She denies any hallucinations or delusions.

* Motor Examination: The patient has no signs of tremors, rigidity, or bradykinesia. Her gait is normal, and she has no signs of ataxia or dysmetria.

* Sensory Examination: The patient has no sensory deficits. She has a normal sense of touch, pain, and temperature perception.

* Cranial Nerve Examination: No abnormalities were noted in the patient's cranial nerves.

* Neuropsychiatric Examination: The patient's mood is euthymic, and she denies any suicidal or homicidal ideation. Her affect is appropriate to her age and medical condition. She has no signs of depression, anxiety, or mania.

Assessment:

* The patient's history of schizophrenia and multifocal epilepsy is consistent with her current symptoms.

* The recent increase in seizure activity is concerning and may be related to the ongoing randomized controlled trial.

* The patient's cognitive function remains intact, and her mental status is stable.

Plan:

1. Continue the patient's current medications, including lithium, levetiracetam, Lamotrigine, and valproic acid.

2. Add the investigational drug for epilepsy to the patient's regimen for a period of 3 months.

3. Monitor the patient's seizure activity closely during the trial period.

4. Schedule follow-up visits every 2 weeks to assess the patient's progress and make any necessary adjustments to her medications.

5. Encourage the patient to maintain a consistent sleep schedule, avoid alcohol and caffeine, and engage in regular physical activity to minimize seizure activity.

I have discussed the above plan with the patient, and she understands the purpose and potential risks of the investigational drug. She has given her informed consent to participate in the randomized controlled trial.

----------------END OF PATIENT ENCOUNTER---------------

-------------- PATIENT ID 180 -- VISIT 2 -----------

<<SYS>><<BR>> <<BR>>

Patient ID: 180

Date: 2023-02-22

Identification:

We present the clinic note for Patient ID 180, a 94-year-old female with a history of schizophrenia and multifocal epilepsy. This patient has been enrolled in a randomized controlled trial (RCT) evaluating the investigational drug ID X13737 for the treatment of epilepsy.

Highlights of the Patient's History:

* Schizophrenia (diagnosed at age 60)

* Multifocal epilepsy ( diagnosed at age 70)

* Previous admission to our facility for status epilepticus

* Currently taking levetiracetam, lamotrigine, and valproate for seizure control

* Recently discontinued ID X13737 as part of the RCT

Subjective:

The patient reported no symptoms since her last visit. She denied any recent seizures or changes in mental status. She was cooperative and alert during the examination, with a normal mood and affect. The patient's speech was clear and coherent, without any tangential thinking or hallucinations.

Medications:

* Levetiracetam (Keppra) 500mg BID

* Lamotrigine (Lamictal) 200mg OD

* Valproate (Depakote) 1000mg TID

* ID X13737 (discontinued as part of the RCT)

Allergies:

* No known allergies

Social History:

* Married with two adult children and four grandchildren

* Retired school teacher

* Active in local community and church groups

Review of Systems:

* General: Denied fever, chills, or weakness

* Neurological: No changes in seizure frequency or severity since last visit

* Psychiatric: No changes in mood or thought content

General Physical Examination:

The patient's vital signs were as follows: blood pressure 120/80 mmHg, heart rate 70 beats per minute, respiratory rate 14 breaths per minute, and temperature 98.6 degrees Fahrenheit. Her weight was 55 kg, and her body mass index was 23. The patient's head was normocephalic, and her eyes were normo-ocular. Her speech was clear and coherent, without any tangential thinking or hallucinations. She had a normal gait and station, with no ataxia or tremors.

Comprehensive Neurologic Exam:

The patient's neurological examination was unremarkable, with no focal deficits or abnormalities noted. Her speech was clear and coherent, without any tangential thinking or hallucinations. She had a normal gait and station, with no ataxia or tremors. The patient's pupils were equal and reactive to light, with no signs of afferent pupillary defect. Her funduscopic examination was unremarkable, without any evidence of retinal hemorrhage or exudate.

Labs:

* Complete Blood Count (CBC): Within normal limits

* Electrolyte Panel (EVP): Within normal limits

* Renal Function Panel (RFP): Within normal limits

* Thyroid-stimulating Hormone (TSH): Within normal limits

Imaging:

* Brain Computerized Tomography (CT) scan performed on 2023-02-15 revealed no evidence of acute intracranial injury or abnormalities.

Assessment:

The patient is a 94-year-old female with a history of schizophrenia and multifocal epilepsy. She has been enrolled in a randomized controlled trial evaluating the investigational drug ID X13737 for the treatment of epilepsy. The patient reported no symptoms since her last visit, and there were no changes in her neurological examination. The patient's seizure frequency has been well-controlled on her current medications, with no recent seizures or changes in mental status. Given the completion of the RCT, the patient will be discontinuing ID X13737 and continuing her current medication regimen.

Plan:

* Continue levetiracetam, lamotrigine, and valproate as prescribed

* Discontinue ID X13737 as part of the RCT

* Monitor the patient's seizure frequency and mental status at follow-up visits

Signature:

[Your Name]

Neurologist

Date: 2023-02-22

Note: This clinic note is a fictional example for educational purposes only. It should not be used as a basis for actual patient care or medical decision-making.

----------------END OF PATIENT ENCOUNTER---------------

-------------- PATIENT ID 181 -- VISIT 1 -----------

Patient ID: 181

Age: 63 yrs

History: Dermatitis, Hypothyroidism, Schizophrenia, Occipital Lobe Epilepsy

Seizure History: 13 seizures in past 2 months

Current Medications: None (randomized controlled trial)

Adding Investigational Drug for Epilepsy (3 months)

Neurological Examination:

* Alert and oriented x3

* No focal deficits

* Normal muscle strength and tone

* Normal sensation in extremities

* No abnormal reflexes

* No dysmetria or tremors

Pertinent Positives:

* No changes in seizure frequency or severity since starting trial

* No adverse effects from investigational drug

Assessment and Plan:

* Continue with randomized controlled trial for 3 months

* Monitor seizure frequency and adverse effects of investigational drug closely

* Consider dose adjustments as needed based on tolerability and efficacy

* Follow-up visit in 1 month to reassess seizure control and monitor for any adverse effects.

----------------END OF PATIENT ENCOUNTER---------------

-------------- PATIENT ID 181 -- VISIT 2 -----------

---

Clinic Note #2, Patient ID 181, 2023-02-24

Identification:

Name: John Doe

Age: 63 years

Gender: Male

Patient ID: 181

Date of Birth: January 15, 1959

Address: 123 Main St, Anytown, USA

HPI:

Chief Complaint: Follow-up visit for randomized controlled trial of investigational drug ID X13737 for epilepsy.

History of Present Illness: Patient was enrolled in a randomized controlled trial for the investigational drug ID X13737 for the treatment of occipital lobe epilepsy. The patient has been taking the medication for three months and has had 29 seizures during that time. The patient reports no symptoms.

Subjective:

Patient reported no complaints or symptoms other than the seizures.

Medications:

1. ID X13737 (Investigational drug for epilepsy) - Discontinued due to completion of study

2. Levetiracetam (Keppra) - 500 mg oral twice daily

3. Lamotrigine (Lamictal) - 100 mg oral once daily

4. Levothyroxine (Synthroid) - 25 mcg oral once daily

Allergies:

None reported.

Social History:

Married with two adult children. Patient is a retired mechanic and enjoys woodworking in his free time.

Review of Systems:

General: No fever, chills, or weakness.

Neurological: No change in seizure activity or frequency.

Musculoskeletal: No joint pain or swelling.

Cardiovascular: No chest pain or shortness of breath.

Respiratory: No cough or difficulty breathing.

Gastrointestinal: No nausea, vomiting, or diarrhea.

Urogenital: No urinary frequency, hesitancy, or pain.

Head and Neck: No headaches, facial weakness, or vision changes.

General Physical Examination:

Vital Signs:

BP: 120/80 mmHg

HR: 70 bpm

T: 98.6°F

RR: 14 breaths/min

Weight: 75 kg

Height: 175 cm

General: Patient appears well-nourished and well-groomed. No obvious skin lesions or abnormalities.

Comprehensive Neurological Examination:

Mental Status: Alert and oriented x3. Patient is able to name the current President of the United States (Joe Biden) and the date (February 24, 2023).

Cranial Nerves:

II - Optic disc margins are normal.

III - No facial weakness or paresis.

IV - No weakness or paresis of the tongue.

V - No difficulty with swallowing or sensation in the throat.

VI - No hearing loss or vestibular abnormalities.

VII - No facial or bulbar signs of weakness.

VIII - No dysfunction of the vestibulo-ocular reflex.

IX - No abnormality of the olfactory nerve.

X - No hemiparesis or weakness of the left arm and leg.

Sensory:

No sensory deficits in the arms and legs.

Motor:

No motor deficits in the arms and legs.

Reflexes:

Biceps reflex is 2+ (normal is 1-4+).

Evoked Potentials:

No abnormality in the brainstem auditory evoked potentials.

Labs:

Complete Blood Count (CBC) - Normal

Blood Chemistry - Normal

Imaging:

MRI of the brain - Normal

Assessment:

The patient is a 63-year-old male with a history of dermatitis, hypothyroidism, schizophrenia, and occipital lobe epilepsy. The patient has been participating in a randomized controlled trial for the investigational drug ID X13737 for the treatment of occipital lobe epilepsy. The patient has had 29 seizures during the past three months, with no change in seizure frequency or severity. The patient reports no symptoms other than the seizures. The investigational drug will be discontinued due to completion of the study.

Plan:

1. Continue levetiracetam (Keppra) and lamotrigine (Lamictal) as prescribed.

2. Discontinue ID X13737 (Investigational drug for epilepsy) due to completion of study.

3. Follow-up visit in one month to assess seizure activity and medication adherence.

4. Consider initiation of additional anticonvulsant medications if seizure activity persists or worsens.

5. Monitor patient for signs of hypothyroidism and adjust levothyroxine (Synthroid) as needed.

6. Encourage patient to continue woodworking hobby and avoid activities that may exacerbate seizures.

7. Re-evaluate patient in one month for changes in seizure activity or medication adherence.

Signature:

[Your Signature]

Date: February 24, 2023

----------------END OF PATIENT ENCOUNTER---------------

-------------- PATIENT ID 182 -- VISIT 1 -----------

CLINIC NOTE DATE: 2022-03-13

PATIENT ID: 182

AGE: 93 years old

SEX: Male

CHIEF COMPLAINT: Recurrent seizures

HISTORY OF PRESENT ILLNESS (HPI): The patient presents with a history of recurrent seizures over the past two months. He has experienced 11 seizures during this time, with no significant triggers or warnings. The patient has a history of parietal lobe epilepsy and has been undergoing a randomized controlled trial for the past year.

SUBJECTIVE: The patient reports no recent changes in his symptoms, other than the ongoing seizures. He denies any new medications or substances that may be contributing to the seizures. He is currently taking levetiracetam and sodium valproate for his epilepsy, and these medications have been unchanged for the past year.

MEDICATIONS:

* Levetiracetam (Keppra) 500mg orally BID

* Sodium valproate (Depakote) 500mg orally TID

ALLERGIES: None reported

SOCIAL HISTORY: The patient is a retired accountant and enjoys spending time with his family. He denies any recent travel or exposure to potential triggers for his seizures.

REVIEW OF SYSTEMS:

* General: The patient appears well-oriented and alert, with no significant changes in his general condition.

* Neurological: The patient reports ongoing seizures, with no recent changes in frequency or severity. He denies any new symptoms or weaknesses.

* Cardiovascular: The patient has a history of hypertension, which is well-controlled with medication.

* Respiratory: The patient has a history of chronic obstructive pulmonary disease (COPD), which is also well-controlled with medication.

GENERAL PHYSICAL EXAMINATION:

* Vital signs: Blood pressure 120/80 mmHg, heart rate 70 bpm, temperature 98.6°F (37°C)

* General appearance: The patient appears well-nourished and well-groomed, with no significant changes in his physical condition.

* Head and neck: No significant abnormalities noted.

* Extremities: No significant abnormalities noted.

* Chest and abdomen: No significant abnormalities noted.

* Neurological examination: The patient exhibits a normal gait and station, with no signs of ataxia or dysmetria. His sensory examination is intact, with no signs of paresthesias or numbness. His cognitive function is intact, with no signs of confusion or disorientation.

COMPREHENSIVE NEUROLOGIC EXAMINATION:

* Mental status: The patient appears alert and oriented to person, place, and time. His speech is clear and concise, with no signs of aphasia or dysarthria.

* Cranial nerves: The patient has normal visual acuity, with no signs of diplopia or abnormal eye movements. His hearing is normal, with no signs of conductive or sensorineural hearing loss. His facial movements are normal, with no signs of weakness or paresis.

* Motor examination: The patient exhibits a normal gait and station, with no signs of ataxia or dysmetria. His muscle strength is normal, with no signs of weakness or wasting. His reflexes are normal, with no signs of hyporeflexia or hyperreflexia.

* Sensory examination: The patient's sensory examination is intact, with no signs of paresthesias or numbness.

LABS:

* Complete blood count (CBC): Normal

* Electrolyte panel: Normal

* Thyroid-stimulating hormone (TSH): Normal

* Urinalysis: Normal

IMAGING:

* Computed tomography (CT) scan of the head: No significant abnormalities noted.

* Magnetic resonance imaging (MRI) of the brain: No significant abnormalities noted.

ASSESSMENT:

* The patient presents with a history of recurrent seizures, with no recent changes in his symptoms. He is currently participating in a randomized controlled trial for his epilepsy and has been taking levetiracetam and sodium valproate for the past year. There are no significant changes in his laboratory or imaging findings. The patient's neurological examination is normal, with no signs of ataxia, dysmetria, or weakness.

* Given the patient's history of parietal lobe epilepsy and ongoing seizures, it is important to continue his current medications and monitor his condition closely. The patient should be counseled on the potential risks and benefits of the investigational drug being used in the randomized controlled trial.

* No changes will be made to the patient's current medication regimen at this time, other than adding the investigational drug for the next 3 months. The patient should be closely monitored for any changes in his symptoms or adverse effects from the medications.

PLAN:

* Continue levetiracetam and sodium valproate as prescribed

* Add investigational drug for the next 3 months

* Close monitoring of the patient's symptoms and adverse effects from the medications

* Follow-up visit in 1 month to assess the patient's condition and adjust the treatment plan as needed

SIGNATURE:

[Your name and title]

Date: 2022-03-13

----------------END OF PATIENT ENCOUNTER---------------

-------------- PATIENT ID 182 -- VISIT 2 -----------

Clinic Note: Patient ID 182, Visit #2 (2022-04-13)

Identification:

Patient Name: John Doe

Patient ID: 182

Date of Birth: January 12, 1929

Gender: Male

Age: 93 years

History of Present Illness (HPI):

Mr. Doe is a 93-year-old male who presents with a history of parietal lobe epilepsy and a recent trial of the investigational drug ID X13737 for the treatment of his seizures. He has been experiencing gait disturbance and has had eight seizures over the past three months since initiating the medication.

Subjective:

On today's visit, Mr. Doe reports a worsening of his gait disturbance and difficulty walking short distances without assistance. He also reports a decrease in seizure frequency since discontinuing the investigational drug. He denies any recent changes in medications or substance use.

Medications:

ID X13737 (Investigational Drug): 3 months ago, 4 times daily

Levetiracetam (Keppra): Currently, 2 times daily

Lamotrigine (Lamictal): Currently, 1 time daily

Allergies:

No known allergies.

Social History:

Married with adult children.

Retired from manufacturing.

Hobbies include gardening and reading.

Review of Systems:

General: No recent illnesses or surgeries.

Neurological: History of parietal lobe epilepsy, recent trial of ID X13737, current medications include levetiracetam and lamotrigine.

Cardiovascular: Hypertension, well-controlled on medication.

Respiratory: Chronic obstructive pulmonary disease (COPD), well-controlled on medication.

General Physical Examination:

Vital signs:

Temperature: 98.6°F

Pulse: 70 beats per minute

Respiration: 12 breaths per minute

Blood Pressure: 130/80 mmHg

Comprehensive Neurological Examination:

Motor: Mr. Doe exhibits a mild right-sided hemiparesis with weakness in the right arm and leg. His left limbs are unaffected. He has difficulty walking short distances without assistance and displays a broad-based gait with a tendency to lean to the right.

Sensory: No sensory deficits were noted.

Cerebellar Function: Mr. Doe's cerebellar function is normal.

Labs:

Blood chemistry profile: Within normal limits.

Urinalysis: Negative for protein or blood.

Electroencephalogram (EEG): Performed 2 months ago, showed a normal background activity with no evidence of seizure activity.

Imaging:

Computed Tomography (CT) Scan: Performed 6 months ago, revealed a history of left parietal lobe infarction.

Magnetic Resonance Imaging (MRI): Performed 3 months ago, showed no evidence of active inflammation or structural abnormalities.

Assessment:

Mr. Doe is a 93-year-old male with a history of parietal lobe epilepsy who has been participating in a randomized controlled trial for the investigational drug ID X13737. Over the past three months, he has experienced eight seizures and has reported worsening gait disturbance. The investigational drug will be discontinued due to the completion of the study. His current medications include levetiracetam and lamotrigine. His neurological examination reveals a mild right-sided hemiparesis with difficulty walking short distances without assistance. His laboratory results are within normal limits, and his imaging studies show no evidence of active inflammation or structural abnormalities.

Plan:

1. Discontinue the investigational drug ID X13737 due to completion of the study.

2. Monitor Mr. Doe's seizure frequency and adjust medications as needed.

3. Refer him to physical therapy for gait disturbance and mobility issues.

4. Schedule a follow-up appointment in 3 months.

Signature:

[Your Signature]

Date: [Today's Date]

Note: This clinic note is a fictional example and does not represent any real patient or study. It is meant to demonstrate the format of a comprehensive neurological clinic note for a 93-year-old male with parietal lobe epilepsy participating in a randomized controlled trial.

----------------END OF PATIENT ENCOUNTER---------------

-------------- PATIENT ID 183 -- VISIT 1 -----------

Visit #1 (2022-10-30)

Patient ID: 183

Age: 23 years

Female

History:

* Multifocal epilepsy

* 144 seizures in past 2 months

* Randomized controlled trial (RCT)

Neurological Examination:

* Head: normocephalic, normal hemispheric symmetry

* Eyes: normal visual acuity, no nystagmus or gaze deviations

* Face: normal facial movements and expression

* Limbs: normal tone and reflexes, no atrophy or weakness

* Sensory: intact sensation in all extremities

* Mental Status: alert and oriented to person, place, and time

Pertinent Positives:

* Multifocal seizures

* Increased frequency of seizures in past 2 months

* Enrolled in RCT for investigational drug for epilepsy

Assessment and Plan:

* Continue with current medications for multifocal epilepsy

* Add investigational drug for epilepsy (RCT) for next 3 months

* Monitor seizure frequency and severity during trial period

* Follow-up visit in 2 weeks to assess tolerability and efficacy of new medication.

----------------END OF PATIENT ENCOUNTER---------------

-------------- PATIENT ID 183 -- VISIT 2 -----------

Visit #2: November 30, 2022

Patient ID 183, a 23-year-old female with a history of multifocal epilepsy, was seen in clinic today for her second visit. The patient has been participating in a randomized controlled trial (RCT) evaluating the investigational drug ID X13737 for the treatment of epilepsy. Over the past three months, the patient has taken the drug and has experienced 63 seizures during this time. Notably, the patient reported experiencing dizziness during the study period.

On examination, the patient's neurological status was as follows:

* Mental Status: The patient's cognitive function and mood were both normal, with no evidence of any clouding or impairment in her mental status.

* Speech: The patient's speech was clear and without any abnormalities.

* Motor Function: The patient had a normal strength and tone in all of her muscle groups, with no evidence of weakness, atrophy, or spasticity. Her reflexes were also normal.

* Sensation: The patient reported no significant changes in her sensory perception, including vision, hearing, touch, taste, or smell.

* Cranial Nerves: The patient's cranial nerves were normal, with no evidence of any dysfunction or abnormalities.

Given the completion of the RCT and the patient's experience of 63 seizures during the study period, it has been decided to discontinue the investigational drug ID X13737. The patient will be transitioned back to her previous medication regimen and will be closely monitored for any changes in her seizure activity. It is important to note that the patient's dizziness may be related to the medication or the study itself, and further evaluation and management will be necessary to address this symptom. Overall, the patient remains in good health and her neurological status is stable.

----------------END OF PATIENT ENCOUNTER---------------

-------------- PATIENT ID 184 -- VISIT 1 -----------

CLINIC NOTE #1

DATE: 2021-09-13

PATIENT ID: 184

CHIEF COMPLAINT:

The patient, a 53-year-old female with a history of epilepsy, presents with a chief complaint of increased seizure frequency over the past two months. She reports having experienced nine seizures during this time. The patient is currently enrolled in a randomized controlled trial and is taking no changes in her medications other than adding an investigational drug for epilepsy, which she will be taking for three months.

MEDICAL HISTORY:

The patient has a history of epilepsy and has been previously treated with levetiracetam and lamotrigine. She has experienced multiple seizures over the years, with an average frequency of one to two seizures per month. However, she has noticed an increase in seizure frequency over the past two months, with nine seizures occurring during this time.

PHYSICAL EXAMINATION:

On examination, the patient appears well-oriented and alert. Her speech is clear and her language is normal. There are no signs of weakness or paralysis. The patient's visual fields appear intact, with no signs of hemianopia or other visual field defects. The patient's extraocular movements are full and without any abnormalities.

Neurological examination reveals:

* Mildly decreased sensation to light touch and pinprick over the bilateral temporal lobes

* Bilateral papilledema, suggestive of increased intracranial pressure

* Normal tone and reflexes in all four limbs

* No signs of ataxia or dysmetria

* No signs of weakness or paralysis

ASSESSMENT AND PLAN:

Based on the patient's history of increased seizure frequency and the results of her neurological examination, it is suspected that the patient may be experiencing a worsening of her epilepsy. The patient's participation in the randomized controlled trial will continue as planned, with the addition of the investigational drug for epilepsy. The patient will be monitored closely over the next three months to assess the effectiveness of this treatment and to determine if any changes need to be made to her current medication regimen.

The patient will be advised to continue taking her current medications as prescribed, with the addition of the investigational drug for epilepsy. The patient will be instructed to report any changes in her seizure frequency or severity to her primary care physician or the study team immediately. The patient will also be advised to avoid any activities that may increase her risk of injury or harm, such as driving or operating heavy machinery, until her seizure frequency has been stabilized.

The patient will be scheduled for follow-up appointments at one month and three months after the start of the investigational drug to assess her response to treatment and to make any necessary adjustments to her medication regimen.

ORDERS:

1. Continue current medications as prescribed, with the addition of the investigational drug for epilepsy.

2. Monitor the patient closely over the next three months to assess the effectiveness of this treatment and to determine if any changes need to be made to her current medication regimen.

3. Schedule follow-up appointments at one month and three months after the start of the investigational drug to assess the patient's response to treatment and to make any necessary adjustments to her medication regimen.

----------------END OF PATIENT ENCOUNTER---------------

-------------- PATIENT ID 184 -- VISIT 2 -----------

---

Clinic Note #2, Patient ID 184, Date: 2021-10-13

Identification:

Patient name: [Patient Name]

Date of birth: [Date of Birth]

Gender: Female

Age: 53 years

History of Present Illness (HPI):

The patient presents with a history of epilepsy and has been participating in a randomized controlled trial (RCT) for the investigational drug ID X13737. The patient has been taking the medication for the past 3 months and has experienced 24 seizures during this time. The patient has also reported dizziness.

Subjective:

The patient reports a history of epilepsy, with a frequency of seizures that has increased over the past year. She also reports experiencing dizziness and vertigo. The patient's medication adherence has been excellent, taking her medications as prescribed without any deviations.

Medications:

* ID X13737 (investigational drug for epilepsy) - discontinued as of today

* Levetiracetam (Keppra) - 500mg oral twice daily

* Lamotrigine (Lamictal) - 200mg oral once daily

Allergies:

No known allergies.

Social History:

The patient is a married woman with two children, ages 12 and 15. She works as an accountant and reports no significant stressors in her life at this time. The patient denies any history of substance abuse or psychiatric illness.

Review of Systems:

* General: Denies fever, chills, or night sweats.

* Neurological: Reports increased seizure frequency over the past year, with a total of 24 seizures during the past 3 months while taking the investigational drug ID X13737.

* Cardiovascular: Denies chest pain, shortness of breath, or irregular heartbeat.

* Respiratory: Denies cough, difficulty breathing, or shortness of breath.

* Gastrointestinal: Reports no changes in bowel habits or appetite.

* Genitourinary: Denies dysuria, frequency, or urgency.

General Physical Examination:

* Vital signs: Blood pressure: 120/80mmHg, Heart rate: 70bpm, Temperature: 98.6°F (37°C)

* Head and neck: Normal gross motor function, no weakness or paresis.

* Extremities: Normal strength and tone, no atrophy or deformity.

* Sensory: No loss of sensation to light touch, pain, or temperature.

* Coordination and balance: Normal coordination and balance, no evidence of gait disturbances.

Comprehensive Neurologic Exam:

* Mental Status: Alert and oriented x3, no evidence of clouding or confusion.

* Cranial Nerves: II-XII normal, no deficits.

* Motor Exam: No gross motor deficits, 5/5 strength in all extremities.

* Sensory Exam: No sensory deficits to light touch, pain, or temperature.

* Reflexes: Normal reflexes, no hyporeflexia or hyperreflexia.

Labs:

* Complete Blood Count (CBC): WBC 4.5, Neutrophils 60%, Lymphocytes 30%, Monocytes 10%, Eosinophils 2%

* Electrolyte Panel: Sodium 138, Potassium 4.2, Chloride 102, Carbon Dioxide 22

* Liver Function Tests (LFTs): AST 25, ALT 30, Alkaline Phosphatase 125, Total Bilirubin 0.6

Imaging:

No recent imaging studies have been performed.

Assessment:

* The patient presents with a history of increased seizure frequency while taking the investigational drug ID X13737.

* The patient reports dizziness and vertigo, which may be related to the medication or her underlying epilepsy.

* The patient's neurological examination is unremarkable, with no evidence of focal deficits or abnormalities.

* Laboratory results are within normal limits, except for a mildly elevated liver function test (LFT) panel.

Plan:

* Discontinue the investigational drug ID X13737 as per study protocol.

* Continue levetiracetam (Keppra) and lamotrigine (Lamictal) as previously prescribed.

* Monitor the patient's seizure frequency and adverse effects closely over the next few months.

* Schedule a follow-up appointment in 3 months to reassess the patient's condition and adjust medications as needed.

Signature:

[Insert signature of healthcare provider]

Note: This clinic note is for a fictional patient and is not based on any real patient or study. It is meant to illustrate an example of a comprehensive neurological clinic note, including a detailed history, physical examination, laboratory results, and assessment.

----------------END OF PATIENT ENCOUNTER---------------

-------------- PATIENT ID 185 -- VISIT 1 -----------

Visit #1 (2021-09-10)

Patient ID: 185

Age: 34 years

History: Depression, temporal lobe epilepsy (TLE)

HPI:

* 14 seizures in past 2 months

* Randomized controlled trial (RCT)

Neurologic Examination:

* Head: normocephalic

* Eyes: normal visual acuity, no nystagmus or abnormal eye movements

* Face: normal facial movements and expression

* Mouth: normal oral hygiene, no tongue fasciculations

* Extremities: normal strength and tone, no atrophy or weakness

* Sensation: intact

* Reflexes: normal

Pertinent Positives:

* No changes in medications other than investigational drug for epilepsy

* Seizure frequency remains unchanged

Assessment and Plan:

* Continue RCT with no changes in medications except for investigational drug for epilepsy

* Monitor seizure frequency and adverse effects every 2 weeks

* Schedule follow-up visit in 2 months.

----------------END OF PATIENT ENCOUNTER---------------

-------------- PATIENT ID 185 -- VISIT 2 -----------

Clinic Note - Visit #2 (2021-10-10)

Patient ID: 185

Age: 34 years

Chief Complaint: None

History of Present Illness:

The patient is a 34-year-old male with a history of depression and temporal lobe epilepsy. He has been participating in a randomized controlled trial (RCT) and has been taking the investigational drug ID X13737 for epilepsy over the past 3 months. The patient has had 27 seizures during this time, but reports no symptoms. The RCT is now complete, and the investigational drug will be discontinued.

Review of Systems:

The patient denies any recent changes in his mental health or any new symptoms. He reports that he has been experiencing some fatigue, but this is not a significant change from his baseline.

Past Medical History:

Depression

Temporal lobe epilepsy

Medications:

ID X13737 (investigational drug for epilepsy)

Social History:

The patient denies any significant social or occupational changes. He reports that he is married and has two children, and he is currently working full-time as an accountant.

Family History:

No significant family history of neurological disorders.

General Examination:

Vital Signs:

* Temperature: 98.6°F (37°C)

* Blood Pressure: 120/80 mmHg

* Pulse: 70 bpm

* Respiration Rate: 14 breaths/min

Head and Neck Examination:

* Mental status: Alert and oriented to person, place, and time.

* Speech: Clear and appropriate.

* Language: Intact.

* Cranial Nerve Examination: All cranial nerves are intact and normal.

* Motor Examination: All muscle groups are intact and normal, with no evidence of weakness or atrophy.

* Sensory Examination: All sensory modalities are intact and normal, including vision, hearing, and somatosensation.

* Coordination and Balance Examination: The patient is able to perform all activities of daily living without difficulty.

Neurological Examination (extremities):

* Right and left upper and lower extremity motor strength are 5/5.

* Sensation in the upper and lower extremities is intact and normal.

* Reflexes are 2+ (moderately increased) in all extremities.

Neurological Examination (special senses):

* Ophthalmoscopic examination reveals no abnormalities.

* Fundoscopic examination reveals no abnormalities.

* Auditory system examination reveals no abnormalities.

* Vestibular system examination reveals no abnormalities.

Assessment and Plan:

The patient is a 34-year-old male with a history of depression and temporal lobe epilepsy who has been participating in a randomized controlled trial (RCT) and has been taking the investigational drug ID X13737 for epilepsy over the past 3 months. The patient has had 27 seizures during this time, but reports no symptoms. The RCT is now complete, and the investigational drug will be discontinued.

The patient's neurological examination reveals no significant abnormalities. However, given the patient's history of temporal lobe epilepsy and the recent seizures during the RCT, it is important to continue close follow-up to monitor for any changes in seizure activity or other neurological symptoms.

The patient will be discontinued from the investigational drug and will be transitioned back to standard of care antiepileptic medications. The patient will be followed up in clinic every 3-6 months for the next year to monitor for any changes in seizure activity or other neurological symptoms. Additionally, the patient will be advised to continue all current medications and to avoid any potential triggers for seizures (e.g., alcohol, sleep deprivation).

The patient's mental health status will also be monitored closely, given his history of depression, and he will be referred to a mental health professional if necessary.

The patient has been informed of the results of the RCT and the plan for ongoing care, and he has been advised to contact the clinic immediately if he experiences any changes in seizure activity or other neurological symptoms.

----------------END OF PATIENT ENCOUNTER---------------

-------------- PATIENT ID 186 -- VISIT 1 -----------

Clinic Note: Visit #1

Date: 2022-11-22

Patient ID: 186

Identification:

Name: John Doe

Age: 72 years

Gender: Male

Contact Information:

Address: 123 Main St, Anytown, USA

Phone Number: 555-555-5555

Chief Complaint:

Patient presents with a history of dermatitis and temporal lobe epilepsy, currently experiencing an increase in seizure activity over the past two months. The patient has had six seizures during this time frame.

History of Present Illness (HPI):

The patient reports a worsening of his seizure activity over the past two months, with a total of six seizures occurring during this time frame. The patient's dermatitis has also been exacerbated, leading to increased discomfort and sleep disturbances. The patient is participating in a randomized controlled trial for the treatment of epilepsy and has not experienced any changes in medication other than the addition of the investigational drug, which will be taken for a period of three months.

Subjective:

The patient reports experiencing more frequent seizures over the past two months, with no noticeable triggers or warning signs. The patient's dermatitis has been persistent and exacerbated, leading to increased discomfort and sleep disturbances. The patient expresses concern about the potential side effects of the investigational drug and the long-term effects on his health.

Medications:

1. Levetiracetam (Keppra) - 2500mg orally BID

2. Lamotrigine (Lamictal) - 100mg orally QD

3. Investigational drug (name not disclosed) - 500mg orally BID for the past three months as part of a randomized controlled trial

Allergies:

The patient reports no known allergies.

Social History:

Married with two adult children, both living locally. The patient is a retired mechanical engineer and currently enjoys hiking and woodworking in his free time. The patient's spouse has been supportive of the patient's medical treatment and has accompanied him to all appointments.

Review of Systems:

The patient reports no significant changes in his review of systems, including his sleep patterns, appetite, or mood. The patient denies any recent travel or exposure to illness.

General Physical Examination:

The patient's vital signs are within normal limits. His general physical examination reveals no significant abnormalities, including no edema, lymphadenopathy, or skin rashes. The patient's neurological examination is significantly perturbed by the presence of seizures and dermatitis.

Comprehensive Neurologic Exam:

The patient's comprehensive neurologic examination reveals the following findings:

1. Cranial Nerves: The patient's cranial nerves are intact, with no signs of weakness or paresis.

2. Motor Exam: The patient's motor exam is significantly perturbed by the presence of seizures and dermatitis. There is no evidence of hemiparesis or other focal neurological deficits.

3. Sensory Exam: The patient's sensory exam is within normal limits, with no signs of sensory loss or abnormalities.

4. Coordination and Balance: The patient's coordination and balance are significantly perturbed due to the presence of seizures and dermatitis. The patient reports difficulty with walking and standing.

5. Reflexes: The patient's reflexes are within normal limits.

Labs:

The patient's laboratory results are as follows:

1. Complete Blood Count (CBC): WBC 4.5, Neutrophils 70%, Lymphocytes 20%, Monocytes 8%, Eosinophils 3%

2. Electrolyte Panel: Sodium 136, Potassium 4.2, Chloride 98, Carbon Dioxide 22

3. Liver Function Tests (LFTs): AST 25, ALT 30, Alkaline Phosphatase 125, Gamma-Glutamyl Transferase (GGT) 25

4. Renal Function Tests: Creatinine 1.2, Urinalysis: No significant abnormalities

Imaging:

The patient's imaging results are as follows:

1. Magnetic Resonance Imaging (MRI): The patient underwent an MRI of the brain, which revealed no significant abnormalities, including no evidence of structural brain lesions or tumors.

2. Electroencephalogram (EEG): The patient underwent an EEG, which revealed no significant abnormalities, including no evidence of seizure activity.

Assessment:

The patient's recent increase in seizure activity and persistent dermatitis are concerning and warrant further evaluation. The patient's participation in a randomized controlled trial for the treatment of epilepsy is also a significant consideration. Based on the patient's history, physical examination, laboratory results, and imaging findings, the following diagnoses are considered:

1. Temporal lobe epilepsy

2. Dermatitis

3. Randomized controlled trial-related adverse events (AEs)

Plan:

The patient will continue to participate in the randomized controlled trial, and no changes will be made to his current medication regimen other than the addition of the investigational drug for a period of three months. The patient's seizure activity will be closely monitored, and any significant changes will be reported to the study team immediately. The patient will also continue to receive dermatitis treatment as needed.

Signature:

[Your Signature]

Date: [2022-11-22]

----------------END OF PATIENT ENCOUNTER---------------

-------------- PATIENT ID 186 -- VISIT 2 -----------

---

Clinic Note #2 - Patient ID 186 - 2022-12-22

Chief Complaint:

The patient presents with a history of dermatitis and temporal lobe epilepsy, currently enrolled in a randomized controlled trial evaluating the investigational drug ID X13737 for the treatment of epilepsy. The patient has experienced 17 seizures over the past 3 months while taking the investigational drug. Additionally, the patient reports experiencing vertigo, falling, and headaches.

History of Present Illness:

The patient was initially diagnosed with dermatitis and temporal lobe epilepsy at age 65. He has been managed with anticonvulsant medications and topical creams for his skin condition. The patient was recently enrolled in a clinical trial evaluating the investigational drug ID X13737 for the treatment of epilepsy. The patient reports that over the past 3 months, he has experienced an increase in seizure activity, with 17 seizures occurring during this time. Additionally, the patient reports experiencing vertigo, falling, and headaches.

Review of Systems:

General: The patient appears well-nourished and well-hydrated. He is alert and oriented to person, place, and time.

Neurological: The patient exhibits a steady gait with no notable tremors or rigidity. His speech is clear and coherent. There is no evidence of facial weakness or abnormalities. His extraocular movements are full and without difficulty. His hearing is intact, and he has no complaints of ear fullness or pain. The patient reports no recent changes in his vision.

Musculoskeletal: The patient has no notable muscle weakness or atrophy. He has a normal range of motion in all joints without any signs of inflammation or redness.

Psychiatric: The patient appears to be cognitively intact and does not report any recent changes in his mood or mental status.

Social: The patient is married and lives with his spouse. He reports no significant social or environmental stressors.

Medications:

The patient is currently taking levetiracetam 500mg twice daily, lamotrigine 200mg once daily, and prednisone 1mg/kg once daily for his dermatitis and epilepsy. He has been advised to discontinue the investigational drug ID X13737 due to completion of the clinical trial.

Assessment:

The patient presents with a history of dermatitis and temporal lobe epilepsy, currently managed with anticonvulsant medications and topical creams. Over the past 3 months, the patient has experienced an increase in seizure activity while taking the investigational drug ID X13737 for the treatment of epilepsy. Additionally, the patient reports experiencing vertigo, falling, and headaches. The patient's neurological examination reveals no notable abnormalities, and his cognitive function appears intact.

Plan:

1. Discontinue the investigational drug ID X13737 due to completion of the clinical trial.

2. Continue current anticonvulsant medications and topical creams for dermatitis management.

3. Monitor the patient's seizure activity and adjust medications as needed.

4. Follow-up in 1 month to assess for any changes in seizure activity or adverse effects from medication discontinuation.

---

Note: This clinic note is a fictional example and does not reflect any real patient or study. It is intended to provide an illustration of a neurologist's clinic note style and content, and should not be used as a basis for actual clinical decision-making or patient care.

----------------END OF PATIENT ENCOUNTER---------------

-------------- PATIENT ID 187 -- VISIT 1 -----------

Patient ID: 187

Age: 41 years

History: Autism, temporal lobe epilepsy

Seizure History: 14 seizures in past 2 months

Current Medications: none (part of randomized controlled trial)

Adding Investigational Drug: Yes, for 3 months

Neurological Examination:

* Mental status: Alert and cooperative

* Head: Normal shape and proportions

* Eyes: Pupils equal and reactive to light, no nystagmus or gaze abnormalities

* Ears: No discharge or otorrhea

* Nose: No discharge or deformity

* Mouth: Normal size and shape, no tremors or anomalies

* Extremities: No weakness, wasting, or paresis

* Sensation: Intact

* Reflexes: Normal

Pertinent Positives:

* No changes in medications other than investigational drug

* History of autism and temporal lobe epilepsy

* Increased seizure activity in past 2 months

Assessment and Plan:

* Continue participating in randomized controlled trial

* Add investigational drug for 3 months to manage increased seizure activity

* Monitor for adverse effects or changes in seizure frequency/pattern

* Schedule follow-up visit in 1 month to assess response to treatment and make any necessary adjustments.

----------------END OF PATIENT ENCOUNTER---------------

-------------- PATIENT ID 187 -- VISIT 2 -----------

---

Patient ID: 187

Date: 2021-09-11

Identification:

Name: [Patient Name]

DOB: [Date of Birth]

Gender: Female

Highlights of the History of Present Illness (HPI):

* Autism spectrum disorder

* Temporal lobe epilepsy

* Participated in a randomized controlled trial for investigational drug ID X13737 for epilepsy

* Took the drug for 3 months with a reported 15 seizures during that time

* Experienced dizziness

Subjective:

* Patient reports no change in seizure frequency or severity since last visit

* Reports persistent dizziness, which is mild but bothersome

* No changes in medications or allergies to report

Medications:

* Levetiracetam 500mg bid

* Lamotrigine 200mg qd

* Valproic acid 500mg tid

Allergies: None

Social History:

* Married with two children

* No changes in living situation or social support since last visit

Review of Systems (ROS):

* No changes in sleep pattern, appetite, or energy level since last visit

* No recent illnesses or hospitalizations

General Physical Examination:

* Vital signs: T 98.6°F, HR 72 bpm, BP 120/80 mmHg, RR 14 bpm

* Weight: 58 kg (last visit: 57 kg)

* Height: 163 cm (last visit: 162 cm)

* Body Mass Index (BMI): 21.3 (last visit: 20.9)

Comprehensive Neurologic Examination:

* Cranial nerves: Intact

* Motor examination: Normal

* Sensory examination: Normal

* Reflexes: Normal

* Coordination and balance: Normal

Labs:

* Complete Blood Count (CBC) within normal limits

* Electrolyte panel within normal limits

* Liver function tests (LFTs) within normal limits

* Thyroid-stimulating hormone (TSH) level within normal limits

Imaging:

* No changes in magnetic resonance imaging (MRI) of the brain since last visit

Assessment:

* Autism spectrum disorder with co-morbid temporal lobe epilepsy

* Patient has experienced no significant changes in seizure frequency or severity since last visit

* Dizziness is a mild but bothersome symptom

* Discontinuation of investigational drug ID X13737 due to completion of the randomized controlled trial

Plan:

* Discontinue investigational drug ID X13737

* Monitor seizure frequency and severity closely

* Consider dose adjustments to current medications as needed

* Follow up in 3 months for re-evaluation of symptoms and medication adherence

Signature:

[Your Signature]

---

Note: This is just an example clinic note, and the content may vary based on the specific patient and their condition.

----------------END OF PATIENT ENCOUNTER---------------

-------------- PATIENT ID 188 -- VISIT 1 -----------

CLINIC NOTE #1 (2021-07-17)

Patient ID: 188

Name: [Mr./Mrs./Ms.] [Last Name]

Age: 94 years

Gender: Male

Chief Complaint: Seizures

History of Present Illness:

The patient, a 94-year-old male with a history of autism and temporal lobe epilepsy, has been experiencing an unusual increase in seizure activity over the past two months. He has had 58 seizures during this time frame, which is significantly higher than his usual frequency. The patient is currently enrolled in a randomized controlled trial investigating a new medication for the treatment of epilepsy.

Review of Systems:

The patient's medical history is significant for autism and temporal lobe epilepsy, as previously mentioned. He has a history of seizures dating back to his early 20s, with a previous diagnosis of juvenile myoclonic epilepsy. The patient has been treated with levetiracetam and sodium valproate for the past several years, with minimal side effects reported. He has no history of allergies or sensitivities to any medications.

Social History:

The patient resides in a long-term care facility and is accompanied by his primary caregiver during clinic visits. He is well-cared for and has no notable social or family history. The patient is unable to communicate effectively due to his autism diagnosis, but he appears to be content and cooperative during the clinic visit.

Medications:

The patient's current medication regimen includes levetiracetam 500 mg twice daily and sodium valproate 500 mg twice daily. He has been adherent to his medication regimen, with no reported missed doses or side effects.

Allergies:

The patient has no known allergies to any medications.

Labs:

Recent laboratory studies, including a complete blood count and electrolyte panel, have been unremarkable. The patient's most recent electroencephalogram (EEG) revealed a marked increase in spike wave discharges, consistent with his diagnosis of temporal lobe epilepsy.

Imaging:

The patient's most recent magnetic resonance imaging (MRI) scan of the brain revealed no acute changes or lesions. There is evidence of previous scar tissue in the temporal lobes, consistent with his history of seizures.

Assessment:

Given the patient's history of autism and temporal lobe epilepsy, as well as his recent increase in seizure activity, it is necessary to continue monitoring his condition closely. The patient's current medication regimen will be maintained, with the addition of the investigational drug for epilepsy. This trial medication will be taken for a period of three months, and the patient's response will be closely monitored.

Plan:

1. Continue the patient's current medication regimen, including levetiracetam 500 mg twice daily and sodium valproate 500 mg twice daily.

2. Add the investigational drug for epilepsy, to be taken for a period of three months.

3. Monitor the patient's seizure activity and adjust medications as needed.

4. Schedule follow-up appointments every two months to assess the patient's response to the trial medication and monitor for any adverse effects.

5. Encourage the patient to maintain a consistent sleep schedule, avoid alcohol and caffeine, and engage in regular physical activity to minimize seizure activity.

Signature:

[Your Name]

Date: 2021-07-17

Note: This clinic note is a sample and should not be used as an actual clinical note. It is important to include all relevant information and follow proper clinical documentation guidelines when creating a clinic note for a patient.

----------------END OF PATIENT ENCOUNTER---------------

-------------- PATIENT ID 188 -- VISIT 2 -----------

Here is your clinic note:

Patient ID: 188

Date: 2021-08-17

Identification:

I am pleased to present the second visit of Patient ID 188, a 94-year-old male with an exceptional history of autism and temporal lobe epilepsy. This patient has been participating in a randomized controlled trial (RCT) investigating the efficacy and safety of the novel compound ID X13737 for treating intractable temporal lobe epilepsy.

History of Present Illness (HPI):

The patient has been adherent to the study protocol, having received ID X13737 daily for the past three months. During this time, he has experienced an unfortunate increase in seizure frequency, with 43 seizures documented over the past 90 days (a 38% increase compared to the baseline period).

Subjective:

At today's visit, the patient reported a pervasive sense of somnolence and lethargy, which he attributes to the investigational drug. He also complained of progressive memory loss and difficulty concentrating, both of which may be related to the study medication or underlying temporal lobe pathology.

Medications:

The patient has been compliant with the ID X13737 dosing regimen (250mg QD x 3 months). He has also been taking levetiracetam (500mg BID) and lacosamide (200mg BID) as adjunctive therapy for his refractory temporal lobe epilepsy.

Allergies:

The patient has no known allergies or sensitivities to any medications or substances.

Social History:

The patient resides in a long-term care facility and is well cared for by the staff. He has limited social interaction due to his advanced age and cognitive limitations. The patient's family members are active in his care, visiting regularly and communicating with the study team as needed.

Review of Systems (ROS):

The patient has a history of autism, which manifests as significant social and communication deficits. He also experiences recurrent temporal lobe seizures, which have been challenging to control despite prior trials of various anticonvulsants. The patient reports no other relevant medical or psychiatric history.

General Physical Examination:

On today's examination, the patient appeared well-nourished and well-groomed, with no acute distress or discomfort. His vital signs were within normal limits (BP 120/80 mmHg, HR 72 beats per minute, T 98.6°F). A comprehensive neurological examination was performed and revealed the following:

* Cranial nerves: The patient demonstrated a mild left-sided hemiparesis, with subtle weakness of the left arm and leg. There was no evidence of abnormal reflexes or sensory deficits.

* Motor examination: The patient exhibited a gradual decrease in muscle tone and power, particularly in the left upper limb. There were no signs of ataxia or dysmetria.

* Sensory examination: The patient showed normal sensation to light touch, pinprick, and temperature on both sides of the body. No sensory deficits were identified.

Comprehensive Neurological Exam:

The patient underwent a comprehensive neurological exam, including assessment of cognitive function, mood, and behavior. The following observations were made:

* Cognition: The patient demonstrated significant cognitive impairment, with difficulty remembering recent events and learning new information. His performance on the Mini-Mental State Examination (MMSE) was 18/30, indicating moderate dementia.

* Mood: The patient reported a flat affect and displayed limited emotional responsiveness during the examination. There were no signs of overt mood disorder or psychosis.

* Behavior: The patient exhibited significant difficulty with social interaction and communication, consistent with his diagnosed autism spectrum disorder. He demonstrated no aggressive or violent behavior during the visit.

Labs:

The patient's recent laboratory results are as follows:

* Complete Blood Count (CBC): WBC 4.5 x 109/L, NEUT 3.2 x 109/L, LYMP 1.1 x 109/L, MONO 6.8 x 109/L, EOSIN 11.5%.

* Blood Chemistry: Sodium 138 mmol/L, Potassium 4.2 mmol/L, Chloride 102 mmol/L, CO2 22 mmol/L, Glucose 150 mg/dL, BUN 20 mg/dL, Creatinine 1.5 mg/dL.

* Urinalysis: Clear yellow urine with no visible blood or pyuria.

Imaging:

The patient has undergone serial magnetic resonance imaging (MRI) scans to assess for changes in brain structure and function over time. The most recent MRI scan revealed mild atrophy of the temporal lobes, consistent with the patient's history of autism and temporal lobe epilepsy.

Assessment:

In light of the patient's recent increase in seizure frequency while taking ID X13737, it is my professional opinion that discontinuing this medication is warranted at this time. Additionally, the patient's cognitive impairment and flat affect may be related to the study medication or underlying temporal lobe pathology, and further evaluation and management are necessary to address these issues. The patient's autism spectrum disorder and refractory temporal lobe epilepsy pose ongoing challenges for optimal clinical management.

Plan:

Given the patient's recent seizure increase while taking ID X13737, we will discontinue this medication immediately. The patient will be transitioned to standard therapy for refractory temporal lobe epilepsy, consisting of levetiracetam and lacosamide. We will closely monitor the patient's seizure frequency and adjust the dosages of these medications as needed. The patient will undergo regular follow-up evaluations to assess for changes in cognitive function, mood, and behavior.

Signature:

[Your name and credentials]

Date: 2021-08-17

----------------END OF PATIENT ENCOUNTER---------------

-------------- PATIENT ID 189 -- VISIT 1 -----------

Patient ID: 189

Date of Visit: 2022-04-21

Identification:

Age: 47 years

Gender: Female

History of Present Illness:

The patient, a 47-year-old female with a history of bipolar disorder, ADHD, and occipital lobe epilepsy, presented to the clinic today with a complaint of increased seizure activity in the past two months. She has experienced 60 seizures during this time, which is a significant increase from her usual frequency of 1-2 seizures per month. The patient is currently participating in a randomized controlled trial (RCT) and has been adherent to her medications as prescribed.

Highlights of the Patient's History:

* Bipolar disorder, ADHD, and occipital lobe epilepsy

* Previous history of 1-2 seizures per month

* Currently participating in an RCT

* No change in medications will be made other than adding the investigational drug for epilepsy for the next 3 months

Subjective:

The patient reported that she has been experiencing more frequent seizures over the past two months, with a total of 60 seizures during this time. She also noted that her seizures have been more intense and prolonged than usual. The patient mentioned that she has been able to manage her symptoms with her current medications, but is concerned about the increased frequency and severity of her seizures.

Medications:

* Lamotrigine (Lamictal) 200mg orally BID

* Valproic acid (Depakote) 500mg orally TID

* Levetiracetam (Keppra) 1000mg orally QID

* Topiramate (Topamax) 200mg orally BID

Allergies:

* None reported

Social History:

* Married with two children

* Works as a teacher

* Denies smoking and alcohol consumption

Review of Systems:

* General: No fever, chills, or weight loss

* Neurological: Increased seizure activity, more intense and prolonged than usual

* Cardiovascular: No history of cardiac disease

* Respiratory: No history of respiratory disease

* Gastrointestinal: No history of gastrointestinal disease

* Musculoskeletal: No history of musculoskeletal pain or injury

General Physical Examination:

* Vital signs: BP 120/80, HR 80, T 98.6

* Head: Normocephalic, normal scalp hair

* Eyes: No dilated pupils, no visual field defects

* Ears: No discharge, no hearing loss

* Nose: No discharge, no nasal congestion

* Throat: Normal throat without any exudate

* Chest: Clear to auscultation, no murmurs or rubs

* Abdomen: Normal contours, no visible masses

* Extremities: No clubbing or cyanosis of the fingers and toes

Comprehensive Neurologic Examination:

* Cranial nerves: Intact, with no signs of weakness or paresis

* Motor examination: No signs of weakness or atrophy

* Sensory examination: No sensory deficits

* Reflexes: 2+ (moderate)

* Gait and station: Normal gait and station, with no signs of imbalance or instability

Labs:

* Complete Blood Count (CBC): Within normal limits

* Serum electrolytes: Within normal limits

* Liver function tests (LFTs): Within normal limits

* Thyroid-stimulating hormone (TSH): Within normal limits

Imaging:

* Magnetic Resonance Imaging (MRI) of the brain: No evidence of acute or chronic changes

* Computerized Tomography (CT) scan of the head: No evidence of acute or chronic changes

Assessment:

The patient presents with an increase in seizure activity over the past two months, which is consistent with the recent changes in her medications. The patient's neurological examination reveals no new deficits or abnormalities. The patient's medical history and current medications do not suggest any other possible causes for her increased seizure activity.

Plan:

1. Continue the patient's current medication regimen, with no changes other than adding the investigational drug for epilepsy for the next 3 months.

2. Monitor the patient's seizure activity closely and adjust medications as needed to control her seizures.

3. Re-evaluate the patient's medication regimen at the end of the 3-month trial period to determine if any changes are necessary.

Signature:

[Your name and title]

Date: 2022-04-21

----------------END OF PATIENT ENCOUNTER---------------

-------------- PATIENT ID 189 -- VISIT 2 -----------

CLINIC NOTE #2

DATE: 2022-05-21

PATIENT ID: 189

AGE: 47 years old

SEX: Female

CHIEF COMPLAINT:

Patient is a 47-year-old female with a history of bipolar disorder, ADHD, and occipital lobe epilepsy. She is currently participating in a randomized controlled trial evaluating the safety and efficacy of investigational drug ID X13737 for the treatment of epilepsy.

MEDICAL HISTORY:

Patient has a history of bipolar disorder, ADHD, and occipital lobe epilepsy. She has been experiencing seizures over the past 3 months while taking investigational drug ID X13737 as part of the clinical trial. The patient reported no symptoms.

REASON FOR VISIT:

The patient is here for a follow-up visit to assess her condition after completing the clinical trial. The investigational drug will be discontinued now that the study is complete.

PHYSICAL EXAMINATION:

General:

* Alert and oriented x3

* No significant changes in weight or appearance since last visit

Head:

* Normocephalic, normotriglyceridemic

* No focal deficits or abnormalities detected

Eyes:

* No visual field defects or abnormalities detected

Ears:

* No hearing loss or abnormalities detected

Mouth:

* Normocephalic, normotriglyceridemic

* No signs of oral lesions or abnormalities detected

Neck:

* No cervical spine tenderness or abnormalities detected

Extremities:

* No significant tremors or rigidity detected

Sensory:

* No sensory deficits or abnormalities detected

Cerebellar:

* No cerebellar ataxia or abnormalities detected

Motor:

* No motor deficits or abnormalities detected

Assessment and Plan:

Based on the patient's history and physical examination, it is determined that the investigational drug ID X13737 has been effective in reducing seizure frequency over the past 3 months. However, since the study is now complete and the drug will be discontinued, the patient will need to be closely monitored for any changes in her condition.

The patient will follow up with the neurology clinic every 2 months for the next 6 months to assess her seizure frequency and overall health. If the patient experiences any changes in her symptoms or has any concerns, she should contact the clinic immediately.

In addition, the patient will continue her current medications for bipolar disorder and ADHD as prescribed by her primary care physician. She will also be referred to a specialist for ongoing management of her occipital lobe epilepsy.

The patient understands the risks and benefits of the investigational drug and has been informed that it is not approved by the FDA for use in epilepsy. She has also been informed that there are no guarantees that the drug will be effective in reducing seizures or that it will be safe for long-term use.

The patient has signed an informed consent form and has been enrolled in the clinical trial. The patient's participation in the study is voluntary, and she may withdraw her consent at any time without penalty.

----------------END OF PATIENT ENCOUNTER---------------

-------------- PATIENT ID 200 -- VISIT 1 -----------

Patient ID: 200

Date: 2022-05-24

Visit #1

Identification:

Name: [Patient Name]

Age: 38 years

Gender: Female

ID Number: [Patient ID number]

HPI (History of Present Illness):

Chief Complaint: Recurrent seizures

Medical History: Frontal lobe epilepsy, previous history of 28 seizures in the past 2 months.

Current Medications: Levetiracetam, Lamotrigine, Valproate

Allergies: None known

Social History: Married, no children, works as an accountant, lives with spouse.

Review of Systems:

General: No fever, fatigue, or weight loss.

Neurological: Recurrent seizures, no focal weakness or sensory deficits.

Psychiatric: No history of psychiatric disorders.

Medication Adherence: Fair, patient reports missing doses occasionally.

Subjective:

Patient reported recent increase in seizure frequency, experiencing 2-3 seizures per week.

Patient reports difficulty with concentration and memory.

Patient denies any changes in sleep pattern or appetite.

Objective:

General Physical Examination:

Vital signs: Blood pressure 120/80 mmHg, heart rate 70 bpm, temperature 98.6 degrees F.

Head and Neck: Normocephalic, normochromatic. No focal swelling or tenderness.

Extremities: Proximal and distal muscle strength 5/5, no atrophy or weakness.

Sensory: Full range of sensation in all extremities.

Comprehensive Neurologic Examination:

Cranial Nerves: Intact, including visual fields and funduscopic examination.

Motor: Normal strength and tone in all extremities.

Sensory: Full range of sensation in all extremities.

Reflexes: Normocaloric, normoaxic.

Labs:

Complete Blood Count (CBC): Within normal limits.

Electrolyte Panel: Within normal limits.

Imaging:

Computed Tomography (CT) scan of the head: No evidence of acute brain injury or lesions.

Magnetic Resonance Imaging (MRI) of the brain: No evidence of structural abnormalities.

Assessment:

Seizure frequency has increased in the past 2 months, with recent seizures occurring 2-3 times per week.

Patient reports difficulty with concentration and memory.

No changes in sleep pattern or appetite.

Plan:

Continue current medications.

Add investigational drug for epilepsy (IDEE) for a period of 3 months.

Monitor seizure frequency and adjust medication regimen as needed.

Follow-up visit in 2 weeks to assess progress.

Signature: [Your Signature]

Note: This clinic note is fictional and should not be used for actual patient care. It is meant for educational purposes only.

----------------END OF PATIENT ENCOUNTER---------------

-------------- PATIENT ID 200 -- VISIT 2 -----------

Patient ID: 200, Female, Age 38

History: Frontal lobe epilepsy, randomized controlled trial (ID X13737)

Neurologic Exam:

* Mild dizziness

Pertinent Positives:

* 19 seizures over past 3 months while taking investigational drug ID X13737

Assessment and Plan:

* Discontinue investigational drug ID X13737 due to completion of study

* Monitor for seizure activity and dizziness in follow-up visits

* Consider alternative medications or therapies as needed.

----------------END OF PATIENT ENCOUNTER---------------

-------------- PATIENT ID 201 -- VISIT 1 -----------

---

Clinic Note #1 - Patient ID: 201, Date: 2021-12-18

Identification:

Name: John Doe

Date of Birth: 01/01/1930

Patient ID: 201

History of Present Illness (HPI):

Mr. Doe, a 92-year-old male, presents with a history of anxiety, bipolar disorder, and epilepsy. He has been experiencing an increased frequency of seizures over the past two months, with a total of 24 seizures. He is currently participating in a randomized controlled trial (RCT) for the treatment of his epilepsy.

Subjective:

Mr. Doe reports that he has been experiencing increased anxiety and depression over the past few months, which he believes may be related to the recent increase in seizure activity. He also mentions that he has been having difficulty sleeping due to frequent nightmares. He denies any recent changes in his medications or any new medical conditions.

Medications:

Current Medications:

* Lamotrigine (LTG) 200mg po QD

* Valproate (VPA) 500mg po TID

* Levetiracetam (LEV) 1000mg po BID

Allergies:

* None reported

Social History:

Mr. Doe is a retired engineer and lives with his wife of 65 years. He has two adult children who live out of state. He denies any history of substance abuse or smoking. He reports that he exercises regularly, including daily walks and light weightlifting. He also mentions that he enjoys reading and spending time with his family.

Review of Systems (ROS):

Cardiovascular: None reported

Respiratory: None reported

Gastrointestinal: None reported

Genitourinary: None reported

Musculoskeletal: None reported

Neurological: Frequent seizures (24 over the past two months), increased anxiety and depression, nightmares

Psychiatric: Anxiety, bipolar disorder

General Physical Examination:

Mr. Doe appears well-nourished and in no acute distress. His vital signs are as follows:

* Blood pressure: 120/80 mmHg

* Heart rate: 70 beats per minute

* Temperature: 98.6°F (37°C)

* Weight: 65 kg

Comprehensive Neurologic Examination:

Mr. Doe's neurological examination reveals the following:

* Mental status: Alert and oriented to person, place, and time

* Speech: Clear and understandable

* Memory: Intact

* Cranial nerves: Normal

* Motor function: Normal

* Sensory function: Normal

* Reflexes: 2+ (brisk)

Labs:

* Complete Blood Count (CBC): To be performed in the near future

* Electrolyte Panel (EVP): To be performed in the near future

* Liver Function Tests (LFTs): To be performed in the near future

Imaging:

* Computed Tomography (CT) scan of the head: To be performed in the near future

Assessment:

Mr. Doe presents with a history of anxiety, bipolar disorder, and epilepsy, and has been experiencing an increased frequency of seizures over the past two months. He is currently participating in a RCT for the treatment of his epilepsy. His neurological examination reveals no significant abnormalities. Based on his history and current symptoms, it is possible that Mr. Doe may be experiencing a worsening of his epilepsy or a side effect of the investigational drug being used in the RCT. It is important to monitor his seizure activity and medication adherence closely over the next few months.

Plan:

1. Continue current medications as prescribed, including LTG, VPA, and LEV.

2. Monitor Mr. Doe's seizure activity and medication adherence closely over the next few months.

3. Perform CBC, EVP, and LFTs in the near future to assess for any changes in his laboratory values.

4. Perform a CT scan of the head in the near future to assess for any changes in his imaging studies.

5. Continue to participate in the RCT as planned.

Signature:

---

[Your Name]

Neurologist

----------------END OF PATIENT ENCOUNTER---------------

-------------- PATIENT ID 201 -- VISIT 2 -----------

Visit #2 - 2022-01-18

Patient ID: 201

Age: 92 years old

History: Anxiety, Bipolar Disorder, Epilepsy (randomized controlled trial)

Investigational Drug: ID X13737 (discontinued)

Neurological Examination:

* Mental Status: Alert and oriented

* Speech: Normal

* Motor Exam: Normal

* Sensory Exam: Normal

* Cranial Nerves: Normal

* Reflexes: 2+ (normal)

* Temperature: Normal

Pertinent Positives:

* No symptoms reported

* 35 seizures in past 3 months while taking investigational drug ID X13737

Assessment and Plan:

* Discontinue investigational drug ID X13737

* Monitor for seizure activity and adverse effects

* Follow up visit in 2 weeks to assess for any changes in symptoms or seizure activity.

----------------END OF PATIENT ENCOUNTER---------------

-------------- PATIENT ID 202 -- VISIT 1 -----------

Clinic Note #1

Patient ID: 202

Date: 2021-04-06

Identification:

Patient Name: Jane Doe

DOB: 04/01/1950

Gender: Female

Highlights of Patient's History:

* Seasonal allergies

* Depression

* Schizophrenia

* Multifocal epilepsy (5 seizures in the past 2 months)

Patient's Chief Complaint (CC): Seizures

Subjective:

* Patient reports experiencing 5 seizures over the past two months, with a frequency of approximately every two weeks.

* Patient denies any recent changes in medications or significant life events that may be contributing to the increased seizure activity.

* Patient expresses concern about the potential impact of the seizures on her daily life and relationships.

Medications:

* Levetiracetam (Keppra) 500mg twice daily

* Lamotrigine (Lamictal) 200mg once daily

* Valproic acid (Depakote) 500mg twice daily

* Investigational drug for epilepsy (randomized controlled trial)

Allergies:

* None known

Social History:

* Married with adult children

* Retired from her job as a teacher

* Active in local community theater

Review of Systems:

* No recent illnesses or injuries

* No significant changes in sleep, appetite, or mood

* No history of substance abuse

General Physical Examination:

* Vital signs within normal limits

* Head and neck normal

* Extremities without any deformity or swelling

* Abdominal examination reveals soft, normoactive bowel sounds

* Neurological examination (see below)

Comprehensive Neurologic Exam:

* Mental status: Alert and oriented to person, place, and time. No evidence of cognitive decline or psychosis.

* Cranial nerves: Intact, including visual and facial movements.

* Motor examination: Normal strength and tone in all extremities. No evidence of ataxia or tremors.

* Sensory examination: Intact sensation to light touch, pain, and temperature over all body regions.

* Coordination and balance: Normal, with no evidence of ataxia or dysmetria.

Labs:

* Complete Blood Count (CBC): Within normal limits

* Electrolyte panel: Within normal limits

* Renal function tests (RFTs): Within normal limits

* Thyroid-stimulating hormone (TSH): 4.5 mIU/L (normal 0.4-4.0 mIU/L)

Imaging:

* Computed Tomography (CT) scan of the brain without contrast: No evidence of acute bleeding or ischemia. Mild diffuse cerebral atrophy noted.

* Magnetic Resonance Imaging (MRI) of the brain with and without contrast: No evidence of acute lesions or abnormalities. Mild diffuse cerebral atrophy and mild degenerative changes in the spinal cord noted.

Assessment:

* Patient presents with a history of increased seizure activity over the past two months, with no recent changes in medications or significant life events that may be contributing to the increased seizure activity.

* Mild diffuse cerebral atrophy and mild degenerative changes in the spinal cord are noted on imaging studies.

* Patient is participating in a randomized controlled trial for the treatment of multifocal epilepsy, with no changes in medications other than the addition of the investigational drug.

Plan:

* Continue current medications (Levetiracetam, Lamotrigine, Valproic acid) as prescribed.

* Add the investigational drug for epilepsy to the patient's regimen for a period of 3 months.

* Monitor the patient closely for any changes in seizure activity or adverse effects of the medications.

* Schedule follow-up appointments every two months to assess the patient's condition and adjust the treatment plan as needed.

Signature:

Jane Doe, MD

Note: This clinic note is for a fictional patient and is not based on any real patient or medical condition. It is intended solely for educational purposes and should not be used to guide actual medical decision-making.

----------------END OF PATIENT ENCOUNTER---------------

-------------- PATIENT ID 202 -- VISIT 2 -----------

Patient ID: 202

Date: 2021-05-06

Identification:

* Name: Mary Johnson

* Date of Birth: 03/22/1949

* Sex: Female

* Height: 168 cm

* Weight: 65 kg

* Hair: Gray

* Eyes: Brown

History of Present Illness (HPI):

* The patient presents with a history of seasonal allergies, depression, schizophrenia, and multifocal epilepsy.

* She has been participating in a randomized controlled trial for the investigational drug ID X13737 for her epilepsy over the past 3 months.

* The patient reported no symptoms during this time, but has experienced three seizures.

Subjective:

* The patient reports feeling well and states that she has not experienced any symptoms since her last visit.

* She mentions that she has been taking the investigational drug ID X13737 as prescribed and has noticed no changes in her condition.

* She reports that she has been adherent to her medication regimen, including the investigational drug.

Medications:

* ID X13737 (Investigational drug for epilepsy): The patient has been taking this medication for the past 3 months as part of a randomized controlled trial.

* Levetiracetam (anticonvulsant): The patient has been taking this medication for her multifocal epilepsy.

* Venlafaxine (antidepressant): The patient has been taking this medication for her depression.

* Quetiapine (antipsychotic): The patient has been taking this medication for her schizophrenia.

Allergies:

* None reported.

Social History:

* Married with two adult children and four grandchildren.

* Retired nurse practitioner.

* Enjoys gardening, reading, and spending time with family.

* Denies smoking and alcohol use.

Review of Systems (ROS):

* General: No complaints of fever, chills, or fatigue.

* Neurological: No changes in her seizure activity since her last visit.

* Musculoskeletal: No joint pain or weakness.

* Cardiovascular: Denies any recent cardiovascular symptoms.

* Respiratory: No shortness of breath or cough.

General Physical Examination (GPE):

* Vital signs: Temperature 98.6°F, blood pressure 120/80 mmHg, heart rate 70 beats per minute, respiratory rate 14 breaths per minute.

* Head and neck: Normal head shape and symmetry, normal facial expressions and gaze. No focal deficits or abnormal movements.

* Extremities: No swelling or deformity of the extremities.

* Neurological: The patient is alert and oriented x3. No evidence of seizure activity or abnormal movement.

Comprehensive Neurologic Exam (CNE):

* Cranial nerves: Intact, including visual, auditory, and sensory functions.

* Motor examination: The patient has normal strength and tone in all extremities, with no focal deficits or abnormal movements.

* Sensory examination: The patient has normal sensation to light touch, pain, and temperature over all extremities.

* Reflexes: Normal reflexes are present in all extremities.

Laboratory Findings:

* Complete Blood Count (CBC): Within normal limits.

* Electrolyte panel: Within normal limits.

* Thyroid-stimulating hormone (TSH) level: 2.5 mIU/L (normal 0.4-4.0 mIU/L).

* Lipid profile: Within normal limits.

* Renal function tests: Within normal limits.

* Liver function tests: Within normal limits.

Imaging Findings:

* Magnetic Resonance Imaging (MRI) of the brain: No evidence of structural abnormalities or lesions.

* Computed Tomography (CT) scan of the head and neck: No evidence of abnormalities.

Assessment:

* The patient is a 72-year-old female with a history of seasonal allergies, depression, schizophrenia, and multifocal epilepsy.

* She has been participating in a randomized controlled trial for the investigational drug ID X13737 for her epilepsy over the past 3 months.

* The patient reported no symptoms during this time, but has experienced three seizures.

* The patient is adherent to her medication regimen and denies any recent cardiovascular symptoms.

* The patient's neurological examination is normal, with no evidence of seizure activity or abnormal movement.

* The patient's laboratory findings are within normal limits, and there is no evidence of structural abnormalities or lesions on imaging studies.

Plan:

* Discontinue the investigational drug ID X13737 now that the study is complete.

* Continue the patient's current medication regimen, including levetiracetam, venlafaxine, and quetiapine.

* Monitor the patient's seizure activity closely and adjust her medication regimen as needed.

* Schedule a follow-up appointment in 3 months to assess the patient's condition and adjust her medication regimen as needed.

Signature:

[Patient's signature]

[Date]

Note: This is a fictional patient and the information provided is for illustrative purposes only. It should not be used as a basis for actual patient care.

----------------END OF PATIENT ENCOUNTER---------------

-------------- PATIENT ID 203 -- VISIT 1 -----------

Patient ID: 203

Visit #1: 2021-08-11

History:

* Bipolar disorder

* Parietal lobe epilepsy

* 15 seizures in past 2 months

* Randomized controlled trial (no change in medications except for investigational drug for epilepsy)

Neurological Examination:

* Mental status: Alert and oriented

* Head: Normal head circumference, normocephalic

* Eyes: No abnormalities

* Visual fields: Full

* Extremities: Normal tone, no atrophy or weakness

* Sensation: Intact

* Reflexes: 2+ (moderate)

* Muscle strength: 5/5 (normal)

* Gait: Normal

Pertinent Positives:

* No change in medications except for investigational drug for epilepsy

* History of bipolar disorder and parietal lobe epilepsy

* Recent increase in seizures (15 in past 2 months)

Assessment and Plan:

* Continue randomized controlled trial with no change in medications except for investigational drug for epilepsy

* Monitor seizure activity and adjust plan as needed

* Follow up in 3 months for re-evaluation.

----------------END OF PATIENT ENCOUNTER---------------

-------------- PATIENT ID 203 -- VISIT 2 -----------

Clinic Note: Visit #2 - 2021-09-11

Patient ID: 203

Age: 56 years

Gender: Male

Chief Complaint: Bipolar disorder and parietal lobe epilepsy

History of Present Illness: The patient is a 56-year-old male with a history of bipolar disorder and parietal lobe epilepsy. He has been participating in a randomized controlled trial evaluating the investigational drug ID X13737 for epilepsy over the past 3 months. During this time, he has experienced 64 seizures, but reports no other symptoms. The study is now complete, and the investigational drug will be discontinued.

Identification:

* Name: [Patient's Name]

* Date of Birth: [Date of Birth]

* Contact Information: [Contact Information]

Highlighted Patient Information (HPI):

* Patient has a history of bipolar disorder and parietal lobe epilepsy

* Patient has been participating in a randomized controlled trial evaluating the investigational drug ID X13737 for epilepsy over the past 3 months

* Patient has experienced 64 seizures during this time, but reports no other symptoms

Subjective:

* The patient reports no subjective complaints at this time

Medications:

* ID X13737 (investigational drug for epilepsy) - discontinued as of today's visit due to completion of the study

* Lamotrigine (mood stabilizer) - ongoing

* Levetiracetam (anticonvulsant) - ongoing

Allergies:

* None reported

Social History:

* Marital status: Married

* Occupation: Retired

* Education level: College-educated

* Living situation: Patient lives with his wife in a private residence

Review of Systems:

* General: No complaints other than seizures (see above)

* Neurological: No complaints other than seizures (see above)

* Psychiatric: None reported

General Physical Examination:

* Vital signs:

+ Temperature: 98.6°F (37°C)

+ Blood pressure: 120/80 mmHg

+ Heart rate: 70 beats per minute

+ Respiratory rate: 14 breaths per minute

* Head and neck: No abnormalities noted

* Extremities: No abnormalities noted

* Abdomen: No abnormalities noted

* Neurological: No abnormalities noted other than those related to the patient's history of parietal lobe epilepsy (see below)

Comprehensive Neurologic Examination:

* Mental status: The patient appears to be alert and oriented to person, place, and time. He is cooperative and responds appropriately to questions and instructions.

* Cranial nerves: No abnormalities noted

* Motor examination: The patient has no significant muscle weakness or atrophy. His muscle tone is within normal limits. There is no evidence of tremors, rigidity, or other abnormal movements.

* Sensory examination: No significant sensory deficits or abnormalities noted. The patient responds appropriately to touch and pinprick sensation in all extremities.

* Coordination and balance: The patient has no significant difficulties with coordination or balance. He is able to walk normally and maintain his balance without difficulty.

Labs:

* Complete blood count (CBC): Within normal limits

* Electrolyte panel: Within normal limits

* Liver function tests (LFTs): Within normal limits

* Thyroid-stimulating hormone (TSH): Within normal limits

Imaging:

* Magnetic resonance imaging (MRI) of the brain: No abnormalities noted other than those related to the patient's history of parietal lobe epilepsy (see below)

Assessment:

* The patient has a history of bipolar disorder and parietal lobe epilepsy, and has been participating in a randomized controlled trial evaluating the investigational drug ID X13737 for epilepsy over the past 3 months. During this time, he has experienced 64 seizures, but reports no other symptoms. The study is now complete, and the investigational drug will be discontinued.

* The patient's neurological examination is within normal limits, with the exception of those findings related to his history of parietal lobe epilepsy (see below).

* The patient has no other symptoms or complaints at this time.

Plan:

* Discontinue the investigational drug ID X13737 as of today's visit, due to completion of the study.

* Continue the patient's current medications for bipolar disorder and epilepsy.

* Monitor the patient's seizure activity and adjust medications as needed to control seizures.

* Schedule follow-up appointments as necessary to monitor the patient's condition and adjust treatment as needed.

Signature:

[Patient's Signature]

[Date]

----------------END OF PATIENT ENCOUNTER---------------

-------------- PATIENT ID 204 -- VISIT 1 -----------

Patient ID: 204, Female, 78 years old

History: Asthma, seasonal allergies, parietal lobe epilepsy (5 seizures in past 2 months)

Randomized controlled trial (no change in medications except for investigational drug for epilepsy)

Neurological Examination:

* Mild weakness in right arm and leg

* Slight diplopia

* Normal sensation in all extremities

* Normal coordination and balance

Pertinent Positives:

* Seizures (5 in past 2 months)

* No change in medications except for investigational drug for epilepsy

Assessment:

* Continue randomized controlled trial with no changes to medications except for investigational drug for epilepsy

* Monitor seizure activity and adjust plan as needed

Plan:

* Add investigational drug for epilepsy for 3 months

* Follow-up appointment in 2 weeks for monitoring of seizure activity and adjustment of plan as needed.

----------------END OF PATIENT ENCOUNTER---------------

-------------- PATIENT ID 204 -- VISIT 2 -----------

---

CLINIC NOTE VISIT #2

Patient ID: 204

Date: 2021-09-03

Age: 78 years old

Gender: Female

Chief Complaint: None

History of Present Illness:

* History of asthma and seasonal allergies

* Parietal lobe epilepsy

* Currently enrolled in a randomized controlled trial for investigational drug ID X13737 for epilepsy (drug discontinued as study is complete)

* Patient has experienced 8 seizures over the past 3 months while taking the investigational drug

Review of Systems:

* No symptoms reported

Neurological Examination:

* Mental status: Alert and oriented x3

* Speech: Normal

* Motor function: Normal

* Sensation: Normal

* Reflexes: Normal

* Cranial nerves: Normal

* Extremities: Normal

* Gait: Normal

Assessment and Plan:

* The patient has completed the randomized controlled trial for investigational drug ID X13737 and has experienced 8 seizures during the study period.

* Given the patient's history of parietal lobe epilepsy and the recent seizure activity, it is recommended that the patient continue to be monitored closely for any changes in their seizure frequency or severity.

* The investigational drug will be discontinued as the study is complete.

* No changes to the patient's current medications are recommended at this time.

* Patient will be scheduled for a follow-up visit in 3 months to monitor for any changes in their seizure activity.

---

Note: This clinic note is written in the style of a neurologist and includes a complete neurologic examination, a brief assessment, and a plan for future care. The patient is a 78-year-old female with a history of asthma, seasonal allergies, and parietal lobe epilepsy, who has been participating in a randomized controlled trial for investigational drug ID X13737. The patient has experienced 8 seizures over the past 3 months while taking the drug, but has no other symptoms. The assessment and plan are based on the patient's history and recent clinical findings.

----------------END OF PATIENT ENCOUNTER---------------

-------------- PATIENT ID 205 -- VISIT 1 -----------

Clinic Note #1

Date: 2023-02-23

Patient ID: 205

Age: 18 years

Chief Complaint: Recent increase in seizure frequency

History of Present Illness:

The patient is an 18-year-old male with a history of parietal lobe epilepsy. He has experienced 70 seizures over the past 2 months, with a significant increase in frequency compared to previous years. The patient is currently enrolled in a randomized controlled trial (RCT) for the treatment of his epilepsy.

Review of Systems:

The patient reports no recent changes in medications or other health issues. He denies any new symptoms or side effects related to the RCT.

Physical Examination:

General: Alert and cooperative, with normal speech and language.

Neurological:

* Mental status: Normal cognitive function, with no evidence of impairment in attention, memory, or executive functions.

* Cranial nerves: Intact, with no abnormalities detected in visual, auditory, or olfactory functions.

* Motor examination: No abnormalities detected in strength, tone, or coordination.

* Sensory examination: Intact sensation to light touch, pinprick, and temperature on all extremities.

* Reflexes: Normocephalic and normochromic.

* Coordination and balance: Normal.

Assessment and Plan:

Based on the patient's history of parietal lobe epilepsy and recent increase in seizure frequency, it is necessary to continue monitoring the patient's condition closely. As the patient is participating in an RCT, no changes will be made to his medications at this time, except for the addition of the investigational drug for epilepsy. The patient will take this medication for a period of 3 months, and his seizure frequency and other symptoms will be closely monitored during this time.

The patient will be seen again in 1 month for a follow-up appointment to assess his condition and review any changes in his symptoms or seizure frequency. In the meantime, the patient should continue to keep a detailed seizure log to track any changes in his seizure activity. Additionally, the patient should be aware of any potential side effects of the investigational drug and report them immediately to his healthcare provider.

Order:

Continue current medications for epilepsy, with the addition of the investigational drug for a period of 3 months.

Follow-up appointment in 1 month to assess patient's condition and review any changes in symptoms or seizure frequency.

Discharge instructions:

* Continue to keep a detailed seizure log.

* Be aware of potential side effects of the investigational drug and report them immediately to your healthcare provider.

* Follow up with your healthcare provider in 1 month for a follow-up appointment.

----------------END OF PATIENT ENCOUNTER---------------

-------------- PATIENT ID 205 -- VISIT 2 -----------

Clinic Note #2 - 2023-03-23

Patient ID: 205

Age: 18 years old

Chief Complaint: Fatigue, upper respiratory infection

History of Present Illness:

The patient is an 18-year-old male with a history of parietal lobe epilepsy. He has been participating in a randomized controlled trial for the investigational drug ID X13737 for the treatment of his epilepsy. The patient has been taking the medication for the past 3 months and has experienced 47 seizures during this time. He now presents with fatigue and upper respiratory infection.

Review of Systems:

The patient reports no changes in his seizure activity or frequency since his last visit. He does report feeling more tired than usual and has had a recent upper respiratory infection.

Physical Examination:

General: The patient appears well-nourished and well-developed, with no signs of distress or discomfort.

Neurological:

* Mental Status: The patient's cognitive function is intact, with normal memory, attention, and concentration.

* Cranial Nerves: All cranial nerves are grossly normal.

* Motor Examination: The patient has full range of motion in all extremities, with no signs of weakness or atrophy.

* Sensory Examination: There is no sensory loss or abnormality noted.

* Reflexes: All reflexes are normal.

* Gait and Coordination: The patient walks normally, with no signs of abnormal gait or coordination.

Assessment and Plan:

Assessment:

* The patient's fatigue and upper respiratory infection are likely related to the investigational drug ID X13737 and will be monitored closely.

* The patient's epilepsy remains stable, with no changes in seizure frequency or severity.

Plan:

* Discontinue the investigational drug ID X13737 now that the study is complete.

* Monitor the patient's fatigue and upper respiratory infection closely and initiate appropriate treatment as needed.

* Schedule a follow-up visit in 2 weeks to assess the patient's condition and monitor for any changes in his epilepsy.

* The patient will be advised to continue his current medications for his epilepsy and to report any changes in his symptoms or seizure activity promptly.

----------------END OF PATIENT ENCOUNTER---------------

-------------- PATIENT ID 206 -- VISIT 1 -----------

Here's the clinic note for Patient ID 206:

Patient Info:

* Age: 81 years old

* Female

* History of diabetes, schizophrenia, asthma, and epilepsy

* 2 seizures in past 2 months

Randomized Controlled Trial:

* Investigational drug for epilepsy will be added for 3 months

* No changes to existing medications

Neurological Examination:

* Head: normocephalic, atraumatic

* Eyes: normal visual acuity, no abnormalities noted

* Ears: normal hearing, no discharge or infection

* Mouth: normal oral hygiene, no lesions or discoloration

* Neck: normal range of motion, no swelling or masses

* Extremities: normal strength and tone, no deformities or lesions

* Sensory: normal sensation to light touch, pinprick, and temperature

* Motor: normal movement, no weakness or ataxia

Pertinent Positives:

* Seizures in past 2 months

* History of diabetes, schizophrenia, asthma, and epilepsy

Assessment and Plan:

* Monitor seizure activity and adjust medications as needed

* Continue existing medications for diabetes, schizophrenia, and asthma

* Add investigational drug for epilepsy for 3 months

* Schedule follow-up appointment in 1 month to reassess seizure activity and adjust medications as needed.

----------------END OF PATIENT ENCOUNTER---------------

-------------- PATIENT ID 206 -- VISIT 2 -----------

Clinic Note: Visit #2 (2022-09-05)

Patient ID: 206

Age: 81 years

Gender: Female

Chief Complaint: Epilepsy, dizziness, somnolence

History of Present Illness:

The patient is an 81-year-old female with a history of diabetes, schizophrenia, asthma, and epilepsy. She has been participating in a randomized controlled trial and has been taking the investigational drug ID X13737 for the past three months. During this time, she has experienced 9 seizures and has reported dizziness and somnolence. The investigational drug will be discontinued now that the study is complete.

Past Medical History:

* Diabetes mellitus type 2

* Schizophrenia

* Asthma

* Epilepsy

Medication List:

* Metformin (glucose lowering agent)

* Lithium carbonate (mood stabilizer)

* Inhaler (beta-agonist and inhaled corticosteroid)

* Levetiracetam (anticonvulsant)

* Lacosamide (anticonvulsant)

Social History:

* Married, lives with spouse

* No known allergies

* Non-smoker

* Moderate alcohol consumption

Review of Systems:

* No recent changes in symptoms or medications

Neurological Examination:

* Mental Status: Alert and oriented x3. No evidence of cognitive decline or psychosis.

* Speech: Clear and coherent. No slurred or abnormal speech noted.

* Motor Examination: No significant weakness, ataxia, or tremors observed. No evidence of seizure activity.

* Sensory Examination: Intact sensation to light touch, pain, temperature, and vibration.

* Cranial Nerve Examination: No abnormal findings.

* Reflexes: Normal.

* Gait and Balance: Stable and normal gait observed.

Assessment and Plan:

The patient's recent seizure activity, despite being on anticonvulsant medications, is a concern. The decision has been made to discontinue the investigational drug ID X13737 due to the potential risks outweighing the benefits. The patient will be transitioned back to standard anticonvulsant therapy and closely monitored for seizure activity. Additionally, the patient's dizziness and somnolence will be addressed with further evaluation and possible medication adjustments.

The patient will follow up in one month for a status check and to assess the effectiveness of the new medication regimen.

Additional orders:

* Lab work to monitor liver enzymes and complete blood count

* Transition to standard anticonvulsant therapy

* Follow-up appointment in one month

----------------END OF PATIENT ENCOUNTER---------------

-------------- PATIENT ID 207 -- VISIT 1 -----------

On February 15, 2022, I conducted visit #1 with Patient ID 207, a 22-year-old male who presented with a history of ADHD, dermatitis, asthma, and parietal lobe epilepsy. The patient has had an alarming increase in seizures over the past two months, with a total of 34 seizures reported. Notably, this increase in seizures coincides with the patient's participation in a randomized controlled trial (RCT) investigating a new drug for epilepsy.

Upon examination, the patient's neurological status was found to be stable, with no notable changes since the previous visit. The patient reported no recent medication changes or triggers that may have contributed to the increased seizure activity. However, the patient did report experiencing more frequent and intense seizures since initiating the RCT, which is a common side effect of the investigational drug being used.

As part of the RCT, the patient has been prescribed the investigational drug for epilepsy for a period of three months. The addition of this drug to the patient's existing medication regimen is intended to evaluate its efficacy in managing seizures. However, no changes will be made to the patient's current medications other than the addition of the investigational drug. This decision was made after careful consideration of the potential risks and benefits of the new drug, as well as consultation with the patient's primary care physician and neurologist.

Overall, the patient's condition remains stable, but the recent increase in seizures is a cause for concern. Close monitoring and regular follow-up visits will be crucial to assess the effectiveness of the investigational drug and manage any potential side effects or complications. Additionally, it will be essential to continue evaluating the patient's neurological status and adjusting their medication regimen as needed to ensure optimal seizure control and overall health.

----------------END OF PATIENT ENCOUNTER---------------

-------------- PATIENT ID 207 -- VISIT 2 -----------

---

Clinic Note #2 (dated March 15, 2022)

for Patient ID 207 (a 22-year-old male)

Identification:

* Name: [Patient's Name]

* Date of Birth: [Date of Birth]

* Patient ID: 207

HPI (History of Present Illness):

* Presents with a history of ADHD, dermatitis, asthma, and parietal lobe epilepsy

* Participated in a randomized controlled trial for investigational drug ID X13737 for epilepsy over the past 3 months

* Experienced 118 seizures during the study period, with no symptoms reported

Subjective:

* Patient reports no current symptoms or concerns

* Reports having been compliant with the investigational drug regimen throughout the study period

* Denies any recent changes in sleep, appetite, mood, or cognitive function

Medications:

* ID X13737 (investigational drug for epilepsy), discontinued as of today due to completion of the randomized controlled trial

* Previous medications for ADHD, dermatitis, and asthma as documented in previous clinic notes

Allergies:

* None reported

Social History:

* Married with two children (ages 4 and 6)

* Works as a software engineer

* Denies any recent travel or exposure to illness

Review of Systems:

* No significant changes in the patient's history of ADHD, dermatitis, asthma, or seizures since previous clinic note (dated February 15, 2022)

* Reports no new medications or supplements initiated since the last visit

General Physical Examination:

* Vital signs within normal limits

* Head and neck examination reveals normal facial movements and reactivity to light and sound

* Extremities show normal tone and reflexes

* No signs of skin rash or exfoliation

* No evidence of laryngeal or pulmonary edema

Comprehensive Neurologic Examination:

* Cranial nerves: Intact (including visual fields, extraocular movements, and facial movements)

* Motor examination: Normal strength and tone in all extremities with normal coordination and balance

* Sensory examination: Intact (including sensation to light touch, pain, and temperature)

* Reflexes: Normal (including knee jerk, ankle jerk, and patellar reflexes)

* Coordination and balance: Normal

Labs:

* Complete blood count (CBC): Within normal limits

* Basic metabolic panel (BMP): Within normal limits

* Thyroid-stimulating hormone (TSH): 4.2 mIU/L (normal range: 0.4 - 4.0 mIU/L)

Imaging:

* No recent imaging studies have been performed

Assessment:

* Patient has a history of ADHD, dermatitis, asthma, and parietal lobe epilepsy, with no significant changes in symptoms or medications since previous clinic note (dated February 15, 2022)

* Patient recently completed a randomized controlled trial for investigational drug ID X13737 for epilepsy, with no reported symptoms and 118 seizures experienced during the study period

* No significant changes in the patient's neurological examination or laboratory results since previous clinic note (dated February 15, 2022)

Plan:

* Continue current medications as previously prescribed, including ID X13737 (discontinued as of today due to completion of the randomized controlled trial) and any other medications as needed for ADHD, dermatitis, asthma, and seizures

* Schedule follow-up clinic note in 3 months to assess for any changes in symptoms or medication needs

Signature:

[Your Signature]

Date: March 15, 2022

----------------END OF PATIENT ENCOUNTER---------------

-------------- PATIENT ID 208 -- VISIT 1 -----------

On November 7th, 2022, I had the pleasure of conducting visit #1 with Patient ID 208, an 87-year-old male who presented with a history of generalized epilepsy and recent increase in seizure activity. Over the past two months, the patient has experienced seven seizures, which is a significant change from their previous frequency of one to two seizures per month.

During the examination, I conducted a thorough neurological assessment of the patient's cognitive and motor functions, including checks for memory, speech, language, balance, coordination, and reflexes. The patient's cognitive function was found to be intact, with no signs of decline or impairment. However, their motor function showed mild weakness on the left side of their body, which may be related to the patient's history of seizures.

The patient is currently participating in a randomized controlled trial for the treatment of their epilepsy, and no changes will be made to their medication regimen at this time, other than the addition of the investigational drug for epilepsy. This new medication will be administered for a period of three months, after which its effectiveness will be re-evaluated. The patient has been instructed to continue with their regular seizure medications and to report any changes in their seizure activity or side effects from the new medication.

In conclusion, visit #1 with Patient ID 208 revealed a patient with a history of generalized epilepsy and increased seizure activity over the past two months. While the patient's cognitive function remains intact, mild weakness on the left side of their body was noted. The patient will continue to participate in the randomized controlled trial with no changes to their medication regimen at this time, other than the addition of the investigational drug for epilepsy. Regular follow-up appointments have been scheduled to monitor the patient's seizure activity and any potential side effects from the new medication.

----------------END OF PATIENT ENCOUNTER---------------

-------------- PATIENT ID 208 -- VISIT 2 -----------

---

Clinic Visit #2 (2022-12-07)

Patient ID: 208

Age: 87 years old

History: Generalized epilepsy, randomized controlled trial (RCT) for investigational drug ID X13737

Chief Complaint: Seizures and nystagmus

Past Medical History: None

Medications: None (previously taking ID X13737 as part of the RCT)

Social History: None

Review of Systems: Negative for fever, chills, or other symptoms

Physical Examination:

Neurological:

* General: Alert and oriented x3

* Mental status: Normal cognitive function

* Speech: Clear and coherent

* Motor: No evidence of weakness or paralysis

* Sensory: Full range of sensation in all extremities

* Coordination/ balance: Stable

* Reflexes: Present and normal

* Muscle strength: Normal

* Nystagmus: Present, sustained and oscillating

* Ophthalmoplegia: None

Systemic:

* Vital signs: Blood pressure 120/80 mmHg, heart rate 70 bpm, temperature 98.6°F (37°C)

* Head, Eyes, Ears, Nose, and Throat (HEENT): Normocephalic, normotrophic, no evidence of mass lesions or swelling

* Cardiovascular: Regular rhythm, normal S1 and S2 sounds, no murmurs or rubs

* Respiratory: Clear to auscultation, no wheezing or crackles

* Abdominal: Soft, normoactive bowel sounds

* Extremities: No evidence of weakness or paralysis, normal tone and reflexes

Assessment:

The patient is an 87-year-old male with a history of generalized epilepsy who has been participating in a randomized controlled trial for investigational drug ID X13737. Over the past 3 months, the patient has experienced 14 seizures and has reported nystagmus. The patient's neurological examination reveals sustained and oscillating nystagmus, but no other abnormalities.

Plan:

1. Discontinue investigational drug ID X13737 as the study is complete.

2. Monitor the patient for any changes in seizure frequency or severity.

3. Consider initiating alternative anti-seizure medication if seizures persist or worsen.

4. Schedule follow-up appointments as needed to monitor the patient's condition and adjust treatment plan accordingly.

---

Note: This clinic note is fictional and for educational purposes only. Any resemblance to real patients, events, or medical practices is purely coincidental.

----------------END OF PATIENT ENCOUNTER---------------

-------------- PATIENT ID 209 -- VISIT 1 -----------

On May 3rd, 2022, I conducted a clinic visit with Patient ID 209, an 80-year-old male who has been diagnosed with schizophrenia, diabetes, and epilepsy. The patient has had a significant increase in seizure activity over the past two months, with 19 seizures reported during this time period. As part of a randomized controlled trial, the patient will be adding an investigational drug for epilepsy to their current medication regimen, which will be taken for three months.

During the clinic visit, I performed a complete neurological examination on the patient. This included an assessment of their cognitive function, including memory and attention, as well as an evaluation of their speech and language abilities. Additionally, I conducted a thorough examination of the patient's nervous system, including their sensory systems, reflexes, and coordination. The results of the examination showed no significant changes or abnormalities in the patient's neurological functioning.

The patient reported that they have been experiencing increased seizure activity over the past two months, with a total of 19 seizures during this time period. The patient also reported that their current medication regimen has not had a significant impact on reducing the frequency or severity of these seizures. As part of the randomized controlled trial, the patient will be adding an investigational drug for epilepsy to their current medication regimen, which will be taken for three months. The patient was informed that this new medication may help reduce the frequency and severity of their seizures, but it is also possible that the medication may not have any effect on their condition.

In conclusion, Patient ID 209 presented to the clinic with a history of schizophrenia, diabetes, and epilepsy, as well as a significant increase in seizure activity over the past two months. The patient will be participating in a randomized controlled trial, during which they will be adding an investigational drug for epilepsy to their current medication regimen. A complete neurological examination was conducted, and the results showed no significant changes or abnormalities in the patient's neurological functioning. The patient was informed of the potential risks and benefits of the new medication, and they were instructed to follow up with the clinic in three months to assess the effectiveness of the treatment.

----------------END OF PATIENT ENCOUNTER---------------

-------------- PATIENT ID 209 -- VISIT 2 -----------

Patient ID: 209

Date: 2022-06-03

HPI:

* 80 year old male

* History of schizophrenia, diabetes, and epilepsy

* Currently participating in a randomized controlled trial (RCT) for epilepsy

Neurological Examination:

* Vitals: BP 120/80, HR 70, T 98.6

* General: Alert and oriented x3

* Mental Status: Intact, no hallucinations or delusions

* Cranial Nerves: Normal

* Motor: No tremors or rigidity

* Sensory: Normal sensation in all extremities

* Reflexes: 2+ (normal)

Pertinent Positives:

* Dizziness

Assessment and Plan:

* Discontinue investigational drug ID X13737 due to high seizure frequency (120 seizures in 3 months)

* Monitor for withdrawal symptoms from discontinuation of drug

* Address dizziness with appropriate medication or referral to specialist if necessary

* Schedule follow-up appointment in 1 month to assess for any changes in seizure frequency or other neurological symptoms.

----------------END OF PATIENT ENCOUNTER---------------

-------------- PATIENT ID 220 -- VISIT 1 -----------

Patient ID: 220

Visit #1 (2021-05-05)

* History: Arthritis, occipital lobe epilepsy (9 seizures in past 2 months)

* Randomized controlled trial (no change in medications except for investigational drug for epilepsy)

Neurological Examination:

* Mental status: Alert and oriented x3

* Speech: Intelligible, normal volume and tone

* Motor function: Decreased mobility in left arm and leg (arthritis)

* Sensation: Normal in all extremities

* Reflexes: 2+ (knee jerk and ankle jerk)

* Coordination: Decreased on the left side (arthritis)

Pertinent Positives:

* Seizures: 9 in past 2 months (occipital lobe epilepsy)

* No change in medications except for investigational drug for epilepsy

Assessment and Plan:

* Monitor seizure activity and adjust trial medication as needed

* Continue with randomized controlled trial

* Schedule follow-up visit in 3 months to assess response to investigational drug and monitor for any changes in seizure activity.

----------------END OF PATIENT ENCOUNTER---------------

-------------- PATIENT ID 220 -- VISIT 2 -----------

Clinic Note Vist #2

Patient ID: 220

Date: 2021-06-05

Identification:

Name: John Smith

DOB: 1924-01-01

ID Number: 220

History of Present Illness (HPI):

Mr. Smith, a 97-year-old male, presents with a history of arthritis and occipital lobe epilepsy. He has been participating in a randomized controlled trial (RCT) evaluating the safety and efficacy of the investigational drug ID X13737 for the treatment of epilepsy. Over the past 3 months, Mr. Smith has taken the investigational drug and has experienced 17 seizures. He reports no symptoms.

Subjective:

Mr. Smith denies any recent changes in his health or medication adherence. He mentions that he has been experiencing some mild joint pain, which is consistent with his known history of arthritis.

Medications:

ID X13737 (investigational drug for epilepsy) - discontinued due to completion of RCT

Levodopa (for parkinsonism)

Ondansetron (for nausea and vomiting)

Aspirin (for arthritis)

Allergies:

None reported

Social History:

Mr. Smith is a retired farmer who enjoys reading and spending time with his family. He lives alone and has a support system in place. He denies any recent travel or exposure to illness.

Review of Systems:

General: No complaints of fever, chills, or fatigue.

Cardiovascular: No chest pain, shortness of breath, or palpitations.

Respiratory: No cough, shortness of breath, or wheezing.

Gastrointestinal: No nausea, vomiting, or diarrhea.

Musculoskeletal: Mild joint pain consistent with known history of arthritis.

Neurological: No recent changes in seizure activity.

Psychiatric: No recent changes in mood or cognition.

General Physical Examination:

Vital Signs:

* Temperature: 98.6°F (37°C)

* Blood Pressure: 120/80 mmHg

* Pulse: 70 beats per minute

* Respiratory Rate: 14 breaths per minute

Head:

* No focal neurological deficits

* Normal cranial nerve examination

Eyes:

* No gaze evoked nystagmus or other abnormal eye movements

* Normal visual acuity and fundus examination

Ears:

* No ear discharge or abnormal hearing

Mouth:

* No oral lesions or abnormalities

Neck:

* No neck mass or stiffness

Extremities:

* Mild joint pain in the hands and knees consistent with known history of arthritis

Comprehensive Neurologic Exam:

Mr. Smith underwent a comprehensive neurologic examination, including an evaluation of his cognitive function, mood, and balance. His speech is clear and concise, and he is able to follow simple commands. His memory and attention span are within normal limits. His mood is euphoric, and he denies any recent changes in his emotional state. His balance is intact, and he is able to walk independently without assistance.

Labs:

* Complete Blood Count (CBC): Within normal limits

* Electrolyte Panel: Within normal limits

* Renal Function Panel: Within normal limits

* Thyroid-stimulating Hormone (TSH) level: Within normal limits

Imaging:

* Computed Tomography (CT) scan of the brain: No abnormalities noted

Assessment:

Mr. Smith is a 97-year-old male with a history of arthritis and occipital lobe epilepsy who has been participating in a randomized controlled trial evaluating the safety and efficacy of the investigational drug ID X13737 for the treatment of epilepsy. Over the past 3 months, Mr. Smith has taken the investigational drug and has experienced 17 seizures. He reports no symptoms. The investigational drug will be discontinued now that the study is complete.

Plan:

1. Discontinue ID X13737

2. Monitor Mr. Smith's seizure activity closely and adjust medication as needed

3. Continue levodopa, ondansetron, and aspirin as prescribed

4. Schedule follow-up appointments as needed to monitor his condition

Signature:

[Your Signature]

Date: 2021-06-05

----------------END OF PATIENT ENCOUNTER---------------

-------------- PATIENT ID 221 -- VISIT 1 -----------

CLINIC NOTE

Visit #1

Date: 2022-04-02

Patient ID: 221

Chief Complaint:

The patient presents with a history of ADHD, arthritis, anxiety, and generalized epilepsy. He reports having experienced 27 seizures in the past 2 months. He is currently enrolled in a randomized controlled trial and will be taking the investigational drug for epilepsy for the next 3 months.

Past Medical History:

The patient has a history of ADHD, arthritis, anxiety, and generalized epilepsy. He has been experiencing an increase in seizure activity over the past 2 months.

Medication List:

The patient is currently taking levothyroxine, lisinopril, metformin, meloxicam, and venlafaxine. He will be added to the investigational drug for epilepsy.

Social History:

The patient denies any recent travel, illnesses, or surgeries. He is a non-smoker and does not drink alcohol. He lives with his wife and children and works as an accountant.

Family History:

The patient's father had a history of seizures and died at the age of 52 due to a sudden cardiac death. His mother has a history of hypertension and hyperlipidemia.

Review of Systems:

The patient denies any recent changes in sleep, appetite, or mood. He reports no recent infections or illnesses. He does report ongoing anxiety and difficulty concentrating.

Neurological Examination:

The patient's neurological examination was unremarkable except for the following:

* Mild tremors were noted in both hands.

* The patient had a slight limp due to arthritis.

* There was no evidence of sensory or motor deficits.

* The patient's speech was clear and coherent.

Assessment and Plan:

Based on the patient's history, physical examination, and laboratory results, I have made the following assessments and recommendations:

1. The patient's increased seizure activity is likely related to his generalized epilepsy.

2. The patient's anxiety and difficulty concentrating may be related to his ADHD and seizure activity.

3. The patient's arthritis may be contributing to his limp and mild tremors.

4. The patient will continue to participate in the randomized controlled trial for the next 3 months, taking the investigational drug for epilepsy as prescribed.

5. The patient's medications will remain unchanged except for the addition of the investigational drug for epilepsy.

6. The patient will be monitored closely for any changes in seizure activity or adverse effects from the investigational drug.

7. The patient will be scheduled for follow-up visits every 2 months to assess his seizure activity, anxiety, and overall health.

Signature:

[Patient's signature]

Date:

[Current date]

Note: This is a fictional case and the patient information, medical history, and examination results are not real. The purpose of this note is to demonstrate the style of a neurologist clinic note for a patient with a history of ADHD, arthritis, anxiety, and generalized epilepsy.

----------------END OF PATIENT ENCOUNTER---------------

-------------- PATIENT ID 221 -- VISIT 2 -----------

Clinic Note - Visit #2

Patient ID: 221

Date: 2022-05-02

Chief Complaint:

The patient presents with a history of ADHD, arthritis, anxiety, and generalized epilepsy. He is currently enrolled in a randomized controlled trial evaluating the investigational drug ID X13737 for the treatment of epilepsy. The patient has been taking the medication for the past 3 months and has reported experiencing somnolence.

History of Present Illness:

The patient was previously prescribed the investigational drug ID X13737 for the treatment of his epilepsy. He has been adherent to the medication regimen and has taken the medication as directed. Over the past 3 months, the patient has experienced 15 seizures. He reports that the somnolence he experienced while taking the medication was significant and interfered with his daily activities.

Past Medical History:

The patient has a history of ADHD, arthritis, anxiety, and generalized epilepsy. He has been previously treated with various medications, including antidepressants, anti-anxiety medications, and seizure medications.

Medication List:

Current Medications:

* ID X13737 (investigational drug for epilepsy) - discontinued due to completion of the clinical trial

* Levodopa (for Parkinson's disease)

* Meloxicam (for arthritis)

* Lorazepam (for anxiety)

* Lamotrigine (for epilepsy)

Social History:

The patient is a 40-year-old male who works as an accountant. He is married and has two children. He denies any significant social or environmental stressors. He reports that he has been adherent to his medication regimen and has not experienced any notable side effects.

Family History:

The patient's family history is unremarkable for neurological disorders.

Review of Systems:

The patient denies any recent changes in his vision, hearing, or speech. He reports no recent muscle weakness, atrophy, or paralysis. He denies any recent difficulty with swallowing, speaking, or breathing. He reports no recent joint pain or swelling.

Neurological Examination:

The patient's neurological examination was significant for:

1. Cranial nerves: The patient's cranial nerves were normal, with no evidence of abnormalities in the vision, hearing, or facial movements.

2. Motor examination: The patient had a normal motor examination, with no evidence of weakness, atrophy, or paralysis.

3. Sensory examination: The patient's sensory examination was normal, with no evidence of numbness, tingling, or loss of sensation in the arms and legs.

4. Coordination and balance: The patient had a normal coordination and balance, with no evidence of ataxia or dizziness.

5. Reflexes: The patient's reflexes were normal, with no evidence of hyporeflexia or hyperreflexia.

Assessment and Plan:

Based on the patient's history and neurological examination, it is my assessment that the patient has experienced somnolence while taking the investigational drug ID X13737 for his epilepsy. The medication will be discontinued now that the clinical trial is complete. I recommend follow-up appointments to monitor the patient's seizure activity and adjust his medication regimen as needed. Additionally, I recommend lifestyle modifications such as regular exercise, a balanced diet, and stress management techniques to improve the patient's overall health and well-being.

Plan:

1. Discontinue the investigational drug ID X13737.

2. Monitor the patient's seizure activity at follow-up appointments.

3. Adjust the patient's medication regimen as needed.

4. Recommend lifestyle modifications such as regular exercise, a balanced diet, and stress management techniques.

5. Schedule follow-up appointments as needed.

----------------END OF PATIENT ENCOUNTER---------------

-------------- PATIENT ID 222 -- VISIT 1 -----------

---

Clinic Note - Visit #1 (2022-09-21)

Patient ID: 222 (Female, 29 years old)

History of Present Illness:

The patient is a 29-year-old female with a history of alcoholism and parietal lobe epilepsy. She has experienced 8 seizures in the past 2 months, which are consistent with her previous baseline frequency. The patient is currently enrolled in a randomized controlled trial (RCT) and is taking no changes in medication other than the addition of the investigational drug for epilepsy, which she will take for 3 months.

Neurological Examination:

General: The patient appears well-oriented and alert. Speech is clear and concise.

Head: No focal deficits or abnormalities.

Eyes: Pupils are symmetrical and react to light. Extremely sensitive to light, particularly blue light (as reported by the patient). No nystagmus or gaze evoked nystagmus.

Cranial Nerves:

* Olfactory: Intact (patient reports no change in sense of smell)

* Optic: Intact (patient reports blurred vision, particularly when exposed to blue light)

* Ocular Motility: Full range of motion, no nystagmus or abnormalities

* Facial: Intact (patient reports no weakness or paresis)

* Tongue: Intact (patient reports no weakness or paresis)

* Mandibular: Intact (patient reports no weakness or paresis)

Motor:

* Upper Limb: No significant weakness or atrophy noted in the upper limbs. Patient reports some mild weakness in the right hand and wrist, but it does not interfere with daily activities.

* Lower Limb: No significant weakness or atrophy noted in the lower limbs.

Sensory:

* Touch: Intact (patient reports no numbness or tingling sensations)

* Pain: Intact (patient reports no changes in pain perception)

* Temperature: Intact (patient reports no changes in temperature sensation)

* Proprioception: Intact (patient reports no changes in joint position sense)

Assessment and Plan:

Based on the patient's history of alcoholism and parietal lobe epilepsy, it is essential to closely monitor her condition during the RCT. The patient has experienced an increase in seizures over the past 2 months, which may be related to the study medication or other factors. Therefore, we will continue to monitor her seizure activity and adjust the study medication accordingly.

The patient's neurological examination revealed no significant deficits or abnormalities, except for extreme sensitivity to blue light. This may be related to her previous history of parietal lobe epilepsy. We will continue to monitor her visual function and adjust the study medication accordingly.

We will also continue to monitor the patient's alcohol consumption, as it may have a significant impact on her seizure activity and overall health. The patient reports no changes in alcohol consumption since enrolling in the RCT.

Plan:

* Continue to monitor the patient's seizure activity and adjust the study medication accordingly.

* Monitor the patient's visual function and adjust the study medication accordingly.

* Continue to monitor the patient's alcohol consumption and address any concerns or issues that arise.

* Follow up with the patient in 2 weeks for a follow-up examination and to review the study medication and its effects.

----------------END OF PATIENT ENCOUNTER---------------

-------------- PATIENT ID 222 -- VISIT 2 -----------

Visit #2 - 2022-10-21

Patient ID: 222

Age: 29 years

Gender: Female

History:

* Alcoholism

* Parietal lobe epilepsy

* Randomized controlled trial (RCT) participant (ID X13737) for epilepsy (last 3 months)

Current Status:

* Seizures: 17 (over past 3 months)

* No symptoms reported

Neurological Examination:

* Mental status: Alert and oriented

* Head: Normal head circumference, normocephalic

* Eyes: No abnormalities noted

* Visual fields: Full

* Extremities: Normal strength and tone

* Sensation: Intact

* Coordination: Normal

Assessment:

* Discontinue investigational drug ID X13737 (RCT)

* Monitor for seizure activity and any adverse effects

Plan:

* Follow-up visit in 2 weeks to assess for any changes in seizure activity or adverse effects

* Consider alternative treatments for epilepsy if necessary

----------------END OF PATIENT ENCOUNTER---------------

-------------- PATIENT ID 223 -- VISIT 1 -----------

Patient ID: 223

Visit #1: 2022-02-01

History:

* 72-year-old female

* History of anxiety, diabetes, schizophrenia, and parietal lobe epilepsy

* 65 seizures in the past 2 months

* Currently enrolled in a randomized controlled trial

Neurological Examination:

* Mental status: Alert and oriented

* Speech: Clear and normal volume

* Motor function: No tremors or rigidity noted

* Sensation: Intact

* Reflexes: 2+ (mild increase in muscle tone)

* Coordination and balance: Normal

Pertinent Positives:

* Recent seizure activity (65 seizures in past 2 months)

* No change in medications other than addition of investigational drug for epilepsy

Assessment and Plan:

* Continue with current medications, including the investigational drug for epilepsy

* Monitor seizure activity and adjust medications as needed

* Follow up in 1 month to assess efficacy of treatment and make any necessary changes.

----------------END OF PATIENT ENCOUNTER---------------

-------------- PATIENT ID 223 -- VISIT 2 -----------

Clinic Note - Visit #2 (2022-03-01)

Patient ID: 223

Age: 72 years old (female)

History: Anxiety, Diabetes, Schizophrenia, Parietal Lobe Epilepsy

Randomized Controlled Trial: The patient has been participating in a randomized controlled trial for the investigational drug ID X13737 for epilepsy over the past 3 months.

Neurological Examination:

General: The patient appears well-oriented and alert. Speech is clear and comprehensible.

Motor: No noticeable tremors or weakness in the extremities.

Sensory: No deficits in sensation to light touch, pain, or temperature.

Cerebellar Function: The patient's cerebellar function is within normal limits.

Reflexes: Normal reflexes are present and appropriate for age.

Muscle Strength: Muscle strength is normal.

Coordination and Balance: The patient has a normal range of motion and balance.

Cognitive Function: The patient's cognitive function appears to be intact, with no evidence of impairment in attention, memory, or executive function.

Psychiatric Status: The patient reports ongoing anxiety and fatigue. No signs of psychosis or mania are present.

Medical History: The patient has a history of anxiety, diabetes, schizophrenia, and parietal lobe epilepsy. She has been taking the investigational drug ID X13737 for epilepsy over the past 3 months and has experienced 75 seizures during this time.

Plan: The patient will be discontinued from the investigational drug ID X13737 as the study is complete. Her anxiety and fatigue will be monitored and addressed as needed. The patient's ongoing care for her medical conditions, including diabetes, schizophrenia, and epilepsy, will continue as usual.

----------------END OF PATIENT ENCOUNTER---------------

-------------- PATIENT ID 224 -- VISIT 1 -----------

Clinic Note Visit #1

Date: 2023-01-16

Patient ID: 224

Identification:

Name: John Doe

DOB: 01/01/1935

Highlights of the History of Present Illness (HPI):

Mr. Doe, an 88-year-old male, presents with a history of alcoholism and parietal lobe epilepsy. He has had 82 seizures in the past 2 months, with increasing frequency and severity. His condition has significantly impacted his daily activities and quality of life.

Subjective:

Mr. Doe reports a persistent headache, which is exacerbated by visual stimuli and has been present for the past 6 months. He also notes difficulty with speech and language processing, which has been ongoing for the past year. Additionally, he experiences frequent dizziness and nausea, especially after seizures. He denies any recent changes in medications or significant life events.

Medications:

* Levetiracetam 500mg twice daily (as directed by primary care physician)

* Lamotrigine 200mg once daily (as directed by primary care physician)

Allergies:

None reported.

Social History:

Mr. Doe is a retired engineer and lives with his wife in an assisted living facility. He has a history of alcoholism, which he has been abstinent from for the past 10 years. He reports no significant social or recreational activities due to his health status. His family is supportive and involved in his care.

Review of Systems:

General: No recent illnesses or injuries.

Cardiovascular: No history of cardiovascular disease, but he does report occasional shortness of breath.

Respiratory: No history of respiratory diseases.

Gastrointestinal: No history of gastrointestinal diseases.

Genitourinary: No history of genitourinary diseases.

Musculoskeletal: No history of musculoskeletal diseases.

Neurological: History of parietal lobe epilepsy, with 82 seizures in the past 2 months.

Psychiatric: No history of psychiatric illnesses.

General Physical Examination:

Vital signs:

* Temperature: 98.6°F (37°C)

* Blood Pressure: 120/80 mmHg

* Heart Rate: 70 beats per minute

* Respiratory Rate: 14 breaths per minute

Head and Neck:

* Normal head shape and symmetry.

* No signs of trauma or deformity.

* No focal neurological deficits.

Extremities:

* Normal tone, power, and reflexes in all extremities.

Comprehensive Neurologic Examination:

Cranial Nerves:

* I (olfactory) - normal

* II (optic) - normal

* III (oculomotor) - normal

* IV (trochlear) - normal

* V (trigeminal) - normal

* VI (abducens) - normal

* VII (facial) - normal

* VIII (vestibulo-cochlear) - normal

* IX (glossopharyngeal) - normal

* X (vagus) - normal

Motor:

* Mr. Doe demonstrates normal motor function, with no signs of weakness or atrophy.

Sensory:

* No significant sensory deficits were identified.

Labs:

* Complete Blood Count (CBC): Within normal limits.

* Serum Electrolytes: Within normal limits.

* Liver Function Tests (LFTs): Within normal limits.

* Renal Function Tests (RFTs): Within normal limits.

Imaging:

* No significant abnormalities were identified on CT scan of the head.

Assessment:

* Parietal lobe epilepsy with increased seizure frequency and severity in the past 2 months.

* History of alcoholism, which may have contributed to the development of parietal lobe epilepsy.

* No significant changes in medications will be made at this time, other than adding the investigational drug for epilepsy that will be taken for 3 months.

* Mr. Doe is randomized to receive the investigational drug as part of a controlled trial.

Plan:

* Continue levetiracetam and lamotrigine as directed by primary care physician.

* Add investigational drug for epilepsy, as part of the randomized controlled trial.

* Monitor seizure frequency and severity closely over the next 3 months.

* Schedule follow-up appointment in 2 weeks to assess tolerance to medications and changes in seizure activity.

Signature:

[Your signature here]

Date: 2023-01-16

Note: This clinic note is a sample and should not be used as an actual patient note without proper documentation and medical knowledge.

----------------END OF PATIENT ENCOUNTER---------------

-------------- PATIENT ID 224 -- VISIT 2 -----------

Visit #2: February 16, 2023

Patient ID 224, an 88-year-old male with a history of alcoholism and parietal lobe epilepsy, was seen in the clinic for his second visit. The patient has been participating in a randomized controlled trial (RCT) and has been taking the investigational drug ID X13737 for epilepsy over the past three months.

According to the patient's report, he has experienced 141 seizures during this time period, but fortunately, he has not experienced any symptoms. The investigational drug will be discontinued now that the study is complete.

During the neurological examination, the patient was found to have a stable cognitive status and no evidence of seizure activity at the time of the visit. His motor function was intact, with normal strength and tone in all extremities. His sensory examination revealed normal sensation in all modalities, including pain and temperature. The patient's reflexes were appropriate for his age, and there were no signs of ataxia or other abnormalities.

The patient's medical history, including his history of alcoholism and parietal lobe epilepsy, was reviewed, and it was noted that the patient has been compliant with the study medication and has not experienced any adverse effects. The patient's laboratory results were within normal limits, and there were no signs of any other medical conditions.

In light of the patient's stable condition and the completion of the RCT, it is recommended that the investigational drug ID X13737 be discontinued, and the patient be monitored closely for any changes in his seizure activity or other symptoms. The patient will be followed up with regular visits to ensure his continued well-being and to address any concerns or questions that may arise.

----------------END OF PATIENT ENCOUNTER---------------

-------------- PATIENT ID 225 -- VISIT 1 -----------

On November 14th, 2022, Patient ID 225, a 64-year-old female, visited our clinic for her first follow-up visit since being enrolled in a randomized controlled trial. The patient has a history of diabetes, dermatitis, and occipital lobe epilepsy, with one seizure occurring in the past two months.

Upon examination, the patient's neurological status was assessed as follows: Her general appearance was normal, with no signs of weakness or ataxia. Her speech was clear and comprehensible, with no evidence of dysarthria or aphasia. Her motor function was intact, with no signs of weakness, paralysis, or tremors. Her sensory examination revealed decreased sensation to light touch and pinprick over the left hemiparesis, consistent with her history of occipital lobe epilepsy. Her reflexes were normal, with no signs of hyporeflexia or hyperreflexia.

The patient reported no new seizures since her last visit, and her current medication regimen, including the investigational drug for epilepsy, remains unchanged. The patient will continue to take this medication for the next three months as part of the randomized controlled trial. The patient's vital signs were normal, with a blood pressure of 120/80 mmHg, a heart rate of 70 beats per minute, and a temperature of 98.6 degrees Fahrenheit.

In conclusion, Patient ID 225 had a follow-up visit on November 14th, 2022, during which her neurological status was assessed as normal, with no signs of new seizures or adverse effects from the investigational drug for epilepsy. The patient will continue to participate in the randomized controlled trial and will be monitored closely for any changes in her condition.

----------------END OF PATIENT ENCOUNTER---------------

-------------- PATIENT ID 225 -- VISIT 2 -----------

---

Clinic Note - Visit #2 (2022-12-14)

Patient ID: 225

Demographics:

* Age: 64 years

* Gender: Female

Chief Complaint:

* History of diabetes, dermatitis, and occipital lobe epilepsy

* Recent fall

History of Present Illness (HPI):

* The patient was enrolled in a randomized controlled trial (RCT) investigating the efficacy and safety of the novel drug ID X13737 for the treatment of occipital lobe epilepsy.

* The patient has been taking the investigational drug for the past 3 months and has experienced 11 seizures during this time.

* The patient reported falling on [date], which may be related to the seizure activity.

Subjective:

* The patient reports persistent symptoms of diabetes, including polyuria, polydipsia, and blurred vision.

* The patient experiences frequent skin rashes and dryness, which are consistent with dermatitis.

* The patient reports ongoing seizure activity, with an average frequency of 1-2 seizures per week.

* The patient describes the seizures as brief, generalized tonic-clonic events lasting approximately 30 seconds to 1 minute.

* The patient reports no significant changes in their vision or hearing.

Medications:

* ID X13737 (investigational drug) - discontinued due to completion of RCT

* Levetiracetam (Keppra) - ongoing

* Metformin (Glucophage) - ongoing

Allergies:

* None reported

Social History:

* Married with two adult children

* Retired from a career in education

* Active in local community organizations and charities

* No significant travel or exposure to medications or substances

Review of Systems (ROS):

* General: denies fever, chills, or weight loss

* Cardiovascular: negative for chest pain, shortness of breath, or palpitations

* Respiratory: denies cough, wheezing, or shortness of breath

* Gastrointestinal: denies abdominal pain, nausea, vomiting, or diarrhea

* Genitourinary: denies urgency, frequency, or burning during urination

* Musculoskeletal: denies joint pain or swelling

* Neurological: reports ongoing seizure activity and recent fall

General Physical Examination:

* Vital signs: blood pressure 120/80 mmHg, heart rate 70 beats per minute, temperature 98.6 degrees Fahrenheit

* Head: atraumatic fall with no evidence of trauma or deformity

* Eyes: no visual disturbances or abnormalities

* Ears: no hearing difficulties or discharge

* Nose: no nasal congestion or discharge

* Throat: no sore throat or difficulty swallowing

* Chest: clear lung sounds, no wheezing or rales

* Abdomen: soft, normoactive bowel sounds

* Extremities: no clubbing or cyanosis

Comprehensive Neurological Examination:

* Mental status: alert and oriented to person, place, and time

* Cranial nerves: intact (II-XII)

* Motor examination: no signs of weakness, atrophy, or tremors

* Sensory examination: no sensory deficits or abnormalities

* Coordination and balance: normal

* Reflexes: normal (1+/5)

Labs:

* Complete Blood Count (CBC): within normal limits (WNL)

* Basic Metabolic Panel (BMP): WNL

* Thyroid-stimulating hormone (TSH): 4.2 mIU/L (normal 0.5-5.0 mIU/L)

* Lipid profile: WNL

Imaging:

* Magnetic Resonance Imaging (MRI) of the brain: no evidence of acute or chronic changes in the occipital lobe

Assessment:

* The patient's history of diabetes, dermatitis, and occipital lobe epilepsy, along with recent seizure activity and fall, suggest a complex medical presentation.

* The patient's participation in a randomized controlled trial investigating the efficacy and safety of ID X13737 for the treatment of occipital lobe epilepsy adds an additional layer of complexity to the assessment.

* The patient's ongoing seizure activity, despite discontinuation of the investigational drug, is a cause for concern and warrants further evaluation and management.

Plan:

* Continue levetiracetam (Keppra) therapy as prescribed

* Discontinue ID X13737 (investigational drug) due to completion of RCT

* Increase frequency of follow-up appointments to monitor seizure activity and adjust medications as needed

* Consider initiation of additional anticonvulsant medications or alternative treatments for occipital lobe epilepsy

Signature:

[Your Signature]

Date: [Today's Date]

Time: [Today's Time]

----------------END OF PATIENT ENCOUNTER---------------

-------------- PATIENT ID 226 -- VISIT 1 -----------

Patient ID: 226

Date: 2022-02-21

HPI:

* 56-year-old male

* History of arthritis and parietal lobe epilepsy

* 40 seizures in past 2 months

* Randomized controlled trial (RCT) participant

Neurological Examination:

* Head: normocephalic, atraumatic

* Eyes: normal visual acuity and field, no diplopia or abnormal movements

* Face: normal facial sensation and motor function

* Neck: normal range of motion and no lymphadenopathy

* Extremities: normal strength and tone, no atrophy or deformity

* Sensory examination: intact

* Coordination and balance: normal

Pertinent Positives:

* No change in seizure frequency or severity since starting RCT

* Patient reports no new symptoms or concerns

Assessment and Plan:

* Continue RCT medication as prescribed

* Monitor patient's seizure frequency and severity every 2 weeks

* Schedule follow-up appointment in 2 months to assess patient's progress and potential side effects of RCT medication.

----------------END OF PATIENT ENCOUNTER---------------

-------------- PATIENT ID 226 -- VISIT 2 -----------

---

Clinic Note #2 (2022-03-21)

Patient ID: 226

Age: 56 years old

History of Present Illness:

The patient is a 56-year-old male with a history of arthritis and parietal lobe epilepsy. He has been participating in a randomized controlled trial for the investigational drug ID X13737 for his epilepsy over the past 3 months. During this time, he has experienced 143 seizures. Additionally, the patient reported experiencing diplopia.

Past Medical History:

The patient has a history of arthritis and parietal lobe epilepsy. He has been managed with medication for his arthritis and has had multiple episodes of seizures over the years.

Medications:

The patient is currently taking levetiracetam, valproate, and lamotrigine for his epilepsy. He also takes meloxicam for his arthritis.

Family History:

The patient does not have any significant family history of neurological disorders.

Social History:

The patient is a retired mechanic and denies any significant exposure to toxins or injuries. He lives with his wife and has two adult children. He denies any history of substance abuse or smoking.

Review of Systems:

General: The patient appears well-developed and well-nourished. He does not report any fever, chills, or weight loss.

Neurological: The patient reports diplopia, which is confirmed on examination. He also reports a history of seizures over the past 3 months, with a total of 143 seizures during this time. There is no history of headaches, weakness, or numbness/tingling.

Ocular: The patient has diplopia, which is confirmed on examination. He does not report any other ocular symptoms.

Motor: The patient has a normal range of motion in all extremities. There is no evidence of weakness or atrophy.

Sensory: The patient denies any sensory deficits.

Cerebral-vascular: The patient does not report any history of cerebral-vascular disease.

Musculoskeletal: The patient has a history of arthritis, which is confirmed on examination.

Psychiatric: The patient denies any significant psychiatric symptoms.

Lab Results:

All laboratory results are within normal limits, including electrolyte panels, liver function tests, and a complete blood count.

Imaging Studies:

The patient has had no recent imaging studies.

Neurological Examination:

The patient is alert and awake, with normal speech and language. His cognitive function appears intact, with no evidence of decline or confusion. The patient has a normal gait and station. He has no difficulty with tandem walking or heel-to-toe walking. His muscle strength is within normal limits, with no evidence of weakness or wasting. The patient's sensation is intact, with no evidence of numbness or tingling. His reflexes are normal, with no evidence of hyperreflexia or hyporeflexia. The patient has a normal funduscopic examination, with no evidence of retinal pathology. The patient has diplopia, which is confirmed on examination.

Assessment and Plan:

Based on the patient's history of seizures and diplopia, it is likely that he has a neurological disorder related to his parietal lobe epilepsy. The investigational drug ID X13737 will be discontinued due to the completion of the randomized controlled trial. The patient's seizures will be managed with existing medications, and his diplopia will be monitored closely. The patient will be referred to a neuro-ophthalmologist for further evaluation and management of his diplopia. The patient will also be scheduled for a follow-up appointment in 2 weeks to assess his seizure control and monitor his overall condition.

---

I hope this clinic note meets your needs! Let me know if there's anything else I can help with.

----------------END OF PATIENT ENCOUNTER---------------

-------------- PATIENT ID 227 -- VISIT 1 -----------

Patient ID: 227

Visit Date: 2021-05-11

Chief Complaint: The patient presents with a history of arthritis, bipolar disorder, and multifocal epilepsy. He has experienced 61 seizures in the past two months, and is currently enrolled in a randomized controlled trial.

Past Medical History: The patient has been diagnosed with arthritis for the past 5 years, and has been managing his symptoms with medication. He was also diagnosed with bipolar disorder 10 years ago, and has been receiving treatment for this condition as well. The patient's seizure history began 3 years ago, and he has been under the care of a neurologist since then.

Current Medications: The patient is currently taking levothyroxine (T4) for his hypothyroidism, lisinopril for his hypertension, celecoxib for his arthritis, and valproic acid for his seizures. He has been advised to continue these medications as prescribed.

Reason for Visit: The patient has been experiencing an increased frequency of seizures over the past two months, and has been referred to our clinic for further evaluation and management.

Neurological Examination: On examination, the patient appears well-developed and well-nourished, with no obvious signs of distress or discomfort. His speech is clear and concise, and his cognitive function seems intact. There is no evidence of weakness or paralysis on motor examination. The patient's sensory examination reveals decreased sensation to light touch and pinprick over the left hemiparesis, consistent with his history of seizures. His reflexes are normal, and there is no evidence of ataxia or abnormal gait.

Plan: The patient will continue his current medications as prescribed, and will be started on the investigational drug for epilepsy. The patient will be monitored closely over the next three months to assess the efficacy and safety of the new medication. No changes in the patient's medication regimen will be made at this time, other than the addition of the investigational drug. The patient will be seen again in one month for follow-up.

Additional Notes: The patient reported a recent increase in stress due to a family conflict, which may be contributing to his increased seizure activity. He has been advised to keep a seizure diary to track any changes in his symptoms and to contact the clinic immediately if he experiences any new or worsening symptoms.

----------------END OF PATIENT ENCOUNTER---------------

-------------- PATIENT ID 227 -- VISIT 2 -----------

---

Clinic Note #2 (dated 2021-06-11) for Patient ID 227:

Identification:

Name: John Doe

DOB: 01/01/1948

ID Number: 227

HPI (History of Present Illness):

Mr. Doe, a 73-year-old male, presents with a history of multifocal epilepsy, bipolar disorder, and arthritis. He has been participating in a randomized controlled trial for the investigational drug ID X13737 for his epilepsy over the past 3 months.

Subjective:

Mr. Doe reports no symptoms since his last clinic visit. He states that he has experienced 66 seizures during the past 3 months while taking the investigational drug, ID X13737. He denies any recent changes in his mood or mental status.

Medications:

ID X13737 (investigational drug for epilepsy) - discontinued as of today's date due to completion of the randomized controlled trial

Lamotrigine (for bipolar disorder and seizures) - ongoing

Methotrexate (for arthritis) - ongoing

Allergies:

None reported

Social History:

Married, living with spouse

No smoking or alcohol use

No known allergies to medications

Review of Systems:

General: No fever, chills, or fatigue.

Neurological: No change in seizure activity since last clinic visit. No new symptoms such as weakness, numbness, or difficulty with speech or vision.

Cardiovascular: No chest pain, shortness of breath, or palpitations.

General Physical Examination:

Vital signs:

* Blood pressure: 120/80 mmHg

* Pulse: 70 beats per minute

* Temperature: 98.6 degrees Fahrenheit

Neurological examination:

Mr. Doe presents with a normal gait and station. His speech is clear and concise, with no evidence of aphasia or dysarthria. He demonstrates intact sensation in all extremities, including touch, pain, temperature, and vibratory sensation. His cranial nerves are grossly intact, with no evidence of abnormalities in his visual, auditory, or olfactory functions. His motor examination reveals normal strength and tone in all muscle groups, with no signs of ataxia, tremor, or rigidity.

Comprehensive Neurologic Exam:

* Cranial nerves: Intact (including visual, auditory, olfactory)

* Motor examination: Normal strength and tone in all muscle groups

* Sensory examination: Intact sensation in all extremities (touch, pain, temperature, vibratory)

* Coordination and balance: Normal

Labs:

* Complete Blood Count (CBC): Within normal limits

* Electrolyte panel: Within normal limits

* Liver function tests (LFTs): Within normal limits

* Renal function tests (RFTs): Within normal limits

Imaging:

* Magnetic Resonance Imaging (MRI) of the brain: Within normal limits

* Computed Tomography (CT) scan of the head and neck: Within normal limits

Assessment:

Mr. Doe presents with a history of multifocal epilepsy, bipolar disorder, and arthritis. He has been participating in a randomized controlled trial for the investigational drug ID X13737 over the past 3 months. During this time, he has experienced 66 seizures, but reports no symptoms since his last clinic visit. His neurological examination is unremarkable, with intact sensation and motor function. The investigational drug will be discontinued as of today's date due to completion of the randomized controlled trial.

Plan:

1. Discontinue the investigational drug, ID X13737, as of today's date.

2. Monitor Mr. Doe closely for any changes in his seizure activity or symptoms.

3. Continue lamotrigine and methotrexate as prescribed.

4. Schedule a follow-up clinic visit in 2 weeks to assess for any changes in Mr. Doe's condition.

Signature:

[Your name and title]

---

I hope this helps! Please let me know if you have any further questions or if there's anything else I can do to assist you.

----------------END OF PATIENT ENCOUNTER---------------

-------------- PATIENT ID 228 -- VISIT 1 -----------

Date: December 1, 2022

Patient ID: 228

Age: 61 years

History: The patient is a 61-year-old male with a history of autism and epilepsy. He has been experiencing an increased frequency of seizures over the past two months, with a total of 18 seizures during this time. The patient is currently enrolled in a randomized controlled trial to assess the efficacy of an investigational drug for the treatment of epilepsy.

Neurological Examination:

On examination, the patient's vital signs were normal, with a temperature of 98.6 degrees Fahrenheit, blood pressure of 120/80 mmHg, and a heart rate of 70 beats per minute. The patient's neurological examination revealed no significant abnormalities, including a normal cognitive function, normal sensory examination, and normal motor function. However, the patient did exhibit some subtle abnormalities in his speech and language, which are consistent with his history of autism.

Plan: The patient will continue to receive the investigational drug for epilepsy for a period of three months. No changes will be made to the patient's current medications other than the addition of the investigational drug. The patient will be closely monitored for any adverse effects or changes in his seizure frequency during this time. The patient will be followed up with at the end of the three-month period to assess the efficacy of the investigational drug and make any necessary adjustments to his treatment plan.

Note: Throughout the course of this clinical trial, it is essential that the patient adhere to the study medication and follow-up schedule to ensure accurate data collection and meaningful results. The patient will be informed of the potential risks and benefits of the investigational drug and will be asked to provide written informed consent before starting the trial.

----------------END OF PATIENT ENCOUNTER---------------

-------------- PATIENT ID 228 -- VISIT 2 -----------

Patient ID: 228

Visit #2: 2023-01-01

Age: 61 years

History: Autism, Epilepsy (investigational drug ID X13737 for past 3 months)

Neurological Examination:

* Mental status: Alert and aware

* Cranial nerves: Normal

* Motor function: Normal

* Sensory examination: Normal

* Reflexes: Normal

* Coordination and balance: Normal

Pertinent Positives:

* No symptoms reported

* 50 seizures experienced during the past 3 months while taking investigational drug ID X13737

Assessment and Plan:

* Investigational drug ID X13737 to be discontinued

* Patient to be monitored for any changes in seizure activity or other symptoms

* Patient to be referred for continued management of epilepsy and autism.

----------------END OF PATIENT ENCOUNTER---------------

-------------- PATIENT ID 229 -- VISIT 1 -----------

---

Patient ID: 229; Date of Visit: 2023-02-06

Identification:

Name: [insert name here]

DOB: [insert date of birth here]

Gender: Female

Age: 29 years

HPI (History of Present Illness):

Chief Complaint: Recurrent seizures

Medical History: Depression, hypothyroidism, alcoholism, frontal lobe epilepsy

Social History: Currently participating in a randomized controlled trial

Subjective:

Patient reports experiencing 8 seizures over the past two months. These seizures have been characterized as simple partial seizures with no loss of consciousness, and have been accompanied by auras such as lightheadedness and confusion. Patient has noticed an increase in stress and sleep deprivation in recent weeks, which may be contributing to the increased frequency of seizures.

Medications:

Current medications include:

* Levetiracetam 500mg BID

* Lamotrigine 200mg OD

* Sertraline 50mg OD

Allergies: None reported

Social History:

Patient is a college-educated female currently residing in [insert location here]. She reports having a stable living situation and social support network. She has been participating in the randomized controlled trial for the past 3 months, and has been compliant with all study-related procedures and medications.

Review of Systems:

Patient reports no significant changes in her general health or constitutional symptoms since her last visit. She denies any new complaints or concerns.

General Physical Examination:

Vital signs: BP 120/80, HR 70, T 98.6, R 18

Head: Normocephalic, normal facial features. No papilledema or visual field defects noted.

Eyes: Pupils are equally reactive to light and there is no evidence of afferent pupillary defect. Extremity motor function is 5/5.

Ears: No discharge or fluid in the external auditory canals. No cerumen impaction. Hearing is normal to whispered voice.

Mouth: No mouth sores or lesions. No evidence of facial weakness or paresis.

Thorax: Clear to auscultation over all lobes. No added sounds or wheezing noted.

Abdomen: Soft and nontender to palpation throughout. No masses or organomegaly appreciated.

Extremities: Full range of motion in all joints. No swelling, erythema, or warmth observed.

Comprehensive Neurological Examination:

Cerebellar function: Normal

Motor function: 5/5 in all extremities

Sensory function: Intact to light touch, pain, and temperature

Reflexes: 2+ in all extremities

Coordination and balance: Normal

Labs:

Results of recent laboratory tests include:

* WBC count: 4.8 x 10^9/L (normal range: 4.5-11 x 10^9/L)

* Neutrophil count: 2.6 x 10^9/L (normal range: 2.0-7.0 x 10^9/L)

* Lymphocyte count: 1.2 x 10^9/L (normal range: 1.0-4.0 x 10^9/L)

* ESR: 35 mm/hr (normal range: 0-20 mm/hr)

* CRP: 7.8 mg/L (normal range: 0-6.0 mg/L)

Imaging:

Results of recent imaging studies include:

* CT scan of head, neck, and spine: No acute or chronic abnormalities noted.

* MRI of brain: No evidence of structural or metabolic abnormalities detected.

Assessment:

Patient presents with a history of recurrent seizures, which have increased in frequency over the past two months. The patient is participating in a randomized controlled trial and has been compliant with all study-related procedures and medications. Laboratory and imaging results are within normal limits. Based on the patient's history and current presentation, it is reasonable to suspect that the increased seizure activity may be related to the underlying epilepsy diagnosis.

Plan:

1. Continue all current medications as prescribed.

2. Add investigational drug for epilepsy (levetiracetam) at a dose of 500mg BID for a period of 3 months.

3. Monitor patient's seizure activity and adjust medications as needed.

4. Schedule follow-up visit in 1 month to assess patient's response to treatment and monitor for any changes in symptoms or laboratory results.

Signature: [insert signature here]

Date: 2023-02-06

----------------END OF PATIENT ENCOUNTER---------------

-------------- PATIENT ID 229 -- VISIT 2 -----------

Clinic Note Vist #2

Patient ID: 229

Date: 2023-03-06

Identification:

Name: [Name], [First Name]

DOB: [Date of Birth]

Gender: Female

ID Number: [Patient ID Number]

HPI:

The patient, a 29-year-old female with a history of depression, hypothyroidism, alcoholism, and frontal lobe epilepsy, presented for her second visit to the clinic. The patient has been participating in a randomized controlled trial (RCT) examining the efficacy and safety of the investigational drug ID X13737 for the treatment of epilepsy.

Subjective:

The patient reported no symptoms since her last visit. She did, however, note some changes in her mood, citing a perceived improvement in her overall sense of well-being. The patient attributed this to the investigational drug, which she had been taking for the past three months. During this time, she experienced 26 seizures, which she reported were well-controlled and did not significantly impact her daily activities.

Medications:

ID X13737 (investigational drug): The patient has been taking 500mg of ID X13737 twice daily for the past three months as part of the RCT. The drug will be discontinued now that the study is complete.

Allergies:

The patient reported no known allergies.

Social History:

The patient denies any history of substance abuse or other medical conditions. She does, however, report occasional alcohol consumption.

Review of Systems:

The patient reported no significant changes in her review of systems since her last visit.

General Physical Examination:

The patient's general physical examination was unremarkable, with no signs of illness or distress.

Comprehensive Neurological Examination:

Cranial nerves: The patient's cranial nerves were normal, with no evidence of weakness or abnormal function.

Motor function: The patient had full range of motion in all extremities, with no signs of weakness or atrophy.

Sensory function: The patient's sensory examination was unremarkable, with no deficits in sensation to light touch, pain, temperature, or vibration.

Coordination and balance: The patient demonstrated normal coordination and balance, with no signs of instability or dizziness.

Reflexes: The patient's reflexes were normal, with no signs of hyperreflexia or hyporeflexia.

Labs:

The patient's laboratory results were as follows:

* Complete Blood Count (CBC): Normal

* Electrolyte Panel: Normal

* Liver Function Tests (LFTs): Normal

* Renal Function Tests (RFTs): Normal

* Thyroid-stimulating hormone (TSH): Elevated (3.4 mIU/L)

Imaging:

The patient's imaging results were as follows:

* Computed Tomography (CT) scan of the brain: Normal

* Magnetic Resonance Imaging (MRI) of the brain: Normal

Assessment:

Based on the patient's history, physical examination, and laboratory and imaging results, the following assessments can be made:

* The patient has a history of depression, hypothyroidism, alcoholism, and frontal lobe epilepsy.

* The patient has been participating in an RCT examining the efficacy and safety of ID X13737 for the treatment of epilepsy.

* The patient reported no symptoms since her last visit, with no significant changes in her review of systems.

* The patient's comprehensive neurological examination was unremarkable, with no signs of weakness or abnormal function.

* The patient's laboratory results were normal, with the exception of an elevated TSH level (3.4 mIU/L).

* The patient's imaging results were also normal.

Plan:

Given the patient's history and current status, the following plan is recommended:

* Discontinue ID X13737 as part of the RCT.

* Monitor the patient's symptoms and seizure activity closely.

* Perform regular follow-up visits to assess the patient's ongoing health and any changes in her condition.

* Consider initiating thyroid hormone replacement therapy to address the patient's elevated TSH level.

Signature:

[Patient Signature]

[Date]

----------------END OF PATIENT ENCOUNTER---------------

### CLINIC NOTES - DRUG ARM

-------------- PATIENT ID 10 -- VISIT 1 -----------

On July 20, 2022, Patient ID 10, a 30-year-old male with a history of hypothyroidism and multifocal epilepsy, presented to the clinic for his first follow-up visit since enrolling in a randomized controlled trial. The patient had experienced 43 seizures in the past two months, a significant increase from his previous seizure activity.

During the examination, the patient was alert and coherent, with no signs of confusion or disorientation. His speech was clear and well-spoken, with no observable tremors or weakness in his extremities. The patient's neck was stable, and there were no signs of muscle wasting or weakness. His reflexes were normal, and there was no evidence of sensory deficits or abnormalities.

The patient's neurological examination revealed no significant changes from the previous visit. However, the patient did report an increase in seizure activity since beginning the trial, with a total of 43 seizures in the past two months. The patient also reported some mild side effects, including headaches and fatigue, which were tolerable and not significantly impacting his daily activities.

In light of the patient's ongoing participation in the randomized controlled trial, no changes will be made to his current medications other than adding the investigational drug for epilepsy. The patient will continue to take this medication for a period of three months, after which time we will reassess his seizure activity and adjust his treatment plan as necessary.

----------------END OF PATIENT ENCOUNTER---------------

-------------- PATIENT ID 10 -- VISIT 2 -----------

On August 20, 2022, I conducted a routine follow-up visit with Patient ID 10, a 30-year-old male who has been participating in a randomized controlled trial for the investigational drug ID X93461. The patient has a history of hypothyroidism and multifocal epilepsy. Over the past three months, the patient has taken the investigational drug and has reported experiencing 23.0 seizures during this time period. Notably, the patient has also reported feeling fatigued.

Upon conducting a complete neurological examination, I observed that the patient's cognitive function and memory appear to be intact. However, there was evidence of mild bilateral sensorimotor axial hyperreflexia, which may be indicative of the patient's multifocal epilepsy. The patient's speech was clear and coherent, with no evidence of dysarthria or aphasia. The patient's language skills were intact, and they were able to communicate effectively.

Given the results of the study and the patient's experience with the investigational drug, it has been determined that the drug will be discontinued. The patient will be transitioned back to their standard medication regimen for their epilepsy, and we will monitor their condition closely to assess the effectiveness of this change. Additionally, we will continue to monitor the patient's fatigue levels and adjust their treatment plan as needed to ensure their quality of life is maintained.

In conclusion, Patient ID 10 has completed their participation in the randomized controlled trial for the investigational drug ID X93461. The results of the study suggest that the drug may be effective in reducing seizure frequency, but further research is needed to confirm these findings. The patient will be transitioned back to their standard medication regimen, and we will continue to monitor their condition closely to ensure their ongoing health and well-being.

----------------END OF PATIENT ENCOUNTER---------------

-------------- PATIENT ID 11 -- VISIT 1 -----------

Patient Identification:

Name: John Doe

Date of Birth: 01/01/1992

Patient ID: 11

Gender: Male

Age: 29 years

Identification Number: 123456789

Highlights of Patient History:

Diabetes mellitus type I (since 2000)

Occipital lobe epilepsy (since 2010)

Seizures: 44 in the past two months

Randomized controlled trial participant (currently on investigational drug for epilepsy)

Medication Adherence:

Patient reports taking medications as prescribed, with no deviations or omissions.

Subjective:

Patient presents with a complaint of increased seizure frequency in the past two months. He mentions that he has experienced an average of 2-3 seizures per week, with no significant triggers or warning signs. The patient also reports no changes in his medication regimen, and no recent illnesses or injuries.

Medications:

Current medications:

1. Levetiracetam (Keppra) 500mg BID

2. Lamotrigine (Lamictal) 200mg OD

3. Investigational drug for epilepsy (IDEE-234) 500mg TID for the past three months as part of a randomized controlled trial.

Allergies:

No known allergies.

Social History:

Married with two children (ages 7 and 9)

Occupation: Software engineer

Lives with spouse and children in a suburban house

Review of Systems:

General: Denies fever, chills, or night sweats.

Neurological: Denies weakness, numbness, or paresthesias. Reports increased seizure activity over the past two months.

Psychiatric: No history of psychiatric disorders or hospitalizations.

Medication History:

No recent changes in medications.

Family and Social History:

No significant family medical history.

General Physical Examination:

Vital signs:

Temperature: 98.6°F (37°C)

Pulse: 70/min

Respiratory Rate: 12/min

Blood Pressure: 120/80 mmHg

Height: 5'10" (178 cm)

Weight: 160 lbs (72 kg)

Head, Eyes, Ears, Nose, and Throat (HEENT):

No abnormalities noted.

Comprehensive Neurological Examination:

Motor: No weakness or paralysis. Normal muscle tone and bulk.

Sensory: No sensory deficits or abnormalities.

Cerebellar: No ataxia or dysmetria noted.

Extrapyramidal: No signs of Parkinsonism or dystonia observed.

Psychiatric: No signs of depression, anxiety, or psychosis detected.

Labs:

Blood workup:

Glucose: 120 mg/dL (normal: 70-110 mg/dL)

HbA1c: 7.5% (normal: <6.5%)

Creatinine: 1.1 mg/dL (normal: 0.8-1.2 mg/dL)

Imaging:

CT scan of the head: No abnormalities noted.

Assessment:

The patient presents with an increased frequency of seizures over the past two months, despite no changes in medication regimen. The patient is currently participating in a randomized controlled trial for investigational drug for epilepsy. A complete neurological examination reveals no abnormalities. Laboratory results show mild hyperglycemia and mild renal impairment. The patient's history of diabetes and occipital lobe epilepsy, combined with the recent increase in seizure frequency, suggest a possible exacerbation of his condition.

Plan:

1. Continue the current medication regimen, including levetiracetam, lamotrigine, and investigational drug for epilepsy.

2. Monitor the patient's seizure frequency and adjust the medication regimen as needed.

3. Schedule a follow-up appointment in two months to reassess the patient's condition and review the results of any additional diagnostic tests or procedures.

4. Encourage the patient to maintain strict blood glucose control and monitor for any changes in renal function.

Signature:

[Your Name]

Date: 2021-06-30

----------------END OF PATIENT ENCOUNTER---------------

-------------- PATIENT ID 11 -- VISIT 2 -----------

Visit #2 - 2021-07-30

Patient ID: 11, Male, Age 29

History: Diabetes, Occipital Lobe Epilepsy (study participant)

Neurological Examination:

* Mental Status: Alert and oriented

* Cranial Nerves: Normal

* Motor Exam: Mild ataxia, gait disturbance

* Sensory Exam: Intact

* Reflexes: 2+ (normal)

Pertinent Positives:

* Dizziness

* Ataxia

* Gait disturbance

Assessment and Plan:

* Discontinue investigational drug ID X93461 due to completion of study

* Monitor for withdrawal seizures and adjust anticonvulsant medication as needed

* Refer patient to physical therapy for gait disturbances and ataxia

* Schedule follow-up appointment in 2 weeks to reassess neurological status.

----------------END OF PATIENT ENCOUNTER---------------

-------------- PATIENT ID 12 -- VISIT 1 -----------

Patient ID: 12

Visit #1 (2022-04-24)

* 87 year old female

* History of anxiety and parietal lobe epilepsy

* 6.0 seizures in past 2 months

* Randomized controlled trial (no change in medications except adding investigational drug for epilepsy)

Neurological Examination:

* Mental status: alert and oriented

* Speech: normal

* Motor exam: normal

* Sensory exam: normal

* Reflexes: normal

* Gait: normal

Pertinent Positives:

* No change in seizure frequency or severity

* Patient reports no side effects from current medications

Assessment and Plan:

* Continue current medications as prescribed

* Add investigational drug for epilepsy for 3 months

* Monitor patient closely for any changes in seizure frequency, severity, or side effects

* Follow up in 2 weeks to assess effectiveness of new medication and make any necessary adjustments.

----------------END OF PATIENT ENCOUNTER---------------

-------------- PATIENT ID 12 -- VISIT 2 -----------

---

Clinic Visit #2: May 24, 2022

Patient ID: 12

Age: 87 years

Gender: Female

History: Anxiety, parietal lobe epilepsy

Current Medications: None (previously took investigational drug ID X93461 for epilepsy)

Chief Complaint: Ataxia

Past Medical History: Parietal lobe epilepsy, anxiety

Social History: None

Family History: None

Review of Systems: None

General: The patient appears well-oriented and in no distress. Speech is clear and comprehensible.

Neurological Examination:

Motor:

* Muscle strength: 5/5 (normal)

* Muscle tone: 2/5 (increased)

* Reflexes: 2+ (normal)

* Coordination and balance: fair (ataxic)

Sensory:

* Visual: No visual field defects or abnormalities noted.

* Auditory: Normal hearing sensitivity and no abnormal sounds detected.

* Tactile: Intact sensation to light touch and pinprick.

* Proprioception: Fair (ataxic)

Cranial Nerves:

* Ophthalmologic: Pupils are symmetrical and react appropriately to light. Extraocular movements are intact.

* Facial: No weakness or paralysis of facial muscles.

* Auditory: No abnormalities detected.

* Glossopharyngeal: Normal strength and function.

* Vestibulocochlear: Normal hearing sensitivity and no abnormal sounds detected.

Extremities:

* Right upper limb: Normal motor strength and sensation.

* Left upper limb: Normal motor strength and sensation.

* Right lower limb: Fair motor strength, ataxic gait.

* Left lower limb: Fair motor strength, ataxic gait.

Assessment and Plan:

The patient has reported ataxia, which is consistent with the adverse event profile of ID X93461. As the patient has completed the randomized controlled trial and the investigational drug will be discontinued, the ataxia is likely related to the study medication. The patient's anxiety and parietal lobe epilepsy remain stable.

Plan:

* Continue regular follow-up appointments to monitor the patient's condition and address any concerns or questions.

* Consider referring the patient to physical therapy to improve balance and coordination.

* Monitor the patient's seizure activity and adjust medication as needed.

Disposition: The patient will be discharged home with a follow-up appointment scheduled in 2 weeks.

----------------END OF PATIENT ENCOUNTER---------------

-------------- PATIENT ID 13 -- VISIT 1 -----------

Patient ID: 13

Visit #1: 2021-10-03

History:

* Schizophrenia

* ADHD

* Epilepsy (11 seizures in past 2 months)

* Randomized controlled trial (no change in medications except for investigational drug for epilepsy)

Neurological Examination:

* Mental status: alert and oriented

* Speech: clear and logical

* Motor exam: normal strength and tone, no tremors or rigidity

* Sensory exam: intact

* Reflexes: normal

* Cranial nerves: normal

Pertinent Positives:

* 11 seizures in past 2 months

* No change in medications except for investigational drug for epilepsy

Assessment and Plan:

* Continue randomized controlled trial with no changes to medication regimen except for investigational drug for epilepsy

* Monitor seizure activity and adjust plan as needed

* Schedule follow-up visit in 3 months to assess efficacy of investigational drug and monitor for any adverse effects.

----------------END OF PATIENT ENCOUNTER---------------

-------------- PATIENT ID 13 -- VISIT 2 -----------

Clinic Note: Visit #2 - November 3, 2021

Patient ID: 13

Age: 64 years old

Gender: Male

Chief Complaint:

* History of schizophrenia, ADHD, and epilepsy

* Participating in a randomized controlled trial for investigational drug ID X93461 for epilepsy

History of Present Illness:

* Patient has been taking the investigational drug ID X93461 for epilepsy for the past 3 months

* Patient has experienced a total of 4.0 seizures since starting the medication

* Patient reported somnolence, fatigue, constipation, vomiting, and fall

Neurological Examination:

* Mental status: Alert and oriented to person, place, and time

* Speech: Clear and coherent

* Language: Intact

* Motor examination: No significant tremors or rigidity noted

* Sensory examination: No significant sensory deficits noted

* Cerebellar function: Normal

* Reflexes: 2+ (moderate)

* Muscle strength: 5/5 (normal)

* Gait: Normal

Assessment and Plan:

* The patient has completed the randomized controlled trial for investigational drug ID X93461 for epilepsy, and the medication will be discontinued due to the study being complete.

* The patient has experienced somnolence, fatigue, constipation, vomiting, and fall during the trial period, which may be related to the investigational drug.

* It is recommended that the patient follow up with their primary care physician for further management of these symptoms.

* The patient's schizophrenia, ADHD, and epilepsy should also be monitored closely and managed accordingly.

* No changes to the patient's current medications are recommended at this time.

Note: This clinic note is based on a fictional patient and is for educational purposes only. It is not intended to substitute professional medical advice, diagnosis, or treatment.

----------------END OF PATIENT ENCOUNTER---------------

-------------- PATIENT ID 14 -- VISIT 1 -----------

Patient ID: 14

Visit Date: 2021-06-18

History of Present Illness:

The patient is a 50-year-old male with a history of schizophrenia, bipolar disorder, and parietal lobe epilepsy. He has been experiencing an increased frequency of seizures over the past 2 months, with 13.0 seizures reported. The patient is currently enrolled in a randomized controlled trial (RCT) and is being treated with the investigational drug for epilepsy. No changes will be made to the patient's medication regimen other than the addition of the investigational drug for the next 3 months.

Neurological Examination:

Mental Status: The patient appears to be alert and oriented to person, place, and time. He has a clear and goal-directed speech, and his thought processes are logical and coherent. There is no evidence of hallucinations or delusions.

Cranial Nerve Examination:

* Visual acuity: 20/20 OU

* Extraocular movements: Full range of motion, without any abnormalities

* Pupils: Equal and reactive to light and accommodation

* Facial movements: Normal

* Hearing: Normal

* Tongue movements: Normal

Motor Examination:

* Strength: 5/5 bilaterally

* Reflexes: 2+ (moderate) bilaterally

* Coordination and balance: Normal

Sensory Examination:

* Somatosensory: Intact to light touch, pinprick, and temperature stimuli

* Visual: Intact to visual stimuli

* Auditory: Intact to auditory stimuli

Neuropsychological Assessment:

The patient's cognitive function appears to be within normal limits. He is able to concentrate and pay attention, and he demonstrates normal memory and executive function abilities. There is no evidence of deficits in language, visuospatial skills, or attention.

Assessment and Plan:

Based on the patient's history and current symptoms, it is diagnosed that the patient has an increased frequency of seizures due to parietal lobe epilepsy. The patient will continue to participate in the RCT and will be treated with the investigational drug for the next 3 months. No changes will be made to the patient's medication regimen other than the addition of the investigational drug. Close monitoring of seizure frequency and severity will be done during this time. Regular follow-up appointments will be scheduled to assess the patient's progress and make any necessary adjustments to the treatment plan.

Note: This is a sample clinic note for educational purposes only, and it should not be used as a substitute for professional medical advice, diagnosis, or treatment.

----------------END OF PATIENT ENCOUNTER---------------

-------------- PATIENT ID 14 -- VISIT 2 -----------

Clinic Note: Visit #2 - July 18, 2021

Patient ID: 14

Age: 50 years old

History: Schizophrenia, Bipolar Disorder, Parietal Lobe Epilepsy

Investigational Drug History: X93461 for epilepsy - last 3 months (discontinued)

Chief Complaint: Somnolence, fatigue, vomiting

Neurological Examination:

1. Mental Status: Alert and oriented x3. Speech is clear and fluent. No evidence of psychotic symptoms or cognitive impairment.

2. Motor Function: No tremors or rigidity noted. There is no evidence of weakness or paralysis on the right side of the body. The patient walks normally without any signs of ataxia or dysmetria.

3. Sensory Function: No sensory deficits were detected in the examination. The patient has normal hearing and vision.

4. Cranial Nerves: Pupils are equal, round, and reactive to light. Extraocular movements are intact. There is no evidence of abducens nerve weakness or oculomotor nerve dysfunction. The patient has normal facial expressions and can speak clearly.

5. Reflexes: Normoreflexia is noted in the upper and lower extremities.

6. Gait: Normal gait with no signs of ataxia, dysmetria, or weakness.

7. Muscle Strength: The patient has normal muscle strength in all four limbs.

8. Coordination: The patient performs normally on the finger-to-nose and heel-to-shin tests, with no signs of ataxia or dysmetria.

9. Balance: The patient maintains a steady balance during the examination.

10. Cerebellar Function: No evidence of cerebellar dysfunction is noted in the examination.

Assessment and Plan:

1. Somnolence, fatigue, and vomiting are likely related to the investigational drug X93461 and will resolve now that the medication has been discontinued.

2. The patient's schizophrenia, bipolar disorder, and parietal lobe epilepsy remain stable and under good control with current medications.

3. No changes are recommended to the patient's current medication regimen at this time.

4. Monitoring for seizure activity and adverse effects will be necessary in the coming weeks, given the recent discontinuation of the investigational drug.

5. The patient will follow up with the study team for further assessment of the drug's effectiveness and any potential long-term side effects.

6. Regular follow-up appointments with the neurologist will be necessary to monitor the patient's condition and address any concerns or complications that may arise.

Note: The patient has been informed of the study's completion and the discontinuation of the investigational drug. The patient understands the rationale for this decision and agrees with the plan moving forward.

----------------END OF PATIENT ENCOUNTER---------------

-------------- PATIENT ID 15 -- VISIT 1 -----------

Patient ID: 15

Visit #1: 2022-11-07

History:

* 21 year old female with history of epilepsy

* 43 seizures in past 2 months

* Currently enrolled in randomized controlled trial

Neurological Examination:

* Alert and oriented x3

* No focal neurological deficits

* Normal muscle strength and tone

* Normal sensation in all extremities

* No evidence of seizure activity

Pertinent Positives:

* History of frequent seizures (43 in past 2 months)

* Enrolled in randomized controlled trial

Assessment and Plan:

* Continue with current medication regimen

* Add investigational drug for epilepsy for 3 months

* Monitor seizure frequency and adverse effects

* Follow-up visit in 1 month

----------------END OF PATIENT ENCOUNTER---------------

-------------- PATIENT ID 15 -- VISIT 2 -----------

CLINIC NOTE VISIT #2 - DATE: 2022-12-07

Patient ID: 15

Age: 21 years old

Gender: Female

Chief Complaint: Epilepsy

History of Present Illness: The patient has been participating in a randomized controlled trial for the investigational drug ID X93461 for the treatment of epilepsy. The patient has been taking the drug for the past 3 months and has reported experiencing 76.0 seizures during this time period. The patient has also complained of dizziness and somnolence.

Neurological Examination:

* General: Alert and oriented x3

* Mental Status: Intact, no evidence of clouding or confusion

* Cranial Nerves: Normal

* Motor System: No noticeable tremors or rigidity

* Sensory System: No deficits in sensation to light touch, pinprick, or vibration

* Reflexes: 2+ (moderate)

* Coordination and Balance: Normal

Assessment:

The patient has been taking the investigational drug ID X93461 for epilepsy over the past 3 months and has reported experiencing a significant number of seizures during this time. The patient has also complained of dizziness and somnolence. Based on the patient's history and physical examination, it is determined that the investigational drug will be discontinued due to the completion of the study. The patient will be advised to seek alternative medication for the management of her epilepsy.

Plan:

1. Discontinue the investigational drug ID X93461.

2. Provide the patient with information on alternative medications for the management of her epilepsy.

3. Schedule a follow-up appointment to monitor the patient's condition and adjust the treatment plan as necessary.

----------------END OF PATIENT ENCOUNTER---------------

-------------- PATIENT ID 16 -- VISIT 1 -----------

On March 8th, 2022, Patient ID 16, a 25-year-old female with a history of autism, seasonal allergies, schizophrenia, and multifocal epilepsy presented for her first clinic visit. The patient has experienced 14 seizures in the past two months, which is a significant increase from her previous seizure frequency. As part of a randomized controlled trial, the patient will be adding an investigational drug for epilepsy to her current medication regimen for a period of three months.

During the clinic visit, a complete neurological examination was conducted. The patient's cognitive function was found to be within normal limits, and there were no signs of weakness or paralysis. The patient's speech was clear and comprehensible, with no apparent difficulty in language processing. The patient's sensory examination revealed normal hearing and vision, with no abnormalities detected.

The patient's seizure activity was also evaluated during the visit. The patient reported experiencing 14 seizures over the past two months, with no significant changes in frequency or severity. The patient's EEG results were within normal limits, and there was no evidence of any other neurological disorders.

Based on the results of the clinic visit, it is recommended that the patient continue with her current medication regimen, including the investigational drug for epilepsy, for a period of three months. The patient will be closely monitored during this time to assess the efficacy of the new medication and any potential side effects. Regular follow-up appointments will be scheduled to ensure that the patient's seizure activity and overall health remain stable.

----------------END OF PATIENT ENCOUNTER---------------

-------------- PATIENT ID 16 -- VISIT 2 -----------

Patient ID: 16

Visit #2: April 8, 2022

Chief Complaint: Balance disorder, multifocal epilepsy, seasonal allergies, autism

History of Present Illness:

The patient is a 25-year-old female with a history of autism, seasonal allergies, schizophrenia, and multifocal epilepsy. She has been participating in a randomized controlled trial and has been taking the investigational drug ID X93461 for the past 3 months. During this time, she has experienced 8.0 seizures. The patient has now completed the study and the investigational drug will be discontinued. She reports balance disorder and is concerned about her recent seizure activity.

Past Medical History:

* Autism

* Seasonal allergies

* Schizophrenia

* Multifocal epilepsy

Medication List:

* ID X93461 (investigational drug) - discontinued

* Levetiracetam (keppra) - ongoing

* Lamotrigine (lamictal) - ongoing

Social History:

* No tobacco or alcohol use

* Denies illicit drug use

* Lives independently with family support

Review of Systems:

* No fever or chills

* No recent travel or sick contacts

* No changes in appetite or weight

Neurological Examination:

General: Alert and oriented x3. Speech is clear and fluent.

Cranial Nerves:

* II - Pupils are equal, round, and reactive to light (2+/2+)

* III - Vertical gaze is maintained

* IV - Extreme lateral gaze is maintained

* VI - No abnormalities detected

Motor:

* Right and left arm and leg strength is 5/5

* No ataxia or tremors noted

Sensory:

* No sensory deficits detected

Balance:

* Patient reports difficulty with balance and recent falls

Assessment and Plan:

The patient's history of multifocal epilepsy, autism, and seasonal allergies, along with her recent participation in a randomized controlled trial and discontinuation of the investigational drug ID X93461, contribute to her balance disorder and seizure activity. Based on the patient's history and examination, I recommend the following:

1. Monitoring of seizure activity and adjustment of medications as needed to control seizures.

2. Physical therapy to address balance issues and prevent falls.

3. Follow-up appointments to monitor progress and adjust treatment plan as necessary.

I will follow up with the patient in 2 weeks to assess her condition and adjust her treatment plan accordingly.

Signature: [Your Name]

Date: April 8, 2022

----------------END OF PATIENT ENCOUNTER---------------

-------------- PATIENT ID 17 -- VISIT 1 -----------

Patient ID: 17

Date: 2021-03-22

HPI:

* 81-year-old male

* History of ADHD, schizophrenia, and frontal lobe epilepsy

* 221.0 seizures in the past 2 months

* Currently participating in a randomized controlled trial

Neurological Examination:

* Mental status: Alert and oriented

* Cranial nerves: Intact

* Motor exam: Normal

* Sensory exam: Intact

* Reflexes: 2+ (brisk)

* Gait: Normal

Pertinent Positives:

* No change in medications except for addition of investigational drug for epilepsy

* Seizure frequency has increased over the past 2 months (221.0 seizures)

Assessment and Plan:

* Continue with randomized controlled trial

* Add investigational drug for epilepsy

* Monitor seizure frequency and adjust medications as needed

* Schedule follow-up appointment in 3 months.

----------------END OF PATIENT ENCOUNTER---------------

-------------- PATIENT ID 17 -- VISIT 2 -----------

Visit #2, 2021-04-22

Patient ID: 17

Age: 81 y/o

Male

History: ADHD, schizophrenia, frontal lobe epilepsy

Randomized controlled trial (ID X93461)

Current Medications:

* Investigational drug ID X93461 (discontinued)

Pertinent Positives:

* 128.0 seizures over past 3 months

* Back pain

Neurological Examination:

* Mild weakness in left arm and leg

* Hyperreflexia

* Sensory deficits in L2, L3, and L4 dermatomes

* Difficulty with fine motor skills

Assessment:

* Discontinue investigational drug ID X93461

* Manage back pain with non-steroidal anti-inflammatory drugs (NSAIDs) as needed

* Monitor for seizure activity and adjust medications as needed

Plan:

* Follow-up visit in 2 weeks to assess seizure control and adjust medications as needed

* Consider referral to physical therapy for fine motor skill difficulties

----------------END OF PATIENT ENCOUNTER---------------

-------------- PATIENT ID 18 -- VISIT 1 -----------

Clinic Note - Visit #1

Date: 2021-05-10

Patient ID: 18

Chief Complaint: The patient presents with a history of ADHD, hypothyroidism, autism, and generalized epilepsy. He has experienced 11.0 seizures in the past 2 months.

History of Present Illness: The patient is an 82-year-old male who has been diagnosed with ADHD, hypothyroidism, autism, and generalized epilepsy. He has been experiencing increased seizure activity over the past 2 months, with 11.0 seizures reported. He is currently participating in a randomized controlled trial and will be taking the investigational drug for epilepsy for the next 3 months.

Past Medical History: The patient has a history of ADHD, hypothyroidism, autism, and generalized epilepsy. He has experienced seizures throughout his life, with the most recent one occurring 2 months ago. He has been treated with various medications, including anticonvulsants and mood stabilizers, but has not achieved complete seizure control.

Medication List: The patient is currently taking levothyroxine (T4) for hypothyroidism, methylphenidate for ADHD, and lamotrigine for epilepsy. He is also taking risperidone for autism-related behavioral issues.

Allergies: The patient has no known allergies.

Social History: The patient lives with his wife and adult child. He reports no significant social or occupational changes recently. He denies alcohol, tobacco, or illicit substance use.

Review of Systems: The patient reports no recent changes in sleep, appetite, or mood. He denies any joint pain or swelling. He has experienced some difficulty with memory and concentration, but this has been ongoing for several years.

Physical Examination:

Vital Signs:

* Blood pressure: 120/80 mmHg

* Pulse: 75 beats per minute

* Temperature: 98.6 degrees Fahrenheit

* Weight: 160 lbs

* Height: 5'10"

Neurological Examination:

* Mental status: The patient is alert and oriented, but he appears somewhat lethargic and has difficulty concentrating.

* Cranial nerves: All cranial nerves are intact, with no abnormalities detected.

* Motor examination: The patient exhibits a slight tremor in both hands, and his finger-to-nose test is positive on the right side. His strength, tone, and reflexes are normal.

* Sensory examination: The patient has normal sensation to light touch, pinprick, and temperature. He does not report any paresthesias or numbness.

* Coordination and balance: The patient exhibits a slight unsteadiness while walking, but he is able to maintain his balance without assistance.

Assessment and Plan:

The patient's history of ADHD, hypothyroidism, autism, and generalized epilepsy, along with his recent increase in seizure activity, suggest a complex neurological disorder that may be challenging to treat. The patient is currently participating in a randomized controlled trial, which limits the ability to adjust his medications at this time. Therefore, the plan will be to continue the patient's current medication regimen and closely monitor his seizure activity over the next 3 months. The patient will be advised to return for follow-up visits every 2 weeks to assess his seizure control and monitor any potential side effects of the investigational drug. Additionally, the patient's memory and concentration difficulties will be addressed with cognitive behavioral therapy and other supportive measures as needed.

During this visit, the patient reported no significant changes in his symptoms or medication side effects. He was advised to continue taking his medications as prescribed and to return for follow-up visits as scheduled. The patient's next appointment is set for 2021-05-24.

----------------END OF PATIENT ENCOUNTER---------------

-------------- PATIENT ID 18 -- VISIT 2 -----------

Clinic Note - Visit #2

Patient ID: 18

Date: 2021-06-10

History of Present Illness:

The patient is an 82-year-old male with a history of ADHD, hypothyroidism, autism, and generalized epilepsy. He has been participating in a randomized controlled trial (RCT) evaluating the safety and efficacy of investigational drug ID X93461 for the treatment of his epilepsy. The patient has been taking the drug for the past 3 months and has reported experiencing 9.0 seizures during this time. He has also reported symptoms of dizziness, somnolence, fall, and gait disturbance.

Review of Systems:

The patient reports no significant changes in his medical history since his last visit. He denies any recent travel or exposure to medications or substances that could be contributing to his symptoms. He also denies any new allergies or sensitivities.

Neurological Examination:

General appearance: The patient appears alert and oriented x3.

Head: Normal head circumference, normocephalic.

Eyes: Pupils are equally reactive to light and accommodation, with no evidence of afferent pupillary defect or abnormal eye movements. Extraocular muscles are intact.

Ears: No ear discharge or odor. No mass or lesion is palpable in the ears. Hearing is normal to speech and whisper.

Nose: No nasal discharge or bleeding. No masses or lesions are visible on the nasal cavity.

Mouth: Normal dentition and oral hygiene. No evidence of tremors or ataxia.

Neck: No mass or lesion is palpable in the neck. No cervical spine tenderness to palpation.

Extremities: No weakness or paralysis is evident in the upper or lower extremities. No sensory deficits are noted. No tremors or ataxia are observed.

Gait: The patient ambulates with a normal gait, without any signs of instability or imbalance.

Sensory: No significant sensory deficits or abnormalities are noted.

Cerebellar function: No evidence of cerebellar dysfunction is observed.

Mental Status: The patient appears alert and oriented x3. His speech is clear and coherent, with no evidence of aphasia or dysarthria. His thought process is logical and goal-directed.

Assessment:

The patient's symptoms of dizziness, somnolence, fall, and gait disturbance are likely related to the investigational drug ID X93461, which will be discontinued now that the study is complete. The patient's history of ADHD, hypothyroidism, autism, and generalized epilepsy may also contribute to these symptoms.

Plan:

1. Discontinue investigational drug ID X93461.

2. Monitor the patient's seizure activity and symptoms closely in the coming weeks.

3. Consider alternative therapies for the patient's epilepsy, as needed.

4. Follow up with the patient in 1-2 weeks to assess for any changes in his symptoms and to monitor the effectiveness of any interventions implemented.

Note: The patient's participation in the RCT is being discontinued, but he will continue to receive regular follow-up care for his epilepsy and other comorbid conditions.

----------------END OF PATIENT ENCOUNTER---------------

-------------- PATIENT ID 19 -- VISIT 1 -----------

On December 12th, 2021, I conducted a clinic visit with Patient ID 19, a 31-year-old male with a history of schizophrenia, dermatitis, and temporal lobe epilepsy. The patient has had 21.0 seizures in the past two months, and is currently participating in a randomized controlled trial. The purpose of this visit was to assess the patient's condition and monitor his progress in the trial.

Upon examination, the patient appeared alert and oriented, with normal speech and thought processes. His mood was euthymic, and he did not report any significant changes in his symptoms since the last visit. The patient's dermatitis appeared mild, with only a few scattered papules and plaques on his face and arms. His temporal lobe epilepsy was stable, with no recent seizures reported.

I performed a complete neurological examination, including an assessment of the patient's cognitive function, sensory systems, and motor function. The patient's cognitive function was within normal limits, and he demonstrated intact memory and attention. His sensory systems were also normal, with no deficits in vision, hearing, or sensation. The patient's motor function was slightly weakened, but this was attributed to his history of temporal lobe epilepsy.

In light of the patient's history and current condition, I have decided to continue the investigational drug for epilepsy for an additional three months. This medication has been effective in controlling the patient's seizures, and it is important to continue it in order to maintain his stability. The patient will be monitored closely throughout the course of the trial, with regular follow-up visits to assess his progress and adjust the treatment as needed.

----------------END OF PATIENT ENCOUNTER---------------

-------------- PATIENT ID 19 -- VISIT 2 -----------

Visit #2, dated 2022-01-12, for Patient ID 19, a 31-year-old male with a history of schizophrenia, dermatitis, and temporal lobe epilepsy. The patient has been participating in a randomized controlled trial (RCT) evaluating the investigational drug ID X93461 for the treatment of epilepsy. Over the past three months, the patient has taken the medication and has reported experiencing 19.0 seizures during this time. Additionally, the patient has noticed fatigue and diplopia (double vision).

Upon examination, the patient's neurological status was assessed. The patient was alert and oriented, with a normal mood and affect. The patient's speech was clear and coherent, with no evidence of any language or cognitive impairment. The patient's motor function was normal, with no signs of weakness, ataxia, or tremors. The patient's sensory examination revealed no abnormalities in vision, hearing, or somatosensory function.

The patient's RCT medication, ID X93461, will be discontinued now that the study is complete. The patient has been advised to continue with their regular anticonvulsant medication and to follow-up with their primary care physician for any further medical needs. The patient has been instructed to contact the study team if they experience any further seizures or any other concerning symptoms.

In conclusion, this clinic note documents the patient's second visit during the RCT evaluation of ID X93461 for the treatment of epilepsy. The patient reported a moderate number of seizures during the study period and experienced some mild side effects. However, the patient's overall neurological status remains stable, and the medication will be discontinued as planned. The patient has been advised to follow-up with their primary care physician for any further medical needs.

----------------END OF PATIENT ENCOUNTER---------------

-------------- PATIENT ID 30 -- VISIT 1 -----------

On May 22nd, 2021, Patient ID 30, a 92-year-old female, presented to our clinic for her first visit. The patient has a history of seasonal allergies, dermatitis, autism, and parietal lobe epilepsy, and has experienced 6.0 seizures in the past two months. As part of a randomized controlled trial, the patient will be taking an investigational drug for epilepsy for the next three months, with no changes made to her current medications.

Upon examination, the patient's neurological status was as follows: General cognitive function and awareness were normal, with intact memory and attention. The patient's speech was clear and coherent, with normal tone and volume. There were no notable deficits in visual fields or extraocular movements. The patient had a normal gait and station, with no signs of ataxia or tremors.

The patient's dermatitis was noted to be mild, with only slight redness and flaking observed on the skin. No new lesions were seen, and the patient reported no increased itching or discomfort. The patient's seasonal allergies were also mild, with only minor symptoms of sneezing and congestion noted.

The patient's autism was well-controlled, with no significant changes in behavior or mood observed since the previous visit. The patient's parietal lobe epilepsy remained stable, with no new seizures reported since the last visit. The patient will continue to be monitored closely throughout the course of the randomized controlled trial, with regular follow-up appointments scheduled to assess the effectiveness of the investigational drug and any potential side effects or complications.

----------------END OF PATIENT ENCOUNTER---------------

-------------- PATIENT ID 30 -- VISIT 2 -----------

Clinic Note #2

Date: 2021-06-22

Patient ID: 30

Age: 92 years old

Gender: Female

Chief Complaint: Seasonal allergies, dermatitis, autism, and parietal lobe epilepsy

History of Present Illness: The patient has been participating in a randomized controlled trial for the investigational drug ID X93461 for epilepsy over the past 3 months. She has reported experiencing 6.0 seizures during this time, as well as dizziness and somnolence. The investigational drug will be discontinued now that the study is complete.

Past Medical History: Seasonal allergies, dermatitis, autism, and parietal lobe epilepsy

Medications: None currently

Social History: None relevant

Family History: None relevant

Review of Systems: The patient reports no significant changes in her symptoms since the last clinic visit. She continues to experience seasonal allergies and dermatitis, which are well-controlled with current medications. Her autism and epilepsy remain stable.

Neurological Examination:

General: The patient appears alert and in no distress.

Cranial Nerves:

* Visual: 20/20 in both eyes, with full fields of vision

* Auditory: Normal hearing sensitivity and discrimination

* Olfactory: Intact sense of smell

* Taste: Intact sense of taste

* Motor: No weakness or atrophy noted in the limbs

* Sensory: Intact sensation to light touch, pinprick, and temperature

* Reflexes: Normal reflexes with no hyporeflexia or hyperreflexia noted

Motor Examination:

* Right-sided weakness: None noted

* Left-sided weakness: None noted

* Muscle strength: 5/5 in all muscle groups

* Muscle tone: Within normal limits

Gait and Station: The patient ambulates independently with a normal gait and station.

Dementia Assessment: No evidence of dementia or cognitive decline noted.

Assessment and Plan:

Based on the patient's history and current symptoms, it is my assessment that the patient has seasonal allergies, dermatitis, autism, and parietal lobe epilepsy. The patient has experienced a slight increase in seizure activity since initiating the investigational drug ID X93461 for epilepsy, but this is likely due to the drug's discontinuation. I recommend continuing current medications for seasonal allergies and dermatitis, as well as ongoing monitoring of seizure activity and adjustments to medications as needed. The patient will be followed up in clinic every 3 months for continued assessment and management of her condition.

Signature: [Your Signature]

Date: [Today's Date]

----------------END OF PATIENT ENCOUNTER---------------

-------------- PATIENT ID 31 -- VISIT 1 -----------

Patient ID: 31, a 77-year-old male, presented to our clinic on 2022-07-18 for his first visit. The patient has a significant medical history, including asthma and epilepsy. Recently, the patient has experienced an increase in seizure activity, with 8.0 seizures occurring over the past two months.

Upon examination, the patient's neurological status was assessed using a complete neurological examination. The patient's consciousness and alertness were normal, with no evidence of altered mental status or confusion. His speech was clear and comprehensible, with no signs of dysarthria or slurring. The patient's motor function was normal, with no signs of weakness, ataxia, or tremors. The patient's sensory examination revealed intact sensation in all modalities, including vision, hearing, and somatosensory.

The patient's medical history, including his diagnosis of asthma and epilepsy, was reviewed, and the patient reported no changes in his symptoms or medications since his last visit. The patient is currently enrolled in a randomized controlled trial (RCT) investigating an investigational drug for the treatment of epilepsy. As part of the RCT, the patient will be taking the investigational drug for the next three months. No changes in the patient's current medications will be made during this time period.

The patient's seizure activity was discussed with the patient and his caregiver, and they were instructed to continue recording any seizures that occur and to contact the clinic immediately if there are any changes in the patient's condition. The patient was advised to follow up with the clinic in three months for a status check on his seizure activity and to assess the effectiveness of the investigational drug.

----------------END OF PATIENT ENCOUNTER---------------

-------------- PATIENT ID 31 -- VISIT 2 -----------

Clinic Visit #2

Patient ID: 31

Date: 2022-08-18

History of Present Illness:

The patient is a 77-year-old male with a history of asthma and epilepsy. He has been participating in a randomized controlled trial for the investigational drug ID X93461 for his epilepsy over the past 3 months. During this time, he has experienced 7.0 seizures, and has reported dizziness. The study is now complete, and the investigational drug will be discontinued.

Neurological Examination:

Mental Status: The patient is alert and oriented to person, place, and time. He is able to speak in full sentences and answers questions appropriately.

General Examination:

The patient appears well-nourished and well- hydrated. His vital signs are as follows: blood pressure 120/80 mmHg, heart rate 70 beats per minute, temperature 98.6 degrees Fahrenheit. He has a normal appearance of his skin, hair, and nails. His neck is supple, and there is no evidence of mass lesions or edema.

Systemic Examination:

The patient's respiratory examination reveals clear lungs, with no wheezing or rhonchi. His cardiovascular examination shows a regular rate and rhythm, with no murmurs or abnormal sounds. His abdominal examination is unremarkable, with no tenderness or guarding. His extremities show normal strength and tone.

Neurological Examination:

The patient's neurological examination reveals the following:

* Cranial Nerves: The patient's cranial nerves are intact, with no abnormal findings.

* Motor System: The patient has normal muscle strength and tone, with no evidence of weakness or atrophy.

* Sensory System: The patient has intact sensation to light touch, pinprick, and temperature. There is no evidence of numbness or paresthesias.

* Coordination and Gait: The patient's coordination and gait are normal, with no evidence of ataxia or dysmetria.

* Reflexes: The patient's reflexes are normal, with no evidence of hyporeflexia or hyperreflexia.

Assessment and Plan:

Based on the patient's history of epilepsy and his recent experience of 7.0 seizures during the investigational drug trial, it is clear that he has a persistent seizure disorder. His reported dizziness may be related to the medication or his underlying condition. Discontinuing the investigational drug will likely lead to a decrease in seizure frequency, but the patient's seizure control should be closely monitored.

The patient's asthma should also be monitored and managed appropriately. His current respiratory examination is reassuring, but his history of asthma suggests that he may require ongoing therapy to prevent exacerbations.

The patient will be scheduled for follow-up appointments to monitor his seizure control and asthma management. He will also be referred to an appropriate specialist for ongoing care and management.

Keywords: epilepsy, seizures, dizziness, asthma, investigational drug, randomized controlled trial, discontinue medication.

----------------END OF PATIENT ENCOUNTER---------------

-------------- PATIENT ID 32 -- VISIT 1 -----------

CLINIC NOTE #1

Date: 2022-01-01

Patient ID: 32

Age: 76 years

Gender: Male

Chief Complaint: The patient presents with a history of asthma and epilepsy, with an increase in seizure frequency over the past two months (6.0 seizures).

Past Medical History: The patient has been diagnosed with asthma and epilepsy for the past 10 years. He has been under treatment with inhalers and oral medications, respectively. The patient has been stable on these medications until the recent increase in seizures.

Medication List: The patient is currently taking the following medications:

* Inhaler (fluticasone propionate/salmeterol xinafoate) twice daily

* Levetiracetam 1000mg PO QID for epilepsy

* Lorazepam 1mg PO BID as needed for seizures

Social History: The patient is a retired accountant, married with two adult children. He denies any recent travel, sick contacts, or alcohol/drug use. The patient reports that he has been experiencing more frequent seizures over the past two months, which have been associated with an increase in stress due to work-related reasons.

Family History: The patient's family history is unremarkable for neurological conditions.

Review of Systems: The patient reports no other symptoms or complaints other than the increase in seizures.

Physical Examination:

* General: The patient appears well-nourished and well-developed, with no signs of distress or discomfort.

* Neurological:

+ Cranial nerves: All cranial nerves are intact, with no signs of weakness or paresis.

+ Motor: There is no evidence of motor weakness or atrophy.

+ Sensory: The patient reports no sensory complaints or abnormalities.

+ Coordination and balance: The patient has normal coordination and balance.

* Cardiovascular: The patient has a regular rhythm, with a pulse rate of 70 beats per minute and blood pressure of 120/80 mmHg.

* Respiratory: The patient has a clear lungs, with no signs of distress or wheezing.

* Abdominal: The patient has a soft, normal-sized abdomen, with no tenderness or masses palpable.

Assessment and Plan:

* The patient's increase in seizures over the past two months is concerning and warrants further evaluation.

* The patient will continue to participate in the randomized controlled trial for epilepsy, with the addition of the investigational drug for the next 3 months.

* The patient will be monitored closely for any changes in seizure frequency or severity, and the effects of the investigational drug on seizure control will be evaluated.

* The patient will be advised to maintain a consistent regimen of inhalers and oral medications for asthma and epilepsy, respectively.

* The patient will be encouraged to keep a seizure diary to track the frequency and severity of seizures, and to report any changes or concerns to the clinic immediately.

Note: This is a fictional patient and not based on any real patient.

----------------END OF PATIENT ENCOUNTER---------------

-------------- PATIENT ID 32 -- VISIT 2 -----------

Visit #2 (2022-02-01)

Patient ID: 32

Age: 76 years

History: Asthma, epilepsy (randomized controlled trial participant)

Investigational Drug: ID X93461 (epilepsy)

Dizziness reported

Seizure Count: 9.0 in past 3 months (while taking investigational drug)

Neurological Examination:

* Mental Status: Alert and oriented

* Cranial Nerves: Normal

* Motor Examination: No abnormalities detected

* Sensory Examination: No abnormalities detected

* Coordination and Balance: Normal

* Reflexes: Normal

Pertinent Positives:

* Dizziness

* Increased seizure frequency while taking investigational drug

Assessment and Plan:

* Discontinue investigational drug due to increased seizure frequency and dizziness

* Monitor for any changes in seizure frequency or new symptoms

* Consider alternative treatments for epilepsy as needed

----------------END OF PATIENT ENCOUNTER---------------

-------------- PATIENT ID 33 -- VISIT 1 -----------

Clinic Note Visit #1

Patient ID: 33

Date: 2021-06-07

Chief Complaint:

The patient presents with a history of hypertension and generalized epilepsy. He has experienced 22.0 seizures in the past two months. He is currently participating in a randomized controlled trial evaluating an investigational drug for the treatment of epilepsy. The patient has not experienced any changes in medications other than the addition of the investigational drug, which he will take for the next three months.

Past Medical History:

The patient has a history of hypertension and generalized epilepsy. He has been previously treated with levetiracetam and valproic acid without significant improvement in seizure control.

Current Medications:

The patient is currently taking levetiracetam 500mg twice daily and valproic acid 2000mg twice daily. He has been added to the investigational drug, which he will take for the next three months.

Social History:

The patient lives alone and reports no significant changes in his social support system or living situation. He denies any recent travel or exposure to medications or substances that may have triggered his seizures.

Review of Systems:

The patient reports no recent changes in his sleep patterns, appetite, or mood. He has experienced some recent fatigue, which he attributes to the increasing frequency of his seizures.

Physical Examination:

Vital Signs:

* Temperature: 98.6°F (37°C)

* Pulse: 70/min

* Blood Pressure: 140/90 mmHg

* Respiration Rate: 16/min

Neurological Examination:

The patient is alert and oriented to person, place, and time. He has a normal gross motor examination with no signs of weakness or ataxia. His fine motor examination reveals a mild tremor in his hands, which is more prominent on the right side. The patient has a normal sensory examination, including visual and auditory functions. His cranial nerve examination is normal, except for a mild weakness of the left gaze.

The patient's speech is clear and comprehensible, with no signs of dysarthria or aphasia. His language function is intact, with no signs of difficulty with understanding or producing language. The patient has a normal gait and station, with no signs of ataxia or weakness.

Assessment:

Based on the patient's history and physical examination, he is diagnosed with hypertension and generalized epilepsy. He is currently experiencing an increased frequency of seizures, which may be related to his participation in the randomized controlled trial evaluating an investigational drug for epilepsy. The patient's neurological examination reveals a mild tremor in his hands and a weakness of the left gaze, which may be related to his seizure activity.

Plan:

The patient will continue to participate in the randomized controlled trial evaluating an investigational drug for epilepsy. He will take the medication as directed by the study team. The patient's seizure activity will be closely monitored, and any changes in his medications will be made based on the results of the study. The patient will be seen for follow-up visits every two months to assess his seizure control and any adverse effects of the medication.

The patient's hypertension will also be closely monitored, and any changes in his blood pressure medications will be made as needed. The patient will be encouraged to maintain a regular exercise program and a healthy diet to help manage his hypertension and seizure activity.

The patient will be advised to avoid any activities that may trigger his seizures, such as driving or operating heavy machinery. He will also be advised to carry a medical ID card and to seek immediate medical attention if he experiences any signs of seizure activity or other adverse effects of the medication.

The patient will be referred to physical therapy to address his mild tremor and weakness of the left gaze, as well as to improve his overall balance and coordination. The patient will also be referred to speech therapy to address any potential dysarthria or aphasia.

The patient will be seen for follow-up visits every two months to assess his seizure control, hypertension management, and any adverse effects of the medication. Any changes in his medications or study protocol will be made as needed based on the results of these follow-up visits.

----------------END OF PATIENT ENCOUNTER---------------

-------------- PATIENT ID 33 -- VISIT 2 -----------

Clinic Note - Visit #2 dated 2021-07-07 for Patient ID 33

Patient Summary:

This is a 98-year-old male patient with a history of hypertension and generalized epilepsy. The patient has been participating in a randomized controlled trial (RCT) evaluating the safety and efficacy of investigational drug ID X93461 for the treatment of epilepsy.

Neurological Examination:

On today's visit, the patient reported experiencing 12.0 seizures over the past three months since initiating treatment with ID X93461. The patient also complained of dizziness and headaches, as well as diplopia (double vision). These symptoms were mild to moderate in severity.

On examination, the patient's vital signs were within normal limits. There was no evidence of focal neurological deficits or sensory loss. The patient had a stable gait and there were no signs of ataxia or dysmetria. However, the patient did exhibit mild diplopia when attempting to focus on near objects.

The results of the neurological examination suggest that the investigational drug ID X93461 has been associated with some adverse effects, including seizures and visual disturbances. These findings are consistent with previous reports of the drug's efficacy in reducing seizure frequency but also causing mild to moderate side effects such as dizziness and diplopia.

Course of Treatment:

Given the results of the neurological examination and the patient's history of hypertension and generalized epilepsy, it has been determined that the investigational drug ID X93461 will be discontinued. The study has now been completed and the patient will be transitioned back to standard care for their condition. The patient will be monitored closely over the coming weeks to ensure a smooth transition and to address any residual effects of the investigational drug.

----------------END OF PATIENT ENCOUNTER---------------

-------------- PATIENT ID 34 -- VISIT 1 -----------

On April 18th, 2021, I had the pleasure of conducting a clinic visit with Patient ID 34, an 80-year-old female who has been grappling with depression, schizophrenia, hypertension, and temporal lobe epilepsy for quite some time now. As per her medical history, the patient has experienced 22.0 seizures over the past two months alone, a concerning trend that warranted further evaluation and management.

Upon entering the examination room, I was struck by the patient's frail appearance and soft-spoken demeanor. Her speech was clear and concise, but her voice lacked the vigor and confidence one would expect from someone of her age and background. As I began my examination, I noticed that the patient had difficulty maintaining eye contact, often looking down or away when engaged in conversation. This could be attributed to her history of schizophrenia and depression, which can manifest in social anxiety and decreased self-esteem.

As part of the neurological examination, I conducted a thorough inspection of the patient's head and neck, paying close attention to any abnormalities or asymmetries. Her cranial nerves were intact, with no signs of weakness or paresis. The patient's tone was normal, with no evidence of rigidity or spasticity. I also conducted a series of cognitive tests to assess her memory, attention, and executive function, which revealed a few areas of concern that will need to be monitored closely over the coming weeks and months.

In light of the patient's history and recent seizure activity, I have decided to add the investigational drug for epilepsy to her treatment plan for the next three months. This decision was made after careful consideration of the potential risks and benefits, as well as consultation with the patient's primary care physician and specialist colleagues. As part of this trial, I will closely monitor the patient's seizure activity, cognitive function, and overall well-being to assess the efficacy and safety of the investigational drug. No changes will be made to her existing medications at this time, except for the addition of the new drug as mentioned above. The patient was informed of the rationale behind this decision, and she expressed her willingness to participate in the trial and adhere to the prescribed treatment regimen.

----------------END OF PATIENT ENCOUNTER---------------

-------------- PATIENT ID 34 -- VISIT 2 -----------

Clinic Note, Visit #2, Date: 2021-05-18

Patient ID: 34, Female, 80 years old

Chief Complaint: The patient is an 80-year-old female with a history of depression, schizophrenia, hypertension, and temporal lobe epilepsy. She is currently participating in a randomized controlled trial for the investigational drug ID X93461 for the treatment of her epilepsy.

History of Present Illness: The patient has been taking the investigational drug ID X93461 for the past three months and has experienced 12.0 seizures during this time. She has also reported symptoms of dizziness, somnolence, and dysarthria.

Neurological Examination:

The patient's neurological examination reveals the following:

* Mental Status: The patient is alert and oriented to person, place, and time. Her speech is clear and her thought process is logical.

* Motor Examination: The patient's motor function is normal, with no signs of weakness, atrophy, or tremors.

* Sensory Examination: The patient's sensory examination reveals decreased sensation to light touch and pinprick sensation on the right side of her body.

* Cranial Nerve Examination: The patient's cranial nerve examination is normal, with no signs of abnormality.

* Reflexes: The patient's reflexes are normal, with no signs of hyperreflexia or hyporeflexia.

Plan:

1. Discontinue the investigational drug ID X93461, as the study is now complete.

2. Monitor the patient's seizure activity and adjust her medication regimen as needed.

3. Reassess the patient's neurological examination in six months to determine if there have been any changes in her condition.

4. Follow up with the patient's primary care physician to address any other medical concerns.

Note: This clinic note is a fictional example and should not be used as a basis for actual patient care. It is meant only to illustrate the style of a clinic note in the narrative form.

----------------END OF PATIENT ENCOUNTER---------------

-------------- PATIENT ID 35 -- VISIT 1 -----------

On July 22nd, 2021, I had the pleasure of conducting an initial clinic visit with Patient ID 35, a 35-year-old male who presented with a history of Attention Deficit Hyperactivity Disorder (ADHD) and temporal lobe epilepsy. The patient has been experiencing 14.0 seizures over the past two months, which is a significant increase from his previous seizure frequency. As part of a randomized controlled trial, we have enrolled the patient to evaluate the efficacy of an investigational drug for epilepsy, with no changes made to his current medications other than adding the study drug for a period of three months.

During the examination, I observed that the patient's vital signs were within normal limits, with a blood pressure of 120/80 mmHg, a heart rate of 75 beats per minute, and a temperature of 98.6 degrees Fahrenheit. His mental status was alert and focused, with no evidence of clouding or confusion. I conducted a thorough neurological examination, which revealed the following findings:

* Cranial nerves: The patient's cranial nerves were intact, with normal visual fields, hearing, and facial movements.

* Motor function: The patient had full range of motion in all extremities, with no evidence of weakness or ataxia.

* Sensory examination: The patient demonstrated normal sensation to light touch, pinprick, and temperature on all extremities. However, he did report some numbness in his left hand, which was not present during the previous examination.

* Coordination and balance: The patient's coordination and balance were intact, with no signs of ataxia or dizziness.

Based on the patient's history, physical examination findings, and current medication regimen, I have made the following diagnoses:

* ADHD (attention deficit hyperactivity disorder)

* Temporal lobe epilepsy

* Increased seizure frequency over the past two months

I have recommended that the patient continue his current medication regimen and participate in the randomized controlled trial to evaluate the efficacy of the investigational drug for epilepsy. I will closely monitor the patient's seizure frequency, cognitive function, and overall well-being during the three-month treatment period. Additionally, I have scheduled a follow-up visit for August 20th, 2021, to assess the patient's progress and make any necessary adjustments to his treatment plan.

----------------END OF PATIENT ENCOUNTER---------------

-------------- PATIENT ID 35 -- VISIT 2 -----------

Clinic Note: Visit #2 - Patient ID 35

Date: 2021-08-22

Identification:

Patient Name: Patient ID 35 (Mr./Ms./Mrs./Dr. [Last Name])

DOB: [Date of Birth]

Gender: Male

Age: 35 years old

HPI (History of Present Illness):

Patient presents with a history of ADHD and temporal lobe epilepsy. He has been participating in a randomized controlled trial (RCT) for the investigational drug ID X93461 for the past 3 months. The patient has reported experiencing somnolence and diplopia during this time period.

Subjective:

Patient reports that he has experienced a decrease in seizure frequency since starting the investigational drug, with only 10.0 seizures in the past 3 months. However, he also reports feeling excessively sleepy and experiencing double vision (diplopia). He denies any other symptoms or concerns.

Medications:

ID X93461 (investigational drug) - discontinued due to study completion

Vyvanse (lisdexamfetamine dimesylate) - ADHD medication (currently held due to patient's participation in RCT)

Allergies:

None reported

Social History:

Patient lives with his spouse and two children. He works as an engineer and reports no significant stressors or changes in his life recently. He denies any history of substance abuse or psychiatric illness.

Review of Systems:

General: Denies fever, chills, or weight loss. Reports somnolence and double vision (diplopia).

Neurological: Denies any recent headaches, weakness, or numbness/tingling. Reports continued seizure activity despite medication adherence.

Past Medical History: ADHD, temporal lobe epilepsy

Medication Adherence: Patient reports good adherence to medications, including ID X93461 and Vyvanse, during the past 3 months.

Labs:

None ordered at this time

Imaging: None ordered at this time

Assessment:

The patient presents with a history of ADHD and temporal lobe epilepsy, and has been participating in an RCT for the investigational drug ID X93461. Despite reporting somnolence and diplopia, the patient has experienced a decrease in seizure frequency during the past 3 months. However, given the patient's history of ADHD and temporal lobe epilepsy, as well as his current symptoms of somnolence and diplopia, it is possible that these symptoms may be related to the investigational drug or another underlying condition.

Plan:

1. Discontinue ID X93461 due to study completion.

2. Re-initiate Vyvanse (lisdexamfetamine dimesylate) for ADHD medication.

3. Monitor patient's symptoms and seizure activity closely, with regular follow-up appointments scheduled every 3 months.

4. Consider referral to an ophthalmologist for further evaluation of diplopia.

5. Educate patient on potential side effects of ADHD medications and the importance of adherence to medication regimen.

Signature: [Your Signature]

Date: [Today's Date]

Note: This clinic note is a fictional example for illustrative purposes only, and should not be used as a basis for actual patient care.

----------------END OF PATIENT ENCOUNTER---------------

-------------- PATIENT ID 36 -- VISIT 1 -----------

**Neurology Clinic Note - Visit #1 (2022-05-19) - Patient ID: 36, Age: 87 years, Male**

Chief Complaint:

The patient presents with a history of bipolar disorder and parietal lobe epilepsy. He has had 27.0 seizures in the past 2 months and is currently enrolled in a randomized controlled trial. The patient reports no changes in medications other than the addition of the investigational drug for epilepsy, which he will take for 3 months.

Past Medical History:

The patient has a history of bipolar disorder and parietal lobe epilepsy. He has been treated with lamotrigine, valproate, and levetiracetam in the past. He has experienced 27.0 seizures in the past 2 months.

Current Medications:

The patient is currently taking lamotrigine, valproate, and levetiracetam. He has been advised to continue these medications as prescribed. Additionally, he will be taking the investigational drug for epilepsy for 3 months as part of the randomized controlled trial.

Family History:

The patient reports a family history of bipolar disorder and seizures. His father had bipolar disorder and experienced seizures.

Social History:

The patient is married and has two adult children. He works as an accountant and enjoys reading and spending time with his family in his free time.

Review of Systems:

The patient reports no recent changes in his mood or mental status. He denies any recent symptoms of depression, anxiety, or mania. He reports that he has experienced 27.0 seizures in the past 2 months, with no change in their frequency or severity.

Neurological Examination:

General: The patient appears alert and awake. He is dressed appropriately and responds appropriately to verbal commands.

Cranial Nerves:

* Visual fields: Full fields, with no defects noted.

* Extremity movements: Intact, with no weakness or ataxia observed.

* Sensation: Intact, with no deficits in sensation to light touch, pain, or temperature.

* Tongue movement: Intact, with no weakness or atrophy noted.

* Facial expressions: Intact, with no signs of weakness or paresis observed.

Motor Examination:

* Upper limbs: Intact, with no weakness or ataxia observed.

* Lower limbs: Intact, with no weakness or ataxia observed.

* Muscle strength: 5/5 in all muscle groups.

* Reflexes: Normal, with no signs of hyperexcitability or diminished reflexes.

Gait Examination:

The patient ambulates independently and without assistance. His gait is normal, with no signs of ataxia or weakness observed.

Assessment:

Based on the patient's history and neurological examination, he is diagnosed with bipolar disorder and parietal lobe epilepsy. He is currently experiencing an increase in seizure frequency, which may be related to the randomized controlled trial medication. The patient will continue his current medications and add the investigational drug for epilepsy for 3 months. Close monitoring of seizure frequency and severity will be necessary to assess the effectiveness of the new medication.

Plan:

1. Continue current medications (lamotrigine, valproate, and levetiracetam) as prescribed.

2. Add investigational drug for epilepsy for 3 months.

3. Monitor seizure frequency and severity closely over the next 3 months to assess the effectiveness of the new medication.

4. Follow up with the patient in 1 month to assess his progress and adjust the treatment plan as necessary.

5. Encourage the patient to maintain a consistent sleep schedule, avoid alcohol and drugs, and engage in regular physical activity to reduce stress and promote overall health.

----------------END OF PATIENT ENCOUNTER---------------

-------------- PATIENT ID 36 -- VISIT 2 -----------

Clinic Note #2

Patient ID: 36

Date: 2022-06-19

Identification:

Name: John Doe

Age: 87 years

Gender: Male

Highlights of the Patient's History of Present Illness (HPI):

* Patient with a history of bipolar disorder and parietal lobe epilepsy

* Participated in a randomized controlled trial (RCT) for the investigational drug ID X93461 for epilepsy over the past 3 months

* Reported no symptoms during this time period

* Investigational drug will be discontinued now that the study is complete

Subjective:

* Patient reports no current symptoms

* Reports a reduction in seizure frequency since starting ID X93461 (14.0 seizures over 3 months)

* No changes in mood or cognitive function noted

* Denies any recent medication non-adherence or changes in social history

Medications:

* ID X93461 (investigational drug for epilepsy, discontinued as of today)

* Lithium carbonate (mood stabilizer)

* Levetiracetam (anticonvulsant)

* Gabapentin (anticonvulsant)

Allergies:

* None reported

Social History:

* Married with adult children

* Retired engineer

* Denies any recent travel or changes in living situation

Review of Systems:

* No significant medical or surgical history

* No current symptoms of anxiety, depression, or other psychiatric concerns

* No changes in appetite, weight, or sleep patterns

* No complaints of fatigue, fever, or night sweats

General Physical Examination:

* Vital signs within normal limits (temperature 98.6°F, blood pressure 120/80 mmHg, heart rate 70 beats per minute, respiratory rate 14 breaths per minute)

* No evidence of focal neurological deficits or abnormal muscle strength

* No signs of meningeal irritation or cranial nerve dysfunction

* No cardiovascular abnormalities noted (no murmurs, rubs, or gallops)

Comprehensive Neurologic Examination:

* Mental status: Alert and oriented to person, place, and time

* Cognitive function: Intact, including attention, memory, and language skills

* Speech: Normal, without dysarthria or aphasia

* Motor function: Intact, with normal tone and strength in all muscle groups

* Sensory function: Intact, with normal sensation to light touch, pain, temperature, and vibratory stimuli in all extremities

* Coordination and balance: Normal, without ataxia or dysmetria

* Reflexes: Normal, without hyperreflexia or hyporeflexia

Labs:

* Complete blood count (CBC) within normal limits:

+ White blood cell count (WBC): 4.5 x 10^9/L

+ Neutrophils: 2.8 x 10^9/L

+ Lymphocytes: 1.2 x 10^9/L

+ Monocytes: 0.6 x 10^9/L

+ Hemoglobin (Hb): 13.5 g/dL

+ Hematocrit (Hct): 40.8%

+ Mean corpuscular volume (MCV): 83.2 fl

+ Mean corpuscular hemoglobin (MCH): 27.6 pg

+ Mean corpuscular hemoglobin concentration (MCHC): 32.9%

* Electrolyte panel within normal limits:

+ Sodium: 140 mmol/L

+ Potassium: 3.8 mmol/L

+ Chloride: 100 mmol/L

+ Carbon dioxide (CO2): 22 mmol/L

* Renal function tests within normal limits:

+ Blood urea nitrogen (BUN): 19 mg/dL

+ Creatinine: 0.8 mg/dL

* Thyroid-stimulating hormone (TSH) level: 4.5 mU/L (normal range: 0.4-4.0 mU/L)

Imaging:

* No significant abnormalities on previous MRI scans of the brain and spine

Assessment:

* The patient is an 87-year-old male with a history of bipolar disorder and parietal lobe epilepsy who has participated in a randomized controlled trial for the investigational drug ID X93461. The patient has reported no symptoms during this time period, and the medication will be discontinued now that the study is complete.

* There are no significant changes in the patient's neurological examination or laboratory results at this time.

* The patient's history of bipolar disorder and epilepsy, as well as his age, may contribute to ongoing cognitive and motor function deficits, and close monitoring will be necessary to ensure that any changes in his condition are promptly addressed.

Plan:

* Continue regular follow-up appointments to monitor the patient's neurological status and medication adherence

* Consider adjusting the patient's medication regimen as needed to manage ongoing symptoms of bipolar disorder and epilepsy

* Encourage the patient to continue reporting any changes in his condition or symptoms to the clinic team promptly

Signature:

[Your name and credentials]

----------------END OF PATIENT ENCOUNTER---------------

-------------- PATIENT ID 37 -- VISIT 1 -----------

Patient ID: 37

Visit #1: 2022-05-23

History:

* Temporal lobe epilepsy

* 33 seizures in past 2 months

* Randomized controlled trial (no change in medications)

Neurological Examination:

* alert and oriented

* normal motor function

* normal sensation

* normal reflexes

* no tremors or rigidity

Pertinent Positives:

* No changes in seizure frequency or severity

* No new symptoms or complaints

Assessment and Plan:

* Continue randomized controlled trial

* Add investigational drug for epilepsy for 3 months

* Monitor seizure frequency and severity

* Follow-up visit in 2 weeks

----------------END OF PATIENT ENCOUNTER---------------

-------------- PATIENT ID 37 -- VISIT 2 -----------

Patient ID: 37

Visit #2: 2022-06-23

History: Temporal lobe epilepsy, currently in randomized controlled trial for investigational drug ID X93461

Investigational Drug History:

* Took ID X93461 for 3 months

* Experienced 29.0 seizures during this time

* Reported dizziness, somnolence, vertigo as adverse effects

Neurological Examination:

* Mild tremors in hands and feet

* Slowed reaction time

* No focal deficits or signs of altered sensorium

Pertinent Positives:

* No seizures during the examination

* Dizziness, somnolence, and vertigo reported as adverse effects

Assessment and Plan:

* Discontinue investigational drug ID X93461 due to completion of study

* Monitor for withdrawal seizures and adjust anticonvulsant medication as needed

* Recommend follow-up visit in 2 weeks to assess for any changes in seizure activity or adverse effects.

----------------END OF PATIENT ENCOUNTER---------------

-------------- PATIENT ID 38 -- VISIT 1 -----------

Neurologist Clinic Note

Visit #1: 2022-11-18

Patient ID: 38

Chief Complaint:

The patient presents with a history of schizophrenia, seasonal allergies, and epilepsy. He reports that he has experienced 22.0 seizures in the past two months. The patient is currently enrolled in a randomized controlled trial (RCT) and will be taking the investigational drug for epilepsy for the next three months.

Past Medical History:

The patient has a history of schizophrenia, seasonal allergies, and epilepsy. He has been on the following medications: risperidone (Risperdal) 2mg orally BID for schizophrenia, loratadine (Claritin) 10mg orally OD for seasonal allergies, and levetiracetam (Keppra) 500mg orally BID for epilepsy.

Current Medications:

The patient is currently taking risperidone (Risperdal) 2mg orally BID, loratadine (Claritin) 10mg orally OD, and levetiracetam (Keppra) 500mg orally BID.

Social History:

The patient is a 24-year-old male who works as a software engineer. He denies smoking, alcohol use, or illicit drug use. He reports that he lives with his parents and has a supportive social network.

Family History:

The patient's family history is significant for schizophrenia in his mother, who passed away when the patient was 10 years old due to complications from the disease. The patient does not have any siblings or children.

Review of Systems:

The patient denies fever, chills, or recent infections. He reports that he has been experiencing more frequent seizures over the past two months, which prompted him to seek medical attention.

Neurological Examination:

Mental Status:

The patient's mental status is normal, and he appears alert and oriented. He denies any hallucinations or delusions. His speech is clear and coherent.

Cranial Nerves:

The patient's cranial nerves are intact, and there is no evidence of abnormality in the visual, auditory, or sensory systems.

Motor Examination:

The patient's motor examination reveals no significant weakness, atrophy, or tremors. His muscle tone is normal, and there are no signs of pseudobulbar or other types of movement disorders.

Sensory Examination:

The patient's sensory examination reveals no evidence of numbness, tingling, or loss of sensation in any part of his body. His reflexes are normal, and there is no evidence of sensory ataxia or other types of sensory abnormalities.

Gait Examination:

The patient's gait examination reveals no signs of ataxia or other types of gait abnormalities. He walks normally and has a normal stride length.

Abdominal Examination:

The patient's abdominal examination reveals no evidence of mass, tenderness, or guarding. His bowel sounds are normal, and there is no evidence of obstipation or other types of gastrointestinal abnormalities.

Assessment and Plan:

Based on the patient's history, physical examination, and laboratory results, I recommend the following:

1. Continue the patient's current medications (risperidone, loratadine, and levetiracetam) for the treatment of schizophrenia, seasonal allergies, and epilepsy.

2. Add an investigational drug for epilepsy to the patient's regimen for the next three months as part of the RCT.

3. Monitor the patient's seizure activity closely during this time period and adjust the medication regimen as needed.

4. Repeat neurological examinations at regular intervals to assess the patient's response to treatment and monitor for any changes in his condition.

5. Encourage the patient to continue with his current therapies, including risperidone and levetiracetam, as these medications are effective in managing his symptoms and preventing seizures.

Note: The patient will be seen again in three months for a follow-up visit to assess his response to treatment and monitor for any changes in his condition.

----------------END OF PATIENT ENCOUNTER---------------

-------------- PATIENT ID 38 -- VISIT 2 -----------

Patient ID: 38

Visit #2: 2022-12-18

History: Schizophrenia, seasonal allergies, epilepsy (randomized controlled trial)

Investigational Drug: ID X93461 (epilepsy) - discontinued due to study completion

Seizure frequency: 25.0 seizures over past 3 months

Subjective Symptoms:

* Dizziness

* Ataxia

Neurological Examination:

* Mental status: Normal

* Cranial nerves: Normal

* Motor exam: Normal

* Sensory exam: Normal

* Coordination and balance: Ataxic

* Reflexes: Hyperreflexia

Pertinent Positives:

* Dizziness

* Ataxia

Plan:

* Discontinue investigational drug ID X93461

* Monitor seizure frequency and report any changes

* Address dizziness and ataxia with appropriate therapy

----------------END OF PATIENT ENCOUNTER---------------

-------------- PATIENT ID 39 -- VISIT 1 -----------

Clinic Note - Visit #1

Date: 2021-10-01

Patient ID: 39

Age: 87 years old

Gender: Female

Identification:

* Name: Jane Doe

* Date of Birth: January 1, 1934

* Contact Information: (address, phone number)

History of Present Illness (HPI):

* Chief Complaint: Temporal lobe epilepsy with increased seizure frequency in the past 2 months.

* Duration of symptoms: 2 months

* Patient's description of symptoms: The patient reports experiencing 18.0 seizures in the past 2 months, with increasing frequency and severity. She also mentions difficulty with concentration and memory.

Subjective:

* Patient's overall health: Fair (despite being prescribed medications for epilepsy, the patient has experienced an increase in seizure frequency)

* Current medications and dosages: Levetiracetam 500mg twice daily, Lamotrigine 200mg once daily, Valproate 500mg twice daily (all prescribed for epilepsy)

* Adherence to medication regimen: Poor (patient reports forgetting to take medications at times)

* Allergies: None known

* Social history: The patient lives alone, has a supportive family network, and enjoys reading and spending time with her grandchildren.

Review of Systems:

* General: Denies fever, chills, or other constitutional symptoms.

* Neurological: Reports increased seizure frequency and severity, with associated confusion and memory difficulty.

* Musculoskeletal: No musculoskeletal complaints.

* Cardiovascular: Denies chest pain, shortness of breath, or other cardiovascular symptoms.

* Respiratory: No respiratory complaints.

* Gastrointestinal: Denies abdominal pain, nausea, vomiting, or diarrhea.

* Genitourinary: Denies urinary frequency, urgency, or other genitourinary symptoms.

General Physical Examination:

* Vital signs: Blood pressure 120/80mmHg, Heart rate 70bpm, Temperature 98.6°F (37°C), Weight 55kg

* General appearance: The patient appears well-nourished and in no acute distress.

* Head and neck: No abnormalities detected.

* Extremities: No abnormalities detected.

* Neurological examination:

+ Mental status: The patient is alert and oriented to person, place, and time. However, she appears slightly confused and has difficulty recalling recent events.

+ Cranial nerves: All cranial nerves are intact.

+ Motor function: No motor weakness or atrophy is noted.

+ Sensory function: No sensory deficits are detected.

+ Coordination and balance: No abnormalities detected.

Comprehensive Neurologic Exam:

* Cerebellar function: The patient's cerebellar function is normal.

* Motor function: The patient's motor function is normal.

* Sensory function: The patient's sensory function is intact.

* Coordination and balance: The patient's coordination and balance are normal.

Labs:

* Complete Blood Count (CBC): Within normal limits.

* Electrolyte Panel: Within normal limits.

* Liver Function Tests (LFTs): Within normal limits.

* Renal Function Tests (RFTs): Within normal limits.

* Thyroid-stimulating hormone (TSH) level: Within normal limits.

Imaging:

* Computed Tomography (CT) scan of the brain: No abnormalities detected.

* Magnetic Resonance Imaging (MRI) of the brain: No abnormalities detected.

Assessment:

* The patient presents with a history of temporal lobe epilepsy and an increase in seizure frequency in the past 2 months, despite being prescribed medications for epilepsy.

* The patient's recent increase in seizure frequency may be related to poor adherence to her medication regimen or other factors.

* The patient is participating in a randomized controlled trial and will be taking the investigational drug for epilepsy for 3 months. No change in medications will be made other than adding the investigational drug.

Plan:

* Continue the patient's current medication regimen, with the addition of the investigational drug for epilepsy for 3 months.

* Encourage the patient to improve adherence to her medication regimen.

* Monitor the patient's seizure frequency and adjust the medication regimen as needed.

* Schedule a follow-up appointment in 2 months to assess the patient's progress and adjust the medication regimen accordingly.

Signature:

[Your signature]

Date: 2021-10-01

Note: This clinic note is a sample and should not be used for actual patients. It is important to customize each clinic note to the specific patient and their unique medical history, symptoms, and circumstances.

----------------END OF PATIENT ENCOUNTER---------------

-------------- PATIENT ID 39 -- VISIT 2 -----------

Clinic Note #2 - Patient ID 39 (87 year old female) - 2021-11-01

Identification:

Patient ID 39, an 87-year-old female, is a participant in a randomized controlled trial investigating the safety and efficacy of the investigational drug ID X93461 for the treatment of temporal lobe epilepsy.

History of Present Illness (HPI):

The patient presents with a history of temporal lobe epilepsy, which has been well-controlled with levetiracetam and lamotrigine for the past several years. However, over the past 3 months, the patient has experienced an increase in seizure activity, with 18.0 seizures during this time period. The patient reports feeling fatigued and experiencing balance disorders.

Subjective:

During today's visit, the patient reported feeling generally well, but noted ongoing fatigue and balance difficulties. She also reported difficulty sleeping due to increased seizure activity. The patient expressed concerns about the potential side effects of the investigational drug and its impact on her overall health.

Medications:

The patient has been taking levetiracetam 500mg twice daily and lamotrigine 200mg once daily for the past several years. In addition, the patient has been receiving the investigational drug ID X93461, which was initiated 3 months ago.

Allergies:

The patient has no known allergies to medications or other substances.

Social History:

The patient is a widow and lives alone. She reports a supportive social network, including her children and grandchildren, who assist with her daily needs. The patient enjoys reading and spending time with her family, but has had to limit these activities due to ongoing seizure activity.

Review of Systems:

The patient reported no significant changes in her medical history since her last visit. She continues to experience occasional muscle weakness and memory difficulties, which are consistent with her longstanding history of temporal lobe epilepsy. The patient denied any recent travel, illnesses, or injuries.

General Physical Examination:

Vital signs were within normal limits. The patient's neurological examination was significant for a slight decline in cognitive function, as evidenced by decreased attention and memory difficulties. There were no other abnormal findings.

Comprehensive Neurologic Exam:

The patient underwent a comprehensive neurologic exam, which included an evaluation of her sensory motor functions, balance, and coordination. The exam revealed mild cognitive decline, consistent with the patient's history of temporal lobe epilepsy. There were no other abnormal findings.

Labs:

The patient's recent laboratory results included:

* Blood urea nitrogen (BUN): 20mg/dL

* Creatinine: 1.5mg/dL

* Glucose: 120mg/dL

* Cholesterol: 180mg/dL

* Hemoglobin A1c (HbA1c): 6.8%

Imaging:

The patient's recent imaging results included:

* Computed tomography (CT) scan of the brain: no significant abnormalities were noted.

* Magnetic resonance imaging (MRI) of the brain: no significant abnormalities were noted.

Assessment:

Based on the patient's history, physical examination, and laboratory and imaging results, it is my assessment that the patient is experiencing a slight decline in cognitive function and ongoing seizure activity, consistent with her history of temporal lobe epilepsy. The patient's recent increase in seizures may be related to the discontinuation of the investigational drug ID X93461, which was initiated 3 months ago.

Plan:

Given the patient's ongoing seizure activity and slight decline in cognitive function, it is my recommendation that the patient continue levetiracetam and lamotrigine as prescribed, with close monitoring of seizure activity and adjustments made as needed. The patient will be transitioned off of the investigational drug ID X93461, as the study has been completed. The patient will be scheduled for follow-up visits every 2 months to monitor her seizure activity and cognitive function.

Signature:

[Your signature here]

Date:

[Today's date]

----------------END OF PATIENT ENCOUNTER---------------

-------------- PATIENT ID 50 -- VISIT 1 -----------

---

Clinic Note #1 (2023-01-25)

Patient ID: 50

Age: 40 years old

History: Temporal lobe epilepsy, 28 seizures in the past 2 months. Currently participating in a randomized controlled trial. No change in medications will be made other than adding the investigational drug for epilepsy that will be taken for 3 months.

Examination:

1. General: Alert and oriented, dressed appropriately.

2. Neurological: No focal deficits or abnormalities noted.

3. Cranial Nerves: Intact, including visual fields, facial movements, and sensation.

4. Motor: No weakness or ataxia observed.

5. Sensory: Normal sensation in all extremities.

6. Coordination: Normal coordination and balance.

7. Reflexes: Normal reflexes bilaterally.

8. Gait: Normal gait and stride length.

9. Station: Normal station and stability.

10. Language: Normal language function.

11. Memory: Normal memory function.

12. Attention: Normal attention and concentration.

Assessment:

1. Temporal lobe epilepsy is controlled with current medication regimen.

2. No changes in medications will be made other than adding the investigational drug for epilepsy that will be taken for 3 months.

3. Patient is participating in a randomized controlled trial and is adherent to the study protocol.

4. No significant neurological findings were noted on examination.

Plan:

1. Continue current medication regimen, including the investigational drug for epilepsy for 3 months.

2. Monitor patient's seizure activity and adjust medications as needed.

3. Schedule follow-up appointment in 3 months to assess response to treatment and monitor for any changes in neurological status.

4. Encourage patient to adhere to study protocol and report any concerns or questions to the study team.

----------------END OF PATIENT ENCOUNTER---------------

-------------- PATIENT ID 50 -- VISIT 2 -----------

Visit #2 - 2023-02-25

Patient ID: 50, 40-year-old male

History: Temporal lobe epilepsy

Randomized controlled trial (RCT) with investigational drug ID X93461 for epilepsy (last 3 months)

Current medications: None

Chief complaints:

* Dizziness

* Fatigue

* Nausea

* Balance disorder

Neurological examination:

* Mental status: Alert and oriented

* Cranial nerves: Normal

* Motor exam: Normal

* Sensory exam: Normal

* Reflexes: Normal

Pertinent positives:

* Seizure frequency: 23.0 seizures in the past 3 months (while taking ID X93461)

Assessment:

* Discontinue investigational drug ID X93461 due to completion of RCT

* Monitor for withdrawal seizures and adjust medication as needed

* Address ongoing symptoms (dizziness, fatigue, nausea, balance disorder) with appropriate therapies

Plan:

* Discontinue ID X93461

* Schedule follow-up visit in 2 weeks to monitor for withdrawal seizures and assess symptoms

* Consider initiation of alternative medication(s) as needed

----------------END OF PATIENT ENCOUNTER---------------

-------------- PATIENT ID 51 -- VISIT 1 -----------

---

Clinic Note - Visit #1 (2021-08-07)

Patient: Patient ID 51, 91 year old female

History: Generalized epilepsy with 48.0 seizures in the past 2 months. Randomized controlled trial underway, no changes to medications other than addition of investigational drug for epilepsy for 3 months.

Physical Examination:

* Vital Signs: Blood pressure 120/80 mmHg, pulse 70 bpm, temperature 98.6°F (37°C), weight 55 kg.

* General Appearance: Alert and oriented, dressed in own clothes, no apparent distress.

* Neurological Examination:

+ Mental Status: No significant cognitive impairment detected. Patient able to name current president, recall recent events (e.g., COVID-19 pandemic), and perform serial sevens subtraction task without difficulty.

+ Cranial Nerves: Intact except for mild right facial weakness noted during examination.

+ Motor Examination: No significant weakness or atrophy detected in the upper or lower limbs. Patient able to perform heel-to-toe walking and stand without assistance.

+ Sensory Examination: Intact except for decreased sensation to light touch and pinprick over the left hemiparesis.

+ Coordination and Balance: No significant abnormalities detected during the finger-to-nose test or heel-to-toe walking examination.

* Review of Systems: None reported.

Assessment:

* Patient is a 91-year-old female with a history of generalized epilepsy, currently experiencing 48.0 seizures in the past 2 months.

* Randomized controlled trial underway, no changes to medications other than addition of investigational drug for epilepsy for 3 months.

* Neurological examination reveals mild right facial weakness and decreased sensation to light touch and pinprick over the left hemiparesis.

Plan:

* Continue current medications as prescribed, with addition of investigational drug for epilepsy starting immediately and titrated to target dose over 3 months.

* Monitor patient's seizure frequency and adjust medication regimen as needed.

* Schedule follow-up visit in 2 weeks to assess for any changes in seizure activity or adverse effects from investigational drug.

---

Note: The information provided in this clinic note is fictional and does not reflect any real patient or actual medical practice. It is intended solely for educational purposes and should not be used as a basis for actual clinical decision-making.

----------------END OF PATIENT ENCOUNTER---------------

-------------- PATIENT ID 51 -- VISIT 2 -----------

---

Clinic Note #2, Patient ID 51, 9/7/2021

Identification:

* Patient ID: 51

* Name: [Patient's Name]

* Date of Birth: [Date of Birth]

* Gender: Female

* Age: 91 years old

History of Present Illness (HPI):

* Chief complaint: Decreased appetite and ongoing seizure activity despite discontinuation of investigational drug ID X93461 for epilepsy.

* History of generalized epilepsy with 155.0 seizures over the past 3 months while taking the investigational drug.

Subjective:

* Patient reports decreased appetite and ongoing seizure activity despite discontinuation of the investigational drug.

* Patient denies any recent changes in medications or significant life events that may be contributing to her symptoms.

Medications:

* ID X93461 (investigational drug for epilepsy) - discontinued as of today's date

* Levetiracetam 500mg BID

* Lamotrigine 200mg OND

Allergies:

* No known allergies

Social History:

* Lives alone with assistance from caregiver

* Denies any recent travel or exposure to medications or substances that may be contributing to her symptoms.

* Reports a history of generalized epilepsy and has been seizure-free for the past 5 years prior to initiating the investigational drug.

Review of Systems (ROS):

* General: Denies fever, chills, or any recent illnesses.

* Neurological: Reports ongoing seizure activity despite discontinuation of the investigational drug.

* Musculoskeletal: No musculoskeletal complaints.

* Cardiovascular: Denies any cardiovascular complaints.

* Respiratory: No respiratory complaints.

* Gastrointestinal: Reports decreased appetite.

General Physical Examination:

* Vital signs: Blood pressure 120/80 mmHg, heart rate 70 bpm, temperature 98.6°F (37°C), weight 55 kg

* Head: Normocephalic, ataxic gait

* Eyes: No ocular complaints or abnormalities noted.

* Ears: No ear complaints or discharge noted.

* Nose: No nasal complaints or discharge noted.

* Throat: No throat complaints or swelling noted.

* Chest: Clear lung sounds, no signs of distress or effusion noted.

* Abdomen: Soft and non-tender, no masses or tenderness noted.

* Extremities: No clubbing or cyanosis noted.

Comprehensive Neurologic Examination:

* Mental status: Alert and oriented to person, place, and time.

* Cranial nerves: Intact, including visual fields, facial movements, and tongue strength.

* Motor examination: No motor weakness or ataxia noted.

* Sensory examination: No sensory complaints or deficits noted.

* Reflexes: 2+ (moderate) reflexes bilaterally.

* Coordination and balance: Intact, but with a slight tremor in the hands.

Labs:

* Complete Blood Count (CBC): WBC 4.5 x 10^9/L, Neutrophils 2.8 x 10^9/L, Lymphocytes 1.2 x 10^9/L, Monocytes 0.4 x 10^9/L, Hemoglobin 12.5 g/dL, Hematocrit 36.8%

* Electrolyte Panel: Sodium 138 mmol/L, Potassium 3.9 mmol/L, Chloride 98 mmol/L, Carbon Dioxide 22 mmol/L

* Renal Function Panel: Creatinine 0.65 mg/dL, Glomerular Filtration Rate (GFR) estimated at 65 mL/min/1.73m^2

Imaging:

* Computed Tomography (CT) scan of the brain with and without contrast - no acute or subacute changes noted.

Assessment:

* Decreased appetite and ongoing seizure activity despite discontinuation of the investigational drug ID X93461 for epilepsy.

* No significant changes in neurological examination or laboratory results since previous visit.

* Considering potential causes of decreased appetite, such as depression or anxiety, and ongoing seizure activity despite discontinuation of the investigational drug.

Plan:

* Continue levetiracetam 500mg BID and lamotrigine 200mg OND as prescribed.

* Consider referral to a mental health professional for further evaluation and potential treatment of depression or anxiety.

* Monitor seizure activity and appetite closely over the next several weeks, with follow-up visit scheduled in 4 weeks.

Signature:

---

[Patient's Name]

Date: 9/7/2021

Time: 10:30 AM

----------------END OF PATIENT ENCOUNTER---------------

-------------- PATIENT ID 52 -- VISIT 1 -----------

On May 17th, 2022, Patient ID 52, a 75-year-old female with a history of temporal lobe epilepsy, presented for her first clinic visit in a randomized controlled trial. The patient has experienced 19.0 seizures in the past two months, which is consistent with her previous medical history.

Upon examination, the patient's neurological status was found to be within normal limits, with no focal deficits or signs of altered consciousness. Her speech was clear and coherent, and her gross motor function was intact. The patient's cognitive function was also evaluated using a series of tests, including the Mini-Mental State Examination (MMSE), which revealed no significant impairment in her cognitive abilities.

The patient's medication regimen remains unchanged at this time, with the exception of the addition of the investigational drug for epilepsy, which will be taken for a period of three months. This decision was made based on the patient's history of intractable temporal lobe epilepsy and her willingness to participate in the randomized controlled trial.

Overall, the patient's first clinic visit was uneventful, with no significant changes in her medical condition or neurological status. The patient will be monitored closely throughout the duration of the study, with regular follow-up visits scheduled to assess her progress and response to the investigational drug.

----------------END OF PATIENT ENCOUNTER---------------

-------------- PATIENT ID 52 -- VISIT 2 -----------

Clinic Note Visit #2

Date: 2022-06-17

Patient ID: 52

Age: 75 years old

Gender: Female

History of Present Illness:

The patient, a 75-year-old female with a history of temporal lobe epilepsy, was referred to our clinic for participation in a randomized controlled trial (RCT) investigating the efficacy and safety of the investigational drug ID X93461 for the treatment of epilepsy. The patient has been taking the medication for the past 3 months and has reported experiencing 10.0 seizures during this time. Additionally, the patient has reported experiencing dizziness and gait disturbance.

Neurological Examination:

On examination, the patient was alert and oriented to person, place, and time. Her speech was clear and coherent, and her language was intact. The patient's gross motor function was normal, with no evidence of gait disturbance or ataxia. Fine motor function was also normal, with no evidence of tremors or rigidity. The patient's sensory examination revealed decreased sensation to light touch and pinprick over the left hemiparesis, consistent with her history of temporal lobe epilepsy. The patient's reflexes were normal, with no signs of spasticity or hyperreflexia.

The patient's recent seizure activity was evaluated using the Patient-Generated Index (PGI), which revealed a total of 10.0 seizures over the past 3 months. The patient reported that these seizures were mild to moderate in intensity and did not require any medical interventions. The patient's neurological status was otherwise unchanged, with no evidence of progression or regression of her disease.

Plan:

Given the patient's history of temporal lobe epilepsy and the recent seizure activity, it is recommended that the patient continue to receive regular follow-up care in our clinic to monitor her condition and adjust her medications as needed. The investigational drug ID X93461 will be discontinued now that the study is complete. The patient will be advised to maintain a seizure diary to track any changes in her seizure activity and to report any adverse effects or concerns promptly.

Key Points:

* The patient has been taking the investigational drug ID X93461 for the past 3 months and has reported experiencing 10.0 seizures during this time.

* The patient has also reported dizziness and gait disturbance.

* Neurological examination revealed decreased sensation to light touch and pinprick over the left hemiparesis, consistent with her history of temporal lobe epilepsy.

* The patient's recent seizure activity was evaluated using the Patient-Generated Index (PGI), which revealed a total of 10.0 seizures over the past 3 months.

* The investigational drug ID X93461 will be discontinued now that the study is complete, and the patient will be advised to maintain a seizure diary to track any changes in her seizure activity.

----------------END OF PATIENT ENCOUNTER---------------

-------------- PATIENT ID 53 -- VISIT 1 -----------

On July 21st, 2021, I conducted a clinic visit with Patient ID 53, a 93-year-old male who has been experiencing an increased frequency of seizures over the past two months. The patient has a history of alcoholism, hypothyroidism, and generalized epilepsy, which has been well-controlled on current medications. However, due to the recent escalation in seizure activity, the patient has been enrolled in a randomized controlled trial to investigate a new investigational drug for the treatment of epilepsy.

During the clinic visit, I conducted a thorough neurological examination to assess the patient's current condition and monitor any changes or progression of their disease. The patient's vital signs were within normal limits, with a temperature of 98.6 degrees Fahrenheit, blood pressure of 120/80 mmHg, and a heart rate of 70 beats per minute. The patient's mental status was clear, with no evidence of confusion or disorientation.

The patient reported a total of 78 seizures in the past two months, with an average frequency of approximately 3-4 seizures per week. The seizures have been primarily generalized tonic-clonic in nature, with some occasional partial seizures. The patient has experienced no significant changes in their sensory or motor function, but has reported increased fatigue and weakness due to the frequent seizure activity.

Given the patient's history of alcoholism, hypothyroidism, and generalized epilepsy, as well as the recent escalation in seizure activity, I have decided to maintain the current medication regimen with no changes other than adding the investigational drug for epilepsy. The patient will be instructed to take the investigational drug for a period of three months, and their response to the treatment will be closely monitored during follow-up visits.

----------------END OF PATIENT ENCOUNTER---------------

-------------- PATIENT ID 53 -- VISIT 2 -----------

Patient ID: 53

Visit #2: 2021-08-21

HPI: 93yo male with history of alcoholism, hypothyroidism, and generalized epilepsy. Enrolled in randomized controlled trial for investigational drug ID X93461.

Neurological Examination:

* Cranial nerves: normal

* Motor exam: normal

* Sensory exam: normal

* Reflexes: normal

* Gait and station: stable

Pertinent Positives:

* Seizure frequency: 34.0 over past 3 months (while taking investigational drug)

Assessment and Plan:

* Discontinue investigational drug ID X93461 as study is complete

* Monitor seizure frequency and adjust medication as needed

* Schedule follow-up visit in 6 weeks to assess seizure control and medication side effects.

----------------END OF PATIENT ENCOUNTER---------------

-------------- PATIENT ID 54 -- VISIT 1 -----------

Patient ID: 54

Age: 81 years

History: Occipital lobe epilepsy

Seizures: 14.0 in past 2 months

Randomized controlled trial (RCT)

Neurological Examination:

* Mental status: Alert and oriented

* Speech: Clear and concise

* Motor: No tremors or rigidity

* Sensation: Intact

* Reflexes: Normal

Pertinent Positives:

* No change in medications except for investigational drug for epilepsy

* Addition of investigational drug for 3 months

Assessment and Plan:

* Continue RCT as planned

* Monitor seizure activity and adjust treatment as needed

* Schedule follow-up visit in 3 months.

----------------END OF PATIENT ENCOUNTER---------------

-------------- PATIENT ID 54 -- VISIT 2 -----------

---

Patient ID: 54; Date of Visit: 2023-02-23

Identification:

Age: 81 years old

Gender: Male

History of Present Illness (HPI):

The patient presents with a history of occipital lobe epilepsy and has been participating in a randomized controlled trial for the investigational drug ID X93461 for the past 3 months. During this time, the patient has experienced 3.0 seizures. The patient has also reported experiencing dizziness, constipation, nystagmus, headaches, and gait disturbance.

Subjective:

The patient reports a worsening of symptoms over the past few weeks, including increased frequency and severity of seizures, as well as new-onset dizziness and constipation. The patient also reports experiencing headaches and gait disturbance, which have been persistent throughout the study period.

Medications:

The patient has been adherent to the investigational drug ID X93461 for the past 3 months, as well as his usual antiepileptic medications.

Allergies: None

Social History:

The patient denies any significant social history, including any history of smoking or alcohol use.

Review of Systems:

The patient reports no recent changes in sleep pattern, appetite, or mood. The patient does report experiencing dizziness and constipation, which have been persistent over the past few weeks.

General Physical Examination:

Vital signs:

* Temperature: 98.6°F (37°C)

* Blood pressure: 120/80 mmHg

* Heart rate: 70 bpm

* Respiratory rate: 14 breaths/min

Physical examination revealed a frail elderly male with a steady gait and no evidence of focal weakness or sensory deficits. The patient had a normal neurological examination, including no signs of seizure activity. There was no evidence of dysarthria, hemiparesis, or other cognitive changes.

Comprehensive Neurologic Exam:

* Mental Status: The patient was alert and oriented to person, place, and time. He had a normal memory and attention span.

* Cranial Nerves: There was no evidence of abnormality in the cranial nerves, including no signs of paresis or weakness in the face, arms, or legs.

* Motor Exam: The patient had a steady gait and normal power in all extremities. There was no evidence of tremors, rigidity, or other abnormal movements.

* Sensory Exam: The patient had a normal sensation to light touch, pain, and temperature in all extremities.

* Coordination and Balance: The patient had a normal coordination and balance.

Labs:

* Complete Blood Count (CBC): Within normal limits

* Electrolyte Panel: Within normal limits

* Renal Function Tests (RFTs): Within normal limits

* Liver Function Tests (LFTs): Within normal limits

* Thyroid-stimulating hormone (TSH): Within normal limits

Imaging:

* Magnetic Resonance Imaging (MRI) of the brain: No evidence of any structural abnormalities or lesions.

Assessment:

The patient presents with a history of occipital lobe epilepsy and has been participating in a randomized controlled trial for the investigational drug ID X93461 for the past 3 months. The patient has experienced 3.0 seizures during this time, as well as new-onset dizziness and constipation. The patient also reports experiencing headaches and gait disturbance. Based on the patient's history and physical examination, it is possible that the investigational drug may be contributing to these symptoms. However, it is also possible that these symptoms may be related to other factors, such as the patient's age or underlying medical conditions.

Plan:

* Discontinue the investigational drug ID X93461 due to the completion of the randomized controlled trial.

* Increase monitoring for seizures and adverse effects.

* Consider initiating a new antiepileptic medication to control seizures and manage symptoms.

Signature:

[Insert signature of healthcare provider]

Note: This clinic note is for visit #2, and the patient has been in the randomized controlled trial for the investigational drug ID X93461 for the past 3 months. The patient has experienced 3.0 seizures during this time, as well as new-onset dizziness and constipation. The patient also reports experiencing headaches and gait disturbance. Based on the patient's history and physical examination, it is possible that the investigational drug may be contributing to these symptoms. However, it is also possible that these symptoms may be related to other factors, such as the patient's age or underlying medical conditions.

----------------END OF PATIENT ENCOUNTER---------------

-------------- PATIENT ID 55 -- VISIT 1 -----------

**Patient Information:**

Name: Patient ID 55

Age: 30 years old

Gender: Male

Chief Complaint: Seizures

**History of Present Illness:**

The patient is a 30-year-old male with a history of schizophrenia, arthritis, anxiety, and generalized epilepsy. He has experienced 8.0 seizures in the past 2 months. He is currently enrolled in a randomized controlled trial and is being treated with the investigational drug for epilepsy for the next 3 months.

**Review of Systems:**

The patient reports no changes in his mental status or cognitive function. He denies any recent travel, illnesses, or medication changes. He does report ongoing joint pain and stiffness, which is consistent with his history of arthritis.

**Neurological Examination:**

The patient's neurological examination reveals the following:

* Mental status: The patient appears alert and oriented to person, place, and time. His speech is clear and fluent, and he denies any auditory or visual hallucinations.

* Cranial nerves: All cranial nerves are intact, with no evidence of abnormality in the facial expressions, eye movements, or hearing.

* Motor function: The patient has a normal range of motion in all extremities, and there is no evidence of weakness, atrophy, or fasciculations.

* Sensory function: The patient has a normal sensation in all extremities, with no evidence of numbness, tingling, or loss of sensation.

* Coordination and balance: The patient has a normal coordination and balance, with no evidence of ataxia or dysmetria.

**Assessment and Plan:**

Based on the patient's history and neurological examination, it is assessed that the patient is experiencing ongoing seizures as part of his generalized epilepsy. The addition of the investigational drug for epilepsy is expected to help reduce the frequency and severity of these seizures.

Plan:

1. Continue the investigational drug for epilepsy for the next 3 months, as per the randomized controlled trial protocol.

2. Monitor the patient's seizure activity closely, and adjust the dosage or add additional medications as needed to control seizures.

3. Schedule follow-up appointments every 2 weeks to assess the patient's seizure activity and monitor for any adverse effects of the investigational drug.

4. Encourage the patient to maintain a consistent sleep schedule, avoid alcohol and caffeine, and engage in regular physical activity to help manage his epilepsy.

5. Schedule an MRI scan to assess for any structural brain abnormalities that may be contributing to the seizures.

**Discussion:**

The patient was informed of the results of his neurological examination and the plan for ongoing management of his epilepsy. He was advised to continue with the investigational drug for the next 3 months, and to follow up regularly to monitor for any changes in his seizure activity or adverse effects of the medication. The patient was also counseled on the importance of maintaining a consistent sleep schedule, avoiding alcohol and caffeine, and engaging in regular physical activity to help manage his epilepsy.

----------------END OF PATIENT ENCOUNTER---------------

-------------- PATIENT ID 55 -- VISIT 2 -----------

Date: 2022-12-20

Patient ID: 55

Chief Complaint: The patient presents with a history of schizophrenia, arthritis, anxiety, and generalized epilepsy. He has been participating in a randomized controlled trial for the investigational drug ID X93461 for his epilepsy over the past 3 months.

Past Medical History: The patient has been diagnosed with schizophrenia, arthritis, and anxiety. He has also experienced recurrent seizures for which he has been taking medication.

Current Medications: The patient is currently taking the investigational drug ID X93461 for his epilepsy, as well as medications for his schizophrenia, arthritis, and anxiety.

Neurological Examination:

On examination, the patient's speech is dysarthric with a weak voice and slurred words. His eye movements are slow and deliberate, and he experiences diplopia (double vision). There is no significant tremor or rigidity in his extremities. His reflexes are normal, and there is no evidence of ataxia or dysmetria. The patient's coordination and balance are intact.

Assessment:

The patient has been participating in a randomized controlled trial for the investigational drug ID X93461 for his epilepsy over the past 3 months. During this time, he has experienced 3.0 seizures. His dysarthria and diplopia are likely related to the drug's side effects. As the study is now complete, the investigational drug will be discontinued, and the patient will be referred back to his primary care physician for ongoing management of his conditions.

Plan:

1. Discontinue the investigational drug ID X93461.

2. Refer the patient back to his primary care physician for ongoing management of his conditions.

3. Monitor the patient's seizure activity and adjust medications as needed.

4. Consider referral to a speech therapist for management of dysarthria.

----------------END OF PATIENT ENCOUNTER---------------

-------------- PATIENT ID 56 -- VISIT 1 -----------

Clinic Note Visit #1

Patient ID: 56

Date: 2023-02-05

Identification:

Name: Mary Johnson

DOB: 1951-02-15

Gender: Female

Age: 72 years

Address: 123 Main Street, Anytown, USA

Phone Number: 555-555-5555

Social History:

Mary Johnson is a 72-year-old female who presents with a history of diabetes, anxiety, and occipital lobe epilepsy. She has experienced 5.0 seizures in the past two months, which is an increase from her usual frequency of 2-3 seizures per year. Mary is currently enrolled in a randomized controlled trial for the treatment of her epilepsy.

Subjective:

Mary reports that she has been experiencing more frequent and severe seizures over the past two months. She also notes that she has been experiencing memory difficulties and has had several falls due to loss of consciousness during her seizures. Mary's anxiety has been increasing, and she has been experiencing nightmares related to her seizures.

Objective:

On examination, Mary's vital signs are as follows:

BP: 120/80 mmHg

HR: 80 beats per minute

T: 98.6°F (37°C)

WEIGHT: 56 kg

Mary's neurological examination is significant for the following findings:

* Cranial nerve assessment reveals no deficits in vision, hearing, or facial movements.

* Motor examination demonstrates a moderate degree of hemiparesis on the left side, with weakness of the left arm and leg.

* Sensory examination reveals decreased sensation to light touch and pinprick on the left side of the body.

* Coordination and balance assessment shows a decrease in stability and an increased risk of falls.

Review of Systems:

Mary reports no significant changes in her medical history, including no recent travel or exposure to illness. She denies any recent allergic reactions or adverse effects from her medications. Mary is currently taking the following medications:

* Levetiracetam (Keppra) 500 mg orally twice daily

* Lamotrigine (Lamictal) 200 mg orally once daily

* Valproate (Depakote) 500 mg orally twice daily

Allergies:

Mary has no known allergies to medications.

Social History:

Mary is a retired teacher and lives with her husband of 45 years. She enjoys spending time with her grandchildren, gardening, and reading. Mary reports that she has been experiencing difficulty sleeping due to her seizures and anxiety.

Lab Results:

Mary's laboratory results are as follows:

* Complete Blood Count (CBC): WBC 4.5 x 10^9/L, Neutrophils 2.8 x 10^9/L, Lymphocytes 1.2 x 10^9/L, Monocytes 0.6 x 10^9/L, Eosinophils 0.3 x 10^9/L, Hemoglobin 12.5 g/dL, Hematocrit 38.5%

* Electrolyte Panel: Sodium 141 mmol/L, Potassium 4.2 mmol/L, Chloride 102 mmol/L, Carbon Dioxide 22 mmol/L

* Glucose: 120 mg/dL

* Lipid Panel: Total Cholesterol 180 mg/dL, HDL-C 45 mg/dL, LDL-C 120 mg/dL, Triglycerides 150 mg/dL

Imaging Results:

Mary's imaging results are as follows:

* MRI of the brain with and without contrast: No significant abnormalities were noted.

Assessment:

Based on Mary's history, physical examination, laboratory, and imaging results, it is my assessment that she is experiencing an increase in seizure activity due to her occipital lobe epilepsy. The recent changes in her medication regimen may have contributed to this increase in seizures. Mary's anxiety and sleep disturbances are also significant concerns, as they can exacerbate her seizures and reduce her quality of life.

Plan:

1. Continue Mary's current medication regimen, with the addition of the investigational drug for epilepsy.

2. Monitor Mary's seizure activity closely over the next three months to assess the efficacy of the new medication.

3. Consider adjusting Mary's medication regimen if her seizures do not improve or if she experiences adverse effects from the investigational drug.

4. Encourage Mary to continue her current medications as prescribed and to follow up with me in three months for a subsequent clinic visit.

Signature:

[Your Name]

Date: 2023-02-05

----------------END OF PATIENT ENCOUNTER---------------

-------------- PATIENT ID 56 -- VISIT 2 -----------

Patient ID 56, a 72-year-old female, presented for her second clinic visit on March 5th, 2023. The patient has a history of diabetes, anxiety, and occipital lobe epilepsy, and is currently enrolled in a randomized controlled trial evaluating the investigational drug ID X93461 for the treatment of epilepsy.

Over the past three months, the patient has taken the investigational drug and has experienced 11.0 seizures. Additionally, the patient has reported experiencing dizziness and somnolence during this time. Due to the completion of the study, the investigational drug will be discontinued.

On examination, the patient's neurological status was as follows:

* Mental Status: The patient was alert and oriented x3, with no evidence of cognitive impairment or change in mood.

* Motor Function: The patient had a normal range of motion and strength in all extremities, with no signs of weakness, ataxia, or tremor.

* Sensory Function: The patient had intact sensation in all modalities, including pain, temperature, and touch.

* Cranial Nerves: The patient had intact visual fields, with no evidence of gaze deviation or abnormal eye movements. Hearing was also normal.

* Reflexes: The patient had normal deep tendon reflexes in all extremities.

Overall, the patient's neurological examination was unremarkable, and there were no significant changes since the last visit. The patient will be discontinued from the investigational drug and will be referred to her primary care physician for ongoing management of her diabetes and anxiety. Regular follow-up appointments will be scheduled to monitor her epilepsy and any potential side effects from the discontinuation of the investigational drug.

----------------END OF PATIENT ENCOUNTER---------------

-------------- PATIENT ID 57 -- VISIT 1 -----------

Patient ID: 57

Visit #1: 2021-08-08

History:

* 60 year old male

* History of occipital lobe epilepsy

* 28 seizures in past 2 months

* Randomized controlled trial (RCT)

Neurological Examination:

* Mild cognitive decline

* No significant motor deficits

* Sensory examination within normal limits

* Cranial nerve examination within normal limits

* Coordination and balance within normal limits

Pertinent Positives:

* Increased frequency of seizures in past 2 months

Assessment and Plan:

* Continue RCT medication as prescribed

* Monitor seizure activity and adjust plan as needed

* Schedule follow-up visit in 3 months to assess medication effectiveness and modify plan as necessary.

----------------END OF PATIENT ENCOUNTER---------------

-------------- PATIENT ID 57 -- VISIT 2 -----------

Clinic Note - Visit #2

Patient ID: 57

Date: 2021-09-08

Identification:

* Name: John Doe

* Date of Birth: 01/01/1961

* Patient ID: 57

History of Present Illness (HPI):

* Chief Complaint: Epilepsy

* History of occipital lobe epilepsy

* Patient has been participating in a randomized controlled trial for the investigational drug ID X93461 for the past 3 months

* Patient has experienced 17.0 seizures during this time period

Subjective:

* Patient reports somnolence and back pain

* No changes in sleep or appetite

* No recent travel or illness

Medications:

* ID X93461 (investigational drug for epilepsy) - discontinued due to study completion

* Levetiracetam (keppra) 500mg BID

* Lamotrigine (lamictal) 200mg OD

Allergies:

* None reported

Social History:

* Married with two children

* Works as an accountant

* Denies smoking, alcohol use, or illicit drug use

Review of Systems:

* No changes in symptoms since last visit

* No recent travel or illness

General Physical Examination:

* Vital signs: BP 120/80, HR 70, T 98.6

* Head: normocephalic, ataxic

* Eyes: Pupils equal and reactive to light, no gaze deviations or abnormal movements

* Ears: No cerumen or debris, canals clear

* Nose: No nasal discharge, turbinate bones normal

* Throat: Normal

* Neck: Normocephalic, no mass or tenderness

* Chest: Clear to auscultation, no adventitious sounds

* Abdomen: Soft, normoactive bowel sounds

* Extremities: No clubbing or cyanosis, normal tone and reflexes

Comprehensive Neurologic Examination:

* Mental Status: Alert and oriented to person, place, and time

* Cranial Nerves: II-XII normal

* Motor Exam: Normal tone and strength, no ataxia or tremors

* Sensory Exam: Intact sensation to light touch, pinprick, and temperature

* Coordination and Balance: No ataxia or difficulty with balance

Labs:

* CBC and chemistry panel within normal limits

* Thyroid function tests (T3, T4, TSH) within normal limits

Imaging:

* MRI brain without contrast - no new lesions or abnormalities

Assessment:

* Patient has a history of occipital lobe epilepsy and has been participating in a randomized controlled trial for the investigational drug ID X93461 for the past 3 months

* Patient has experienced 17.0 seizures during this time period, and the medication will be discontinued due to study completion

* The patient reports somnolence and back pain, which may be related to the investigational drug or other factors

* No changes in symptoms since last visit, no recent travel or illness

Plan:

* Discontinue ID X93461 and monitor for withdrawal seizures

* Increase monitoring of somnolence and back pain

* Schedule follow-up appointment in 2 weeks to assess for withdrawal seizures and monitor for any changes in symptoms

Signature:

[Your signature]

Date: [Today's date]

----------------END OF PATIENT ENCOUNTER---------------

-------------- PATIENT ID 58 -- VISIT 1 -----------

Clinic Note: Visit #1 (2021-12-03)

Patient ID: 58

Age: 68 years old

Gender: Female

Chief Complaint: Seizures

History of Present Illness:

The patient, a 68-year-old female with a history of hypertension, arthritis, autism, and parietal lobe epilepsy, presents with a complaint of increased seizure activity over the past two months. She has experienced 15.0 seizures during this time, with no change in medications other than the addition of an investigational drug for epilepsy as part of a randomized controlled trial.

Past Medical History:

* Hypertension

* Arthritis

* Autism

* Parietal lobe epilepsy

Medications:

* No changes other than the addition of investigational drug for epilepsy

Social History:

* Married

* No smoking or alcohol use

Review of Systems:

* None reported

Neurological Examination:

* Mental status: Alert and oriented x3

* Speech: Normal rate and volume

* Facies: Normal

* Head: Normocephalic, normoglyceridemic

* Eyes: Pupils equal, round, and reactive to light

* Vision: 20/20 in each eye

* Hearing: Normal hearing sensitivity

* Extremities: Normal tone and reflexes

* Sensory: Intact sensation to light touch, pain, and temperature over all body regions

* Motor: Normal strength and movement in all four limbs

Assessment and Plan:

* The patient's recent increase in seizure activity is concerning, particularly given her history of parietal lobe epilepsy.

* Given the patient's participation in a randomized controlled trial, no changes will be made to her medications at this time other than the addition of the investigational drug for epilepsy.

* The patient will continue to receive the same medication regimen as before, with close monitoring of seizure activity and adjustments to be made as needed.

* The patient will return for follow-up visits every 2 weeks to assess her condition and make any necessary adjustments to her treatment plan.

* The patient's hypertension and arthritis will also be monitored and managed as needed.

I hereby certify that this document is a true and accurate record of the patient's visit and that all information has been documented in accordance with professional and ethical standards.

Signature: ______________

Date: 2021-12-03

Note: The above clinic note is a sample and should not be used for actual clinical practice. It is important to follow the specific guidelines and protocols of your healthcare institution and the randomized controlled trial in which the patient is participating.

----------------END OF PATIENT ENCOUNTER---------------

-------------- PATIENT ID 58 -- VISIT 2 -----------

Visit #2 - 2022-01-03

Patient ID: 58

Age: 68 years old

Female

History: Hypertension, Arthritis, Autism, Parietal Lobe Epilepsy (study participant)

Investigational Drug: ID X93461 (discontinued)

Seizure Frequency: 23.0 seizures over past 3 months

Adverse Effects:

* Somnolence

* Balance disorder

* Back pain

* Gait disturbance

Neurological Examination:

* Mental status: alert and oriented

* Speech: normal

* Motor exam: normal (except for gait disturbance)

* Sensory exam: normal

* Reflexes: normal

* Gait disturbance noted (ataxic)

Assessment:

* Study drug ID X93461 discontinued due to completion of randomized controlled trial

* Patient reports adverse effects of somnolence, balance disorder, back pain, and gait disturbance

* Neurological examination reveals ataxic gait disturbance

Plan:

* Monitor patient for ongoing adverse effects and seizure frequency

* Consider alternative medications to manage epilepsy and address gait disturbance

* Schedule follow-up visit in 4-6 weeks to assess patient's condition and adjust plan as needed.

----------------END OF PATIENT ENCOUNTER---------------

-------------- PATIENT ID 59 -- VISIT 1 -----------

Visit #1 - 2021-06-19

Patient ID: 59

Age: 99 years old

Female

History:

* Generalized epilepsy

* 11 seizures in past 2 months

* Randomized controlled trial (RCT) participant

Neurological Examination:

* Alert and oriented x3

* No focal deficits

* Normal motor function

* Normal sensation

* No cranial nerve abnormalities

Pertinent Positives:

* Seizures have increased in frequency over past 2 months

* RCT participant

Assessment and Plan:

* Continue RCT medication as prescribed

* Monitor seizure activity and adjust plan as needed

* Schedule follow-up visit in 3 months.

----------------END OF PATIENT ENCOUNTER---------------

-------------- PATIENT ID 59 -- VISIT 2 -----------

Visit #2 - 2021-07-19

Patient ID: 59, Female, 99 years old

Generalized Epilepsy with Ataxia

The patient, a 99-year-old female with a history of generalized epilepsy, presented for her second visit in the randomized controlled trial. Over the past three months, the patient has been administered the investigational drug ID X93461 for her epilepsy. During this time, she has experienced 6.0 seizures, with ataxia being reported as a side effect.

Neurological Examination:

Upon examination, the patient's vital signs were within normal limits. Her mental status was clear, and she was able to communicate effectively. The patient's speech was slightly slurred, likely due to the ataxia. The patient's neck muscles were stable, and there was no evidence of weakness or paralysis. The patient's sensation was intact, and there were no signs of peripheral neuropathy.

The patient's cognitive function was assessed using the Mini-Mental State Examination (MMSE), which revealed a score of 28 out of 30, indicating near-normal cognitive function. The patient's memory was intact, and she was able to recall recent events without difficulty. The patient's language skills were also normal, with no evidence of aphasia or dysarthria.

The patient's motor function was evaluated using the Unified Parkinson's Disease Rating Scale (UPDRS), which revealed a score of 2 out of 4, indicating mild ataxia. The patient had difficulty with gait and balance, and she experienced occasional tremors in her hands. However, there was no evidence of rigidity or bradykinesia.

The investigational drug ID X93461 will be discontinued now that the study is complete. The patient's seizure activity will continue to be monitored, and any changes in her condition will be documented accordingly. The patient will be followed up with regular visits to assess her condition and ensure that she remains stable.

----------------END OF PATIENT ENCOUNTER---------------

-------------- PATIENT ID 70 -- VISIT 1 -----------

Clinic Note - Visit #1

Patient ID: 70

Date: 2021-09-26

Chief Complaint:

The patient presents with a history of schizophrenia, diabetes, and generalized epilepsy. She has experienced 74 seizures in the past two months. She is currently participating in a randomized controlled trial and has been instructed not to make any changes to her medications unless explicitly authorized by the study team.

History of Present Illness:

The patient reports experiencing an increased frequency of seizures over the past two months, with an average of 3-4 seizures per week. She denies any recent changes in her symptoms or medications. She reports that her seizures have been well-controlled on her current regimen but has noticed some increase in the intensity of her seizures recently.

Past Medical History:

The patient has a history of schizophrenia, diabetes, and generalized epilepsy. She has been previously treated with a variety of medications for these conditions, including antipsychotics, anticonvulsants, and insulin therapy.

Medication List:

The patient is currently taking the following medications:

1. Risperidone (Risperdal) 2mg oral daily

2. Lamotrigine (Lamictal) 50mg oral daily

3. Levetiracetam (Keppra) 2000mg oral daily

4. Metformin (Glucophage) 1000mg oral daily

5. Insulin glargine (Lantus) 10 units subcutaneous daily

Review of Systems:

The patient denies any recent changes in her symptoms or medical history. She reports that she has been experiencing some increased fatigue and weight gain over the past few months, but these symptoms have been stable and do not seem to be related to her seizure activity.

Neurological Examination:

The patient's neurological examination was unremarkable, with no evidence of focal deficits or abnormal reflexes. Her speech is clear and coherent, and her language is intact. She does not exhibit any signs of dysarthria or aphasia. Her cranial nerves are normal, and she has no signs of facial weakness or paresis. Her coordination and balance are also normal, and she does not exhibit any signs of ataxia or tremors.

Assessment and Plan:

Based on the patient's history and examination, it is my assessment that the patient is experiencing an increased frequency of seizures likely related to her generalized epilepsy. The recent addition of the investigational drug for epilepsy may be contributing to this increase in seizure activity. I recommend that the patient continue her current medication regimen, with the addition of the investigational drug for the next three months as part of the randomized controlled trial. I also recommend that the patient return for regular follow-up visits to monitor her seizure activity and adjust her medications as needed.

Plan:

1. Continue current medication regimen, including risperidone, lamotrigine, levetiracetam, metformin, and insulin glargine.

2. Add investigational drug for epilepsy for the next three months as part of the randomized controlled trial.

3. Schedule regular follow-up visits every two weeks to monitor seizure activity and adjust medications as needed.

4. Instruct the patient not to make any changes to her medications unless explicitly authorized by the study team.

----------------END OF PATIENT ENCOUNTER---------------

-------------- PATIENT ID 70 -- VISIT 2 -----------

On October 26th, 2021, I conducted a clinic visit with Patient ID 70, a 38-year-old female with a history of schizophrenia, diabetes, and generalized epilepsy. The patient has been participating in a randomized controlled trial (RCT) and has been receiving the investigational drug ID X93461 for the past three months to treat her epilepsy.

According to the patient's report, she has experienced 25.0 seizures during this time period, which is a significant improvement compared to her previous baseline. However, the patient has also reported experiencing somnolence, which may be related to the medication. The patient's neurological examination revealed no changes in her cognitive function or motor skills, but she did exhibit mild tremors and rigidity, which are consistent with her pre-existing condition of generalized epilepsy.

Given that the RCT has now been completed, the investigational drug ID X93461 will be discontinued. The patient will be transitioned back to her standard of care medication and will be closely monitored for any changes in her seizure frequency or severity. It is important to note that the patient's somnolence may be a side effect of the investigational drug, but it could also be a symptom of another underlying condition. Therefore, close monitoring and regular follow-up appointments will be crucial in managing her condition effectively.

In conclusion, this clinic visit provided an opportunity to assess the patient's response to the investigational drug ID X93461 and to monitor her overall health. While the medication has shown some efficacy in reducing seizure frequency, it has also been associated with somnolence. As the RCT comes to an end, it is important to carefully transition the patient back to her standard of care medication and to continue monitoring her closely for any changes in her condition.

----------------END OF PATIENT ENCOUNTER---------------

-------------- PATIENT ID 71 -- VISIT 1 -----------

Patient ID: 71

Date: 2022-03-09

* History: Depression, temporal lobe epilepsy (TLE)

* Current medications: None (randomized controlled trial)

* Seizure history: 32 seizures in past 2 months

Neurological Examination:

* Mental status: Alert and cooperative

* Speech: Normal

* Language: Normal

* Memory: Intact

* Attention/concentration: Normal

* Executive function: Normal

* Sensory: No defects

* Motor: No deficits or tremors

* Reflexes: 2+ (normal)

* Gait: Normal

Pertinent Positives:

* No changes in medications other than investigational drug for epilepsy

* Randomized controlled trial

* Increased seizure frequency (32 seizures in past 2 months)

Assessment and Plan:

* Monitor seizure activity and side effects of investigational drug

* Continue randomized controlled trial

* Reassess medication needs after 3-month trial period.

----------------END OF PATIENT ENCOUNTER---------------

-------------- PATIENT ID 71 -- VISIT 2 -----------

Visit #2: April 9th, 2022

Patient ID 71, a 26-year-old female with a history of depression and temporal lobe epilepsy, was seen in the clinic for her second visit. The patient has been participating in a randomized controlled trial (RCT) evaluating the investigational drug ID X93461 for the treatment of epilepsy. Over the past three months, the patient has taken the drug and has reported a decrease in seizure frequency. However, she has also experienced dizziness and somnolence as adverse effects.

On examination, the patient's neurological status was stable. She had no focal deficits or signs of weakness/paresis. Her speech was clear and comprehensible, with no evidence of aphasia or dysarthria. Her cranial nerves were intact, with no abnormalities detected in the visual, auditory, or vestibular systems. Her reflexes were normal, with no signs of hyperreflexia or hyporeflexia. The patient's gait was normal, with no signs of ataxia or spasticity.

The patient reported a total of 13 seizures during the past three months, with no significant changes in her seizure frequency compared to previous visits. She also reported experiencing dizziness and somnolence, which were mild and transient. These adverse effects have been well-tolerated by the patient, and there have been no significant changes in her overall health status.

Given that the RCT is now complete, the investigational drug ID X93461 will be discontinued. The patient will continue to receive standard antiepileptic medications as needed to control her seizures. Her next follow-up visit is scheduled for three months from today's date.

----------------END OF PATIENT ENCOUNTER---------------

-------------- PATIENT ID 72 -- VISIT 1 -----------

---

Clinic Note #1 - Patient ID: 72 - Date: 2021-08-13

Identification:

Name: Mary Johnson

DOB: 03/01/1940

Gender: Female

Age: 81 years old

History of Present Illness (HPI):

Mary Johnson, an 81-year-old female, presents with a history of frontal lobe epilepsy and has been experiencing an increase in seizure activity over the past two months. She has had 13.0 seizures during this time frame. Mary is currently participating in a randomized controlled trial (RCT) examining the efficacy of an investigational drug for epilepsy.

Subjective:

Mary reports experiencing frequent seizures, with an average frequency of 13.0 seizures per month over the past two months. She also reports difficulty with memory and concentration, which has been affecting her daily activities. Mary denies any recent changes in medications or treatments.

Objective:

Medications:

* Levetiracetam (Keppra) 500mg PO QID

* Lacosamide (Vimpat) 100mg PO BID

* Ethotoin (Peganone) 200mg PO QHS

Allergies:

* None reported

Social History:

* Married with adult children

* Retired from her job as a teacher

* Enjoys reading and spending time with family

Review of Systems:

* General: No fever, chills, or night sweats.

* Neurological: Frequent seizures, difficulty with memory and concentration.

* Medication adherence: Denies any recent changes in medications or treatments.

General Physical Examination:

Vital signs:

* Temperature: 98.6°F

* Pulse: 70/min

* Blood Pressure: 120/80 mmHg

* Respiration Rate: 14/min

Neurological Examination:

Mary's neurological examination revealed the following:

* Cranial nerves: Intact, with no signs of abnormalities.

* Motor function: No signs of weakness or ataxia.

* Sensory function: No signs of paresthesias or sensory loss.

* Coordination and balance: Normal.

Comprehensive Neurologic Exam:

* Mental status: Alert and oriented x3.

* Language: Intact, with no signs of aphasia.

* Memory: Poor, with difficulty recalling recent events.

* Attention and concentration: Difficulty maintaining focus.

* Executive function: Difficulty with decision-making and problem-solving.

Labs:

* Complete Blood Count (CBC): Within normal limits.

* Basic Metabolic Panel (BMP): Within normal limits.

* Electrolyte Panel: Within normal limits.

* Thyroid Function Tests (TFTs): Within normal limits.

Imaging:

* Brain Magnetic Resonance Imaging (MRI): No abnormalities noted.

Assessment:

* Mary's frequent seizures and poor memory suggest ongoing frontal lobe epilepsy.

* Her participation in an RCT examining the efficacy of an investigational drug for epilepsy may impact her current symptoms.

* No changes will be made to her current medications, other than adding the investigational drug for the next 3 months.

Plan:

1. Continue current medications as prescribed.

2. Add the investigational drug for the next 3 months.

3. Monitor seizure activity and adjust medications as needed.

4. Follow up in 3 months to reassess symptoms and adjust treatment plan as necessary.

Signature:

---

Mary Johnson's clinic note #1, dated 2021-08-13, reveals a comprehensive assessment of her neurological symptoms and history of frontal lobe epilepsy. The addition of the investigational drug to her current medications is intended to improve her seizure control and address her difficulty with memory and concentration. Regular follow-up appointments will be essential to monitor her progress and adjust her treatment plan as needed.

----------------END OF PATIENT ENCOUNTER---------------

-------------- PATIENT ID 72 -- VISIT 2 -----------

Neurology Clinic Note

Patient ID: 72

Date of Visit: 2021-09-13

History of Present Illness:

The patient is an 81-year-old female with a history of frontal lobe epilepsy. She has been participating in a randomized controlled trial (RCT) for the investigational drug ID X93461 for epilepsy over the past 3 months. During this time, she has experienced 7.0 seizures. The patient has also reported constipation.

Review of Systems:

The patient denies any recent changes in her medical history, including no new medications, surgeries, or illnesses. She does report some difficulty with constipation over the past few months.

Physical Examination:

Vital Signs:

* Blood pressure: 120/80 mmHg

* Heart rate: 75 beats per minute

* Temperature: 98.6 degrees Fahrenheit

* Weight: 140 pounds (down from previous visit)

Neurological Examination:

The patient's neurological examination was unremarkable, with no signs of weakness, paralysis, or sensory deficits. Her speech and language are intact, and her cognitive function appears to be normal. The patient does exhibit a mild tremor in her hands, which is consistent with her previous examinations.

Assessment:

Based on the patient's history of frontal lobe epilepsy and recent seizure activity, it is clear that the investigational drug ID X93461 has had a beneficial effect in reducing the patient's seizures. However, the patient's report of constipation may be related to the medication, and further monitoring and management may be necessary.

Plan:

Given the completion of the RCT, the patient will discontinue taking ID X93461. The patient will continue to be monitored for seizure activity and any changes in her constipation symptoms. If necessary, additional medications or interventions may be recommended to address these issues. Regular follow-up appointments will be scheduled to ensure the patient's continued well-being and to monitor her condition.

The patient will also be informed of the study results and any future treatment options that may be available to her. Additionally, the patient will be referred to a gastroenterologist for further evaluation and management of her constipation symptoms.

Note: This clinic note is fictional and for illustrative purposes only. It does not reflect any actual patient or study.

----------------END OF PATIENT ENCOUNTER---------------

-------------- PATIENT ID 73 -- VISIT 1 -----------

---

Clinic Note #1 (2021-04-08)

Patient ID: 73

Age: 18 years old

Gender: Male

Chief Complaint: Seasonal allergies, arthritis, ADHD, and frontal lobe epilepsy with increased seizure activity in the past 2 months.

History of Present Illness (HPI): The patient presents with a history of seasonal allergies, arthritis, ADHD, and frontal lobe epilepsy. He has been experiencing an increase in seizure activity over the past 2 months, with 18.0 seizures reported. The patient is currently enrolled in a randomized controlled trial for the treatment of his epilepsy.

Subjective:

* Patient reports worsening seizure activity over the past 2 months, with an average of 3-4 seizures per week.

* Patient notes that his seizures have been more frequent and longer in duration compared to previous years.

* The patient experiences occasional diplopia (double vision) during seizures.

* He denies any recent changes in medications or significant life events that may be contributing to his increased seizure activity.

Medications:

* Levetiracetam (Keppra) 500mg twice daily

* Lamotrigine (Lamictal) 200mg once daily

* Methylphenidate (Ritalin) 20mg once daily for ADHD

Allergies:

* Seasonal allergies (pollen, dust mites)

* No known drug allergies

Social History:

* Patient lives with his parents and has a younger sister.

* He is currently enrolled in college and is studying to be a software engineer.

* The patient denies any history of substance abuse or psychiatric illness.

Review of Systems (ROS):

* Respiratory: No respiratory symptoms reported.

* Cardiovascular: No cardiovascular symptoms reported.

* Gastrointestinal: Patient reports occasional abdominal pain, but no changes in bowel movements or appetite.

* Musculoskeletal: Patient reports joint pain and stiffness in the knees and wrists, consistent with his diagnosis of arthritis.

* Neurological: Patient reports worsening seizure activity over the past 2 months, including diplopia (double vision) during seizures.

General Physical Examination:

* Vital signs: Blood pressure 120/80mmHg, heart rate 70bpm, temperature 98.6°F, weight 150lbs.

* Head: No focal neurological deficits. No evidence of trauma or mass lesions.

* Eyes: Pupils are equal and reactive to light, no abnormalities noted.

* Ears: No abnormalities noted.

* Nose: No nasal discharge or deformities.

* Throat: No throat symptoms reported.

* Chest: No abnormalities noted.

* Abdomen: No abnormalities noted.

* Extremities: No abnormalities noted.

Comprehensive Neurologic Exam:

* Mental Status: The patient is alert and oriented to person, place, and time. He is cooperative and able to communicate his symptoms effectively.

* Cranial Nerves: All cranial nerves are intact, with no abnormalities noted.

* Motor Exam: No significant motor deficits or tremors observed.

* Sensory Exam: No sensory deficits or abnormalities noted.

* Reflexes: Normal reflexes are present, with no signs of spasticity or hyporeflexia.

Labs:

* Complete Blood Count (CBC): Within normal limits.

* Electrolyte Panel: Within normal limits.

* Liver Function Tests (LFTs): Within normal limits.

* Renal Function Tests (RFTs): Within normal limits.

* Thyroid-stimulating hormone (TSH): Within normal limits.

Imaging:

* Computed Tomography (CT) scan of the head: No abnormalities noted.

Assessment:

* Patient presents with a history of seasonal allergies, arthritis, ADHD, and frontal lobe epilepsy, with increased seizure activity over the past 2 months.

* The patient is currently enrolled in a randomized controlled trial for the treatment of his epilepsy, with no changes in medications other than the addition of the investigational drug.

* Patient reports worsening seizure activity over the past 2 months, with an average of 3-4 seizures per week, and occasional diplopia (double vision) during seizures.

* No significant neurological deficits or abnormalities noted on comprehensive neurologic exam.

Plan:

1. Continue the patient's current medications for epilepsy and ADHD, with the addition of the investigational drug for the next 3 months.

2. Monitor the patient's seizure activity closely and adjust medications as needed.

3. Schedule follow-up appointments every 2 weeks to assess the patient's progress and adjust the treatment plan as necessary.

4. Encourage the patient to continue his current therapies for seasonal allergies and arthritis.

Signature:

[Your Name]

---

Note: This clinic note is fictional and does not reflect any actual patient or medical condition. It is intended as an example of a comprehensive neurological examination and assessment for a patient with a history of seasonal allergies, arthritis, ADHD, and frontal lobe epilepsy, and should not be used as a basis for actual clinical decision-making.

----------------END OF PATIENT ENCOUNTER---------------

-------------- PATIENT ID 73 -- VISIT 2 -----------

Patient ID 73, 18 y/o male

History: Seasonal allergies, arthritis, ADHD, frontal lobe epilepsy (study participant)

Investigational Drug: ID X93461 (epilepsy) - discontinued due to study completion

Seizure History: 17.0 seizures in past 3 months

Symptoms: Dizziness, fatigue, vomiting, headaches

Neurological Examination:

* General: Alert and oriented x3

* Cranial Nerves: Intact

* Motor: Normal strength and tone

* Sensory: Normal sensation

* Coordination: Normal

* Reflexes: Normal

Pertinent Positives:

* Dizziness

* Fatigue

* Vomiting

* Headaches

Assessment and Plan:

* Discontinue investigational drug ID X93461 due to study completion

* Monitor for seizure activity and address with anticonvulsants as needed

* Address symptoms of dizziness, fatigue, vomiting, and headaches with appropriate medications and therapies

* Schedule follow-up appointment in 2 weeks for continued monitoring and assessment.

----------------END OF PATIENT ENCOUNTER---------------

-------------- PATIENT ID 74 -- VISIT 1 -----------

Patient ID: 74

Date of Visit: August 5, 2022

Chief Complaint: Temporal lobe epilepsy with increased seizure frequency in the past two months.

History of Present Illness:

The patient is a 22-year-old female with a history of hypertension, diabetes, depression, and temporal lobe epilepsy. She has been experiencing an increase in seizure frequency over the past two months, with 11.0 seizures reported. The patient is currently enrolled in a randomized controlled trial (RCT) and is taking her regular medications as prescribed.

Past Medical History:

The patient has a history of hypertension, diabetes, and depression, all of which are well-controlled on current medications. She has a history of temporal lobe epilepsy and has been experiencing an increase in seizure frequency over the past two months.

Medications:

The patient is currently taking levetiracetam 1000mg twice daily, lamotrigine 200mg once daily, and valproic acid 500mg twice daily for her epilepsy. She is also taking metformin 1000mg twice daily for her diabetes and amlodipine 5mg once daily for her hypertension.

Neurological Examination:

General: The patient appears well-nourished and in no acute distress.

Head and Eyes:

* No gaze evoked nystagmus

* No ocular flutter

* No diplopia

* Pupils are equal and reactive to light

Cranial Nerves:

* II-XII cranial nerves are intact

Motor Examination:

* No weakness or paralysis

* Normal muscle tone and bulk

* No tremors or rigidity

Sensory Examination:

* No sensory deficits

* Normal sensation to light touch, pain, and temperature

Coordination and Balance:

* Normal coordination and balance

Assessment and Plan:

The patient's increased seizure frequency over the past two months is concerning, and further evaluation is warranted. The patient will continue to participate in the RCT and will be taking the investigational drug for epilepsy for the next three months. No changes will be made to her current medications at this time. The patient will be monitored closely for any changes in seizure frequency or severity.

Recommendations:

1. Continue with current medications and add the investigational drug for epilepsy.

2. Monitor the patient's seizure frequency and severity closely over the next three months.

3. Schedule a follow-up appointment in one month to assess the patient's progress.

----------------END OF PATIENT ENCOUNTER---------------

-------------- PATIENT ID 74 -- VISIT 2 -----------

Clinic Note: Visit #2 (2022-09-05) - Patient ID 74

I. Identification:

* Name: [Patient Name]

* Date of Birth: [Date of Birth]

* Patient ID: 74

II. History of Present Illness (HPI):

* The patient is a 22-year-old female with a history of hypertension, diabetes, depression, and temporal lobe epilepsy.

* She has been participating in a randomized controlled trial for the investigational drug ID X93461 for epilepsy over the past 3 months.

* The patient has had 9.0 seizures during this time period, with no symptoms reported.

III. Subjective:

* The patient reports no symptoms or concerns at this time.

IV. Medications:

* ID X93461 (investigational drug for epilepsy) - discontinued due to completion of the randomized controlled trial

* Levetiracetam (seizure medication) - ongoing

* Lisinopril (hypertension medication) - ongoing

* Metformin (diabetes medication) - ongoing

V. Allergies:

* None reported

VI. Social History:

* Marital status: Single

* Occupation: Student

* Living situation: With parents

VII. Review of Systems:

* General: No significant complaints or concerns.

* Neurological: No recent seizure activity or other neurological symptoms reported.

* Cardiovascular: Hypertension is well-controlled with current medications.

* Pulmonary: No respiratory symptoms or concerns.

* Gastrointestinal: No significant gastrointestinal symptoms or concerns.

VIII. General Physical Examination:

* Vital signs: Temperature 98.6°F, Blood Pressure 120/80 mmHg, Heart Rate 70 bpm, Respiratory Rate 14 bpm.

* Head: Normocephalic (head circumference within normal limits), no focal tenderness or masses.

* Eyes: No visual disturbances or abnormalities.

* Ears: No ear discharge or pain.

* Nose: No nasal congestion or discharge.

* Throat: No sore throat or tonsillitis.

* Chest: No signs of respiratory distress or effusion.

* Abdomen: Soft and non-tender, no masses or organomegaly.

* Extremities: No swelling, deformity, or weakness.

IX. Comprehensive Neurological Examination:

* Cranial nerves: Intact (no abnormalities detected).

* Motor function: No significant motor weakness or wasting.

* Sensory function: No significant sensory deficits or abnormalities.

* Coordination and balance: Normal.

* Reflexes: Normocellular (reflexes within normal limits).

X. Labs:

* Complete Blood Count (CBC): Within normal limits.

* Electrolyte Panel: Within normal limits.

* Liver Function Tests (LFTs): Within normal limits.

* Renal Function Tests (RFTs): Within normal limits.

* Thyroid-stimulating Hormone (TSH): Within normal limits.

XI. Imaging:

* No recent imaging studies have been performed.

XII. Assessment:

* The patient has a history of hypertension, diabetes, depression, and temporal lobe epilepsy.

* She has been participating in a randomized controlled trial for the investigational drug ID X93461 for epilepsy over the past 3 months, with no significant adverse effects or seizure activity reported.

* The patient is currently stable and there are no significant neurological or systemic concerns at this time.

XIII. Plan:

* Continue levetiracetam (seizure medication) as prescribed.

* Discontinue ID X93461 (investigational drug for epilepsy) due to completion of the randomized controlled trial.

* Monitor blood pressure and diabetes control regularly.

* Schedule follow-up appointment in 3 months for ongoing care and monitoring.

XIV. Signature:

[Signature of healthcare provider]

Note: This clinic note is a fictional example and does not represent any real patient or study. It is intended to demonstrate the format and content of a comprehensive neurological examination and assessment for a patient with a history of hypertension, diabetes, depression, and temporal lobe epilepsy participating in a randomized controlled trial.

----------------END OF PATIENT ENCOUNTER---------------

-------------- PATIENT ID 75 -- VISIT 1 -----------

On August 6th, 2021, Patient ID 75, a 23-year-old female, presented to our clinic for her first visit. The patient has a history of frontal lobe epilepsy and has experienced 12.0 seizures in the past two months. She is currently participating in a randomized controlled trial (RCT) and will be taking the investigational drug for epilepsy for the next three months.

Upon examination, the patient's neurological status was as follows:

* General appearance: Alert and oriented to person, place, and time.

* Mental status: Cooperative and responding appropriately to questions.

* Speech: Clear and concise, with no observed difficulties.

* Motor function: No gross motor deficits or ataxia.

* Sensory function: No obvious sensory deficits or abnormalities.

* Cranial nerves: Intact, including visual and auditory functions.

The patient reported no significant changes in her seizure activity since the previous visit. She denied any new medication allergies or side effects. Her current medications include levetiracetam and lacosamide, which will remain unchanged during the RCT. The patient was instructed to continue with her regular dosing schedule and to contact the clinic if she experiences any changes in her seizure activity or medication side effects.

The patient's family history, social history, and review of systems were all unremarkable. No red flags or warning signs for seizure activity were noted. The patient was informed about the RCT and the potential risks and benefits of the investigational drug. She was advised to maintain a seizure log to track any changes in her seizure activity during the study period.

In conclusion, Patient ID 75 presented for her first clinic visit with a history of frontal lobe epilepsy and recent increase in seizure activity. She is currently participating in an RCT and will be taking the investigational drug for epilepsy for the next three months. No changes to her medication regimen were made other than adding the investigational drug. A complete neurological examination was conducted, and no significant abnormalities were noted. The patient was educated about the RCT and instructed to monitor her seizure activity closely during the study period.

----------------END OF PATIENT ENCOUNTER---------------

-------------- PATIENT ID 75 -- VISIT 2 -----------

Clinic Note #2 - Patient ID 75 - 2021-09-06

Identification:

Name: Patient ID 75

DOB: 03/01/1998

Gender: Female

HPI:

The patient, a 23-year-old female with a history of frontal lobe epilepsy, has been participating in a randomized controlled trial (RCT) evaluating the safety and efficacy of the investigational drug ID X93461 for the treatment of epilepsy. The patient has been receiving the medication for the past 3 months and has reported a total of 5.0 seizures during this time period. The patient has also experienced somnolence, which is a known side effect of the medication.

Subjective:

During today's clinic visit, the patient presented with a complaint of ongoing somnolence and decreased cognitive function. She reported that her seizure activity has remained stable over the past 3 months, with no changes in frequency or severity. The patient also mentioned that she has been experiencing difficulty concentrating and memory issues, which have affected her daily activities.

Medications:

ID X93461 (investigational drug) - discontinued due to completion of RCT

Levetiracetam 500mg BID - stable dosage

Lamotrigine 200mg OD - stable dosage

Allergies:

None reported

Social History:

The patient resides with her parents and has a supportive social network. She is currently enrolled in college and is studying to become a nurse. The patient denies any history of substance abuse or psychiatric illness.

Review of Systems:

General: Denies fever, chills, or recent illness.

Cardiovascular: Denies chest pain, shortness of breath, or palpitations.

Respiratory: Denies cough, chest tightness, or wheezing.

Gastrointestinal: Denies abdominal pain, nausea, or diarrhea.

Genitourinary: Denies urinary frequency, urgency, or hesitancy.

Musculoskeletal: Denies joint pain or muscle weakness.

Neurological: Denies headaches, dizziness, or blurred vision.

Psychiatric: Denies depression, anxiety, or suicidal ideation.

General Physical Examination:

Vital Signs:

* Temperature: 98.6°F (37°C)

* Blood Pressure: 120/80 mmHg

* Heart Rate: 80 beats per minute

* Respiratory Rate: 14 breaths per minute

Comprehensive Neurologic Examination:

* Mental Status: Alert and oriented x3.

* Cranial Nerves: Intact, including visual fields.

* Motor Function: Normal strength and tone in all extremities.

* Sensory Function: Intact sensation to light touch, pinprick, and temperature.

* Reflexes: Normocephalic, normochromic.

* Coordination and Balance: Normal.

Labs:

* Complete Blood Count (CBC) - WNL

* Electrolyte Panel - WNL

* Liver Function Tests (LFTs) - WNL

* Thyroid-stimulating Hormone (TSH) - WNL

Imaging:

* Computed Tomography (CT) scan of the brain - WNL

Assessment:

The patient presents with a history of frontal lobe epilepsy and has been participating in an RCT evaluating the safety and efficacy of ID X93461. The patient has reported a total of 5.0 seizures during the 3-month treatment period, and has experienced somnolence as a known side effect of the medication. The patient's neurological examination is unremarkable, and all laboratory and imaging studies are within normal limits. Given the completion of the RCT, the investigational drug ID X93461 will be discontinued, and the patient will be transitioned back to her previous medication regimen.

Plan:

1. Discontinue ID X93461.

2. Continue levetiracetam 500mg BID and lamotrigine 200mg OD as previously prescribed.

3. Monitor the patient's seizure activity and adjust medications as needed.

4. Schedule follow-up clinic visit in 1 month to assess the patient's status.

Signature:

[Your Name]

Neurologist

----------------END OF PATIENT ENCOUNTER---------------

-------------- PATIENT ID 76 -- VISIT 1 -----------

Clinic Note Visit #1

Patient ID: 76

Date: 2022-02-11

Identification:

Name: John Doe

DOB: 02-01-1933

Address: 123 Main St, Anytown, USA

Issues:

1. History of hypothyroidism

2. Frontal lobe epilepsy

3. Recent increase in seizure activity (10.0 seizures in past 2 months)

4. Participating in a randomized controlled trial

HPI:

Mr. John Doe, an 89-year-old male, presented to the clinic today for his first visit. He reported a history of hypothyroidism and frontal lobe epilepsy, as well as a recent increase in seizure activity. Specifically, he has experienced 10.0 seizures in the past two months. The patient is currently participating in a randomized controlled trial to assess the efficacy of an investigational drug for epilepsy.

Subjective:

Mr. Doe reported that his seizure activity has been increasing steadily over the past two months, with an average of 2-3 seizures per week. He also noted that he has been experiencing more frequent episodes of confusion and disorientation, particularly in the morning hours. Additionally, the patient reported difficulty with memory recall and word-finding difficulties.

Medications:

* Levothyroxine (T4) 25 mcg oral daily

* Lamotrigine (LTG) 200 mg oral daily

* Valproic acid (VPA) 500 mg oral twice daily

Allergies:

* None reported

Social history:

* Married with adult children

* Retired from his job as an accountant

* Enjoys reading, gardening, and spending time with his family

Review of Systems:

* General: No significant changes in overall health status

* Neurological: Seizure activity (10.0 seizures in past 2 months), confusion, disorientation, memory recall difficulties, word-finding difficulties

* Cardiovascular: No reported chest pain or shortness of breath

* Respiratory: No reported cough or difficulty breathing

* Gastrointestinal: No reported nausea, vomiting, or abdominal pain

* Musculoskeletal: No reported joint pain or weakness

General Physical Examination:

Vital signs:

* Blood pressure: 120/80 mmHg

* Heart rate: 75 beats per minute

* Temperature: 98.6°F (37°C)

Neurological examination:

Mr. Doe was alert and oriented to person, place, and time. His speech was clear and concise, with no evidence of aphasia or dysarthria. No significant abnormalities were observed in his cranial nerves, motor function, or sensory examination. However, the patient did exhibit some mild tremors in his hands and feet, which is consistent with his history of frontal lobe epilepsy.

Comprehensive Neurologic Exam:

* Cerebral cortex: No significant abnormalities observed

* Basal ganglia: No significant abnormalities observed

* Thalamus: No significant abnormalities observed

* Brainstem: No significant abnormalities observed

* Spinal cord: No significant abnormalities observed

Labs:

* Complete Blood Count (CBC): Within normal limits

* Basic Metabolic Panel (BMP): Within normal limits

* Thyroid-stimulating hormone (TSH): 2.5 mU/L (normal range: 0.4-4.0 mU/L)

* Free thyroxine (FT4): 1.2 ng/dL (normal range: 0.8-1.8 ng/dL)

Imaging:

* Computerized tomography (CT) scan of the head: No significant abnormalities observed

Assessment:

Mr. Doe's recent increase in seizure activity, coupled with his history of hypothyroidism and frontal lobe epilepsy, suggests a possible connection between these conditions. Additionally, the patient's participation in a randomized controlled trial may be contributing to his increased seizure activity. Based on the patient's current medications and laboratory results, no changes will be made to his medication regimen at this time, except for the addition of the investigational drug for epilepsy. The patient will be closely monitored for any changes in his seizure activity or other neurological symptoms.

Plan:

1. Continue current medications as prescribed

2. Add investigational drug for epilepsy (dose and frequency to be determined by the study protocol)

3. Monitor patient closely for changes in seizure activity or other neurological symptoms

4. Schedule follow-up visit in 2 weeks to assess medication adherence and seizure activity

Signature:

[Your Signature]

Date: [Today's Date]

Time: [Today's Time]

----------------END OF PATIENT ENCOUNTER---------------

-------------- PATIENT ID 76 -- VISIT 2 -----------

Date: 2022-03-11

Patient ID: 76

Age: 89 years old

Chief Complaint: Fatigue, headaches, diplopia

History of Present Illness: The patient is an 89-year-old male with a history of hypothyroidism and frontal lobe epilepsy. He has been participating in a randomized controlled trial for the investigational drug ID X93461 for epilepsy over the past 3 months. During this time, he has experienced 2.0 seizures and has reported fatigue, headaches, and diplopia.

Past Medical History: The patient has a history of hypothyroidism and frontal lobe epilepsy. He has been on medication for these conditions for many years.

Medications: The patient is currently taking levothyroxine sodium (T4) for his hypothyroidism and valproic acid for his epilepsy. He has also been receiving the investigational drug ID X93461 for the past 3 months.

Neurological Examination:

General: The patient appears generally well-oriented and alert. He has a normal speech and language.

Cranial Nerves: The patient has no signs of abnormal cranial nerve function, including no evidence of diplopia or other ocular abnormalities.

Motor: The patient has no significant weakness or atrophy of the muscles. His strength and tone are normal.

Sensory: The patient has no significant sensory deficits. He has a normal sense of touch, pain, temperature, and vibratory sensation.

Coordination and Balance: The patient has no signs of ataxia or difficulty with balance. He is able to walk normally and performs heel-to-toe gait without assistance.

Results of Investigations:

Laboratory Results: The patient's most recent blood work, including thyroid function tests and liver enzymes, are within normal limits.

Imaging Studies: The patient has had no recent imaging studies.

Plan:

The patient will discontinue the investigational drug ID X93461 now that the study is complete. He will continue to receive valproic acid for his epilepsy and levothyroxine sodium (T4) for his hypothyroidism. The patient will be followed up in clinic to monitor for any changes in his condition and to address any concerns or side effects that may arise.

The patient's fatigue, headaches, and diplopia will be monitored closely, and appropriate management will be provided as needed. The patient will also be advised on the importance of compliance with his medication regimen and follow-up appointments to ensure optimal seizure control and overall health.

----------------END OF PATIENT ENCOUNTER---------------

-------------- PATIENT ID 77 -- VISIT 1 -----------

On January 5th, 2022, I had the pleasure of conducting a clinic visit with Patient ID 77, a 91-year-old male who has been experiencing an increased frequency of seizures over the past two months. As part of a randomized controlled trial, the patient will be taking an investigational drug for the next three months to assess its effectiveness in managing their multifocal epilepsy.

Upon entering the examination room, I was greeted by the patient who appeared alert and in no apparent distress. After introducing myself and the purpose of my visit, I began the neurological examination. The patient's mentation was clear and focused, with no evidence of clouding or confusion. Their speech was normal in rate and rhythm, with no observable tremors or abnormalities.

The patient's cranial nerves were intact, with no signs of deficits or weaknesses. Their extraocular movements were smooth and coordinated, and their pupillary responses were normal. The patient's motor function was assessed through a series of gross motor movements, including flexion, extension, abduction, adduction, and rotation. There were no observable deficits or weaknesses in any of these areas.

The patient's sensory examination revealed intact pinprick and light touch sensation over all areas of the body. However, there was a noticeable decrease in vibratory sensation in the lower extremities, consistent with their long-standing history of diabetes. The patient's reflexes were symmetrical and normal in amplitude, with no signs of hyporeflexia or hyperreflexia.

In conclusion, the neurological examination conducted on January 5th, 2022, revealed no significant changes or abnormalities in Patient ID 77's condition. The patient's cognitive function remains intact, and their motor and sensory functions are stable. As part of the randomized controlled trial, the patient will be taking the investigational drug for the next three months to assess its effectiveness in managing their multifocal epilepsy. Regular follow-up appointments have been scheduled to monitor the patient's progress and adjust medications as needed.

----------------END OF PATIENT ENCOUNTER---------------

-------------- PATIENT ID 77 -- VISIT 2 -----------

Visit #2 (2022-02-05)

Patient ID: 77

Age: 91 years

History:

* Arthritis

* Diabetes

* Seasonal allergies

* Multifocal epilepsy

* Randomized controlled trial (RCT)

Investigational Drug:

* ID X93461 (epilepsy)

* Taken for 3 months

* 2.0 seizures during this time

* Dizziness, gait disturbance reported

Neurological Examination:

* Mild decreased sensation in left arm and leg

* Bilateral extensor plantar responses

* No gaze evoked nystagmus or intention tremor

* Cerebellar function normal

* No abnormalities on rapid alternating movement (RAM) testing

Pertinent Positives:

* Decreased sensation in left arm and leg

* Bilateral extensor plantar responses

Assessment and Plan:

* Discontinue investigational drug ID X93461 due to completion of RCT

* Monitor for seizure activity and adjust medication as needed

* Address dizziness and gait disturbance with physical therapy and/or medication

* Follow up in 2 weeks for reassessment.

----------------END OF PATIENT ENCOUNTER---------------

-------------- PATIENT ID 78 -- VISIT 1 -----------

On March 3rd, 2022, Patient ID 78, a 64-year-old female, presented to our clinic for her first visit. The patient has a history of hypothyroidism, depression, asthma, and epilepsy, with a recent increase in seizure activity. Over the past two months, the patient has experienced 59.0 seizures, which is concerning and warranted further evaluation.

During the clinical examination, the patient's vital signs were within normal limits. She appeared alert and oriented, with no significant changes in her mental status or behavior. The patient's speech was clear and coherent, with no evidence of aphasia or dysarthria. Her motor function was intact, with no noticeable weakness, tremors, or ataxia. The patient's reflexes were normal, and she had no sensory deficits.

The patient's neurological examination revealed no significant abnormalities, other than a mild tremor in her hands, which is consistent with her history of hypothyroidism. Her cognitive function was intact, with no evidence of decline or impairment. The patient's recent seizure activity was carefully evaluated, and no new or unusual symptoms were noted.

Given the patient's history of epilepsy and recent increase in seizure activity, we have decided to enroll her in a randomized controlled trial investigating a new medication for the treatment of epilepsy. The patient will be taking the investigational drug for three months, and no changes will be made to her current medications other than the addition of the study drug. We will closely monitor her seizure activity and adjust the study drug dosage as needed to optimize her treatment and minimize any potential side effects.

----------------END OF PATIENT ENCOUNTER---------------

-------------- PATIENT ID 78 -- VISIT 2 -----------

Here's the clinic note for Patient ID 78:

Visit #2, dated 2022-04-03:

Patient ID 78, 64-year-old female:

* History: Hypothyroidism, depression, asthma, epilepsy

* Currently in a randomized controlled trial

* Took investigational drug ID X93461 for epilepsy (last 3 months)

+ Reported 29.0 seizures during this time

+ Experienced headaches

* Study now complete; drug to be discontinued

Neurological Examination:

* Head: Normocephalic, normotensive

* Eyes: Normal visual acuity, no abnormalities noted

* Ears: No ear discharge or odor

* Nose: No nasal discharge or facial swelling

* Throat: No stridor or abnormal breathing sounds

* Neck: No masses or asymmetry

* Extremities: Normal tone, no spasticity or flaccidity

* Sensory: Normal sensation to light touch, pinprick, and temperature

* Motor: Normal strength and range of motion in all extremities

Pertinent Positives:

* Seizures (29.0)

* Headaches

Assessment and Plan:

* Discontinue investigational drug ID X93461

* Monitor seizure activity and adjust medication as needed

* Address headaches as needed

* Schedule follow-up visit in 4-6 weeks for reassessment.

----------------END OF PATIENT ENCOUNTER---------------

-------------- PATIENT ID 79 -- VISIT 1 -----------

**Clinic Note - Visit #1**

Patient ID: 79

Date of Visit: 2022-09-08

Chief Complaint:

The patient presents with a history of ADHD, dermatitis, and temporal lobe epilepsy. He has experienced 16.0 seizures in the past 2 months. The patient is currently enrolled in a randomized controlled trial.

Past Medical History:

The patient has a history of ADHD, dermatitis, and temporal lobe epilepsy. He has been diagnosed with juvenile myoclonic epilepsy and has had multiple seizures over the years. The patient has been managed with medications and has had some improvements.

Medication History:

The patient is currently taking levtiracetam 1000mg BID, lamotrigine 200mg OD, and prednisone 5mg QAM. He has been advised to continue these medications as prescribed.

Family History:

The patient's family history is negative for neurological disorders.

Social History:

The patient denies alcohol or drug use. He works as an accountant and reports that he has had no changes in his job or living situation recently.

Review of Systems:

The patient reports no recent illnesses or surgeries. He denies any new symptoms, such as fever, headache, or weight loss.

Physical Examination:

General: The patient appears well-nourished and alert. He has no evidence of tremors or rigidity.

Head and Neck: No focal neurological deficits were found. The patient had normal tone and range of motion in all extremities. There was no evidence of facial weakness or paresis.

Extremities: The patient's extremities were normal, with no signs of weakness or atrophy.

Sensory: The patient had normal sensation in all extremities.

Cerebellar Function: The patient had normal cerebellar function, including rapid alternating movements and heel-to-toe walking.

Motor: The patient had normal motor function, with no signs of weakness or paresis.

Reflexes: The patient's reflexes were normal, with equal and appropriate response to stimuli.

Assessment:

The patient is a 37-year-old male with a history of ADHD, dermatitis, and temporal lobe epilepsy. He has experienced 16.0 seizures in the past 2 months, which is an increase from his previous seizure frequency. The patient is currently enrolled in a randomized controlled trial and will be taking the investigational drug for epilepsy for the next 3 months.

Plan:

1. Continue all current medications as prescribed.

2. Monitor the patient's seizure activity closely and report any changes to the study team immediately.

3. Schedule follow-up appointments every 2 weeks to assess the patient's condition and adjust the plan as needed.

4. Encourage the patient to continue his current lifestyle and avoid any potential triggers for seizures, such as stress or lack of sleep.

5. Advise the patient to inform his study team of any changes in his condition or medications.

----------------END OF PATIENT ENCOUNTER---------------

-------------- PATIENT ID 79 -- VISIT 2 -----------

Clinic Note #2: Patient ID 79

Date: October 8th, 2022

Identification:

Patient ID 79 is a 37-year-old male who presented to our clinic for a follow-up visit. He was referred by Dr. Smith, the principal investigator of the randomized controlled trial (RCT) in which he has been participating over the past 3 months. The patient has a history of ADHD, dermatitis, and temporal lobe epilepsy.

Highlights of the Patient's History:

* Patient reported a history of ADHD symptoms since childhood, including difficulty sustaining attention, hyperactivity, and impulsivity.

* He also reported a history of dermatitis, specifically atopic dermatitis, which has been well-controlled with topical corticosteroids.

* Patient was diagnosed with temporal lobe epilepsy 5 years ago, and has been on levetiracetam monotherapy since then. He has experienced an average of 4.0 seizures per year, which have been well-controlled with the current medication regimen.

Subjective:

During today's visit, patient reported somnolence and headaches as adverse effects from the investigational drug ID X93461, which he has been taking for the past 3 months as part of the RCT. He also complained of increased seizure activity during this time period, with an average of 4.0 seizures per month compared to his usual average of 2.5 seizures per year. However, patient reported that these adverse effects have been tolerable and have not significantly impacted his daily activities or quality of life.

Medications:

* Levetiracetam (Keppra) 500 mg PO QD

* Lamotrigine (Lamictal) 200 mg PO QD

* ID X93461 (investigational drug) 500 mg PO BID (discontinued as of today due to completion of RCT)

Allergies:

* No known allergies

Social History:

* Married with two children (ages 8 and 10)

* Employed as a software engineer

* Denies smoking, alcohol consumption, or illicit drug use

* Reports occasional caffeine use (2-3 cups of coffee per day)

Review of Systems:

* General: Denies fever, weight loss, or night sweats.

* Neurological: Denies weakness, numbness, or paresthesias. Reports increased seizure activity as described above.

* Psychiatric: Denies depression, anxiety, or suicidal ideation.

* Musculoskeletal: Denies joint pain or muscle weakness.

General Physical Examination:

* Vital signs: Blood pressure 120/80 mmHg, heart rate 70 bpm, temperature 98.6°F (37°C), weight 75 kg

* Head: Normocephalic, normotectal. No focal deficits or abnormalities.

* Eyes: Pupils are equal and reactive to light, no nystagmus or afferent defects.

* Ears: No ear discharge or odor. Hearing is normal to speech and tone.

* Nose: No nasal flaring or rhinorrhea.

* Throat: No swelling or exudate.

* Chest: Clear to auscultation.

* Abdomen: Soft, normoactive bowel sounds. No rebound tenderness or guarding.

* Extremities: Normal tone and reflexes. No deformities or lesions.

Comprehensive Neurological Examination:

* Mental status: Alert and oriented x3. Reports no confusion, disorientation, or memory impairment.

* Cranial nerves: Intact, including visual fields and funduscopic examination.

* Motor examination: No focal weakness or atrophy. Normal muscle tone and reflexes.

* Sensory examination: No sensory deficits or abnormalities.

* Coordination and balance: No difficulties with gait or balance.

Labs:

* Complete Blood Count (CBC): WBC 4.5 x10^9/L, NEUT 2.8 x10^9/L, LYMP 1.1 x10^9/L, MONO 600/L, EOSIN 300/L, RBC 4.2 x10^12/L, HEMAT 15.8 g/dL, PLATELETS 275 x10^9/L

* Blood chemistry: Na 138 mmol/L, K 4.4 mmol/L, Cl 102 mmol/L, CO2 26 mmol/L, BUN 18 mg/dL, Creatinine 1.05 mg/dL, Glucose 94 mg/dL

* Urinalysis: No red blood cells or white blood cells on dipstick. No protein or ketones in urine.

Imaging:

* CT scan of the brain (done as part of RCT) shows no abnormalities or lesions.

Assessment:

* Patient presents with a history of ADHD, dermatitis, and temporal lobe epilepsy. He has been participating in an RCT for the past 3 months, during which time he reported somnolence and headaches as adverse effects, as well as increased seizure activity compared to his usual baseline. However, patient reported that these adverse effects have been tolerable and have not significantly impacted his daily activities or quality of life. Based on the results of today's examination and review of systems, patient's neurological status is stable.

* Given the patient's history of epilepsy and recent increase in seizure activity during the RCT, it is essential that he continues to receive appropriate anticonvulsant therapy to control his seizures and maintain his quality of life.

Plan:

* Continue levetiracetam monotherapy at current dosage.

* Consider adding an additional anticonvulsant medication, such as lamotrigine or carbamazepine, to further control seizure activity.

* Monitor patient's seizure activity and adjust medications as needed.

* Schedule follow-up appointment in 3 months for reassessment of seizure activity and medication regimen.

Signature:

[Your Name]

Neurologist

----------------END OF PATIENT ENCOUNTER---------------

-------------- PATIENT ID 90 -- VISIT 1 -----------

Patient ID: 90

Visit #1: 2022-02-17

* 96 year old male

* History of ADHD, alcoholism, multifocal epilepsy

* 13 seizures in past 2 months

* Randomized controlled trial (RCT)

Neurological Examination:

* Alert and oriented x3

* No signs of fatigue or confusion

* Mild tremors in hands and feet

* Brisk reflexes

* No sensory deficits

* No ataxia or dysmetria

Pertinent Positives:

* Seizures continue despite current medication regimen

* No changes to medications will be made except for addition of investigational drug for epilepsy

* Patient is participating in RCT

Assessment and Plan:

* Continue current medication regimen with addition of investigational drug for epilepsy

* Monitor seizure activity closely during 3-month trial period

* Schedule follow-up visit in 4 weeks to assess efficacy and tolerability of investigational drug

* Encourage patient to maintain detailed seizure log to track frequency and severity of seizures.

----------------END OF PATIENT ENCOUNTER---------------

-------------- PATIENT ID 90 -- VISIT 2 -----------

Patient ID: 90

Visit #2: 2022-03-17

History:

* ADHD

* Alcoholism

* Multifocal epilepsy

* Randomized controlled trial (RCT) for investigational drug ID X93461

Neurological Examination:

* Ataxia

Pertinent Positives:

* 7.0 seizures in past 3 months while taking investigational drug ID X93461

Assessment and Plan:

* Discontinue investigational drug ID X93461 due to completion of RCT

* Monitor for seizure activity and ataxia

* Consider alternative treatments for epilepsy and ataxia as needed.

----------------END OF PATIENT ENCOUNTER---------------

-------------- PATIENT ID 91 -- VISIT 1 -----------

Patient ID: 91

Visit #1: 2022-03-14

History:

* Bipolar disorder

* Occipital lobe epilepsy (16 seizures in past 2 months)

* Randomized controlled trial

Neurological Examination:

* Normal tone, strength, and reflexes

* Intact sensation

* No motor or sensory deficits

* Normal coordination and balance

Pertinent Positives:

* No change in medications except for investigational drug for epilepsy

* Randomized controlled trial

Assessment:

* Epilepsy persistently active despite previous treatments

* Considering investigational drug as a new treatment option

Plan:

* Continue current medications

* Add investigational drug for epilepsy (3 months)

* Monitor seizure activity and adjust plan as needed

* Follow-up visit in 2 months.

----------------END OF PATIENT ENCOUNTER---------------

-------------- PATIENT ID 91 -- VISIT 2 -----------

Visit #2 (2022-04-14)

Patient ID: 91

Age: 49 years

History: Bipolar disorder, occipital lobe epilepsy (study participant in randomized controlled trial)

Investigational Drug History: X93461 for epilepsy (last 3 months)

Neurological Examination:

* Somnolence (reported by patient)

* Fatigue (reported by patient)

* Ataxia (observed)

Pertinent Positives:

* Seizure frequency: 6.0 seizures in the past 3 months

Assessment and Plan:

* Discontinue investigational drug X93461 due to study completion

* Monitor patient for withdrawal symptoms or recurrent seizures

* Schedule follow-up visit in 2 weeks to assess patient's condition and adjust treatment plan as needed.

----------------END OF PATIENT ENCOUNTER---------------

-------------- PATIENT ID 92 -- VISIT 1 -----------

Patient ID: 92, Male, Age: 95 years old

Visit #1 - Date: 2021-07-05

Chief Complaint: The patient presents with a history of diabetes, arthritis, asthma, and multifocal epilepsy. He has experienced 9.0 seizures in the past two months. He is currently enrolled in a randomized controlled trial to evaluate the effectiveness of an investigational drug for his epilepsy.

Past Medical History: The patient has been diagnosed with diabetes, arthritis, and asthma for several years. He has been managed with medications and lifestyle modifications. He also has a history of multifocal epilepsy and has experienced recurrent seizures over the past few months.

Current Medications: The patient is currently taking metformin, sulfasalazine, fluticasone propionate, and montelukast sodium for his diabetes, arthritis, and asthma. He has also been prescribed levetiracetam and lacosamide for his epilepsy.

Present Illness: The patient presents with a recent increase in seizure frequency over the past two months. He has experienced 9.0 seizures during this time, which is higher than his usual frequency. His most recent seizure occurred yesterday morning, and he reports that it was similar in intensity and duration to previous seizures.

Neurological Examination:

General: The patient appears alert and oriented to person, place, and time. He has a calm demeanor and speaks clearly with no signs of dysarthria or slurred speech.

Cranial Nerves: There is no evidence of abnormality in the cranial nerves. Pupils are equal, round, and react to light with no signs of asymmetry or abnormal movement.

Motor: The patient has a normal range of motion in all extremities without any signs of weakness or atrophy. There is no evidence of tremors or rigidity.

Sensory: The patient has a normal sensation in all extremities without any signs of numbness or tingling. He does not report any unusual sensations or pain.

Coordination and Balance: The patient has a normal coordination and balance without any signs of unsteadiness or instability.

Assessment: Based on the patient's history, physical examination, and current medications, it is determined that he is suitable for participation in the randomized controlled trial. His recent increase in seizure frequency may be related to the investigational drug being studied, but further evaluation and monitoring are necessary to confirm this association. The patient will continue to take his current medications as prescribed and will also receive the investigational drug for a period of three months. Regular follow-up appointments will be scheduled to monitor his seizure activity and assess the effectiveness of the investigational drug.

----------------END OF PATIENT ENCOUNTER---------------

-------------- PATIENT ID 92 -- VISIT 2 -----------

---

Patient ID: 92

Clinic Visit #2 (2021-08-05)

Identification:

Name: John Doe

Age: 95 years old

Gender: Male

HPI:

John Doe, a 95-year-old male with a history of diabetes, arthritis, asthma, and multifocal epilepsy, presents for his second clinic visit. He has been participating in a randomized controlled trial (RCT) evaluating the investigational drug ID X93461 for the treatment of epilepsy. Over the past 3 months, John has been taking the investigational drug and has reported experiencing 8.0 seizures during this time. He also complains of dizziness.

Subjective:

John reports that he has been feeling generally well since his last clinic visit, with no significant changes in his symptoms or medication adherence. However, he does report some mild dizziness that has been persistent over the past few days. He denies any new symptoms or seizures.

Medications:

* ID X93461 (investigational drug): discontinued as of today due to completion of RCT

* Levetiracetam (Keppra): 2000mg/day

* Lamotrigine (Lamictal): 500mg/day

* Metformin (Glucophage): 1000mg/day

Allergies:

* None reported

Social History:

John lives with his wife of 65 years and has two adult children. He reports no significant changes in his social or occupational activities since his last clinic visit. He does mention that he has been experiencing some mild dizziness, which has affected his ability to drive and perform daily activities.

Review of Systems:

* General: Denies fever, chills, or other constitutional symptoms

* Neurological: Reports mild dizziness, no changes in seizure activity or medication adherence

* Cardiovascular: No significant changes in blood pressure or heart rate

* Respiratory: Denies shortness of breath or other respiratory symptoms

General Physical Examination:

Vital signs:

* Blood pressure: 120/80 mmHg

* Heart rate: 70 beats per minute

* Temperature: 98.6°F (37°C)

Head and Neck:

* No significant changes in facial or extremity movements

* No signs of meningism or cranial nerve deficits

Extremities:

* No signs of weakness, wasting, or sensory loss

Comprehensive Neurologic Examination:

John's comprehensive neurologic examination reveals no significant changes in his symptoms since his last clinic visit. His speech and language are intact, and he is able to perform simple motor tasks without any notable weakness or sensory deficits. His funduscopic examination is within normal limits, and there is no evidence of retinal hemorrhages or other abnormalities.

Labs:

* Complete Blood Count (CBC): Within normal limits

* Electrolyte Panel: Within normal limits

* Renal Function Panel: Within normal limits

* Thyroid Function Panel: Within normal limits

Imaging:

* No significant changes in brain imaging studies since last clinic visit

Assessment:

John's mild dizziness and reported seizure activity (8.0 seizures over the past 3 months) suggest that the investigational drug ID X93461 may be effective in reducing seizure frequency, but further study is needed to confirm these findings. However, due to the completion of the RCT, John will be discontinued from the investigational drug and his seizures will be monitored on a regular basis to assess for any changes in seizure activity. His dizziness should also be monitored and managed as needed.

Plan:

* Discontinue ID X93461 as of today

* Increase monitoring of seizure activity and dizziness

* Consider initiation of other medications or therapies as needed to manage dizziness and seizures

Signature:

John Doe

---

Note: This clinic note is a fictional example and should not be used as a basis for actual patient care.

----------------END OF PATIENT ENCOUNTER---------------

-------------- PATIENT ID 93 -- VISIT 1 -----------

Clinic Note Visit #1

Patient ID: 93

Date: 2022-05-17

Chief Complaint: The patient presents with a history of frontal lobe epilepsy and has experienced 20.0 seizures in the past two months. He is currently participating in a randomized controlled trial to evaluate the efficacy of an investigational drug for epilepsy.

History of Present Illness: The patient reports that he has been experiencing increased frequency of seizures over the past two months, with an average of 2-3 seizures per week. He denies any recent changes in his medications or other medical conditions. He is aware that he is participating in a clinical trial and is taking the investigational drug as prescribed.

Review of Systems: The patient reports no recent changes in his sleep patterns, appetite, or mood. He denies any recent travel or exposure to illness. He is aware of the potential risks and benefits of the clinical trial and has signed informed consent.

Neurological Examination:

General: The patient appears alert and awake, with no evidence of altered mental status or confusion.

Cranial Nerves: The patient's cranial nerves are intact, with no evidence of abnormalities in vision, hearing, or facial movements.

Motor System: The patient's motor system is intact, with no evidence of weakness, atrophy, or tremors.

Sensory System: The patient's sensory system is intact, with no evidence of numbness, tingling, or loss of sensation.

Seizure Activity: The patient reports that he has experienced 20.0 seizures in the past two months, with an average of 2-3 seizures per week. He denies any recent changes in his seizure activity or frequency.

Plan: The patient will continue to participate in the randomized controlled trial and will take the investigational drug for the next three months. No change in medications will be made other than adding the investigational drug. The patient will be monitored closely for any changes in his seizure activity or adverse effects. Follow-up appointments will be scheduled as needed.

----------------END OF PATIENT ENCOUNTER---------------

-------------- PATIENT ID 93 -- VISIT 2 -----------

Neurology Clinic Note - Visit #2 (2022-06-17)

Patient Information:

* Patient ID: 93

* Age: 66 years old

* Gender: Male

* History of: Frontal lobe epilepsy

Reason for Visit:

* The patient is in a randomized controlled trial and has been taking the investigational drug ID X93461 for epilepsy over the past 3 months.

Chief Complaints:

* Somnolence

* Fatigue

Past Medical History:

* Frontal lobe epilepsy

Current Medications:

* ID X93461 (investigational drug) (discontinued)

* Levetiracetam ( Keppra )

Social History:

* No significant social history

Family History:

* No significant family history

Review of Systems:

* General: No fever, weight loss, or other constitutional symptoms.

* Neurological: No new neurological symptoms. The patient reports continued seizure activity with 15.0 seizures over the past 3 months while taking the investigational drug.

* Other: No other significant symptoms.

Physical Examination:

* General: The patient appears well-nourished and well-oriented.

* Neurological:

+ Mental status: The patient's mental status is intact, with no evidence of cognitive decline or confusion.

+ Cranial nerves: All cranial nerves are intact, with no abnormalities noted.

+ Motor examination: There is no evidence of weakness, atrophy, or other motor abnormalities.

+ Sensory examination: There is no evidence of sensory loss or abnormalities.

+ Coordination and balance: The patient's coordination and balance are intact.

* Other: No other significant physical examination findings.

Assessment and Plan:

* The patient has been participating in a randomized controlled trial and has experienced continued seizure activity while taking the investigational drug ID X93461 over the past 3 months.

* Based on the patient's history of frontal lobe epilepsy and continued seizure activity, it is recommended that the patient continue with current medications (Levetiracetam) and undergo regular monitoring for seizure control.

* The investigational drug ID X93461 will be discontinued due to the completion of the study.

* The patient will be scheduled for follow-up appointments to monitor seizure activity and adjust medications as needed.

Note: This clinic note is a fictional example and does not reflect any real patient or medical condition.

----------------END OF PATIENT ENCOUNTER---------------

-------------- PATIENT ID 94 -- VISIT 1 -----------

Patient ID: 94

Visit Date: 2021-12-10

Identification:

* Name: [Name], [Initials]

* Date of Birth: [Date of Birth]

* Gender: Female

* Age: 49 years

History of Present Illness (HPI):

* The patient presents with a history of diabetes and multifocal epilepsy.

* She has been participating in a randomized controlled trial for the treatment of her epilepsy.

* She has had no seizures in the past two months.

Subjective:

* The patient reports that she has been feeling well since her last visit.

* She denies any changes in her symptoms or medications.

* She mentions that she has been experiencing some fatigue, but this is not a significant change from her previous state.

Medications:

* Levetiracetam (Keppra) 500mg QID

* Lamotrigine (Lamictal) 200mg OD

* Valproic acid (Depakote) 1000mg BID

Allergies:

* None reported

Social History:

* Married with two children

* Works as a teacher

* Denies any recent travel or illness

Review of Systems:

* No significant changes in the patient's medical history, social history, or review of systems.

General Physical Examination:

* Vital signs:

+ Blood pressure: 120/80 mmHg

+ Heart rate: 70 bpm

+ Temperature: 98.6°F (37°C)

* Head and neck examination:

+ No abnormalities noted

* Extremity examination:

+ No abnormalities noted

* Sensory examination:

+ No abnormalities noted

* Mental status examination:

+ Alert and oriented x3

Comprehensive Neurologic Exam:

* Cranial nerves:

+ II-XII intact

* Motor examination:

+ No abnormalities noted

* Sensory examination:

+ No abnormalities noted

* Coordination and balance examination:

+ No abnormalities noted

* Reflexes:

+ 2+ (moderate) extensor reflexes in upper and lower extremities

Labs:

* Complete Blood Count (CBC):

+ WBC: 4.5 x 10^9/L (normal range: 4.5-11 x 10^9/L)

+ Hemoglobin: 12.0 g/dL (normal range: 12.0-16.0 g/dL)

+ Hematocrit: 38.0% (normal range: 35.0-50.0%)

* Electrolyte panel:

+ Sodium: 142 mmol/L (normal range: 136-145 mmol/L)

+ Potassium: 4.2 mmol/L (normal range: 3.5-5.0 mmol/L)

+ Chloride: 98 mmol/L (normal range: 95-110 mmol/L)

* Renal function panel:

+ Creatinine: 0.7 mg/dL (normal range: 0.6-1.2 mg/dL)

+ Urinalysis:

- No significant abnormalities noted

Imaging:

* Most recent MRI of the brain:

+ No significant abnormalities noted

Assessment:

* The patient presents with a history of diabetes and multifocal epilepsy, and is currently participating in a randomized controlled trial for the treatment of her epilepsy.

* She has had no seizures in the past two months.

* Her neurological examination reveals no significant changes.

* Her laboratory results are within normal limits.

* Her imaging studies show no significant abnormalities.

Plan:

* Continue the patient's current medications for her diabetes and epilepsy, including levetiracetam, lamotrigine, and valproic acid.

* Add the investigational drug for epilepsy, as per the randomized controlled trial protocol.

* Monitor the patient's seizure activity and adjust her medications as needed.

* Schedule a follow-up visit in two months to assess her progress and make any necessary changes to her treatment plan.

Signature: [Your Signature]

Date: 2021-12-10

----------------END OF PATIENT ENCOUNTER---------------

-------------- PATIENT ID 94 -- VISIT 2 -----------

Clinic Note: Visit #2 - 2022-01-10

Patient ID: 94

Age: 49 years

Gender: Female

Chief Complaint: Gait disturbance, history of diabetes and multifocal epilepsy

History of Present Illness: The patient has been participating in a randomized controlled trial for the investigational drug ID X93461 for epilepsy over the past 3 months. She has experienced 3.0 seizures during this time. The patient also reports gait disturbance.

Past Medical History: Diabetes mellitus type 2, multifocal epilepsy

Medications: Levetiracetam (500mg BID), Lamotrigine (100mg QD), Metformin (1000mg BID)

Social History: Denies alcohol and tobacco use. Married with two children.

Review of Systems: Negative for fever, headache, or other neurological symptoms.

Neurological Examination:

General: Alert and oriented x3.

Head: Normocephalic, normochromic. No scalp edema or lesions.

Eyes: Pupils equal and reactive to light. No evidence of afferent pupillary defect. Extraocular movements intact. Funduscopic examination reveals no abnormalities.

Face: Mild weakness of the left facial muscles.

Limbs: Mild ataxia with a wide-based gait. No evidence of tremors or rigidity. Power and reflexes are normal in all extremities.

Sensory: No deficits in sensation to light touch, pain, or temperature. Vibratory sensation is intact.

Cranial Nerves: II-XII are intact.

Motor: Mild weakness of the left facial muscles.

Assessment and Plan:

1. The patient's history of diabetes and multifocal epilepsy, along with her current gait disturbance and mild weakness of the left facial muscles, suggest a possible diagnosis of diabetic neuropathy or seizure-related peripheral nerve injury.

2. The patient's recent participation in a randomized controlled trial for ID X93461 for epilepsy may have contributed to her current symptoms. However, since the study is now complete and the investigational drug will be discontinued, we will monitor the patient closely for any changes in her condition.

3. The patient's seizure activity will be closely monitored, and she will be advised to continue her current anticonvulsant medications.

4. The patient will undergo further evaluation and testing to confirm the diagnosis and determine the appropriate course of treatment. This may include electromyography (EMG), nerve conduction studies (NCS), or imaging studies such as magnetic resonance imaging (MRI) or computed tomography (CT).

5. The patient will be scheduled for follow-up appointments to monitor her condition and adjust her treatment plan as needed.

Note: This clinic note is fictional and does not reflect any real patient or study. It is meant solely for educational purposes and should not be used for actual patient care.

----------------END OF PATIENT ENCOUNTER---------------

-------------- PATIENT ID 95 -- VISIT 1 -----------

On March 23rd, 2022, Patient ID 95, a 47-year-old female with a history of alcoholism and multifocal epilepsy, visited our clinic for her first visit. The patient has experienced 14.0 seizures in the past two months, which is a significant increase from her previous seizure frequency. As part of a randomized controlled trial, the patient will be adding an investigational drug for epilepsy to her current medication regimen for a period of three months.

During the neurological examination, the patient was alert and cooperative. She reported no significant changes in her symptoms since her last visit, although she did mention experiencing some increased tremors in her hands and feet. Her speech was clear and concise, and there were no noticeable deficits in her cognitive function. The patient's mood was euthymic, and she denied any suicidal or homicidal ideation.

The patient's vital signs were within normal limits, with a blood pressure of 120/80 mmHg, a heart rate of 70 beats per minute, and a temperature of 98.6 degrees Fahrenheit. Her examination revealed no abnormalities in her head, eyes, ears, nose, throat, or skin. Her neurological reflexes were normal, and there was no evidence of ataxia or dysmetria. The patient's muscle strength and tone were within normal limits, and she did not exhibit any signs of weakness or wasting.

In conclusion, Patient ID 95 presented with a significant increase in seizure frequency, which is consistent with the patient's history of multifocal epilepsy. The addition of an investigational drug for epilepsy to her current medication regimen is expected to improve her seizure control and overall health outcomes. The patient's neurological examination revealed no significant changes or abnormalities, and her vital signs were within normal limits.

----------------END OF PATIENT ENCOUNTER---------------

-------------- PATIENT ID 95 -- VISIT 2 -----------

Clinic Note Visit #2, Patient ID 95, dated 2022-04-23:

Chief Complaint: The patient presents today with no complaints of seizures or symptoms since her last visit on 2022-03-18. She is currently participating in a randomized controlled trial evaluating the investigational drug ID X93461 for the treatment of multifocal epilepsy.

Past Medical History: The patient has a history of alcoholism and multifocal epilepsy, with a previous diagnosis of generalized seizures. She has been undergoing treatment with the investigational drug ID X93461 for the past 3 months, during which time she has experienced 13.0 seizures.

Physical Examination: On examination today, the patient's neurological status is normal, with no evidence of tremors, ataxia, or other abnormalities. Her speech is clear and coherent, and her eyes are normal. There is no evidence of weakness or paresis in her limbs. Her vital signs are within normal limits, including her blood pressure (120/80 mmHg), heart rate (70 beats per minute), respiratory rate (14 breaths per minute), and temperature (36.8°C).

Plan: The patient will discontinue the investigational drug ID X93461 now that the study is complete. Her seizure activity will continue to be monitored, and she will return for follow-up visits as needed. Her blood work and other laboratory tests will also be checked as necessary.

----------------END OF PATIENT ENCOUNTER---------------

-------------- PATIENT ID 96 -- VISIT 1 -----------

Clinic Note Visit #1

Patient ID: 96

Date: 2022-12-05

Identification:

Name: John Doe

Age: 59 years

Gender: Male

Chief Complaint:

Mr. Doe presents with a history of schizophrenia, bipolar disorder, diabetes, and frontal lobe epilepsy. He has been experiencing an increased frequency of seizures over the past 2 months, with 59.0 seizures reported.

History of Present Illness (HPI):

Mr. Doe reports that he has been experiencing more frequent seizures in the past 2 months, with an average of 3-4 seizures per week. He also notes that he has had no recent changes in his medications, including his anticonvulsant regimen. Additionally, Mr. Doe informs that he is currently participating in a randomized controlled trial for the treatment of his epilepsy.

Subjective:

During today's visit, Mr. Doe appears to be in no acute distress and is able to communicate clearly and effectively. He reports that he has been experiencing some confusion and disorientation during his seizures, but has not experienced any aura or warning signs prior to their onset.

Medications:

Mr. Doe is currently prescribed the following medications:

* Lithium carbonate (600 mg orally BID) for bipolar disorder

* Olanzapine (10 mg orally QD) for schizophrenia

* Levetiracetam (500 mg orally BID) and valproic acid (500 mg orally BID) for frontal lobe epilepsy

* Insulin glargine (1 unit/kg subcutaneously QD) for diabetes

Allergies:

Mr. Doe reports no known allergies to any medications.

Social History:

Mr. Doe is a retired mechanic and currently resides with his wife of 30 years. He denies any history of substance abuse or smoking. He reports that he has a strong support system, including his spouse and adult children.

Review of Systems:

Mr. Doe reports no significant changes in his review of systems since his last visit. He denies any current symptoms of depression, anxiety, or other psychiatric disturbances.

General Physical Examination:

On today's examination, Mr. Doe appears to be in no acute distress and has a normal appearance. His vital signs are as follows:

* Blood pressure: 120/80 mmHg

* Pulse: 70 beats per minute

* Temperature: 98.6°F (37°C)

* Weight: 175 lbs (79.4 kg)

Comprehensive Neurological Examination:

Mr. Doe underwent a comprehensive neurological examination, including assessment of his cognitive function, sensory motor function, and reflexes. His examination revealed the following findings:

* Cognitive function: Mr. Doe demonstrates normal cognitive function, with intact memory, attention, and concentration.

* Sensory motor function: Mr. Doe has intact sensory motor function, with no signs of weakness or paresis.

* Reflexes: Mr. Doe's reflexes are normal, with no signs of hyperexplexia or other abnormalities.

Labs:

Mr. Doe's laboratory results are as follows:

* Complete blood count (CBC): Within normal limits

* Electrolyte panel: Within normal limits

* Renal function panel: Within normal limits

* Thyroid-stimulating hormone (TSH) level: 4.2 mU/L (normal: 0.4-4.0 mU/L)

Imaging:

Mr. Doe has undergone imaging studies, including a computed tomography (CT) scan of his brain and an electroencephalogram (EEG), which have revealed the following findings:

* CT scan of the brain: No evidence of any acute or chronic changes in brain structure.

* EEG: No evidence of any seizure activity or other abnormalities.

Assessment:

Based on Mr. Doe's history, physical examination, and laboratory results, he is diagnosed with schizophrenia, bipolar disorder, diabetes, and frontal lobe epilepsy. His increased frequency of seizures over the past 2 months is concerning, and further evaluation and management are necessary to address this issue.

Plan:

* Mr. Doe will continue his current medication regimen, with no changes made at this time.

* He will participate in a randomized controlled trial for the treatment of his epilepsy, as previously discussed.

* His seizure frequency will be closely monitored, and any changes will be addressed promptly.

* His diabetes management will be optimized, with close monitoring of his blood sugar levels and any necessary adjustments to his medication regimen.

Signature:

[Your signature]

Date: 2022-12-05

----------------END OF PATIENT ENCOUNTER---------------

-------------- PATIENT ID 96 -- VISIT 2 -----------

Visit #2: January 5, 2023

Patient ID 96, a 59-year-old male with a history of schizophrenia, bipolar disorder, diabetes, and frontal lobe epilepsy, was seen in the clinic for his second visit. The patient has been participating in a randomized controlled trial (RCT) evaluating the investigational drug ID X93461 for the treatment of epilepsy. Over the past three months, the patient has taken the drug and has reported no symptoms.

During the neurological examination, the patient was alert and oriented with no evidence of seizure activity or other neurological deficits. His speech was clear and coherent, and his motor function was normal. The patient's cognitive function was also assessed using a series of tests, including the Mini-Mental State Examination (MMSE), and results indicated that his cognitive function is intact.

The patient has experienced 25.0 seizures since initiating treatment with ID X93461, which is within the expected range for this type of study. However, as the RCT is now complete, the investigational drug will be discontinued. The patient will be closely monitored for any adverse effects or changes in his condition.

In conclusion, visit #2 revealed no significant changes in the patient's neurological function or cognitive status. The patient has tolerated the investigational drug well and has experienced a reasonable number of seizures during the study period. As the RCT is complete, the drug will be discontinued, and the patient will be closely monitored for any changes in his condition.

----------------END OF PATIENT ENCOUNTER---------------

-------------- PATIENT ID 97 -- VISIT 1 -----------

Clinic Note Visit #1

Patient ID: 97

Date: 2022-11-22

Identification:

Mr. John Doe, a 60-year-old male, presents with a history of intractable partial seizures with secondary generalization and parietal lobe epilepsy. He has experienced 17.0 seizures in the past two months.

Highlights of the Patient's History:

* Patient has a history of intractable partial seizures with secondary generalization

* Patient has parietal lobe epilepsy

* Patient has experienced 17.0 seizures in the past two months

* Patient is currently enrolled in a randomized controlled trial (RCT) for the treatment of epilepsy

Subjective:

During today's visit, Mr. Doe reported that he has been experiencing frequent seizures over the past two months. He also noted that he has been experiencing some cognitive difficulties, including memory lapses and difficulty with concentration. Additionally, he reported that he has been experiencing some mood changes, including irritability and anxiety.

Medications:

* Levetiracetam (Keppra) 500mg BID

* Lamotrigine (Lamictal) 200mg OD

* Valproic acid (Depakote) 1000mg TID

Allergies:

* None reported

Social History:

* Married with two adult children

* Occupation: Retired

* Living situation: Patient lives alone with his spouse

Review of Systems:

* General: No recent illnesses or hospitalizations

* Neurological: Frequent seizures over the past two months, including both partial and secondary generalization seizures

* Musculoskeletal: No reported musculoskeletal complaints

* Cardiovascular: No reported cardiovascular complaints

* Respiratory: No reported respiratory complaints

General Physical Examination:

* Vital signs: Blood pressure 120/80 mmHg, heart rate 70 bpm, temperature 98.6°F (37°C), and weight 75 kg

* Head: No focal deficits or abnormalities noted

* Eyes: No abnormalities noted

* Ears: No abnormalities noted

* Nose: No abnormalities noted

* Throat: No abnormalities noted

* Neck: No abnormalities noted

* Chest: No abnormalities noted

* Abdominal: No abnormalities noted

* Extremities: No abnormalities noted

Comprehensive Neurologic Examination:

* Mental status: Alert and oriented to person, place, and time

* Cranial nerves: Intact

* Motor function: No signs of weakness or paralysis

* Sensory function: No signs of sensory loss or dysfunction

* Coordination and balance: No signs of ataxia or dizziness

Labs:

* Complete Blood Count (CBC): Within normal limits

* Basic Metabolic Panel (BMP): Within normal limits

* Thyroid-stimulating hormone (TSH): 2.5 mIU/L (normal 0.4-4.0 mIU/L)

Imaging:

* Computed Tomography (CT) scan of the brain: No abnormalities noted

Assessment:

* Mr. Doe is a 60-year-old male with a history of intractable partial seizures with secondary generalization and parietal lobe epilepsy. He has experienced 17.0 seizures in the past two months and is currently enrolled in an RCT for the treatment of epilepsy.

* The patient's neurological examination is within normal limits, with no focal deficits or abnormalities noted.

* The patient's laboratory results are within normal limits, with the exception of slightly elevated TSH levels.

* The patient's imaging studies are within normal limits, with no abnormalities noted.

Plan:

* Continue the current medication regimen, including levetiracetam (Keppra) 500mg BID, lamotrigine (Lamictal) 200mg OD, and valproic acid (Depakote) 1000mg TID.

* Add the investigational drug for epilepsy, lacosamide (Vimpat), at a dose of 200mg BID for a period of three months.

* Monitor the patient's seizure activity and adjust the medication regimen as needed.

Signature:

[Your Signature]

Date: 2022-11-22

----------------END OF PATIENT ENCOUNTER---------------

-------------- PATIENT ID 97 -- VISIT 2 -----------

Patient ID: 97

Visit #2: 2022-12-22

History:

* Parietal lobe epilepsy

* History of seizures (12.0) over past 3 months while taking investigational drug ID X93461

* Reported balance disorder

Neurological Examination:

* Mild weakness in left arm and leg

* Sensory deficits in the form of hypesthesia (decreased sensation) in the left arm and leg

* Negative Romberg test (indicates unsteadiness)

* Positive Hoffmann's reflex (indicates increased muscle tone) in both arms

* Normal cerebellar function

Assessment:

* Seizure activity likely related to investigational drug ID X93461

* Balance disorder possibly related to seizure activity or drug side effects

Plan:

* Discontinue investigational drug ID X93461

* Monitor seizure activity and balance problems closely

* Consider alternative treatments for epilepsy if seizures persist

----------------END OF PATIENT ENCOUNTER---------------

-------------- PATIENT ID 98 -- VISIT 1 -----------

Visit #1 (2022-01-01)

Patient ID: 98

Age: 74 years

Female

History:

* Autism

* Bipolar disorder

* Alcoholism

* Parietal lobe epilepsy (7.0 seizures in past 2 months)

* Randomized controlled trial (no change in medications except addition of investigational drug for epilepsy)

Neurological Examination:

* Mental status: alert and oriented

* Speech: clear and coherent

* Motor examination: normal gross motor skills, brisk reflexes

* Sensory examination: normal sensation to light touch and pinprick in all extremities

* Cranial nerve examination: normal

* Coordination and balance: normal

Pertinent Positives:

* No change in seizure activity since last visit

* Patient reports no side effects from investigational drug

Assessment and Plan:

* Continue investigational drug for epilepsy for 3 months

* Monitor patient's seizure activity and side effects closely

* Schedule follow-up visit in 1 month to assess response to treatment.

----------------END OF PATIENT ENCOUNTER---------------

-------------- PATIENT ID 98 -- VISIT 2 -----------

Clinic Note - Visit #2

Patient ID: 98

Date: 2022-02-01

This is a follow-up visit for Patient ID 98, a 74-year-old female with a history of autism, bipolar disorder, alcoholism, and parietal lobe epilepsy. The patient has been participating in a randomized controlled trial (RCT) and has been receiving the investigational drug ID X93461 for the past three months to control her seizures.

During this visit, the patient reported experiencing 3.0 seizures since starting the medication. She also complained of dizziness, somnolence, fatigue, dysarthria, diplopia, and gait disturbance. These side effects are consistent with the known adverse effects of ID X93461.

On examination, the patient had a complete neurological examination which revealed no significant changes in her cognitive or motor functions compared to the previous visit. However, there was mild weakness in her left arm and leg, which may be related to the drug's side effects. Her speech was also slightly slurred, which may indicate dysarthria.

Given that the RCT is now complete, it has been decided to discontinue the investigational drug ID X93461 for Patient ID 98. The patient will be closely monitored for any changes in her seizure activity and other symptoms over the next few weeks. Any further treatment or interventions will be based on the results of this monitoring period.

----------------END OF PATIENT ENCOUNTER---------------

-------------- PATIENT ID 99 -- VISIT 1 -----------

Patient ID: 99

Date: 2023-02-23

History:

* 76 year old male

* History of parietal lobe epilepsy

* 55 seizures in past 2 months

* Randomized controlled trial (no change in medications except for investigational drug for epilepsy)

Neurological Examination:

* Alert and oriented

* No significant headache, fever, or other complaints

* Normal hearing and vision

* Normal strength and sensation in all extremities

* No tremors or rigidity

* No abnormal reflexes

Pertinent Positives:

* No change in seizure activity since last visit

* Continues to experience 5-10 seizures per day

Assessment and Plan:

* Continue randomized controlled trial with investigational drug for epilepsy

* Monitor seizure activity and adjust dosage as needed

* Follow up in 1 month to assess efficacy and tolerability of investigational drug.

----------------END OF PATIENT ENCOUNTER---------------

-------------- PATIENT ID 99 -- VISIT 2 -----------

On March 23rd, 2023, I evaluated Patient ID 99, a 76-year-old male with a history of parietal lobe epilepsy, during visit #2 of our randomized controlled trial. Over the past three months, the patient has taken the investigational drug ID X93461 for his epilepsy and has experienced 33.0 seizures during this time. The patient has also reported constipation and diplopia as adverse effects of the medication.

Upon examination, the patient's neurological status was as follows: His speech was clear and coherent, with no evidence of aphasia or dysarthria. His motor function was intact, with no signs of weakness or paralysis. His sensory function was normal, with no deficits in vision, hearing, or somatosensation. His reflexes were normal and there was no evidence of ataxia or dyskinesia. His cranial nerve examination revealed no abnormalities, including no evidence of diplopia.

During the study, the patient experienced 33 seizures, which is a significant increase from his baseline rate. This finding suggests that the investigational drug ID X93461 may not be effective in controlling the patient's epilepsy. In light of this result, we have decided to discontinue the medication now that the study is complete. The patient will be transitioned back to his previous medication and will be monitored closely for any changes in his seizure activity.

In conclusion, visit #2 of our randomized controlled trial has provided valuable information about the effectiveness and safety of the investigational drug ID X93461 for the treatment of parietal lobe epilepsy. Although the medication showed promise in controlling the patient's seizures, the adverse effects of constipation and diplopia were significant. Based on these findings, we have decided to discontinue the medication and transition the patient back to his previous therapy. Further analysis of the data will be necessary to determine the long-term efficacy and safety of this medication for this indication.

----------------END OF PATIENT ENCOUNTER---------------

-------------- PATIENT ID 110 -- VISIT 1 -----------

**Patient Information:**

Patient ID: 110

Date of Visit: 2021-06-28

Age: 25 years

Gender: Female

**Chief Complaint:**

The patient presents with a history of seasonal allergies, anxiety, and parietal lobe epilepsy. She has experienced 69.0 seizures in the past 2 months.

**Past Medical History:**

The patient has a history of parietal lobe epilepsy and has been receiving treatment with levetiracetam and lacosamide. She also has a history of seasonal allergies and anxiety.

**Current Medications:**

The patient is currently taking levetiracetam and lacosamide for her epilepsy, and alprazolam for her anxiety.

**Social History:**

The patient is currently participating in a randomized controlled trial for her epilepsy. She denies any recent travel or exposure to illness.

**Physical Examination:**

General: The patient appears well-nourished and well-developed. Her speech is clear and she has a normal tone of voice. Her affect is anxious.

Neurological:

* Mental Status: The patient's mental status is normal. She is able to answer questions appropriately and follows commands.

* Cranial Nerves: The patient's cranial nerves are intact, including visual fields, hearing, taste, and smell.

* Motor: The patient has a normal range of motion in all extremities. Her muscle strength is 5/5 in all groups.

* Sensory: The patient has a normal sensation in all extremities.

* Coordination and Balance: The patient's coordination and balance are normal.

* Reflexes: The patient's reflexes are normal.

**Assessment and Plan:**

Based on the patient's history of parietal lobe epilepsy, seasonal allergies, and anxiety, as well as her recent increase in seizures, I recommend the following:

1. Continue current medications for epilepsy and anxiety.

2. Add an investigational drug for epilepsy for a 3-month trial period.

3. Monitor the patient's seizure activity and adjust medications as needed.

4. Refer the patient to a specialist for further management of her anxiety and seasonal allergies.

5. Schedule a follow-up appointment in 3 months to assess the patient's progress and adjust her treatment plan as needed.

I have discussed the risks and benefits of the investigational drug with the patient, and she has given informed consent for its use. I will be monitoring the patient's seizure activity closely during the trial period and will make any necessary adjustments to her medications.

----------------END OF PATIENT ENCOUNTER---------------

-------------- PATIENT ID 110 -- VISIT 2 -----------

Clinic Note: Visit #2 - July 28, 2021

Patient ID: 110 (25 year old female)

Chief Complaint: Seasonal allergies, anxiety, and parietal lobe epilepsy

History of Present Illness: The patient has been participating in a randomized controlled trial for the investigational drug ID X93461 for epilepsy over the past 3 months. During this time, she has experienced 30.0 seizures and reported somnolence and balance disorder. The study is now complete and the investigational drug will be discontinued.

Past Medical History: Seasonal allergies, anxiety, parietal lobe epilepsy

Medications: none

Social History: none

Family History: none

Review of Systems:

Neurological: The patient reports ongoing somnolence and balance disorder. She has experienced 30.0 seizures over the past 3 months, with no changes in frequency or severity.

Allergic: Seasonal allergies are well-controlled with over-the-counter medication.

Musculoskeletal: None

Cardiovascular: None

Respiratory: None

Gastrointestinal: None

Urinary: None

Neurological Examination:

Mental Status: Alert and oriented x3

Speech: Normal rate and tone

Language: Intact

Memory: Intact

Attention and Concentration: Intact

Executive Function: Intact

Cerebellar Function: Intact

Sensory Examination:

Visual: Full fields of vision, no abnormalities noted

Auditory: Normal hearing in both ears

Tactile: Intact

Proprioception: Intact

Interoception: Intact

Motor Examination:

Upper extremities: Intact

Lower extremities: Intact

Gait and balance: Unsteady, with reported balance disorder

Reflexes: 2+ (brisk)

Assessment and Plan:

The patient's ongoing somnolence and balance disorder are concerning and may be related to the investigational drug ID X93461. The patient has experienced a significant increase in seizure frequency during the study, which may also be related to the drug. Further evaluation and management of these symptoms will be necessary.

Given the completion of the study, the investigational drug ID X93461 will be discontinued. The patient's epilepsy medication will be re-evaluated and adjusted as needed.

The patient will be scheduled for a follow-up appointment in 2-3 weeks to assess for any changes in symptoms and to discuss further management of her epilepsy and balance disorder. In the interim, the patient is advised to continue with her current medications and to monitor her seizure frequency closely.

The patient will also be referred to a specialist for ongoing management of her balance disorder.

Note: The patient's seasonal allergies are well-controlled with over-the-counter medication, but should be re-evaluated in the spring to ensure optimal control.

----------------END OF PATIENT ENCOUNTER---------------

-------------- PATIENT ID 111 -- VISIT 1 -----------

---

Clinic Note - Visit #1 (2022-08-02)

Patient ID: 111

Name: [Patient's Name]

Age: 27 years old

Gender: Male

Chief Complaint: Recent increase in seizure activity

History of Present Illness:

The patient, a 27-year-old male with a history of autism and occipital lobe epilepsy, has experienced an alarming increase in seizure activity over the past two months. He has had 30.0 seizures during this time period, significantly exceeding his usual frequency. The patient is currently enrolled in a randomized controlled trial (RCT) and has been adherent to his medication regimen without any changes. However, due to the recent increase in seizure activity, it has been decided that the patient will be added to the investigational drug for epilepsy for a period of three months.

Identification:

Patient ID: 111

Name: [Patient's Name]

Date of Birth: [Patient's Date of Birth]

Contact Information: [Patient's Contact Information]

Highlights of the Patient's History:

* Autism diagnosis at age 3

* Occipital lobe epilepsy diagnosis at age 10

* Previous seizure frequency was 5-6 per month

* Currently enrolled in an RCT for epilepsy medication

Highlights of the Patient's Current Symptoms:

* Increased seizure activity over the past two months (30.0 seizures)

* No changes in medications other than addition of investigational drug for epilepsy

Subjective:

The patient reports an increased frequency of seizures over the past two months, with an average of 3-4 seizures per week. He also mentions that he has been experiencing more frequent and intense auras preceding his seizures. The patient denies any recent changes in medications or other potential triggers for his seizure activity.

Medications:

* Levetiracetam (Keppra) 500mg BID

* Lamotrigine (Lamictal) 200mg OD

* Valproate (Depakote) 1000mg TID

Allergies:

* None reported

Social History:

* Married with two children (ages 4 and 6)

* Works as a graphic designer

* Denies alcohol or drug use

* Reports good sleep hygiene (7-8 hours per night)

Review of Systems:

* No significant changes in the patient's systemic symptoms or medical history

General Physical Examination:

* Vital signs within normal limits

* Head, neck, and extremities examination reveals no abnormalities

* Full neurological examination is documented below

Comprehensive Neurologic Examination:

* Mental status: Alert and oriented x3

* Cranial nerve examination: Normal

* Motor examination: No significant abnormalities

* Sensory examination: Normal

* Coordination and balance examination: Normal

* Reflexes: 2+ (normal)

Labs:

* Complete Blood Count (CBC): Within normal limits

* Electrolyte Panel: Within normal limits

* Liver Function Tests (LFTs): Within normal limits

* Thyroid-stimulating hormone (TSH): Within normal limits

Imaging:

* Magnetic Resonance Imaging (MRI) of the brain performed on [Date] revealed no significant abnormalities or changes in the patient's occipital lobe epilepsy.

Assessment:

The patient's recent increase in seizure activity is concerning, and it is essential to investigate the cause and potential triggers for this increase. The addition of the investigational drug for epilepsy will help determine its efficacy in reducing seizure frequency and severity. The patient's current medications will remain unchanged, as they have been effective in managing his seizures thus far. The patient's cognitive function and mental status are within normal limits.

Plan:

* Continue the patient's current medication regimen without any changes

* Add the investigational drug for epilepsy for a period of three months

* Monitor the patient's seizure activity and adjust medications as needed

* Schedule follow-up appointments at [Frequency] to assess the patient's progress and adjust the treatment plan accordingly

Signature:

[Your Signature]

Date: [Current Date]

Time: [Current Time]

---

I hope this helps! Let me know if you have any further questions or need any additional assistance.

----------------END OF PATIENT ENCOUNTER---------------

-------------- PATIENT ID 111 -- VISIT 2 -----------

Patient ID: 111

Visit #2: 2022-09-02

Identifying Information:

* 27 year old male

* History of autism and occipital lobe epilepsy

* In a randomized controlled trial (ID X93461)

Highlights of the Visit:

* Patient has been taking investigational drug ID X93461 for epilepsy over the past 3 months

* Reported 16.0 seizures during this time

* Experienced somnolence and vomiting as adverse effects

Neurological Examination:

* Alert and oriented

* No focal neurological deficits

* Normal strength and tone in all muscle groups

* Normal sensation in all extremities

Pertinent Positives:

* No changes in seizure frequency or severity since last visit

* Continued somnolence and vomiting as adverse effects

Assessment and Plan:

* Discontinue investigational drug ID X93461 due to completion of study

* Monitor for any changes in seizure frequency or severity

* Address ongoing somnolence and vomiting with appropriate medication adjustments.

----------------END OF PATIENT ENCOUNTER---------------

-------------- PATIENT ID 112 -- VISIT 1 -----------

On September 29th, 2021, Patient ID 112, a 42-year-old female, visited our clinic for her first follow-up visit since enrolling in a randomized controlled trial for the treatment of her parietal lobe epilepsy. The patient has a history of seasonal allergies and has experienced 28.0 seizures in the past two months.

Upon examination, the patient was alert and coherent, with no evidence of any recent seizure activity. Her neurological examination revealed no abnormalities, including no focal neurological deficits, cranial nerve abnormalities, or muscle weakness. The patient's speech was clear and comprehensible, and her language skills were intact.

The patient reported no changes in her symptoms since the previous visit, and there were no significant changes in her medication regimen. The investigational drug for epilepsy was added to her regimen, and the patient will take this medication for a period of three months as part of the randomized controlled trial.

In conclusion, Patient ID 112 had a normal neurological examination, with no evidence of recent seizure activity or any other abnormalities. The addition of the investigational drug for epilepsy to her medication regimen was deemed appropriate, and the patient will continue to be monitored closely as part of the randomized controlled trial.

----------------END OF PATIENT ENCOUNTER---------------

-------------- PATIENT ID 112 -- VISIT 2 -----------

---

Clinic Note - Visit #2 (2021-10-29)

Patient ID: 112 (Female, 42 years old)

Identification:

* Name: [Insert name here]

* Date of birth: [Insert date of birth here]

* Contact information: [Insert contact information here]

HPI:

* Patient presents with a history of seasonal allergies and parietal lobe epilepsy.

* Patient has been participating in a randomized controlled trial for the investigational drug ID X93461 for epilepsy over the past 3 months.

* Patient reported experiencing dizziness, nausea, ataxia, and falling on multiple occasions.

Subjective:

* Patient reports experiencing 17.0 seizures since starting the investigational drug.

* Patient describes feeling dizzy and nauseous, with a sense of ataxia (impaired coordination and balance) and falling on multiple occasions.

* Patient reports no changes in sleep patterns or appetite.

Medications:

* ID X93461 (investigational drug for epilepsy) - discontinued due to completion of the study.

* Levetiracetam (Keppra) - maintained at 2000mg/day.

* Lorazepam (Ativan) - as needed for seizure activity.

Allergies:

* None reported.

Social History:

* Married with two children (ages 8 and 10).

* Occupation: Elementary school teacher.

* Denies history of substance abuse or psychiatric illness.

* Lives in a two-story house with carpeting and no pets.

Review of Systems:

* General: Patient appears well-nourished and well-hydrated.

* Neurological: No changes in seizure activity or medication adherence.

* Musculoskeletal: No joint pain or muscle weakness.

* Cardiovascular: No chest pain or shortness of breath.

* Respiratory: No shortness of breath or cough.

* Gastrointestinal: No abdominal pain or diarrhea.

General Physical Examination:

* Vital signs:

+ Temperature: 98.6°F (37°C)

+ Blood pressure: 120/80 mmHg

+ Heart rate: 70 beats per minute

+ Respiratory rate: 14 breaths per minute

* Head: Normocephalic, normal symmetrical facial features.

* Eyes: No abnormalities noted.

* Ears: No abnormalities noted.

* Nose: No nasal discharge or deformities.

* Mouth: Normal dentition and oral hygiene.

* Neck: Normalsized, no lymphadenopathy or masses.

* Chest: Clear to auscultation, no adventitious sounds.

* Abdomen: Soft and normoactive.

* Extremities: No clubbing or cyanosis.

Comprehensive Neurologic Examination:

* Cranial nerves: Intact.

* Motor exam: No abnormalities noted.

* Sensory exam: No abnormalities noted.

* Reflexes: Normal.

* Gait: Normal.

Labs:

* Complete Blood Count (CBC): Within normal limits.

* Basic Metabolic Panel (BMP): Within normal limits.

* Thyroid-stimulating hormone (TSH): Within normal limits.

Imaging:

* No imaging studies performed or ordered at this time.

Assessment:

* Patient presents with a history of seasonal allergies and parietal lobe epilepsy, and has been participating in a randomized controlled trial for the investigational drug ID X93461 for epilepsy over the past 3 months.

* Patient experienced dizziness, nausea, ataxia, and falling on multiple occasions, which may be related to the investigational drug.

* Patient's seizure activity remains stable, with no changes in medication adherence or sleep patterns.

* Patient is a good candidate for continued participation in the study, and the investigational drug will be discontinued now that the study is complete.

Plan:

* Continue Levetiracetam (Keppra) at 2000mg/day.

* Discontinue ID X93461 (investigational drug for epilepsy).

* Monitor patient's seizure activity and medication adherence closely.

* Schedule follow-up appointment in 3 months to assess for any changes in seizure activity or medication adherence.

Signature:

[Insert signature here]

---

Note: The above clinic note is an example of a comprehensive neurological examination and assessment for a patient with parietal lobe epilepsy, and should not be used as a basis for actual patient care. It is important to consult with a qualified healthcare professional before making any decisions or recommendations based on this information.

----------------END OF PATIENT ENCOUNTER---------------

-------------- PATIENT ID 113 -- VISIT 1 -----------

Clinic Note #1 (2023-02-20)

Patient ID: 113

Age: 83 years

Gender: Female

Chief Complaint: Recent increase in seizure frequency.

History of Present Illness: The patient, a 83-year-old female with a history of hypothyroidism, autism, dermatitis, and frontal lobe epilepsy, has experienced an increase in seizure frequency over the past two months, with 5.0 seizures occurring since her last visit on December 10, 2022. She is currently participating in a randomized controlled trial (RCT) and has been adherent to her medication regimen.

Identification:

Name: [Patient Name]

Date of Birth: [Date of Birth]

Contact Information: [Contact Information]

HPI: The patient presents with a long-standing history of frontal lobe epilepsy, hypothyroidism, autism, and dermatitis. She has been experiencing an increase in seizure frequency over the past two months, with 5.0 seizures occurring since her last visit on December 10, 2022.

Subjective:

The patient reports no changes in her medication regimen since her last visit. She has been adherent to her medications and has not experienced any significant side effects. She denies any recent illnesses or injuries that may be contributing to her seizure activity.

Medications:

* Levothyroxine (T4) 25 mcg PO daily

* Lamotrigine (LTG) 100 mg PO daily

* Valproic acid (VPA) 500 mg PO twice daily

* Rifampin (RIF) 600 mg PO daily

* Ethotoin (Pegloticase) 200 mg PO daily (as needed for seizure control)

Allergies:

* None reported

Social History:

The patient lives alone and has a support system consisting of her adult children. She is currently participating in an RCT and is under the care of a neurologist at another institution.

Review of Systems:

General: The patient appears well-preserved for her age, with no significant weight loss or gain. She reports no fever, chills, or other constitutional symptoms.

Neurological: The patient reports no new onset of numbness, weakness, or paresthesias. She denies any recent changes in her cognitive function or memory.

Musculoskeletal: The patient reports no joint pain or swelling. She does not have any significant muscle weakness or wasting.

Cardiovascular: The patient reports no chest pain, shortness of breath, or palpitations. Her blood pressure is 120/80 mmHg, and her heart rate is 70 beats per minute.

Respiratory: The patient reports no shortness of breath or wheezing. Her respiratory examination reveals clear lung sounds, with no evidence of rhonchi or wheezes.

Gastrointestinal: The patient reports no abdominal pain, nausea, or vomiting. She denies any changes in her bowel habits or appetite.

Urological: The patient reports no urinary frequency, urgency, or incontinence. Her last pap smear was negative for any abnormal cells.

General Physical Examination:

The patient appears well-preserved for her age, with no significant signs of decline in her cognitive or physical functioning. Her speech is clear and comprehensible, with no evidence of dysarthria or aphasia. Her motor examination reveals no signs of weakness, ataxia, or tremors. Her reflexes are normal, with no evidence of hyperreflexia or hyporeflexia.

Comprehensive Neurologic Exam:

The patient's neurological examination is largely unremarkable, with no signs of focal deficits or abnormalities in her sensory, motor, or cerebellar functions. Her gait is normal, with no evidence of ataxia or weakness. Her speech is clear and comprehensible, with no evidence of dysarthria or aphasia. Her cognitive function appears intact, with no signs of decline or impairment in her memory, attention, or executive functions.

Labs:

* Complete Blood Count (CBC): Within normal limits

* Basic Metabolic Panel (BMP): Within normal limits

* Thyroid Function Tests (TFTs): T4 5.0 mcg/dL (normal 4.5-11.5), T3 1.0 ng/dL (normal 0.7-2.0)

* Lipid Panel: Within normal limits

* Vitamin D Level: 20 ng/dL (normal 32-100)

Imaging:

* Computed Tomography (CT) scan of the brain: No evidence of intracranial abnormalities or lesions.

* Magnetic Resonance Imaging (MRI) of the brain: No evidence of structural abnormalities or lesions.

Assessment:

The patient presents with a history of frontal lobe epilepsy, hypothyroidism, autism, and dermatitis. She has been experiencing an increase in seizure frequency over the past two months, with 5.0 seizures occurring since her last visit on December 10, 2022. Her neurological examination is largely unremarkable, with no signs of focal deficits or abnormalities in her sensory, motor, or cerebellar functions. Her laboratory results are within normal limits, and her imaging studies do not reveal any intracranial abnormalities or lesions. Considering the patient's history of frontal lobe epilepsy and recent increase in seizure frequency, it is reasonable to assume that her seizures may be related to her underlying neurological condition.

Plan:

1. Continue the patient's current medication regimen, with close monitoring of her seizure activity and adjustments as needed.

2. Add the investigational drug (ID) for epilepsy, which will be taken for 3 months, to assess its efficacy in reducing seizure frequency.

3. Schedule a follow-up visit in two months to reassess the patient's seizure activity and adjust her medication regimen as needed.

4. Encourage the patient to maintain a consistent sleep schedule, avoid stimuli that may trigger seizures, and engage in regular physical activity to improve her overall health and well-being.

Signature:

[Your Signature]

Date: [Today's Date]

Note: This is just an example clinic note for a patient with a history of frontal lobe epilepsy, hypothyroidism, autism, and dermatitis. The specific details and content may vary depending on the patient's individual circumstances and the care provided.

----------------END OF PATIENT ENCOUNTER---------------

-------------- PATIENT ID 113 -- VISIT 2 -----------

Clinic Note: Visit #2, March 20, 2023

Patient ID: 113

Demographics:

* Age: 83 years old

* Gender: Female

Chief Complaint (CC):

* History of hypothyroidism, autism, dermatitis, and frontal lobe epilepsy

History of Present Illness (HPI):

* The patient has been participating in a randomized controlled trial for the investigational drug ID X93461 for epilepsy over the past 3 months

* The patient reported a decrease in seizure frequency from 5-6 per month to 3.0 per month during the study period

* The patient experienced dizziness and fell once during the study, resulting in a minor injury

Subjective:

* Patient reports difficulty with memory and concentration, especially during the study period

* Patient reports feeling "spacey" at times and experiencing visual disturbances

* Patient denies any recent changes in mood or behavior

Medications:

* ID X93461 (investigational drug for epilepsy) - discontinued as of today's visit

* Levothyroxine (thyroid hormone replacement) - ongoing

* Melatonin (sleep aid) - ongoing

Allergies:

* None reported

Social History:

* Married with adult children

* Retired teacher

* No significant travel or exposure history

Review of Systems (ROS):

* General: The patient appears well-appeared and well-nourished. She is alert and oriented to person, place, and time.

* Neurological: The patient reports difficulty with memory and concentration, feeling "spacey" at times, and experiencing visual disturbances.

* Musculoskeletal: The patient denies any musculoskeletal pain or limitations.

* Cardiovascular: The patient denies any cardiovascular symptoms or history of cardiovascular disease.

* Respiratory: The patient denies any respiratory symptoms or history of respiratory disease.

* Gastrointestinal: The patient denies any gastrointestinal symptoms or history of gastrointestinal disease.

General Physical Examination:

* Vital signs: Blood pressure - 120/80 mmHg, Pulse - 70 bpm, Respiratory rate - 14 breaths/min, Temperature - 98.6°F (37°C)

* Head: Normocephalic (normal head size), no focal swelling or tenderness

* Eyes: No abnormalities noted in the visual acuity or extraocular movements

* Ears: No abnormalities noted in the external auditory canals or tympanic membranes

* Nose: No abnormalities noted in the nasal passages or sinuses

* Throat: No abnormalities noted in the pharynx or larynx

* Chest: Clear to auscultation, no signs of respiratory distress or compromise

* Abdomen: Soft and non-tender, no masses or lesions appreciated

* Extremities: No clubbing or cyanosis noted in the fingers or toes

* Neurological Examination:

+ Cranial Nerves: Intact (including visual, olfactory, and somatosensory functions)

+ Motor Examination: No signs of weakness, atrophy, or abnormal movements

+ Sensory Examination: No signs of sensory loss or abnormalities

+ Reflexes: Present and normal

Comprehensive Neurological Exam:

* Cerebral hemispheres: No focal lesions or atrophy noted

* Cerebellum: Normocephalic, no signs of ataxia or dysmetria

* Brainstem: No signs of midline shift or focal lesions

* Spinal Cord: Not examined

Labs:

* Complete Blood Count (CBC): Within normal limits

* Electrolyte Panel (EKP): Within normal limits

* Thyroid Function Tests (TFTs): Levothyroxine levels within therapeutic range

Imaging:

* Computed Tomography (CT) scan of the head and spine - no abnormalities noted

Assessment:

* The patient's history of hypothyroidism, autism, dermatitis, and frontal lobe epilepsy, along with the recent participation in a randomized controlled trial for the investigational drug ID X93461, contribute to her complex and multifaceted neurological presentation.

* The patient's difficulty with memory and concentration, feeling "spacey" at times, and experiencing visual disturbances are concerning for potential cognitive and seizure-related effects of the investigational drug.

* The patient's history of falls and minor injuries is a concern for her overall safety and mobility.

* The patient's current medication regimen, including levothyroxine and melatonin, is appropriate for her diagnosed conditions.

Plan:

* Discontinue the investigational drug ID X93461 as of today's visit

* Continue levothyroxine and melatonin as prescribed

* Monitor the patient's seizure frequency and cognitive function closely over the next few months to assess for any potential effects of the discontinued drug

* Consider referral to a specialist (e.g., neuropsychologist, physical therapist) for further evaluation and management of the patient's cognitive and mobility issues

Signature:

[Your name and title]

Neurologist

----------------END OF PATIENT ENCOUNTER---------------

-------------- PATIENT ID 114 -- VISIT 1 -----------

Patient Name: John Doe

Patient ID: 114

Date of Visit: 2022-10-19

Chief Complaint:

Mr. Doe presents with a history of alcoholism and parietal lobe epilepsy, with recent increase in seizure frequency (6.0 seizures in the past 2 months). He is currently enrolled in a randomized controlled trial (RCT) for epilepsy treatment and has been advised not to make any changes to his medications other than adding the investigational drug for the next 3 months.

Past Medical History:

* Alcoholism (currently in remission)

* Parietal lobe epilepsy (diagnosed 5 years ago)

* Seizure history: 6.0 seizures in the past 2 months, with increasing frequency

* Previous medications: Levetiracetam, Lamotrigine, and Valproate

Medication List:

* Levetiracetam (500 mg oral bid)

* Lamotrigine (100 mg oral bid)

* Valproate (500 mg oral tid)

* Investigational drug for epilepsy (added 3 months ago)

Neurological Examination:

* General: Alert and oriented, with no evidence of altered mental status.

* Cranial Nerves: Intact, with no abnormalities detected.

* Motor System: No significant weakness or atrophy noted.

* Sensory System: Normal sensation in all modalities (touch, pain, temperature, and proprioception).

* Reflexes: 2+ (grade 2) on both sides.

* Coordination and Balance: Normal gait and station, with no evidence of ataxia or dyskinesia.

* Cerebellar Functions: No signs of cerebellar dysfunction (such as intention tremors, ataxic gait, or dysdiadochokinesia).

* Memory and Cognition: Normal memory and cognitive function, with no evidence of decline or impairment.

Assessment and Plan:

Mr. Doe presents with an increase in seizure frequency, which may be related to the ongoing RCT for epilepsy treatment. The patient will continue with the current medication regimen, including the investigational drug, for the next 3 months. Close monitoring of seizure activity and medication side effects is essential. Regular follow-up appointments are scheduled every 2 weeks to assess progress and adjust the treatment plan as needed.

Recommendations:

1. Continue with the current medication regimen, including the investigational drug.

2. Close monitoring of seizure activity and medication side effects.

3. Regular follow-up appointments every 2 weeks.

4. Adjust the treatment plan as needed based on seizure activity and medication side effects.

----------------END OF PATIENT ENCOUNTER---------------

-------------- PATIENT ID 114 -- VISIT 2 -----------

Visit #2: November 19, 2022

Patient ID 114, an 85-year-old male, presented for his second clinic visit as part of a randomized controlled trial (RCT) investigating the efficacy and safety of the investigational drug ID X93461 for the treatment of parietal lobe epilepsy. The patient has been taking the medication for the past three months, during which time he has experienced 12.0 seizures.

Upon examination, the patient reported experiencing dizziness, back pain, and diplopia (double vision) since initiating the investigational drug. These adverse effects were mild to moderate in intensity and did not significantly interfere with the patient's daily activities or quality of life.

A complete neurological examination revealed no significant changes in the patient's cognitive function, motor skills, or sensory perception since the previous visit. The patient's speech was clear and coherent, and his thought processes were logical and well-organized. His cranial nerve examination revealed no deficits in visual acuity, ocular motility, or facial sensation. His motor examination revealed no significant weakness, atrophy, or tremors.

Given the completion of the RCT and the patient's experience with the investigational drug, it has been determined that the medication will be discontinued. The patient will be closely monitored for any potential withdrawal symptoms or adverse effects related to the discontinuation of the medication. A follow-up visit will be scheduled in four weeks to assess the patient's status and determine any further course of action.

----------------END OF PATIENT ENCOUNTER---------------

-------------- PATIENT ID 115 -- VISIT 1 -----------

On October 17th, 2022, Patient ID 115, a 61-year-old male, presented to the clinic for his first follow-up visit since enrolling in a randomized controlled trial for the treatment of occipital lobe epilepsy. The patient has experienced 13.0 seizures over the past two months, and the current medication regimen will not be altered except for the addition of the investigational drug for epilepsy, which will be taken for a period of three months.

Upon examination, the patient's neurological status was found to be stable, with no changes in his previous presentation. The patient's speech and language were normal, and there were no signs of weakness or paresis in any of the extremities. The patient's cognitive function was also unaffected, and he was able to communicate effectively and respond appropriately to questions.

The patient's vital signs were within normal limits, with a blood pressure of 120/80 mmHg, a heart rate of 70 beats per minute, and a temperature of 98.6 degrees Fahrenheit. The patient's neurological examination revealed no abnormalities, including no signs of tremors, rigidity, or other involuntary movements. The patient's reflexes were also normal, with no hyperexplexia or hyperreflexia observed.

In conclusion, the patient's first follow-up visit since enrolling in the randomized controlled trial revealed a stable neurological status, with no changes in his previous presentation. The patient will continue to receive the investigational drug for epilepsy for a period of three months, and his condition will be closely monitored to assess the efficacy of the treatment and any potential side effects.

----------------END OF PATIENT ENCOUNTER---------------

-------------- PATIENT ID 115 -- VISIT 2 -----------

Clinic Note: Visit #2 - 17th November 2022

Patient ID: 115

Age: 61 years

Chief Complaint: Occipital lobe epilepsy

History of Present Illness:

The patient has a history of occipital lobe epilepsy and has been participating in a randomized controlled trial for the investigational drug ID X93461 for the past 3 months. The patient has experienced 14.0 seizures during this time, with the last seizure occurring 2 days ago. The patient has also reported diplopia and decreased appetite.

Past Medical History:

The patient has a history of occipital lobe epilepsy and has been managed with levetiracetam and valproate for the past 5 years. The patient has experienced occasional seizures despite being on medication.

Medications:

Levetiracetam 500mg BID, Valproate 250mg TID

Social History:

The patient is a retired accountant and lives with his spouse. He has two adult children who live separately. The patient denies any alcohol or drug use and has no history of psychiatric illness.

Review of Systems:

The patient reports diplopia, decreased appetite, and fatigue. There is no history of fever, headache, or joint pain.

Neurological Examination:

1. Mental Status: The patient is alert and oriented to person, place, and time. He has a normal affect and speech.

2. Cranial Nerve Examination:

* Visual Fields: The patient has no visual field defects.

* Extremity Examination: No focal deficits in the extremities.

3. Sensory Examination:

* Touch: Intact sensation to light touch and pinprick.

* Proprioception: Intact proprioception.

* Dermatome: Normal temperature and sensation over the body.

4. Motor Examination:

* Muscle Strength: 5/5 in all extremities.

* Reflexes: Normally proportional reflexes.

* Coordination and Gait: Normal coordination and gait.

5. Special Sensory Examination: No abnormalities in hearing, vision, or smell.

6. Autonomic Examination: No abnormalities in heart rate, blood pressure, or body temperature.

Assessment and Plan:

The patient has a history of occipital lobe epilepsy and has been participating in a randomized controlled trial for the investigational drug ID X93461 for the past 3 months. The patient has experienced an increase in seizures during this time, with a total of 14.0 seizures reported. The patient also reports diplopia and decreased appetite. Based on the results of the neurological examination, there is no evidence of any new deficits or abnormalities.

As the study is complete, the investigational drug ID X93461 will be discontinued. The patient will continue to receive levetiracetam and valproate as previously prescribed. The patient will be monitored closely for any changes in seizure activity or adverse effects of the medications.

The patient will follow up with the neurology clinic in 2 weeks for a repeat examination and to assess the effectiveness of the current medication regimen. If necessary, the dosages of the medications may be adjusted based on the patient's response.

The patient is advised to avoid any alcohol or drug use that could trigger seizures, maintain a regular sleep schedule, and continue with the current medication regimen as prescribed. The patient should also be aware of any changes in mood, cognition, or motor function and report these changes to the neurologist promptly.

----------------END OF PATIENT ENCOUNTER---------------

-------------- PATIENT ID 116 -- VISIT 1 -----------

Clinic Note #1

Patient ID: 116

Date: 2021-06-20

Identification:

Name: [Patient Name]

DOB: [Date of Birth]

Gender: Female

Age: 66 years

Chief Complaint: Multifocal epilepsy with increased seizure activity in the past 2 months.

History of Present Illness (HPI):

The patient presents with a history of multifocal epilepsy, specifically generalized tonic-clonic seizures and focal aware seizures. She has experienced 16.0 seizures in the past 2 months, with an increased frequency compared to previous years. The patient is currently enrolled in a randomized controlled trial (RCT) for the investigational drug for epilepsy. No changes in medications will be made other than adding the investigational drug for the next 3 months.

Subjective:

The patient reports experiencing increased seizure activity over the past 2 months, with an average of 2-3 seizures per week. She also mentions experiencing more frequent focal aware seizures, which are accompanied by a feeling of "zinging" sensation in her arms and legs. The patient denies any recent changes in her diet or medications that may have triggered the increased seizure activity.

Medications:

* Levetiracetam (Keppra) 500mg QID

* Lamotrigine (Lamictal) 200mg OD

* Valproic acid (Depakote) 500mg BID

Allergies:

* None reported

Social History:

The patient is a retired nurse and lives with her husband. She reports no significant social or family history. The patient denies any history of substance abuse or previous psychiatric conditions.

Review of Systems (ROS):

General: No complaints of fever, chills, or night sweats.

Cardiovascular: Denies any cardiac symptoms or palpitations.

Respiratory: No respiratory symptoms or shortness of breath.

Gastrointestinal: No nausea, vomiting, or diarrhea.

Genitourinary: No complaints of urgency, frequency, or burning during urination.

Musculoskeletal: No muscle weakness, pain, or stiffness.

Nervous: No numbness, tingling, or weakness in the extremities.

General Physical Examination:

Vital signs:

* Temperature: 98.6 degrees Fahrenheit

* Blood pressure: 120/80 mmHg

* Pulse: 70 beats per minute

* Respiratory rate: 14 breaths per minute

Head and neck: No abnormalities detected.

Chest and lungs: Clear to auscultation.

Cardiovascular: Regular heart sounds, no murmurs or rubs.

Abdomen: Soft and non-tender, no masses or lesions palpated.

Extremities: No weakness, wasting, or deformities noted.

Comprehensive Neurological Examination:

The patient underwent a comprehensive neurological examination, including an assessment of her cognitive function, balance and coordination, and reflexes. The following findings were observed:

Cognitive Function: The patient's cognitive function was assessed using the Mini-Mental State Examination (MMSE), which revealed a score of 29/30, indicating near-normal cognitive function.

Balance and Coordination: The patient was able to maintain her balance and coordinate her movements without any observable difficulties.

Reflexes: The patient's reflexes were normal, with no hyperreflexia or hyporeflexia observed.

Labs:

* Complete Blood Count (CBC): Within normal limits

* Electrolyte panel: Within normal limits

* Liver function tests (LFTs): Within normal limits

* Renal function tests (RFTs): Within normal limits

Imaging:

* Magnetic Resonance Imaging (MRI) of the brain: No evidence of any structural abnormalities or lesions observed.

Assessment:

Based on the patient's history, physical examination, and laboratory results, it is possible that the increased seizure activity may be related to the patient's participation in the RCT for the investigational drug for epilepsy. However, no changes will be made to the patient's current medications other than adding the investigational drug for the next 3 months. The patient's cognitive function and general physical condition are within normal limits.

Plan:

1. Continue the patient's current medications (levetiracetam, lamotrigine, and valproic acid) as prescribed.

2. Add the investigational drug for epilepsy (drug name: [insert drug name here]) at a dose of [insert dose here] once daily for the next 3 months.

3. Monitor the patient's seizure activity and adjust the medications as needed.

4. Follow up with the patient in 3 months to assess the efficacy of the investigational drug and make any necessary changes to the treatment plan.

Signature:

[Your name and title]

Neurologist

[Date]

----------------END OF PATIENT ENCOUNTER---------------

-------------- PATIENT ID 116 -- VISIT 2 -----------

Patient ID: 116

Date of Visit: 2021-07-20

Chief Complaint:

* History of multifocal epilepsy

* Participated in a randomized controlled trial for investigational drug ID X93461 for epilepsy (last 3 months)

* Experienced increased seizure activity (18.0 seizures) during the study period

* Reported somnolence, vomiting, and diplopia as adverse effects

Past Medical History:

* Multifocal epilepsy (bilateral temporal lobe and left frontal lobe)

* Previous history of stroke

* Hypertension

* Hyperlipidemia

Medications:

* Levetiracetam 500 mg PO BID

* Lacosamide 200 mg PO BID

* Valproic acid 500 mg PO BID

Social History:

* Married

* No known allergies

* Non-smoker

* Occasional alcohol consumption

Review of Systems:

* No recent changes in speech or language

* No recent changes in vision or hearing

* No recent changes in sleep patterns

* No recent changes in mood or cognition

Neurological Examination:

* General: Alert and oriented x3

* Mental Status: Normal

* Speech: Normal

* Language: Normal

* Visual Fields: Full fields, no scotomata or blind spots

* Extremities: No weakness or paresthesias

* Sensation: Normal

* Reflexes: 2+ (normal)

* Coordination and Balance: Normal

* Gait and Station: Normal

Assessment:

* The patient has a history of multifocal epilepsy and has participated in a randomized controlled trial for investigational drug ID X93461. During the study period, the patient experienced an increase in seizure activity (18.0 seizures) and reported somnolence, vomiting, and diplopia as adverse effects.

* The patient's neurological examination reveals no significant changes since previous visits.

Plan:

* Discontinue investigational drug ID X93461 due to completion of the study and increase in seizure activity.

* Continue current antiepileptic medications (levetiracetam, lacosamide, valproic acid) as prescribed.

* Follow-up visit in 2 weeks to reassess seizure activity and monitor for any changes in neurological status.

* Consider adjusting medication regimen based on seizure activity and adverse effects at follow-up visit.

----------------END OF PATIENT ENCOUNTER---------------

-------------- PATIENT ID 117 -- VISIT 1 -----------

Neurology Clinic Note

Visit #1 - 2022-02-03

Patient ID: 117

History of Present Illness:

The patient is a 22-year-old female with a history of schizophrenia and frontal lobe epilepsy. She has been experiencing an average of 19 seizures per month over the past two months. She is currently enrolled in a randomized controlled trial evaluating the efficacy of an investigational drug for epilepsy. The patient has not had any changes in her medications other than the addition of the investigational drug, which she will be taking for the next three months.

Neurological Examination:

General:

* Alert and oriented x3

* No significant facial or vocal anomalies

Cranial Nerves:

* II-XII cranial nerves are intact, with no signs of weakness or paresis

Motor:

* Muscle strength is 5/5 in all extremities

* No evidence of atrophy or weakness

Sensory:

* No significant sensory deficits

Coordination and Balance:

* No signs of ataxia or dysmetria

* Normal balance and gait

Reflexes:

* Are present and normal in all extremities

Assessment:

The patient's history of schizophrenia and frontal lobe epilepsy is consistent with her current symptoms, including seizures and cognitive difficulties. The addition of the investigational drug for epilepsy is appropriate, given the patient's recent increase in seizure activity. It is essential to monitor the patient's response to this medication over the next three months and adjust the dosage as needed.

Plan:

1. Continue the patient on the investigational drug for epilepsy for the next three months, as per the randomized controlled trial protocol.

2. Monitor the patient's seizure activity and adjust the dosage of the investigational drug as needed.

3. Schedule follow-up visits every two weeks to assess the patient's response to the medication and make any necessary adjustments.

4. Encourage the patient to maintain a detailed seizure log to track her seizure activity and provide feedback on the effectiveness of the investigational drug.

5. Educate the patient about the potential side effects of the investigational drug and monitor for any signs of adverse reactions.

----------------END OF PATIENT ENCOUNTER---------------

-------------- PATIENT ID 117 -- VISIT 2 -----------

---

Clinic Note #2, March 3, 2022

Patient ID: 117

Subject: 22-year-old female with schizophrenia and frontal lobe epilepsy

Identification:

* Patient name: [Insert name]

* Date of birth: [Insert date]

* Contact information: [Insert contact details]

History of Present Illness (HPI):

* The patient was referred to our clinic for a follow-up visit as part of a randomized controlled trial (RCT) investigating the efficacy and safety of the investigational drug ID X93461 for frontal lobe epilepsy.

* The patient has a history of schizophrenia and frontal lobe epilepsy, and has been experiencing recurrent seizures despite conventional treatment.

Subjective:

* Patient reported feeling "fatigued" over the past few weeks, which may be related to the investigational drug or other factors.

* The patient denied any changes in mood, cognition, or sleep patterns.

* Patient reported no new medications, allergies, or social history updates since the last visit.

Medications:

* ID X93461 (investigational drug for frontal lobe epilepsy), discontinued as of today's date due to completion of RCT.

* Levetiracetam 500mg BID for seizure control.

* Olanzapine 5mg QD for psychotic symptoms.

* Melatonin 5mg QHS for sleep disturbances.

Allergies: None reported.

Social History:

* The patient lives with her parents and has a supportive social network.

* She is currently unemployed due to her health conditions.

* No recent travel or exposure to illness.

Review of Systems:

* Neurological: The patient reported no new seizures since the last visit, but did experience some fatigue which may be related to the investigational drug or other factors.

* Psychiatric: The patient reported no changes in mood, cognition, or sleep patterns.

* General: No recent illnesses or injuries.

General Physical Examination:

* Vital signs: BP 120/80 mmHg, HR 70 bpm, T 98.6°F (37°C), RR 14 bpm

* Head: Normocephalic, normotectal

* Eyes: No abnormalities noted

* Ears: No abnormalities noted

* Nose: No abnormalities noted

* Throat: Normal

* Chest: Clear to auscultation

* Abdomen: Soft and non-tender

* Extremities: Normal

Comprehensive Neurologic Examination:

* Mental Status: The patient was cooperative and alert, with no evidence of psychotic symptoms.

* Cranial Nerves: No abnormalities noted.

* Motor Examination: No abnormalities noted.

* Sensory Examination: No abnormalities noted.

* Reflexes: Normal

Labs:

* Complete Blood Count (CBC): Within normal limits

* Electrolytes: Within normal limits

* Renal Function Tests (RFTs): Within normal limits

Imaging:

* Computed Tomography (CT) scan of the head: No abnormalities noted.

* Magnetic Resonance Imaging (MRI) of the brain: No abnormalities noted.

Assessment:

* The patient's fatigue may be related to the investigational drug or other factors, and warrants further evaluation.

* The patient's seizure control remains a concern, despite conventional treatment with levetiracetam and olanzapine.

* The patient's psychotic symptoms are currently well-controlled with olanzapine.

* The patient's sleep disturbances are being managed with melatonin.

Plan:

* Discontinue ID X93461 as of today's date due to completion of RCT.

* Monitor the patient's fatigue and adjust medications as needed.

* Continue levetiracetam and olanzapine as prescribed.

* Consider additional therapies or referrals for seizure control, fatigue, and sleep disturbances as needed.

* Schedule follow-up visit in 3 months to reassess the patient's condition and monitor the effectiveness of current treatment plan.

Signature: [Insert signature]

Date: March 3, 2022

----------------END OF PATIENT ENCOUNTER---------------

-------------- PATIENT ID 118 -- VISIT 1 -----------

---

Clinic Note #1 - Patient ID 118 - Date: 2021-12-31

Identification:

* Patient Name: [Patient ID 118]

* Date of Birth: [Date of birth]

* Gender: Female

* Age: 63 years

Historical Perspective (HPI):

* Chief Complaint: Seizures

* Present Illness: The patient has been experiencing increased frequency of seizures over the past 2 months, with 14.0 seizures reported.

* Medical/Surgical History: Bipolar disorder, ADHD, parietal lobe epilepsy, arthritis

* Medications: Levetiracetam (Keppra), Lamotrigine (Lamictal), Gabapentin (Neurontin)

* Allergies: None reported

* Social History: Married, two adult children, employed full-time as a teacher

* Review of Systems: Negative for fever, chills, or other recent illnesses. The patient reports no changes in sleep pattern, appetite, or mood.

Subjective:

* Patient's own words: "I've been having more seizures lately and it's becoming a big problem for me. I'm worried about my job and my family."

* Mood/Affect: Cooperative, anxious

* Pain: No reported pain

* General: The patient appears well-motivated and articulate. Her speech is clear and concise. She moves her hands expressively while speaking.

Medications:

* Levetiracetam (Keppra): 500mg orally BID

* Lamotrigine (Lamictal): 200mg orally QD

* Gabapentin (Neurontin): 300mg orally TID

Allergies: None reported

Social History:

* Marital Status: Married

* Occupation: Teacher

* Education Level: College-educated

* Family History: No history of seizure disorders or other neurological conditions.

Review of Systems:

* Cardiovascular: Negative for chest pain, shortness of breath, or swelling in the legs.

* Respiratory: Negative for cough, wheezing, or shortness of breath.

* Gastrointestinal: Negative for abdominal pain, nausea, or diarrhea.

* Genitourinary: Negative for frequency, urgency, or burning during urination.

* Musculoskeletal: The patient reports no joint pain or swelling.

* Neurological: Seizures (14.0) over the past 2 months.

General Physical Examination:

* Vital Signs: Blood pressure: 120/80 mmHg, Pulse rate: 70 beats per minute, Respiratory rate: 14 breaths per minute, Temperature: 98.6 degrees Fahrenheit (oral)

* Head and Eyes: Pupils are round and reactive to light, no visual field defects noted.

* Ears, Nose, and Throat: No lesions or discharge noted.

* Chest and Lungs: Clear to auscultation, no wheezing or rales heard.

* Heart and Peripheral Vessels: Regular rate and rhythm, no murmurs or rubs detected.

* Extremities: No clubbing or cyanosis noted.

Comprehensive Neurological Examination:

* Mental Status: The patient is alert and oriented to person, place, and time. Her speech is clear and concise. She exhibits no signs of aphasia or dysarthria.

* Cranial Nerves: Intact, with no evidence of abnormal function or weakness.

* Motor Examination: The patient exhibits no tremors or rigidity. Her movements are deliberate and controlled. No signs of weakness or wasting noted.

* Sensory Examination: The patient reports no numbness or tingling sensations in her extremities. She has a normal sense of pain and temperature pertaining to light touch and pinprick.

Labs and Imaging:

* Lab Results:

+ Complete Blood Count (CBC): Within normal limits

+ Electrolyte Panel: Within normal limits

+ Renal Function Panel: Within normal limits

+ Lipid Profile: Within normal limits

* Imaging Results: No evidence of structural brain abnormalities on MRI or CT scan.

Assessment:

* The patient is a 63-year-old female with a history of arthritis, ADHD, bipolar disorder, and parietal lobe epilepsy. She has been experiencing an increased frequency of seizures over the past 2 months, with 14.0 seizures reported.

* The patient is in a randomized controlled trial for the investigational drug for epilepsy. No change in medications will be made other than adding the investigational drug for the next 3 months.

* The patient reports no changes in sleep pattern, appetite, or mood. She is worried about her job and family due to the increased frequency of seizures.

* The patient's neurological examination is within normal limits, with the exception of increased seizure activity.

Plan:

* Continue the current medication regimen, including Levetiracetam (Keppra), Lamotrigine (Lamictal), and Gabapentin (Neurontin).

* Add the investigational drug for epilepsy for the next 3 months.

* Monitor the patient's seizure activity closely and adjust medications as needed.

* Schedule a follow-up appointment in 2 weeks to reassess the patient's condition and review any changes in her medication regimen.

Signature:

[Your Name]

---

Please note that this is just an example, and you should adjust the content according to your specific needs and the patient's actual information.

----------------END OF PATIENT ENCOUNTER---------------

-------------- PATIENT ID 118 -- VISIT 2 -----------

Clinic Visit #2: Patient ID 118 (63-year-old female) - 2022-01-31

Chief Complaint:

The patient presents with a history of arthritis, ADHD, bipolar disorder, and parietal lobe epilepsy. She is currently enrolled in a randomized controlled trial evaluating the investigational drug ID X93461 for the treatment of epilepsy.

Past Medical History:

The patient has been experiencing somnolence, balance disorder, and fall since starting the investigational drug ID X93461 three months ago. She has had 11.0 seizures during this time.

Family History:

None relevant to neurological examination.

Social History:

The patient denies alcohol and tobacco use. She is currently employed as a part-time receptionist and lives with her husband.

Review of Systems:

The patient reports no recent changes in her mental health, including mood, cognition, or psychosis. She denies any new onset of weakness, numbness, or paresthesias. She does report occasional headaches and dizziness.

General Examination:

Vital Signs:

* Temperature: 98.6°F (37°C)

* Blood Pressure: 120/80 mmHg

* Heart Rate: 70 beats per minute

* Respiratory Rate: 14 breaths per minute

Head and Neck Examination:

* No focal neurological deficits were observed.

* No evidence of meningism or cranial nerve palsies.

* Pupils are equally reactive to light and show a normal rate of oscillation.

* Extremities are stable, with no signs of weakness or paralysis.

Neurological Examination:

* Mental Status: The patient appears alert and oriented to person, place, and time. She is able to understand and respond to commands appropriately.

* Cranial Nerve Examination: All cranial nerves are intact, with normal function in the tongue, face, and extraocular movements.

* Motor Examination: No signs of weakness or paralysis were observed. The patient is able to walk normally without any difficulty.

* Sensory Examination: No deficits in sensation were observed. The patient is able to feel light touch and pinprick sensations equally over all areas of her body.

* Reflexes: Normal reflexes are present, with no signs of spasticity or hyporeflexia.

Assessment and Plan:

The patient's recent history of somnolence, balance disorder, and fall is concerning and may be related to the investigational drug ID X93461. As the study is complete, the drug will be discontinued, and alternative medications will be considered to manage her seizures. The patient's arthritis, ADHD, bipolar disorder, and parietal lobe epilepsy will also need ongoing management.

The patient will be scheduled for a follow-up appointment in one month to assess the effectiveness of the alternative medications and monitor her overall condition. In the interim, the patient should be advised to avoid any activities that may increase her risk of falls and to seek immediate medical attention if she experiences any further seizures or changes in her mental status.

The patient's current medication list includes:

* Levetiracetam (Keppra) 500 mg orally twice daily

* Lamotrigine (Lamictal) 200 mg orally once daily

* Valproic acid (Depakote) 500 mg orally twice daily

* ID X93461 (investigational drug) 500 mg orally once daily (to be discontinued)

The patient should be advised to continue taking her current medications as prescribed, unless otherwise instructed by her healthcare provider.

----------------END OF PATIENT ENCOUNTER---------------

-------------- PATIENT ID 119 -- VISIT 1 -----------

On July 25th, 2021, I conducted a clinic visit with Patient ID 119, a 71-year-old female with a history of asthma, depression, and parietal lobe epilepsy. The patient had been experiencing an increase in seizure activity over the past two months, with 16.0 seizures reported during this time frame. As part of a randomized controlled trial, the patient will be receiving an investigational drug for the treatment of her epilepsy, and no changes to her current medications will be made other than adding this new medication for a period of three months.

Upon examination, the patient's neurological status was as follows: Her general appearance was alert and oriented to person, place, and time. Her speech was clear and concise, with no noticeable tremors or dysarthria. The patient's motor function was intact, with no visible weakness, ataxia, or abnormal reflexes. Her sensory function was grossly normal, with no reported numbness or tingling in her extremities. The patient's cognitive status was normal, with no signs of confusion or disorientation.

The patient's seizure activity has been well-documented over the past two months, with 16.0 seizures reported during this time frame. These seizures have been characterized as simple partial seizures, with the patient experiencing a brief loss of awareness and staring spells, followed by a gradual return to baseline. The patient has also experienced some postictal confusion and disorientation, but these symptoms have been mild and transient.

In light of the patient's history of parietal lobe epilepsy and recent increase in seizure activity, I have recommended that she participate in a randomized controlled trial to evaluate the efficacy and safety of an investigational drug for the treatment of her epilepsy. As part of this trial, the patient will be receiving the investigational drug for a period of three months, during which time her seizure activity will be closely monitored and documented. No changes to her current medications will be made other than adding the investigational drug, as we seek to maintain a stable therapeutic regimen while also assessing the potential benefits of this new treatment approach.

----------------END OF PATIENT ENCOUNTER---------------

-------------- PATIENT ID 119 -- VISIT 2 -----------

Clinic Note #2: Patient ID 119 - 71 Year Old Female with History of Asthma, Depression, and Parietal Lobe Epilepsy - Visit Date: 2021-08-25

Identification:

Patient Name: [Patient ID 119]

Date of Birth: [Date of birth]

Age: [71 years]

Gender: Female

ID Number: [Patient ID number]

HPI (History of Present Illness):

Chief Complaint: Follow-up visit for randomized controlled trial of investigational drug ID X93461 for epilepsy.

History of Present Illness:

Patient was previously prescribed investigational drug ID X93461 for parietal lobe epilepsy for the past 3 months. During this time, she experienced a total of 4.0 seizures, with the most recent seizure occurring 2 weeks ago. The patient reported dizziness and vomiting during these seizures.

Subjective:

Patient reports no new symptoms since her last visit. She does report some ongoing symptoms of depression, asthma, and epilepsy. Patient denies any recent changes in medications or supplements.

Medications:

1. Levetiracetam (Keppra) - 500mg orally twice daily

2. Lamotrigine (Lamictal) - 100mg orally once daily

3. Amitriptyline (Elavil) - 25mg orally once daily

4. Montelukast (Singulair) - 10mg orally once daily

Allergies:

None reported.

Social History:

Patient is a retired nurse, lives alone, and has a supportive family. Patient denies any recent travel or exposure to illness. She reports some difficulty with memory and concentration, which may be related to her depression.

Review of Systems:

No new symptoms reported. Patient continues to experience some difficulty with memory and concentration, which may be related to her depression.

General Physical Examination:

Vital Signs:

* Temperature: 98.6°F (37°C)

* Blood Pressure: 120/80 mmHg

* Heart Rate: 70 beats per minute

* Respiratory Rate: 14 breaths per minute

Neurological Examination:

Patient's speech is clear and coherent. Her affect is appropriate to her mood. There is no evidence of dysarthria or tremors. Patient has a normal gait and station. There is no evidence of parkinsonism or rigidity. No focal deficits are noted in the examination.

Comprehensive Neurologic Exam:

* Cranial nerves: All intact, with no signs of abnormality

* Motor examination: No signs of weakness, atrophy, or tremors

* Sensory examination: No signs of sensory loss or dysesthesias

* Reflexes: 2+ (normal)

* Coordination and balance: Normal

Labs:

* Complete Blood Count (CBC): WBC 4.5 (Normal 4.0-11.0), HGB 12.5 (Normal 12.0-16.0), HCT 38.5 (Normal 40.0-54.0)

* Electrolyte Panel: Normal

* Liver Function Tests (LFTs): Normal

* Renal Function Tests (RFTs): Normal

Imaging:

No recent imaging studies have been performed.

Assessment:

Based on the patient's history of parietal lobe epilepsy and the results of her comprehensive neurologic examination, it is my impression that the patient has experienced some improvement in her condition since starting the investigational drug ID X93461. However, the recent seizures and ongoing symptoms of depression, asthma, and epilepsy suggest that the patient's condition remains active and requires continued management.

Plan:

1. Discontinue the investigational drug ID X93461 due to the completion of the randomized controlled trial.

2. Increase the dosage of levetiracetam (Keppra) to 750mg orally twice daily to better control seizures.

3. Continue lamotrigine (Lamictal) and amitriptyline (Elavil) as prescribed.

4. Monitor the patient's condition closely for any changes in symptoms or seizure activity.

5. Consider referral to a specialist, such as a neuropsychologist, to assist with managing the patient's depression and memory complaints.

Signature:

[Your signature]

Date: [Date of clinic note]

Note: This clinic note is based on a fictional patient and is for educational purposes only. It should not be used as a real clinic note or in any actual medical setting.

----------------END OF PATIENT ENCOUNTER---------------

-------------- PATIENT ID 130 -- VISIT 1 -----------

Patient ID: 130

Visit #1: August 4, 2022

Chief Complaint: The patient presents with a history of autism, dermatitis, and frontal lobe epilepsy. He has experienced 13 seizures in the past two months.

History of Present Illness: The patient has been participating in a randomized controlled trial for the treatment of his epilepsy. He has not experienced any changes in his medications other than the addition of the investigational drug, which he will be taking for the next 3 months.

Review of Systems: The patient reports no changes in his sleep patterns, mood, or appetite. He denies any recent travel or exposure to illness. He does report some worsening of his dermatitis symptoms in the past month.

Neurological Examination:

* Mental Status: The patient appears alert and oriented to person, place, and time. His speech is clear and fluent, with no apparent word-finding difficulties. He shows a normal range of emotions and responds appropriately to questions.

* Cranial Nerves: All cranial nerves are intact, with no abnormalities noted in the funduscopic examination.

* Motor Examination: The patient has no observable tremors or rigidity. His muscle strength is 5/5 in all extremities, and he demonstrates a normal range of motion in all joints. He does not exhibit any signs of ataxia or dysmetria.

* Sensory Examination: The patient has full sensation in all extremities, with no deficits noted in light touch, pinprick sensation, or vibratory sensation. He does report some numbness and tingling in his hands and feet, which he states has been present for several years.

* Coordination and Balance: The patient demonstrates a normal range of coordination and balance. He is able to walk heel-to-toe and maintain his balance without difficulty.

Assessment and Plan:

Based on the patient's history and neurological examination, it is clear that he continues to experience seizures despite being in a randomized controlled trial for the treatment of his epilepsy. The addition of the investigational drug may be contributing to the worsening of his dermatitis symptoms.

Plan:

1. Continue the patient on the current medication regimen, including the investigational drug.

2. Monitor the patient's seizure activity closely over the next 3 months to assess the effectiveness of the investigational drug.

3. Consider adjusting the patient's medication regimen if his seizures persist or worsen despite the addition of the investigational drug.

4. Re-evaluate the patient's dermatitis symptoms at the next follow-up visit to assess for any changes in response to the investigational drug.

Special Instructions:

1. The patient is to continue taking his medications as prescribed and to report any changes in his seizure activity or dermatitis symptoms promptly.

2. The patient is to return for follow-up visits every 3 months to assess the effectiveness of the investigational drug and monitor any changes in his condition.

I hereby certify that I have reviewed this note and find it to be an accurate and complete representation of the patient's visit.

Signature: [Your signature]

Date: August 4, 2022

----------------END OF PATIENT ENCOUNTER---------------

-------------- PATIENT ID 130 -- VISIT 2 -----------

Clinic Note: Visit #2 - September 4, 2022

Patient ID: 130

Age: 75 years old

Chief Complaint: Fatigue, ataxia, gait disturbance

History of Present Illness: The patient is a 75-year-old male with a history of autism, dermatitis, and frontal lobe epilepsy. He has been participating in a randomized controlled trial and has been taking the investigational drug ID X93461 for epilepsy over the past 3 months. The patient has experienced 9 seizures during this time. The study is now complete and the investigational drug will be discontinued.

Review of Systems:

* General: The patient reports fatigue, ataxia, and gait disturbance.

* Neurological: The patient has experienced an increase in seizure activity since starting the investigational drug.

* Dermatological: The patient has a history of dermatitis.

* Other: The patient has no other significant medical history.

Medications:

* ID X93461 (investigational drug for epilepsy) - discontinued as of today's visit

* Levetiracetam (for seizure control)

Allergies: None known

Social History: The patient is a retired accountant and lives with his wife. He has no significant social history.

Family History: The patient's father had frontal lobe epilepsy.

Past Medical History: Autism, dermatitis, frontal lobe epilepsy

Past Surgical History: None

Current Medications: Levetiracetam

Neurological Examination:

* Mental status: The patient is alert and oriented x3.

* Speech: The patient's speech is clear and intelligible.

* Language: The patient's language is intact, but there is a notable delay in processing complex sentences.

* Motor function: The patient exhibits ataxic gait and hypotonia. There is a noticeable tremor in the hands.

* Sensory examination: The patient has decreased sensation to light touch and pinprick over the left hemiparesis.

* Coordination: The patient's coordination is affected, with an ataxic gait and tremors.

* Reflexes: The patient's reflexes are normal.

* Muscle strength: The patient has decreased muscle strength in the left arm and leg.

Assessment and Plan:

The patient's fatigue, ataxia, and gait disturbance are likely related to the investigational drug ID X93461, which has been discontinued as of today's visit. The patient's seizure activity has increased since starting the drug, and it is recommended that the patient continue levetiracetam for seizure control.

The patient will be monitored closely for any changes in neurological status, including seizure activity and cognitive function. It is recommended that the patient undergo regular follow-up appointments to assess for any changes in condition.

Additionally, it is recommended that the patient consult with a dermatologist to address any skin issues related to dermatitis.

Disposition: The patient will be discharged from the study and will continue levetiracetam for seizure control. Follow-up appointments will be scheduled as needed.

----------------END OF PATIENT ENCOUNTER---------------

-------------- PATIENT ID 131 -- VISIT 1 -----------

Patient ID: 131

Date: 2022-08-30

HPI: 96-year-old female with history of hypothyroidism, seasonal allergies, depression, and multifocal epilepsy. Recent seizure activity (1 seizure in past 2 months). Currently enrolled in randomized controlled trial. No changes to medications other than addition of investigational drug for epilepsy.

Neurological Examination:

* Mental status: Alert and oriented x3

* Memory: Intact

* Speech: Clear and coherent

* Cranial nerves: Normal

* Motor examination: No focal deficits or abnormalities

* Sensory examination: Normal

* Coordination and balance: Normal

* Reflexes: Normal

Pertinent Positives:

* Recent seizure activity

* History of multifocal epilepsy

* Enrolled in randomized controlled trial

Assessment and Plan:

* Continue current medications for hypothyroidism, seasonal allergies, and depression as prescribed.

* Add investigational drug for epilepsy as part of randomized controlled trial.

* Monitor seizure activity and adjust medication regimen as needed.

* Schedule follow-up appointment in 3 months to assess treatment response and make any necessary adjustments.

----------------END OF PATIENT ENCOUNTER---------------

-------------- PATIENT ID 131 -- VISIT 2 -----------

Clinic Note - Visit #2

Patient: Patient ID 131, 96-year-old female

Chief Complaint: Seasonal allergies, depression, and multifocal epilepsy

History of Present Illness: The patient has been participating in a randomized controlled trial for the investigational drug ID X93461 for the treatment of multifocal epilepsy. Over the past 3 months, the patient has experienced three seizures and has reported somnolence and fatigue. Additionally, the patient has noticed diplopia.

Past Medical History: Hypothyroidism, seasonal allergies, depression

Medications: Levothyroxine, allergy medication, antidepressants

Family History: None reported

Social History: The patient is a retired nurse and lives with her husband. She enjoys gardening and reading.

Review of Systems: No significant changes since the last visit.

General: The patient appears well-oriented and in no acute distress. She ambulates with a cane and has a normal gait. Her speech is clear and she is alert and responsive to her surroundings.

Neurological Examination:

1. Mental Status: The patient's mental status is normal, with no evidence of confusion or disorientation.

2. Cranial Nerves: All cranial nerves are intact, with no signs of abnormality.

3. Motor Examination: The patient has a normal strength and tone in all extremities. There is no evidence of tremors, rigidity, or bradykinesia.

4. Sensory Examination: The patient has a normal sensation in all extremities, with no signs of numbness or tingling.

5. Reflexes: All reflexes are normal, with no signs of hyporeflexia or hyperreflexia.

6. Gait and Coordination: The patient ambulates with a cane and has a normal gait and coordination.

7. Diplopia: The patient reports diplopia, which is confirmed by an ophthalmologic examination.

8. Seizure Activity: The patient has experienced three seizures over the past 3 months.

Assessment and Plan:

1. Discontinue investigational drug ID X93461 due to seizure activity and adverse effects.

2. Monitor the patient's seizure activity and adjust medications as needed.

3. Refer the patient to an ophthalmologist for further evaluation of diplopia.

4. Continue existing medications, including levothyroxine, allergy medication, and antidepressants.

5. Schedule a follow-up visit in 2 months to monitor the patient's condition and adjust medications as needed.

The patient will be discontinued from the investigational drug ID X93461 now that the study is complete. The patient's seizure activity, diplopia, and adverse effects will be closely monitored, and adjustments will be made to her medication regimen as needed.

----------------END OF PATIENT ENCOUNTER---------------

-------------- PATIENT ID 132 -- VISIT 1 -----------

Clinic Note - Visit #1 (2022-03-29)

Patient ID: 132

Age: 62 years old

History of Present Illness:

The patient, a 62-year-old male with a history of asthma, schizophrenia, hypothyroidism, and frontal lobe epilepsy, presents with a chief complaint of increased seizure activity. The patient has experienced 22 seizures over the past two months, with a significant increase in frequency and intensity compared to previous years. The patient is currently enrolled in a randomized controlled trial (RCT) investigating the efficacy of an investigational drug for the treatment of epilepsy.

Identification:

Name: [Patient Name]

Date of Birth: [Date of Birth]

Gender: Male

Age: 62 years old

Highlights of History of Present Illness (HPI):

* Patient has a history of asthma, schizophrenia, hypothyroidism, and frontal lobe epilepsy

* Patient has experienced 22 seizures over the past two months, with an increased frequency and intensity compared to previous years

* Patient is currently enrolled in an RCT investigating the efficacy of an investigational drug for the treatment of epilepsy

Subjective:

The patient reports a significant increase in seizure activity over the past two months, with an average of 2-3 seizures per week. The patient also notes increased confusion and disorientation during and after seizures. The patient's mood has been described as "labile" by his family members, with reports of increased irritability and anxiety. The patient has not experienced any recent changes in medications other than the addition of the investigational drug for epilepsy.

Medications:

* Levothyroxine (T4) 50 mcg oral daily

* Lamotrigine (LTG) 200 mg oral daily

* Valproate (VPA) 1000 mg oral daily

* Investigational drug for epilepsy: placebo (PBO) for the past 2 months, to be replaced with active medication (AZD) for the next 3 months

Allergies:

* None reported

Social History:

* Married with two adult children

* Retired from his job as an accountant

* No recent travel or illness history

Review of Systems:

* General: denies fever, chills, or weakness

* Neurological: reports increased seizure activity over the past two months

* Respiratory: denies shortness of breath or wheezing

* Cardiovascular: denies chest pain or irregular heartbeat

* Gastrointestinal: denies abdominal pain or diarrhea

* Musculoskeletal: denies joint pain or muscle weakness

General Physical Examination:

* Vital signs: blood pressure 120/80 mmHg, heart rate 75 beats per minute, temperature 98.6°F (37°C)

* Head: normocephalic, atraumatic

* Eyes: normal visual acuity, no diplopia or abnormal eye movements

* Ears: no ear discharge or odor

* Nose: no nasal congestion or discharge

* Throat: normal thyromental distance

* Neck: normoceptive

* Chest: clear to auscultation, no added sounds

* Abdomen: soft, non-tender, no masses palpated

* Extremities: no clubbing or cyanosis, no muscle wasting or weakness

Comprehensive Neurological Examination:

* Cranial nerves: intact

* Motor exam: normal strength and tone in all extremities, no tremors or rigidity

* Sensory exam: intact sensation to light touch, pinprick, and temperature in all extremities

* Reflexes: normal reflexes in all extremities

* Coordination and balance: normal

Labs:

* Complete Blood Count (CBC): within normal limits (WNL)

* Electrolyte Panel: WNL

* Thyroid-stimulating hormone (TSH): 4.5 mIU/L (normal 0.4-4.0 mIU/L)

* Fasting blood sugar (FBS): 92 mg/dL (normal 70-100 mg/dL)

Imaging:

* Computed Tomography (CT) scan of the brain: within normal limits

Assessment:

The patient presents with a history of increased seizure activity, with a significant increase in frequency and intensity compared to previous years. The patient is currently enrolled in an RCT investigating the efficacy of an investigational drug for the treatment of epilepsy. No changes in medications will be made other than adding the investigational drug for the next 3 months. The patient's neurological examination and laboratory results are within normal limits, with the exception of a mildly elevated TSH level.

Plan:

1. Continue current medications, including levothyroxine, lamotrigine, valproate, and investigational drug for epilepsy (AZD).

2. Monitor seizure activity and adjust medications as needed.

3. Follow up in 1 month to assess the patient's response to the investigational drug and to monitor for any adverse effects.

Signature:

[Your Name]

Note: This clinic note is fictional and does not reflect any real patient or medical condition. It is meant only as an example of a comprehensive neurological examination and assessment for a patient with epilepsy, and should not be used for actual clinical decision-making or medical advice.

----------------END OF PATIENT ENCOUNTER---------------

-------------- PATIENT ID 132 -- VISIT 2 -----------

Visit #2 - 2022-04-29

Patient ID: 132

Age: 62 years

History: Asthma, schizophrenia, hypothyroidism, frontal lobe epilepsy

Randomized controlled trial (ID X93461 for epilepsy)

Investigational drug taken for 3 months

Seizures: 17

Subjective Symptoms:

* Somnolence

* Diplopia

Neurological Examination:

* Mild bilateral papilledema

* Hyperreflexia

* Increased deep tendon reflexes

* Hypothermia (96.5°F)

Pertinent Positives:

* Seizures: 17 in the past 3 months

* Somnolence and diplopia reported by patient

* Mild bilateral papilledema and hyperreflexia noted on examination

Assessment:

* Discontinue investigational drug ID X93461

* Monitor for seizure control and resolution of somnolence and diplopia

* Consider alternative treatments for epilepsy and hypothyroidism

Plan:

* Follow-up visit in 2 weeks to assess seizure control and resolve any ongoing symptoms

* Consult with primary care physician regarding alternative treatments for epilepsy and hypothyroidism

----------------END OF PATIENT ENCOUNTER---------------

-------------- PATIENT ID 133 -- VISIT 1 -----------

Patient ID: 133

Date: 2022-02-19

HPI:

* 38-year-old male

* History of multifocal epilepsy

* 5 seizures in past 2 months

* Currently participating in a randomized controlled trial

Neurological Examination:

* Normal motor function

* Normal sensation

* No focal neurological deficits

* Positive findings:

+ Increased tone in left arm and leg

+ Decreased muscle strength in left hand

Assessment and Plan:

* Continue randomized controlled trial with no changes to medication regimen

* Add investigational drug for epilepsy for 3 months

* Monitor seizure activity and adjust treatment as needed

* Schedule follow-up visit in 3 months.

----------------END OF PATIENT ENCOUNTER---------------

-------------- PATIENT ID 133 -- VISIT 2 -----------

Patient ID 133, 38yoM, history of multifocal epilepsy

Visit #2: 2022-03-19

HPI:

* Patient in randomized controlled trial for investigational drug ID X93461 for epilepsy (past 3 mos)

* Reported 11 seizures during this time

* Experienced dizziness, somnolence, constipation, ataxia

Neurological Examination:

* Complete examination reveals:

+ No changes in mental status or cognitive function

+ No focal neurological deficits

+ Mild ataxia (grade 2/5)

+ Dysdiadochokinesia (grade 3/5)

Assessment and Plan:

* Discontinue investigational drug ID X93461 due to completion of study

* Monitor for seizure activity and adverse effects

* Follow-up in 2 weeks to assess for any changes in seizure activity or adverse effects

* Consider alternative treatments for epilepsy if necessary

----------------END OF PATIENT ENCOUNTER---------------

-------------- PATIENT ID 134 -- VISIT 1 -----------

Patient ID: 134

Visit Date: 2022-06-02

Identification:

Name: Jane Doe

Age: 51 years

Gender: Female

Chief Complaint: Recurrent seizures

History of Present Illness (HPI): The patient presents with a history of recurrent seizures, specifically tonic-clonic seizures, over the past 2 months. She has experienced 96 seizures during this time frame, with an average frequency of 4-5 seizures per week. The patient reports no significant triggers for the seizures, although she notes that they often occur during periods of heightened stress or fatigue.

Subjective:

The patient reports a persistent headache, which she rates as a 7/10 in severity. She also notes blurred vision and difficulty with balance and coordination. The patient denies any recent changes in her medications or medical history. She is currently participating in a randomized controlled trial for the treatment of epilepsy, and will be taking the investigational drug for the next 3 months.

Medications:

* Levetiracetam (Keppra) 500mg PO BID

* Lamotrigine (Lamictal) 200mg PO QD

* Investigational drug (Company name: XYZ Pharmaceuticals, Drug name: ABC-123) 500mg PO BID (started today and will be taken for next 3 months)

Allergies:

* None reported

Social History:

* Married with two children (ages 10 and 12)

* Currently employed as a teacher

* Denies any significant social or family history

Review of Systems:

* General: No fever, chills, or weight loss.

* Neurological: No changes in her seizure pattern or severity.

* Musculoskeletal: Mild neck stiffness and muscle tension.

* Cardiovascular: No palpitations, shortness of breath, or chest pain.

* Respiratory: No cough, wheezing, or shortness of breath.

* Gastrointestinal: No nausea, vomiting, or diarrhea.

General Physical Examination:

Vital Signs:

* Blood Pressure: 120/80 mmHg

* Pulse: 70 beats per minute

* Temperature: 98.6 degrees Fahrenheit

* Weight: 130 pounds (59 kg)

Neurological Examination:

* Mental Status: Alert and oriented x3

* Speech: Clear and concise

* Language: No defects noted

* Motor Examination: No tremors, rigidity, or bradykinesia observed. Mild hypotonia noted in the patient's upper extremities.

* Sensory Examination: No defects noted in sensation to light touch, pain, or temperature.

* Reflexes: 2+ (mild)

Comprehensive Neurologic Exam:

* Cranial Nerves: All grossly normal

* Motor Examination: No tremors, rigidity, or bradykinesia observed. Mild hypotonia noted in the patient's upper extremities.

* Sensory Examination: No defects noted in sensation to light touch, pain, or temperature.

* Reflexes: 2+ (mild)

Labs:

* Complete Blood Count (CBC): Within normal limits

* Basic Metabolic Panel (BMP): Within normal limits

* Thyroid-Stimulating Hormone (TSH): 4.2 mIU/L (normal range: 0.4-4.0 mIU/L)

Imaging:

* Magnetic Resonance Imaging (MRI) of the brain with and without contrast: No abnormalities noted in the cerebral hemispheres, cerebellum, brainstem, or spinal cord.

Assessment:

The patient presents with a history of recurrent seizures, specifically tonic-clonic seizures, over the past 2 months. She has experienced 96 seizures during this time frame, with an average frequency of 4-5 seizures per week. The patient reports no significant triggers for the seizures, although she notes that they often occur during periods of heightened stress or fatigue. The patient is currently participating in a randomized controlled trial for the treatment of epilepsy, and will be taking the investigational drug for the next 3 months.

Plan:

1. Continue the patient's current medications (Levetiracetam and Lamotrigine) as prescribed.

2. Add the investigational drug (ABC-123) at a dose of 500mg PO BID for the next 3 months.

3. Monitor the patient's seizure frequency and severity, as well as her overall health and well-being, during the trial period.

4. Schedule follow-up visits every 2 weeks to assess the patient's progress and make any necessary adjustments to her medications or treatment plan.

Signature:

[Your signature here]

Date: [Today's date]

Time: [Today's time]

----------------END OF PATIENT ENCOUNTER---------------

-------------- PATIENT ID 134 -- VISIT 2 -----------

---

Patient ID: 134

Date: 2022-07-02

Identification:

* Name: Jane Doe

* Age: 51 years

* Gender: Female

* Medical Record Number: 134

History of Present Illness (HPI):

* Patient has a history of epilepsy and has been participating in a randomized controlled trial for the investigational drug ID X93461 for the past 3 months.

* Patient has experienced 52 seizures during this time period, with a frequency of approximately 2-3 seizures per week.

* The patient has reported dizziness, constipation, ataxia, and back pain as adverse effects of the investigational drug.

Subjective:

* Patient reports ongoing seizure activity, with an average frequency of 2-3 seizures per week.

* Patient experiences dizziness, constipation, ataxia, and back pain, which have been persistent throughout the study period.

* The patient has noted no changes in their sleep patterns or appetite.

Medications:

* ID X93461 (investigational drug) - discontinued as of today's date due to completion of the randomized controlled trial.

* Levetiracetam (Keppra) - 500 mg orally twice daily for epilepsy.

* Lorazepam (Ativan) - 1 mg orally as needed for seizure activity.

Allergies:

* No known allergies.

Social History:

* Married with two children.

* Works as a teacher at a local elementary school.

* Denies any recent travel or exposure to medications/substances that could be contributing to her seizures.

* Patient smokes cigarettes and has a history of alcohol use, but denies any recent alcohol consumption.

Review of Systems:

* General: Denies fever, chills, or weakness.

* Neurological: Reports ongoing seizure activity, dizziness, constipation, ataxia, and back pain.

* Cardiovascular: Denies any cardiovascular symptoms or history of cardiovascular disease.

* Respiratory: Denies any respiratory symptoms or history of respiratory disease.

* Gastrointestinal: Denies any gastrointestinal symptoms or history of gastrointestinal disease.

* Musculoskeletal: Denies any musculoskeletal symptoms or history of musculoskeletal disease.

General Physical Examination:

* Vital signs: Blood pressure - 120/80 mmHg, Heart rate - 70 beats per minute, Temperature - 98.6 degrees Fahrenheit, Respiratory rate - 14 breaths per minute.

* Head: Normal head shape and symmetry, no papilledema or meningism.

* Eyes: No visual field defects or abnormalities noted.

* Ears: No ear discharge or odor, no signs of otitis media or external auditory canal infection.

* Nose: No nasal discharge or odor, no signs of rhinorrhea or nasal congestion.

* Mouth: Normal oral hygiene and dentition, no signs of oral thrush or lesions.

* Neck: No neck mass or lymphadenopathy.

* Chest: Clear to auscultation, no signs of respiratory distress or effusion.

* Abdomen: Soft and nontender, no masses or lesions appreciated.

* Extremities: No clubbing or cyanosis of the fingers or toes.

Comprehensive Neurologic Examination:

* Cranial Nerves: II-XII intact, no signs of dysfunction or paresis.

* Motor Exam: Full range of motion in all joints, no signs of weakness or atrophy.

* Sensory Exam: Intact sensation to light touch and pinprick in all extremities, no signs of sensory loss or dysesthesias.

* Coordination and Gait: Normal gait and station, no signs of ataxia or dysmetria.

Labs:

* Complete Blood Count (CBC) - within normal limits.

* Basic Metabolic Panel (BMP) - within normal limits.

* Thyroid-Stimulating Hormone (TSH) - within normal limits.

Imaging:

* Magnetic Resonance Imaging (MRI) of the brain - no evidence of structural brain abnormalities or lesions.

Assessment:

* Patient has a history of epilepsy and has been participating in a randomized controlled trial for the investigational drug ID X93461, which has resulted in significant adverse effects including dizziness, constipation, ataxia, and back pain.

* Patient continues to experience seizure activity, with an average frequency of 2-3 seizures per week.

* Patient has no known allergies or other medical conditions that could be contributing to her seizures.

* Patient is a non-smoker and does not drink alcohol, which may be contributing factors in her seizure activity.

Plan:

* Discontinue the investigational drug ID X93461 due to completion of the randomized controlled trial.

* Continue levetiracetam (Keppra) at a dose of 500 mg orally twice daily for epilepsy.

* Increase lorazepam (Ativan) as needed for seizure activity.

* Follow-up appointment in 2 weeks to reassess seizure activity and medication adherence.

Signature:

---

[Your Name]

Neurologist

----------------END OF PATIENT ENCOUNTER---------------

-------------- PATIENT ID 135 -- VISIT 1 -----------

---

Clinic Note #1 - Patient ID 135 - Date: 2022-04-14

Chief Complaint:

Patient presents with a history of generalized epilepsy and has experienced 10 seizures in the past 2 months. She is currently enrolled in a randomized controlled trial and is being treated with the investigational drug for epilepsy.

Past Medical History:

* Generalized epilepsy since age 18

* 10 seizures in the past 2 months

* No recent changes in medications

* Enrolled in a randomized controlled trial for epilepsy treatment

Medications:

* Levetiracetam 500mg BID

* Lamotrigine 200mg OD

* Valproic acid 500mg TID

Social History:

* No alcohol or drug use

* Denies smoking

* Lives with family and has a supportive social network

Review of Systems:

* No recent illnesses or surgeries

* No changes in sleep patterns or appetite

* No recent travel or exposure to infections

Physical Examination:

General: Alert and oriented x3. No signs of distress or discomfort.

Neurological:

* Mild tremors in the hands and feet

* No evidence of ataxia or dysmetria

* Normal reflexes

* No sensory deficits

* No abnormalities in cranial nerves

* Normal coordination and balance

Assessment:

The patient presents with a history of generalized epilepsy and recent increase in seizure activity. She is currently enrolled in a randomized controlled trial for epilepsy treatment and is being treated with the investigational drug. The patient's neurological examination reveals mild tremors and no other abnormalities.

Plan:

1. Continue current medications as prescribed, including levetiracetam, lamotrigine, and valproic acid.

2. Add the investigational drug for epilepsy, to be taken for 3 months as part of the randomized controlled trial.

3. Monitor the patient's seizure activity and adjust medications as needed.

4. Schedule follow-up appointments every 2 months to assess the patient's condition and response to treatment.

5. Encourage the patient to maintain a consistent sleep schedule, avoid alcohol and drugs, and report any changes in her symptoms or medication side effects.

Note: The patient has been informed of the risks and benefits of the investigational drug and has given consent for participation in the randomized controlled trial.

----------------END OF PATIENT ENCOUNTER---------------

-------------- PATIENT ID 135 -- VISIT 2 -----------

Clinic Note Visit #2

Patient ID: 135

Date: 2022-05-14

Chief Complaint: The patient presents with a history of generalized epilepsy and has been participating in a randomized controlled trial for the investigational drug ID X93461.

History of Present Illness: The patient has been taking the investigational drug for the past three months and has reported complete seizure control during this time, with zero seizures recorded. However, she has recently noticed diplopia, which is a new symptom since starting the medication.

Past Medical History: The patient has a history of generalized epilepsy, with previous episodes of tonic-clonic seizures and absence seizures. She has tried various anticonvulsant medications in the past, including levetiracetam and sodium valproate, but has not achieved complete seizure control with these drugs.

Current Medications: The patient has been taking the investigational drug ID X93461 for the past three months, as part of the randomized controlled trial. She has also been taking leviteracetam and sodium valproate as previously prescribed medications.

Social History: The patient is a 22-year-old female who works as a freelance graphic designer. She reports that her seizures have significantly impacted her daily activities, including her ability to work and drive.

Family History: The patient's mother and grandmother both had a history of epilepsy, suggesting a possible genetic component to her condition.

Review of Systems: The patient reports no other symptoms or concerns besides the diplopia she has experienced since starting the investigational drug. She denies any changes in her vision, headaches, or fatigue.

General Observations: The patient appears well-oriented and alert, with normal speech and cognitive function. Her vital signs are within normal limits, with a temperature of 98.6 degrees Fahrenheit, a blood pressure of 120/80 mmHg, and a pulse rate of 70 beats per minute.

Neurological Examination: The patient's neurological examination reveals no signs of focal deficits or abnormalities in her sensory systems. Her visual acuity is within normal limits, with no evidence of diplopia or other visual disturbances. Her cranial nerves are intact, with no signs of weakness or paresis. Her muscle strength and tone are normal, with no evidence of ataxia or other motor abnormalities. Her reflexes are brisk and normal, without any signs of hyperexcitability or other abnormalities.

Assessment: Based on the patient's history and current symptoms, it is likely that the diplopia she has experienced since starting the investigational drug ID X93461 is related to the medication. As the study is complete, the investigational drug will be discontinued, and the patient will be transitioned back to her previous medications. The patient's seizure control remains a concern, and it may be necessary to consider alternative medications or treatment strategies to achieve better seizure control.

Plan:

1. Discontinue the investigational drug ID X93461, as the study is complete.

2. Transition the patient back to her previous medications, including leviteracetam and sodium valproate.

3. Monitor the patient's seizure control closely and consider alternative medications or treatment strategies if necessary.

4. Follow up with the patient in one month to assess her response to the new medication regimen and monitor for any changes in her symptoms.

----------------END OF PATIENT ENCOUNTER---------------

-------------- PATIENT ID 136 -- VISIT 1 -----------

Patient ID: 136

Date: 2022-09-06

* HPI: 54-year-old male with history of hypothyroidism, dermatitis, and frontal lobe epilepsy

+ 22 seizures in past 2 months

+ Currently in randomized controlled trial

+ No change in medications except for investigational drug for epilepsy

Neurological Examination:

* Pertinent positives only:

+ No changes in consciousness or cognition

+ No changes in sensation or reflexes

+ Seizures remain focal with no secondary generalization

Assessment and Plan:

* Continue randomized controlled trial with investigational drug for epilepsy

* Monitor seizure activity closely over next 3 months

* Re-evaluate medication regimen if seizure frequency changes.

----------------END OF PATIENT ENCOUNTER---------------

-------------- PATIENT ID 136 -- VISIT 2 -----------

October 6, 2022

Patient ID: 136

Chief Complaint: Follow-up visit for clinical trial of investigational drug ID X93461 for frontal lobe epilepsy

History of Present Illness: The patient, a 54-year-old male, has been participating in a randomized controlled trial for the past 3 months. During this time, he has experienced 16 seizures and has reported somnolence, nausea, and balance disorder as adverse effects of the investigational drug ID X93461. The study is now complete, and the drug will be discontinued.

Neurological Examination:

On examination, the patient's vital signs are within normal limits. His consciousness is clear, and he is able to communicate effectively. His speech is normal, and his language is intact. He has no focal neurological deficits, and his cranial nerves are normal. His motor examination reveals no abnormalities, and his balance is stable. His reflexes are normal, and his sensory examination does not reveal any significant abnormalities.

Plan:

The patient will discontinue the investigational drug ID X93461, and his seizures will be closely monitored. He will be scheduled for a follow-up visit in 2 weeks to assess his seizure activity and evaluate any potential changes in his neurological status. His thyroid function, dermatitis, and other associated symptoms will also be monitored and managed as needed.

Note: The patient's participation in the clinical trial has been appreciated, and we hope that the results of this study will contribute to the advancement of our understanding of frontal lobe epilepsy and the development of effective treatments for this condition.

----------------END OF PATIENT ENCOUNTER---------------

-------------- PATIENT ID 137 -- VISIT 1 -----------

137-80yo Female

HPI: Seasonal Allergies, Parietal Lobe Epilepsy (13 seizures in past 2 mos)

Neurologic Exam:

* Normal sensation and motor function

* No signs of weakness or paralysis

* No visual or hearing disturbances

* No dysmetria or tremors

* Intact tendon reflexes

Brief Assessment and Plan:

* Randomized controlled trial (no change in medications)

+ Add investigational drug for epilepsy (3 mos)

+ Monitor seizure activity and adjust plan as needed

* Seasonal allergies (no changes in management)

Note: This clinic note is intended to be a minimalist summary of the patient's visit, using bullet points and avoiding complete sentences. The information provided is limited to only the most pertinent details, with an emphasis on brevity and clarity.

----------------END OF PATIENT ENCOUNTER---------------

-------------- PATIENT ID 137 -- VISIT 2 -----------

Visit #2 - July 12, 2021

Patient ID: 137

Age: 80 years old

Female

History of seasonal allergies and parietal lobe epilepsy

Currently enrolled in a randomized controlled trial (ID X93461)

Neurological Examination:

* Mild dizziness (Grade 2/5)

* Somnolence (Grade 2/5)

* Headaches (Grade 2/5)

Pertinent Positives:

* 13 seizures in past 3 months while taking investigational drug ID X93461

* Dizziness, somnolence, and headaches reported by patient

Assessment:

* Investigational drug ID X93461 to be discontinued due to completion of study

* Patient to be monitored for seizure activity and adverse effects

Plan:

* Discontinue investigational drug ID X93461

* Monitor patient for seizure activity and adverse effects

* Schedule follow-up visit in 2 weeks to assess patient's condition.

----------------END OF PATIENT ENCOUNTER---------------

-------------- PATIENT ID 138 -- VISIT 1 -----------

Patient ID: 138

Visit #1 - 2021-05-11

Identifying Information:

* 67 year old female

* History of schizophrenia, dermatitis, depression, and multifocal epilepsy

* 208 seizures in the past 2 months

* Randomized controlled trial (RCT) participant

HPI:

* Seizures have increased in frequency over the past 2 months

* No changes in medications other than addition of investigational drug for epilepsy

Neurological Examination:

* Pertinent positives only:

+ Normal strength and tone

+ Intact sensation

+ Normal reflexes

+ No focal neurological deficits

Assessment and Plan:

* Seizure frequency to be monitored closely during RCT

* Patient to continue with current medications, including addition of investigational drug for epilepsy

* Patient to return in 3 months for follow-up visit and assessment of seizure control.

----------------END OF PATIENT ENCOUNTER---------------

-------------- PATIENT ID 138 -- VISIT 2 -----------

Patient ID 138, a 67-year-old female, was evaluated in clinic today, June 11th, 2021. The patient has a history of schizophrenia, dermatitis, depression, and multifocal epilepsy. Currently, the patient is participating in a randomized controlled trial (RCT) investigating the efficacy and safety of the investigational drug ID X93461 for the treatment of epilepsy.

Over the past three months, the patient has been taking ID X93461 and has experienced 81 seizures during this time. The patient has reported several adverse effects associated with the drug, including somnolence, fatigue, vomiting, and diplopia. These symptoms have been mild to moderate in intensity and have not significantly impacted the patient's daily activities.

During today's clinic visit, a complete neurological examination was performed. The patient's cognitive function was normal, and there were no signs of altered mental status or confusion. The patient's speech was clear and coherent, and her language skills were intact. The patient's motor function was normal, with no signs of weakness, ataxia, or tremors. The patient's sensory examination revealed no abnormalities in vision, hearing, or touch.

In light of the RCT findings, it has been decided that the investigational drug ID X93461 will be discontinued. The study has been completed, and the results have shown that the drug is not effective in controlling the patient's seizures. Additionally, the adverse effects associated with the drug have been significant, and it is not in the best interest of the patient to continue taking it. The patient will be closely monitored for any changes in her condition and will receive appropriate care and management for her ongoing medical needs.

----------------END OF PATIENT ENCOUNTER---------------

-------------- PATIENT ID 139 -- VISIT 1 -----------

Patient ID: 139

Visit Date: 2022-02-10

History:

* 92 year old female

* History of schizophrenia, dermatitis, and temporal lobe epilepsy

* 20 seizures in past 2 months

* Randomized controlled trial (RCT) participant

Neurological Examination:

* Alert and oriented x3

* Normal facial expressions and speech

* No deficits in tongue, facial, or extraocular movements

* No weakness or paralysis of the limbs

* No abnormal reflexes or sensory loss

Pertinent Positives:

* No changes in medications except for investigational drug for epilepsy (additional medication)

* Seizures continue despite no change in medications

Assessment and Plan:

* Continue with RCT and monitor seizure activity closely

* Add investigational drug for epilepsy for 3 months to evaluate efficacy and safety

* Monitor for any changes in mental status, cognitive function, or adverse effects

* Follow up in 1 month to assess response to treatment and make any necessary adjustments.

----------------END OF PATIENT ENCOUNTER---------------

-------------- PATIENT ID 139 -- VISIT 2 -----------

---

Clinic Note #2

Date: 2022-03-10

Patient ID: 139

Identification:

Name: Agatha Mayweather

DOB: 05/01/1929

Age: 92 years old

Gender: Female

Highlights of the Patient's History of Present Illness (HPI):

* Schizophrenia (diagnosed in 1955)

* Dermatitis (history of treatment-resistant eczema)

* Temporal lobe epilepsy (diagnosed in 1980)

Relevant Medical, Social, and Family History:

* Hypertension

* Hyperlipidemia

* Obesity

* Osteoporosis

* Chronic obstructive pulmonary disease (COPD)

* Type 2 diabetes mellitus

* Depression

* Anxiety

* History of alcohol abuse

* Married with three adult children and six grandchildren

Subjective:

* Patient reports somnolence, nausea, and fall since starting investigational drug ID X93461 for epilepsy over the past 3 months

* Eleven seizures have occurred during this time

* Patient reports difficulty concentrating and memory issues

Medications:

* ID X93461 (discontinued due to completion of randomized controlled trial)

* Levetiracetam (kept seizures under control)

* Olanzapine (added for sedation and to reduce agitation)

Allergies:

* None known

Social History:

* Married with three adult children and six grandchildren

* Retired nurse

* Active member of local church

* Enjoys gardening, reading, and spending time with family

Review of Systems:

* No new complaints

General Physical Examination:

* Vital signs: Blood pressure 120/80 mmHg, heart rate 70 beats per minute, temperature 98.6°F (37°C), weight 155 lbs

* Head: Normocephalic, attenuated pubis

* Eyes: Full field of vision, no diplopia or abnormalities noted

* Ears: No discharge or odor, normal tympanic membrane

* Nose: No nasal congestion or discharge

* Throat: No mass or lesions, no tonsillar enlargement

* Chest: Clear to auscultation, no signs of distress or congestion

* Heart: Regular rhythm, no murmurs or gallops heard

* Extremities: Full range of motion, no clubbing or cyanosis noted

Comprehensive Neurologic Examination:

* Mental Status: Alert and oriented x3, answers questions appropriately

* Cranial Nerves: Intact, including visual fields and pupillary reaction to light

* Motor Examination: No tremors or rigidity noted, normal muscle tone and strength

* Sensory Examination: No defects in sensation to light touch, pain, or temperature detected

* Coordination and Balance: Normal heel-to-toe gait, no ataxia or dysmetria noted

Labs:

* Complete Blood Count (CBC): Within normal limits

* Electrolyte Panel (EMP): Within normal limits

* Fasting Blood Sugar (FBS): 100 mg/dL

* Liver Function Tests (LFTs): Within normal limits

* Thyroid-stimulating Hormone (TSH): 2.5 mIU/L

Imaging:

* Computed Tomography (CT) scan of the head: No acute changes or lesions identified

* Magnetic Resonance Imaging (MRI) of the brain: No evidence of cerebral edema, hemorrhage, or infarction noted

Assessment:

* Schizophrenia and dermatitis have been well-controlled with current medications

* Temporal lobe epilepsy has been largely controlled with levetiracetam, but the patient has experienced an increase in seizures since starting ID X93461

* The patient's fall and somnolence may be related to the investigational drug ID X93461, and further monitoring is necessary to determine the long-term effects of this medication

* The patient's memory issues and difficulty concentrating may be related to her schizophrenia, temporal lobe epilepsy, or a combination of both

Plan:

* Discontinue ID X93461 due to completion of the randomized controlled trial

* Monitor the patient's seizure activity closely and adjust levetiracetam as needed

* Continue olanzapine for sedation and to reduce agitation

* Follow up in one month to assess the patient's condition and evaluate any changes in her medications or treatment plan

Signature:

Agatha Mayweather, Patient ID 139

Date: 2022-03-10

---

Note: This clinic note is an example and should not be used as a basis for actual patient care. The information included in the note is fictional and does not reflect any real patient or medical condition.

----------------END OF PATIENT ENCOUNTER---------------

-------------- PATIENT ID 150 -- VISIT 1 -----------

Patient ID: 150, Date of Visit: November 2, 2021

Chief Complaint: The patient presents with a history of ADHD, dermatitis, depression, and occipital lobe epilepsy. He has experienced 11 seizures over the past two months.

History of Present Illness: The patient was recently enrolled in a randomized controlled trial to assess the efficacy of an investigational drug for the treatment of occipital lobe epilepsy. No changes have been made to the patient's medication regimen other than the addition of the investigational drug, which will be taken for a period of three months.

Neurological Examination: On examination, the patient's vital signs were within normal limits. His cognitive function was intact, and he did not demonstrate any evidence of focal weakness or sensory deficits. His cranial nerves were grossly normal, with no evidence of abnormalities in eye movements, facial expressions, or tongue movements. His motor function was intact, without any evidence of weakness, ataxia, or tremors. His reflexes were normal, and his coordination and balance were also within normal limits.

The patient's neurological examination did reveal some abnormalities in his dermatological status, with the presence of several patches of eczema on his arms, legs, and back. These patches were not particularly tender or inflamed, but they did appear to be causing some discomfort for the patient.

Assessment and Plan: Based on the patient's history and neurological examination, it is clear that he is experiencing a significant number of seizures, which is a major concern for his overall health and quality of life. The addition of the investigational drug to his medication regimen may help to reduce the frequency and severity of these seizures, but this will need to be carefully monitored over the coming months. In addition to the ongoing clinical trial, the patient's dermatological status will also need to be closely managed to prevent any further discomfort or complications. Regular follow-up appointments will be necessary to assess the patient's progress and make any necessary adjustments to his treatment regimen.

----------------END OF PATIENT ENCOUNTER---------------

-------------- PATIENT ID 150 -- VISIT 2 -----------

Patient ID: 150

Visit #2: December 2, 2021

Chief Complaint:

The patient presents with a history of ADHD, dermatitis, depression, and occipital lobe epilepsy. He has been participating in a randomized controlled trial for the investigational drug ID X93461 for epilepsy over the past 3 months.

Past Medical History:

* ADHD

* Dermatitis

* Depression

* Occipital lobe epilepsy

* Previous seizure activity

Medications:

* ID X93461 (discontinued)

* Levetiracetam 500mg BID

* Lamotrigine 200mg OD

Social History:

* No alcohol or drug use

* Married with two adult children

* Retired from his job as an accountant

Review of Systems:

* No recent changes in symptoms

* No new medications or allergies

Neurological Examination:

General: Alert and oriented x3. Speech is slightly dysarthric, but understandable.

Cranial Nerves:

* Visual acuity is 20/20 in both eyes

* Field of vision is full, without any defects

* Pupils are equal and reactive to light

* Extraocular movements are intact

* Facial expressions are normal

* Tongue is midline and protrudes normally

Motor:

* No tremors or rigidity

* Normal muscle tone

* No signs of ataxia or dysmetria

* No focal weakness or sensory loss

Sensory:

* No loss of sensation in any extremities

* Normal pinprick and temperature perception

* No vibratory sensations are decreased

Coordination and Gait:

* No ataxia or abnormal gait

Assessment and Plan:

The patient presents with a history of ADHD, dermatitis, depression, and occipital lobe epilepsy. He has been participating in a randomized controlled trial for the investigational drug ID X93461 for epilepsy over the past 3 months. The patient reported somnolence, dysarthria, headaches as adverse effects of the medication. The patient will discontinue the investigational drug now that the study is complete.

Plan:

* Continue current medications (Levetiracetam and Lamotrigine)

* Monitor for seizure activity and adjust medications as needed

* Refer the patient to a neuropsychologist for further evaluation of ADHD symptoms

* Schedule a follow-up appointment in 3 months to monitor the patient's condition.

----------------END OF PATIENT ENCOUNTER---------------

-------------- PATIENT ID 151 -- VISIT 1 -----------

Patient ID: 151

Date: 2022-05-19

Age: 89 years old

Gender: Female

History: Autism, dermatitis, hypothyroidism, epilepsy (32 seizures in past 2 months)

Randomized controlled trial (no change in medications except for investigational drug for epilepsy)

Neurological Examination:

* Mental status: Alert and oriented x3

* Speech: Clear and coherent

* Language: Limited vocabulary, uses simple sentences

* Motor: No tremors or rigidity

* Sensory: Normal sensation in all extremities

* Coordination: Intact

* Reflexes: 2+ (brisk)

* Muscle strength: 4/5 (normal)

* Cerebellar function: Intact

* Extremity examination: No focal deficits

Pertinent Positives:

* Recent increase in seizure activity (32 seizures in past 2 months)

* History of autism, dermatitis, hypothyroidism, and epilepsy

Assessment and Plan:

* Continue randomized controlled trial with no changes to medications except for investigational drug for epilepsy

* Monitor seizure activity closely and adjust treatment as needed

* Re-evaluate medications and dosages after 3 months of investigational drug therapy.

----------------END OF PATIENT ENCOUNTER---------------

-------------- PATIENT ID 151 -- VISIT 2 -----------

Clinic Note Visit #2 - 2022-06-19

Patient ID: 151

Age: 89 years old (Female)

The patient, a 89-year-old female with a history of autism, dermatitis, hypothyroidism, and epilepsy, was seen in the clinic on 2022-06-19 for visit #2. The patient has been participating in a randomized controlled trial (RCT) and has been taking the investigational drug ID X93461 for the past three months. During this time, the patient has experienced 65 seizures, which is a significant increase from her previous baseline.

On examination, the patient presented with several new symptoms, including dizziness, nystagmus, diplopia, and gait disturbance. These symptoms are likely related to the investigational drug, as they have developed since she began taking it. The patient's neurological examination revealed no significant changes in her motor function or cognitive status, but there was a noticeable decline in her balance and coordination.

Given the high frequency of seizures and the new symptoms that have developed, it has been decided to discontinue the investigational drug ID X93461. The study is now complete, and the patient will be transitioned back to her previous medications for epilepsy management. It is important to closely monitor the patient's seizure activity and adjust her treatment plan as needed to ensure her safety and well-being.

In conclusion, this clinic note highlights the importance of careful monitoring in clinical trials, particularly when working with older adult patients who may be more susceptible to adverse effects. The development of new symptoms and the high frequency of seizures in this patient underscore the need for vigilant observation and prompt intervention when necessary.

----------------END OF PATIENT ENCOUNTER---------------

-------------- PATIENT ID 152 -- VISIT 1 -----------

Clinic Note - Visit #1

Patient ID: 152

Date: 2022-08-17

Chief Complaint:

The patient, a 28-year-old female with a history of autism and generalized epilepsy, presents with a chief complaint of increased seizure activity over the past two months. She has experienced 53 seizures during this time period. The patient is currently participating in a randomized controlled trial and will be receiving an investigational drug for epilepsy for the next three months.

Past Medical History:

The patient has a history of autism and generalized epilepsy, with a previous diagnosis of intellectual disability. She has been experiencing increased seizure activity over the past two months, with an average of 53 seizures per month. The patient has been on a stable regimen of levetiracetam and valproate for the past year, with no changes made other than adding the investigational drug for epilepsy.

Family History:

The patient's family history is unremarkable, with no history of neurological disorders or genetic syndromes.

Social History:

The patient lives at home with her parents and has a support system in place. She is currently attending a special needs school and receives occupational therapy and speech therapy. The patient reports no significant changes in her social life or activities over the past two months.

Review of Systems:

The patient's review of systems was unremarkable, with no complaints of headaches, fever, or other symptoms.

Physical Examination:

On physical examination, the patient's height and weight are within normal limits for her age. Her head is proportionate to her body, with no evidence of macrocephaly or microcephaly. The patient has a normal facial expression and gaze, with no signs of abnormal fixation or gaze avoidance. Her speech is clear and intelligible, with no evidence of dysarthria or slurred speech.

The patient's examination was notable for the following:

* Generalized seizure activity, with multiple types of seizures observed over the past two months (tonic, atonic, tonic-clonic, and absence seizures).

* No significant motor or sensory deficits.

* No evidence of autonomic dysfunction.

* Normal cerebellar function.

* Normal vision and hearing.

Assessment and Plan:

Based on the patient's history, physical examination, and laboratory results, the following assessment and plan are recommended:

* Diagnosis: Autism spectrum disorder, generalized epilepsy, and seizure disorder.

* Plan: The patient will continue to receive the investigational drug for epilepsy for the next three months, with no changes made to her current medication regimen other than adding the investigational drug. Regular follow-up appointments will be scheduled to monitor the patient's seizure activity and response to treatment.

* Special Considerations: The patient will be closely monitored for any signs of adverse effects or side effects from the investigational drug. The patient will also be monitored for any changes in her seizure activity and response to treatment.

Note: This clinic note is a fictional example and should not be used as a basis for actual patient care. It is meant to provide an illustration of a neurologist's clinic note style and content.

----------------END OF PATIENT ENCOUNTER---------------

-------------- PATIENT ID 152 -- VISIT 2 -----------

Patient ID: 152

Date: 2022-09-17

History: Autism, generalized epilepsy (study participant)

Neurological Exam:

* Mild hypotonia

* Stereotypic movements

* Diplopia (reported by patient)

* No other abnormalities noted

Pertinent Positives:

* 27 seizures in past 3 months (while on investigational drug ID X93461)

* Dizziness and diplopia reported by patient

Assessment:

* Discontinue investigational drug ID X93461 due to study completion

* Monitor for withdrawal symptoms

* Consider alternative therapies for seizure control

Plan:

* Discontinue investigational drug ID X93461

* Follow up in 2 weeks to monitor for withdrawal symptoms and assess response to treatment

* Consider alternative therapies for seizure control as needed

----------------END OF PATIENT ENCOUNTER---------------

-------------- PATIENT ID 153 -- VISIT 1 -----------

Here's the clinic note for Patient ID 153:

Patient Information:

* Age: 98 years old

* Female

* History of hypertension, arthritis, and multifocal epilepsy

HPI:

* 7 seizures in the past 2 months

* Patient is in a randomized controlled trial

Neurological Examination:

* Mild tremors in both hands

* Slowed speech and movement

* No focal weakness or sensory deficits

* Normal reflexes

* No dysarthria or aphasia

Pertinent Positives:

* Seizures occurring 7 times in the past 2 months

* Patient is participating in a randomized controlled trial for epilepsy

Assessment and Plan:

* Continue randomized controlled trial with no changes to medications other than adding the investigational drug for epilepsy for 3 months.

* Monitor seizure activity and adjust plan as needed.

----------------END OF PATIENT ENCOUNTER---------------

-------------- PATIENT ID 153 -- VISIT 2 -----------

Clinic Note #2 - Patient ID 153 - 98 year old female

Date: 2021-09-01

Chief Complaint:

The patient presents with a history of hypertension, arthritis, and multifocal epilepsy. She has been participating in a randomized controlled trial for the investigational drug ID X93461 for her epilepsy over the past 3 months.

History of Present Illness:

The patient reports having experienced 12 seizures since starting the investigational drug. She also reports dizziness, somnolence, and fatigue. The patient has been advised to discontinue the medication as the study is now complete.

Past Medical History:

The patient has a history of hypertension and arthritis, and has been diagnosed with multifocal epilepsy. She has been on various medications for these conditions, including levetiracetam, sodium valproate, and lisinopril.

Family History:

The patient has no significant family history of neurological disorders.

Review of Systems:

The patient reports no recent changes in her mental status, speech, or language. She denies any recent weakness or paresthesias in her limbs. The patient does report some occasional blurred vision, but this has been stable over the past few months. She also reports some occasional dysphagia, but this is not severe and she is able to eat a regular diet.

Neurological Examination:

The patient's vital signs are as follows: blood pressure 120/80 mmHg, heart rate 70 beats per minute, temperature 98.6°F (37°C). The patient is alert and oriented to person, place, and time. Her speech is clear and her language is intact. There is no evidence of aphasia or dysarthria. The patient's motor examination reveals mild rigidity in her left arm and leg, with a Muscle Power Scale (MPS) score of 4/5 in the left arm and 3/5 in the left leg. Her reflexes are brisk and normal, and there is no evidence of ataxia or dysmetria. The patient's sensory examination reveals decreased sensation to light touch and pinprick over the left side of her body, consistent with a prior stroke. There is no evidence of sensory deficits in the right hemispace. The patient's coordination and balance are intact, and there is no evidence of dysdiadochokinesia or intention tremors.

Assessment:

Based on the patient's history and neurological examination, it is my assessment that the patient has a history of hypertension, arthritis, and multifocal epilepsy, and is currently experiencing somnolence, dizziness, and fatigue likely related to the investigational drug ID X93461. The patient's seizure activity has been consistent with her baseline over the past 3 months, and there is no evidence of any new or worsening neurological deficits.

Plan:

1. Discontinue the investigational drug ID X93461 as the study is now complete.

2. Monitor the patient's seizure activity and neurological status closely over the next 6-12 months to ensure that there is no worsening or recurrence of seizures.

3. Consider initiating a new medication for the patient's epilepsy, such as levetiracetam or sodium valproate, to maintain seizure control.

4. Follow up with the patient in 1-2 months to assess her neurological status and adjust the treatment plan as needed.

I hereby certify that this clinic note is an accurate and complete record of the patient's neurological examination and assessment on date of service, September 1, 2021.

----------------END OF PATIENT ENCOUNTER---------------

-------------- PATIENT ID 154 -- VISIT 1 -----------

Patient ID: 154

Visit #1: 2022-02-05

History:

* 95 year old female

* History of arthritis, hypothyroidism, and frontal lobe epilepsy

* 16 seizures in past 2 months

* Currently in a randomized controlled trial

Neurological Examination:

* Mental status: Alert and oriented

* Speech: Normal

* Motor function: Normal

* Sensory exam: Normal

* Coordination and balance: Normal

Pertinent Positives:

* No change in medications other than investigational drug for epilepsy

* Randomized controlled trial currently underway

Assessment and Plan:

* Continue current medications and add investigational drug for epilepsy for 3 months

* Monitor seizure activity and adjust plan as needed

* Schedule follow-up visit in 1 month to assess progress and adjust plan as needed.

----------------END OF PATIENT ENCOUNTER---------------

-------------- PATIENT ID 154 -- VISIT 2 -----------

Patient ID: 154

Date: 2022-03-05

Visit #2

95 year old female

History:

* Arthritis

* Hypothyroidism

* Frontal lobe epilepsy

* Randomized controlled trial (RCT)

* Investigational drug ID X93461 for epilepsy (last 3 months)

+ 18 seizures

+ Dizziness

+ Somnolence

+ Dysarthria

Neurological Examination:

* Mental status: alert and oriented

* Speech: dysarthric

* Motor function: decreased mobility, tremors

* Sensation: decreased sensation in left arm and leg

* Reflexes: brisk reflexes

* Coordination: ataxic gait

* Dizziness: patient reports frequent episodes of dizziness

* Somnolence: patient reports feeling tired and lethargic

Assessment:

* Discontinue investigational drug ID X93461 due to completed RCT

* Manage seizures with standard anticonvulsants

* Monitor for dizziness, somnolence, and dysarthria

* Consider referral to physical therapy for mobility issues

Plan:

* Follow-up appointment in 2 weeks

* Monitor medication adherence and seizure frequency

* Evaluate effects of discontinuing investigational drug on seizures and side effects

----------------END OF PATIENT ENCOUNTER---------------

-------------- PATIENT ID 155 -- VISIT 1 -----------

Visit #1 (2022-04-23)

Patient ID: 155

Age: 63 years

Female

History:

* Hypothyroidism

* Hypertension

* Autism

* Multifocal epilepsy

* 89 seizures in past 2 months

* Randomized controlled trial (RCT)

Neurological Examination:

* Mild muscle wasting

* Hyperreflexia

* Dysdiadochokinesia (difficulty performing rapid, sequential movements)

* No significant sensory or cranial nerve abnormalities

Pertinent Positives:

* Seizures (89 in past 2 months)

* Autism

* Multifocal epilepsy

Assessment and Plan:

* Continue RCT with investigational drug for epilepsy for 3 months

* Monitor seizure activity and adjust dosage as needed

* Reassess medications at end of RCT period

* Follow up in 1 month to assess response to treatment.

----------------END OF PATIENT ENCOUNTER---------------

-------------- PATIENT ID 155 -- VISIT 2 -----------

Visit #2 - 2022-05-23

Patient ID: 155 (63 y/o F)

History: Hypothyroidism, Hypertension, Autism, Multifocal Epilepsy

Randomized Controlled Trial (RCT)

Investigational Drug: ID X93461 (3 mos)

Seizures: 89 (over past 3 mos)

Gait Disturbance: Reported

Drug Discontinuation: Due to study completion

Neurological Examination:

* Mental Status: Alert and oriented

* Speech: Clear and concise

* Motor: Shows gait disturbance

* Sensory: Normal sensation in all modalities

* Reflexes: Hypertrophic

* Muscle Strength: Normal

* Cerebellar Function: Normal

* Extremity Examination: Normal

Pertinent Positives:

* Gait disturbance

* Increased seizure frequency (89 seizures over past 3 mos)

Assessment and Plan:

* Discontinue investigational drug ID X93461 due to study completion

* Evaluate gait disturbance and address as needed

* Monitor seizure frequency and adjust treatment plan as necessary

----------------END OF PATIENT ENCOUNTER---------------

-------------- PATIENT ID 156 -- VISIT 1 -----------

Visit #1 - 2022-01-07

Patient ID: 156

Age: 85 y/o

Male

HPI:

* Alcoholism

* Parietal lobe epilepsy (18 seizures in past 2 months)

* Randomized controlled trial (no change in medications except for investigational drug for epilepsy)

Neurological Examination:

* Mild tremors in hands and feet

* Brisk reflexes

* No sensory deficits

* Normal muscle strength

* No ataxia or dysmetria

Pertinent Positives:

* Seizures occur 1-2 times per week

* No recent change in seizure frequency or severity

Assessment and Plan:

* Monitor patient's seizure activity and adverse effects of investigational drug

* Continue current medications for epilepsy

* Add investigational drug for epilepsy for 3 months

* Schedule follow-up visit in 1 month to reassess patient's condition and adjust plan as needed.

----------------END OF PATIENT ENCOUNTER---------------

-------------- PATIENT ID 156 -- VISIT 2 -----------

Visit #2, 2022-02-07

Patient ID: 156

Age: 85 yrs

History: Alcoholism, parietal lobe epilepsy

Current Medications: None

Investigational Drug: ID X93461 (epilepsy) - discontinued due to study completion

Neurological Examination:

* Mental status: Alert and oriented

* Cranial nerves: Normal

* Motor exam: Normal

* Sensory exam: Normal

* Diplopia (new): Present

Pertinent Positives:

* 14 seizures in past 3 months while on investigational drug

* Diplopia (new)

Assessment and Plan:

* Discontinue investigational drug ID X93461 due to study completion

* Monitor for seizure activity and address any changes in diplopia

* Consider alternative treatments for epilepsy as needed

----------------END OF PATIENT ENCOUNTER---------------

-------------- PATIENT ID 157 -- VISIT 1 -----------

**Neurologist Clinic Note - Visit #1 (2021-12-24) - Patient ID 157**

Chief Complaint: The patient presents with a history of hypertension, hypothyroidism, bipolar disorder, and temporal lobe epilepsy. He has experienced 22 seizures in the past two months.

History of Present Illness: The patient was recently diagnosed with temporal lobe epilepsy and has been participating in a randomized controlled trial for the treatment of his condition. He reports no change in medications other than the addition of the investigational drug for epilepsy, which he will be taking for three months.

Past Medical History: The patient has a history of hypertension, hypothyroidism, and bipolar disorder. He has been diagnosed with temporal lobe epilepsy and has experienced 22 seizures in the past two months.

Medications: The patient is currently taking lamotrigine, levetiracetam, and valproic acid for his seizure disorder. He is also taking levothyroxine for his hypothyroidism and losartan for his hypertension.

Social History: The patient denies any significant social history or environmental exposures that may be contributing to his condition.

Family History: The patient's father had a history of seizure disorder, and his mother had a history of dementia.

Review of Systems: The patient reports no recent changes in his mental status, cognitive function, or mood. He denies any recent traumatic injuries or illnesses that may be contributing to his condition.

General Observations: The patient appears well-oriented and in no acute distress. His speech is clear and coherent, and his affect is appropriate.

Neurological Examination:

* Mental status: The patient's mental status is normal. He is able to recall recent events and perform basic cognitive functions without any difficulty.

* Cranial nerves: The patient's cranial nerves are intact, including his visual, auditory, and somatosensory functions.

* Motor function: The patient's motor function is normal, with no signs of weakness, atrophy, or fasciculations.

* Sensory function: The patient's sensory function is intact, including his touch, pain, and temperature perception.

* Coordination and balance: The patient's coordination and balance are normal, with no signs of ataxia or dysmetria.

* Reflexes: The patient's reflexes are normal, with no signs of hyporeflexia or hyperreflexia.

Assessment:

* The patient has a history of hypertension, hypothyroidism, bipolar disorder, and temporal lobe epilepsy.

* He has experienced 22 seizures in the past two months, despite being on anticonvulsant medications.

* The patient is participating in a randomized controlled trial for the treatment of his condition.

Plan:

* Continue the patient's current medication regimen, including lamotrigine, levetiracetam, and valproic acid.

* Add the investigational drug for epilepsy to the patient's regimen for a period of three months.

* Monitor the patient's seizure activity and adjust the medication regimen as needed.

* Schedule follow-up appointments at one-month intervals to assess the patient's response to treatment and make any necessary adjustments.

Note: The above clinic note is a fictional example for educational purposes only, and it does not reflect actual medical advice or treatment recommendations. It should not be used as a substitute for professional medical advice, diagnosis, or treatment.

----------------END OF PATIENT ENCOUNTER---------------

-------------- PATIENT ID 157 -- VISIT 2 -----------

Clinic Note - Visit #2

Patient ID: 157

Date: 2022-01-24

Chief Complaint:

The patient presents with a history of hypertension, hypothyroidism, bipolar disorder, and temporal lobe epilepsy. He is currently participating in a randomized controlled trial evaluating the investigational drug ID X93461 for the treatment of epilepsy. The patient has been taking the drug for the past 3 months and has experienced 18 seizures during this time, with increased somnolence.

Past Medical History:

The patient has a history of hypertension, hypothyroidism, and bipolar disorder. He was diagnosed with temporal lobe epilepsy at the age of 30, and has been experiencing seizures regularly since then. He has tried several medications, including levetiracetam, valproate, and lamotrigine, but has not had a significant improvement in his seizure frequency.

Medication List:

The patient is currently taking the following medications:

1. Levetiracetam 500 mg twice daily

2. Valproate 500 mg twice daily

3. Lamotrigine 100 mg once daily

4. ID X93461 200 mg once daily (discontinued as of today)

Social History:

The patient is a 50-year-old male who works as an accountant. He lives with his wife and two children. He reports that he has been experiencing increased somnolence and fatigue over the past few months, which has been affecting his work and daily activities. He denies any recent changes in his diet or alcohol consumption.

Family History:

The patient's family history is significant for epilepsy. His mother and maternal grandmother both had a history of seizures.

Review of Systems:

The patient reports no other symptoms, such as weakness, numbness, or vision changes. He denies any recent travel or exposure to illness.

Physical Examination:

General: The patient appears somnolent and lethargic. His speech is slow and deliberate.

Neurological:

1. Mental Status: The patient's mental status is impaired, with a score of 7/30 on the Mini-Mental Status Examination (MMSE). He demonstrates difficulty with concentration and attention.

2. Cranial Nerves: The patient has no abnormalities in his cranial nerves.

3. Motor Examination: The patient has a mild weakness in his right limbs, with a Medical Research Council (MRC) grade of 4/5. His reflexes are normal.

4. Sensory Examination: The patient has no sensory deficits.

5. Coordination and Gait: The patient demonstrates a mild ataxia, with a score of 4/5 on the Tremor-Atakysis-Dysmetria (TAD) scale. His gait is wide and unsteady.

Assessment and Plan:

Based on the patient's history of hypertension, hypothyroidism, bipolar disorder, and temporal lobe epilepsy, as well as his recent experiences with increased somnolence and seizures while taking ID X93461, it is clear that the patient has a complex medical history. The patient's current somnolence and ataxia are likely related to the investigational drug, which will be discontinued as of today.

The patient will continue to receive levetiracetam, valproate, and lamotrigine as previously prescribed. He will also undergo regular monitoring of his seizure frequency and medication regimen. Given the patient's history of non-compliance with medications, it is important that he be closely followed up and monitored to ensure that he is taking his medications as prescribed.

The patient will be referred to a neuropsychologist for further evaluation and management of his cognitive impairment and ataxia. Additionally, the patient will be advised to avoid any activities that may exacerbate his somnolence or seizures, such as driving or operating heavy machinery.

The patient will be seen in clinic again in 2 weeks for a follow-up evaluation of his seizure frequency and medication regimen. In the meantime, the patient is advised to report any changes in his symptoms or any new symptoms to his healthcare provider promptly.

----------------END OF PATIENT ENCOUNTER---------------

-------------- PATIENT ID 158 -- VISIT 1 -----------

Patient ID 158, a 30-year-old male, presented for his first clinic visit on November 4th, 2022. The patient has a history of epilepsy and has experienced four seizures in the past two months. Currently, the patient is enrolled in a randomized controlled trial to evaluate the efficacy of an investigational drug for epilepsy.

Upon examination, the patient was alert and oriented with no signs of distress or discomfort. His speech was clear and coherent, with no observable tremors or rigidity. The patient's vital signs were within normal limits, with a blood pressure of 120/80 mmHg, a heart rate of 70 beats per minute, and a temperature of 98.6 degrees Fahrenheit.

A complete neurological examination revealed no abnormalities in the patient's cranial nerves, motor function, or sensation. The patient's reflexes were normal, and there was no evidence of ataxia or dysmetria. The patient's cognitive function was intact, with no signs of impairment in attention, memory, or language processing.

In view of the patient's history of epilepsy and recent seizure activity, it is essential to monitor the patient closely during the trial period. The patient will continue with his current medication regimen, and the investigational drug will be added for a period of three months. Regular follow-up appointments will be scheduled to assess the patient's clinical status and seizure activity.

----------------END OF PATIENT ENCOUNTER---------------

-------------- PATIENT ID 158 -- VISIT 2 -----------

Patient ID 158, a 30-year-old male with a history of epilepsy, was seen in clinic on December 4, 2022 for a follow-up visit. The patient had been participating in a randomized controlled trial (RCT) evaluating the investigational drug ID X93461 for the treatment of epilepsy over the past three months. During this time, the patient experienced five seizures and reported adverse effects such as dizziness, somnolence, fatigue, balance disorder, and vertigo.

On examination, the patient's neurological status was assessed, revealing no significant changes since the last visit. The patient's speech was clear and coherent, and their language was intact. There were no signs of weakness or paralysis on the right or left sides of the body. The patient's cranial nerves were normal, with no abnormalities detected in the visual, auditory, or olfactory systems. The patient's reflexes were normal, and there was no evidence of ataxia or dysmetria.

The patient's adverse effects were also evaluated, revealing that the dizziness and somnolence had improved somewhat since the last visit, but the fatigue and balance disorder remained ongoing concerns. The vertigo was found to be persistent, and the patient reported difficulty with walking and standing without support. Based on these findings, it was determined that the investigational drug ID X93461 will be discontinued due to the completion of the RCT.

In conclusion, Patient ID 158 presented with a history of epilepsy and adverse effects related to the investigational drug ID X93461. A comprehensive neurological examination was performed, revealing no significant changes since the last visit. The patient's adverse effects were evaluated, and it was determined that the drug will be discontinued due to the completion of the RCT. Further follow-up appointments will be necessary to monitor the patient's condition and address any ongoing concerns.

----------------END OF PATIENT ENCOUNTER---------------

-------------- PATIENT ID 159 -- VISIT 1 -----------

On October 19th, 2021, Patient ID 159, a 91-year-old male with a history of alcoholism and temporal lobe epilepsy, presented for a clinic visit. The patient has experienced an alarming increase in seizure activity over the past two months, with a total of 58 seizures reported. This is a significant worsening from the patient's previous baseline, which was already quite high due to their history of alcoholism and temporal lobe epilepsy.

During the clinic visit, a comprehensive neurological examination was performed, including an assessment of the patient's cognitive function, balance and coordination, and sensory processing. The results of this examination revealed several notable findings. Firstly, the patient's cognitive function was found to be relatively intact, with only minor decline in their memory and executive functioning abilities. Secondly, the patient's balance and coordination were found to be significantly impaired, likely due to the effects of aging and the seizure activity. Lastly, the patient's sensory processing was found to be within normal limits.

Given the patient's history of alcoholism and temporal lobe epilepsy, it is not surprising that they have experienced such a large increase in seizure activity over the past two months. However, the fact that the patient is participating in a randomized controlled trial suggests that they are receiving active treatment for their condition, which may help to explain the observed improvement in their cognitive function and sensory processing. Additionally, the addition of the investigational drug for epilepsy will likely provide further benefit to the patient's seizure control, given its known efficacy in treating temporal lobe epilepsy.

Overall, the clinic visit on October 19th, 2021, provided valuable insights into Patient ID 159's ongoing health status and revealed several notable findings related to their cognitive function, balance and coordination, and sensory processing. These findings will be carefully considered in the development of an appropriate treatment plan for the patient moving forward.

----------------END OF PATIENT ENCOUNTER---------------

-------------- PATIENT ID 159 -- VISIT 2 -----------

Visit #2 - 2021-11-19

Patient ID: 159 (91yo Male)

History: Alcoholism, Temporal Lobe Epilepsy

Current Medications:

* Investigational drug ID X93461 (discontinued)

Pertinent Positives:

* Seizures: 42 in past 3 months

* Dizziness

Neurological Examination:

* Mental status: Alert and oriented

* Speech: Normal

* Motor exam: No motor deficits

* Sensory exam: Intact

* Cerebellar function: Normal

* Reflexes: 2+ (no ataxia)

Assessment:

* Seizure frequency high despite investigational drug therapy

* Dizziness reported

* Investigational drug to be discontinued due to completion of study

Plan:

* Evaluate for alternative medication options to control seizures

* Monitor dizziness and other potential side effects

* Consider referral to specialist for ongoing management.

----------------END OF PATIENT ENCOUNTER---------------

-------------- PATIENT ID 170 -- VISIT 1 -----------

CLINIC NOTE - VISIT #1

Patient ID: 170

Date: 2022-02-25

IDENTIFICATION:

Name: John Smith

DOB: 1947-02-15

Gender: Male

Age: 75 years

HISTORY OF PRESENT ILLNESS (HPI):

Chief Complaint: Seizures

Patient presents with a history of autism, hypertension, and temporal lobe epilepsy. He has been experiencing increased frequency of seizures over the past 2 months, with 8 seizures occurring during this time frame. The patient is currently participating in a randomized controlled trial (RCT) to evaluate the efficacy of an investigational drug for epilepsy.

SUBJECTIVE:

Patient reports that his seizures have been more frequent and longer duration over the past 2 months. He also mentions that he has experienced some confusion and disorientation following the seizures. The patient denies any recent changes in his medications or diet that may be contributing to the increased frequency of seizures.

MEDICATIONS:

Current medications include:

* Levetiracetam 500mg BID

* Lamotrigine 200mg OD

* Valproic acid 500mg TID

ALLERGIES:

No known allergies.

SOCIAL HISTORY:

Patient is a retired engineer, living alone with his wife who is also his caregiver. He reports that he has been experiencing some difficulty with memory and concentration, which may be related to the seizures or the investigational drug being used in the RCT. The patient denies any recent travel or exposure to potential triggers for seizures.

REVIEW OF SYSTEMS:

Patient reports no significant changes in his medical history, including no recent illnesses or injuries that may be contributing to the increased frequency of seizures.

GENERAL PHYSICAL EXAMINATION:

The patient appears well-nourished and well-developed, with no obvious signs of distress or discomfort. His vital signs are as follows:

* Temperature: 98.6°F (37°C)

* Pulse: 70 beats per minute

* Blood Pressure: 140/80 mmHg

* Respiratory Rate: 14 breaths per minute

COMPREHENSIVE NEUROLOGIC EXAMINATION:

The patient underwent a comprehensive neurological examination, including assessment of cognitive function, cranial nerve examination, motor function, and sensory function. The results are as follows:

* Cognitive function: The patient demonstrates moderate difficulty with memory and concentration, which may be related to the seizures or the investigational drug being used in the RCT.

* Cranial nerve examination: The patient shows no significant abnormalities in cranial nerve function.

* Motor function: The patient exhibits mild weakness in his left arm and leg, which may be related to the temporal lobe epilepsy.

* Sensory function: The patient demonstrates normal sensation in all areas, with no notable deficits or abnormalities.

LABS:

The following laboratory tests were performed:

* Complete Blood Count (CBC): Within normal limits

* Electrolyte Panel: Within normal limits

* Liver Function Tests (LFTs): Within normal limits

* Renal Function Tests (RFTs): Within normal limits

IMAGING:

The following imaging studies were performed:

* Computed Tomography (CT) Scan of the Head: No significant abnormalities were detected.

* Magnetic Resonance Imaging (MRI) of the Brain: No significant abnormalities were detected.

ASSESSMENT:

Based on the patient's history, physical examination, and laboratory and imaging findings, it is assessed that the patient is experiencing increased frequency of seizures due to the temporal lobe epilepsy. The investigational drug being used in the RCT may be contributing to the increased frequency of seizures, but this cannot be definitively determined at this time. The patient's cognitive function and memory appear to be mildly affected, which may be related to the seizures or the investigational drug.

PLAN:

The patient will continue to participate in the RCT and receive the investigational drug for the next 3 months. No changes will be made to his current medications, other than adding the investigational drug. The patient will be closely monitored for any changes in seizure frequency or severity, as well as for any potential adverse effects of the investigational drug.

SIGNATURE:

[Your Signature]

Date: 2022-02-25

Note: This clinic note is a sample and should not be used as a real clinic note. It is important to always use proper medical terminology and adhere to HIPAA regulations when writing clinical notes.

----------------END OF PATIENT ENCOUNTER---------------

-------------- PATIENT ID 170 -- VISIT 2 -----------

Clinic Note - Visit #2

Patient ID: 170

Date: 2022-03-25

Chief Complaint:

* Decreased seizure frequency since starting investigational drug ID X93461 (now complete)

* Somnolence

* Back pain

* Diplopia

History of Present Illness:

* 75-year-old male with history of autism, hypertension, and temporal lobe epilepsy

* Participated in a randomized controlled trial for ID X93461 for epilepsy over the past 3 months

* Reported 12 seizures during the study period

* Stopped taking investigational drug as study is now complete

Past Medical History:

* Autism

* Hypertension

* Temporal lobe epilepsy

* Previous history of seizures

Medications:

* Levetiracetam

* Lamotrigine

* Valproic acid

Social History:

* No alcohol or drug use

* Married with adult children

* Retired from his job as an accountant

Review of Systems:

* No fever, chills, or other constitutional symptoms

* No headache, nausea, or vomiting

* No recent changes in vision or hearing

* No recent travel or exposure to illness

Neurological Examination:

* Mental status: Alert and oriented x3

* Speech: Clear and fluent

* Motor exam: No signs of weakness, atrophy, or tremors

* Sensory exam: Intact sensation in all extremities

* Cranial nerve exam: No abnormal findings

* Reflexes: 2+ (moderate) throughout

* Gait: Steady and normal

Assessment and Plan:

* Based on the patient's history of autism, hypertension, and temporal lobe epilepsy, as well as his recent participation in a randomized controlled trial for ID X93461, it is likely that the patient's seizures are related to his underlying neurological conditions.

* Discontinuation of the investigational drug should help reduce the frequency and severity of seizures.

* The patient will continue on current medications (levetiracetam, lamotrigine, valproic acid) and be monitored closely for changes in seizure activity and side effects.

* Patient will follow up with clinic in 2-4 weeks to assess response to medication and discuss any changes to the treatment plan as needed.

* Patient will also have a telemedicine follow-up in 1 month to monitor for any changes in seizure activity or side effects.

Note: The information in this clinic note is fictional and does not represent any actual patient or study.

----------------END OF PATIENT ENCOUNTER---------------

-------------- PATIENT ID 171 -- VISIT 1 -----------

Patient ID: 171

Date of Visit: 2022-05-28

Chief Complaint: The patient presents with a history of generalized epilepsy and has experienced seven seizures over the past two months. She is currently enrolled in a randomized controlled trial (RCT) and will be taking the investigational drug for epilepsy for three months.

Neurological Examination:

On examination, the patient appears well-oriented and alert. Her speech is clear and concise, with no evident cognitive impairment. The patient's vital signs are within normal limits, with a blood pressure of 120/80 mmHg, a heart rate of 70 beats per minute, and a temperature of 98.6 degrees Fahrenheit.

The patient's neurological examination reveals no focal deficits or abnormalities in her sensory, motor, or cranial nerves. Her tone is normal, with no signs of rigidity or tremors. Her reflexes are brisk and normal, and her muscle strength is 5/5. The patient's coordination and balance are intact, and she is able to walk normally without any assistive devices.

The patient's past medical history includes a diagnosis of generalized epilepsy, for which she has been taking levetiracetam and lamotrigine. She has experienced seven seizures over the past two months, with no evidence of any recent seizure activity. The patient reports no significant changes in her medications or other medical conditions.

Plan:

The patient will continue to take her current medications, including levetiracetam and lamotrigine, as prescribed by her primary care physician. In addition, she will begin taking the investigational drug for epilepsy as part of the RCT. The patient will be closely monitored over the next three months to assess the efficacy and safety of the investigational drug. No changes in her medications or dosages are anticipated during this time period. The patient will be followed up with every two weeks to assess her seizure activity and any potential side effects from the medication.

----------------END OF PATIENT ENCOUNTER---------------

-------------- PATIENT ID 171 -- VISIT 2 -----------

---

Clinic Note #2 - Patient ID 171 - 28 year old female with generalized epilepsy - 2022-06-28

Identification:

Patient ID 171, a 28-year-old female, was seen in clinic today for a follow-up visit related to her history of generalized epilepsy and participation in a randomized controlled trial (RCT) evaluating the investigational drug ID X93461 for epilepsy.

HPI:

The patient presented with a history of generalized epilepsy, with a recent seizure occurrence on [date] at [time]. The patient reported somnolence, fatigue, and diplopia as adverse effects related to the investigational drug ID X93461, which was administered for the past 3 months.

Subjective:

The patient reported a decrease in seizure frequency during the RCT period, with only one seizure occurring during the past month. However, she also reported experiencing somnolence, fatigue, and diplopia as adverse effects related to the investigational drug. The patient expressed concern about the potential long-term effects of the drug and its impact on her quality of life.

Medications:

The patient has been compliant with her medication regimen, including levetiracetam (Keppra) and lacosamide (Vimpat), as prescribed. However, she will be discontinuing the investigational drug ID X93461 as part of the study protocol.

Allergies:

The patient has no known allergies to any medications or substances.

Social History:

The patient denies any significant social history, including smoking, alcohol consumption, or illicit drug use. She reports a supportive family environment and good social support network.

Review of Systems:

The patient's review of systems was unremarkable, with no complaints of respiratory, cardiovascular, gastrointestinal, or other systemic symptoms.

General Physical Examination:

On examination, the patient was alert and oriented, with a normal appearance and demeanor. Her vital signs were within normal limits, including temperature, blood pressure, and heart rate. Her neurological examination revealed no abnormalities, including no evidence of seizure activity or focal deficits. The patient's speech was clear and coherent, with no signs of dysarthria or aphasia. Her eye movements were full and without any evidence of nystagmus or strabismus.

Comprehensive Neurologic Exam:

The patient underwent a comprehensive neurological examination, including an evaluation of her cognitive function, memory, and executive function. The results of this examination were unremarkable, with no evidence of significant cognitive impairment or decline.

Labs:

The patient's laboratory results, including complete blood count (CBC), electrolyte panel, and liver function tests (LFTs), were within normal limits.

Imaging:

The patient's imaging studies, including magnetic resonance imaging (MRI) of the brain and computed tomography (CT) scans, were unremarkable, with no evidence of structural abnormalities or lesions.

Assessment:

Based on the patient's history, physical examination, and laboratory results, it is my assessment that the patient has a history of generalized epilepsy and has tolerated the investigational drug ID X93461 well over the past 3 months. However, the patient has reported somnolence, fatigue, and diplopia as adverse effects related to the drug, and these effects may be of concern for long-term use. Additionally, the patient's seizure frequency has decreased during the RCT period, but the long-term efficacy and safety of the drug are yet to be determined.

Plan:

Given the patient's history of generalized epilepsy and participation in the RCT, it is my plan to continue monitoring the patient's seizure activity and adverse effects related to the investigational drug. The patient will be discontinued from the study drug and her medication regimen will be adjusted as needed to manage her seizures effectively. Regular follow-up appointments will be scheduled to assess the patient's progress and address any concerns or issues that may arise.

Signature:

[Your signature]

Date:

2022-06-28

----------------END OF PATIENT ENCOUNTER---------------

-------------- PATIENT ID 172 -- VISIT 1 -----------

Clinic Note Visit #1

Patient ID: 172

Date: 2021-05-05

Identification:

Name: Agatha May Smith

Age: 97 years old

Gender: Female

Address: 123 Main St, Anytown, USA

Contact Information: (555) 123-4567

Chief Complaint:

Agatha presents with a history of arthritis, autism, seasonal allergies, and multifocal epilepsy. She has been experiencing an increase in seizure activity over the past two months, with 24 seizures reported. The patient is currently enrolled in a randomized controlled trial (RCT) for the treatment of her epilepsy.

Highlights of the Patient's History:

* Arthritis: Agatha has been diagnosed with osteoarthritis and rheumatoid arthritis, with chronic pain and stiffness in her joints.

* Autism: The patient has a history of autism, with difficulty communicating and interacting with others.

* Seasonal Allergies: Agatha experiences seasonal allergies, with symptoms including sneezing, congestion, and itchy eyes.

* Multifocal Epilepsy: The patient has a history of multifocal epilepsy, with seizures originating from multiple foci in the brain.

Subjective Complaints:

Agatha reports an increase in seizure activity over the past two months, with 24 seizures reported. She also complains of continued chronic pain and stiffness in her joints, as well as difficulty sleeping due to nighttime seizure activity. The patient mentions feeling "very tired" and experiencing "brain fog" intermittently throughout the day.

Medication Adherence:

Agatha reports taking her medications as prescribed, with no missed doses or alterations in medication schedule. She is currently taking levitiracetam, valproic acid, and lamotrigine for her epilepsy, as well as meloxicam for her arthritis pain.

Allergies:

The patient has no known allergies.

Social History:

Agatha lives with her daughter, who is her primary caregiver. She has a supportive social network and enjoys spending time with family and friends. The patient is active in her community and participates in local art classes.

Review of Systems:

Cardiovascular: No history of cardiovascular disease or risk factors.

Respiratory: No history of respiratory disease or risk factors.

Gastrointestinal: No history of gastrointestinal disease or risk factors.

Neurological: Patient has a history of multifocal epilepsy with seizures originating from multiple foci in the brain.

Musculoskeletal: Patient has osteoarthritis and rheumatoid arthritis, with chronic pain and stiffness in her joints.

Psychiatric: The patient has a history of autism, with difficulty communicating and interacting with others.

General Physical Examination:

Vital Signs:

* Temperature: 97.8°F (36.6°C)

* Blood Pressure: 120/80 mmHg

* Heart Rate: 75 beats per minute

* Respiratory Rate: 14 breaths per minute

Head:

* Normocephalic (normal head size)

* No deformities or lesions noted

Eyes:

* Full visual fields, with no signs of gaze evoked nystagmus or abnormal saccadic movements

* No evidence of papilledema or retinal hemorrhages

Ears:

* No signs of otitis media or external ear canal infections

Nose:

* No nasal polyps or masses noted

Mouth:

* Normal oral hygiene and no signs of oral lesions

Thorax:

* Normally proportioned chest with no deformities or lesions noted

Abdomen:

* Soft, normocephalic (normal size), and nontender abdomen

Extremities:

* No deformities or lesions noted in the upper or lower extremities

Comprehensive Neurological Examination:

Cranial Nerves:

* Visual fields normal

* No signs of gaze evoked nystagmus or abnormal saccadic movements

* No evidence of papilledema or retinal hemorrhages

* Facial expressions and smile intact

* No signs of ophthalmoplegia or abducens palsy

* Eye movements normal

* No signs of Horner's syndrome or other autonomic dysfunction

Motor Examination:

* Full range of motion in all joints, with no signs of muscle weakness or wasting

* Normal tone and reflexes

* No signs of ataxia or dysmetria

Sensory Examination:

* Intact sensation to light touch, pain, and temperature over all extremities

Labs:

* Complete Blood Count (CBC): Within normal limits

* Electrolyte Panel: Within normal limits

* Liver Function Tests (LFTs): Within normal limits

* Renal Function Tests (RFTs): Within normal limits

* Thyroid Function Tests (TFTs): Within normal limits

* Urinalysis: Within normal limits

Imaging:

* No recent imaging studies available for review

Assessment:

* Multifocal epilepsy with seizures originating from multiple foci in the brain

* Chronic pain and stiffness in joints due to osteoarthritis and rheumatoid arthritis

* History of autism, with difficulty communicating and interacting with others

* No changes in medications will be made other than adding the investigational drug for epilepsy, which will be taken for 3 months as part of the RCT.

Plan:

1. Continue current medications as prescribed, including levitiracetam, valproic acid, and lamotrigine.

2. Add the investigational drug for epilepsy, which will be taken for 3 months as part of the RCT.

3. Monitor seizure activity and adjust medications as needed.

4. Schedule follow-up appointments every 2 months to assess progress and make any necessary adjustments to the treatment plan.

Signature:

Agatha May Smith, MD

Date: 2021-05-05

----------------END OF PATIENT ENCOUNTER---------------

-------------- PATIENT ID 172 -- VISIT 2 -----------

CLINIC NOTE VISIT #2

Patient ID: 172

Date: 2021-06-05

Identification:

Age: 97 years

Gender: Female

History of Present Illness:

The patient, a 97-year-old female with a history of arthritis, autism, seasonal allergies, and multifocal epilepsy, presented for a routine follow-up visit. She has been participating in a randomized controlled trial (RCT) investigating the efficacy and safety of the investigational drug ID X93461 for the treatment of epilepsy. The patient has been taking the drug for the past three months and has experienced 22 seizures during this time period.

Highlights of Patient's Medical History:

* Arthritis (status post-total knee replacement)

* Autism

* Seasonal allergies

* Multifocal epilepsy (previously treated with levetiracetam and lamotrigine)

Subjective Complaints:

The patient reported experiencing dizziness, somnolence, balance disorder, dysarthria, and gait disturbance during the past three months. These symptoms have been ongoing since starting the investigational drug.

Medications:

* Levetiracetam (Keppra) 500mg BID

* Lamotrigine (Lamictal) 200mg OD

* ID X93461 (investigational drug) 500mg QD (discontinued as of today)

Allergies:

* No known allergies

Social History:

* Married with adult children and grandchildren

* Retired teacher

* Enjoys reading, gardening, and spending time with family

Review of Systems:

* General: Denies fever, chills, or recent illness. Reports ongoing dizziness and somnolence.

* Neurological: Denies numbness or tingling in the extremities. Reports balance disorder, dysarthria, and gait disturbance.

* Cardiovascular: Denies chest pain, shortness of breath, or palpitations.

* Respiratory: Denies shortness of breath or wheezing.

* Gastrointestinal: Denies abdominal pain, nausea, or vomiting.

General Physical Examination:

The patient was alert and oriented to person, place, and time. She had a normal appearance, with no signs of distress or discomfort. Her speech was slightly slurred, with noticeable dysarthria. Her gait was ataxic, with a wide-based, unsteady stance. The patient's balance was unstable, and she demonstrated difficulty walking without assistance.

Comprehensive Neurologic Examination:

The patient's neurological examination revealed the following findings:

* Cranial nerves: Intact, with no abnormalities noted.

* Motor function: The patient had ataxic movements and a wide-based, unsteady gait. Her muscle strength was normal.

* Sensory function: The patient had intact sensation to light touch, pain, and temperature. She reported no numbness or tingling in the extremities.

* Reflexes: Hypoactive, with a few brisk reflexes noted.

Labs:

* Complete Blood Count (CBC) within normal limits

* Basic Metabolic Panel (BMP) within normal limits

* Thyroid-stimulating hormone (TSH) level: 4.5 mIU/L (normal: 0.4-4.0 mIU/L)

Imaging:

* Magnetic Resonance Imaging (MRI) of the brain: No abnormalities noted.

Assessment:

The patient's symptoms of dizziness, somnolence, balance disorder, dysarthria, and gait disturbance are likely related to the investigational drug ID X93461. The drug has been associated with these side effects in previous studies. Given the patient's history of multifocal epilepsy and the recent onset of seizures while taking the drug, it is possible that the drug may be contributing to her seizure activity.

Plan:

1. Discontinue the investigational drug ID X93461 as of today.

2. Monitor the patient's symptoms closely and adjust medications as needed to manage her dizziness, somnolence, balance disorder, dysarthria, and gait disturbance.

3. Schedule a follow-up visit in four weeks to assess the patient's condition and review any changes in her symptoms or medications.

Signature:

[Your Name]

Neurologist

----------------END OF PATIENT ENCOUNTER---------------

-------------- PATIENT ID 173 -- VISIT 1 -----------

Clinic Note: Visit #1 (2022-12-20)

Patient ID: 173

Age: 95 years old

History of Present Illness:

The patient, a 95-year-old male, presents with a history of refractory parietal lobe epilepsy, having experienced 18 seizures in the past two months. He is currently participating in a randomized controlled trial (RCT) to assess the efficacy of an investigational drug for the treatment of epilepsy. The patient has been adherent to his current medication regimen and has not experienced any changes in his seizure frequency or severity.

Identification:

Name: John Doe

Date of Birth: January 1, 1927

Patient Number: 173

Location: Clinic 1

HPI:

The patient presents with a history of parietal lobe epilepsy, which has been refractory to previous medication regimens. He has experienced 18 seizures in the past two months, with no significant changes in frequency or severity. The patient is currently participating in an RCT evaluating the efficacy of an investigational drug for the treatment of epilepsy.

Subjective:

The patient reports no changes in his seizure frequency or severity since his last visit. He denies any new symptoms or complaints and reports a stable mood and cognitive function. The patient is generally healthy and active, with no significant medical comorbidities.

Medications:

Current medications include:

* Levetiracetam 2000mg PO QID

* Lamotrigine 200mg PO BID

* Valproic acid 500mg PO TID

* Investigational drug (X123456) 500mg PO BID (added to regimen for RCT)

Allergies:

The patient has no known allergies.

Social History:

The patient is a retired engineer and lives with his spouse. He denies any significant social or environmental stressors. The patient reports a stable living situation and adequate support from family members.

Review of Systems:

Neurological: The patient reports no new neurological symptoms, including no changes in seizure frequency or severity. He denies any new headaches, weakness, or numbness.

General: The patient reports no significant general complaints, including no changes in sleep quality, appetite, or energy level.

Comprehensive Neurologic Exam:

Mental Status: The patient is alert and oriented x3. He demonstrates normal cognitive function and responds appropriately to questions and commands.

Cranial Nerves: The patient's cranial nerve examination reveals no abnormalities, including normal visual fields, facial movements, and hearing sensitivity.

Motor Function: The patient demonstrates normal motor strength and tone, with no observable tremors or rigidity. He is able to perform a full range of motion without difficulty.

Sensory Function: The patient's sensory examination reveals no deficits in vision, hearing, touch, pain, or temperature perception.

Coordination and Gait: The patient demonstrates normal coordination and gait, with no observable ataxia or dysmetria.

Reflexes: The patient's reflexes are normal, including normal knee jerk and ankle jerk reflexes.

Labs:

Complete Blood Count (CBC) and basic metabolic panel (BMP) within normal limits.

Imaging:

Magnetic Resonance Imaging (MRI) of the brain with contrast reveals no evidence of acute or chronic changes in the brain parenchyma, including no signs of infarction, hemorrhage, or tumor.

Assessment:

The patient presents with a history of refractory parietal lobe epilepsy and has been participating in an RCT evaluating the efficacy of an investigational drug for the treatment of epilepsy. He reports no changes in his seizure frequency or severity since his last visit. The patient's comprehensive neurologic examination reveals no abnormalities, including normal cognitive function and motor strength. The patient's laboratory results are within normal limits, and imaging studies do not show any evidence of acute or chronic changes in the brain parenchyma. Based on the patient's history and current symptoms, it is appropriate to continue the current medication regimen and add the investigational drug for the treatment of epilepsy.

Plan:

1. Continue the current medication regimen, including levetiracetam, lamotrigine, valproic acid, and the investigational drug (X123456) 500mg PO BID.

2. Monitor the patient's seizure frequency and severity closely and report any changes to the prescribing physician.

3. Schedule follow-up visits every 2 months to assess the patient's response to therapy and monitor for any adverse effects of the medications.

4. Encourage the patient to maintain a consistent sleep schedule, avoid alcohol and drugs, and engage in regular exercise to optimize seizure control.

Signature:

[Your signature]

Date: [Today's date]

----------------END OF PATIENT ENCOUNTER---------------

-------------- PATIENT ID 173 -- VISIT 2 -----------

Clinic Note - Visit #2 (2023-01-20) for Patient ID 173, 95 year old male with history of parietal lobe epilepsy.

Today, I had the pleasure of examining Patient ID 173, a 95-year-old male who has been participating in a randomized controlled trial for the investigational drug ID X93461 for his parietal lobe epilepsy. Over the past three months, the patient has been taking this medication and has reported experiencing 18 seizures during this time. Additionally, the patient has reported feeling dizziness, somnolence, and fatigue since starting the medication.

On examination, the patient's neurological status was as follows: The patient's speech was clear and coherent, and his thought processes were logical and goal-directed. His motor function was intact, with no noticeable weakness or paresis. However, the patient did exhibit a mild dysmetria on the right side of his body, which is likely related to the seizures he has experienced. The patient's reflexes were normal, and his sensation was intact.

Given the patient's history of parietal lobe epilepsy and the significant number of seizures he has experienced while taking the investigational drug ID X93461, it is my recommendation that the medication be discontinued at this time. While the drug has shown promise in reducing seizure frequency, the potential risks associated with its use, including dizziness, somnolence, and fatigue, outweigh any potential benefits for this patient.

In conclusion, today's examination revealed a mild dysmetria on the right side of the patient's body, likely related to the seizures he has experienced while taking the investigational drug ID X93461. The medication will be discontinued given the patient's history of parietal lobe epilepsy and the significant number of seizures he has experienced while taking the drug. Further monitoring and management of the patient's condition will be necessary to ensure his continued health and well-being.

----------------END OF PATIENT ENCOUNTER---------------

-------------- PATIENT ID 174 -- VISIT 1 -----------

---

Clinic Note #1 - Patient ID 174

Date: 2021-08-07

Chief Complaint: Seizures

History of Present Illness: The patient, a 46-year-old male, presents with a history of seasonal allergies, ADHD, alcoholism, and epilepsy. He has experienced 7 seizures in the past 2 months. He is currently enrolled in a randomized controlled trial (RCT) and is taking no changes in medications except for adding the investigational drug for epilepsy which he will be taking for 3 months.

Review of Systems:

Allergies: Seasonal allergies

Medications: Current medications include:

* Levothyroxine (T4)

* Lisinopril

* Alprazolam (as needed)

* Clonazepam (as needed)

* Lamotrigine (for epilepsy)

Social History: The patient reports occasional alcohol use and denies any illicit drug use. He lives with his spouse and children.

Family History: No relevant medical history in the family.

Past Medical History: Seasonal allergies, ADHD, alcoholism, epilepsy

Current Medications: Levothyroxine (T4), Lisinopril, Alprazolam (as needed), Clonazepam (as needed), Lamotrigine (for epilepsy)

Allergies: Seasonal allergies

Social History: Occasional alcohol use, denies illicit drug use

Review of Systems: Negative for any relevant medical history

Neurological Examination:

Motor:

* Right and left arm and leg strength 5/5

* No tremors or rigidity noted

* Normal reflexes

Sensory:

* Normal sensation in all extremities

* No evidence of sensory loss or dysesthesias

Cerebellar Function:

* Heel-to-toe gait

* Normal finger-to-nose test

* No ataxia or dysmetria noted

Extraocular Movements:

* Full range of motion in all directions

Cranial Nerves:

* II-XII intact

Muscle Strength and Reflexes: Normal muscle strength and reflexes noted in all extremities.

Assessment and Plan:

The patient presents with a history of seasonal allergies, ADHD, alcoholism, and epilepsy. He has experienced 7 seizures in the past 2 months. The patient is currently enrolled in an RCT and taking no changes in medications except for adding the investigational drug for epilepsy which he will be taking for 3 months.

The neurological examination revealed normal motor strength, sensory function, cerebellar function, extraocular movements, and cranial nerves. The patient's muscle strength and reflexes were also normal in all extremities.

Plan:

1. Continue the current medications, including Lamotrigine for epilepsy, as prescribed by the RCT protocol.

2. Monitor the patient's seizure activity and adjust the medication regimen as needed.

3. Schedule follow-up appointments every 2 months to assess the patient's progress and adjust the medication regimen as needed.

4. Counsel the patient on the safe use of alcohol and the importance of adherence to the study protocol.

Note: The patient's history of seasonal allergies, ADHD, alcoholism, and epilepsy may impact his ability to participate in the RCT and may require additional monitoring and adjustments to his medication regimen. Additionally, the patient's history of seizures may be related to his epilepsy or other factors such as sleep deprivation or alcohol use. It is important to continue monitoring the patient's seizure activity and adjusting his medication regimen as needed to optimize seizure control.

----------------END OF PATIENT ENCOUNTER---------------

-------------- PATIENT ID 174 -- VISIT 2 -----------

Clinic Note #2 - Patient ID 174

Date: 2021-09-07

Chief Complaint: Balance disorder and seizures

History of Present Illness:

The patient, a 46-year-old male with a history of seasonal allergies, ADHD, alcoholism, and epilepsy, presents for a follow-up visit. He has been participating in a randomized controlled trial and has been taking the investigational drug ID X93461 for epilepsy over the past 3 months. During this time, he has experienced 9 seizures. The patient also reports balance disorder.

Past Medical History:

The patient has a history of seasonal allergies, ADHD, alcoholism, and epilepsy. He has been receiving treatment for these conditions and has had occasional seizures in the past.

Medications:

The patient is currently taking levetiracetam for his epilepsy and has been taking ID X93461 as part of the randomized controlled trial.

Social History:

The patient reports that he has been drinking alcohol on a regular basis and has smoked cigarettes in the past. He denies any illicit drug use.

Family History:

The patient's father had a history of epilepsy.

Review of Systems:

General: The patient reports no significant changes in his overall health.

Neurological: The patient reports balance disorder and recent seizures.

Musculoskeletal: The patient denies any musculoskeletal pain or weakness.

Psychiatric: The patient reports no significant changes in his mood or cognitive function.

General Physical Examination:

Vital Signs:

* Temperature: 98.6°F (37°C)

* Pulse: 70/min

* Blood Pressure: 120/80 mmHg

* Respiration Rate: 14/min

Head and Neck Examination:

* Head is normocephalic and atraumatic

* Eyes are normal, with full range of motion

* Ears are normal, with no discharge or infection

* Mouth and throat are normal, with no lesions or mass

Neurological Examination:

* Cranial Nerves: All grossly normal, with no deficits in vision, hearing, taste, or smell.

* Motor Examination: Normal strength and tone, with no ataxia or tremors.

* Sensory Examination: Full range of sensation in all extremities, with no deficits or loss of sensation.

* Coordination and Balance: The patient exhibits a balance disorder, with a history of recent seizures.

Assessment and Plan:

The patient's balance disorder and recent seizures are likely related to the investigational drug ID X93461, which will be discontinued now that the study is complete. The patient will be referred to physical therapy for balance rehabilitation and will be monitored closely for any changes in his neurological status. The patient's alcohol use and history of ADHD will also be addressed in his ongoing care.

Differential Diagnosis:

* Balance disorder

* Seizures

* Drug-related adverse effects

Plan:

1. Discontinue ID X93461

2. Refer to physical therapy for balance rehabilitation

3. Monitor closely for any changes in neurological status

4. Address alcohol use and ADHD in ongoing care

----------------END OF PATIENT ENCOUNTER---------------

-------------- PATIENT ID 175 -- VISIT 1 -----------

Clinic Note Visit #1

Date: 2021-09-18

Patient ID: 175

Name: John Doe

Age: 93 years

Gender: Male

Chief Complaint:

* Recent onset of seizures

* History of dermatitis, schizophrenia, and temporal lobe epilepsy

History of Present Illness (HPI):

* The patient has experienced 18 seizures over the past two months, with a frequency of approximately every other day.

* The patient reports a history of dermatitis, schizophrenia, and temporal lobe epilepsy.

* The patient is currently participating in a randomized controlled trial.

* No changes will be made to the patient's medications at this time, with the exception of adding an investigational drug for epilepsy.

Subjective:

* The patient reports feeling "a little off" and experiencing intermittent confusion.

* He denies any recent traumatic brain injuries or infections.

* He reports that his seizures are generally brief, lasting approximately 30 seconds to 1 minute, but he has noticed an increase in their frequency over the past two months.

* The patient's family members have reported worsening dementia-like symptoms, including confusion and disorientation.

Medications:

* Levetiracetam (Keppra) 500mg BID

* Lamotrigine (Lamictal) 200mg QD

* Valproic acid (Depakote) 500mg BID

* Investigational drug for epilepsy (placebo) added, to be taken for 3 months.

Allergies:

* None reported.

Social History:

* The patient is a retired accountant and lives with his wife of 65 years.

* He has two adult children and four grandchildren.

* He denies any recent travel or exposure to potential seizure triggers.

* The patient reports enjoying reading, gardening, and spending time with family.

Review of Systems:

* General: Denies fever, chills, or recent illness.

* Neurological: Reports intermittent confusion, dizziness, and seizures.

* Psychiatric: History of schizophrenia, but currently asymptomatic.

* Musculoskeletal: Denies joint pain or weakness.

* Cardiovascular: No history of heart disease or hypertension.

* Respiratory: Denies shortness of breath or cough.

* Gastrointestinal: Reports occasional constipation, but no recent changes.

General Physical Examination:

* Vital signs: Blood pressure 120/80 mmHg, heart rate 70 bpm, temperature 98.6°F (37°C), weight 75 kg.

* Head and neck: No masses or lesions, normal neck range of motion.

* Extremities: No weakness or paresthesias, normal muscle strength.

* Sensory: Normal sensation to light touch and pinprick in all extremities.

* Coordination and balance: Normal gait and station, no ataxia or dysmetria.

Comprehensive Neurological Examination:

* Mental Status: The patient is alert and oriented to person, place, and time. He denies any recent changes in cognitive function or mood.

* Cranial Nerves: Normal visual acuity and field, normal hearing sensitivity.

* Motor Exam: No muscle weakness or atrophy, normal tone and reflexes.

* Sensory Exam: Intact sensation to light touch and pinprick in all extremities.

* Coordination and Balance Exam: Normal gait and station, no ataxia or dysmetria.

Labs:

* Complete Blood Count (CBC): Within normal limits.

* Basic Metabolic Panel (BMP): Within normal limits.

* Thyroid-stimulating hormone (TSH): 4.5 mIU/L (normal 0.5-4.5 mIU/L).

* Vitamin B12 and Folate: Normal levels.

Imaging:

* Computed Tomography (CT) scan of the brain: No acute lesions or abnormalities detected.

* Magnetic Resonance Imaging (MRI) of the brain: No acute lesions or abnormalities detected.

Assessment:

* The patient presents with a history of dermatitis, schizophrenia, and temporal lobe epilepsy, as well as recent onset of seizures.

* He is currently participating in a randomized controlled trial and will be taking an investigational drug for epilepsy for the next 3 months.

* No changes will be made to the patient's medications at this time.

* The patient's seizures are generally brief, but there has been an increase in their frequency over the past two months.

* The patient reports intermittent confusion and dizziness.

* The patient has a history of schizophrenia, but is currently asymptomatic.

* The patient denies any recent travel or exposure to potential seizure triggers.

Plan:

* Continue the patient's current medications for epilepsy and psychiatric symptoms.

* Add the investigational drug for epilepsy and monitor the patient's response over the next 3 months.

* Recommend regular follow-up appointments to assess the patient's seizure frequency and overall health status.

Signature:

John Doe, MD

Neurologist

----------------END OF PATIENT ENCOUNTER---------------

-------------- PATIENT ID 175 -- VISIT 2 -----------

CLINIC NOTE #2

Patient ID: 175

Date: 2021-10-18

Chief Complaint: Dizziness and seizures

History of Present Illness:

The patient is a 93-year-old male with a history of dermatitis, schizophrenia, and temporal lobe epilepsy. He has been participating in a randomized controlled trial evaluating the investigational drug ID X93461 for the treatment of epilepsy. Over the past 3 months, the patient has experienced 9 seizures while taking the medication. The patient has also reported feeling dizzy.

Past Medical History:

The patient has a history of dermatitis, schizophrenia, and temporal lobe epilepsy. He has been treated for these conditions with various medications over the years.

Current Medications:

The patient is currently taking no medications other than the investigational drug ID X93461, which will be discontinued as part of the study.

Social History:

The patient lives at home and has a caregiver who assists with his daily activities. He denies any recent travel or exposure to illness.

Review of Systems:

The patient reports no significant changes in his symptoms since his last visit. He continues to experience occasional seizures, and he has been experiencing dizziness over the past few weeks.

Neurological Examination:

Mental Status: The patient is alert and oriented to person, place, and time. His speech is clear and his thought processes are logical.

Cranial Nerves:

* Visual acuity: 20/20 in both eyes

* Extraocular movements: Full range of motion in all directions

* Ocular motility: Full range of motion in all directions

* Facial expressions: Intact

* Tongue movements: Intact

Motor Examination:

* Strength: 5/5 in all muscle groups

* Reflexes: 2+ in all extremities

* Coordination and balance: Intact

Sensory Examination:

* Somatosensory: Full range of sensation in all extremities

* Dermatological: No notable changes in the patient's skin lesions

Assessment and Plan:

The patient's recent seizures and dizziness suggest a possible adverse effect of the investigational drug ID X93461. As part of the study, the medication will be discontinued, and the patient will be monitored closely for any changes in his symptoms. The patient's neurological examination is largely unchanged since his last visit, with the exception of a slight decline in cognitive function.

Plan:

1. Discontinue the investigational drug ID X93461 and monitor the patient for any changes in his symptoms.

2. Schedule follow-up appointments as needed to assess the patient's status.

3. Consider alternative medications or treatments for the patient's epilepsy and dizziness, as appropriate.

I hereby attest that this clinic note is an accurate representation of the patient's current condition and treatment plan.

Signature: ________________________________

Date: ________________________________

----------------END OF PATIENT ENCOUNTER---------------

-------------- PATIENT ID 176 -- VISIT 1 -----------

---

Patient: Patient ID 176, an 81-year-old female

Chief Complaint: Seasonal allergies and frontal lobe epilepsy with increased seizure frequency in the past 2 months (8 seizures)

History of Present Illness: The patient has a history of seasonal allergies and frontal lobe epilepsy. She has been experiencing an increased frequency of seizures over the past 2 months, with 8 seizures occurring despite no changes in her medications. The patient is currently enrolled in a randomized controlled trial for the investigational drug for epilepsy and will be taking this medication for the next 3 months.

Review of Systems:

* Allergies: Seasonal allergies are well-controlled with over-the-counter medications.

* Medications: The patient is currently taking levetiracetam, lamotrigine, and valproate for her epilepsy. She has not experienced any side effects or changes in medication effectiveness.

* Family History: No history of neurological disorders or seizures in the family.

Neurologic Examination:

* Mental Status: The patient is alert and oriented to person, place, and time. She has no evidence of cognitive impairment or changes in her mental status.

* Cranial Nerves: All cranial nerves are intact, with normal strength and reflexes.

* Motor Examination: The patient has normal strength and tone in all extremities. There is no evidence of ataxia or tremors.

* Sensory Examination: All sensory modalities are intact, with normal sensation to light touch, pain, and temperature.

* Coordination and Gait: The patient has a normal gait and there is no evidence of ataxia or abnormal coordination.

* Reflexes: All reflexes are normal.

Assessment and Plan:

* Assessment: The patient's seasonal allergies are well-controlled, and her frontal lobe epilepsy is stable. However, there has been an increase in seizure frequency over the past 2 months, which may be related to the ongoing randomized controlled trial.

* Plan: The patient will continue to take her current medications for epilepsy (levetiracetam, lamotrigine, and valproate) and will also begin taking the investigational drug for epilepsy as part of the randomized controlled trial. The patient will be closely monitored over the next 3 months to assess the effectiveness and safety of the investigational drug. Regular follow-up appointments will be scheduled to monitor the patient's seizure frequency, medication effectiveness, and any potential side effects of the investigational drug.

---

Note: The above clinic note is a sample and should not be used for actual clinical decision-making or as a basis for patient care.

----------------END OF PATIENT ENCOUNTER---------------

-------------- PATIENT ID 176 -- VISIT 2 -----------

Visit #2: July 6, 2022

Patient ID 176, an 81-year-old female, presented for her second visit in the randomized controlled trial. The patient has a history of seasonal allergies and frontal lobe epilepsy. Over the past three months, the patient has been taking the investigational drug ID X93461 for her epilepsy. Unfortunately, during this time, the patient has experienced 16 seizures despite being on the medication. Additionally, the patient reported experiencing diplopia, a condition characterized by seeing double.

On examination, the patient's neurological status was assessed. Her consciousness and alertness were normal, and she was able to communicate effectively. There were no signs of dysarthria or aphasia. The patient's motor function was intact, with no evidence of weakness, paralysis, or tremors. Her sensory examination revealed no deficits in hearing, vision, or sensation. However, the patient did exhibit diplopia when attempting to focus on objects.

The investigational drug ID X93461 will be discontinued now that the study is complete. The patient will be monitored closely for any changes in her condition, and alternative treatments will be explored to manage her epilepsy and diplopia. It is important to note that the patient's history of seasonal allergies may also be contributing to her symptoms, and appropriate steps will be taken to address these issues as well. The patient will be scheduled for follow-up visits to assess her progress and adjust her treatment plan as needed.

----------------END OF PATIENT ENCOUNTER---------------

-------------- PATIENT ID 177 -- VISIT 1 -----------

Clinic Note #1

Date: 2022-02-15

Patient ID: 177

Identification:

Name: Mary Johnson

Age: 90 years old

Gender: Female

Chief Complaint: Mary presents with a history of alcoholism and occipital lobe epilepsy, with a recent increase in seizure frequency.

History of Present Illness (HPI):

Mary has been experiencing an increased frequency of seizures over the past two months, with 9 episodes reported since January 2022. These seizures have been characterized as occipital lobe epilepsy, with Mary describing a sensation of "seeing stars" followed by loss of consciousness. She has also noted some confusion and disorientation following the seizures.

Subjective:

Mary reports no changes in her medications since her last clinic visit, although she has been adherent to her prescribed regimen. She denies any recent alcohol consumption. Mary expresses concern about the recent increase in seizures and its potential impact on her cognitive function.

Objective:

Mary's vital signs are within normal limits. Her neurological examination reveals no significant abnormalities, with the exception of a mild tremor in her hands. The remainder of her physical examination is unremarkable.

Laboratory Findings:

Mary's complete blood count (CBC) and electrolyte panels are within normal limits. Her serum alcohol level is undetectable, indicating that she has not consumed alcohol recently.

Imaging Findings:

Recent MRI scans of Mary's brain reveal no significant abnormalities or changes since her last scan.

Assessment:

Given Mary's history of occipital lobe epilepsy and recent increase in seizure frequency, it is possible that the investigational drug being used in the randomized controlled trial may be contributing to this change. However, without any changes in her medications, it is also possible that other factors may be contributing to the increased seizure activity. The mild tremor observed in Mary's hands may be a sign of underlying parkinsonism or other extrapyramidal disorders. Further evaluation and monitoring are necessary to determine the cause of Mary's seizures and to assess the efficacy of the investigational drug.

Plan:

1. Continue with current medications, including the investigational drug, for a period of 3 months.

2. Monitor Mary's seizure activity closely, with follow-up visits scheduled every 2 weeks.

3. Evaluate Mary's cognitive function and potential alcohol withdrawal symptoms during each follow-up visit.

4. Consider additional imaging studies or other diagnostic tests as needed to further evaluate the cause of Mary's seizures.

Signature:

[Your Name]

Attending Neurologist

Note: This clinic note is a fictional example and should not be used as a real clinical document. It is important to consult with a qualified healthcare professional before making any medical decisions or changes to your treatment plan.

----------------END OF PATIENT ENCOUNTER---------------

-------------- PATIENT ID 177 -- VISIT 2 -----------

Patient ID: 177

Date of Visit: 2022-03-15

Chief Complaint: Fatigue and seizures

History of Present Illness: The patient, a 90-year-old female with a history of alcoholism and occipital lobe epilepsy, has been participating in a randomized controlled trial for the investigational drug ID X93461 for the past three months. During this time, she has experienced three seizures and reported feeling fatigued.

Neurological Examination:

On examination, the patient's vital signs were normal, with a temperature of 98.6°F (37°C), blood pressure of 120/80 mmHg, and a pulse rate of 70 beats per minute. Her mental status was clear, and she was able to communicate effectively. The patient's motor function was normal, with no signs of weakness or paralysis. Her sensory examination revealed no deficits in vision, hearing, or touch.

The patient's neurological examination also revealed no abnormalities in her cranial nerves, including the visual and auditory systems. Her reflexes were normal, with no signs of hyporeflexia or hyperreflexia. The patient's gait was steady and normal, with no signs of ataxia or tremors.

Plan:

Based on the patient's history of alcoholism and occipital lobe epilepsy, as well as her recent experiences with seizures and fatigue during the randomized controlled trial, it is recommended that the patient discontinue taking the investigational drug ID X93461. The patient will be monitored closely for any changes in her condition, and appropriate medications will be prescribed to manage any residual symptoms. Regular follow-up appointments will be scheduled to assess the patient's progress and adjust the treatment plan as needed.

----------------END OF PATIENT ENCOUNTER---------------

-------------- PATIENT ID 178 -- VISIT 1 -----------

Here's the clinic note for Patient ID 178, an 82-year-old female with a history of parietal lobe epilepsy:

* Patient ID: 178

* Date: 2021-03-06

* History:

+ Parietal lobe epilepsy

+ 44 seizures in past 2 months

+ Randomized controlled trial (no change in medications except for investigational drug for epilepsy)

* Neurological Examination:

+ Mild weakness on right side of body

+ Sensory deficits in right arm and leg

+ No aura or postictal phenomena

+ Pertinent positives:

- Seizures are predominantly generalized tonic-clonic with some features of secondary generalization

- No significant cognitive changes

* Assessment:

+ Continue randomized controlled trial with investigational drug for epilepsy

+ Monitor seizure frequency and severity

+ Evaluate for potential side effects of investigational drug

* Plan:

+ Administer investigational drug for 3 months

+ Follow-up in 1 month to assess response and address any concerns or side effects

+ Reassess seizure frequency and severity after 3 months of treatment

----------------END OF PATIENT ENCOUNTER---------------

-------------- PATIENT ID 178 -- VISIT 2 -----------

Clinic Note: Visit #2 - Patient ID 178 - 82 year old female with parietal lobe epilepsy - Date: 2021-04-06

Chief Complaints:

* Recent history of seizure activity (13 seizures over the past 3 months)

* Dizziness

* Somnolence

* Fatigue

* Diplopia

History of Present Illness:

The patient was enrolled in a randomized controlled trial for the investigational drug ID X93461 for epilepsy. She has been taking the medication for the past 3 months and has experienced an increase in seizure activity during this time, with 13 seizures reported. The patient also reported dizziness, somnolence, fatigue, and diplopia.

Review of Systems:

The patient denies any recent changes in her mental status, speech, or language. She reports no recent traumatic events or infections. She does report some difficulty with balance and coordination, which she attributes to the dizziness and somnolence.

Physical Examination:

General: The patient appears somewhat disheveled and lethargic, with a slowed speech and deliberate movements. Her vital signs are within normal limits.

Neurological: The patient has a regular temporal relationship between her eye movements and the movement of her extremities. She does not have any significant motor weakness or atrophy. Her sensory examination reveals decreased sensation to light touch and pinprick in the right hemiparesis, consistent with previous diagnosis of parietal lobe epilepsy. The patient has no evidence of facial weakness or abnormalities.

Ocular: The patient has a subtle horizontal gaze palsy, with a mild esophoria. Her fundus examination reveals mild tortuosity of the retinal vessels, consistent with previous diagnosis of parietal lobe epilepsy.

Musculoskeletal: The patient has some mild rigidity in her muscles, particularly in her neck and shoulders. Her reflexes are brisk and symmetrical.

Assessment and Plan:

Given the patient's history of parietal lobe epilepsy and recent increase in seizure activity while taking the investigational drug ID X93461, it is recommended that the drug be discontinued. The patient will be referred to an epileptologist for further management of her seizures.

In the meantime, the patient will be monitored closely for any changes in her symptoms or seizure activity. Her medications will be adjusted as needed to control her seizures and manage any residual side effects.

The patient will also undergo regular follow-up appointments with the neurology clinic to assess her condition and make any necessary adjustments to her treatment plan.

Disposition: The patient will be discharged home with instructions to follow up with an epileptologist for further management of her seizures. She will also be prescribed a new medication regimen to control her seizures and manage any residual side effects.

----------------END OF PATIENT ENCOUNTER---------------

-------------- PATIENT ID 179 -- VISIT 1 -----------

Patient ID: 179

Age: 59 y/o

Chief Complaint: Increased seizure activity

History of Present Illness:

* Depression

* Occipital lobe epilepsy

* Randomized controlled trial (RCT)

* No change in medications except for investigational drug for epilepsy

Pertinent Positives:

* 113 seizures in past 2 months

* No change in medications

Neurological Examination:

* Mental status: Alert and oriented

* Cranial nerves: Normal

* Motor exam: Normal

* Sensory exam: Normal

* Reflexes: Normal

Assessment and Plan:

* Continue RCT medication as prescribed

* Monitor seizure activity and adjust plan as needed

* Schedule follow-up appointment in 3 months for reassessment.

----------------END OF PATIENT ENCOUNTER---------------

-------------- PATIENT ID 179 -- VISIT 2 -----------

On October 23rd, 2022, I conducted a routine follow-up visit with Patient ID 179, a 59-year-old male participant in a randomized controlled trial. The patient has a history of depression and occipital lobe epilepsy, for which he has been taking the investigational drug ID X93461 for the past three months. During this time, the patient has experienced 98 seizures, but has reported no symptoms.

Upon examination, the patient's neurological status was found to be stable. There were no signs of altered mental status, and his mood was euthymic. His speech was clear and intelligible, with no evidence of dysarthria or aphasia. His motor function was intact, with no weakness or paresis noted in any of the extremities. The patient's sensory examination revealed no deficits in sensation to light touch, pinprick, or vibration. His reflexes were normal and symmetrical.

The patient's ocular examination revealed no abnormalities in his visual acuity, extraocular movements, or funduscopic examination. His hearing examination was also within normal limits, with no evidence of conductive or sensorineural hearing loss. The patient's balance and coordination were intact, with no signs of ataxia or dizziness.

Given the completion of the randomized controlled trial, the investigational drug ID X93461 will be discontinued. The patient will be transitioned back to his previous medication regimen, and will be monitored closely for any changes in his seizure activity or neurological status. Overall, the patient remains in good health, with no significant changes noted during this follow-up visit.

----------------END OF PATIENT ENCOUNTER---------------

-------------- PATIENT ID 190 -- VISIT 1 -----------

Clinic Note #1

Date: 2022-06-19

Patient ID: 190

Identification:

I am pleased to present the following clinic note for Patient ID 190, a 77-year-old female who graced our clinic with her presence on 2022-06-19. This esteemed patient has been under our care for several months, and we have had the pleasure of witnessing her remarkable resilience and determination in the face of various health challenges.

History of Present Illness (HPI):

The patient presents with a history of dermatitis, diabetes, and parietal lobe epilepsy. She has experienced an unfortunate increase in seizure activity over the past two months, with a total of 10 seizures reported. This distressing development has undoubtedly had a profound impact on her quality of life, and we must acknowledge the considerable emotional toll that this ordeal has taken on both our patient and her loved ones.

Subjective:

During today's encounter, the patient reported experiencing frequent seizures, which have been characterized as absence seizures with simple partial secondary features. She also expressed concern about the potential risks of driving while experiencing these seizures, as well as the possibility of accidental ingestion of medications. Additionally, she revealed a heightened awareness of her surroundings and an increased sensitivity to light and sound, which she attributes to the ongoing seizure activity.

Medications:

The patient is currently prescribed the following medications:

* Levetiracetam (Keppra) 500 mg BID

* Lamotrigine (Lamictal) 100 mg QD

* Valproic acid (Depakote) 250 mg BID

Allergies:

The patient has reported no known allergies to any medications or substances.

Social History:

Our esteemed patient is a retired nurse who enjoys spending time with her family and engaging in needlework. She is currently participating in a randomized controlled trial (RCT) evaluating the efficacy of an investigational drug for epilepsy. The patient has been advised that no changes will be made to her current medication regimen during the RCT, with the exception of adding the investigational drug for a three-month period.

Review of Systems:

The patient's review of systems revealed no remarkable findings, other than the recent onset of seizure activity. She reported experiencing occasional dizziness and nausea, but these symptoms are not severe enough to interfere with her daily activities. The patient also mentioned that she has been experiencing some difficulty sleeping due to nighttime seizures.

General Physical Examination:

Upon general physical examination, the patient's vital signs were as follows:

* Temperature: 98.6°F (37°C)

* Blood Pressure: 120/80 mmHg

* Pulse: 75 beats per minute

* Respiratory Rate: 16 breaths per minute

Comprehensive Neurologic Examination:

The patient's neurological examination revealed the following findings:

* Cranial Nerves: All cranial nerves were intact, with the exception of mild weakness in the right arm.

* Motor Examination: The patient demonstrated a slight tremor in both hands and a mild weakness in the right arm.

* Sensory Examination: The patient exhibited decreased sensation to light touch and pinprick on the right side of her body, consistent with previous seizure activity.

* Coordination and Balance: The patient displayed a slight stagger while walking, but was able to maintain her balance without difficulty.

Labs:

The following laboratory tests were ordered and are currently within normal limits:

* Complete Blood Count (CBC)

* Comprehensive Metabolic Panel (CMP)

* Thyroid-Stimulating Hormone (TSH)

* Electrolyte Panel

* Urinalysis

Imaging:

The patient's recent imaging studies, including a computed tomography (CT) scan and magnetic resonance imaging (MRI), have revealed no remarkable findings consistent with seizure activity. The patient's previous MRI studies have shown evidence of cerebral atrophy and gliosis, which are consistent with her history of parietal lobe epilepsy.

Assessment:

In consideration of the patient's history of dermatitis, diabetes, and parietal lobe epilepsy, as well as her recent increase in seizure activity, we must acknowledge the complex interplay of factors that contribute to her overall health profile. The patient's participation in a randomized controlled trial further complicates her medical situation, as we must be mindful of the potential risks and benefits associated with the investigational drug being used in this study.

Plan:

In light of the above findings, we will continue to monitor the patient's seizure activity and adjust her medication regimen as necessary to control her symptoms. The patient will be advised to continue taking her current medications as prescribed, with the exception of adding the investigational drug for a three-month period during the RCT. Additionally, we will work closely with the research team to ensure that the patient's safety and well-being are protected throughout the duration of the study.

Signature:

---

Please note that this clinic note is fictional and does not reflect any actual patient or medical condition.

----------------END OF PATIENT ENCOUNTER---------------

-------------- PATIENT ID 190 -- VISIT 2 -----------

Patient ID: 190

Visit #2: 2022-07-19

HPI:

* 77 year old female

* History of dermatitis, diabetes, and parietal lobe epilepsy

* Currently participating in a randomized controlled trial for investigational drug ID X93461

Neurologic Examination:

* Pertinent positives only:

+ No symptoms reported

+ Seizures decreased from 21/month at baseline to 17/month over the past 3 months while taking investigational drug

Assessment and Plan:

* Discontinue investigational drug ID X93461 since study is complete

* Monitor seizure frequency and report any changes in future visits

* No additional interventions needed at this time.

----------------END OF PATIENT ENCOUNTER---------------

-------------- PATIENT ID 191 -- VISIT 1 -----------

Clinic Note: Visit #1

Patient ID: 191

Date: 2022-11-12

Identification:

Name: Mary Johnson

DOB: 03/01/1932

Height: 5'4"

Weight: 120 lbs

Reason for Visit: Follow-up appointment for ongoing management of depression and parietal lobe epilepsy.

History of Present Illness (HPI):

Mary Johnson, a 90-year-old female, presents with a history of depression and parietal lobe epilepsy. She has been experiencing recurrent seizures over the past two months, with approximately 15 episodes reported. Her symptoms include:

* Intermittent episodes of confusion, disorientation, and sudden loss of consciousness lasting between 30 seconds to 2 minutes.

* Auras: visual hallucinations (seeing colors and shapes) and sensory disturbances (hearing buzzing sounds).

* Postictal state: drowsiness, confusion, and headache for several hours after seizure activity.

Medication History:

* Currently prescribed: Levetiracetam 1000mg QID, Lamotrigine 200mg OND, Venlafaxine 75mg OD.

* Medication adherence: generally good, but some missed doses reported in the past week due to forgetfulness.

Allergies: None reported.

Social History:

* Married with three adult children and six grandchildren.

* Retired homemaker.

* Active member of local church community.

* Denies any recent significant life stressors or changes in social support network.

Review of Systems (ROS):

* General: No recent weight loss, fever, or other symptoms.

* Neurological: Continues to experience recurrent seizures as described above.

* Cardiovascular: Hypertension controlled with Lisinopril 20mg OD.

* Respiratory: No significant respiratory symptoms.

* Musculoskeletal: Mild arthritis in hands and knees, but no recent exacerbations.

General Physical Examination:

* Vital signs: Blood pressure 120/80 mmHg, heart rate 75 bpm, temperature 98.6°F (37°C), weight 120 lbs.

* Head and neck: Normocephalic, ataxic (slightly unsteady gait).

* Extremities: No significant abnormalities.

* Neurological: Presents with a clear sense of confusion and disorientation during the seizure activity, and mild aphasia postictally.

Comprehensive Neurologic Examination:

* Cranial nerves: Intact except for slight right gaze preference.

* Motor examination: No significant abnormalities.

* Sensory examination: No significant deficits.

* Coordination and balance: Mild ataxia noted during seizure activity.

Labs and Imaging:

* Complete Blood Count (CBC): WBC 4.8 x 10^9/L, Neutrophils 2.5 x 10^9/L, Lymphocytes 1.3 x 10^9/L, Hemoglobin 11.8 g/dL, Hematocrit 34%

* Electroencephalogram (EEG): Performed on 2022-11-08, showing frequent generalized spike-wave discharges consistent with parietal lobe epilepsy.

* Magnetic Resonance Imaging (MRI): Brain MRI performed on 2022-11-05, demonstrating a well-circumscribed lesion in the left posterior parietal lobe consistent with the diagnosis of mesial temporal lobe epilepsy.

Assessment:

* Parietal lobe epilepsy with recurrent seizures and mild cognitive decline.

* Depression (mild) with poor sleep quality.

* No significant changes in medication regimen at this time, except for the addition of the investigational drug for epilepsy as part of the randomized controlled trial.

Plan:

1. Continue current medications as prescribed, including Levetiracetam 1000mg QID, Lamotrigine 200mg OND, and Venlafaxine 75mg OD.

2. Add the investigational drug for epilepsy (Study Drug) at a dose of 500mg BID for a period of 3 months, as part of the randomized controlled trial.

3. Monitor seizure activity and adjust medication regimen as needed.

4. Re-evaluate medications and consider other therapeutic options if seizures persist or worsen despite current treatment.

5. Refer patient to neuropsychology for cognitive assessment and potential cognitive behavioral therapy (CBT) interventions to address mild depression and poor sleep quality.

Signature:

[Patient's name] has been informed of the risks and benefits of the investigational drug for epilepsy, and has given informed consent to participate in the randomized controlled trial.

[Your name and credentials]

----------------END OF PATIENT ENCOUNTER---------------

-------------- PATIENT ID 191 -- VISIT 2 -----------

Patient ID: 191

Date: 2022-12-12

HPI: 90yo F, history of depression and parietal lobe epilepsy, currently in randomized controlled trial (ID X93461 for epilepsy)

Neurological Examination:

* Mental status: Alert and oriented x3

* Cranial nerves: Normal

* Motor exam: Normal

* Sensory exam: Normal

* Coordination and balance: Normal

Pertinent Positives:

* Somnolence

* Constipation

Assessment and Plan:

* Discontinue investigational drug ID X93461 due to completion of study

* Monitor for seizure activity and adjust anticonvulsant medications as needed

* Address somnolence and constipation with appropriate medications and/or lifestyle modifications.

----------------END OF PATIENT ENCOUNTER---------------

-------------- PATIENT ID 192 -- VISIT 1 -----------

Clinic Note #1

Patient: Patient ID 192, 90 year old male

Chief Complaint: Recent increase in seizure activity

History of Present Illness: The patient is a 90-year-old male with a history of autism, depression, anxiety, and parietal lobe epilepsy. He has had three seizures in the past two months, which is an increase from his previous seizure activity. The patient is currently participating in a randomized controlled trial for the treatment of his epilepsy.

Past Medical History: The patient has a history of parietal lobe epilepsy and has been receiving treatment with levetiracetam and sodium valproate. He has also been diagnosed with autism, depression, and anxiety.

Medications: Levetiracetam 500mg twice daily, sodium valproate 500mg twice daily, and lamotrigine 200mg once daily.

Social History: The patient is a retired engineer and lives with his wife. He has no significant social or environmental stressors.

Family History: No significant neurological or psychiatric disorders in the patient's family.

Review of Systems: No other symptoms or complaints.

Neurological Examination:

Motor: The patient had a normal strength and tone in all four limbs. He was able to perform a full range of motion without any restrictions.

Sensory: The patient had a normal sensation in all extremities, including touch, pain, temperature, and vibration.

Cerebellar: The patient had a normal cerebellar function, with no signs of ataxia or dysmetria.

Extrapyramidal: The patient had a normal extrapyramidal function, with no signs of parkinsonism or dystonia.

Psychiatric: The patient was alert and oriented to person, place, and time. His mood was euthymic and his affect was appropriate. He denied any suicidal or homicidal ideation.

Assessment and Plan:

Assessment: The patient's recent increase in seizure activity is concerning, and it may be related to the ongoing randomized controlled trial. However, the patient's neurological examination was largely unremarkable, with no significant changes in his motor, sensory, or cerebellar function.

Plan: The patient will continue to participate in the randomized controlled trial and receive the investigational drug for epilepsy for the next three months. His medications will remain unchanged, except for the addition of the investigational drug. The patient will be closely monitored for any changes in his seizure activity or neurological function. Regular follow-up appointments will be scheduled to assess his progress and adjust the treatment plan as needed.

Special Instructions: The patient should continue to avoid any potential triggers for seizures, such as alcohol, caffeine, or lack of sleep. He should also report any changes in his symptoms or medication side effects promptly.

----------------END OF PATIENT ENCOUNTER---------------

-------------- PATIENT ID 192 -- VISIT 2 -----------

Clinic Note - Visit #2 (2021-08-19)

Patient ID: 192

Age: 90 years old

Gender: Male

Chief Complaint: Autism, depression, anxiety, and parietal lobe epilepsy

Background: The patient has been participating in a randomized controlled trial for the investigational drug ID X93461 for epilepsy over the past 3 months. During this time, the patient has experienced 15 seizures with no reported symptoms. The study is now complete and the investigational drug will be discontinued.

Neurological Examination:

The patient presents with a history of autism, depression, anxiety, and parietal lobe epilepsy. On examination, the patient appears alert and cooperative. The patient's speech is clear and fluent, with no evidence of dysarthria or apraxia of speech. The patient's cognitive function is intact, with no evidence of impairment in attention, memory, or executive functions.

The patient's sensorium is normal, with no evidence of hallucinations, delusions, or disorganized thinking. The patient's mood is euthymic, with no evidence of depression or anxiety. The patient's affect is appropriate to the situation, with no evidence of flat or inappropriate affect.

The patient's motor function is intact, with no evidence of weakness, tremor, ataxia, or dyskinesias. The patient's reflexes are normal, with no evidence of hyperreflexia or hyporeflexia. The patient's sensory examination reveals no deficits in vision, hearing, or tactile sensation.

The patient's neurological examination is unremarkable, with no evidence of focal deficits or abnormalities. The patient's epilepsy remains well-controlled on the current medication regimen. Based on the patient's history and examination findings, it is recommended that the investigational drug ID X93461 be discontinued, and the patient continue on their current medication regimen.

Recommendations:

1. Discontinue the investigational drug ID X93461.

2. Continue the patient's current medication regimen to control seizures.

3. Monitor the patient's seizure activity and adjust medication as needed.

4. Schedule a follow-up appointment in 3 months to reassess the patient's neurological function and seizure activity.

----------------END OF PATIENT ENCOUNTER---------------

-------------- PATIENT ID 193 -- VISIT 1 -----------

Patient ID: 193

Visit #1: 2022-10-29

History:

* Hypothyroidism

* Depression

* Parietal lobe epilepsy (15 seizures in past 2 months)

* Randomized controlled trial (no change in medications except for investigational drug for epilepsy)

Neurological Examination:

* Alert and oriented x3

* No focal deficits

* Normal muscle strength and tone

* Intact sensation (including pain, temperature, and vibratory sensation)

* Normal coordination and balance

* No dysdiadochokinesia or ataxia

Pertinent Positives:

* 15 seizures in past 2 months

* No change in medications except for investigational drug for epilepsy

Assessment and Plan:

* Continue randomized controlled trial with no change in medications except for investigational drug for epilepsy

* Monitor seizure activity and adjust plan as needed

* Schedule follow-up visit in 3 months to assess efficacy and safety of investigational drug.

----------------END OF PATIENT ENCOUNTER---------------

-------------- PATIENT ID 193 -- VISIT 2 -----------

Visit #2: 2022-11-29

Patient ID: 193

Age: 69 yo

Hx: Hypothyroidism, depression, parietal lobe epilepsy

Randomized controlled trial (RCT)

Investigational drug: ID X93461 (epilepsy)

Last 3 months:

* Took ID X93461

* Experienced 14 seizures

* Reported somnolence

Neurological examination:

* Mental status: Alert and oriented

* Speech: Normal

* Motor: No tremors or rigidity

* Sensation: Normal

* Reflexes: Normal

Pertinent positives:

* Seizures (14) in past 3 months

* Somnolence

Assessment and plan:

* Discontinue investigational drug ID X93461

* Monitor for seizure control and side effects

* Consider alternative treatments for epilepsy

* Schedule follow-up visit in 2 weeks.

----------------END OF PATIENT ENCOUNTER---------------

-------------- PATIENT ID 194 -- VISIT 1 -----------

Visit #1: November 13, 2021

Patient ID: 194

Chief Complaint: The patient presents with a history of anxiety, diabetes, hypertension, and temporal lobe epilepsy. She has experienced 12 seizures in the past two months.

History of Present Illness: The patient has been experiencing increased frequency and severity of seizures over the past two months. She has a history of well-controlled hypertension and diabetes, but her anxiety has been exacerbated by her seizure activity.

Past Medical History: The patient has a history of temporal lobe epilepsy and has been managing her condition with medication. She has been compliant with her medications and has not experienced any significant adverse effects.

Current Medications: The patient is currently taking levetiracetam 500mg twice daily, lamotrigine 200mg once daily, and gabapentin 300mg once daily. She has been instructed to continue her current medication regimen without any changes.

Neurological Examination:

* Mental Status: The patient is alert and oriented x3. She appears anxious and reports difficulty sleeping due to increased seizure activity.

* Cranial Nerve Examination: The patient's cranial nerve examination reveals no abnormalities.

* Motor Examination: The patient's motor examination reveals no signs of weakness, atrophy, or tremors.

* Sensory Examination: The patient's sensory examination reveals no abnormalities.

* Coordination and Balance Examination: The patient's coordination and balance examination reveals no signs of instability or dizziness.

Plan:

1. Continue the patient's current medication regimen without any changes.

2. Monitor the patient's seizure activity closely over the next three months.

3. Schedule a follow-up visit in two months to assess the patient's progress and make any necessary adjustments to her medication regimen.

4. Consider adding an investigational drug for epilepsy if the patient's seizure activity does not improve with current medications.

----------------END OF PATIENT ENCOUNTER---------------

-------------- PATIENT ID 194 -- VISIT 2 -----------

Vist #2, 2021-12-13

Patient ID: 194, Female, 39 yrs, anxiety, diabetes, hypertension, temporal lobe epilepsy

HPI:

* History of anxiety, diabetes, hypertension, and temporal lobe epilepsy

* Participated in a randomized controlled trial (RCT) for epilepsy (ID X93461) over past 3 months

* Experienced 8 seizures during the study

* Reported somnolence, ataxia, gait disturbance

Neurologic Examination:

* Vitals: BP 120/80 mmHg, HR 70 bpm, T 98.6°F (37°C)

* General: Alert and oriented x3, cooperative

* Cranial Nerves: Intact

* Motor: Mild ataxia, gait disturbance

* Sensory: Normal sensation

* Reflexes: 2+ (mild)

Pertinent Positives:

* Seizures have increased in frequency since starting ID X93461

* Somnolence, ataxia, and gait disturbance reported

Assessment:

* Discontinue ID X93461 due to seizure risk

* Monitor for any changes in seizure activity or adverse effects

Plan:

* Follow-up visit in 1 month to assess seizure activity and adverse effects

* Consider alternative treatments for epilepsy if necessary

* Provide education on seizure management and emergency preparedness.

----------------END OF PATIENT ENCOUNTER---------------

-------------- PATIENT ID 195 -- VISIT 1 -----------

Patient ID: 195

Date: 2021-11-21

HPI:

* 59-year-old male

* History of arthritis and occipital lobe epilepsy

* 16 seizures in past 2 months

* Currently in randomized controlled trial

Neurological Examination:

* Normal sensation and reflexes

* No weakness or atrophy noted

* No changes in cranial nerve examination

* No changes in coordination and balance

Pertinent Positives:

* No change in seizure frequency since last visit

* Patient reports continued difficulty with memory and concentration

Assessment and Plan:

* Continue randomized controlled trial medication as prescribed

* Add investigational drug for epilepsy for 3 months

* Monitor patient's seizure frequency and adverse effects of medications at follow-up visits

* Evaluate patient's memory and concentration at future visits.

----------------END OF PATIENT ENCOUNTER---------------

-------------- PATIENT ID 195 -- VISIT 2 -----------

---

Clinic Note #2 - Patient ID 195 - Date: 2021-12-21

Identification:

* Name: John Doe

* Age: 59 years old

* Gender: Male

* Patient ID: 195

Highlights of the History of Present Illness (HPI):

* Chief complaint: Seizures and dizziness

* History of arthritis and occipital lobe epilepsy

* Currently participating in a randomized controlled trial for investigational drug ID X93461 for epilepsy

* Has been taking the drug for 3 months and has experienced 27 seizures during that time

* Reports dizziness, fatigue, and gait disturbance

Subjective:

* Patient reports increased frequency of seizures over the past 3 months

* Describes dizziness as "a spinning feeling"

* Reports fatigue and difficulty walking due to gait disturbance

* Denies any other symptoms or concerns

Medications:

* ID X93461 (investigational drug for epilepsy) - discontinued as of today's visit

* Levetiracetam (keppra) - ongoing

* Meloxicam (mobic) - ongoing

Allergies:

* None reported

Social History:

* Married with two adult children

* Employed as an accountant

* No significant travel or exposure to known allergens

Review of Systems:

* General: Denies fever, chills, or weakness

* Neurological: Reports increased seizure activity and dizziness

* Musculoskeletal: Reports joint pain and stiffness consistent with arthritis

* Cardiovascular: Denies chest pain, shortness of breath, or palpitations

* Respiratory: Denies cough, wheezing, or shortness of breath

General Physical Examination:

* Vital signs: Blood pressure 120/80 mmHg, heart rate 70 bpm, temperature 98.6°F (37°C), weight 175 lbs

* Head and neck: Normal facial features, no hemiparesis or aphasia

* Extremities: No clubbing or cyanosis of the hands and feet

* Abdomen: Soft and nontender, no mass or organomegaly

* Neurological Examination: (See below for comprehensive neuro exam)

Comprehensive Neurologic Exam:

* Mental status: Alert and oriented x3

* Cranial nerves: Intact (visual, auditory, olfactory, gustatory, tactile)

* Motor function: No hemiparesis or weakness, normal tone and reflexes

* Sensory function: Normal sensation to light touch, pinprick, and temperature on all extremities

* Coordination and balance: Normal

* Gait and station: Normal

Labs:

* Complete Blood Count (CBC) - pending

* Electrolyte Panel - pending

* Liver Function Tests (LFTs) - pending

* Thyroid-stimulating hormone (TSH) - pending

Imaging:

* Magnetic Resonance Imaging (MRI) of the brain with and without contrast - normal findings

Assessment:

* Seizure activity continues to be a significant concern for this patient, particularly given the recent increase in frequency and the potential impact on his quality of life.

* The patient's history of arthritis and occipital lobe epilepsy suggest a possible etiology for his seizures, but further investigation is needed to confirm this relationship.

* The patient's current medications, including levetiracetam and meloxicam, may be contributing to his seizure activity and should be carefully monitored and adjusted as needed.

* The patient's recent participation in a randomized controlled trial for investigational drug ID X93461 may have influenced his seizure activity, and the discontinuation of this drug may have a positive impact on his condition.

Plan:

* Discontinue investigational drug ID X93461 as of today's visit

* Increase monitoring of current medications, including levetiracetam and meloxicam, to ensure proper dosing and minimize seizure activity

* Consider additional therapies or interventions as needed to manage seizure activity and improve quality of life, such as:

+ Vagus nerve stimulation (VNS)

+ Stereotactic surgery for drug-resistant epilepsy

+ Other anticonvulsant medications

* Schedule follow-up appointment in 2 weeks to reassess seizure activity and adjust treatment plan as needed.

Signature:

[Your signature here]

Date: 2021-12-21

----------------END OF PATIENT ENCOUNTER---------------

-------------- PATIENT ID 196 -- VISIT 1 -----------

Patient ID: 196

Visit #1 - 2023-01-01

History:

* Autism

* Occipital lobe epilepsy

* 4 seizures in past 2 months

* Randomized controlled trial (no change in medications)

Neurological Examination:

* Mental status: Alert and cooperative

* Speech: Clear and appropriate

* Motor exam: No abnormal movements or tremors

* Sensory exam: Normal sensation in all extremities

* Cranial nerves: Intact

* Reflexes: Normal

Pertinent Positives:

* No change in seizure activity since last visit

* Patient is compliant with trial medication

Assessment and Plan:

* Monitor seizure activity every 2 weeks

* Continue trial medication for 3 months

* Follow up visit on 2023-01-15 to reassess seizure activity and monitor trial medication tolerability.

----------------END OF PATIENT ENCOUNTER---------------

-------------- PATIENT ID 196 -- VISIT 2 -----------

Clinic Note Visit #2

Patient ID: 196

Date: 2023-02-01

Chief Complaint:

The patient presents with a history of autism and occipital lobe epilepsy. She has been participating in a randomized controlled trial for the investigational drug ID X93461 for the past 3 months. The patient reports experiencing somnolence and has had six seizures during this time.

Past Medical History:

The patient has a history of autism and occipital lobe epilepsy. She has been on levetiracetam and sodium valproate for seizure control.

Medications:

Levetiracetam 500mg BID

Sodium valproate 500mg TID

Social History:

The patient is a 63-year-old female who lives with her husband and adult child. She reports enjoying spending time with her family and watching TV.

Family History:

The patient's mother and grandfather had a history of seizures.

Review of Systems:

The patient reports no changes in her vision, hearing, or sleep patterns. She denies any recent infections or illnesses.

Neurological Examination:

General: The patient appears somnolent and has a flat affect. Her speech is slow and deliberate.

Cranial Nerves:

* Visual: The patient's visual acuity is normal, and her pupils are equally reactive to light. She has no abnormalities in her visual fields.

* Ocular Motility: The patient's ocular motility is full and without dysconjugacy or nystagmus.

* Facial: The patient's facial muscles are intact, and she has no weakness or paralysis.

* Auditory: The patient's hearing is normal, and she has no complaints of tinnitus or ear fullness.

* Optic: The patient's optic discs are normal, and there is no swelling or redness.

* Extremities: The patient's extremities are without atrophy or weakness.

Assessment:

The patient has a history of autism and occipital lobe epilepsy, and she has been participating in a randomized controlled trial for the investigational drug ID X93461. The patient reports somnolence and has had six seizures during this time. Based on the patient's neurological examination and medical history, it is recommended that the investigational drug be discontinued due to the risk of seizures.

Plan:

1. Discontinue the investigational drug ID X93461.

2. Monitor the patient's seizure activity closely and adjust her medications as needed.

3. Schedule a follow-up appointment in one month to reassess the patient's neurological status and adjust her treatment plan as necessary.

Note: The patient has been informed of the risks and benefits of the investigational drug and has given her informed consent to participate in the study. The patient has also been advised of the potential for seizures and has been instructed on how to manage them if they occur.

----------------END OF PATIENT ENCOUNTER---------------

-------------- PATIENT ID 197 -- VISIT 1 -----------

Patient ID: 197

Date: 2022-08-01

HPI:

* 50 year old male

* History of anxiety, autism, and multifocal epilepsy

* 29 seizures in past 2 months

* Randomized controlled trial (RCT)

Neurologic Examination:

* Alert and oriented x3

* No significant muscle weakness or atrophy

* Normal sensation in all extremities

* Intact reflexes

* Normal gait and station

Pertinent Positives:

* Seizures continue despite current medication regimen

* No change in medications other than adding investigational drug for epilepsy

* RCT is ongoing

Assessment and Plan:

* Monitor seizure activity and adjust medication regimen as needed

* Continue RCT and assess efficacy of investigational drug

* Schedule follow-up visit in 3 months to reassess seizure activity and medication regimen.

----------------END OF PATIENT ENCOUNTER---------------

-------------- PATIENT ID 197 -- VISIT 2 -----------

Visit #2 - September 1, 2022

Patient ID: 197

Age: 50 years old

History: Anxiety, Autism, Multifocal Epilepsy (randomized controlled trial)

Investigational Drug: ID X93461 (epilepsy) - discontinued due to study completion

Neurological Examination:

* Somnolence (reported by patient)

* Constipation (reported by patient)

* Back pain (reported by patient)

Pertinent Positives:

* 22 seizures over past 3 months while taking investigational drug

Assessment and Plan:

* Discontinue investigational drug due to study completion

* Monitor for withdrawal symptoms

* Discuss alternative therapies for anxiety, autism, and epilepsy with patient and primary care physician

* Schedule follow-up appointment in 2 weeks for reassessment.

----------------END OF PATIENT ENCOUNTER---------------

-------------- PATIENT ID 198 -- VISIT 1 -----------

On January 24th, 2022, Patient ID 198, a 20-year-old male, visited our clinic for his first visit. The patient presented with a history of asthma, hypertension, and parietal lobe epilepsy. He had experienced 8 seizures in the past two months, which was a significant increase from his previous seizure activity.

During the clinical examination, the patient's vital signs were within normal limits. His neurological examination revealed no evidence of focal deficits or abnormalities in sensation or reflexes. However, he did exhibit a mild tremor in his hands and feet, which is consistent with his known history of parietal lobe epilepsy. The patient's language and cognitive function were normal, and he was able to communicate effectively.

The patient was informed about the randomized controlled trial that he would be participating in, and he was aware of the potential risks and benefits associated with the investigational drug for epilepsy. He was advised that no changes would be made to his current medications, other than the addition of the investigational drug, which would be taken for a period of three months. The patient was also counseled on the importance of adherence to his medication regimen and the need to monitor any changes in his seizure activity.

Overall, the patient's clinical presentation and neurological examination revealed no significant changes since his last visit. However, his increased seizure activity was a cause for concern, and the patient was advised to continue with his current medications and to closely monitor any changes in his symptoms. The patient was scheduled for a follow-up visit in three months to assess his progress and evaluate any potential changes in his seizure activity.

----------------END OF PATIENT ENCOUNTER---------------

-------------- PATIENT ID 198 -- VISIT 2 -----------

Patient ID: 198

Visit #2: 2022-02-24

History: Asthma, hypertension, parietal lobe epilepsy

Investigational Drug: ID X93461 (discontinued)

Neurological Examination:

* Dizziness

* No changes in seizure activity or frequency

* Normal muscle strength and tone

* Intact sensation and reflexes

* Normal coordination and balance

Pertinent Positives:

* 15 seizures in past 3 months while taking investigational drug

* Dizziness reported by patient

Assessment and Plan:

* Discontinue investigational drug ID X93461

* Monitor seizure activity and dizziness closely

* Consider alternative treatments for epilepsy

* Follow-up visit in 1 month to assess status and adjust plan as needed.

----------------END OF PATIENT ENCOUNTER---------------

-------------- PATIENT ID 199 -- VISIT 1 -----------

On July 14, 2022, Patient ID 199, an 83-year-old female, presented to our clinic for her first visit. The patient has a history of parietal lobe epilepsy and has experienced 14 seizures in the past two months. She is currently participating in a randomized controlled trial (RCT) aimed at evaluating the efficacy of an investigational drug for epilepsy. As part of her RCT, the patient will be taking the investigational drug for three months.

Upon examination, the patient's neurological status was as follows: She had a normal gross motor function, with no signs of weakness or ataxia. Her fine motor skills were intact, and she was able to perform activities of daily living (ADLs) without any difficulties. Her sensory examination revealed a normal sensation in all modalities, including visual, auditory, and tactile. The patient had a normal cognitive function, with no signs of memory loss or confusion.

The patient's past medical history included parietal lobe epilepsy, for which she had been taking levetiracetam and lacosamide for the past three years. She had experienced 14 seizures in the past two months, with no significant changes in her medication regimen. Her family history was unremarkable, and there were no known allergies or sensitivities to any medications.

During the visit, the patient underwent a complete neurological examination, which revealed no significant abnormalities. Her speech was normal, and she had a normal language function. Her cranial nerve examination showed no signs of dysfunction, and her reflexes were normal. The patient's muscle strength and tone were within normal limits, and she had no signs of atrophy or weakness.

In conclusion, Patient ID 199 is an 83-year-old female with a history of parietal lobe epilepsy who presented to our clinic for her first visit. She has experienced 14 seizures in the past two months and is currently participating in an RCT evaluating the investigational drug for epilepsy. Her neurological examination revealed no significant abnormalities, and she will continue with her current medication regimen with the addition of the investigational drug for the next three months.

----------------END OF PATIENT ENCOUNTER---------------

-------------- PATIENT ID 199 -- VISIT 2 -----------

CLINIC NOTE #2

PATIENT ID: 199

DOB: 1938-08-14

SEX: F

CHIEF COMPLAINT: Follow-up visit for epilepsy and adverse effects of investigational drug ID X93461.

HISTORY OF PRESENT ILLNESS:

The patient, a 83-year-old female, presents with a history of parietal lobe epilepsy and has been participating in a randomized controlled trial for the investigational drug ID X93461 for the past 3 months. The patient has experienced 26 seizures during this time and has reported adverse effects such as dizziness, somnolence, constipation, balance disorder, and vertigo.

PAST MEDICAL HISTORY:

The patient has a history of parietal lobe epilepsy and has been on stable doses of levetiracetam and lacosamide for the past 5 years.

MEDICATIONS:

Levetiracetam 2000mg BID, Lacosamide 100mg BID

RECENT DEVELOPMENTS:

The patient has been receiving the investigational drug ID X93461 for the past 3 months as part of a randomized controlled trial. The study is now complete and the investigational drug will be discontinued.

EXAMINATION:

Neurological examination reveals:

* Alert and oriented x3

* No focal deficits or weakness

* Normal tone, no tremors or rigidity

* Normal reflexes

* No sensory deficits

* Bilateral optic disc edema (likely due to long-standing epilepsy)

ASSESSMENT:

The patient's history of parietal lobe epilepsy and recent participation in a randomized controlled trial for the investigational drug ID X93461 are consistent with her reported seizure activity and adverse effects. The patient's neurological examination reveals no significant changes since her last visit.

PLAN:

1. Discontinue the investigational drug ID X93461 as the study is now complete.

2. Monitor the patient's seizure activity and adverse effects closely in the coming months.

3. Continue stable doses of levetiracetam and lacosamide.

4. Follow-up appointment in 2 weeks to assess for any changes in the patient's condition.

DIAGNOSIS:

1. Parietal lobe epilepsy

2. Adverse effects of investigational drug ID X93461 (dizziness, somnolence, constipation, balance disorder, vertigo)

IMPRESSION:

The patient's history of parietal lobe epilepsy and recent participation in a randomized controlled trial for the investigational drug ID X93461 are consistent with her reported seizure activity and adverse effects. The patient's neurological examination reveals no significant changes since her last visit.

RECOMMENDATIONS:

1. Discontinue the investigational drug ID X93461 as the study is now complete.

2. Monitor the patient's seizure activity and adverse effects closely in the coming months.

3. Continue stable doses of levetiracetam and lacosamide.

4. Follow-up appointment in 2 weeks to assess for any changes in the patient's condition.

----------------END OF PATIENT ENCOUNTER---------------

-------------- PATIENT ID 210 -- VISIT 1 -----------

Patient ID 210, an 81-year-old male, presented to the clinic on September 26, 2021, for a routine follow-up visit. The patient has a history of multifocal epilepsy and has experienced three seizures in the past two months. Currently, the patient is participating in a randomized controlled trial (RCT) to evaluate the efficacy of an investigational drug for the treatment of epilepsy.

Upon examination, the patient was alert and oriented, with no signs of distress or discomfort. The patient's speech was clear and coherent, with no noticeable tremors or rigidity. The patient's cognitive function was intact, with no evidence of confusion or memory impairment. The patient's motor function was slightly affected, with a mild degree of ataxia and hypotonia noted on examination.

The patient's neurological examination revealed no significant changes since the last visit. The patient continues to experience occasional seizures, but there has been no marked improvement or worsening of symptoms. The patient's EEG results were unchanged, with persistent activity observed in the right frontal and temporal regions.

The patient will continue to participate in the RCT for the next three months, with no changes made to the patient's medication regimen other than the addition of the investigational drug. The patient will be closely monitored during this time to assess the efficacy of the investigational drug and any potential adverse effects. Regular follow-up appointments have been scheduled to ensure the patient's continued well-being and to evaluate the progress of the RCT.

----------------END OF PATIENT ENCOUNTER---------------

-------------- PATIENT ID 210 -- VISIT 2 -----------

Patient ID: 210

Visit #2: 2021-10-26

History:

* Multifocal epilepsy

* Randomized controlled trial (RCT) participant (investigational drug ID X93461)

Neurological Examination:

* Somnolence: present

* Diplopia: present

Pertinent Positives:

* Seizures: 8 in past 3 months

Assessment and Plan:

* Discontinue investigational drug ID X93461 due to study completion

* Monitor for seizure activity and side effects

* Consider alternative treatments for multifocal epilepsy.

----------------END OF PATIENT ENCOUNTER---------------

-------------- PATIENT ID 211 -- VISIT 1 -----------

---

Clinic Note #1 (2023-02-10)

Patient ID: 211

Age: 87 years old

Gender: Male

Chief Complaint: Generalized epilepsy with 22 seizures in the past 2 months.

History of Present Illness (HPI): The patient presents with a history of generalized epilepsy, which has been poorly controlled over the past two months. He has experienced 22 seizures during this time, with no significant changes in his medication regimen. The patient is currently enrolled in a randomized controlled trial (RCT) investigating the efficacy and safety of an investigational drug for epilepsy.

Subjective:

The patient reports experiencing frequent seizures over the past two months, with no significant improvements in his symptoms despite continued treatment with standard anticonvulsant medications. He has noticed some improvement in his condition since enrolling in the RCT, but notes that his seizures have not been fully controlled. The patient also reports experiencing some cognitive difficulties, including memory loss and confusion, which he attributes to the seizures.

Objective:

On examination, the patient's vital signs are within normal limits. His general physical examination reveals no significant abnormalities. However, a comprehensive neurological examination reveals the following findings:

* Cranial nerve examination: The patient's pupils are equal and reactive to light, with no signs of afferent pupillary defect. His extraocular movements are full and without abnormalities. His facial expressions are normal, with no signs of weakness or paresis.

* Motor examination: The patient's muscle strength is normal, with no signs of atrophy or weakness. His tone is slightly increased, particularly in his upper limbs. His reflexes are normal, with no signs of hyporeflexia or hyperreflexia.

* Sensory examination: The patient's sensation is intact, with no signs of numbness or paresthesias. His proprioception and coordination are normal, with no signs of ataxia or dysmetria.

* Cerebellar function examination: The patient's cerebellar functions are within normal limits, with no signs of ataxia or dysmetria.

Laboratory Findings:

The patient's laboratory results are as follows:

* Complete Blood Count (CBC): White blood cell count (WBC) 4.8 x 10^9/L, hemoglobin (Hb) 13.2 g/dL, hematocrit (Hct) 40.5%, platelet count (Plt) 400 x 10^9/L.

* Electrolytes: Sodium 138 mmol/L, potassium 4.2 mmol/L, chloride 98 mmol/L, carbon dioxide 24 mmol/L.

* Renal Function Tests: Creatinine 0.87 mg/dL, urea 24 mg/dL.

* Liver Function Tests: Aspartate transaminase (AST) 19 U/L, alanine transaminase (ALT) 22 U/L, gamma-glutamyl transferase (GGT) 13 U/L, albumin 4.2 mg/dL.

Imaging Findings:

The patient's imaging results are as follows:

* Computed Tomography (CT) scan of the brain: No significant abnormalities were noted.

* Magnetic Resonance Imaging (MRI) of the brain: No significant abnormalities were noted.

Assessment:

The patient's history of poorly controlled generalized epilepsy, coupled with his recent experience of 22 seizures in the past two months, suggests a complex and refractory condition. The patient's participation in a randomized controlled trial (RCT) investigating an investigational drug for epilepsy is a significant factor in his current management. Given the patient's history of poor medication adherence and the lack of significant improvements in his symptoms despite continued treatment with standard anticonvulsant medications, it is likely that the patient's seizures are not fully controlled by these medications.

Plan:

1. Continue the patient's current medication regimen, including his standard anticonvulsant medications and the investigational drug being tested in the RCT.

2. Monitor the patient's seizure activity and adjust his medication regimen as needed to achieve better seizure control.

3. Recommend close follow-up with the study team to ensure compliance with the RCT protocol and to monitor the patient's response to the investigational drug.

4. Consider other treatment options, such as surgical interventions or alternative medications, if the patient's seizures remain poorly controlled despite continued treatment with standard anticonvulsant medications and the investigational drug.

Signature:

[Your signature here]

Date: 2023-02-10

Time: [Your time here]

----------------END OF PATIENT ENCOUNTER---------------

-------------- PATIENT ID 211 -- VISIT 2 -----------

CLINIC NOTE DATE: 2023-03-10

PATIENT ID: 211

AGE: 87 years old

SEX: Male

HISTORY OF PRESENT ILLNESS:

The patient is an 87-year-old male with a history of generalized epilepsy. He has been participating in a randomized controlled trial (RCT) for the investigational drug ID X93461 for the treatment of his epilepsy. The patient has been taking the medication for the past 3 months and has experienced 16 seizures during this time. The patient has also reported dizziness, somnolence, and vertigo.

REASON FOR VISIT:

The patient is presenting for a follow-up visit to assess the efficacy of the investigational drug and to address the patient's current symptoms.

PAST MEDICAL HISTORY:

The patient has a history of generalized epilepsy and has been experiencing seizures for the past 20 years. He has tried several different medications, including levetiracetam, carbamazepine, and valproic acid, but has not achieved complete seizure control.

MEDICATIONS:

The patient is currently taking the investigational drug ID X93461 as part of the RCT. He has also been taking levetiracetam and carbamazepine in the past to control his seizures.

SOCIAL HISTORY:

The patient lives alone and has a caregiver who assists him with daily activities. He reports no significant social or environmental stressors.

PHYSICAL EXAMINATION:

General: The patient appears generally well-oriented and alert, but appears slightly disheveled.

Neurological:

* Mental status: The patient's mental status is normal, with no evidence of cognitive impairment or clouding.

* Cranial nerves: The patient's cranial nerves are intact, with no abnormal findings.

* Motor: The patient has a normal strength and tone, with no significant tremors or rigidity.

* Sensory: The patient has a normal sensation in all extremities, with no evidence of numbness or tingling.

* Coordination and balance: The patient has a normal coordination and balance, with no evidence of ataxia or dysmetria.

ASSSESSMENT AND PLAN:

Assessment:

* The patient's history of generalized epilepsy and the recent seizures experienced during the RCT suggest that he may be a suitable candidate for continued treatment with ID X93461 or another anticonvulsant medication.

* The patient's current symptoms, including dizziness, somnolence, and vertigo, may be related to the investigational drug and may improve after discontinuation of the medication.

Plan:

* Discontinue the investigational drug ID X93461 as part of the RCT.

* Consider initiating a different anticonvulsant medication to control the patient's seizures, such as levetiracetam or carbamazepine.

* Monitor the patient's seizure activity and symptoms closely and adjust the treatment plan as needed.

* Schedule follow-up appointments as necessary to monitor the patient's condition and adjust the treatment plan.

----------------END OF PATIENT ENCOUNTER---------------

-------------- PATIENT ID 212 -- VISIT 1 -----------

Patient ID: 212

Visit #1: 2021-05-01

History:

* Bipolar disorder

* Alcoholism

* Dermatitis

* Occipital lobe epilepsy

* 52 seizures in past 2 months

* Randomized controlled trial (RCT)

Neurological Examination:

* Mental status: Alert and oriented

* Speech: Clear and appropriate

* Motor exam: No significant tremors or rigidity

* Sensory exam: Intact sensation in all extremities

* Reflexes: 2+ (mild)

* Coordination and balance: Normal

Pertinent Positives:

* No change in medications except for investigational drug for epilepsy

* Randomized controlled trial (RCT)

Assessment and Plan:

* Seizure frequency: 52 seizures in past 2 months

* Neurological examination: Normal

* Assessment: Patient is in a randomized controlled trial, no change in medications except for investigational drug for epilepsy.

* Plan: Continue investigational drug for epilepsy for 3 months. Monitor seizure frequency and neurological status closely. Follow up in 2 weeks.

----------------END OF PATIENT ENCOUNTER---------------

-------------- PATIENT ID 212 -- VISIT 2 -----------

Neurology Clinic Note - Visit #2 (2021-06-01)

Patient Information:

Name: Patient ID 212

Age: 77 years old

Gender: Female

History of Present Illness:

The patient is a 77-year-old female with a history of bipolar disorder, alcoholism, dermatitis, and occipital lobe epilepsy. She has been participating in a randomized controlled trial (RCT) for the investigational drug ID X93461 for epilepsy over the past 3 months. During this time, she has experienced 48 seizures and reported dizziness and fatigue. The RCT is now complete, and the investigational drug will be discontinued.

Past Medical History:

* Bipolar disorder

* Alcoholism

* Dermatitis

* Occipital lobe epilepsy

Medications:

* Levetiracetam (Keppra)

* Lamotrigine (Lamictal)

* Valproic acid (Depakote)

Social History:

* Smoking: Negative

* Alcohol consumption: Negative

* Illicit drug use: Negative

Review of Systems:

* None

Neurological Examination:

* Mental status: Alert and oriented to person, place, and time

* Speech: Clear and coherent

* Language: Intact

* Motor function: Intact, including tone, strength, and coordination

* Sensory examination: Intact, including visual, auditory, and tactile

* Reflexes: Normal

* Cranial nerves: Normal

* Gait and station: Normal

Assessment:

* The patient has a history of bipolar disorder, alcoholism, dermatitis, and occipital lobe epilepsy.

* She has been experiencing an increased frequency of seizures during the past 3 months while participating in a randomized controlled trial for the investigational drug ID X93461.

* The patient reported dizziness and fatigue.

Plan:

* Discontinue the investigational drug ID X93461.

* Monitor the patient's seizure frequency and adjust medications as needed.

* Consider referral to a psychiatrist for management of bipolar disorder.

* Follow up with the patient in 2 weeks to assess her condition and adjust the treatment plan as necessary.

Special Instructions:

* The patient should be advised to avoid any activities that may trigger seizures, such as driving or swimming alone.

* She should also be counseled on the potential risks and benefits of discontinuing the investigational drug and the need for close follow-up.

----------------END OF PATIENT ENCOUNTER---------------

-------------- PATIENT ID 213 -- VISIT 1 -----------

Visit #1: 2022-06-15

Patient ID: 213

Age: 43 years

Chief Complaint: Frontal lobe epilepsy with increased seizure frequency in the past 2 months.

History of Present Illness: The patient, a 43-year-old male, has been experiencing an increase in seizure frequency over the past two months, with a total of 34 seizures during this time. He is currently participating in a randomized controlled trial and will be taking the investigational drug for epilepsy for the next three months. No changes will be made to his current medications.

Neurological Examination:

The patient's neurological examination revealed the following:

* Mental status: Alert and oriented x3

* Head: Normocephalic, atraumatic

* Eyes: No visual disturbances or abnormalities

* Facial movements: Intact

* Motor function: No weakness or paresis, 5/5 strength in all muscle groups

* Sensory examination: Intact sensation to light touch, pinprick, and temperature over all body regions

* Coordination and balance: Normal

* Reflexes: Normocircadian reflexes

Assessment: The patient presents with a history of frontal lobe epilepsy and an increase in seizure frequency over the past two months. He is currently participating in a randomized controlled trial, taking the investigational drug for epilepsy. No changes will be made to his current medications. The neurological examination reveals no significant abnormalities.

Plan:

1. Continue with the current medication regimen and add the investigational drug for epilepsy.

2. Monitor the patient's seizure frequency and adjust the medication regimen as needed.

3. Schedule follow-up appointments every 2 weeks to assess the patient's progress and make any necessary adjustments to the medication regimen.

----------------END OF PATIENT ENCOUNTER---------------

-------------- PATIENT ID 213 -- VISIT 2 -----------

Patient ID 213, 43 y/o male

History: Frontal lobe epilepsy

Investigational drug ID X93461 (3 mos)

Neurologic Exam:

* Dizziness

* Fatigue

* Vomiting

* Upper respiratory infection

* Diplopia

Assessment:

* Discontinue investigational drug (study complete)

* Monitor for seizure activity and medication side effects

Plan:

* Follow-up visit in 1 month to assess for seizure control and medication side effects.

----------------END OF PATIENT ENCOUNTER---------------

-------------- PATIENT ID 214 -- VISIT 1 -----------

Patient ID 214, a 79-year-old female, presented to the clinic on November 5th, 2021, for her first visit in this randomized controlled trial. The patient has a history of hypertension and frontal lobe epilepsy, with a recent increase in seizure activity (10 seizures in the past two months). Her previous medications have remained unchanged, with the exception of adding an investigational drug for epilepsy that will be taken for three months.

Upon examination, the patient's neurological status was as follows:

* Mental status: Alert and oriented to person, place, and time. No evidence of clouding or confusion.

* Speech: Clear and coherent, with normal tone and volume.

* Motor function: No gross motor deficits or ataxia noted. Patient was able to perform a full range of movements without difficulty.

* Sensory function: Normal sensation in all extremities, with no reported numbness or tingling.

* Cranial nerves: II-XII were normal, with no abnormalities detected.

* Reflexes: 2+ (brisk) bilaterally, with normal reactivity to stretch.

* Muscle strength: 5/5 in all extremities, with no evidence of weakness or atrophy.

The patient's seizure activity remains a concern, and we will continue to monitor her closely throughout the trial. The investigational drug is expected to help control her seizures, but it may take some time to see the full effects. We will also be watching for any potential side effects or interactions with her existing medications. Overall, the patient is in good health and spirits, and we are confident that this trial will provide valuable insights into the management of frontal lobe epilepsy.

----------------END OF PATIENT ENCOUNTER---------------

-------------- PATIENT ID 214 -- VISIT 2 -----------

Here's the clinic note for Patient ID 214, a 79-year-old female with a history of hypertension and frontal lobe epilepsy:

Date: December 5, 2021

Patient Identification: 214

Subjective: The patient presented to the clinic on December 5, 2021, for her second visit. She reported experiencing three seizures over the past three months while taking investigational drug ID X93461 as part of a randomized controlled trial (RCT). The patient also complained of dizziness and nausea during that time. The patient has been adherent to her medications, including levetiracetam 500mg twice daily and lisinopril 20mg once daily for hypertension. She has no allergies.

History of Present Illness (HPI): The patient was referred to the clinic for an assessment of her epilepsy after participating in an RCT of ID X93461. The study required her to take the drug for three months, during which time she experienced three seizures.

Medications: The patient is currently taking levetiracetam 500mg twice daily and lisinopril 20mg once daily. She has been adherent to her medications.

Allergies: None reported.

Social History: The patient lives alone, has a support system in place, and is able to manage her daily activities. She reports no significant changes in her social or work life since her last visit.

Review of Systems (ROS): The patient reports no significant changes in her ROS, including no new medications, supplements, or changes in her sleep or appetite patterns.

General Physical Examination: On today's exam, the patient's vital signs were as follows: blood pressure 120/80mmHg, heart rate 70 beats per minute, temperature 98.6 degrees Fahrenheit (oral), and weight 55kg. Her general physical examination revealed no significant abnormalities.

Comprehensive Neurological Examination: The patient's comprehensive neurological examination revealed the following:

* Mental Status: The patient was alert and oriented x3, with normal speech and thought processes.

* Cranial Nerves: The patient had no abnormalities in her cranial nerves.

* Motor Examination: The patient had no signs of weakness, ataxia, or tremors.

* Sensory Examination: The patient had no signs of sensory deficits.

* Coordination and Balance: The patient had normal coordination and balance.

Labs: The patient's recent lab results included the following:

* Complete Blood Count (CBC): White blood cell count 4.5 x10^9/L, hemoglobin 12.3g/dL, hematocrit 36.1%

* Electrolytes: Sodium 138mmol/L, potassium 4.2mmol/L, chloride 104mmol/L

* Renal and Hepatic Function Tests: Creatinine 0.87mg/dL, urea 27.6mmol/L, aspartate transaminase (AST) 15U/L, alanine transaminase (ALT) 13U/L, gamma-glutamyl transferase (GGT) 48U/L

Imaging: The patient's recent imaging results included the following:

* Computed Tomography (CT) scan of the brain without contrast: No significant abnormalities were noted.

Assessment: The patient presents with a history of frontal lobe epilepsy and hypertension, and has been participating in an RCT of ID X93461 for her epilepsy. She experienced three seizures during the study period and reported dizziness and nausea. The patient's comprehensive neurological examination did not reveal any significant abnormalities. The patient's recent lab results and imaging studies were unremarkable. Based on her history of frontal lobe epilepsy, it is possible that the seizures she experienced during the study may be related to her condition. However, given the short duration of the RCT and the limited number of seizures, it is difficult to draw definitive conclusions about the cause of her seizures.

Plan: The patient will discontinue taking ID X93461 as part of the study's completion. Her levetiracetam and lisinopril prescriptions will remain unchanged. The patient will be monitored closely for any changes in her condition. Follow-up appointments will be scheduled as needed.

Signature: [Your Signature]

Date: December 5, 2021

Note: This clinic note is an example and should not be used as a real clinical document without proper authorization and patient consent.

----------------END OF PATIENT ENCOUNTER---------------

-------------- PATIENT ID 215 -- VISIT 1 -----------

Patient ID: 215

Date: 2023-02-02

History:

* Bipolar disorder

* Alcoholism

* Generalized epilepsy (15 seizures in past 2 months)

* Randomized controlled trial (no change in medications except for investigational drug for epilepsy)

Neurological Examination:

* Mental status: Alert and oriented

* Cranial nerves: Normal

* Motor exam: No abnormalities

* Sensory exam: Normal

* Reflexes: Normal

Pertinent Positives:

* 15 seizures in past 2 months

* No change in medications except for investigational drug for epilepsy

Assessment and Plan:

* Continue randomized controlled trial with no change in medications except for investigational drug for epilepsy

* Monitor seizure activity and adjust plan as needed

* Schedule follow-up visit in 3 months to assess efficacy of investigational drug and monitor for any adverse effects.

----------------END OF PATIENT ENCOUNTER---------------

-------------- PATIENT ID 215 -- VISIT 2 -----------

Visit #2: March 2, 2023

Patient ID 215, a 34-year-old female with a history of bipolar disorder, alcoholism, and generalized epilepsy, was seen in clinic for her second visit as part of a randomized controlled trial. Over the past three months, the patient has been taking the investigational drug ID X93461 for her epilepsy, and during this time she has experienced 19 seizures.

On examination, the patient appeared somnolent and fatigued, with a noticeable gait disturbance. Her speech was slow and slurred, and her eyes were droopy. The patient's neck was rigid, and her arms and legs were limp. There was no evidence of tremors or convulsions.

The patient's neurological examination revealed a few notable findings. Her muscle tone was decreased, and her reflexes were diminished. She had difficulty with rapid alternating movements, and her coordination and balance were compromised. However, there were no signs of sensory deficits or cognitive impairment.

In light of the patient's experience with ID X93461, it has been decided to discontinue the investigational drug now that the study is complete. The patient will be transitioned back to her previous medication regimen and will be closely monitored for any changes in her seizure activity or neurological status. Further follow-up visits will be scheduled as needed to assess the patient's ongoing care and treatment.

----------------END OF PATIENT ENCOUNTER---------------

-------------- PATIENT ID 216 -- VISIT 1 -----------

Clinic Note: Visit #1

Patient ID: 216

Date: 2022-08-04

Identification:

Name: Mary Johnson

DOB: 1935-07-23

Age: 86 years

Gender: Female

Address: 123 Main St, Anytown, USA

HPI:

Mary Johnson, an 86-year-old female patient, presents with a history of hypothyroidism and generalized epilepsy. She has experienced 15 seizures in the past two months, which have been well-documented by her family members and caregivers. The patient has a history of poor medication adherence, particularly with regards to her anticonvulsant medications.

Subjective:

During today's visit, Mary reported that she has been experiencing increasing frequency and severity of seizures over the past two months. She also complained of blurred vision, difficulty with balance, and occasional confusion. Mary mentioned that she has noticed a significant increase in her seizure activity since discontinuing her previous anticonvulsant medication six months ago. She is currently participating in a randomized controlled trial (RCT) evaluating the efficacy of an investigational drug for epilepsy.

Medications:

Current Medications:

* Levothyroxine (T4) 50 mcg oral daily

* Lamotrigine (LTG) 200 mg oral daily

* Valproic acid (VPA) 500 mg oral daily

* Investigational drug for epilepsy (ID-E) 500 mg oral daily (randomized controlled trial medication)

Allergies:

* None recorded

Social History:

Mary is a widow and lives alone. She has two adult children who reside in different states. Mary enjoys reading and spending time with her family, but her seizure activity has limited her ability to engage in these activities. She reported difficulty sleeping due to frequent awakenings during the night. Mary also mentioned that she has been experiencing some joint pain and stiffness, particularly in her knees and hips.

Review of Systems:

General: Denies fever, chills, or recent illness.

Neurological: Reports worsening seizure activity over the past two months, with increased frequency and severity. Also reports difficulty with balance, blurred vision, and occasional confusion.

Musculoskeletal: Reports joint pain and stiffness in knees and hips.

Psychiatric: Denies depression, anxiety, or other psychiatric symptoms.

Family History: No history of neurological disorders in first-degree relatives.

General Physical Examination:

Mary appeared well-nourished and well-groomed. Her vital signs were within normal limits. Her neck was flexible, and her fundus examination revealed no abnormalities. Her cardiac and respiratory exams were unremarkable. Mary ambulated with a cane and demonstrated a wide-based gait.

Comprehensive Neurologic Exam:

Mary's neurological exam was significant for the following:

* Seizure activity: Mary experienced a seizure during the exam, which was well-documented by her family members and caregivers. The seizure was characterized by a sudden loss of consciousness, convulsive movements of the arms and legs, and postictal confusion.

* Cranial nerves: Mary's cranial nerve examination revealed no abnormalities.

* Motor exam: Mary demonstrated a mild degree of hypotonia and ataxia, particularly in her upper extremities. Her finger-to-nose test was positive, and her rapid alternating movement (RAM) test was negative.

* Sensory exam: Mary's sensory examination revealed decreased sensation to light touch and pinprick over the left hemiparesis.

* Coordination and balance: Mary demonstrated a wide-based gait with a tendency to stagger, indicating poor balance and coordination.

Labs:

* Complete Blood Count (CBC): WBC 4.5 x 10^9/L, Neutrophils 60%, Lymphocytes 20%, Monocytes 10%, Hemoglobin 12 g/dL, Hematocrit 37%

* Blood Chemistry: Sodium 138 mmol/L, Potassium 4.2 mmol/L, Chloride 102 mmol/L, Carbon Dioxide 22 mmol/L, Glucose 126 mg/dL, BUN 20 mg/dL, Creatinine 1.5 mg/dL

* Urinalysis: No red blood cells or white blood cells on urinalysis.

Imaging:

* MRI brain: No evidence of structural brain abnormalities or tumors.

Assessment:

Mary presents with a history of hypothyroidism and generalized epilepsy, as well as poor medication adherence. Her recent increase in seizure activity is concerning, particularly given her participation in an RCT evaluating the efficacy of an investigational drug for epilepsy. The patient's joint pain and stiffness may be related to her age or hypothyroidism. A thorough workup, including blood tests and imaging studies, has been conducted to evaluate her condition.

Plan:

1. Continue the current medication regimen, including levothyroxine, lamotrigine, valproic acid, and investigational drug for epilepsy.

2. Monitor seizure activity closely and adjust medications as needed.

3. Initiate physical therapy to improve balance and coordination.

4. Consider referral to a pain management specialist for evaluation of joint pain and stiffness.

5. Schedule follow-up appointments as needed.

Signature:

[Your Name]

Date: 2022-08-04

----------------END OF PATIENT ENCOUNTER---------------

-------------- PATIENT ID 216 -- VISIT 2 -----------

2022-09-04

Patient ID 216

86 year old female

HPI:

* History of hypothyroidism and generalized epilepsy

* Currently enrolled in a randomized controlled trial (ID X93461 for epilepsy)

* Took investigational drug for 3 months, experienced 13 seizures

* Reported dizziness, somnolence, back pain

Neurological Examination:

* Pertinent positives only:

+ Dizziness

+ Somnolence

+ Back pain

Assessment and Plan:

* Discontinue investigational drug (ID X93461) since study is complete

* Monitor for seizure activity and adjust medication as needed

* Address dizziness, somnolence, and back pain with appropriate therapy

* Follow-up in 2 weeks to assess for any changes in seizure activity or adverse effects.

----------------END OF PATIENT ENCOUNTER---------------

-------------- PATIENT ID 217 -- VISIT 1 -----------

Clinic Note: Visit #1 - Patient ID 217 - 73 year old female with history of epilepsy

Date: 2022-05-24

Identification:

Patient Name: Patient ID 217 (Mrs. John Smith)

DOB: 02/01/1949

Gender: Female

Age: 73 years old

Chief Complaint:

The patient presents with a history of recurrent seizures over the past two months, with a total of 18 seizures. She has been experiencing frequent episodes of loss of consciousness and confusion, which have significantly impacted her daily activities and quality of life.

HPI (History of Present Illness):

The patient was recently enrolled in a randomized controlled trial for the treatment of epilepsy. The study is examining the efficacy and safety of an investigational drug for the treatment of refractory epilepsy. The patient has been adherent to her medications, including levetiracetam and valproate, but has experienced a significant increase in seizure activity over the past two months.

Subjective:

The patient reports feeling "very anxious" about her seizures and their impact on her daily life. She states that she has been experiencing more frequent seizures at night, which has disrupted her sleep patterns and led to increased daytime sleepiness. The patient also reports difficulty with concentration and memory, which has affected her ability to perform daily tasks and activities.

Medications:

Current medications: Levetiracetam (Keppra), valproate (Depakote)

Allergies: None reported

Social History:

The patient lives with her husband and adult children. She is a homemaker and has limited income. She reports that her seizures have significantly impacted her ability to perform daily activities, including cooking, cleaning, and taking care of her family.

Review of Systems:

The patient denies any recent illnesses or infections. She does report some mild muscle aches and pains, but these are not significant.

General Physical Examination:

Vital Signs:

* Temperature: 98.6°F (37°C)

* Blood Pressure: 120/80 mmHg

* Pulse: 75 beats per minute

* Respiratory Rate: 16 breaths per minute

Physical Examination:

The patient's general physical examination is unremarkable, with no signs of focal neurological deficits or other systemic abnormalities.

Comprehensive Neurologic Examination:

* Mental Status: The patient appears alert and oriented to person, place, and time. Her speech is clear and concise, with no evidence of aphasia or dysarthria.

* Cranial Nerves: The patient's cranial nerves are intact, with normal strength and tone in the muscles of mastication and extraocular movements.

* Motor Examination: The patient has no significant weakness or paresis, with normal tone and movement in all four limbs.

* Sensory Examination: The patient reports some numbness and tingling in her hands and feet, which is consistent with her history of epilepsy. There is no evidence of sensory deficits or abnormalities.

* Coordination and Balance: The patient's coordination and balance are normal, with no signs of ataxia or dysmetria.

Labs:

The patient's lab results are as follows:

* Complete Blood Count (CBC): Normal

* Electrolyte Panel: Normal

* Liver Function Tests (LFTs): Normal

* Thyroid-Stimulating Hormone (TSH): Normal

Imaging:

The patient has undergone a recent MRI of the brain, which revealed no significant abnormalities. There is some mild atrophy of the temporal lobe, which is consistent with her age and history of epilepsy.

Assessment:

Based on the patient's history and current symptoms, it is my assessment that she is experiencing a significant increase in seizure activity, which is likely related to her participation in the randomized controlled trial. The patient's cognitive function appears normal, with no evidence of aphasia or dysarthria. Her physical examination is unremarkable, with no signs of focal neurological deficits or other systemic abnormalities.

Plan:

The patient will continue to participate in the randomized controlled trial, with no changes made to her current medications other than adding the investigational drug for epilepsy. The patient will be monitored closely for any changes in her seizure activity or cognitive function. Regular follow-up appointments will be scheduled to assess her progress and adjust her treatment as needed.

Signature:

[Your signature]

Date: 2022-05-24

----------------END OF PATIENT ENCOUNTER---------------

-------------- PATIENT ID 217 -- VISIT 2 -----------

CLINIC NOTE - VISIT #2

Patient ID: 217

Date: 2022-06-24

History of Present Illness:

This 73-year-old female patient with a history of epilepsy has been participating in a randomized controlled trial (RCT) evaluating the investigational drug ID X93461 for the treatment of epilepsy. The patient has been taking the drug for the past three months and has experienced 21 seizures during this time. The patient has also reported symptoms such as somnolence, fatigue, nausea, dysarthria, upper respiratory infection, and gait disturbance.

Reason for Visit:

The patient has been referred to our clinic for an assessment of the investigational drug's effectiveness and any potential adverse effects. The study is now complete, and the investigational drug will be discontinued.

Neurological Examination:

General: The patient appears tired and somnolent. She has difficulty maintaining eye contact and displays mild dysarthria.

Cranial Nerves:

* Visual fields: Full visual field examination reveals no deficits.

* Extremities: No weakness or atrophy is noted in the extremities.

* Sensation: No sensory deficits are detected.

Motor Examination:

* Mobilization: The patient's limbs are somewhat stiff and resistant to mobilization, particularly in the upper extremities.

* Muscle strength: The patient displays mild weakness in the proximal muscle groups of both upper and lower extremities.

* Gait disturbance: The patient exhibits a broad-based gait with a slight shuffling effect.

Special Senses Examination:

* Ocular motility: Full eye examination shows no deficits in ocular motility.

* Vestibulo-ocular reflex: No abnormalities are detected in the vestibulo-ocular reflex.

Assessment and Plan:

Based on the patient's history of epilepsy and the adverse effects experienced during the RCT, it is determined that the investigational drug ID X93461 is not effective in controlling seizures and is associated with unacceptable side effects. The study is now complete, and the investigational drug will be discontinued.

Plan:

* Discontinue ID X93461 and replace it with a standard antiepileptic drug.

* Monitor the patient's seizure control and adverse effects closely.

* Schedule follow-up visits as needed to assess the patient's response to new medication and address any concerns or side effects that may arise.

This clinic note is a summary of the patient's neurological examination and assessment during visit #2 on June 24, 2022. The patient has been experiencing adverse effects from the investigational drug ID X93461 and will be discontinued from the study. Further monitoring and management of the patient's epilepsy will be necessary to ensure optimal seizure control and minimize potential side effects.

----------------END OF PATIENT ENCOUNTER---------------

-------------- PATIENT ID 218 -- VISIT 1 -----------

Patient ID: 218

Date: 2021-03-07

* History:

+ Parietal lobe epilepsy

+ 13 seizures in past 2 months

+ Randomized controlled trial (RCT) participant

* Neurological Examination:

+ Normal strength and tone

+ Intact sensation in all modalities

+ Normal cranial nerve examination

+ Normal coordination and balance

+ No evidence of seizure activity or focal deficits

* Pertinent Positives:

+ No change in medications other than investigational drug for epilepsy

+ 3-month trial period for investigational drug

* Assessment and Plan:

+ Monitor patient's seizure activity and response to investigational drug during RCT

+ Regular follow-up appointments every 2 weeks to assess patient's status and adjust plan as needed

+ Patient remains in RCT for 3 months, then re-evaluate effectiveness of treatment and make any necessary changes.

----------------END OF PATIENT ENCOUNTER---------------

-------------- PATIENT ID 218 -- VISIT 2 -----------

Clinic Note: Visit #2 (2021-04-07)

Patient ID: 218

Age: 48 years old

History: Patient has a history of parietal lobe epilepsy. He is currently participating in a randomized controlled trial studying the investigational drug ID X93461 for the treatment of epilepsy. The patient has been taking the drug for the past three months and has experienced 7 seizures during this time. He has also reported dizziness, somnolence, and nausea as side effects.

Neurological Examination:

On examination, the patient's vital signs are within normal limits. His consciousness is clear, and he is able to speak freely. There is no evidence of focal weakness or sensory deficits. His cranial nerves are intact, and his coordination and balance are normal. The patient's reflexes are also normal, and there is no evidence of ataxia or dysmetria.

Assessment:

Based on the patient's history and current symptoms, it is evident that the investigational drug ID X93461 has not been effective in controlling his epilepsy. The patient has experienced a significant number of seizures during the study period, and has also reported bothersome side effects. In light of these findings, it is decided to discontinue the investigational drug and explore alternative treatments for the patient's epilepsy.

Plan:

The patient will be advised to discontinue the investigational drug ID X93461 and will be referred to a specialist for further management of his epilepsy. His seizure activity and side effects will be closely monitored, and any necessary adjustments will be made to his treatment plan as needed. Regular follow-up appointments will be scheduled to assess the patient's progress and adjust the treatment plan accordingly.

----------------END OF PATIENT ENCOUNTER---------------

-------------- PATIENT ID 219 -- VISIT 1 -----------

Clinic Note Visit #1

Patient ID: 219

Date: 2022-08-17

Chief Complaint: The patient presents with a history of schizophrenia and frontal lobe epilepsy, with a recent increase in seizure frequency.

History of Present Illness: The patient has experienced 44 seizures over the past two months, with no changes in medication other than the addition of an investigational drug for epilepsy which will be taken for three months as part of a randomized controlled trial.

Neurological Examination:

Mental Status: The patient's mental status is normal, with no evidence of psychosis or clouding of consciousness.

Cranial Nerves: All cranial nerves are intact, with normal strength and sensation in the face and extremities.

Motor Examination: The patient has normal muscle tone and bulk, with full range of motion in all joints. There is no evidence of tremors or rigidity.

Sensory Examination: The patient has normal sensation to light touch, pain, and temperature over the entire body, including the face and extremities. There is no evidence of sensory loss or dysesthesias.

Reflexes: All reflexes are normal, with equal and appropriate resistance to flexion and extension in all joints.

Gait and Coordination: The patient ambulates normally with a steady gait and no signs of ataxia or dysmetria.

Assessment: The patient presents with a history of schizophrenia and frontal lobe epilepsy, with a recent increase in seizure frequency. No changes in medication other than the addition of an investigational drug for epilepsy will be made for the next three months as part of a randomized controlled trial. The patient's neurological examination reveals no abnormalities.

----------------END OF PATIENT ENCOUNTER---------------

-------------- PATIENT ID 219 -- VISIT 2 -----------

---

Patient: Patient ID 219, 70 year old female

Chief Complaint: None

History of Present Illness: The patient is a 70-year-old female with a history of schizophrenia and frontal lobe epilepsy. She has been participating in a randomized controlled trial (RCT) evaluating the investigational drug ID X93461 for the treatment of epilepsy. The patient has been taking the medication for the past 3 months and has reported 18 seizures during this time. The patient has otherwise been asymptomatic.

Past Medical History: Schizophrenia, frontal lobe epilepsy

Medications: ID X93461 (investigational drug), levetiracetam (anticonvulsant)

Social History: None

Review of Systems: None

Neurological Examination:

Motor Examination: The patient's motor function is intact, with no evidence of weakness, atrophy, or tremors.

Sensory Examination: The patient's sensation is intact, with no evidence of numbness, tingling, or loss of sensation.

Cranial Nerve Examination: The patient's cranial nerves are normal, with no evidence of abnormalities in vision, hearing, smell, taste, or facial movements.

Autonomic Examination: The patient's autonomic function is intact, with no evidence of hypotension, tachycardia, or other abnormalities.

Assessment and Plan:

The patient has completed the 3-month RCT evaluating ID X93461 for the treatment of epilepsy. The patient has reported a total of 18 seizures during this time, which is consistent with the expected frequency given the patient's history of frontal lobe epilepsy. The investigational drug will be discontinued now that the study is complete.

The patient's neurological examination reveals no significant abnormalities, and her symptoms are well-controlled with current medications. However, given the patient's history of schizophrenia and frontal lobe epilepsy, it is important to closely monitor her condition and adjust her treatment plan as needed.

The patient will be followed up in 2 weeks for a follow-up appointment to assess her condition and adjust her medications as needed. Additionally, the patient will be advised to continue with her current medications and to report any changes in her symptoms or side effects.

---

Note: This is a fictional patient and the information provided is for educational purposes only.

----------------END OF PATIENT ENCOUNTER---------------

-------------- PATIENT ID 230 -- VISIT 1 -----------

Patient ID: 230

Date: 2021-12-05

HPI:

* 97-year-old female

* History of hypothyroidism, arthritis, bipolar disorder, and occipital lobe epilepsy

* 13 seizures in past 2 months

* Randomized controlled trial (RCT) participant

Neurological Examination:

* Mental status: Alert and oriented x3

* Speech: Clear and coherent

* Motor: No observable tremors or rigidity

* Sensation: Intact

* Reflexes: 2+ (moderate)

* Coordination: Normal

Pertinent Positives:

* Seizures occurring every 2-3 days

* No recent changes in medications

Assessment and Plan:

* Continue RCT medication as prescribed

* Monitor seizure activity and adjust plan as needed

* Reassess medication regimen after 3 months of investigational drug therapy.

----------------END OF PATIENT ENCOUNTER---------------

-------------- PATIENT ID 230 -- VISIT 2 -----------

Patient ID 230, a 97-year-old female, was seen in clinic on January 5, 2022 for her second visit. The patient has a history of hypothyroidism, arthritis, bipolar disorder, and occipital lobe epilepsy. She has been participating in a randomized controlled trial and has been receiving the investigational drug ID X93461 for the past three months to treat her epilepsy.

During this visit, the patient reported experiencing balance disorders, headaches, and decreased appetite over the past few months. The patient also mentioned that she had experienced five seizures since starting the investigational drug. These symptoms are concerning and may be related to the medication. A complete neurological examination was performed, including an assessment of the patient's balance, coordination, and reflexes. Additionally, the patient's cognitive function, memory, and mood were evaluated.

The results of the neurological examination revealed some concerning findings. The patient had difficulty with balance and coordination, and her reflexes were decreased. Her cognitive function was also affected, as she had difficulty recalling recent events and exhibited poor judgment. Furthermore, the patient reported feeling tired and lethargic, which may be related to the medication.

Based on the results of the neurological examination and the patient's symptoms, it is recommended that the investigational drug ID X93461 be discontinued. The patient will be transitioned back to her previous medications, including levothyroxine for hypothyroidism and a non-steroidal anti-inflammatory drug (NSAID) for arthritis. Regular follow-up appointments will be scheduled to monitor the patient's condition and address any concerns or symptoms that may arise in the future.

----------------END OF PATIENT ENCOUNTER---------------

-------------- PATIENT ID 231 -- VISIT 1 -----------

On March 5th, 2021, I had the pleasure of conducting a clinic visit with Patient ID 231, a 32-year-old female who presented with a history of occipital lobe epilepsy. The patient has experienced an alarming increase in seizure frequency over the past two months, with a total of 11 seizures during this timeframe. Given the patient's medical background and current symptoms, it was decided that she would participate in a randomized controlled trial aimed at exploring the efficacy of an investigational drug for epilepsy.

During the clinic visit, I conducted a thorough neurological examination to assess the patient's overall condition and identify any potential triggers or contributing factors to her seizures. The examination revealed no notable abnormalities in the patient's cognitive function, balance, or reflexes. However, I did observe some subtle changes in the patient's speech patterns and eye movements, which may indicate a mild degree of dysfunction in the patient's temporal lobe.

In light of these findings, it is essential that we continue to closely monitor the patient's condition and adjust her treatment plan as needed. The patient has been prescribed an investigational drug for epilepsy, which she will be taking for a period of three months. During this time, it will be crucial to track the patient's seizure frequency and assess the efficacy of the new medication. Additionally, I have advised the patient to maintain a detailed seizure journal to help us better understand the timing and severity of her seizures.

In conclusion, clinic visit #1 with Patient ID 231 was an opportunity for us to assess the patient's current condition and outline a plan for managing her epilepsy moving forward. While some subtle changes in the patient's speech patterns and eye movements were observed, it is essential that we continue to closely monitor her condition and adjust her treatment plan as needed. With the patient participating in a randomized controlled trial and taking an investigational drug for epilepsy, we are optimistic about improving her seizure frequency and overall quality of life.

----------------END OF PATIENT ENCOUNTER---------------

-------------- PATIENT ID 231 -- VISIT 2 -----------

---

Clinic Note - Visit #2

Patient ID: 231

Date: 2021-04-05

Chief Complaint: Balance disorder, history of occipital lobe epilepsy

History of Present Illness:

The patient is a 32-year-old female with a history of occipital lobe epilepsy who has been participating in a randomized controlled trial for the investigational drug ID X93461 for the past 3 months. During this time, she has had 3 seizures. The patient has also reported a balance disorder.

Past Medical History:

* Occipital lobe epilepsy

* History of seizure activity

Medications:

* Levetiracetam (Keppra)

* Lamotrigine (Lamictal)

Social History:

* No significant social history

Family History:

* No significant family history

Review of Systems:

* No other symptoms or complaints

Neurological Examination:

* Mental Status: Alert and oriented

* Head: Normal head shape and size, no focal defects

* Eyes: Pupils are equal and reactive to light, no abnormal movements or deviations

* Ears: No cerumen or ear discharge, no hearing loss or distortion

* Nose: No nasal discharge or obstruction, no anosmia

* Throat: No throat tenderness or swelling, no odynophagia

* Neck: No neck stiffness or masses, normal range of motion

* Extremities: No weakness or paresthesias in the arms or legs, normal tone and reflexes

* Sensory Examination: Normal sensation to light touch, pinprick, and temperature

* Coordination and Balance: The patient has a balance disorder and reports unsteadiness when walking or standing.

Assessment and Plan:

* The patient's history of occipital lobe epilepsy and recent seizure activity suggest ongoing epilepsy that may be refractory to current medications.

* The patient's balance disorder is likely related to the investigational drug ID X93461, which will be discontinued given the completion of the study.

* Given the patient's history of seizure activity and ongoing epilepsy, it is important to continue close monitoring and adjust medications as needed to control seizures.

* The patient's balance disorder should also be addressed with appropriate therapies such as physical therapy or vestibular rehabilitation.

Plan:

* Continue current medications (levetiracetam, lamotrigine) and monitor seizure activity closely.

* Discontinue the investigational drug ID X93461 given the completion of the study.

* Refer the patient to physical therapy or vestibular rehabilitation for balance disorder.

* Schedule a follow-up appointment in 2-3 months to assess seizure control and balance symptoms.

----------------END OF PATIENT ENCOUNTER---------------

-------------- PATIENT ID 232 -- VISIT 1 -----------

On June 3rd, 2021, I conducted visit #1 with Patient ID 232, a 46-year-old female with a history of bipolar disorder and epilepsy. The patient has experienced 124 seizures in the past two months, which is a significant increase from her usual frequency. Given the patient's history and recent seizure activity, it was decided that she will continue to participate in the randomized controlled trial (RCT) for epilepsy. No changes will be made to her current medication regimen other than adding the investigational drug for the next three months.

During the neurological examination, the patient's vital signs were within normal limits. Her speech and language were clear, and she was able to convey her thoughts effectively. The patient's consciousness was intact, and there were no signs of cognitive impairment or altered mental status. Her motor function was generally unimpaired, with the exception of mild weakness in the left extremities. However, the patient did exhibit increased muscle tone and a slight tremor in her hands.

The patient's sensory examination revealed no deficits or abnormalities in hearing, vision, or tactile sensation. Her reflexes were within normal limits, and there was no evidence of ataxia or other abnormal gait patterns. The patient's cranial nerve examination showed no abnormalities, and her cognitive function was intact.

In conclusion, the neurological examination conducted during visit #1 revealed no significant changes in the patient's condition since her last visit. Despite her recent increase in seizure activity, the patient remains stable and will continue to participate in the RCT for epilepsy. The addition of the investigational drug is expected to help manage her seizures and improve her overall health outcomes.

----------------END OF PATIENT ENCOUNTER---------------

-------------- PATIENT ID 232 -- VISIT 2 -----------

---

Clinic Note #2, dated 2021-07-03, for Patient ID 232:

Identification:

Name: [Patient ID 232]

DOB: [Date of Birth]

Gender: Female

Age: 46 years

Highlights of the Present Illness (HPI):

Chief Complaint (CC): The patient presents with a history of bipolar disorder and epilepsy, currently participating in a randomized controlled trial (RCT) for the investigational drug ID X93461 for the treatment of epilepsy.

History of Present Illness (HPI): The patient has been experiencing increased seizure activity over the past 3 months since initiating the investigational drug, with a total of 133 seizures reported during this time. The patient also reports experiencing dizziness and somnolence.

Subjective:

The patient reports feeling generally well, but notes that she has been experiencing an increase in seizure activity since initiating the investigational drug. She also reports feeling dizzy and sleepy.

Medications:

1. ID X93461 (investigational drug for epilepsy) - discontinued as of today's date due to completion of RCT

2. Lamotrigine (for bipolar disorder)

3. Levetiracetam (for seizures)

Allergies:

1. No known allergies

Social History:

1. Married with two children

2. Currently employed as a teacher

3. Denies smoking and alcohol consumption

4. Reports stress related to work and family life

Review of Systems (ROS):

1. Neurological: The patient reports an increase in seizure activity since initiating the investigational drug, with a total of 133 seizures reported during this time. She also reports experiencing dizziness and somnolence.

2. General: No other complaints reported.

General Physical Examination (GPE):

1. Vital Signs: Blood pressure - 120/80 mmHg, Heart rate - 76 beats per minute, Temperature - 98.6°F (37°C)

2. Head: Normocephalic, normal scalp hair

3. Eyes: No evidence of abnormal eye movements or vision changes

4. Ears: No evidence of abnormal hearing or discharge

5. Nose: No evidence of abnormal nasal secretions or deformities

6. Throat: Normopharyngeal, no evidence of thrush or lesions

7. Chest: Clear to auscultation, no signs of distress

8. Abdomen: Soft, non-tender, no rebound tenderness or guarding

9. Extremities: No clubbing or cyanosis, no evidence of joint or muscle pain

10. Neurological Examination (see below)

Comprehensive Neurologic Exam (CNE):

1. Cranial Nerves: Intact, except for a mild right facial weakness

2. Motor Examination: No evidence of weakness or atrophy

3. Sensory Examination: Intact, except for a mild decrease in proprioception on the right side

4. Reflexes: 1+ (hyperreflexia) bilaterally

5. Gait and Station: Normal, with no evidence of ataxia or gait abnormalities

6. Speech and Language: Intact, except for a mild slurring of words

7. Cognitive Examination: No evidence of cognitive impairment or disorientation

Labs:

1. Complete Blood Count (CBC): WBC - 6,500 cells/mm³, NEUT - 3,200 cells/mm³, LYMPH - 800 cells/mm³, RBC - 4.8 million cells/mm³, Hemoglobin - 14.2 g/dL, Hematocrit - 46.5%

2. Electrolyte Panel: Sodium - 139 mmol/L, Potassium - 4.2 mmol/L, Chloride - 100 mmol/L, Carbon Dioxide - 22 mmol/L

3. Renal Function Panel: Creatinine - 1.1 mg/dL, BUN - 25 mg/dL

4. Lipid Panel: Cholesterol - 180 mg/dL, Triglycerides - 120 mg/dL

Imaging:

1. MRI Brain (done on 06-01-2021): No evidence of acute or chronic changes in the brain

2. EEG (done on 06-01-2021): No evidence of seizure activity

Assessment:

The patient is a 46-year-old female with a history of bipolar disorder and epilepsy, currently participating in a randomized controlled trial for the investigational drug ID X93461. She has experienced an increase in seizure activity since initiating the drug, with a total of 133 seizures reported during this time. The patient also reports experiencing dizziness and somnolence. The patient's neurological examination is essentially normal, except for a mild right facial weakness and hyperreflexia bilaterally. The patient's laboratory results are within normal limits, with the exception of a slightly elevated BUN. The patient's imaging studies show no evidence of acute or chronic changes in the brain.

Plan:

1. Discontinue the investigational drug ID X93461 due to completion of RCT

2. Monitor the patient's seizure activity and adjust medications as needed

3. Repeat neurological examination and laboratory studies in 3-6 months to assess for any changes in her condition

Signature:

[Your signature here]

---

Note: This is just an example of a clinic note, and the specific details and organization may vary depending on the needs of your patient and the requirements of your clinic.

----------------END OF PATIENT ENCOUNTER---------------

-------------- PATIENT ID 233 -- VISIT 1 -----------

---

Clinic Note #1 - Patient ID 233 (2021-07-31)

Identification:

Patient Name: [Name], [Initials]

DOB: [Date of Birth]

ID Number: [Patient ID]

HPI:

Chief Complaint: The patient presents with a history of diabetes and frontal lobe epilepsy. She has experienced 11 seizures in the past two months, with the most recent occurring yesterday. The patient is currently enrolled in a randomized controlled trial and is not anticipating any changes to her medication regimen other than the addition of an investigational drug for epilepsy, which she will take for the next three months.

Subjective:

The patient reports experiencing increasing frequency and severity of seizures over the past two months. She denies any recent changes in her diet or medication regimen that may be contributing to the increased seizure activity. The patient notes that she has been feeling generally well, with no new symptoms or concerns.

Medications:

1. Levetiracetam (Keppra) - 500mg bid

2. Lamotrigine (Lamictal) - 200mg qd

3. Valproic acid (Depakote) - 500mg tid

4. Investigational drug for epilepsy (ID-E) - 500mg bid (starting today and continuing for the next three months)

Allergies:

The patient reports no known allergies.

Social History:

Marital status: Married

Occupation: Teacher

Number of dependents: 2 (ages 8 and 10)

Smoking: Does not smoke

Alcohol consumption: Occasional, socially

Recreational drug use: None

Review of Systems:

General: No recent changes in general health or constitutional symptoms.

Cardiovascular: No chest pain, shortness of breath, or palpitations.

Respiratory: No cough, fever, or difficulty breathing.

Gastrointestinal: No abdominal pain, nausea, vomiting, or diarrhea.

Genitourinary: No urinary frequency, hesitancy, or hematuria.

Musculoskeletal: No joint pain or swelling.

Nervous: No recent changes in neurological symptoms other than increased seizure activity.

General Physical Examination:

Vital signs: Blood pressure - 120/80 mmHg, Heart rate - 70 bpm, Temperature - 98.6°F (37°C)

Comprehensive Neurologic Examination:

Mental Status: Alert and oriented x3. No evidence of aphasia, dysarthria, or neglect.

Cranial Nerves:

* Ocular: Pupils are equal and reactive to light, with no evidence of diplopia or abnormal eye movements.

* Facial: No evidence of weakness or paresis in facial muscles.

* Auditory: No evidence of hearing loss or abnormal ear examination.

* Visual: No evidence of visual field defects or abnormal visual acuity.

* Tongue: No evidence of tongue weakness or atrophy.

* Extremities: No evidence of weakness, paresis, or sensory deficits in the extremities.

Laboratory Findings:

1. Complete Blood Count (CBC): Within normal limits

2. Serum electrolytes: Within normal limits

3. Renal function tests: Within normal limits

4. Thyroid-stimulating hormone (TSH) level: With normal limits

5. Lipid profile: Within normal limits

Imaging Findings:

1. Magnetic Resonance Imaging (MRI) of the brain: No evidence of structural abnormalities or lesions.

2. Electroencephalogram (EEG): Shows frequent, high-amplitude spikes consistent with frontal lobe epilepsy.

Assessment:

The patient presents with a history of diabetes and frontal lobe epilepsy, with an increased frequency and severity of seizures over the past two months. The patient is currently enrolled in a randomized controlled trial and is not anticipating any changes to her medication regimen other than the addition of an investigational drug for epilepsy. The patient reports no recent changes in her diet or medication regimen that may be contributing to the increased seizure activity. Based on the patient's history, physical examination, and laboratory and imaging findings, it is assessed that the patient is experiencing frontal lobe epilepsy with no evidence of other etiologies.

Plan:

1. Continue current medication regimen, including levetiracetam, lamotrigine, and valproic acid.

2. Add investigational drug for epilepsy (ID-E) at a dose of 500mg bid starting today and continuing for the next three months.

3. Monitor seizure activity and adjust medication regimen as needed.

4. Repeat EEG and MRI imaging in 3 months to assess response to therapy and monitor for any changes in brain activity or structural abnormalities.

Signature:

[Your Name]

Neurologist

Note: This clinic note is a fictional example and should not be used as a basis for actual patient care. It is meant to illustrate the type of information that may be included in a clinic note, and it should not be considered a comprehensive or definitive guide to clinical practice.

----------------END OF PATIENT ENCOUNTER---------------

-------------- PATIENT ID 233 -- VISIT 2 -----------

Patient ID: 233, Female, Age: 44

History: Diabetes, Frontal Lobe Epilepsy (study participant)

Investigational Drug: ID X93461 (discontinued)

Neurological Examination:

* Dizziness (reported by patient)

Pertinent Positives:

* 16 seizures in past 3 months

Assessment and Plan:

* Discontinue investigational drug ID X93461 due to study completion

* Monitor for seizure activity and dizziness in follow-up visits

* Consider alternative treatment options for epilepsy as needed

----------------END OF PATIENT ENCOUNTER---------------

-------------- PATIENT ID 234 -- VISIT 1 -----------

---

Clinic Note #1 - Patient ID: 234 - Visit Date: 2023-01-08

Chief Complaint: The patient presents with a history of autism, bipolar disorder, hypertension, and occipital lobe epilepsy. She has experienced 5 seizures in the past two months.

Past Medical History: The patient was diagnosed with autism at age 3, bipolar disorder at age 19, and hypertension at age 28. She has a history of occipital lobe epilepsy and has been on levetiracetam for the past 5 years.

Current Medications: Levetiracetam (500mg twice daily), lithium carbonate (600mg twice daily), and olanzapine (10mg once daily)

Social History: The patient is currently enrolled in a randomized controlled trial investigating the efficacy of an investigational drug for the treatment of epilepsy. She has been advised not to change her medications during the trial period, except for the addition of the investigational drug.

Family History: The patient's father and brother have a history of seizures.

Review of Systems: The patient denies any recent changes in mood, appetite, or sleep patterns. She reports no new focal neurological symptoms, such as weakness, numbness, or paresthesias.

General Observations: The patient appears anxious and agitated during the examination. She has a monotone speech pattern and displays repetitive behaviors.

Neurological Examination:

Motor: The patient's motor examination reveals normal strength, tone, and coordination. There is no evidence of any focal neurological deficits.

Sensory: The patient's sensory examination reveals normal sensation to light touch, pain, temperature, and vibration over all extremities. She does not report any sensory deficits or paresthesias.

Cerebellar Function: The patient's cerebellar function is normal, with no evidence of ataxia, dysmetria, or intention tremors.

Visual Function: The patient's visual function is normal, with no evidence of blurring, double vision, or field defects.

Cognitive Function: The patient's cognitive function is normal, with no evidence of impairment in attention, memory, or executive functions.

Assessment and Plan:

Assessment: The patient presents with a history of autism, bipolar disorder, hypertension, and occipital lobe epilepsy. She has experienced 5 seizures in the past two months. Her current medications include levetiracetam, lithium carbonate, and olanzapine. The patient is participating in a randomized controlled trial investigating the efficacy of an investigational drug for the treatment of epilepsy.

Plan:

1. Continue the patient's current medications, including levetiracetam, lithium carbonate, and olanzapine.

2. Add the investigational drug for the treatment of epilepsy, as per the trial protocol.

3. Monitor the patient's seizure activity and adjust the medication regimen as needed.

4. Schedule follow-up visits every 2 weeks to assess the patient's response to the medications and monitor for any adverse effects.

5. Advise the patient to avoid any changes in her medications during the trial period, except for the addition of the investigational drug.

I have discussed the above plan with the patient, and she has agreed to participate in the randomized controlled trial and follow the recommended medication regimen.

---

Note: This clinic note is a fictional example and should not be used as a basis for actual patient care. It is intended only as an educational tool to demonstrate the style and content of a neurologist's clinic note.

----------------END OF PATIENT ENCOUNTER---------------

-------------- PATIENT ID 234 -- VISIT 2 -----------

On February 8th, 2023, I conducted a routine follow-up visit with Patient ID 234, a 43-year-old female with a history of autism, bipolar disorder, hypertension, and occipital lobe epilepsy. The patient has been participating in a randomized controlled trial (RCT) for the investigational drug ID X93461, which she has been taking for the past three months. During this time, she has experienced 17 seizures, and reported mild nausea.

Upon examination, the patient's neurological status was stable, with no significant changes from previous visits. Her speech and language were normal for her age and cognitive ability, and her motor function was intact, with no signs of weakness or ataxia. The patient's sensation was grossly normal, with no evidence of numbness or paresthesias. Her reflexes were appropriate for her age and condition, and her cranial nerves were intact.

The patient's medication regimen included the investigational drug ID X93461, which was discontinued due to the completion of the RCT. The patient's blood pressure was within normal limits, and her electrocardiogram (ECG) showed no signs of arrhythmia or other cardiac abnormalities.

In light of the patient's history of occipital lobe epilepsy and recent seizure activity, it is my recommendation that she continue to receive regular follow-up care with a neurologist to monitor her condition and adjust her medication regimen as needed. Additionally, given the patient's history of bipolar disorder, it is important that she continues to receive psychiatric care to manage her mood and prevent any potential relapses.

----------------END OF PATIENT ENCOUNTER---------------

-------------- PATIENT ID 235 -- VISIT 1 -----------

Patient ID: 235

DOB: 1940

Gender: Female

Age: 82

Chief Complaint: Temporal lobe epilepsy with recent increase in seizure frequency

History of Present Illness:

* 7 seizures in past 2 months

* History of temporal lobe epilepsy

* Currently in randomized controlled trial

Neurological Examination:

* Mild decline in cognitive function

* No focal neurological deficits

* Normal cranial nerve examination

* Normal motor examination

Pertinent Positives:

* Recent increase in seizure frequency

* History of temporal lobe epilepsy

Assessment and Plan:

* Randomized controlled trial ongoing

* No change in medications other than addition of investigational drug for epilepsy

* Drug to be taken for 3 months

* Monitor seizure frequency and cognitive function every 2 weeks

* Re-evaluate medication regimen after 3 months

----------------END OF PATIENT ENCOUNTER---------------

-------------- PATIENT ID 235 -- VISIT 2 -----------

Visit #2: 2023-01-23

Patient ID: 235

Age: 82 y/o

History: temporal lobe epilepsy

Investigational Drug: ID X93461 (3 months)

Seizure Activity:

* 3 seizures during ID X93461 treatment

* Somnolence reported by patient

Neurological Examination:

* General assessment: alert and oriented

* Mental status: normal

* Cranial nerves: normal

* Motor examination: normal

* Sensory examination: normal

* Reflexes: normal

* Gait: normal

Pertinent Positives:

* No new seizures since last visit

* Somnolence resolved

Assessment and Plan:

* Discontinue investigational drug ID X93461 due to study completion

* Monitor patient for any changes in seizure activity or adverse effects

* Schedule follow-up visit in 2 weeks to assess patient's condition.

----------------END OF PATIENT ENCOUNTER---------------

-------------- PATIENT ID 236 -- VISIT 1 -----------

On April 1st, 2021, Patient ID 236, a 59-year-old male, presented to our clinic for his first visit. The patient has a history of anxiety, asthma, seasonal allergies, and epilepsy. He has been experiencing an increase in seizure activity over the past two months, with a total of 8 seizures occurring during this time.

During the neurological examination, the patient's vitals were within normal limits. His consciousness and cognitive function were intact, with no signs of confusion or disorientation. The patient's speech was clear and coherent, without any slurring or difficulty articulating words. His motor functions were grossly normal, with no signs of weakness, tremors, or rigidity.

The patient's neurological examination revealed no abnormalities in his sensory systems, including vision and hearing. His reflexes were within normal limits, and there was no evidence of any muscle weakness or atrophy. The patient's gait was normal, without any signs of imbalance or instability.

The patient has been randomized to receive the investigational drug for epilepsy, which will be administered for a period of 3 months. No changes will be made to the patient's current medications other than adding this new drug. It is important to closely monitor the patient's seizure activity and adjust the treatment plan as needed to ensure optimal control of his condition.

----------------END OF PATIENT ENCOUNTER---------------

-------------- PATIENT ID 236 -- VISIT 2 -----------

Clinic Note - Visit #2

Patient ID: 236

Date: 2021-05-01

Chief Complaint: The patient presents with a history of anxiety, asthma, seasonal allergies and epilepsy. He has been participating in a randomized controlled trial for the investigational drug ID X93461 for epilepsy over the past 3 months.

Past Medical History: The patient has a history of anxiety, asthma, seasonal allergies and epilepsy. He has been experiencing seizures for several years and has tried various medications without significant improvement.

Medication List: The patient is currently taking Levetiracetam, Lamotrigine and Valproate for his epilepsy. He also takes Prednisone for asthma and Fluticasone for seasonal allergies.

Social History: The patient denies any significant social or occupational activities due to his seizure disorder. He lives with his wife and two children.

Family History: The patient's father had a history of seizures and died at the age of 65 due to a seizure-related accident. The patient's mother has hypertension and hyperlipidemia.

Review of Systems: The patient reports no significant changes in his symptoms over the past 3 months. He has experienced nausea and diplopia, which he believes are related to the investigational drug ID X93461.

Neurological Examination:

* General: The patient appears well-oriented and alert.

* Mental Status: The patient's cognitive function is intact.

* Cranial Nerves:

+ Visual acuity: 20/25 OU (right eye, left eye)

+ Extremities: Normal tone and reflexes

+ Sensory: Intact sensation to light touch, pinprick, and temperature

+ Coordination and balance: Normal

* Motor: Normal strength and range of motion in all extremities

* Gait: Normal gait and station

* Reflexes: Normal reflexes bilaterally

* Special Senses:

+ Visual fields: Full visual fields OU

+ Pupils: Equal, round, and reactive to light

+ Intraocular pressure: Within normal limits

* Dental Examination: Normal dentition and oral hygiene

Assessment:

* The patient has a history of anxiety, asthma, seasonal allergies and epilepsy.

* He has been participating in a randomized controlled trial for the investigational drug ID X93461 for epilepsy over the past 3 months and has reported nausea and diplopia.

* The patient's seizure frequency has increased to 15 seizures over the past 3 months while taking the investigational drug, which will be discontinued now that the study is complete.

Plan:

* Discontinue the investigational drug ID X93461

* Increase the dosage of Levetiracetam to 2000mg/day and Lamotrigine to 400mg/day to optimize seizure control

* Schedule a follow-up visit in 2 weeks to reassess seizure frequency and monitor for any adverse effects of the medication change.

* Encourage the patient to continue with his current asthma and seasonal allergy management.

* Provide education on the potential side effects of the medications and encourage the patient to report any changes in symptoms or concerns.

----------------END OF PATIENT ENCOUNTER---------------

-------------- PATIENT ID 237 -- VISIT 1 -----------

Clinic Note #1 - Patient ID 237 - 2021-03-14

Identification:

Name: Patient ID 237 (Ms. A. Smith)

DOB: March 3, 1973

Gender: Female

ID #: 237

HPI: Ms. Smith is a 48-year-old female who presents with a history of diabetes and parietal lobe epilepsy. She has experienced nine seizures over the past two months, which have been well-documented by her family members and caregivers.

Subjective:

Ms. Smith reports experiencing an increase in seizure frequency over the past two months. She also notes difficulty with memory and concentration, as well as recent changes in her mood, including increased irritability and anxiety. Ms. Smith denies any recent changes in her medication regimen or any other significant medical history.

Medications:

Current Medications:

* Levetiracetam (Keppra) 500mg BID

* Lamotrigine (Lamictal) 200mg OD

* Valproate (Depakote) 500mg TID

Allergies:

None reported.

Social History:

Ms. Smith is a married mother of two, with an 18-year-old son and a 13-year-old daughter. She works as a part-time librarian and reports no significant changes in her social or work-related activities over the past two months. Ms. Smith denies any history of substance abuse or illicit drug use.

Review of Systems:

General: Ms. Smith reports feeling generally well, with no recent constitutional symptoms such as fever, chills, or weight loss.

Neurological: Ms. Smith reports worsening seizure activity over the past two months, with an increase in the frequency and severity of her seizures. She also notes difficulty with memory and concentration, as well as recent changes in her mood.

Past Medical History:

Ms. Smith has a history of diabetes and parietal lobe epilepsy. She has been treated with levetiracetam (Keppra), lamotrigine (Lamictal), and valproate (Depakote) for the past five years, with regular monitoring and adjustments as needed.

Family History:

Ms. Smith's mother and maternal grandmother both had a history of epilepsy. Her father died of a heart attack at the age of 55.

General Physical Examination:

Vital Signs:

* Temperature: 98.6°F (37°C)

* Pulse: 80/min

* Respiration Rate: 16/min

* Blood Pressure: 120/80 mmHg

* Weight: 58 kg (128 lbs)

Head and Neck:

* Normal head shape and proportions.

* No signs of trauma or edema.

* Intact hair and scalp.

Extremities:

* No signs of clubbing or cyanosis.

* Normal tone and reflexes in upper and lower limbs.

* No evidence of muscle weakness or atrophy.

Neurological Examination:

Ms. Smith underwent a comprehensive neurological examination, including assessment of her cognitive function, motor function, sensory system, and reflexes.

Cognitive Function:

* Ms. Smith was able to recall recent events and perform basic arithmetic calculations with minimal difficulty. However, she did report difficulty with memory and concentration, which is consistent with her reported history of seizures.

Motor Function:

* Ms. Smith had a normal gait and station, with no signs of ataxia or rigidity. Her muscle strength and tone were within normal limits.

Sensory System:

* Ms. Smith had a normal sensation in all four limbs, with no signs of numbness or tingling. Her vision and hearing were normal.

Reflexes:

* Ms. Smith had normal reflexes in all four limbs, with no signs of hyporeflexia or hyperreflexia.

Labs:

* Complete Blood Count (CBC): Within normal limits

* Electrolyte Panel: Within normal limits

* Lipid Panel: Within normal limits

* Thyroid-Stimulating Hormone (TSH): Within normal limits

Imaging:

* MRI Brain: No evidence of structural abnormalities or lesions.

Assessment:

Ms. Smith presents with a history of parietal lobe epilepsy and recent increases in seizure frequency, which is concerning for potential medication non-adherence or progression of her disease. Her cognitive function and motor function are within normal limits, but she does report difficulty with memory and concentration, as well as recent changes in her mood.

Plan:

1. Continue current medications (levetiracetam, lamotrigine, and valproate) and monitor for any changes in seizure frequency or severity.

2. Add the investigational drug (I.D.) to Ms. Smith's regimen for the next three months, with close monitoring of her clinical status and laboratory results.

3. Repeat MRI Brain in three months to assess for any changes in her brain structure or function.

4. Encourage Ms. Smith to maintain a detailed seizure log to monitor for any changes in her seizure activity.

5. Schedule follow-up appointments every three months to assess her clinical status and make any necessary adjustments to her medication regimen.

Signature:

[Your Signature]

Date: 2021-03-14

----------------END OF PATIENT ENCOUNTER---------------

-------------- PATIENT ID 237 -- VISIT 2 -----------

Patient ID: 237

Visit #2: 2021-04-14

History:

* Diabetes

* Parietal lobe epilepsy

* Randomized controlled trial (RCT) participant

* Took investigational drug ID X93461 for epilepsy (last 3 months)

* Experienced 11 seizures during this time

Neurological Examination:

* General: Alert and oriented

* Mental Status: Normal

* Cranial Nerves: All grossly normal

* Motor Examination: No abnormalities

* Sensory Examination: Normal

* Reflexes: Normal

* Gait and Station: Normal

Pertinent Positives:

* No symptoms reported

Assessment:

* Completed RCT, discontinuing investigational drug ID X93461

Plan:

* Discontinue investigational drug ID X93461

* Monitor for seizure activity and any adverse effects

----------------END OF PATIENT ENCOUNTER---------------

-------------- PATIENT ID 238 -- VISIT 1 -----------

Patient ID: 238

Date: 2022-09-10

HPI:

* 96-year-old male

* History of parietal lobe epilepsy

* Seven seizures in past 2 months

* Randomized controlled trial (RCT) participant

Neurological Examination:

* Mild cognitive decline

* No changes in motor function or coordination

* No focal neurological deficits

* Normal sensation and reflexes

Pertinent Positives:

* No change in seizure frequency or severity since starting RCT

* Patient reports continued difficulty with memory and concentration

Assessment and Plan:

* Continue RCT medication as prescribed

* Monitor seizure frequency and severity every 2 weeks

* Evaluate cognitive function at end of RCT

* Consider adjusting medications based on study results.

----------------END OF PATIENT ENCOUNTER---------------

-------------- PATIENT ID 238 -- VISIT 2 -----------

Visit #2, October 10, 2022

Patient ID: 238

Age: 96 years

History: Parietal lobe epilepsy

Investigational Drug: ID X93461 (randomized controlled trial)

Neurological Examination:

* General: Alert and oriented

* Mental Status: Normal

* Language: Intact

* Motor: No significant weakness or ataxia

* Sensation: Normal

* Reflexes: 2+ (normal)

* Abnormalities: Decreased appetite

Pertinent Positives:

* 4 seizures in the past 3 months while taking ID X93461

* Decreased appetite

Assessment and Plan:

* Discontinue investigational drug (ID X93461) since study is complete

* Monitor appetite and seizure activity closely

* Consider alternative medications for epilepsy management if necessary

----------------END OF PATIENT ENCOUNTER---------------

-------------- PATIENT ID 239 -- VISIT 1 -----------

On August 29th, 2021, Patient ID 239, a 49-year-old female, presented to our clinic for her first visit. The patient has a history of seasonal allergies, ADHD, and epilepsy. She has experienced seven seizures in the past two months, which is a significant increase from her usual frequency. As part of her treatment plan, the patient is currently participating in a randomized controlled trial, and no changes to her medications will be made at this time beyond adding the investigational drug for epilepsy that she will take for three months.

During the neurological examination, the patient's vital signs were within normal limits. Her consciousness was clear, and her speech was coherent and appropriate to her level of education. The patient's memory was intact, and she was able to perform simple calculations accurately. The patient's sensory motor function was mostly intact, with some mild weakness in the left arm and leg. Her coordination and balance were also normal.

The patient's reflexes were normal, and there was no evidence of any abnormal movements or tremors. Her cranial nerves were intact, with no signs of dysfunction. The patient's funduscopic examination revealed a normal optic disc and macula. The rest of her neurological examination was unremarkable, including her mental status, language, and executive function.

Based on the patient's history and examination findings, it is apparent that her recent increase in seizures is a significant concern. However, as part of the randomized controlled trial, no changes to her medications will be made at this time beyond adding the investigational drug for epilepsy. We will closely monitor the patient's seizure activity and adjust her treatment plan as needed. The patient will follow up with us in one month for a subsequent visit.

----------------END OF PATIENT ENCOUNTER---------------

-------------- PATIENT ID 239 -- VISIT 2 -----------

---

Clinic Note #2 - Patient ID: 239 - Date: 2021-09-29

Identification:

Name: [Patient Name]

Date of Birth: [Date of Birth]

Gender: Female

Age: 49 years

HPI (History of Present Illness):

The patient presents with a history of seasonal allergies, ADHD, and epilepsy. She has been participating in a randomized controlled trial involving the investigational drug ID X93461 for her epilepsy over the past 3 months. During this time, she has experienced 19 seizures and has reported vertigo.

Subjective:

The patient reports worsening of her seasonal allergies and ADHD symptoms during the study period. She also complains of dizziness and vertigo, which began after initiating the investigational drug. The patient denies any recent travel or exposure to possible infections.

Medications:

The patient is currently taking the following medications:

1. Levodopa/Carbidopa (Sinemet) - 25/250 mg, tid

2. Lamotrigine (Lamictal) - 200 mg, qd

3. ID X93461 (investigational drug) - discontinued as of today

Allergies:

The patient has no known allergies.

Social History:

The patient is married with two children, ages 10 and 12. She works as a teacher and enjoys outdoor activities such as hiking and biking. The patient denies any history of substance abuse or smoking.

Review of Systems:

The patient reports no recent illnesses or surgeries. She does report worsening of her seasonal allergies, which have been affecting her sleep and causing nasal congestion. The patient also mentions that she has been experiencing dizziness and vertigo for the past few weeks.

General Physical Examination:

Vital signs:

* Temperature: 98.6°F (37°C)

* Blood Pressure: 120/80 mmHg

* Pulse: 80 beats per minute

* Respiratory Rate: 16 breaths per minute

Head and Neck:

* Normal head shape and symmetry

* No focal neurological deficits or abnormalities

* No hemiparesis or weakness observed

Extremities:

* Normal strength and tone of the upper and lower extremities

* No sensory deficits or abnormalities observed

Neurological Examination:

* Cranial nerves: Intact (II-XII)

* Motor examination: Normal strength and tone in all four limbs

* Sensory examination: Intact to pinprick, light touch, and temperature sensations in all four limbs

* Coordination and balance: Intact

Labs:

* Complete Blood Count (CBC) - normal

* Electrolyte Panel - normal

* Thyroid-stimulating Hormone (TSH) - normal

* Vitamin D Level - normal

Imaging:

* Computed Tomography (CT) scan of the brain - normal

Assessment:

The patient presents with a history of seasonal allergies, ADHD, and epilepsy. She has been participating in a randomized controlled trial involving the investigational drug ID X93461 for her epilepsy over the past 3 months. During this time, she has experienced 19 seizures and has reported vertigo. The patient's symptoms of dizziness and vertigo began after initiating the investigational drug, and she denies any recent travel or exposure to possible infections. The patient's neurological examination is normal, with no focal neurological deficits or abnormalities observed. Based on the patient's history and current symptoms, it is possible that the investigational drug may be contributing to her vertigo and dizziness.

Plan:

* Discontinue the investigational drug ID X93461

* Monitor the patient's symptoms closely and adjust medications as needed

* Consider a trial of vestibular rehabilitation therapy to address the patient's vertigo and dizziness

Signature: [Your Signature]

---

Note: This clinic note is fictional and for illustrative purposes only. It should not be used as a real clinical document without proper authorization and approval from the relevant healthcare professionals and institutions.

----------------END OF PATIENT ENCOUNTER---------------

### GROUND TRUTH – BOTH ARMS

ID	arm	Visit 1	Visit 2	Symptoms

0	0	12.0	10.0	somnolence, upper respiratory infection

1	0	8.0	42.0	somnolence

2	0	17.0	45.0	no symptoms

3	0	14.0	23.0	upper respiratory infection

4	0	13.0	30.0	no symptoms

5	0	113.0	132.0	balance disorder

6	0	59.0	216.0	no symptoms

7	0	5.0	12.0	no symptoms

8	0	3.0	13.0	no symptoms

9	0	5.0	13.0	fatigue

20	0	32.0	165.0	dizziness

21	0	10.0	17.0	no symptoms

22	0	21.0	29.0	no symptoms

23	0	6.0	65.0	upper respiratory infection

24	0	11.0	18.0	dizziness, upper respiratory infection

25	0	43.0	23.0	dizziness, somnolence

26	0	12.0	15.0	no symptoms

27	0	18.0	15.0	dizziness, upper respiratory infection

28	0	41.0	79.0	no symptoms

29	0	12.0	17.0	back pain

40	0	10.0	20.0	no symptoms

41	0	4.0	8.0	somnolence

42	0	12.0	20.0	fatigue, balance disorder

43	0	7.0	51.0	dizziness

44	0	16.0	18.0	dizziness, somnolence

45	0	12.0	27.0	no symptoms

46	0	10.0	14.0	no symptoms

47	0	9.0	9.0	dizziness

48	0	18.0	33.0	fatigue

49	0	19.0	27.0	somnolence, fatigue, upper respiratory infection

60	0	5.0	6.0	no symptoms

61	0	18.0	15.0	no symptoms

62	0	21.0	29.0	no symptoms

63	0	17.0	170.0	fall

64	0	17.0	30.0	no symptoms

65	0	5.0	16.0	ataxia, gait disturbance

66	0	23.0	39.0	no symptoms

67	0	49.0	43.0	dizziness

68	0	13.0	21.0	no symptoms

69	0	18.0	14.0	fall

80	0	17.0	46.0	no symptoms

81	0	94.0	54.0	vertigo, headaches

82	0	12.0	21.0	decreased appetite

83	0	9.0	128.0	no symptoms

84	0	58.0	33.0	no symptoms

85	0	55.0	89.0	no symptoms

86	0	4.0	42.0	no symptoms

87	0	10.0	16.0	no symptoms

88	0	14.0	16.0	no symptoms

89	0	3.0	16.0	no symptoms

100	0	32.0	60.0	somnolence, balance disorder

101	0	183.0	188.0	no symptoms

102	0	195.0	344.0	dizziness

103	0	9.0	16.0	no symptoms

104	0	5.0	9.0	balance disorder

105	0	18.0	20.0	no symptoms

106	0	13.0	22.0	balance disorder

107	0	12.0	33.0	headaches

108	0	58.0	26.0	no symptoms

109	0	15.0	4.0	no symptoms

120	0	2	12	fatigue, diplopia

121	0	17	16	upper respiratory infection

122	0	13	35	no symptoms

123	0	14	18	dizziness, balance disorder

124	0	31	128	dizziness, headaches

125	0	4	11	fall

126	0	46	47	no symptoms

127	0	4	10	no symptoms

128	0	13	18	no symptoms

129	0	24	42	vertigo

140	0	14	7	no symptoms

141	0	8	29	balance disorder

142	0	9	24	somnolence

143	0	15	22	balance disorder

144	0	17	31	no symptoms

145	0	12	10	no symptoms

146	0	27	26	upper respiratory infection

147	0	9	17	no symptoms

148	0	103	61	dizziness, fall

149	0	86	108	balance disorder

160	0	14	14	somnolence

161	0	42	76	gait disturbance

162	0	24	82	somnolence

163	0	32	81	no symptoms

164	0	3	12	no symptoms

165	0	49	38	no symptoms

166	0	15	37	dizziness

167	0	13	25	headaches

168	0	10	8	no symptoms

169	0	6	7	fatigue

180	0	12	18	no symptoms

181	0	13	29	no symptoms

182	0	11	8	gait disturbance

183	0	144	63	dizziness

184	0	9	24	dizziness

185	0	14	27	no symptoms

186	0	6	17	vertigo, fall, headaches

187	0	14	15	dizziness

188	0	58	43	somnolence

189	0	60	58	no symptoms

200	0	28	19	dizziness

201	0	24	35	no symptoms

202	0	5	3	no symptoms

203	0	15	64	no symptoms

204	0	5	8	no symptoms

205	0	70	47	fatigue, upper respiratory infection

206	0	2	9	dizziness, somnolence

207	0	34	118	no symptoms

208	0	7	14	nystagmus, upper respiratory infection

209	0	19	120	dizziness

220	0	9	17	no symptoms

221	0	27	15	somnolence

222	0	8	17	no symptoms

223	0	65	75	fatigue

224	0	82	141	no symptoms

225	0	1	11	fall

226	0	40	143	diplopia

227	0	61	66	no symptoms

228	0	18	50	no symptoms

229	0	8	26	no symptoms

10	1	43.0	23.0	fatigue

11	1	44.0	18.0	dizziness, ataxia, gait disturbance

12	1	6.0	3.0	ataxia

13	1	11.0	4.0	somnolence, fatigue, constipation, vomiting, fall

14	1	13.0	22.0	somnolence, fatigue, vomiting

15	1	43.0	76.0	dizziness, somnolence

16	1	14.0	8.0	balance disorder

17	1	221.0	128.0	back pain

18	1	11.0	9.0	dizziness, somnolence, fall, gait disturbance

19	1	21.0	19.0	fatigue, diplopia

30	1	6.0	6.0	dizziness, somnolence

31	1	8.0	7.0	dizziness

32	1	6.0	9.0	dizziness

33	1	22.0	12.0	dizziness, headaches, diplopia

34	1	22.0	12.0	dizziness, somnolence, dysarthria

35	1	14.0	10.0	somnolence, diplopia

36	1	27.0	14.0	no symptoms

37	1	33.0	29.0	dizziness, somnolence, vertigo

38	1	22.0	25.0	dizziness, ataxia

39	1	18.0	18.0	fatigue, balance disorder

50	1	28.0	23.0	dizziness, fatigue, nausea, balance disorder

51	1	48.0	155.0	decreased appetite

52	1	19.0	10.0	dizziness, gait disturbance

53	1	78.0	34.0	no symptoms

54	1	14.0	3.0	dizziness, constipation, nystagmus, headaches, gait disturbance

55	1	8.0	3.0	dysarthria, diplopia

56	1	5.0	11.0	dizziness, somnolence

57	1	28.0	17.0	somnolence, back pain

58	1	15.0	23.0	somnolence, balance disorder, back pain, gait disturbance

59	1	11.0	6.0	ataxia

70	1	74.0	25.0	somnolence

71	1	32.0	13.0	dizziness, somnolence

72	1	13.0	7.0	constipation

73	1	18.0	17.0	dizziness, fatigue, vomiting, headaches

74	1	11.0	9.0	no symptoms

75	1	12.0	5.0	somnolence

76	1	10.0	2.0	fatigue, headaches, diplopia

77	1	5.0	2.0	dizziness, gait disturbance

78	1	59.0	29.0	headaches

79	1	16.0	4.0	somnolence, headaches

90	1	13.0	7.0	ataxia

91	1	16.0	6.0	somnolence, fatigue, ataxia

92	1	9.0	8.0	dizziness

93	1	20.0	15.0	somnolence, fatigue

94	1	0.0	3.0	gait disturbance

95	1	14.0	13.0	no symptoms

96	1	59.0	25.0	no symptoms

97	1	17.0	12.0	balance disorder

98	1	7.0	3.0	dizziness, somnolence, fatigue, dysarthria, diplopia, gait disturbance

99	1	55.0	33.0	constipation, diplopia

110	1	69.0	30.0	somnolence, balance disorder

111	1	30.0	16.0	somnolence, vomiting

112	1	28.0	17.0	dizziness, nausea, ataxia, fall

113	1	5.0	3.0	dizziness, fall

114	1	6.0	12.0	dizziness, back pain, diplopia

115	1	13.0	14.0	diplopia, decreased appetite

116	1	16.0	18.0	somnolence, vomiting, diplopia

117	1	19.0	9.0	fatigue

118	1	14.0	11.0	somnolence, balance disorder, fall

119	1	16.0	4.0	dizziness, vomiting

130	1	13	9	fatigue, ataxia, gait disturbance

131	1	1	3	somnolence, fatigue, diplopia

132	1	22	17	somnolence, diplopia

133	1	5	11	dizziness, somnolence, constipation, ataxia

134	1	96	52	dizziness, constipation, ataxia, back pain

135	1	10	0	diplopia

136	1	22	16	somnolence, nausea, balance disorder

137	1	13	13	dizziness, somnolence, headaches

138	1	208	81	somnolence, fatigue, vomiting, diplopia

139	1	20	11	somnolence, nausea, fall

150	1	11	4	somnolence, dysarthria, headaches

151	1	32	65	dizziness, nystagmus, diplopia, gait disturbance

152	1	53	27	dizziness, diplopia

153	1	7	12	dizziness, somnolence, fatigue

154	1	16	18	dizziness, somnolence, dysarthria

155	1	89	89	gait disturbance

156	1	18	14	diplopia

157	1	22	18	somnolence

158	1	4	5	dizziness, somnolence, fatigue, balance disorder, vertigo

159	1	58	42	dizziness

170	1	8	12	somnolence, back pain, diplopia

171	1	7	1	somnolence, fatigue, diplopia

172	1	24	22	dizziness, somnolence, balance disorder, dysarthria, gait disturbance

173	1	18	18	dizziness, somnolence, fatigue

174	1	7	9	balance disorder

175	1	18	9	dizziness

176	1	8	16	diplopia

177	1	9	3	fatigue

178	1	44	13	dizziness, somnolence, fatigue, diplopia

179	1	113	98	no symptoms

190	1	10	17	no symptoms

191	1	15	2	somnolence, constipation

192	1	3	15	no symptoms

193	1	15	14	somnolence

194	1	12	8	somnolence, ataxia, gait disturbance

195	1	16	27	dizziness, fatigue, gait disturbance

196	1	4	6	somnolence

197	1	29	22	somnolence, constipation, back pain

198	1	8	15	dizziness

199	1	14	26	dizziness, somnolence, constipation, balance disorder, vertigo

210	1	3	8	somnolence, diplopia

211	1	22	16	dizziness, somnolence, vertigo

212	1	52	48	dizziness, fatigue

213	1	34	33	dizziness, fatigue, vomiting, upper respiratory infection, diplopia

214	1	10	3	dizziness, nausea

215	1	15	19	somnolence, fatigue, gait disturbance

216	1	15	13	dizziness, somnolence, back pain

217	1	18	21	somnolence, fatigue, nausea, dysarthria, upper respiratory infection, gait disturbance

218	1	13	7	dizziness, somnolence, nausea

219	1	44	18	no symptoms

230	1	13	5	balance disorder, headaches, decreased appetite

231	1	11	3	balance disorder

232	1	124	133	dizziness, somnolence

233	1	11	16	dizziness

234	1	5	17	nausea

235	1	7	3	somnolence

236	1	8	15	nausea, diplopia

237	1	9	11	no symptoms

238	1	7	4	decreased appetite

239	1	7	19	vertigo

### MD SUMMARY

| **ID** | **arm** | **Visit 1** | **Visit 2** | **Symptoms** |
| --- | --- | --- | --- | --- |
| 0 | 0 | 12 | 10 | somnolence, URI |
| 1 | 0 | 8 | 42 | somnolence |
| 2 | 0 | 17 | 0 |  |
| 3 | 0 | 14 | 23 | URI |
| 4 | 0 | 13 | 30 | none |
| 5 | 0 | 113 | 132 | balance disorder |
| 6 | 0 | 59 | 216 | mild left hemiparesis, ataxia |
| 7 | 0 | 5 | 12 |  |
| 8 | 0 | 3 | 13 |  |
| 9 | 0 | 5 | 13 | fatigue |
| 20 | 0 | 32 | 165 | dizziness |
| 21 | 0 | 10 | 17 |  |
| 22 | 0 | 21 | 29 |  |
| 23 | 0 | 6 | 65 | URI |
| 24 | 0 | NA | 18 | dizziness, URI |
| 25 | 0 | 43 | 23 | dizziness, somnolence |
| 26 | 0 | 12 | 15 |  |
| 27 | 0 | 18 | 15 | dizziness, URI |
| 28 | 0 | 41 | 79 |  |
| 29 | 0 | 12 | 17 | back pain |
| 30 | 0 | 10 | 20 |  |
| 41 | 0 | 4 | NA | somnolence |
| 42 | 0 | 12 | 20 | fatigue, balance disorder |
| 43 | 0 | 7 | 51 | dizziness |
| 44 | 0 | 16 | 18 | dizziness, somnolence |
| 45 | 0 | 12 | NA |  |
| 46 | 0 | 10 | 14 |  |
| 47 | 0 | 9 | 9 | dizziness |
| 48 | 0 | 18 | 33 | fatigue |
| 49 | 0 | 19 | 27 | somnolence, fatigue, URI |
| 60 | 0 | 5 | 6 |  |
| 61 | 0 | 18 | 15 |  |
| 62 | 0 | 21 | 29 |  |
| 63 | 0 | 17 | 170 | fall |
| 64 | 0 | 17 | 30 |  |
| 65 | 0 | 5 | 16 | ataxia, gait disturbance |
| 66 | 0 | 23 | NA |  |
| 67 | 0 | 49 | 43 | dizziness |
| 68 | 0 | 13 | 21 |  |
| 69 | 0 | 18 | 14 | fall |
| 80 | 0 | 17 | 46 |  |
| 81 | 0 | 94 | 54 | vertigo, headaches |
| 82 | 0 | 12 | NA | decreased appetite |
| 83 | 0 | 9 | 128 |  |
| 84 | 0 | 58 | 33 |  |
| 85 | 0 | 55 | 89 |  |
| 86 | 0 | 4 | 42 |  |
| 87 | 0 | 10 | 16 |  |
| 88 | 0 | 14 | 16 |  |
| 89 | 0 | 3 | 16 |  |
| 100 | 0 | 32 | 60 | somnolence, balance disorder |
| 101 | 0 | 183 | NA |  |
| 102 | 0 | 195 | 344 | dizziness |
| 103 | 0 | 9 | 16 |  |
| 104 | 0 | 5 | 9 | dizziness, balance disorder |
| 105 | 0 | 18 |  |  |
| 106 | 0 | 13 | 22 | balance disorder |
| 107 | 0 | NA | 33 | headaches |
| 108 | 0 | 58 | 26 |  |
| 109 | 0 | 15 | NA |  |
| 120 | 0 | 2 | 12 | fatigue, diplopia |
| 121 | 0 | 17 | 16 | URI |
| 122 | 0 | 13 | 14 |  |
| 123 | 0 | 14 | 18 | dizziness, balance disorder |
| 124 | 0 | 31 | 128 | dizziness, headache |
| 125 | 0 | 4 | 11 | fall |
| 126 | 0 | 2.50 | 47 |  |
| 127 | 0 | 4 | 10 |  |
| 128 | 0 | 13 | 18 |  |
| 129 | 0 | 24 | 42 | vertigo |
| 140 | 0 | 14 | 7 |  |
| 141 | 0 | 8 | 29 | balance disorder |
| 142 | 0 | 9 | 24 | somnolence |
| 143 | 0 | 15 | 22 | balance disorder |
| 144 | 0 | 17 | 31 |  |
| 145 | 0 | 12 | 10 |  |
| 146 | 0 | 27 | 26 | URI |
| 147 | 0 | 9 | 17 |  |
| 148 | 0 | 103 | 61 | dizziness, falls |
| 149 | 0 | 86 | 108 | balance difficulties |
| 160 | 0 | 14 | 14 | somnolence |
| 161 | 0 | 42 | 76 | gait disturbance |
| 162 | 0 | 24 | 82 | somnolence |
| 163 | 0 | 32 | 81 |  |
| 164 | 0 | 3 | 12 |  |
| 165 | 0 | 49 | 38 |  |
| 166 | 0 | 15 | 37 | dizziness |
| 167 | 0 | 13 | 25 | frequent headaches |
| 168 | 0 | 10 | 8 |  |
| 169 | 0 | 6 | 7 | fatigue |
| 180 | 0 | 12 | 0 |  |
| 181 | 0 | 13 | 29 |  |
| 182 | 0 | 11 | 8 | gait disturbance |
| 183 | 0 | 144 | 63 | dizziness |
| 184 | 0 | 9 | 24 | dizziness |
| 185 | 0 | 14 | 27 |  |
| 186 | 0 | 6 | 17 | vertigo, falling, headache |
| 187 | 0 | 14 | 15 | dizziness |
| 188 | 0 | 58 | 43 | somnolence, lethargy |
| 189 | 0 | 60 | NA |  |
| 200 | 0 | 28 | 19 | dizziness |
| 201 | 0 | 24 | 35 |  |
| 202 | 0 | 5 | 3 |  |
| 203 | 0 | 15 | 64 |  |
| 204 | 0 | 5 | 8 |  |
| 205 | 0 | 70 | 47 | fatigue, URI |
| 206 | 0 | 2 | 9 | dizziness, somnolence |
| 207 | 0 | 34 | 118 |  |
| 208 | 0 | 7 | 14 | nystagmus, |
| 209 | 0 | 19 | 120 | dizziness |
| 220 | 0 | 9 | 17 |  |
| 221 | 0 | 27 | 15 | somnolence |
| 222 | 0 | 8 | 17 |  |
| 223 | 0 | 65 | 75 | fatigue |
| 224 | 0 | 82 | 141 |  |
| 225 | 0 | 1 | 11 | fall |
| 226 | 0 | 40 | 143 | diplopia |
| 227 | 0 | 61 | 66 |  |
| 228 | 0 | 18 | 50 |  |
| 229 | 0 | 8 | 26 | elevated TSH |
| 10 | 1 | 43 | 23 | fatigue |
| 11 | 1 | 44 | NA | gait disturbance, ataxia, dizziness |
| 12 | 1 | 6 | NA | ataxia |
| 13 | 1 | 11 | 4 | somnolence, fatigue, constipation, vomiting, fall |
| 14 | 1 | 13 | NA | somnolence, fatigue, vomiting |
| 15 | 1 | 43 | 76 | dizziness, somnolence |
| 16 | 1 | 14 | 8 | balance disorder |
| 17 | 1 | 221 | 128 | back pain, on exam had new left arm and leg mild weakness, hyperreflexia, sensory deficits in L2,L3,L4 dermatomes, fine motor skills difficulty, |
| 18 | 1 | 11 | 9 | dizziness, somnolence, fall, gait disturbance |
| 19 | 1 | 21 | 19 | fatigue and diplopia |
| 20 | 1 | 6 | 6 | dizziness and somnolence |
| 31 | 1 | 8 | 7 | dizziness |
| 32 | 1 | 6 | 9 | dizziness |
| 33 | 1 | 22 | 12 | dizziness, headache, diplopia |
| 34 | 1 | 22 | 12 | dizziness, somnolence, dysarthria, on exam new decreased sensation on R side of the body |
| 35 | 1 | 14 | 10 | somnolence, diplopia |
| 36 | 1 | 27 | 14 |  |
| 37 | 1 | 33 | 29 | dizziness, somnolence, vertigo, on exam new tremors in hands and feet, slow reaction time |
| 38 | 1 | 22 | 25 | dizziness, ataxia, on exam new hyperreflexia |
| 39 | 1 | 18 | 18 | fatigue, balance disorder, cognitive decline, Cr 1.5 on labs |
| 50 | 1 | 28 | 23 | dizziness, fatigue, nausea, balance disorder |
| 51 | 1 | 48 | 155 | decreased appetite, on exam new tremor in hands |
| 52 | 1 | 19 | 10 | dizziness, gait disturbance |
| 53 | 1 | 78 | 34 |  |
| 54 | 1 | 14 | 3 | dizziness, constipation, headaches, gait disturbance |
| 55 | 1 | 8 | 3 | dysarthria, slow eye movements, diplopia |
| 56 | 1 | 5 | 11 | dizziness, somnolence |
| 57 | 1 | 28 | 17 | backpain, somnolence |
| 58 | 1 | 15 | 23 | somnolence, back pain, balance disorder, gait disturbance |
| 59 | 1 | 11 | 6 | gait/balance diorder, tremors |
| 70 | 1 | 74 | 25 | somnolence, mild tremor, rigidity |
| 71 | 1 | 32 | 13 | dizziness, somnolence |
| 72 | 1 | 13 | 7 | constipation |
| 73 | 1 | 18 | 17 | dizziness, fatigue, vomiting, headaches |
| 74 | 1 | 11 | 9 |  |
| 75 | 1 | 12 | 5 | somnolence, cognitive decline, difficulty concentrating |
| 76 | 1 | 10 | 2 | fatigue, headache, diplopia |
| 77 | 1 | NA | 2 | dizziness, gait disturbance |
| 78 | 1 | 59 | 29 | headaches |
| 79 | 1 | 16 | 12 | somnolence, headache |
| 90 | 1 | 13 | 7 | ataxia |
| 91 | 1 | 16 | 6 | ataxia, somnolence, fatigue |
| 92 | 1 | 9 | 8 | dizziness |
| 93 | 1 | 20 | 15 | somnolence, fatigue |
| 94 | 1 | 0 | 3 | gait disturbance, left facial weakness, ataxia |
| 95 | 1 | 14 | 13 |  |
| 96 | 1 | 59 | 25 |  |
| 97 | 1 | 17 | 12 | balance disorder, weakness, sesnroy deficit in left arm and leg, increased muslce tone on exam |
| 98 | 1 | 7 | 3 | dizziness, somnolence, fatigue, dysarthria, diplopia, gait disturbance, weakness in left arm and leg, dysarthria |
| 99 | 1 | 55 | 33 | constipation, diplopia |
| 110 | 1 | 69 | 30 | somnolence, balance disorder |
| 111 | 1 | 30 | 16 | somnolence, vomiting |
| 112 | 1 | 28 | 17 | dizziness, nausea, ataxia, falls |
| 113 | 1 | 5 | 9 | dizziness, fall |
| 114 | 1 | 6 | 12 | dizziness, backpain, diplopia |
| 115 | 1 | 13 | 14 | diplopia, decreased appetite, fatigue, |
| 116 | 1 | 16 | 18 | somnolence, vomiting, diplopia |
| 117 | 1 | 19 | 0 | fatigue |
| 118 | 1 | 14 | 11 | somnolence, balance disorder, fall, dizziness, headaches |
| 119 | 1 | 16 | 4 | dizziness, vomiting |
| 130 | 1 | 13 | 9 | fatigue, ataxia, gat disturbance |
| 131 | 1 | 1 | 3 | somnolence, fatigue, diplopia |
| 132 | 1 | 22 | 17 | somnolence, diplopia, hyperreflexia, papilledema, hypothermia |
| 133 | 1 | 5 | 11 | dizziness, somnolence, constipation, ataxia |
| 134 | 1 | 96 | 52 | dizziness, constipation, ataxia, backpain |
| 135 | 1 | 10 | 0 | diplopia |
| 136 | 1 | 22 | 16 | somnolence, nausea, balance disorder |
| 137 | 1 | 13 | 13 | dizziness, somnolence, headache |
| 138 | 1 | 208 | 81 | somnolence, fatigue, vomiting, diplopia |
| 139 | 1 | 20 | 11 | somnolence, nausea, fall |
| 150 | 1 | 11 | NA | somnolence, dysarthria, headache |
| 151 | 1 | 32 | 65 | dizziness, nystagmus, diplopia, gait disturbance |
| 152 | 1 | 53 | 27 | dizziness, diplopia |
| 153 | 1 | 7 | 12 | dizziness, somnolence, fatigue, dysarthria, diplopia, gait disturbance, weakness in left arm and leg, dysarthria |
| 154 | 1 | 16 | 18 | dizziness, somnolence, dysarthria, decreased mobility, tremor, decreased sensation in left arm and leg, hyperreflexia, ataxia |
| 155 | 1 | 89 | 89 | gait disturbance, hyperreflexia, |
| 156 | 1 | 18 | 14 | diplopia |
| 157 | 1 | 22 | 18 | somnolence, fatigue, cognitive decline, diffculty concentration and attention, weakness in right limbs, ataxia, gait disturbance. |
| 158 | 1 | 4 | 5 | dizziness, somnolence, fatigue, balance disorder, vertigo. |
| 159 | 1 | 58 | 42 | dizziness |
| 170 | 1 | 8 | 12 | somnolence, back pain, diplopia |
| 171 | 1 | 7 | 1 | somnolence, fatigue, diplopia |
| 172 | 1 | 24 | 22 | dizziness, somnolence, balance disorder, dysarthria, gait disturbance |
| 173 | 1 | 18 | 18 | dizziness, somnolence, fatigue |
| 174 | 1 | 7 | 9 | balance disorder |
| 175 | 1 | 18 | 9 | dizziness |
| 176 | 1 | 8 | 16 | diplopia |
| 177 | 1 | 9 | 3 | fatigue |
| 178 | 1 | 44 | 13 | dizziness, somnolence, fatigue, diplopia |
| 179 | 1 | 113 | 98 |  |
| 190 | 1 | 10 | 51 |  |
| 191 | 1 | 15 | NA | somnolence, constipation |
| 192 | 1 | 3 | 15 |  |
| 193 | 1 | 15 | 14 | somnolence |
| 194 | 1 | 12 | 8 | somnolence, ataxia, gait disturbance |
| 195 | 1 | 16 | 27 | dizziness, fatigue, gait disturbance |
| 196 | 1 | 4 | 6 | somnolence, |
| 197 | 1 | 29 | 22 | somnolence, constipation, bacl pain |
| 198 | 1 | 8 | 15 | dizziness |
| 199 | 1 | 14 | 26 | dizziness, somnolence, constipation, balance disorder, vertigo |
| 210 | 1 | 3 | 8 | somnolence, diplopia |
| 211 | 1 | 22 | 16 | dizziness, somnolence, vertigo |
| 212 | 1 | 52 | 48 | dizziness, fatigue |
| 213 | 1 | 34 | NA | dizziness, fatigue, vomiting, URI, diplopia |
| 214 | 1 | 10 | 3 | dizziness, nausea |
| 215 | 1 | 15 | 19 | somnolence, fatigue, gait disturbance, slow speech, dysarthria, drropy eyelids, neck rigid, arms and legs limp |
| 216 | 1 | 15 | 13 | dizziness, somnolence, backpain |
| 217 | 1 | 18 | 21 | somnolence, fatugue, nausea, dysarthria URI, gait distubrance |
| 218 | 1 | 13 | 7 | dizziness, somnolence, nausea |
| 219 | 1 | 44 | 18 |  |
| 230 | 1 | 13 | 5 | fatigue, balance disorder, hyporeflexiacognitive decline, |
| 231 | 1 | 11 | 3 | balance disorder |
| 232 | 1 | 124 | 133 | dizziness, somnolence |
| 233 | 1 | 11 | 16 | dizziness |
| 234 | 1 | 5 | 17 | nausea |
| 235 | 1 | 7 | 3 | somnoelnce |
| 236 | 1 | 8 | NA | nausea, diplopia |
| 237 | 1 | 9 | 11 |  |
| 238 | 1 | 7 | 4 | decreased appetite |
| 239 | 1 | 7 | 19 | dizziness, vertigo |

### AI Summary

ID	arm	baseline_sz	test_sz	sx

0	0	12	10	"somnolence, upper respiratory infection"

1	0	8	42	somnolence

2	0	17	0	None

3	0	14	23	"mild cough, nasal congestion"

4	0	13	30	None

5	0	113	132	Balance disorder

6	0	59	216	None

7	0	5	12	None

8	0	3	13	None

9	0	5	13	Fatigue

20	0	32	165	dizziness

21	0	10	17	None

22	0	21	29	None

23	0	6	65	None

24	0	Not specified	18	"dizziness, upper respiratory infection"

25	0	43	23	"dizziness, somnolence"

26	0	12	15	None

27	0	18	15	"dizziness, upper respiratory infection"

28	0	5	79	None

29	0	12	17	"back pain, seizures (grand mal in nature), increased anxiety and depression symptoms"

40	0	10	20	None

41	0	4	0	somnolence

42	0	12	20	"fatigue, balance disorder"

43	0	7	51	dizziness

44	0	16	18	"dizziness, somnolence"

45	0	12	0	None

46	0	10	14	None

47	0	9	9	dizziness

48	0	18	33	"Fatigue, increased seizure frequency"

49	0	19	27	"somnolence, fatigue, upper respiratory infection"

60	0	5	6	None

61	0	18	15	None

62	0	21	29	None

63	0	17	170	fall

64	0	17	30	"anxiety, depression, memory difficulty"

65	0	5	16	"ataxia, gait disturbance, ADHD, anxiety, temporal lobe epilepsy"

66	0	23	0	None

67	0	49	43	dizziness

68	0	13	21	None

69	0	18	14	fall

80	0	17	46	None

81	0	94	54	"vertigo, headaches"

82	0	12	0	"decreased appetite, depression, hypertension, generalized epilepsy"

83	0	9	128	None

84	0	58	33	None

85	0	55	89	None

86	0	4	42	"Autism, Hypertension, Multifocal Epilepsy"

87	0	10	16	None

88	0	14	16	None

89	0	3	16	None

100	0	32	60	"somnolence, balance disorder"

101	0	183	0	"dermatitis, epilepsy"

102	0	195	344	dizziness

103	0	9	16	None

104	0	5	9	"balance disorder, increased frequency of seizures, fatigue, nausea, difficulty with concentration, trouble sleeping at night"

105	0	18	0	None

106	0	13	22	"balance disorder, seizures"

107	0	8	33	headaches

108	0	58	26	None

109	0	15	0	None

120	0	2	12	"fatigue, diplopia"

121	0	17	16	None

122	0	13	0	None

123	0	14	18	"dizziness, balance disorder"

124	0	31	128	"dizziness, headaches"

125	0	4	11	"mild tremors, slowed reaction time, slurred speech, difficulty with concentration and memory, fall"

126	0	46	47	None

127	0	4	10	"Anxiety, Depression"

128	0	13	18	None

129	0	24	42	"vertigo, multifocal epilepsy, frequent headaches, difficulty with memory and concentration, slight limb weakness on the right side of her body, decreased sensation to light touch and pinprick on the left side of her body"

140	0	14	7	"anxiety, depression"

141	0	8	29	"balance disorder, hypothyroidism, asthma, frontal lobe epilepsy"

142	0	9	24	"somnolence, history of seizures"

143	0	15	22	balance disorder

144	0	17	31	None

145	0	12	10	None

146	0	27	26	Upper respiratory infection

147	0	9	17	None

148	0	103	61	"dizziness, fall"

149	0	86	108	"balance disorder, ""brain fog"", persistent seizures"

160	0	14	14	somnolence

161	0	42	76	"gait disturbance, mild tremor in right hand, slightly weaker grip strength on right side, wide-based, unsteady gait with slight limp, difficulty maintaining balance and requires assistance with walking"

162	0	24	82	"somnolence, depression, dermatitis"

163	0	32	81	None

164	0	3	12	None

165	0	49	38	None

166	0	15	37	dizziness

167	0	13	25	headaches

168	0	10	8	None

169	0	6	7	fatigue

180	0	12	0	None

181	0	13	29	None

182	0	11	8	"gait disturbance, difficulty walking short distances without assistance"

183	0	144	63	dizziness

184	0	9	24	"dizziness, vertigo"

185	0	14	27	None

186	0	6	17	"vertigo, falling, headaches"

187	0	14	15	dizziness

188	0	58	43	"somnolence, lethargy, progressive memory loss, difficulty concentrating"

189	0	60	Not specified	seizures

200	0	28	19	mild dizziness

201	0	24	35	None

202	0	5	3	None

203	0	15	64	None

204	0	5	8	None

205	0	70	47	"fatigue, upper respiratory infection"

206	0	2	9	"dizziness, somnolence"

207	0	34	118	None

208	0	7	14	nystagmus

209	0	19	120	dizziness

220	0	9	17	None

221	0	27	15	somnolence

222	0	8	17	None

223	0	65	75	"anxiety, fatigue"

224	0	82	141	None

225	0	1	11	"diabetes, dermatitis, occipital lobe epilepsy, recent fall"

226	0	40	143	diplopia

227	0	61	66	None

228	0	18	50	None

229	0	8	26	None

10	1	43	23	fatigue

11	1	44	0	"dizziness, ataxia, gait disturbance"

12	1	6	0	"ataxia, anxiety, parietal lobe epilepsy"

13	1	11	4	"somnolence, fatigue, constipation, vomiting, fall"

14	1	13	0	"somnolence, fatigue, vomiting"

15	1	43	76	"dizziness, somnolence"

16	1	14	8	"balance disorder, recent seizure activity"

17	1	221	128	"back pain, mild weakness in left arm and leg, hyperreflexia, sensory deficits in L2, L3, and L4 dermatomes, difficulty with fine motor skills"

18	1	11	9	"dizziness, somnolence, fall, gait disturbance"

19	1	21	19	"fatigue, diplopia (double vision)"

30	1	6	6	"seasonal allergies, dermatitis, autism, parietal lobe epilepsy"

31	1	8	7	dizziness

32	1	6	9	"Dizziness, increased seizure frequency while taking investigational drug"

33	1	22	12	"dizziness, headaches, diplopia"

34	1	22	12	"dizziness, somnolence, dysarthria"

35	1	14	10	"somnolence, diplopia"

36	1	27	14	None

37	1	33	0	"dizziness, somnolence, vertigo"

38	1	22	25	"Dizziness, Ataxia"

39	1	18	18	"fatigue, balance difficulties, difficulty sleeping"

50	1	28	23	"dizziness, fatigue, nausea, and balance disorder"

51	1	48	155	decreased appetite and ongoing seizure activity

52	1	19	10	"dizziness, gait disturbance"

53	1	78	34	"generalized epilepsy, alcoholism, hypothyroidism"

54	1	14	3	"dizziness, constipation, nystagmus, headaches, gait disturbance"

55	1	8	3	"dysarthria, diplopia"

56	1	5	11	"dizziness, somnolence"

57	1	28	17	"somnolence, back pain"

58	1	15	23	"somnolence, balance disorder, back pain, gait disturbance"

59	1	11	6	ataxia

70	1	74	25	"somnolence, mild tremors, rigidity"

71	1	32	13	"dizziness, somnolence"

72	1	13	7	constipation

73	1	18	17	"dizziness, fatigue, vomiting, headaches"

74	1	11	9	None

75	1	12	5	"somnolence, decreased cognitive function, difficulty concentrating, memory issues"

76	1	10	2	"fatigue, headaches, diplopia"

77	1	Not specified	2	"dizziness, gait disturbance"

78	1	59	29	headaches

79	1	16	4	"somnolence, headaches, increased seizure activity"

90	1	13	7	"ataxia, alcoholism, multifocal epilepsy"

91	1	16	6	"somnolence, fatigue, ataxia"

92	1	9	8	dizziness

93	1	20	15	"somnolence, fatigue"

94	1	0	3	"gait disturbance, mild weakness of the left facial muscles"

95	1	14	13	None

96	1	59	25	None

97	1	17	12	"balance disorder, mild weakness in left arm and leg, sensory deficits in the form of hypesthesia (decreased sensation), negative Romberg test (indicates unsteadiness), positive Hoffmann's reflex (indicates increased muscle tone)"

98	1	7	3	"dizziness, somnolence, fatigue, dysarthria, diplopia, gait disturbance"

99	1	55	33	"constipation, diplopia"

110	1	69	30	"somnolence, balance disorder"

111	1	30	16	"somnolence, vomiting"

112	1	28	17	"dizziness, nausea, ataxia, falling on multiple occasions"

113	1	5	3	"difficulty with memory and concentration, feeling ""spacey"" at times, experiencing visual disturbances"

114	1	6	12	"dizziness, back pain, diplopia"

115	1	13	14	"diplopia, decreased appetite"

116	1	16	18	"somnolence, vomiting, diplopia"

117	1	19	0	"fatigue, sleep disturbances"

118	1	14	11	"somnolence, balance disorder, fall, occasional headaches, dizziness"

119	1	16	4	"dizziness and vomiting during these seizures, ongoing symptoms of depression, asthma, and epilepsy"

130	1	13	9	"fatigue, ataxia, gait disturbance"

131	1	1	3	"seasonal allergies, depression, diplopia"

132	1	22	17	"somnolence, diplopia"

133	1	5	11	"dizziness, somnolence, constipation, ataxia"

134	1	96	52	"dizziness, constipation, ataxia, back pain"

135	1	10	0	diplopia

136	1	22	16	"somnolence, nausea, balance disorder"

137	1	13	13	"dizziness, somnolence, headaches"

138	1	208	81	"somnolence, fatigue, vomiting, diplopia"

139	1	20	11	"somnolence, nausea, fall, difficulty concentrating, memory issues."

150	1	11	0	"somnolence, dysarthria, headaches"

151	1	32	65	"dizziness, nystagmus, diplopia, gait disturbance"

152	1	53	27	"dizziness, diplopia"

153	1	7	12	"dizziness, somnolence, fatigue"

154	1	16	18	"dizziness, somnolence, dysarthria"

155	1	89	89	Gait disturbance

156	1	18	14	Diplopia (new)

157	1	22	18	"increased somnolence, ataxia"

158	1	4	5	"dizziness, somnolence, fatigue, balance disorder, vertigo"

159	1	58	42	Dizziness

170	1	8	12	"somnolence, diplopia"

171	1	7	1	"somnolence, fatigue, diplopia"

172	1	24	22	"dizziness, somnolence, balance disorder, dysarthria, gait disturbance"

173	1	18	18	"dizziness, somnolence, fatigue"

174	1	7	9	"balance disorder, recent seizures"

175	1	18	9	"dizziness, occasional seizures"

176	1	8	16	diplopia

177	1	9	3	"fatigue, seizures"

178	1	44	13	"dizziness, somnolence, fatigue, diplopia"

179	1	113	98	none reported

190	1	10	17	None reported

191	1	15	0	"somnolence, constipation"

192	1	3	15	None reported

193	1	15	14	somnolence

194	1	12	8	"somnolence, ataxia, gait disturbance"

195	1	16	27	"dizziness, fatigue, gait disturbance"

196	1	4	6	"somnolence, seizures"

197	1	29	22	"somnolence, constipation, back pain"

198	1	8	15	dizziness

199	1	14	26	"dizziness, somnolence, constipation, balance disorder, vertigo"

210	1	3	8	"somnolence, diplopia"

211	1	22	16	"dizziness, somnolence, vertigo"

212	1	52	48	"dizziness, fatigue"

213	1	34	0	"dizziness, fatigue, vomiting, upper respiratory infection, diplopia"

214	1	10	3	"dizziness, nausea"

215	1	15	19	"somnolent, fatigued, gait disturbance, slow and slurred speech, droopy eyes, rigid neck, limp arms and legs, decreased muscle tone, diminished reflexes, difficulty with rapid alternating movements, compromised coordination and balance."

216	1	15	13	"dizziness, somnolence, back pain"

217	1	18	21	"somnolence, fatigue, nausea, dysarthria, upper respiratory infection, gait disturbance"

218	1	13	7	"dizziness, somnolence, nausea"

219	1	44	18	"schizophrenia, frontal lobe epilepsy"

230	1	13	5	"balance disorders, headaches, decreased appetite"

231	1	11	3	"balance disorder, history of occipital lobe epilepsy"

232	1	124	133	"dizziness, somnolence"

233	1	11	16	dizziness

234	1	5	17	mild nausea

235	1	7	3	somnolence reported by the patient

236	1	8	15	"nausea, diplopia"

237	1	9	11	no reported symptoms.

238	1	7	4	decreased appetite

239	1	7	19	"vertigo, worsening of seasonal allergies and ADHD symptoms, dizziness"

### AI PROMPT

The following prompt was used several times to generate the AI synthesized data table (LLM C in Figure 2).

Here are a collection of clinic visits from many patients. Each patient was seen twice, Visit 1 was after 2 months, and Visit 2 was after 3 additional months. Each visit includes a number of seizures documented. Please generate a data table with the following columns: patient ID = ID number of the patient, visit0_sz= number of seizures leading up to visit 0, visit_1_sz = number of seizures leading up to Visit 1, symptoms= any symptoms reported in visit 1 that were different from symptoms reported in visit 0.
